# Integrative GWAS Identifies Novel Loci and Genetic Links Between Psychiatric and Metabolic Factors in Anorexia Nervosa

**DOI:** 10.1101/2025.05.11.25327401

**Authors:** Yingchao Song, Yue Jiang, Joseph Glessner, Hakon Hakonarson, Xiao Chang

**Affiliations:** College of Medical Information and Artificial Intelligence, Shandong First Medical University, Shandong, China; Center for Applied Genomics, The Children’s Hospital of Philadelphia, Pennsylvania, United States

## Abstract

Anorexia nervosa (AN) is a complex psychiatric disorder with both psychiatric and metabolic underpinnings. To explore the genetic architecture of AN, we conducted a meta-analysis of AN GWAS data from European and Finnish populations, identifying a novel genome-wide significant locus near the *SOX5* gene. We further integrated GWAS summary statistics from the UK Biobank and FinnGen, focusing on psychiatric traits, metabolic markers, and lifestyle factors. Using cross-trait analysis methods including linkage disequilibrium score regression, Mendelian randomization and multi-trait analysis of GWAS, we revealed the causal relationships between AN and its risk factors, as well as the pleiotropic loci and shared genetic mechanisms. Gene Co-expression Network Analysis further uncovered key gene modules with both metabolic and neurological functions. These findings illustrate the interplay between psychiatric and metabolic pathways, highlighting their collective importance in shaping the genetic architecture of AN.

## Introduction

Anorexia nervosa (AN) is a severe and potentially life-threatening psychiatric disorder characterized by an excessive preoccupation with body shape and weight, accompanied by abnormal eating behaviors. Patients with AN often engage in self-imposed starvation, leading to significant emaciation^1–3^. This disorder typically begins in early adolescence and exhibits a pronounced difference in prevalence between males and females, with significantly higher rates in females ^4,5^. Since the pathophysiology of AN and the biological mechanisms driving its potentially lethal self-imposed food restrictions remain unclear, further characterization of its pathophysiology is crucial.

Empirical studies indicate that genetic factors play a significant role in AN, with an estimated heritability of 50%–60%^6^. However, pinpointing the precise genetic sources of risk for AN has proven difficult. Challenges in recruiting participants for AN research have limited the scope of previous transcriptomic studies^7^, whole exome sequencing^8^, and genome-wide association studies (GWAS)^3,9–11^, preventing definitive insights into the genetic architecture of AN. Recently, a comprehensive analysis of all available genotyped AN data worldwide (encompassing 16,992 cases and 55,525 controls) led to a discovery of eight loci that reach genome-wide significance^12^. Nevertheless, because the specific loci and genes influencing anorexia nervosa remain largely unknown, ongoing research is crucial to further elucidate the genetic underpinnings of this complex disorder.

The advances of cross-trait analysis of GWAS has provided a robust approach for exploring the genetic overlap and shared causality among complex traits^13^. By performing meta-analyses of GWAS summary statistics for genetically correlated traits, this method improves statistical power and identifies pleiotropic genetic variants that affect related traits. Additionally, it elucidates complex genetic interactions among phenotypes and uncovers the common biological pathways underlying their development. Such cross-trait methods have been effectively applied in the study of AN. For instance, previous studies utilizing conditional/conjunctional false discovery rate (condFDR/conjFDR) analyses have identified numerous pleiotropic genetic loci associated with AN and psychiatric disorders such as schizophrenia (SCZ), bipolar disorder (BIP), and major depression, demonstrating shared genetic structures between AN and other psychiatric conditions^14,15^. Despite these advancements, current cross-trait analyses of anorexia nervosa predominantly focus on psychiatric disorders. Given the complex metabo-psychiatric origins of AN, it is crucial to account for metabolic factors in addition to psychiatric disorders, including metabolic profiles, sex hormones, and lifestyle habits.^12^ Furthermore, the significant gender disparity in AN prevalence, with rates ranging from 0.9% to 4% in females compared to 0.3% in males^16–18^, underscores the need for more nuanced research. Recent increases in the availability of sex-specific GWAS summary data present new opportunities to investigate sex-specific pathogenic risk factors and pleiotropic genetic mechanisms associated with AN.

In this study, we initially performed a meta-analysis of AN GWAS data from European and Finnish cohorts. Subsequently, we integrated a broad range of risk factors from the UK Biobank (UKBB) and FINNGEN, encompassing psychiatric traits, metabolic markers, sex hormones, and lifestyle variables, to conduct an extensive cross-trait analysis. Leveraging approaches including linkage disequilibrium score regression (LDSC), Mendelian randomization (MR), heritability estimation (HESS), and multi-trait analysis of GWAS (MTAG), we identified pleiotropic genetic loci associated with AN and explored their causal relationships. Finally, we applied Weighted Gene Co-expression Network Analysis (WGCNA) to investigate key gene modules implicated in AN, providing deeper insights into the genetic underpinnings of AN.

## Results

### Novel Loci Identified in Anorexia Nervosa Meta-Analysis

We conducted a meta-analysis by integrating summary statistics from the largest available GWAS of AN in European and Finnish populations, identifying three genome-wide significant loci (*P* < 5×10^−8^). Two loci, 11q14.2 (rs10790068) and 3q13.11 (rs9874089), had been previously reported in European cohorts (Table 1). The lead variant rs10771043, situated in an intergenic region near *SOX5*, represents a novel locus associated with AN, marking the first report of its genome-wide significance (*P* < 5×10^−8^) in AN.

**Table 1.**
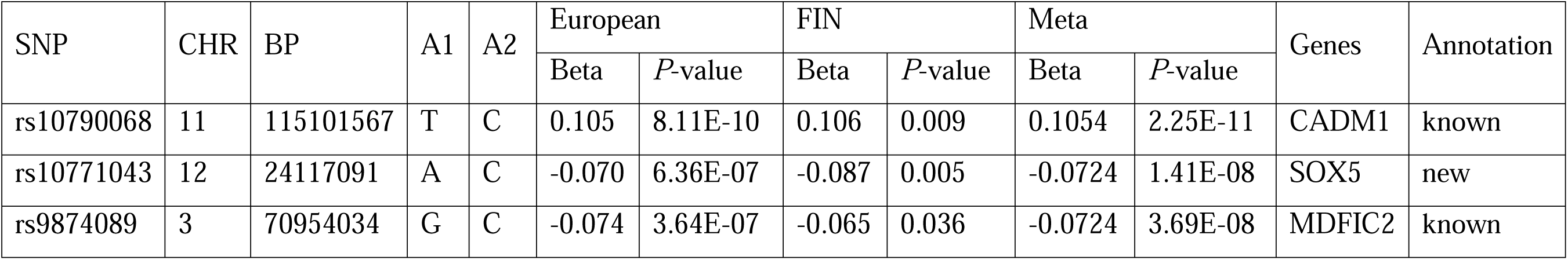
Meta-analysis of anorexia nervosa between European and Finnish populations.

### Global Genetic Correlation between Anorexia Nervosa and related Phenotypes

To elucidate the global genetic correlations between AN and its potential risk factors, we performed linkage disequilibrium score regression (LDSC) analyses on 98 phenotypes from the UK Biobank (UKB), encompassing psychiatric traits, metabolic markers, sex hormones, and lifestyle factors. Our findings revealed significant positive genetic correlations between AN and psychiatric traits (see Table S3, Figure 1), such as obsessive-compulsive disorder (r_g_ = 0.455, *P* = 1.40 × 10^−8^), substance or behavioral addiction (r_g_ = 0.385, *P* = 1.61 × 10^−5^), and worrier / anxious feelings (r_g_ = 0.331, *P* = 3.63 × 10^−19^). Additionally, we observed significant positive genetic correlations between AN and sex hormone-related traits, such as sex hormone-binding globulin (r_g_ = 0.246, *P* = 2.69 × 10^−17^), as well as sleep habits, including snoring (r_g_ = 0.261, *P* = 1.39 × 10^−14^). Conversely, AN showed significant negative genetic correlations with several metabolic traits, including body mass index (BMI) (r_g_ = −0.329, *P* = 4.09 × 10^−35^), basal metabolic rate (r_g_ = −0.195, *P* = 2.58 × 10^−14^), and triglyceride levels (r_g_ = −0.213, P = 1.53 × 10^−9^). Additionally, AN exhibited significant negative genetic correlations with several blood biomarkers, such as C-reactive protein (r_g_ = −0.272, P = 9.90 × 10^−16^), high light scatter reticulocyte count (r_g_ = −0.235, P = 4.86 × 10^−13^), and reticulocyte count (r_g_ = −0.229, P = 2.32 × 10^−12^). To further validate the genetic correlations observed in European populations, we conducted analogous analyses in the Finnish cohort, as detailed in Table S4. Although the limited number of anorexia nervosa cases in this population precluded significant LDSC results, the observed correlations were consistent with those from the European cohort. Specifically, anorexia nervosa demonstrated strong positive genetic correlations with psychiatric traits (all anxiety disorders: r_g_ = 0.907) and strong negative correlations with metabolic markers (obesity: r_g_ = −0.48) in the Finnish population, reflecting similar patterns observed in the broader European sample.

**Figure 1.**
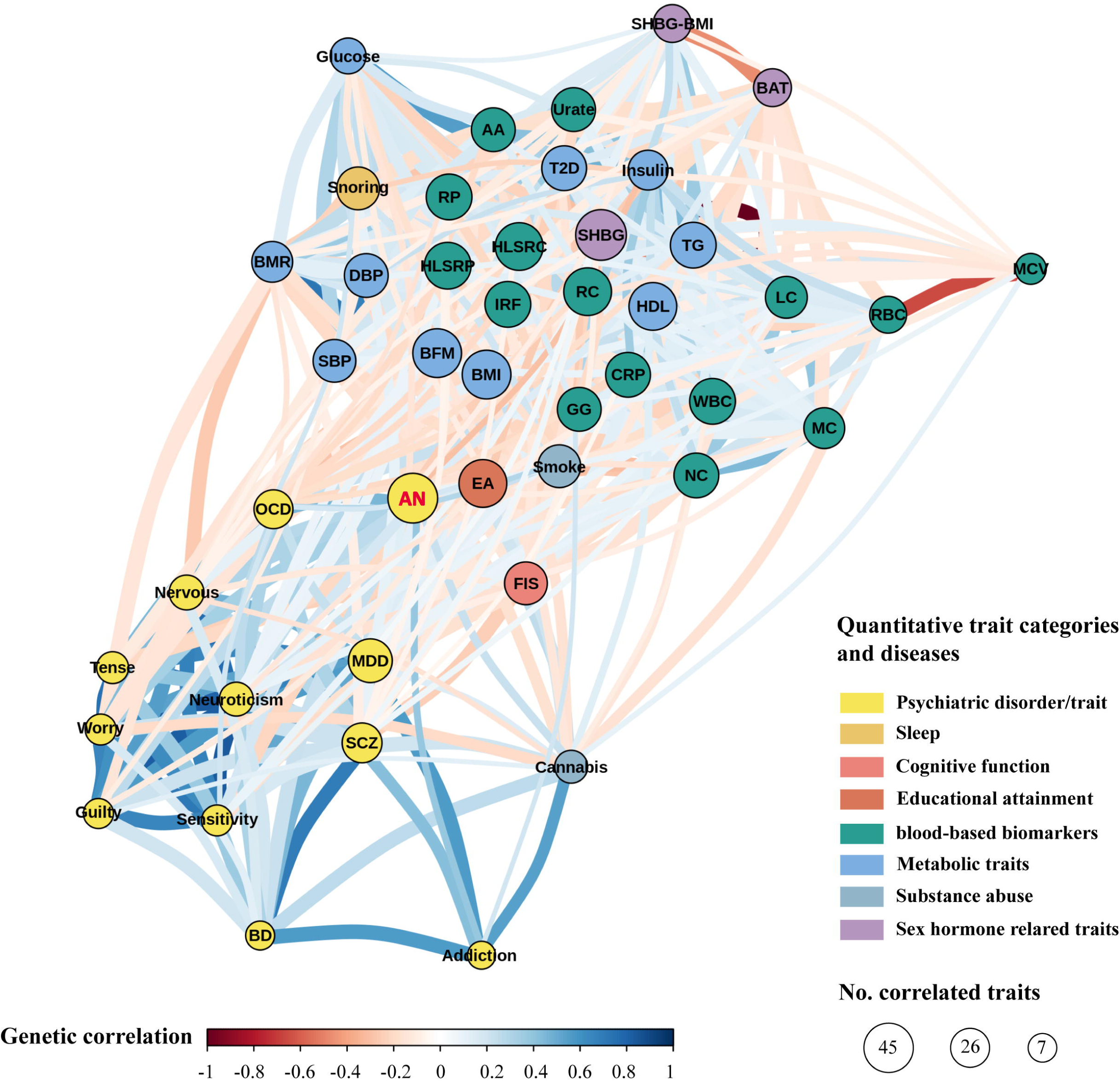
Genetic correlation network between anorexia nervosa and risk factors. Genetically correlated traits and diseases are clustered close together. Each circle represents a trait, and each edge represents a significant genetic correlation (P < 0.001). Positive and negative genetic correlations are indicated by color according to the key. AN, anorexia nervosa; OCD, obsessive-compulsive disorder; MDD, major depressive disorder; SCZ, schizophrenia; BD, bipolar disorder; Guilty, guilty feelings; Addiction, ever addicted to any substance or behaviour; Sensitivity, sensitivity / hurt feelings; Nervous, nervous feelings; Worry, worrier / anxious feelings; Tense, Tense / ‘highly strung’; Neuroticism, neuroticism score; EA, educational attainment (age completed full time education); FIS, Fluid intelligence score; Cannabis, ever taken cannabis; Smoke, age started smoking in former smokers; Snoring, snoring; BAT, Bioavailable testosterone levels; SHBG, Sex hormone-binding globulin levels; SHBG-BMI, Sex hormone-binding globulin levels adjusted for BMI; BMI, body mass index; BFM, whole body fat mass; T2D, type 2 diabetes; BMR, basal metabolic rate; SBP, systolic blood pressure; DBP, diastolic blood pressure; Insulin, fasting blood insulin; HDL, HDL cholesterol; TG, Triglycerides; Glucose, glucose; WBC, white blood cell (leukocyte) count; RBC, red blood cell (erythrocyte) count; MCV, mean corpuscular volume; LC, lymphocyte count; MC, monocyte count; NC, Neutrophill count; RP, reticulocyte percentage; RC, reticulocyte count; IRF, immature reticulocyte fraction; HLSRP, high light scatter reticulocyte percentage; HLSRC, high light scatter reticulocyte count; AA, Alanine aminotransferase (quantile); CRP, C-reactive protein (quantile); GG, gamma glutamyltransferase (quantile); Urate, urate (quantile).

To investigate sex-specific risk factors for AN, we performed LDSC analyses on male- and female-specific GWAS data, enabling us to assess whether genetic correlations in the overall population differ by sex, thereby identifying potential sex-specific genetic risks for AN. The results for males and females are provided in Table S3 and Figures S1 and S2. As anticipated, traits related to sex hormones exhibited pronounced sex differences. For instance, bioavailable testosterone levels were associated with AN only in females (r_g female_ = −0.275, *P* _female_ = 1.22 × 10^−17^; r_g male_ = 0.038, *P* _male_ = 0.293), while SHBG showed much stronger genetic associations in females compared to males (r_g female_ = 0.287, *P* _female_ = 7.19 × 10^−20^; r_g male_ = 0.164, *P* _male_ = 2.61 × 10^−7^). Conversely, total testosterone levels were significantly associated with AN exclusively in males (r_g male_ = 0.166, *P* _male_ = 7.89 × 10^−8^; r_g female_ = −0.095, *P* _female_ = 0.003). In other traits, the results in females generally showed stronger genetic correlations and greater statistical significance. For example, metabolic traits whole body fat mass and HDL cholesterol were correlated with AN in both sexes, but the associations were notably stronger in females (whole body fat mass: r_g female_ = −0.403, *P* _female_ = 2.22 × 10^−41^; r_g male_ = −0.256, *P* _male_ = 2.08 × 10^−16^; HDL: r_g female_ = 0.266, *P* _female_ = 6.76 × 10^−16^; r_g male_ = 0.176, *P* _male_ = 4.42 × 10^−7^). A similar pattern was observed for other blood biochemical traits, such as urate, where the genetic associations were stronger in females than in males (r_g female_ = −0.214, *P* _female_ = 4.26 × 10^−10^; r_g male_ = −0.091, *P* _male_ = 0.009).

### Causal relationship between Anorexia Nervosa and related Phenotypes

To further investigate the causal relationships between AN and the genetically correlated traits identified through LDSC, we conducted a bi-directional Mendelian randomization (MR) analysis using five different methods, with results primarily based on the Inverse Variance Weighted method. The traits analyzed included psychiatric disorders, metabolic traits, sex hormones, and other blood biomarkers. The MR results are summarized in Figure 2, Figures S3, Tables S5 and S6, with Table S5 showing the effect of anorexia nervosa on other phenotypes, and Table S6 displaying the effect of other phenotypes on anorexia nervosa.

**Figure 2.**
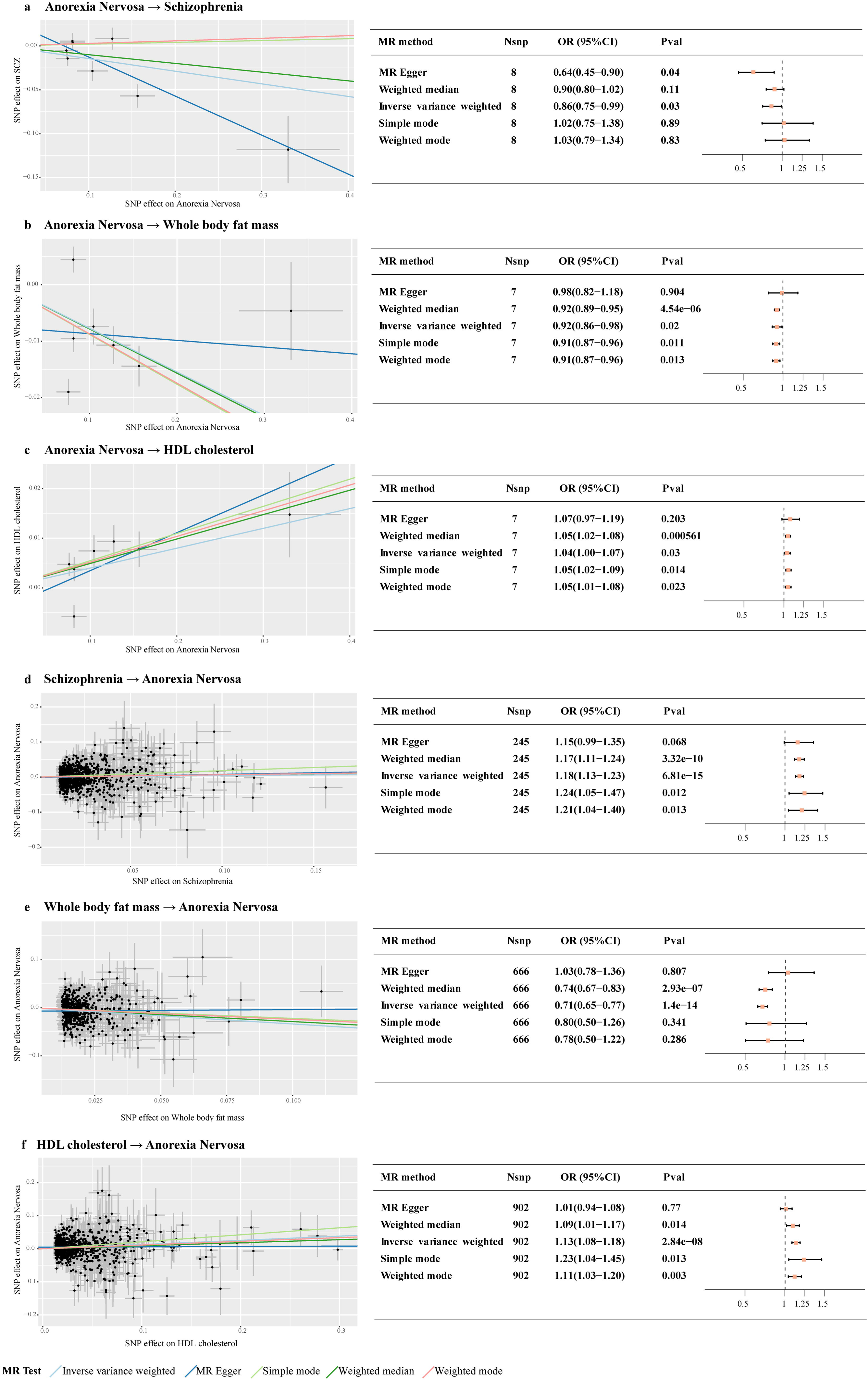
MR plots for the relationship between anorexia nervosa and risk factors. Left panel, scatter plot of SNP effects on anorexia nervosa versus corresponding risk factors, with the slope of each line corresponding to the estimated MR effect per method. Right panel, model-based sensitivity analyses from five methods. The effect estimates represent the odds ratio for outcome per one-point increment in exposure. Data are presented as mean values ± s.e.m. The width of the lines extending from the center point represent the 95% CI. Two-sided unadjusted association P values from five models are given.

Among psychiatric traits, both schizophrenia and neuroticism scores exhibited significant positive causal effects on AN. Specifically, schizophrenia was associated with an increased risk of anorexia nervosa (schizophrenia →AN: OR = 1.18 [95% CI: 1.13, 1.23], p = 6.81×10^−15^), and neuroticism scores were similarly linked to anorexia nervosa (neuroticism scores →AN: OR = 1.60 [95% CI: 1.25, 2.05], p = 1.91×10^−4^). These associations remained robust across multiple MR methods, such as Weighted Median, Simple Mode, and Weighted Mode, further supporting the causal nature of these relationships. These findings align with recent studies, reinforcing the role of psychiatric disorders in the etiology of anorexia nervosa ^14,15^.

In the context of metabolic traits, our analyses revealed a complex bidirectional relationship between AN and body mass index (BMI) related metrics. Whole body fat mass and basal metabolic rate (BMR) exhibited negative causal effects on anorexia nervosa, indicating that reductions in these traits contribute to the disorder’s development (whole body fat mass →AN: OR = 0.71 [95% CI: 0.65, 0.78], p = 1.40×10^−14^; BMR →AN: OR = 0.80 [95% CI: 0.74, 0.87], p = 3.16×10^−7^). Conversely, AN exerted negative causal effects on both BMI and whole body fat mass, suggesting that the disease further exacerbates reductions in these traits. Additionally, we identified a bidirectional positive causal relationship between AN and HDL cholesterol, indicating that elevated HDL cholesterol levels may both result from and contribute to AN (HDL →AN: OR = 1.04 [95% CI: 1.00, 1.08], p = 0.03; AN →HDL: OR = 1.14 [95% CI: 1.09, 1.19], p = 2.84×10^−8^). Moreover, AN showed a positive causal effect on Sex hormone-binding globulin (SHBG) levels, highlighting its influence on hormonal regulation. Several significant causal relationships were identified, indicating that elevated levels of blood biomarkers, such as white blood cell (leukocyte) count (white blood cell →AN: OR = 1.11 [95% CI: 1.06, 1.18], p = 8.54×10^−5^), lymphocyte count (lymphocyte count →AN: OR = 1.15 [95% CI: 1.09, 1.21], p = 2.16×10^−7^), and eosinophil count (eosinophil count →AN: OR = 1.16 [95% CI: 1.08, 1.25], p = 4.28×10^−5^), are associated with an increased risk of developing AN. In contrast, reductions in mean reticulocyte volume and C-reactive protein (CRP) were linked to a higher risk of the disorder (mean reticulocyte volume →AN: OR = 0.93 [95% CI: 0.88, 0.97], p = 3.86×10^−4^; CRP →AN: OR = 0.89 [95% CI: 0.85, 0.94], p = 1.64×10). Furthermore, anorexia nervosa led to decreases in blood parameters like mean corpuscular volume, red blood cell (erythrocyte) distribution width, reticulocyte count, and high light scatter reticulocyte count, indicating widespread impacts on hematological homeostasis.

### Local genetic correlations between Anorexia Nervosa and related Phenotypes

We next conducted a genome-wide scan to identify specific genomic regions contributing to the shared heritability of genetically correlated traits. All significant regions identified in the local genetic correlation analysis are detailed in Table S7. After correcting for multiple testing, 12 significant regions were identified in the local genetic correlation analysis between AN and schizophrenia (Figure S4), six of which represent novel loci not previously reported in AN GWAS. In the analysis of AN and metabolic traits, we uncovered two additional novel regions, with one located at 11p15.3 in the correlation between AN and BMI and another at 12q14.1 associated with HDL cholesterol levels (Figure S5). Moreover, the analysis of AN and sex hormone-binding globulin (SHBG) levels revealed two novel regions at 11p15.1-15.2 and 17q25.3, both validated in a female-specific analysis, suggesting their involvement in sex-specific genetic contributions to these traits (Figure S6). In contrast, only the leukocyte antigen (HLA) region showed significant correlations between AN and blood-based biomarkers, including white blood cell count, reticulocyte count, and urate levels (Figure S7).

### Cell-type-specific enrichment of SNP heritability

We further investigated the genetic architecture of AN and related phenotypes by partitioning SNP heritability across six chromatin marks and nine cell types (Figure S8). The S-LDSC analysis revealed that AN heritability is primarily enriched in tissues linked to the central nervous system, particularly in H3K4me1 and H3K4me3 chromatin marks (Figure S8 d&e). Additionally, significant enrichment was observed in blood and immune-related tissues for chromatin marks such as DNase and H3K36me3 (Figure S8 a&c), suggesting potential involvement of immune pathways. Hierarchical clustering of traits consistently placed AN alongside psychiatric disorders and cognitive traits. For instance, in the H3K36me3 mark, AN clustered closely with traits such as neuroticism, bipolar disorder, major depressive disorder, and fluid intelligence, indicating shared genetic architecture. However, in certain chromatin marks like H3K9ac (Figure S8f), AN displayed a distinct enrichment pattern compared to other psychiatric disorders, reflecting its complex neuro-metabolic origins that differentiate it from traditional models of psychiatric disease.

### Shared and novel loci for AN and related Phenotypes

In light of the genetic correlation findings, we proceeded to explore potential pleiotropic loci shared between AN and related phenotypes. Given the robust associations between AN and psychiatric disorders, we employed multi-trait analysis of GWAS (MTAG) to jointly analyze AN and the four psychiatric traits with the strongest genetic correlations identified in the LDSC analysis—schizophrenia, major depressive disorder, worrier/anxious feelings, and nervous feelings. This analysis uncovered 16 significant loci shared between AN and the psychiatric traits, with 10 of these loci being novel findings for AN (Table S8, Figure 3). The expression quantitative trait loci (eQTLs) for these newly identified pleiotropic loci, derived from the Genotype-Tissue Expression (GTEx) Analysis Release V8, are presented in Table S9. Notably, four novel loci—3p21.1 (rs11713763), 17p13.1 (rs12944041), 11q12.1 (rs2081361), and 5q14.3 (rs1814149)—were identified as eQTLs for neighboring genes in brain tissues. For instance, the rs12944041 functions as an eQTL for the nearby gene *VAMP2* across multiple brain regions, including the hypothalamus, hippocampus, caudate, and putamen. Beyond brain tissues, these novel loci also influence gene expression in gastrointestinal and adipose tissues, with rs2081361 serving as an eQTL for several genes, including *MED19*, *TIMM10*, and *ZDHHC5*, in adipose tissue.

**Figure 3.**
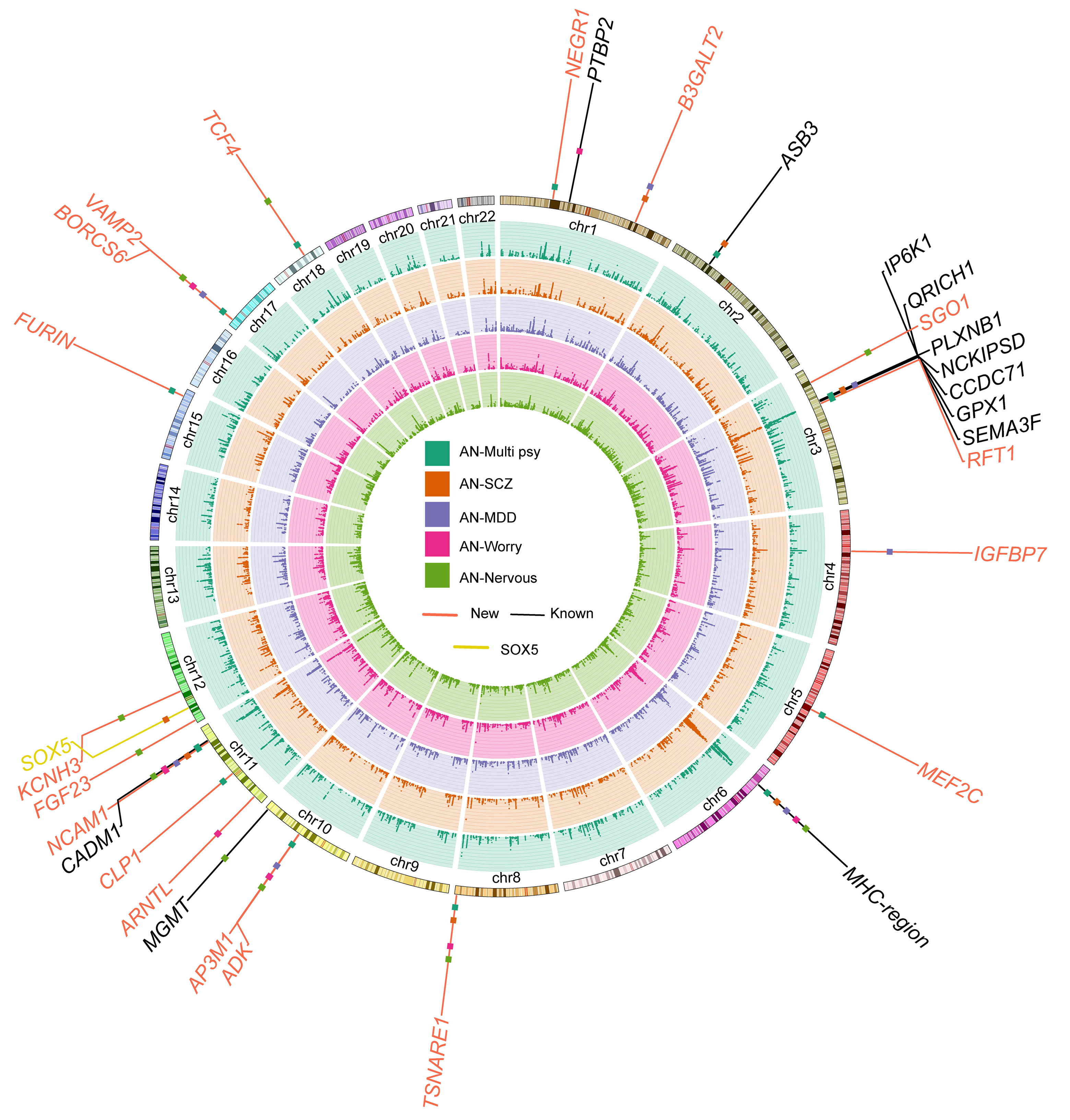
Manhattan plot of the GWAS multi-trait meta-analysis between anorexia nervosa and psychiatric traits. The green circle shows the results of MTAG results of anorexia nervosa and the four psychiatric traits with the strongest genetic correlations identified in the LDSC analysis—schizophrenia, major depressive disorder, worrier/anxious feelings, and nervous feelings; the orange circle shows the results of two-trait MTAG between anorexia nervosa and schizophrenia; the purple circle shows the results of two-trait MTAG between anorexia nervosa and major depressive disorder; the pink circle shows the results of two-trait MTAG between anorexia nervosa and worrier/anxious feelings; the light green circle shows the results of two-trait MTAG between anorexia nervosa and nervous feelings.

To further explore pleiotropic risk loci associated with AN, we transitioned from multi-trait joint analyses to a series of two-trait MTAG analyses, each pairing AN with a relevant phenotype identified through LDSC. Focusing on psychiatric traits with significant genetic correlations to AN, these analyses identified 20 independent loci, 13 of which are novel in the context of AN (Table S10). Notably, five of these loci were also observed in the earlier MTAG analysis involving AN and the four psychiatric traits—schizophrenia, major depressive disorder, worrier/anxious feelings, and nervous feelings. Additionally, the novel locus at 12p12.1, previously discovered in our meta-analysis of AN across European and Finnish cohorts, was replicated in the MTAG analysis of AN and schizophrenia, further supporting the pleiotropic effects of this locus across multiple psychiatric conditions. Another novel locus, 11p15.2 (rs900144), consistently emerged in the MTAG analysis of AN and several psychiatric traits, including nervous feelings, neuroticism score, guilty feelings, and worry too long after embarrassment. This locus is located near the *ARNTL* gene, which is involved in metabolic, immune, and oxidative stress responses, suggesting potential implications for AN pathogenesis through its impact on energy metabolism and stress-related pathways.

In the MTAG analysis of AN and metabolic traits, including measures such as basal metabolic rate, BMI, and HDL cholesterol, we identified a total of 44 significant loci, of which 33 were novel discoveries (Table S11, Figure S9). As anticipated, many of the genes near these newly identified loci are associated with metabolic functions, such as *LPL* (8p21.3), *FADS1*(11q12.2), and *NPIPB6* (16p11.2), which are involved in lipid metabolism and fatty acid processing. Interestingly, several of these loci also encompass genes implicated in psychiatric conditions, cognitive function, and brain phenotypes. For example, genes such as *BDNF* (11p14.1), *PCDH17* (13q21.1), *FANCL* (2p16.1), and *EPHA3* (3p11.1) are essential for neuronal development, synaptic plasticity, and brain structure. Notably, several novel loci function as eQTLs, suggesting potential regulatory roles in gene expression. For instance, the lead SNP rs4751 at the 1p36.33 locus acts as an eQTL for the nearby gene *CFAP74* in the cortex, *MMP23A* in gastrointestinal tissues, and *SLC35E2* in adipose tissues, indicating its potential involvement in AN. Additionally, in the MTAG analysis of AN and sex hormone-binding globulin (SHBG) levels, we identified nine novel loci. Notably, the 22q12.2 locus (rs174723) stood out, as it consistently appeared in MTAG analyses of AN with other biochemical markers, including reticulocyte percentage, immature reticulocyte fraction, and high light scatter reticulocyte count (Figure S10). This locus influences the expression of nearby genes in various tissues, such as *THOC5*, which is expressed in adipose and gastrointestinal tissues, and *RFPL1S*, which shows differential expression in brain tissues. These findings suggest that rs174723 may have a pleiotropic effect, influencing both metabolic and hematological traits, as well as playing a role in neurodevelopmental pathways. Further details on the two-trait MTAG results between AN and other biochemical markers, as well as lifestyle factors, are provided in Table S12.

### Key gene modules in anorexia nervosa identified through transcriptome network analysis

We first conducted TWAS to identify genes whose genetically predicted expression levels are associated with AN across multiple tissues, including brain, adipose, thyroid, and gastrointestinal tissues. Applying a significance threshold of p < 0.01, we identified 640 significant genes in the brain (Table S13), 464 in adipose tissue (Table S14), 364 in the thyroid (Table S15), and 576 in the gastrointestinal tissues (Table S16). We then applied WGCNA to RNA sequencing data from female samples in the GTEx v8 dataset to construct gene co-expression networks for each tissue. To assess the overlap between TWAS-identified gene sets and their corresponding WGCNA modules, we performed Fisher’s exact test. At a significance threshold of p < 0.05, we identified gene modules significantly enriched with TWAS-identified genes (Table S17). Further, gene ontology analysis of these enriched modules revealed their biological functions and pathways, providing deeper insights into the underlying molecular mechanisms related to AN (Table S18). In brain tissue, the TWAS-identified gene sets were significantly enriched in six distinct co-expression modules. Among these, M5 [Cerebellar Hemisphere] and M5 [Frontal Cortex (BA9)] were primarily associated with neurological processes such as nervous system development (GO:0007399), neurogenesis (GO:0022008), and neuron projection (GO:0043005). In contrast, the modules M3 [Hippocampus] and M16 [Hippocampus], while also linked to neurological processes, were more prominently involved in metabolic functions, including oxidative phosphorylation (GO:0006119), respiratory chain complex (GO:0098803), primary metabolic process (GO:0044238) and ATP metabolic process (GO:0046034). Additionally, modules in the brain that were exclusively associated with metabolic functions, such as M10 [Amygdala] and M12 [Amygdala], were strongly linked to the regulation of primary metabolic process (GO:0080090) and regulation of metabolic process (GO:0019222). In non-brain tissues, including adipose and gastrointestinal tissues, the enriched modules also primarily related to both neurological and metabolic functions. For example, modules M12 [Adipose - Subcutaneous] and M12 [Adipose - Visceral (Omentum)] were associated with neurological processes such as synapse (GO:0045202) and neurogenesis (GO:0022008). Meanwhile, modules like M10 [Adipose - Subcutaneous] and M2 [Esophagus - Gastroesophageal Junction] exhibited dual roles, being involved in both neurological functions and metabolic processes. These findings underscore the critical involvement of both neural and metabolic mechanisms, reinforcing the complex interplay between these processes in the development of AN.

To further evaluate the robustness and reproducibility of these co-expression modules across different tissues, we conducted a preservation analysis. This analysis aimed to determine the stability of module structure and gene relationships across tissue types, thereby providing additional insights into the consistency of neuro-metabolic pathways implicated in AN. For modules identified in brain tissues, a strong preservation was almost observed (Z > 10) across all brain regions, while a weak to moderate preservation (2 < Z score < 10) was detected in most non-brain tissues. It is worth mentioning that modules M10 [Amygdala], M12 [Amygdala] and M3 [Hippocampus] not only demonstrated strong preservation within brain tissues but also exhibited comparable conservation in adipose, thyroid and gastrointestinal tissues (Figure 4). In contrast, while most modules identified in adipose and gastrointestinal tissues exhibited high preservation primarily within non-brain tissues, a predominantly weak to moderate preservation was observed across most brain tissues. Notably, modules M10 [Adipose - Subcutaneous] and M14 [Esophagus - Muscularis] demonstrated remarkably high preservation strength across nearly all brain tissues (Figure 4). The observed cross-tissue preservation highlights the importance of shared molecular networks between the brain and peripheral tissues, reinforcing the hypothesis that systemic metabolic dysregulation, in concert with neural processes, plays a critical role in the pathogenesis of AN.

**Figure 4.**
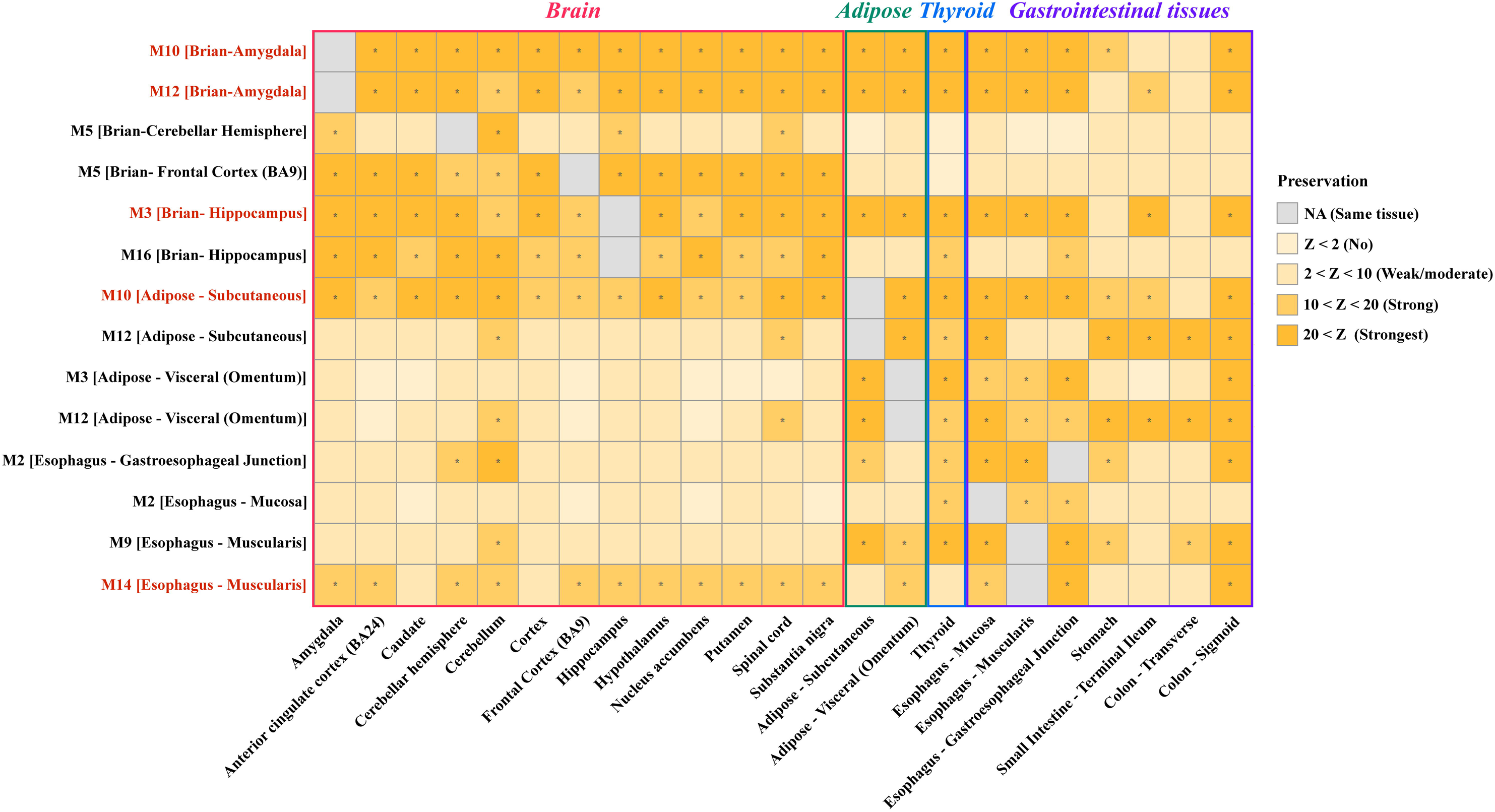
Preservation of co-expression modules across brain and digestive tissues. Colors of the heatmap indicate module preservation, with Z values > 20 representing the strongest evidence, >10 denoting strong evidence, 2-10 suggesting weak to moderate preservation, and < 2 indicating no preservation. On the x-axis, the red box represents brain tissues, the green box represents adipose tissues, the blue box represents thyroid tissue, and the purple box represents digestive tissues. On the y-axis, modules highlighted in red represent those highly preserved across both brain and non-brain tissues.

## Discussion

To our knowledge, this study is the first comprehensive genome-wide multi-trait analysis investigating the genetic correlations, pleiotropic loci, and shared genetic mechanisms between AN and a broad spectrum of risk factors, including psychiatric traits, metabolic markers, sex hormones, and lifestyle variables. First, by integrating summary statistics from the largest available GWAS of AN in European and Finnish populations, we identified a novel locus through inverse-variance weighted meta-analysis. Second, we performed an in-depth examination of 98 distinct phenotypes from the UK Biobank to identify risk factors for AN, with sex-specific analyses revealing gender-specific genetic risks. Third, through MR, S-LDSC, HESS, and MTAG, we further explored the causal relationships between AN and its risk factors, as well as the pleiotropic loci and shared genetic mechanisms. Finally, we identified key gene modules involved in the neurological and metabolic pathways of AN through transcriptome network analysis.

In 2017, the PGC conducted the first anorexia nervosa GWAS (3,495 cases, 10,982 controls), identifying the first significant locus associated with AN at 12q13.2 ^3^. More recently, an expanded GWAS of 16,992 cases and 55,525 controls identified eight additional genome-wide significant loci ^12^. However, compared to other psychiatric disorders like schizophrenia and depression, the loci influencing AN remain largely undiscovered. In this study, we integrated summary statistics from the largest available GWAS in European and Finnish populations and conducted a traditional meta-analysis, identifying a novel genome-wide significant locus at 12q21, which marks the tenth genome-wide significant locus associated with AN. Notably, this locus was also detected in subsequent MTAG analyses of AN and schizophrenia and has been reported in recent condFDR/conjFDR cross-disorder analyses of schizophrenia and AN ^15^, further corroborating its reliability. Additionally, this locus is located near *SOX5*, a gene that plays a crucial role in neurodevelopment and neuronal differentiation ^19–22^. *SOX5* is a transcription factor involved in the development of the central nervous system, particularly in the differentiation of neurons and glial cells ^23,24^. It has been implicated in various neuropsychiatric disorders, including schizophrenia ^25^, depression ^26^ and bipolar disorders ^27^, with its dysregulation contributing to an increased susceptibility to these conditions.

In addition to identifying novel loci through the meta-analysis of AN, we further investigated the genetic correlations and pleiotropic loci shared between AN and other psychiatric disorders. Consistent with previous studies, we observed robust positive genetic correlations between AN and a spectrum of psychiatric traits, such as schizophrenia, bipolar disorder, major depressive disorder, and obsessive-compulsive disorder ^3,12,14^. These findings highlight the complex polygenic architecture of AN, characterized by substantial genetic overlap with these psychiatric conditions^14,15^. Furthermore, our MR analyses provided evidence that several psychiatric disorders, including schizophrenia, neuroticism, and difficulty falling asleep, significantly increase the risk of AN. This reinforces the hypothesis that the observed genetic correlations are not solely attributable to shared pleiotropic loci but also reflect direct causal relationships. These results suggest that specific psychiatric traits may contribute to the pathogenesis of AN, thereby enhancing our understanding of the etiological connections between AN and its comorbid psychiatric traits. Next, we performed MTAG analysis for AN and related psychiatric disorders. We uncovered 16 novel pleiotropic loci shared between AN and four psychiatric traits— schizophrenia, major depressive disorder, worrier/anxious feelings, and nervous feelings—which showed the strongest genetic correlations in the LDSC analysis. In addition, the two-trait MTAG analysis identified eight novel loci shared between AN and psychiatric traits. Notably, several of these newly discovered loci have been previously reported as pleiotropic loci associated with multiple psychiatric disorders. For example, loci at 1p31.1, 5q14.3, 15q26.1, and 18q21.2 were identified in a comprehensive meta-analysis of eight psychiatric disorders ^28^, indicating their critical role in revealing shared genetic pathways underlying psychiatric comorbidity. Moreover, several newly identified loci influence the expression of nearby genes in brain tissues. For instance, the lead SNP rs12944041 of the 17p13.1 locus served as an eQTL for the nearby gene *VAMP2* in seven brain tissues such as caudate, hippocampus, hypothalamus and frontal cortex. *VAMP2* encodes a key component of the synaptic vesicle fusion complex, playing a crucial role in neurotransmitter release at the step between vesicle docking and fusion, which is essential for efficient neural communication and cognitive functions ^29–33^. Interestingly, beyond eQTL effects in brain tissue, several novel loci also regulate gene expression in non-brain tissues, such as adipose, gastrointestinal, and thyroid tissues. For example, the lead SNPs rs4281434 and rs4338470 are significantly associated with the expression level of *ADK*, a gene known for its vital roles in energy metabolism, neuroprotection, and signal transduction. These observations, along with the distinct cell-type-specific enrichment patterns revealed by the S-LDSC analysis— where AN exhibits a unique enrichment profile compared to traditional psychiatric disorders— underscore the complex neuro-metabolic underpinnings of AN. This differentiation from conventional psychiatric disorders motivates further investigation into the metabolic genetic mechanisms contributing to the development of AN.

In our investigation of AN and metabolic-related traits, we identified significant genetic correlations between AN and key metabolic parameters, including BMI, whole body fat mass, basal metabolic rate (BMR), and HDL cholesterol. Unlike the previously mentioned unidirectional influence of psychiatric disorders on AN, the MR results revealed bidirectional causal relationships between AN and these metabolic traits. For instance, elevated HDL cholesterol levels not only increased the risk of developing AN but, conversely, AN also contributed to changes in HDL cholesterol levels. Notably, abnormally elevated HDL cholesterol levels in AN patients have been widely reported, further supporting this finding ^34^. These bidirectional associations highlight the complex interplay between metabolic processes and the pathophysiology of AN. We next conducted a MTAG analysis to uncover pleiotropic variants impacting both AN and metabolic traits. We identified 33 novel pleiotropic loci, five of which, including 10q26.13, 11p15.2, 11q23.3, 4q12, and 6q16.1, were also found in the above mentioned MTAG analysis of AN and psychiatric traits. These shared loci further support the genetic overlap between anorexia nervosa and associated psychiatric and metabolic factors.

Additionally, the lead SNP, rs6265, is a missense variant near the *BDNF* gene. Brain-derived neurotrophic factor (BDNF) is one of the most widely distributed and extensively studied neurotrophins in the mammalian brain. Among its key functions, BDNF regulates neuronal and glial development, provides neuroprotection, and modulates both short-term and long-term synaptic interactions, which are essential for cognition and memory ^35–38^. Additionally, in the MTAG analysis of AN and other blood biochemical markers, we identified 23 novel loci, several of which were identified as eQTLs influencing multiple nearby genes across various tissues. For instance, the variant rs13133311 affects the expression of *HSD17B11* in brain, adipose, and gastrointestinal tissues. HSD17B11, a key member of the 17-β hydroxysteroid dehydrogenase family, plays a critical role in steroid metabolism and hormone regulation. Its proper function is essential for maintaining hormonal and metabolic balance, while dysfunction can lead to endocrine and metabolic disorders. These findings underscore the critical role of metabolic regulation in AN pathophysiology.

In addition to identifying novel pleiotropic loci primarily involved in neurological and metabolic functions, which reflect the metabolic-psychiatric mechanisms underlying AN, our transcriptome network analysis provides further evidence for this connection. By integrating TWAS and WGCNA, we identified key gene modules associated with AN. Functional annotation results revealed that the identified gene modules fall into three categories: (1) neurological functions, exemplified by the M5 [Cerebellar Hemisphere] and M12 [Cerebellar Hemisphere] modules; (2) metabolic functions, represented by the M12 [Amygdala] and M2 [Esophagus - Gastroesophageal Junction]; and (3) both neurological and metabolic functions, including the M3 [Hippocampus] and M10 [Adipose - Subcutaneous]. Furthermore, following preservation analysis revealed that specific modules, such as M10 [Amygdala], M12 [Amygdala], and M3 [Hippocampus] in brain tissues, along with M10 [Adipose - Subcutaneous] and M14 [Esophagus - Muscularis] in non-brain tissues, are highly conserved across both brain and non-brain tissues. These findings elucidate how gene modules that integrate both psychiatric and metabolic functionalities exert a coordinated influence on the pathophysiology of AN through interactions within brain and peripheral tissues.

In conclusion, our integrative analysis provides a comprehensive understanding of the shared genetic mechanisms linking AN with psychiatric disorders and metabolic factors. Through the integration of genetic data and transcriptome network analysis, we revealed novel genetic associations and offered additional insights from gene co-expression network perspectives. Future functional studies are warranted to validate the key gene targets identified in this study, which will deepen our understanding of the intricate interplay between psychiatric and metabolic mechanisms. We hope our study may help guide strategies aimed at improving health outcomes for individuals affected by AN.

## Methods

### GWAS data

The GWAS summary statistics for AN were obtained from the largest GWAS meta-analysis to date, conducted on a European population (16,992 cases and 55,525 controls), using combined data from the Anorexia Nervosa Genetics Initiative (ANGI) ^39,40^ and the Eating Disorders Working Group of the Psychiatric Genomics Consortium (PGC-ED). Additionally, the GWAS summary statistics for AN in the Finnish population were derived from the FinnGen consortium^41^, which included 2,115 AN cases and 400,510 controls.

The GWAS summary statistics for risk factors in Europeans and Finns were sourced from UK Biobank (UKB)^42^ and FinnGen (FG), respectively. UK Biobank is a prospective population-based study primarily involving UK residents aged 40 to 69 years. In contrast, FinnGen includes both population-based cohorts and disease-specific cohorts. We selected potential risk factors for AN, including psychiatric phenotypes, metabolic indicators, sex hormones, and lifestyle factors. Precomputed summary statistics from round 2 UKB GWAS, including both sex-combined and sex-specific data, were obtained from the Neale lab website (http://www.nealelab.is/uk-biobank). FinnGen release 10 data was accessed from the project website (https://r10.finngen.fi). The list of traits analyzed for Europeans is shown in Supplementary Table S1, while the details for Finns are presented in Supplementary Table S2.

### Meta-Analysis of GWAS in Europeans and Finns

We conducted an inverse-variance weighted meta-analysis on the GWAS data for anorexia nervosa in European and Finnish populations using the basic meta-analysis function in PLINK (v1.90b7). Fixed-effect meta-analysis *P* values and fixed-effect beta values were calculated. We prioritized SNPs that achieved genome-wide significance (*P* < 5×10^−8^) in the meta-analysis and those that demonstrated suggestive significance (*P* < 0.05) in the original single-trait GWAS.

### Global genetic correlation analysis

To identify potential risk factors for AN in European populations, we analyzed 98 GWAS summary statistics of deep phenotypes from the UKB, including psychiatric phenotypes, cognitive function, metabolic indicators, blood-based biomarkers, sex hormone-related traits, and lifestyle factors. The genetic correlation (rg) between AN and each risk factor was estimated using linkage disequilibrium (LD) score regression (LDSC)^43^. For this analysis, we utilized pre-computed LD scores for HapMap3 SNPs based on European-ancestry data from the 1000 Genomes Project. To investigate the presence of sex-specific risk factors for AN, the LDSC analyses were also conducted separately on male- and female-specific GWAS data.

Furthermore, to assess the robustness of the LDSC results in European populations, we conducted analogous analyses in Finnish populations. Due to differences in trait availability between the FINNGEN project and the UKB database, we focused on 51 risk factors from FINNGEN, including psychiatric disorders, metabolic traits, and lifestyle indicators. In the LDSC analysis of AN and risk factors in the Finnish population, we used pre-computed LD scores for HapMap3 SNPs based on Finnish-ancestry data from the 1000 Genomes Project.

### Mendelian randomization

To explore the causal relationship between AN and other phenotypes, we conducted Mendelian randomization (MR) analyses on phenotypic pairs with global genetic correlations (LDSC *P* < 0.001). Instrumental variables (IVs) were selected from exposure GWAS data through LD clumping (r2 threshold: 0.01, P-value threshold: 5×10^−8^, window size: 10 MB). Data corresponding to these IVs were then extracted from both exposure and outcome datasets and harmonized. Bidirectional Mendelian randomization analysis was conducted using the five MR methods [Inverse variance weighted (IVW), MR Egger, Weighted median, Simple mode, and Weighted mode] implemented in the R package TwoSampleMR^44,45^. Results with a P-value < 0.001 from any of the MR methods were considered statistically significant, and other results were also presented for comprehensive analysis.

### Local genetic correlation analysis

Given that genetic correlation, as estimated by LDSC, integrates data from all genetic variants across the genome, we further estimated the pairwise local genetic correlation using ρ-HESS (heritability estimation from summary statistics)^46^. ρ-HESS are designed to quantify the local genetic correlation between pairs of traits at each of the 1703 prespecified LD independent segments with an average length of 1.6 Mb. A Bonferroni corrected *P* value less than 0.05/1703 was considered as statistically significant.

### Cell-type-specific enrichment of SNP heritability

S-LDSC was used to detect potential functional categories or cell types contributing disproportionately to the heritability of AN and its associated risk factors (LDSC *P* < 0.001). Annotation data constructed by the Roadmap project for six chromatin marks (DHS, H3K27ac, H3K36me3, H3K4me1, H3K4me3, and H3K9ac) in a set of 88 cell types or tissues were used to partition the SNP heritability of each trait. For each chromatin mark, cell-type-specific annotations were further classified into nine broad groups including adipose, central nervous system, digestive system, cardiovascular, musculoskeletal and connective tissue, immune and blood, liver, pancreas, and other^47^. Annotation-specific enrichment values for each trait were transformed into color scale and visualized by hierarchical clustering.

### Multi-trait analysis of GWAS

Multi-trait GWAS meta-analysis for AN and the corresponding genetically correlated risk factors was performed by the Multi-Trait Analysis of GWAS (MTAG) framework, a generalized meta-analysis method that outputs trait-specific SNP associations^13^. MTAG can increase the power to detect loci from correlated traits by analyzing GWAS summary statistics jointly. It also accounts for sample overlap and incomplete genetic correlation, when comparing with the conventional inverse-variance weighted meta-analysis. The first step of MTAG is to filter variants by removing non common SNPs, duplicated SNPs, or SNPs with strand ambiguity. MTAG then estimates the pairwise genetic correlation between traits using LDSC^48^ and uses these estimates to calibrate the variance-covariance matrix of the random effect component. MTAG next performs a random-effect meta-analysis to generate the SNP-level summary statistics. In MTAG analysis, we consistently designated AN as trait 1and other risk factors as trait 2. The MTAG results reported are specific to trait 1 (AN). We prioritized significant pleiotropic SNPs that reached genome-wide significance (*p* < 5×10^−8^) in the multi-trait analysis and suggestive significance (*p* < 0.05) in the original single-trait GWAS.

### Transcriptome-wide association study

Transcriptome-wide association study (TWAS) analysis is a method used to investigate the relationship between gene expression and a specific trait, enabling the identification of genes that differ from those detected through GWAS analysis. We conducted TWAS using S-PrediXcan^49^ and the GTEx v8 MASHR-M models^50^ to identify genes whose genetically predicted expression levels are associated with AN. The tissues analyzed were selected based on their relevance to AN, including 13 brain tissues, 8 digestive tract tissues, 2 adipose tissues, and 1 thyroid tissue. Given the correlation of eQTLs across tissues, we employed S-MultiXcan to integrate the results within each tissue system, yielding meta-results for the brain, adipose tissue, and digestive tract. Genes with a *P* value lower than 0.01 were considered potentially significant in relation to AN and were subsequently included in gene enrichment analyses.

### Weighted Gene Co-expression Network Analysis (WGCNA)

RNA sequencing data (read counts) from the aforementioned tissues were obtained from the GTEx v8. All samples were exclusively derived from female donors due to the predominance of female patients in the anorexia GWAS. Initially, unexpressed genes (with a read count value of 0) were removed from the analysis. For normalization, the DESeq2 package was utilized to create a DESeq dataset for subsequent differential expression analysis. Variance stability transformation (VST) was applied to achieve approximate homoscedasticity by fitting the dispersion-mean relationship of the dataset, and read counts were normalized by dividing by the size factor. Gene co-expression modules for the aforementioned tissues were independently constructed using the WGCNA package in R^51^. A signed pairwise correlation matrix was calculated using Pearson’s product-moment correlation coefficient, emphasizing strong gene-gene correlations by selecting an appropriate “soft-thresholding” value. This correlation matrix was transformed into an adjacency matrix and normalized using a topological overlap function. Hierarchical clustering and module segmentation were performed using average linkage and the dynamic tree cut algorithm, respectively. Gene ontology analysis for each identified co-expression module was conducted using g:Profiler, with enrichment significance set at FDR < 0.05.

### Module preservation

We employed Fisher’s Exact test to evaluate the overlap between gene sets associated with AN from the TWAS analysis and gene modules identified by WGCNA, with statistical significance defined at a *P*-value threshold of 0.01. Modules that consistently appeared across multiple tissues were designated as key modules. We then assessed the preservation of these key modules across various tissues to determine their stability and relevance. Network module preservation across GTEx tissues was analyzed using the ‘modulePreservation’ function in WGCNA^52^. This approach, involving ‘reference’ and ‘test’ network modules, calculates preservation statistics in three categories: i) density-based, assessing the similarity of gene-gene connectivity patterns; ii) separability-based, determining whether test network modules remain distinct within reference network modules; and iii) connectivity-based, evaluating the similarity of gene connectivity patterns between reference and test networks. Four complementary statistics—median rank, Z_density_, Z_connectivity_, and Z_summary_—were used to determine module preservation. Z_density_ and Z_connectivity_ are standardized measures of density and connectivity, respectively, while Z_summary_ represents their average. Module preservation was established based on Z_summary_ values: a Z_summary_ > 20 indicates the strongest evidence of preservation, >10 indicates strong evidence, 2-10 suggests weak to moderate preservation, and <2 indicates no preservation.

## Data Availability

All data produced in the present study are available upon reasonable request to the authors

## Conflict of interest

The authors report no biomedical financial interests or potential conflicts of interest.

## Acknowledgements

The study was supported by grants from the Special Funds of Taishan Scholar Project, China (tsqn202211224), the National Natural Science Foundation of China (32270661), Excellent Youth Science Fund Project (Overseas) of Shandong China (2023HWYQ-082), Shandong Postdoctoral Science Foundation (SDCX-ZG-202400042) and Shandong Province Higher Education Institution Youth Innovation and Technology Support Program (2023KJ179).

## Notes

### Competing Interest Statement

The authors have declared no competing interest.

