## Supplemental Tables and Figures for "Integrative GWAS Identifies Novel Loci and Genetic Links Between Psychiatric and Metabolic Factors in Anorexia Nervosa"

**Song *et al.***

**Supplementary Online Contents**

**Table S1.** Details on the characteristics of GWAS datasets for Europeans.
**Table S2.** Details on the characteristics of GWAS datasets for Finns.
**Table S3.** Global genetic correlations between anorexia nervosa and potential risk factors in European populations.
**Table S4.** Genetic correlations between anorexia nervosa and potential risk factors in Finnish populations.
**Table S5.** Mendelian randomization results: effect of anorexia nervosa on other phenotypes.
**Table S6.** Mendelian randomization results: effect of other phenotypes on anorexia nervosa.
**Table S7.** Significant genomic regions identified in the local genetic correlation analysis between anorexia nervosa and genetically correlated traits.
**Table S8.** Multi-Trait Meta-Analysis of anorexia nervosa and four psychiatric traits.
**Table S9.** Single-Tissue eQTLs for novel loci.
**Table S10.** Two-trait MTAG of anorexia nervosa and psychiatric traits.
**Table S11.** Two-trait MTAG of anorexia nervosa and metabolic traits.
**Table S12.** Two-trait MTAG of anorexia nervosa and other traits.
**Table S13.** TWAS-identified genes associated with anorexia nervosa in brain tissues.
**Table S14.** TWAS-identified genes associated with anorexia nervosa in adipose tissues.
**Table S15.** TWAS-identified genes associated with anorexia nervosa in the thyroid.
**Table S16.** TWAS-identified genes associated with anorexia nervosa in gastrointestinal tissues.
**Table S17.** Gene co-expression modules enriched with TWAS-identified genes across multiple tissues.
**Table S18.** Enriched GO and KEGG Pathways for co-expression modules identified in each tissue.

**Figure S1.** Genetic correlation network between anorexia nervosa and risk factors in males.
**Figure S2.** Genetic correlation network between anorexia nervosa and risk factors in females.
**Figure S3.** MR plots for the relationship between anorexia nervosa and risk factors.
**Figure S4.** Local genetic correlation between anorexia nervosa and schizophrenia.
**Figure S5.** Local genetic correlation between anorexia nervosa and metabolic traits.
**Figure S6.** Local genetic correlation between anorexia nervosa and sex hormone related traits.
**Figure S7.** Local genetic correlation between anorexia nervosa and blood-based biomarkers.
**Figure S8.** Heat maps showing patterns of cell-type-specific enrichments of SNP-heritability for genetically associated traits across 88 cell types- or tissues-specific annotations.
**Figure S9.** Manhattan plot of the GWAS multi-trait meta-analysis between anorexia nervosa and metabolic traits.
**Figure S10.** Manhattan plot of the GWAS multi-trait meta-analysis between anorexia nervosa and biochemical markers.

**Table S1**. Details on the characteristics of GWAS datasets for Europeans.

| Phenotype | Data source | Combine | | | Female | | | Male | | |
|---|---|---|---|---|---|---|---|---|---|---|
| | | Total | Ncase | Ncontrol | Total | Ncase | Ncontrol | Total | Ncase | Ncontrol |
| Anorexia Nervosa | PMID: 35216811 | 72,517 | 16,992 | 55,525 | NA | NA | NA | NA | NA | NA |
| Schizophrenia | PMID: 35396580 | 122,800 | 50,807 | 71,993 | 54,513 | 17,710 | 36,803 | 68,287 | 33,097 | 35,190 |
| Bipolar Disorder | PMID: 34002096 | 413,466 | 41,917 | 371,549 | NA | NA | NA | NA | NA | NA |
| Major Depressive Disorder | PMID: 30718901 | 446,238 | 118,811 | 327,427 | NA | NA | NA | NA | NA | NA |
| Obsessive-Compulsive Disorde | PMID: 28761083 | 9,725 | 2,688 | 7,037 | NA | NA | NA | NA | NA | NA |
| ADHD | PMID: 36702997 | 77,382 | 38,691 | 38,691 | NA | NA | NA | NA | NA | NA |
| Panic disorder_PGC | PMID: 31712720 | 9,907 | 2,147 | 7,760 | NA | NA | NA | NA | NA | NA |
| ASD | PMID: 38327885 | 46,351 | 18,382 | 27,969 | NA | NA | NA | NA | NA | NA |
| PTSD | PMID: 31594949 | 170,000 | 30,000 | 170,000 | NA | NA | NA | NA | NA | NA |
| White blood cell (leukocyte) count | Neale Lab UKB | 350,470 | NA | NA | 188,074 | NA | NA | 162,396 | NA | NA |
| Red blood cell (erythrocyte) count | Neale Lab UKB | 350,475 | NA | NA | 188,076 | NA | NA | 162,399 | NA | NA |
| Haemoglobin concentration | Neale Lab UKB | 350,474 | NA | NA | 188,076 | NA | NA | 162,398 | NA | NA |
| Haematocrit percentage | Neale Lab UKB | 350,475 | NA | NA | 188,076 | NA | NA | 162,399 | NA | NA |
| Mean corpuscular volume | Neale Lab UKB | 350,473 | NA | NA | 188,075 | NA | NA | 162,398 | NA | NA |
| Mean corpuscular haemoglobin | Neale Lab UKB | 350,472 | NA | NA | 188,074 | NA | NA | 162,398 | NA | NA |
| Mean corpuscular haemoglobin concentration | Neale Lab UKB | 350,468 | NA | NA | 188,072 | NA | NA | 162,396 | NA | NA |
| Red blood cell (erythrocyte) distribution width | Neale Lab UKB | 350,473 | NA | NA | 188,075 | NA | NA | 162,398 | NA | NA |
| Platelet count | Neale Lab UKB | 350,474 | NA | NA | 188,075 | NA | NA | 162,399 | NA | NA |
| Platelet crit | Neale Lab UKB | 350,471 | NA | NA | 188,073 | NA | NA | 162,398 | NA | NA |
| Mean platelet (thrombocyte) volume | Neale Lab UKB | 350,470 | NA | NA | 188,072 | NA | NA | 162,398 | NA | NA |
| Platelet distribution width | Neale Lab UKB | 350,470 | NA | NA | 188,072 | NA | NA | 162,398 | NA | NA |

| | | | | | | | | | | |
|---|---|---|---|---|---|---|---|---|---|---|
| Lymphocyte count | Neale Lab UKB | 349,856 | NA | NA | 187,758 | NA | NA | 162,098 | NA | NA |
| Monocyte count | Neale Lab UKB | 349,856 | NA | NA | 187,758 | NA | NA | 162,098 | NA | NA |
| Neutrophill count | Neale Lab UKB | 349,856 | NA | NA | 187,758 | NA | NA | 162,098 | NA | NA |
| Eosinophill count | Neale Lab UKB | 349,856 | NA | NA | 187,758 | NA | NA | 162,098 | NA | NA |
| Basophill count | Neale Lab UKB | 349,856 | NA | NA | 187,758 | NA | NA | 162,098 | NA | NA |
| Nucleated red blood cell count | Neale Lab UKB | 349,848 | NA | NA | 187,752 | NA | NA | 162,096 | NA | NA |
| Lymphocyte percentage | Neale Lab UKB | 349,861 | NA | NA | 187,760 | NA | NA | 162,101 | NA | NA |
| Monocyte percentage | Neale Lab UKB | 349,861 | NA | NA | 187,760 | NA | NA | 162,101 | NA | NA |
| Eosinophill percentage | Neale Lab UKB | 349,861 | NA | NA | 187,760 | NA | NA | 162,101 | NA | NA |
| Basophill percentage | Neale Lab UKB | 349,861 | NA | NA | 187,760 | NA | NA | 162,101 | NA | NA |
| Reticulocyte percentage | Neale Lab UKB | 344,728 | NA | NA | 184,923 | NA | NA | 159,805 | NA | NA |
| Reticulocyte count | Neale Lab UKB | 344,729 | NA | NA | 184,923 | NA | NA | 159,806 | NA | NA |
| Mean reticulocyte volume | Neale Lab UKB | 344,728 | NA | NA | 184,922 | NA | NA | 159,806 | NA | NA |
| Mean sphered cell volume | Neale Lab UKB | 344,729 | NA | NA | 184,923 | NA | NA | 159,806 | NA | NA |
| Immature reticulocyte fraction | Neale Lab UKB | 344,728 | NA | NA | 184,922 | NA | NA | 159,806 | NA | NA |
| High light scatter reticulocyte percentage | Neale Lab UKB | 344,729 | NA | NA | 184,923 | NA | NA | 159,806 | NA | NA |
| High light scatter reticulocyte count | Neale Lab UKB | 344,729 | NA | NA | 184,923 | NA | NA | 159,806 | NA | NA |
| Alanine aminotransferase (quantile) | Neale Lab UKB | 344,136 | NA | NA | 184,990 | NA | NA | 159,146 | NA | NA |
| Apoliprotein B (quantile) | Neale Lab UKB | 342,590 | NA | NA | 184,377 | NA | NA | 158,213 | NA | NA |
| Aspartate aminotransferase (quantile) | Neale Lab UKB | 342,990 | NA | NA | 184,364 | NA | NA | 158,626 | NA | NA |
| Urea (quantile) | Neale Lab UKB | 344,052 | NA | NA | 184,888 | NA | NA | 159,164 | NA | NA |
| Calcium (quantile) | Neale Lab UKB | 315,153 | NA | NA | 168,077 | NA | NA | 147,076 | NA | NA |
| Creatinine (quantile) | Neale Lab UKB | 344,104 | NA | NA | 184,912 | NA | NA | 159,192 | NA | NA |
| C-reactive protein (quantile) | Neale Lab UKB | 343,524 | NA | NA | 184,664 | NA | NA | 158,860 | NA | NA |
| Gamma glutamyltransferase (quantile) | Neale Lab UKB | 344,104 | NA | NA | 184,927 | NA | NA | 159,177 | NA | NA |

| | | | | | | | | | | |
|---|---|---|---|---|---|---|---|---|---|---|
| Phosphate (quantile) | Neale Lab UKB | 314,658 | NA | NA | 167,812 | NA | NA | 146,846 | NA | NA |
| Total protein (quantile) | Neale Lab UKB | 314,921 | NA | NA | 167,977 | NA | NA | 146,944 | NA | NA |
| Urate (quantile) | Neale Lab UKB | 343,836 | NA | NA | 184,755 | NA | NA | 159,081 | NA | NA |
| Vitamin D (quantile) | Neale Lab UKB | 329,247 | NA | NA | 174,597 | NA | NA | 154,650 | NA | NA |
| Lipoprotein A (quantile) | Neale Lab UKB | 273,896 | NA | NA | 147,684 | NA | NA | 126,212 | NA | NA |
| Body mass index (BMI) | Neale Lab UKB | 359,983 | NA | NA | 193,570 | NA | NA | 166,413 | NA | NA |
| Whole body fat mass | Neale Lab UKB | 354,244 | NA | NA | 190,941 | NA | NA | 163,303 | NA | NA |
| Obesity | Neale Lab UKB | 361,194 | NA | NA | 194,174 | NA | NA | 167,020 | NA | NA |
| Type 2 diabetes | PMID: 28566273 | 143,520 | NA | NA | | NA | NA | | NA | NA |
| Basal metabolic rate | Neale Lab UKB | 354,825 | NA | NA | 190,987 | NA | NA | 163,838 | NA | NA |
| Systolic blood pressure | Neale Lab UKB | 340,159 | NA | NA | 182,645 | NA | NA | 157,514 | NA | NA |
| Diastolic blood pressure | Neale Lab UKB | 340,162 | NA | NA | 182,647 | NA | NA | 157,515 | NA | NA |
| Fasting blood insulin | PMID: 20081858 | 38,238 | NA | NA | NA | NA | NA | NA | NA | NA |
| Glycated hemoglobin levels （HbA1c） | PMID: 34059833 | 200,622 | NA | NA | NA | NA | NA | NA | NA | NA |
| HDL cholesterol (quantile) | Neale Lab UKB | 315,133 | NA | NA | 168,089 | NA | NA | 147,044 | NA | NA |
| Triglycerides (quantile) | Neale Lab UKB | 343,992 | NA | NA | 184,885 | NA | NA | 159,107 | NA | NA |
| Cholesterol (quantile) | Neale Lab UKB | 344,278 | NA | NA | 184,998 | NA | NA | 159,280 | NA | NA |
| LDL direct (quantile) | Neale Lab UKB | 343,621 | NA | NA | 184,689 | NA | NA | 158,932 | NA | NA |
| IGF-1 (quantile) | Neale Lab UKB | 342,439 | NA | NA | 183,995 | NA | NA | 158,444 | NA | NA |
| Glucose (quantile) | Neale Lab UKB | 314,916 | NA | NA | 167,978 | NA | NA | 146,938 | NA | NA |
| Appendicular lean mass | PMID: 33097823 | 450,243 | NA | NA | 244,730 | NA | NA | 205,513 | NA | NA |
| Loneliness, isolation | Neale Lab UKB | 355,583 | NA | NA | 191,014 | NA | NA | 164,569 | NA | NA |
| Guilty feelings | Neale Lab UKB | 351,907 | NA | NA | 188,864 | NA | NA | 163,043 | NA | NA |
| Risk taking | Neale Lab UKB | 116,746 | NA | NA | 65,410 | NA | NA | 51,336 | NA | NA |
| Ever addicted to any substance or behaviour | Neale Lab UKB | 116,746 | NA | NA | 65,410 | NA | NA | 51,336 | NA | NA |

| | | | | | | | | | | |
|---|---|---|---|---|---|---|---|---|---|---|
| Sensitivity / hurt feelings | Neale Lab UKB | 350,821 | NA | NA | 188,917 | NA | NA | 161,904 | NA | NA |
| Nervous feelings | Neale Lab UKB | 351,829 | NA | NA | 188,370 | NA | NA | 163,459 | NA | NA |
| Worrier / anxious feelings | Neale Lab UKB | 351,833 | NA | NA | 189,455 | NA | NA | 162,378 | NA | NA |
| Tense / 'highly strung' | Neale Lab UKB | 350,159 | NA | NA | 187,755 | NA | NA | 162,404 | NA | NA |
| Neuroticism score | Neale Lab UKB | 293,006 | NA | NA | 155,126 | NA | NA | 137,880 | NA | NA |
| Professional informed about anxiety | Neale Lab UKB | 36,366 | NA | NA | 23,304 | NA | NA | 13,062 | NA | NA |
| Professional informed about depression | Neale Lab UKB | 66,302 | NA | NA | 42,185 | NA | NA | 24,117 | NA | NA |
| Bioavailable testosterone levels | PMID: 32042192 | 382,988 | NA | NA | 188,507 | NA | NA | 178,782 | NA | NA |
| Estradiol levels | PMID: 32042192 | NA | NA | NA | NA | NA | NA | 206,927 | NA | NA |
| Sex hormone-binding globulin levels adjusted for BMI | PMID: 32042192 | 368,929 | NA | NA | 188,908 | NA | NA | 180,094 | NA | NA |
| Sex hormone-binding globulin levels | PMID: 32042192 | 370,125 | NA | NA | 189,473 | NA | NA | 180,726 | NA | NA |
| Total testosterone levels | PMID: 32042192 | 425,097 | NA | NA | 230,454 | NA | NA | 194,453 | NA | NA |
| SHBG (quantile) | Neale Lab UKB | 312,215 | NA | NA | 166,235 | NA | NA | 145,980 | NA | NA |
| Testosterone (quantile) | Neale Lab UKB | 312,102 | NA | NA | 154,364 | NA | NA | 157,738 | NA | NA |
| Albumin (quantile) | Neale Lab UKB | 315,268 | NA | NA | 168,146 | NA | NA | 147,122 | NA | NA |
| Time spent doing vigorous physical activity | Neale Lab UKB | 51,427 | NA | NA | 27,958 | NA | NA | 23,469 | NA | NA |
| Time spent doing moderate physical activity | Neale Lab UKB | 51,427 | NA | NA | 27,958 | NA | NA | 23,469 | NA | NA |
| Time spent doing light physical activity | Neale Lab UKB | 51,427 | NA | NA | 27,958 | NA | NA | 23,469 | NA | NA |
| Ever taken cannabis | Neale Lab UKB | 117,911 | NA | NA | 66,042 | NA | NA | 51,869 | NA | NA |
| Maximum frequency of taking cannabis | Neale Lab UKB | 24,798 | NA | NA | 12,103 | NA | NA | 12,695 | NA | NA |
| Age started smoking in former smokers | Neale Lab UKB | 88,898 | NA | NA | 39,933 | NA | NA | 48,965 | NA | NA |
| Number of cigarettes previously smoked daily | Neale Lab UKB | 84,456 | NA | NA | 39,743 | NA | NA | 44,713 | NA | NA |
| Ever stopped smoking for 6+ months | Neale Lab UKB | 87,937 | NA | NA | 39,609 | NA | NA | 48,328 | NA | NA |
| Ever smoked | Neale Lab UKB | 359,751 | NA | NA | 193,408 | NA | NA | 166,343 | NA | NA |
| Frequency of drinking alcohol | Neale Lab UKB | 117,914 | NA | NA | 66,040 | NA | NA | 51,874 | NA | NA |
| Amount of alcohol drunk on a typical drinking day | Neale Lab UKB | 108,256 | NA | NA | 59,472 | NA | NA | 48,784 | NA | NA |
| Fluid intelligence score | Neale Lab UKB | 117,131 | NA | NA | 62,810 | NA | NA | 54,321 | NA | NA |
| Prospective memory result | Neale Lab UKB | 119,729 | NA | NA | 64,129 | NA | NA | 55,600 | NA | NA |

| | | | | | | | | | | |
|---|---|---|---|---|---|---|---|---|---|---|
| Mean time to correctly identify matches | Neale Lab UKB | 358,695 | NA | NA | 192,893 | NA | NA | 165,802 | NA | NA |
| Age completed full time education | Neale Lab UKB | 240,547 | NA | NA | 131,639 | NA | NA | 108,908 | NA | NA |
| Sleep duration | Neale Lab UKB | 359,020 | NA | NA | 192,810 | NA | NA | 166,210 | NA | NA |
| Sleeplessness / insomnia | Neale Lab UKB | 360,738 | NA | NA | 193,987 | NA | NA | 166,751 | NA | NA |
| Snoring | Neale Lab UKB | 336,320 | NA | NA | 178,728 | NA | NA | 157,592 | NA | NA |
| Daytime dozing / sleeping | Neale Lab UKB | 359,752 | NA | NA | 193,472 | NA | NA | 166,280 | NA | NA |
| Trouble falling asleep | Neale Lab UKB | 45,540 | NA | NA | 30,117 | NA | NA | 15,423 | NA | NA |
| Sleeping too much | Neale Lab UKB | 45,540 | NA | NA | 30,117 | NA | NA | 15,423 | NA | NA |

**Table S2**. Details on the characteristics of GWAS datasets for Finns.

| Phenotype | Data source | Total sample size | Ncase | Ncontrol |
|---|---|---|---|---|
| Anorexia (incl.atypical) | FinnGen | 402,625 | 2,115 | 400,510 |
| Alcohol use disorder, Swedish definition | FinnGen | 412,181 | 24,070 | 388,111 |
| Type 2 diabetes, definitions combined | FinnGen | 400,197 | 65,085 | 335,112 |
| Obesity | FinnGen | 412,055 | 23,971 | 388,084 |
| Polycystic ovarian syndrome, consortium definition | FinnGen | 230,310 | 34,388 | 195,922 |
| Disorders of lipoprotein metabolism and other lipidaemias | FinnGen | 395,429 | 43,197 | 352,232 |
| Metabolic disorders | FinnGen | 412,181 | 59,949 | 352,232 |
| Obesity due to excess calories | FinnGen | 405,186 | 16,976 | 388,210 |
| Hyperlipidaemia, other/unspecified | FinnGen | 365,889 | 13,657 | 352,232 |
| Obesity, other/unspecified | FinnGen | 400,498 | 12,288 | 388,210 |
| Sleep disorders (combined) | FinnGen | 408,074 | 49,880 | 358,194 |
| Any mental disorder | FinnGen | 412,181 | 110,302 | 301,879 |
| Anxiety disorders (more control exclusions) | FinnGen | 346,542 | 44,663 | 301,879 |
| Substance abuse (more control exclusions) | FinnGen | 327,338 | 25,459 | 301,879 |
| Suicide or other Intentional self-harm | FinnGen | 412,181 | 9,771 | 402,410 |
| Schizophrenia or delusion (more control exclusions) | FinnGen | 315,617 | 13,738 | 301,879 |
| Personality disorders (more control exclusions) | FinnGen | 312,769 | 10,894 | 301,875 |
| KRA_PSY_EATING (more control exclusions) | FinnGen | 305,000 | 3,121 | 301,879 |
| Any mental disorder, or suicide (or attempt), or psychic disorders complicating pregnancy, partum or puerperium or nerve system disorders (more control exclusions) | FinnGen | 312,989 | 11,873 | 301,116 |
| Nonorganic sleeping disorders (more control exclusions) | FinnGen | 307,843 | 5,964 | 301,879 |
| Body-mass index, inverse-rank normalized | FinnGen | 290,820 | NA | NA |
| Depression or dysthymia | FinnGen | 297,542 | 54,733 | 242,809 |
| Use of hypnotics and sedatives | FinnGen | 412,181 | 71,955 | 340,226 |
| Dementia, including avohilmo | FinnGen | 412,181 | 20,338 | 391,843 |
| Mood [affective] disorders | FinnGen | 412,181 | 52,891 | 359,290 |
| Dementia | FinnGen | 407,717 | 19,157 | 388,560 |
| Depression | FinnGen | 406,986 | 47,696 | 359,290 |
| Recurrent or chronic depression | FinnGen | 265,431 | 19,388 | 246,043 |
| Alcohol use disorder, ICD-based | FinnGen | 412,181 | 17,197 | 394,984 |
| All anxiety disorders | FinnGen | 395,718 | 27,664 | 368,054 |

| | | | | |
|---|---|---|---|---|
| Alcohol dependence | FinnGen | 396,592 | 10,707 | 385,885 |
| Schizophrenia | FinnGen | 405,094 | 6,708 | 398,386 |
| Acute and transient psychotic disorders | FinnGen | 402,850 | 4,464 | 398,386 |
| Disorders of psychological development | FinnGen | 412,181 | 4,233 | 407,948 |
| Mental and behavioural disorders due to multiple drug use and use of multiple drugs and use of other psychoactive substances | FinnGen | 407,440 | 2,491 | 404,949 |
| Kela-code for severe mental illness | FinnGen | 410,331 | 16,578 | 393,753 |
| Insomnia | FinnGen | 410,030 | 4,801 | 405,229 |
| Emotional disorders and disorders of social functioning with onset specific to childhood | FinnGen | 406,952 | 1,799 | 405,153 |
| Persistent delusional disorders | FinnGen | 400,940 | 2,554 | 398,386 |
| Delirium, not induced by alcohol and other psychoactive substances | FinnGen | 391,931 | 3,371 | 388,560 |
| Bipolar affective disorders | FinnGen | 366,859 | 7,569 | 359,290 |
| Other anxiety disorders | FinnGen | 386,957 | 18,903 | 368,054 |
| Other reaction to severe stress, and adjustment disorders | FinnGen | 383,280 | 15,226 | 368,054 |
| Persistent mood disorders | FinnGen | 366,651 | 7,361 | 359,290 |
| Generalized anxiety disorder | FinnGen | 373,334 | 5,280 | 368,054 |
| Phobic anxiety disorders | FinnGen | 372,901 | 4,847 | 368,054 |
| Panic disorder | FinnGen | 373,422 | 5,368 | 368,054 |
| Mixed and other personality disorders | FinnGen | 403,436 | 2,817 | 400,619 |
| Emotionally unstable personality disorder | FinnGen | 405,170 | 4,551 | 400,619 |
| Psychotic depression | FinnGen | 248,501 | 2,149 | 246,352 |
| Behavioural and emotional disorders with onset usually occurring in childhood and adolescence | FinnGen | 412,181 | 7,028 | 405,153 |
| Behavioural syndromes associated with physiological disturbances and physical factors | FinnGen | 412,181 | 11,671 | 400,510 |

**Table S3**. Global genetic correlations between anorexia nervosa and potential risk factors in European populations.

| Catalog | Trait | Combine | | Female | | Male | |
|---|---|---|---|---|---|---|---|
| | | rg | p | rg | p | rg | p |
| Psychiatric disorder/trait | Schizophrenia | **0.223** | **2.59E-17** | **-0.180** | **3.95E-06** | **-0.234** | **4.10E-16** |
| | Bipolar Disorder | **0.221** | **5.81E-12** | | | | |
| | Major Depressive Disorder | **0.260** | **3.42E-16** | | | | |
| | Obsessive-Compulsive Disorde | **0.455** | **1.40E-08** | | | | |
| | Loneliness, isolation | 0.121 | 0.005 | 0.092 | 0.069 | 0.1487 | 0.013 |
| | Guilty feelings | **0.262** | **8.57E-11** | **0.265** | **9.76E-09** | **0.244** | **1.49E-06** |
| | Risk taking | 0.019 | 0.577 | 0.013 | 0.768 | 0.019 | 0.636 |
| | Ever addicted to any substance or behaviour | **0.385** | **1.61E-05** | 0.378 | 0.022 | **0.344** | **2.00E-04** |
| | Sensitivity / hurt feelings | **0.190** | **6.13E-08** | **0.138** | **0.001** | **0.245** | **8.12E-08** |
| | Nervous feelings | **0.290** | **1.87E-17** | **0.288** | **5.58E-13** | **0.274** | **1.81E-08** |
| | Worrier / anxious feelings | **0.331** | **3.63E-19** | **0.326** | **1.42E-13** | **0.307** | **1.78E-11** |
| | Tense / 'highly strung' | **0.306** | **7.76E-15** | **0.301** | **1.51E-11** | **0.293** | **2.83E-08** |
| | Neuroticism score | **0.232** | **7.68E-12** | **0.234** | **2.49E-09** | **0.229** | **1.10E-07** |
| | Professional informed about anxiety | 0.230 | 0.017 | 0.170 | 0.103 | 0.289 | 0.161 |
| | Professional informed about depression | 0.213 | 0.0081 | 0.117 | 0.251 | 0.331 | 0.010 |
| Sex hormone relared traits | Bioavailable testosterone levels | **-0.148** | **5.05E-06** | **-0.275** | **1.22E-17** | 0.038 | 0.293 |
| | Sex hormone-binding globulin levels adjusted for BMI | **0.096** | **2.00E-04** | **0.103** | **2.00E-04** | 0.083 | 0.003 |
| | Sex hormone-binding globulin levels | **0.229** | **6.37E-17** | **0.278** | **2.83E-20** | **0.156** | **1.32E-07** |
| | Total testosterone levels | 0.045 | 0.143 | -0.095 | 0.003 | **0.166** | **7.89E-08** |
| | SHBG (quantile) | **0.246** | **2.69E-17** | **0.287** | **7.19E-20** | **0.164** | **2.610E-07** |
| | Testosterone (quantile) | 0.084 | 0.019 | -0.083 | 0.034 | **0.158** | **3.650E-06** |
| | Albumin (quantile) | 0.028 | 0.347 | 0.069 | 0.056 | -0.020 | 0.595 |
| Sleep | Sleep duration | -0.085 | 0.006 | **-0.122** | **9.00E-04** | -0.039 | 0.346 |
| | Sleeplessness / insomnia | 0.060 | 0.084 | 0.019 | 0.626 | 0.100 | 0.022 |
| | Snoring | **0.261** | **1.39E-14** | **0.272** | **6.54E-13** | **0.233** | **1.10E-06** |
| | Daytime dozing / sleeping (narcolepsy) | -0.066 | 0.094 | -0.069 | 0.133 | -0.064 | 0.218 |
| | Trouble falling asleep | -0.210 | 0.021 | -0.228 | 0.044 | -0.169 | 0.265 |
| | Sleeping too much | -0.002 | 0.984 | -0.066 | 0.648 | 0.106 | 0.542 |
| Metabolic traits | Body mass index (BMI) | **-0.329** | **4.09E-35** | **-0.387** | **1.67E-40** | **-0.235** | **1.76E-15** |

| | | | | | | | |
|---|---|---|---|---|---|---|---|
| | Whole body fat mass | **-0.342** | **1.30E-35** | **-0.403** | **2.22E-41** | **-0.256** | **2.08E-16** |
| | Obesity | -0.164 | 0.092 | -0.179 | 0.101 | NA | NA |
| | Type 2 diabetes | **-0.246** | **5.41E-08** | | | | |
| | Basal metabolic rate | **-0.195** | **2.58E-14** | **-0.252** | **1.24E-18** | **-0.117** | **2.80E-05** |
| | Systolic blood pressure, automated reading | **-0.171** | **1.03E-10** | **-0.183** | **1.16E-10** | **-0.138** | **9.78E-05** |
| | Diastolic blood pressure, automated reading | **-0.198** | **4.65E-11** | **-0.199** | **3.18E-10** | **-0.176** | **1.56E-05** |
| | Fasting blood insulin | **-0.294** | **7.44E-05** | | | | |
| | Glycated hemoglobin levels（HbA1c） | -0.131 | 1.30E-03 | | | | |
| | HDL cholesterol (quantile) | **0.230** | **3.07E-14** | **0.266** | **6.76E-16** | **0.176** | **4.42E-07** |
| | Triglycerides (quantile) | **-0.213** | **1.53E-09** | **-0.220** | **1.07E-10** | **-0.169** | **9.821E-06** |
| | Cholesterol (quantile) | 0.026 | 0.471 | 0.009 | 0.818 | 0.057 | 0.209 |
| | LDL direct (quantile) | -0.033 | 0.416 | -0.082 | 0.108 | 0.037 | 0.491 |
| | IGF-1 (quantile) | 0.019 | 0.470 | -0.018 | 0.542 | 0.048 | 0.106 |
| | Glucose (quantile) | **-0.142** | **2.81E-05** | -0.107 | 0.004 | **-0.157** | **0.001** |
| | Appendicular lean mass | -0.039 | 0.098 | -0.014 | 0.591 | -0.039 | 0.026 |
| blood-based biomarkers | White blood cell (leukocyte) count | **-0.179** | **8.88E-09** | **-0.182** | **1.08E-07** | **-0.171** | **4.708E-06** |
| | Red blood cell (erythrocyte) count | **-0.129** | **2.00E-06** | **-0.150** | **2.04E-07** | -0.099 | 0.002 |
| | Haemoglobin concentration | -0.094 | 0.002 | -0.101 | 0.003 | -0.083 | 0.018 |
| | Haematocrit percentage | -0.092 | 0.002 | -0.090 | 0.007 | -0.089 | 0.013 |
| | Mean corpuscular volume | **0.100** | **1.00E-04** | **0.129** | **6.06E-06** | 0.052 | 0.095 |
| | Mean corpuscular haemoglobin | 0.084 | 0.002 | **0.106** | **3.00E-04** | 0.057 | 0.067 |
| | Mean corpuscular haemoglobin concentration | -0.006 | 0.856 | -0.041 | 0.350 | 0.036 | 0.410 |
| | Red blood cell (erythrocyte) distribution width | -0.055 | 0.046 | -0.064 | 0.036 | -0.051 | 0.113 |
| | Platelet count | -0.017 | 0.456 | -0.025 | 0.339 | -0.003 | 0.910 |
| | Platelet crit | -0.043 | 0.098 | -0.062 | 0.034 | -0.013 | 0.704 |
| | Mean platelet (thrombocyte) volume | -0.027 | 0.217 | -0.045 | 0.068 | -0.013 | 0.616 |
| | Platelet distribution width | 0.018 | 0.440 | 0.017 | 0.547 | 0.026 | 0.351 |
| | Lymphocyte count | **-0.104** | **0.001** | -0.111 | 0.001 | -0.089 | 0.017 |
| | Monocyte count | **-0.106** | **1.00E-04** | -0.095 | 0.003 | **-0.129** | **9.369E-05** |
| | Neutrophill count | **-0.182** | **1.64E-09** | **-0.179** | **1.21E-07** | **-0.171** | **1.848E-06** |
| | Eosinophill count | -0.063 | 0.034 | -0.087 | 0.009 | -0.037 | 0.282 |
| | Basophill count | -0.108 | 0.031 | -0.097 | 0.138 | -0.123 | 0.048 |
| | Nucleated red blood cell count | NA | NA | NA | NA | NA | NA |

| | | | | | | | |
|---|---|---|---|---|---|---|---|
| | Lymphocyte percentage | 0.057 | 0.051 | 0.054 | 0.094 | 0.067 | 0.057 |
| | Monocyte percentage | 0.034 | 0.214 | 0.052 | 0.099 | 0.001 | 0.983 |
| | Eosinophill percentage | 0.000 | 0.998 | -0.018 | 0.562 | 0.015 | 0.595 |
| | Basophill percentage | 0.024 | 0.556 | 0.039 | 0.400 | 0.003 | 0.952 |
| | Reticulocyte percentage | **-0.211** | **5.03E-11** | **-0.217** | **1.84E-09** | **-0.187** | **1.688E-07** |
| | Reticulocyte count | **-0.229** | **2.32E-12** | **-0.239** | **5.53E-11** | **-0.198** | **4.175E-08** |
| | Mean reticulocyte volume | 0.083 | 0.002 | 0.077 | 0.010 | 0.069 | 0.031 |
| | Mean sphered cell volume | 0.048 | 0.138 | 0.089 | 0.001 | **0.109** | **3.000E-04** |
| | Immature reticulocyte fraction | **-0.190** | **1.15E-08** | **-0.212** | **3.15E-08** | **-0.154** | **2.371E-05** |
| | High light scatter reticulocyte percentage | **-0.220** | **5.77E-12** | **-0.231** | **9.67E-11** | **-0.189** | **4.551E-08** |
| | High light scatter reticulocyte count | **-0.235** | **4.86E-13** | **-0.250** | **3.55E-12** | **-0.198** | **1.361E-08** |
| | Alanine aminotransferase (quantile) | **-0.192** | **3.63E-10** | **-0.163** | **6.74E-06** | **-0.204** | **4.836E-09** |
| | Apoliprotein B (quantile) | -0.095 | 0.027 | -0.134 | 0.011 | -0.031 | 0.617 |
| | Aspartate aminotransferase (quantile) | 0.004 | 0.893 | 0.051 | 0.131 | -0.040 | 0.249 |
| | Urea (quantile) | -0.002 | 0.940 | -0.048 | 0.167 | 0.054 | 0.144 |
| | Calcium (quantile) | -0.032 | 0.188 | -0.041 | 0.169 | -0.019 | 0.569 |
| | Creatinine (quantile) | -0.064 | 0.036 | -0.075 | 0.022 | -0.037 | 0.266 |
| | C-reactive protein (quantile) | **-0.273** | **9.90E-16** | **-0.297** | **2.15E-17** | **-0.223** | **6.854E-09** |
| | Gamma glutamyltransferase (quantile) | **-0.151** | **6.94E-09** | **-0.141** | **1.58E-06** | **-0.159** | **3.844E-08** |
| | Phosphate (quantile) | -0.016 | 0.595 | -0.009 | 0.800 | -0.022 | 0.531 |
| | Total protein (quantile) | -0.093 | 0.001 | -0.105 | 0.002 | -0.072 | 0.057 |
| | Urate (quantile) | **-0.176** | **7.87E-09** | **-0.214** | **4.26E-10** | -0.091 | 0.009 |
| | Vitamin D (quantile) | 0.060 | 0.116 | 0.084 | 0.080 | 0.018 | 0.650 |
| | Lipoprotein A (quantile) | -0.111 | 0.371 | -0.139 | 0.584 | NA | NA |
| Substance abuse | Ever taken cannabis | **0.229** | **2.21E-06** | 0.171 | 0.003 | **0.259** | **1.92E-05** |
| | Maximum frequency of taking cannabis | 0.077 | 0.430 | -0.083 | 0.508 | 0.206 | 0.109 |
| | Age started smoking in former smokers | **0.222** | **7.98E-05** | **0.372** | **5.05E-05** | 0.123 | 0.071 |
| | Number of cigarettes previously smoked daily | -0.103 | 0.037 | -0.111 | 0.094 | -0.090 | 0.157 |
| | Ever stopped smoking for 6+ months | 0.116 | 0.120 | 0.143 | 0.125 | 0.084 | 0.537 |
| | Ever smoked | 0.029 | 0.401 | 0.035 | 0.379 | 0.025 | 0.559 |
| | Frequency of drinking alcohol | 0.047 | 0.296 | 0.105 | 0.038 | -0.042 | 0.547 |
| | Amount of alcohol drunk on a typical drinking day | -0.139 | 0.012 | -0.172 | 0.051 | -0.121 | 0.064 |
| Cognitive function | Fluid intelligence score | **0.125** | **0.001** | 0.116 | 0.010 | **0.155** | **4.00E-04** |

| | | | | | | | |
|---|---|---|---|---|---|---|---|
| | Prospective memory result | -0.007 | 0.912 | 0.019 | 0.799 | -0.041 | 0.570 |
| | Mean time to correctly identify matches | 0.049 | 0.123 | 0.057 | 0.148 | 0.037 | 0.329 |
| Educational attainment | Age completed full time education | **0.146** | **2.00E-04** | 0.140 | 0.002 | 0.148 | 0.002 |

**Table S4**. Global genetic correlations between anorexia nervosa and potential risk factors in Finnish populations.

| **category** | **Trait** | **rg** | **p** |
|---|---|---|---|
| Psychiatric disorder/trait | Behavioural syndromes associated with physiological disturbances and physical factors | 0.523 | 0.074 |
| | Other anxiety disorders | 0.941 | 0.091 |
| | Recurrent or chronic depression | 0.707 | 0.093 |
| | All anxiety disorders | **0.907** | **0.093** |
| | Mood [affective] disorders | 0.649 | 0.095 |
| | Psychotic depression | 0.661 | 0.096 |
| | Persistent mood disorders | 0.898 | 0.103 |
| | Depression | 0.448 | 0.111 |
| | Generalized anxiety disorder | 0.983 | 0.117 |
| | Behavioural and emotional disorders with onset usually occurring in childhood and adolescence | 0.794 | 0.128 |
| | Emotionally unstable personality disorder | 0.726 | 0.132 |
| | Mental and behavioural disorders due to multiple drug use and use of multiple drugs and use of other psychoactive substances | 0.763 | 0.133 |
| | Kela-code for severe mental illness | 0.900 | 0.151 |
| | Use of hypnotics and sedatives | 0.449 | 0.156 |
| | Alcohol dependence | 0.444 | 0.157 |
| | Mixed and other personality disorders | 0.865 | 0.159 |
| | Panic disorder | 0.722 | 0.159 |
| | Other reaction to severe stress, and adjustment disorders | 0.635 | 0.167 |
| | Phobic anxiety disorders | 0.629 | 0.183 |
| | Depression or dysthymia | 1.082 | 0.192 |
| | Alcohol use disorder, ICD-based | 0.333 | 0.209 |
| | Bipolar affective disorders | 0.483 | 0.227 |
| | Emotional disorders and disorders of social functioning with onset specific to childhood | 0.629 | 0.269 |
| | Insomnia | 0.605 | 0.269 |
| | Persistent delusional disorders | 0.460 | 0.298 |
| | Disorders of psychological development | 0.489 | 0.381 |
| | Acute and transient psychotic disorders | 0.275 | 0.431 |
| | Schizophrenia | 0.231 | 0.474 |
| | Dementia | 0.255 | 0.513 |

| | | | |
|---|---|---|---|
| | Delirium, not induced by alcohol and other psychoactive substances | -0.355 | 0.621 |
| | Dementia, including avohilmo | 0.139 | 0.682 |
| | Anxiety disorders (more control exclusions) | 0.689 | 0.095 |
| | Any mental disorder, or suicide (or attempt), or psychic disorders complicating pregnancy, partum or puerperium or nerve system disorders (more control exclusions) | 0.610 | 0.099 |
| | KRA_PSY_EATING (more control exclusions) | 0.730 | 0.100 |
| | Personality disorders (more control exclusions) | 0.709 | 0.112 |
| | Any mental disorder | 0.502 | 0.143 |
| | Suicide or other Intentional self-harm | 0.521 | 0.147 |
| | Substance abuse (more control exclusions) | 0.334 | 0.185 |
| | Nonorganic sleeping disorders (more control exclusions) | 0.748 | 0.186 |
| | Schizophrenia or delusion (more control exclusions) | 0.358 | 0.254 |
| Substance abuse | Alcohol use disorder, Swedish definition | 0.194 | 0.361 |
| Metabolic traits | Type 2 diabetes, definitions combined | -0.229 | 0.198 |
| | Body-mass index, inverse-rank normalized | -0.388 | 0.085 |
| | Obesity | **-0.480** | **0.078** |
| | Obesity, other/unspecified | -0.475 | 0.084 |
| | Obesity due to excess calories | -0.423 | 0.130 |
| | Polycystic ovarian syndrome, consortium definition | 0.439 | 0.181 |
| | Disorders of lipoprotein metabolism and other lipidaemias | -0.306 | 0.243 |
| | Hyperlipidaemia, other/unspecified | -0.360 | 0.326 |
| | Metabolic disorders | -0.174 | 0.473 |
| Sleep | Sleep disorders (combined) | 0.137 | 0.479 |

**Table S5**. Mendelian randomization results: effect of anorexia nervosa on other phenotypes.

| **Exposure** | **Outcome_Trait** | **MR_method** | **nsnp** | **b** | **se** | **pval** | **lo_ci** | **up_ci** | **or** | **or_lci95** | **or_uci95** |
|---|---|---|---|---|---|---|---|---|---|---|---|
| Anorexia Nervosa | Body mass index (BMI) | MR Egger | 7 | -0.003 | 0.094 | 0.974 | -0.187 | 0.181 | 0.997 | 0.829 | 1.198 |
| | | Weighted median | | -0.067 | 0.016 | **4.52E-05** | -0.099 | -0.035 | 0.935 | 0.906 | 0.966 |
| | | Inverse variance weighted | | -0.070 | 0.033 | 0.036 | -0.135 | -0.004 | 0.933 | 0.874 | 0.996 |
| | | Simple mode | | -0.084 | 0.024 | 0.014 | -0.132 | -0.036 | 0.919 | 0.876 | 0.964 |
| | | Weighted mode | | -0.081 | 0.026 | 0.020 | -0.131 | -0.031 | 0.922 | 0.877 | 0.970 |
| | Whole body fat mass | MR Egger | 7 | -0.012 | 0.094 | 0.904 | -0.196 | 0.172 | 0.988 | 0.822 | 1.188 |
| | | Weighted median | | -0.078 | 0.017 | **4.54E-06** | -0.112 | -0.045 | 0.925 | 0.894 | 0.956 |
| | | Inverse variance weighted | | -0.077 | 0.033 | 0.020 | -0.142 | -0.012 | 0.926 | 0.867 | 0.988 |
| | | Simple mode | | -0.088 | 0.024 | 0.011 | -0.135 | -0.041 | 0.916 | 0.873 | 0.960 |
| | | Weighted mode | | -0.087 | 0.025 | 0.013 | -0.136 | -0.038 | 0.916 | 0.872 | 0.963 |
| | HDL cholesterol (quantile) | MR Egger | 7 | 0.076 | 0.052 | 0.203 | -0.026 | 0.178 | 1.079 | 0.975 | 1.195 |
| | | Weighted median | | 0.049 | 0.014 | **5.61E-04** | 0.021 | 0.077 | 1.050 | 1.021 | 1.080 |
| | | Inverse variance weighted | | 0.040 | 0.018 | 0.030 | 0.004 | 0.076 | 1.041 | 1.004 | 1.079 |
| | | Simple mode | | 0.055 | 0.016 | 0.014 | 0.024 | 0.086 | 1.056 | 1.024 | 1.090 |
| | | Weighted mode | | 0.052 | 0.017 | 0.023 | 0.018 | 0.085 | 1.053 | 1.018 | 1.089 |
| | SHBG (quantile) | MR Egger | 7 | 0.055 | 0.046 | 0.283 | -0.035 | 0.144 | 1.056 | 0.966 | 1.155 |
| | | Weighted median | | 0.049 | 0.014 | **5.06E-04** | 0.021 | 0.076 | 1.050 | 1.022 | 1.079 |
| | | Inverse variance weighted | | 0.041 | 0.015 | 0.008 | 0.011 | 0.072 | 1.042 | 1.011 | 1.074 |
| | | Simple mode | | 0.054 | 0.017 | 0.020 | 0.020 | 0.089 | 1.056 | 1.021 | 1.093 |
| | | Weighted mode | | 0.051 | 0.015 | 0.016 | 0.021 | 0.080 | 1.052 | 1.021 | 1.084 |
| | Sex hormone-binding globulin levels | MR Egger | 7 | 0.026 | 0.029 | 0.406 | -0.031 | 0.084 | 1.027 | 0.970 | 1.088 |
| | | Weighted median | | 0.028 | 0.006 | **7.52E-06** | 0.016 | 0.040 | 1.028 | 1.016 | 1.041 |
| | | Inverse variance weighted | | 0.024 | 0.010 | 0.013 | 0.005 | 0.043 | 1.024 | 1.005 | 1.044 |
| | | Simple mode | | 0.028 | 0.008 | 0.013 | 0.012 | 0.044 | 1.028 | 1.013 | 1.044 |
| | | Weighted mode | | 0.027 | 0.007 | 0.009 | 0.013 | 0.041 | 1.028 | 1.013 | 1.042 |
| | Mean corpuscular volume | MR Egger | 7 | -0.088 | 0.030 | 0.031 | -0.146 | -0.030 | 0.916 | 0.864 | 0.970 |
| | | Weighted median | | -0.050 | 0.015 | 0.001 | -0.080 | -0.021 | 0.951 | 0.923 | 0.979 |
| | | Inverse variance weighted | | -0.045 | 0.012 | **2.06E-04** | -0.069 | -0.021 | 0.956 | 0.934 | 0.979 |

| | | | | | | | | | | | |
|---|---|---|---|---|---|---|---|---|---|---|---|
| | | Simple mode | | -0.051 | 0.026 | 0.091 | -0.102 | -0.001 | 0.950 | 0.903 | 0.999 |
| | | Weighted mode | | -0.052 | 0.025 | 0.086 | -0.102 | -0.002 | 0.949 | 0.903 | 0.998 |
| | Red blood cell (erythrocyte) distribution width | MR Egger | 7 | -0.087 | 0.029 | 0.030 | -0.144 | -0.030 | 0.916 | 0.866 | 0.970 |
| | | Weighted median | | -0.058 | 0.015 | **1.59E-04** | -0.088 | -0.028 | 0.944 | 0.916 | 0.973 |
| | | Inverse variance weighted | | -0.050 | 0.011 | **4.74E-06** | -0.071 | -0.029 | 0.951 | 0.931 | 0.972 |
| | | Simple mode | | -0.070 | 0.025 | 0.030 | -0.118 | -0.021 | 0.933 | 0.888 | 0.979 |
| | | Weighted mode | | -0.070 | 0.025 | 0.032 | -0.120 | -0.021 | 0.932 | 0.887 | 0.979 |
| | Reticulocyte percentage | MR Egger | 7 | -0.258 | 0.062 | 0.009 | -0.379 | -0.136 | 0.773 | 0.685 | 0.872 |
| | | Weighted median | | -0.072 | 0.020 | **4.53E-04** | -0.112 | -0.032 | 0.931 | 0.894 | 0.969 |
| | | Inverse variance weighted | | -0.113 | 0.031 | **2.78E-04** | -0.174 | -0.052 | 0.893 | 0.840 | 0.949 |
| | | Simple mode | | -0.063 | 0.035 | 0.124 | -0.132 | 0.006 | 0.939 | 0.876 | 1.006 |
| | | Weighted mode | | -0.053 | 0.024 | 0.068 | -0.100 | -0.006 | 0.948 | 0.905 | 0.994 |
| | Reticulocyte count | MR Egger | 7 | -0.237 | 0.062 | 0.012 | -0.358 | -0.116 | 0.789 | 0.699 | 0.890 |
| | | Weighted median | | -0.066 | 0.021 | 0.001 | -0.107 | -0.026 | 0.936 | 0.899 | 0.974 |
| | | Inverse variance weighted | | -0.110 | 0.029 | **1.66E-04** | -0.167 | -0.053 | 0.896 | 0.846 | 0.949 |
| | | Simple mode | | -0.059 | 0.034 | 0.133 | -0.126 | 0.008 | 0.942 | 0.881 | 1.008 |
| | | Weighted mode | | -0.053 | 0.024 | 0.071 | -0.100 | -0.006 | 0.949 | 0.905 | 0.994 |
| | Immature reticulocyte fraction | MR Egger | 7 | -0.153 | 0.049 | 0.027 | -0.249 | -0.056 | 0.858 | 0.779 | 0.945 |
| | | Weighted median | | -0.065 | 0.018 | **2.58E-04** | -0.100 | -0.030 | 0.937 | 0.904 | 0.970 |
| | | Inverse variance weighted | | -0.083 | 0.020 | **3.56E-05** | -0.122 | -0.043 | 0.921 | 0.885 | 0.957 |
| | | Simple mode | | -0.118 | 0.044 | 0.036 | -0.205 | -0.032 | 0.888 | 0.815 | 0.968 |
| | | Weighted mode | | -0.034 | 0.031 | 0.320 | -0.094 | 0.027 | 0.967 | 0.910 | 1.028 |
| | High light scatter reticulocyte percentage | MR Egger | 7 | -0.257 | 0.067 | 0.012 | -0.388 | -0.126 | 0.773 | 0.678 | 0.882 |
| | | Weighted median | | -0.074 | 0.021 | **4.82E-04** | -0.116 | -0.032 | 0.929 | 0.891 | 0.968 |
| | | Inverse variance weighted | | -0.120 | 0.032 | **1.43E-04** | -0.182 | -0.058 | 0.887 | 0.834 | 0.944 |

| | | | | | | | | | | | |
|---|---|---|---|---|---|---|---|---|---|---|---|
| | | Simple mode | | -0.063 | 0.046 | 0.216 | -0.153 | 0.026 | 0.939 | 0.858 | 1.027 |
| | | Weighted mode | | -0.052 | 0.026 | 0.091 | -0.103 | -0.001 | 0.949 | 0.902 | 0.999 |
| | High light scatter reticulocyte count | MR Egger | 7 | -0.244 | 0.068 | 0.016 | -0.377 | -0.110 | 0.784 | 0.686 | 0.896 |
| | | Weighted median | | -0.072 | 0.021 | **7.73E-04** | -0.114 | -0.030 | 0.931 | 0.893 | 0.971 |
| | | Inverse variance weighted | | -0.119 | 0.030 | **9.43E-05** | -0.179 | -0.059 | 0.888 | 0.836 | 0.943 |
| | | Simple mode | | -0.063 | 0.047 | 0.231 | -0.155 | 0.030 | 0.939 | 0.856 | 1.030 |
| | | Weighted mode | | -0.054 | 0.023 | 0.058 | -0.099 | -0.009 | 0.947 | 0.905 | 0.991 |
| | Urate (quantile) | MR Egger | 7 | -0.010 | 0.025 | 0.694 | -0.059 | 0.039 | 0.990 | 0.942 | 1.039 |
| | | Weighted median | | -0.035 | 0.012 | 0.003 | -0.059 | -0.012 | 0.965 | 0.943 | 0.988 |
| | | Inverse variance weighted | | -0.032 | 0.009 | **4.81E-04** | -0.050 | -0.014 | 0.968 | 0.951 | 0.986 |
| | | Simple mode | | -0.040 | 0.017 | 0.055 | -0.073 | -0.007 | 0.961 | 0.930 | 0.993 |
| | | Weighted mode | | -0.038 | 0.018 | 0.076 | -0.072 | -0.003 | 0.963 | 0.930 | 0.997 |

nsnp, number of SNPs used in the MR analysis; lo_ci, the lower limit of the 95% confidence interval, representing the lower bound of the effect estimate; up_ci, the upper limit of the 95% confidence interval, representing the upper bound of the effect estimate; or, Odds Ratio, indicating the ratio relationship between exposure and outcome; or_lci95, the lower limit of the 95% confidence interval for the odds ratio; or_uci95, the upper limit of the 95% confidence interval for the odds ratio.

**Table S6.** Mendelian randomization results: effect of other phenotypes on anorexia nervosa.

| **Exposure** | **Outcome_Trait** | **MR_method** | **nsnp** | **b** | **se** | **pval** | **lo_ci** | **up_ci** | **or** | **or_lci95** | **or_uci95** |
|---|---|---|---|---|---|---|---|---|---|---|---|
| Schizophrenia | Anorexia Nervosa | MR Egger | 245 | 0.148 | 0.081 | 0.068 | -0.010 | 0.306 | 1.158 | 0.990 | 1.356 |
| | | Weighted median | | 0.164 | 0.026 | **3.32E-10** | 0.113 | 0.215 | 1.177 | 1.119 | 1.241 |
| | | Inverse variance weighted | | 0.166 | 0.021 | **6.81E-15** | 0.124 | 0.207 | 1.180 | 1.132 | 1.230 |
| | | Simple mode | | 0.219 | 0.086 | 0.012 | 0.049 | 0.388 | 1.244 | 1.051 | 1.472 |
| | | Weighted mode | | 0.192 | 0.076 | 0.013 | 0.042 | 0.341 | 1.210 | 1.043 | 1.409 |
| Neuroticism score | | MR Egger | 92 | -0.206 | 0.613 | 0.738 | -1.406 | 0.995 | 0.814 | 0.245 | 2.705 |
| | | Weighted median | | 0.372 | 0.148 | 0.012 | 0.082 | 0.662 | 1.451 | 1.086 | 1.938 |
| | | Inverse variance weighted | | 0.472 | 0.126 | **1.91E-04** | 0.224 | 0.720 | 1.603 | 1.251 | 2.054 |
| | | Simple mode | | 0.384 | 0.362 | 0.292 | -0.327 | 1.094 | 1.468 | 0.721 | 2.987 |
| | | Weighted mode | | 0.384 | 0.329 | 0.246 | -0.260 | 1.028 | 1.468 | 0.771 | 2.796 |
| Trouble falling asleep | | MR Egger | 717 | -0.075 | 0.125 | 0.546 | -0.320 | 0.169 | 0.928 | 0.726 | 1.184 |
| | | Weighted median | | -0.249 | 0.053 | **3.04E-06** | -0.353 | -0.144 | 0.780 | 0.703 | 0.866 |
| | | Inverse variance weighted | | -0.319 | 0.041 | **7.24E-15** | -0.399 | -0.238 | 0.727 | 0.671 | 0.788 |
| | | Simple mode | | -0.164 | 0.228 | 0.472 | -0.611 | 0.283 | 0.849 | 0.543 | 1.327 |
| | | Weighted mode | | -0.073 | 0.158 | 0.644 | -0.382 | 0.236 | 0.930 | 0.682 | 1.267 |
| Whole body fat mass | | MR Egger | 666 | 0.034 | 0.139 | 0.807 | -0.239 | 0.307 | 1.035 | 0.787 | 1.360 |
| | | Weighted median | | -0.289 | 0.056 | **2.93E-07** | -0.400 | -0.179 | 0.749 | 0.671 | 0.836 |
| | | Inverse variance weighted | | -0.339 | 0.044 | **1.40E-14** | -0.425 | -0.253 | 0.713 | 0.654 | 0.777 |
| | | Simple mode | | -0.222 | 0.233 | 0.341 | -0.679 | 0.235 | 0.801 | 0.507 | 1.265 |
| | | Weighted mode | | -0.244 | 0.228 | 0.286 | -0.692 | 0.204 | 0.783 | 0.501 | 1.226 |
| Basal metabolic rate | | MR Egger | 1236 | 0.082 | 0.119 | 0.490 | -0.152 | 0.316 | 1.086 | 0.859 | 1.372 |
| | | Weighted median | | -0.122 | 0.058 | 0.035 | -0.236 | -0.008 | 0.885 | 0.790 | 0.992 |
| | | Inverse variance weighted | | -0.220 | 0.043 | **3.16E-07** | -0.304 | -0.136 | 0.802 | 0.738 | 0.873 |
| | | Simple mode | | 0.040 | 0.252 | 0.875 | -0.454 | 0.534 | 1.040 | 0.635 | 1.705 |
| | | Weighted mode | | -0.091 | 0.203 | 0.653 | -0.490 | 0.307 | 0.913 | 0.613 | 1.359 |
| HDL cholesterol (quantile) | | MR Egger | 902 | 0.011 | 0.037 | 0.770 | -0.062 | 0.084 | 1.011 | 0.940 | 1.087 |
| | | Weighted median | | 0.091 | 0.037 | 0.014 | 0.019 | 0.164 | 1.096 | 1.019 | 1.178 |
| | | Inverse variance weighted | | 0.127 | 0.023 | **2.84E-08** | 0.082 | 0.172 | 1.135 | 1.086 | 1.187 |
| | | Simple mode | | 0.211 | 0.084 | 0.013 | 0.045 | 0.376 | 1.235 | 1.046 | 1.457 |
| | | Weighted mode | | 0.112 | 0.038 | 0.003 | 0.038 | 0.186 | 1.119 | 1.039 | 1.204 |
| | | MR Egger | 880 | 0.213 | 0.062 | **5.96E-04** | 0.092 | 0.334 | 1.237 | 1.096 | 1.396 |

| | | | | | | | | | | | |
|---|---|---|---|---|---|---|---|---|---|---|---|
| White blood cell (leukocyte) count | | Weighted median | | 0.160 | 0.040 | **6.37E-05** | 0.082 | 0.238 | 1.173 | 1.085 | 1.269 |
| | | Inverse variance weighted | | 0.108 | 0.027 | **8.54E-05** | 0.054 | 0.162 | 1.114 | 1.056 | 1.176 |
| | | Simple mode | | 0.191 | 0.153 | 0.212 | -0.109 | 0.491 | 1.210 | 0.897 | 1.634 |
| | | Weighted mode | | 0.174 | 0.094 | 0.063 | -0.009 | 0.358 | 1.190 | 0.991 | 1.430 |
| Lymphocyte count | | MR Egger | 925 | 0.248 | 0.060 | **3.87E-05** | 0.130 | 0.366 | 1.282 | 1.139 | 1.441 |
| | | Weighted median | | 0.151 | 0.039 | **1.10E-04** | 0.074 | 0.227 | 1.163 | 1.077 | 1.255 |
| | | Inverse variance weighted | | 0.138 | 0.027 | **2.16E-07** | 0.086 | 0.190 | 1.148 | 1.090 | 1.209 |
| | | Simple mode | | -0.135 | 0.161 | 0.404 | -0.450 | 0.181 | 0.874 | 0.638 | 1.198 |
| | | Weighted mode | | 0.120 | 0.100 | 0.231 | -0.076 | 0.317 | 1.128 | 0.926 | 1.373 |
| Monocyte count | | MR Egger | 1150 | 0.038 | 0.044 | 0.385 | -0.048 | 0.125 | 1.039 | 0.953 | 1.133 |
| | | Weighted median | | 0.042 | 0.034 | 0.221 | -0.025 | 0.109 | 1.043 | 0.975 | 1.115 |
| | | Inverse variance weighted | | 0.083 | 0.022 | **2.03E-04** | 0.039 | 0.127 | 1.087 | 1.040 | 1.135 |
| | | Simple mode | | 0.069 | 0.111 | 0.533 | -0.148 | 0.286 | 1.072 | 0.862 | 1.331 |
| | | Weighted mode | | -0.011 | 0.052 | 0.826 | -0.113 | 0.090 | 0.989 | 0.893 | 1.094 |
| Eosinophill count | | MR Egger | 911 | 0.184 | 0.086 | 0.032 | 0.016 | 0.352 | 1.202 | 1.016 | 1.422 |
| | | Weighted median | | 0.131 | 0.054 | 0.016 | 0.025 | 0.238 | 1.140 | 1.025 | 1.268 |
| | | Inverse variance weighted | | 0.152 | 0.037 | **4.28E-05** | 0.079 | 0.225 | 1.164 | 1.082 | 1.252 |
| | | Simple mode | | 0.115 | 0.185 | 0.536 | -0.248 | 0.477 | 1.121 | 0.780 | 1.612 |
| | | Weighted mode | | 0.091 | 0.112 | 0.413 | -0.127 | 0.310 | 1.096 | 0.880 | 1.363 |
| Mean reticulocyte volume | | MR Egger | 1047 | -0.074 | 0.042 | 0.082 | -0.156 | 0.009 | 0.929 | 0.855 | 1.009 |
| | | Weighted median | | -0.126 | 0.034 | **1.98E-04** | -0.192 | -0.060 | 0.882 | 0.825 | 0.942 |
| | | Inverse variance weighted | | -0.076 | 0.021 | **3.86E-04** | -0.118 | -0.034 | 0.927 | 0.888 | 0.966 |
| | | Simple mode | | -0.149 | 0.097 | 0.125 | -0.340 | 0.041 | 0.861 | 0.712 | 1.042 |
| | | Weighted mode | | -0.131 | 0.050 | 0.009 | -0.230 | -0.032 | 0.877 | 0.795 | 0.968 |
| C-reactive protein (quantile) | | MR Egger | 621 | -0.113 | 0.044 | 0.010 | -0.199 | -0.027 | 0.893 | 0.819 | 0.973 |
| | | Weighted median | | -0.142 | 0.043 | 0.001 | -0.226 | -0.058 | 0.868 | 0.798 | 0.943 |
| | | Inverse variance weighted | | -0.115 | 0.027 | **1.64E-05** | -0.167 | -0.063 | 0.891 | 0.846 | 0.939 |
| | | Simple mode | | -0.085 | 0.105 | 0.418 | -0.292 | 0.121 | 0.918 | 0.747 | 1.129 |
| | | Weighted mode | | -0.144 | 0.042 | 0.001 | -0.228 | -0.061 | 0.865 | 0.796 | 0.941 |
| Total protein (quantile) | | MR Egger | 630 | 0.172 | 0.076 | 0.024 | 0.023 | 0.321 | 1.187 | 1.023 | 1.378 |
| | | Weighted median | | 0.084 | 0.046 | 0.070 | -0.007 | 0.175 | 1.088 | 0.993 | 1.192 |
| | | Inverse variance weighted | | 0.135 | 0.033 | **4.87E-05** | 0.070 | 0.201 | 1.145 | 1.073 | 1.222 |
| | | Simple mode | | -0.047 | 0.189 | 0.803 | -0.418 | 0.323 | 0.954 | 0.658 | 1.382 |
| | | Weighted mode | | -0.047 | 0.101 | 0.640 | -0.245 | 0.151 | 0.954 | 0.782 | 1.163 |

nsnp, number of SNPs used in the MR analysis; lo_ci, the lower limit of the 95% confidence interval, representing the lower bound of the effect estimate; up_ci, the upper limit of the 95% confidence interval, representing the upper bound of the effect estimate; or, Odds Ratio, indicating the ratio relationship between exposure and outcome; or_lci95, the lower limit of the 95% confidence interval for the odds ratio; or_uci95, the upper limit of the 95% confidence interval for the odds ratio.

**Table S7**. Significant genomic regions identified in the local genetic correlation analysis between anorexia nervosa and genetically correlated traits.

| Catalog | Trait | chr | start | end | num_snp | k | local_rhog | var | se | z | p | Annotation |
|---|---|---|---|---|---|---|---|---|---|---|---|---|
| Psychiatric disorder | Schizophrenia | 3 | 47727212 | 49316972 | 963 | 35 | -0.0006 | 8.32E-09 | 0.000091216 | -6.38 | 1.77E-10 | known |
| | | 7 | 142656310 | 144968289 | 2920 | 50 | -0.0006 | 8.88E-09 | 0.000094244 | -6.21 | 5.27E-10 | new |
| | | 8 | 143044914 | 144236881 | 2643 | 50 | -0.0007 | 1.59E-08 | 0.00012601 | -5.81 | 6.23E-09 | new |
| | | 6 | 31571218 | 32682664 | 3874 | 50 | -0.0009 | 2.43E-08 | 0.00015583 | -5.50 | 3.70E-08 | MHC region |
| | | 3 | 49316972 | 51832015 | 1941 | 47 | -0.0006 | 1.30E-08 | 0.00011388 | -5.01 | 5.42E-07 | known |
| | | 6 | 29737971 | 30798168 | 4629 | 50 | -0.0007 | 2.20E-08 | 0.00014847 | -4.90 | 9.64E-07 | MHC region |
| | | 6 | 32682664 | 33236497 | 3087 | 50 | -0.0007 | 1.81E-08 | 0.00013449 | -4.87 | 1.10E-06 | MHC region |
| | | 1 | 71684405 | 74326907 | 3304 | 50 | -0.0006 | 1.61E-08 | 0.00012707 | -4.71 | 2.52E-06 | new |
| | | 6 | 30798168 | 31571218 | 5764 | 50 | -0.0007 | 2.16E-08 | 0.00014701 | -4.66 | 3.15E-06 | MHC region |
| | | 8 | 121201700 | 124861459 | 5741 | 50 | -0.0003 | 4.86E-09 | 0.000069712 | -4.65 | 3.34E-06 | new |
| | | 1 | 244109499 | 245506746 | 2371 | 50 | -0.0004 | 8.42E-09 | 0.00009177 | -4.42 | 9.96E-06 | new |
| | | 3 | 81427310 | 82643886 | 1657 | 50 | -0.0004 | 7.80E-09 | 0.000088291 | -4.33 | 1.46E-05 | new |
| Metabolic traits | Body mass index (BMI) | 3 | 94255316 | 95311971 | 2071 | 43 | -0.0004 | 6.76E-09 | 0.000082213 | -4.64 | 3.51E-06 | known |
| | | 11 | 12564229 | 13373124 | 1726 | 50 | -0.0004 | 9.06E-09 | 0.000095166 | -4.62 | 3.83E-06 | new |
| | Whole body fat mass | 3 | 94255316 | 95311971 | 2071 | 43 | -0.0004 | 7.25E-09 | 0.00008516 | -4.92 | 8.46E-07 | known |
| | | 3 | 49316972 | 51832015 | 2112 | 49 | -0.0005 | 1.41E-08 | 0.00011895 | -4.22 | 2.47E-05 | known |
| | Basal metabolic rate | 3 | 94255316 | 95311971 | 2071 | 43 | -0.0003 | 5.45E-09 | 0.000073838 | -4.41 | 1.02E-05 | known |
| | HDL cholesterol (quantile) | 6 | 29737971 | 30798168 | 5726 | 50 | 0.0005 | 1.33E-08 | 0.00011547 | 4.43 | 9.63E-06 | MHC region |
| | | 12 | 57548860 | 59308666 | 2335 | 50 | 0.0006 | 1.58E-08 | 0.00012557 | 4.39 | 1.15E-05 | new |
| Sex hormone relared traits | Sex hormone-binding globulin levels | 11 | 15742552 | 17578402 | 2723 | 50 | -0.0005 | 1.03E-08 | 0.00010141 | -4.50 | 6.66E-06 | new |
| | | 17 | 78837588 | 80034408 | 2074 | 50 | -0.0005 | 1.15E-08 | 0.0001071 | -4.44 | 8.85E-06 | new |
| | Sex hormone-binding globulin levels (Female) | 11 | 114830666 | 116383348 | 2481 | 50 | -0.0005 | 1.01E-08 | 0.00010042 | -4.78 | 1.79E-06 | known |
| | | 11 | 15742552 | 17578402 | 2723 | 50 | -0.0005 | 1.13E-08 | 0.00010646 | -4.36 | 1.32E-05 | new |
| | | 17 | 78837588 | 80034408 | 2074 | 50 | -0.0005 | 1.51E-08 | 0.0001229 | -4.28 | 1.84E-05 | new |
| blood-based biomarkers | White blood cell (leukocyte) count | 6 | 31571218 | 32682664 | 6062 | 50 | 0.0012 | 8.07E-08 | 0.00028403 | 4.30 | 1.72E-05 | MHC region |

| | | | | | | | | | | | | |
|---|---|---|---|---|---|---|---|---|---|---|---|---|
| | Reticulocyte percentage | 6 | 30798168 | 31571218 | 7457 | 50 | 0.0008 | 2.52E-08 | 0.00015868 | 4.73 | 2.25E-06 | MHC region |
| | Reticulocyte count | 6 | 30798168 | 31571218 | 7457 | 50 | 0.0008 | 2.87E-08 | 0.00016953 | 4.99 | 5.95E-07 | MHC region |
| | High light scatter reticulocyte percentage | 6 | 30798168 | 31571218 | 7457 | 50 | 0.0007 | 2.22E-08 | 0.00014897 | 4.45 | 8.72E-06 | MHC region |
| | High light scatter reticulocyte count | 6 | 30798168 | 31571218 | 7457 | 50 | 0.0007 | 2.49E-08 | 0.00015779 | 4.75 | 2.04E-06 | MHC region |
| | Urate (quantile) | 6 | 29737971 | 30798168 | 5726 | 50 | -0.0006 | 1.79E-08 | 0.00013377 | -4.44 | 8.80E-06 | MHC region |

start, start positions of the locus; end, end positions of the locus; num_snp, number of SNPs in the locus; k, number of eigenvectors used in the truncated-SVD;local_rhog, local genetic covariance estimates; var, variance estimates;
se, standard error; z, z value; p, pvalue.

**Table S8**. Multi-Trait Meta-Analysis of anorexia nervosa and four psychiatric traits.

| | | | | | mtag | | AN | | SCZ | | MDD | | Nervous | | Worry | | | | |
|---|---|---|---|---|---|---|---|---|---|---|---|---|---|---|---|---|---|---|---|
| **SNP** | **CHR** | **BP** | **A1** | **A2** | **beta** | **pval** | **beta** | **pval** | **beta** | **pval** | **beta** | **pval** | **beta** | **pval** | **beta** | **pval** | **Genes** | **Cytogenetic band** | **Annotation** |
| rs10749815 | 1 | 73640129 | A | G | -0.026 | 4.23E-08 | -0.051 | 1.64E-04 | 0.054 | 3.99E-10 | 0.0276 | 1.75E-10 | -0.0015 | 0.129 | -0.0020 | 0.087 | NEGR1 | 1p31.1 | new |
| rs2287348 | 2 | 54039813 | C | T | 0.037 | 8.28E-09 | 0.104 | 5.62E-09 | -0.029 | 0.014 | 0.0015 | 0.795 | 0.0007 | 0.637 | 0.0024 | 0.137 | ASB3 | 2p16.2 | known |
| rs9812977 | 3 | 48720303 | G | A | 0.060 | 1.67E-14 | 0.165 | 8.57E-15 | -0.062 | 9.90E-06 | 0.0050 | 0.475 | 0.0031 | 0.063 | 0.0003 | 0.892 | NCKIPSD | 3p21.31 | known |
| rs11713763 | 3 | 53160360 | G | A | -0.027 | 3.64E-08 | -0.042 | 0.003 | 0.069 | 1.63E-14 | -0.0113 | 0.012 | -0.0045 | 1.64E-05 | -0.0063 | 2.35E-07 | RFT1 | 3p21.1 | new |
| rs1814149 | 5 | 87393746 | A | G | 0.033 | 3.67E-08 | 0.073 | 3.24E-05 | -0.040 | 0.00023 | -0.0144 | 0.009 | 0.0060 | 5.99E-06 | 0.0048 | 0.002 | MEF2C | 5q14.3 | new |
| rs17598658 | 6 | 26175866 | A | C | -0.050 | 1.01E-08 | -0.066 | 0.009 | 0.191 | 3.08E-29 | 0.0466 | 2.87E-10 | -0.0073 | 8.85E-06 | -0.0048 | 0.012 | MHC region | 6p22.2 | |
| rs73396802 | 6 | 32915797 | C | T | -0.058 | 7.30E-11 | -0.101 | 8.34E-05 | 0.164 | 3.24E-21 | -0.0328 | 2.42E-05 | -0.0066 | 1.75E-04 | -0.0080 | 7.50E-05 | MHC region | 6p21.32 | |
| rs545787 | 6 | 33703230 | G | A | -0.028 | 1.16E-08 | -0.059 | 3.56E-05 | 0.053 | 9.03E-09 | -0.0051 | 0.252 | -0.0061 | 6.93E-09 | -0.0034 | 0.005 | MHC region | 6p21.31 | |
| rs4129585 | 8 | 143312933 | A | C | -0.034 | 6.67E-13 | -0.059 | 9.42E-06 | 0.075 | 5.11E-18 | 0.0151 | 5.29E-04 | -0.0062 | 9.44E-10 | -0.0061 | 1.76E-07 | TSNARE1 | 8q24.3 | new |
| rs10824069 | 10 | 75853796 | C | T | -0.030 | 2.40E-08 | -0.078 | 3.80E-07 | -0.004 | 0.710 | -0.0148 | 0.002372 | -0.0033 | 0.004 | -0.0045 | 6.42E-04 | AP3M1 | 10q22.2 | new |
| rs2081361 | 11 | 57411742 | C | T | 0.030 | 1.22E-08 | 0.058 | 1.50E-04 | -0.049 | 3.37E-07 | 0.0202 | 2.68E-05 | 0.0010 | 0.397 | 0.0072 | 3.25E-08 | CLP1 | 11q12.1 | new |
| rs750350 | 11 | 112885342 | G | T | 0.040 | 5.53E-09 | 0.111 | 9.42E-08 | -0.048 | 1.48E-04 | 0.0145 | 0.023 | 0.0031 | 0.034 | 0.0015 | 0.375 | NCAM1 | 11q23.2 | new |
| rs10502199 | 11 | 115120615 | C | T | 0.043 | 1.93E-10 | 0.121 | 2.70E-10 | -0.002 | 0.889 | 0.0003 | 0.958 | 0.0046 | 0.001 | 0.0054 | 0.001 | CADM1 | 11q23.3 | known |
| rs17514846 | 15 | 91416550 | C | A | -0.026 | 3.09E-08 | -0.051 | 1.62E-04 | 0.074 | 5.00E-17 | -0.0174 | 6.36E-05 | -0.0008 | 0.404 | -0.0025 | 0.034 | FURIN | 15q26.1 | new |
| rs12944041 | 17 | 8073097 | G | A | 0.027 | 7.90E-09 | 0.073 | 2.72E-07 | -0.004 | 0.647 | 0.0128 | 0.003 | 0.0032 | 0.001 | 0.0039 | 8.61E-04 | VAMP2 | 17p13.1 | new |
| rs1660237 | 18 | 53013947 | C | T | 0.027 | 1.85E-08 | 0.056 | 5.70E-05 | -0.021 | 0.016 | 0.0189 | 1.56E-05 | 0.0036 | 4.43E-04 | 0.0059 | 6.74E-07 | TCF4 | 18q21.2 | new |

MTAG section of the table presents the MTAG results specific to Trait 1. anorexia nervosa. AN, anorexia nervosa; SCZ, schizophrenia; MDD, major depressive disorder; Nervous, nervous feelings; Worry, worrier / anxious feelings.

**Table S9**. Single-Tissue eQTLs for novel loci.

| Trait | SNP | Gene Symbol | CHR | BP | Cytogenetic band | A1 | A2 | P-Value | NES | Tissue |
|---|---|---|---|---|---|---|---|---|---|---|
| AN & HDL cholesterol (quantile) | rs10105606 | LPL | chr8 | 19970337 | 8p21.3 | C | A | 9.80E-05 | 0.066 | Adipose - Subcutaneous |
| AN & Gamma glutamyltransferase (quantile) | rs10153315 | ACTG1 | chr17 | 81514746 | 17q25.3 | T | C | 1.10E-05 | 0.16 | Esophagus - Gastroesophageal Junction |
| | | FSCN2 | | | | | | 5.90E-07 | 0.45 | Brain - Nucleus accumbens (basal ganglia) |
| | | | | | | | | 4.30E-05 | 0.39 | Brain - Caudate (basal ganglia) |
| | | RP13-766D20.2 | | | | | | 2.60E-07 | 0.44 | Brain - Nucleus accumbens (basal ganglia) |
| | | TSPAN10 | | | | | | 7.80E-05 | -0.17 | Esophagus - Mucosa |
| AN & Alanine aminotransferase (quantile) | rs10733608 | AKNA | chr9 | 114386150 | 9q32 | G | T | 1.60E-26 | -0.35 | Esophagus - Muscularis |
| | | | | | | | | 7.00E-18 | -0.32 | Colon - Sigmoid |
| | | | | | | | | 2.00E-12 | -0.33 | Esophagus - Gastroesophageal Junction |
| | | | | | | | | 1.90E-06 | -0.24 | Brain - Cerebellum |
| | | | | | | | | 2.20E-06 | -0.25 | Brain - Hippocampus |
| | | WHRN | | | | | | 9.70E-07 | 0.28 | Brain - Putamen (basal ganglia) |
| | | | | | | | | 2.60E-05 | -0.38 | Brain - Cerebellum |
| | | | | | | | | 9.20E-05 | 0.15 | Adipose - Subcutaneous |
| AN & Whole body fat mass | rs10788797 | BNIPL | chr1 | 151030595 | 1q21.3 | G | A | 4.90E-42 | -0.74 | Thyroid |
| | | | | | | | | 4.40E-06 | -0.35 | Colon - Sigmoid |
| | | | | | | | | 4.50E-06 | -0.36 | Small Intestine - Terminal Ileum |
| | | CDC42SE1 | | | | | | 2.20E-06 | -0.23 | Colon - Transverse |
| | | | | | | | | 0.00013 | -0.13 | Thyroid |
| | | CERS2 | | | | | | 0.00011 | 0.11 | Adipose - Subcutaneous |
| | | HORMAD1 | | | | | | 4.00E-08 | 0.51 | Esophagus - Mucosa |
| | | | | | | | | 6.60E-05 | 0.34 | Esophagus - Muscularis |
| | | | | | | | | 8.90E-05 | 0.28 | Colon - Transverse |
| | | | | | | | | 0.00014 | 0.31 | Adipose - Subcutaneous |
| | | LYSMD1 | | | | | | 2.00E-06 | -0.26 | Esophagus - Muscularis |
| | | | | | | | | 2.90E-06 | -0.2 | Thyroid |

| | | | | | | | | | | |
|---|---|---|---|---|---|---|---|---|---|---|
| | | | | | | | | 8.80E-05 | -0.17 | Adipose - Subcutaneous |
| | | MLLT11 | | | | | | 0.00023 | -0.15 | Esophagus - Mucosa |
| | | MRPS21 | | | | | | 6.80E-05 | -0.16 | Esophagus - Mucosa |
| | | PRUNE1 | | | | | | 2.80E-12 | -0.25 | Esophagus - Muscularis |
| | | | | | | | | 1.10E-06 | -0.14 | Thyroid |
| | | | | | | | | 2.00E-06 | -0.25 | Esophagus - Mucosa |
| | | RP11-54A4.2 | | | | | | 0.00041 | -0.2 | Thyroid |
| | | RP11-68I18.10 | | | | | | 0.00025 | -0.25 | Thyroid |
| | | SELENBP1 | | | | | | 0.00029 | -0.17 | Thyroid |
| AN & four psychiatric traits<br>AN & Bipolar Disorder<br>AN & Nervous feelings<br>AN & Tense / 'highly strung'<br>AN & Worrier / anxious feelings | rs10824069 | ADK | chr10 | 74094038 | 10q22.2 | C | T | 6.20E-10 | -0.41 | Colon - Sigmoid |
| | | | | | | | | 2.50E-09 | -0.35 | Colon - Transverse |
| | | | | | | | | 2.80E-09 | 0.13 | Esophagus - Mucosa |
| | | | | | | | | 2.70E-05 | -0.16 | Esophagus - Muscularis |
| | | | | | | | | 2.80E-05 | 0.13 | Thyroid |
| | | AP3M1 | | | | | | 3.10E-07 | -0.22 | Colon - Sigmoid |
| | | AP3M1 | | | | | | 0.00012 | -0.15 | Colon - Transverse |
| | | SEC24C | | | | | | 0.00017 | 0.092 | Esophagus - Mucosa |
| | | VCL | | | | | | 7.20E-05 | -0.093 | Esophagus - Muscularis |
| AN & Bipolar Disorder<br>AN & Nervous feelings | rs11001053 | ADK | chr10 | 74487347 | 10q22.2 | C | T | 1.80E-10 | -0.38 | Colon - Transverse |
| | | | | | | | | 4.10E-09 | 0.13 | Esophagus - Mucosa |
| | | | | | | | | 5.00E-09 | -0.4 | Colon - Sigmoid |
| | | | | | | | | 8.20E-06 | -0.17 | Esophagus - Muscularis |
| | | | | | | | | 0.00035 | 0.11 | Thyroid |
| | | AP3M1 | | | | | | 2.10E-06 | -0.2 | Colon - Sigmoid |
| | | AP3M1 | | | | | | 0.00013 | -0.16 | Colon - Transverse |
| | | SEC24C | | | | | | 0.00041 | 0.089 | Esophagus - Mucosa |
| | | VCL | | | | | | 0.00035 | -0.086 | Esophagus - Muscularis |
| AN & Body mass index (BMI)<br>AN & Whole body fat mass | rs11030088 | BDNF-AS | chr11 | 27624700 | 11p14.1 | G | A | 3.90E-06 | -0.16 | Adipose - Visceral (Omentum) |
| | | | | | | | | 1.30E-05 | -0.17 | Adipose - Subcutaneous |
| | | | | | | | | 1.60E-05 | -0.16 | Esophagus - Muscularis |
| | | | | | | | | 3.30E-05 | -0.12 | Thyroid |
| | | LIN7C | | | | | | 2.60E-09 | 0.17 | Thyroid |

| | | | | | | | | | | |
|---|---|---|---|---|---|---|---|---|---|---|
| | | | | | | | | 2.40E-08 | 0.2 | Esophagus - Muscularis |
| | | | | | | | | 2.90E-07 | 0.17 | Adipose - Subcutaneous |
| | | | | | | | | 0.00013 | 0.16 | Esophagus - Gastroesophageal Junction |
| | | | | | | | | 0.00019 | 0.12 | Esophagus - Mucosa |
| AN & Worrier / anxious feelings | rs11169059 | C1QL4 | chr12 | 49531049 | 12q13.12 | A | G | 5.30E-07 | 0.3 | Thyroid |
| | | DNAJC22 | | | | | | 8.10E-06 | -0.17 | Thyroid |
| | | | | | | | | 7.90E-05 | -0.16 | Stomach |
| | | | | | | | | 0.00013 | -0.3 | Esophagus - Muscularis |
| | | KCNH3 | | | | | | 1.60E-07 | -0.22 | Thyroid |
| | | RP11-161H23.8 | | | | | | 1.90E-11 | -0.58 | Brain - Cerebellar Hemisphere |
| | | | | | | | | 5.70E-10 | -0.69 | Brain - Frontal Cortex (BA9) |
| | | | | | | | | 6.30E-07 | -0.62 | Brain - Cortex |
| | | | | | | | | 7.60E-06 | -0.48 | Brain - Hippocampus |
| | | | | | | | | 1.30E-05 | -0.6 | Brain - Anterior cingulate cortex (BA24) |
| | | | | | | | | 2.10E-05 | -0.56 | Brain - Hypothalamus |
| | | | | | | | | 2.20E-05 | -0.49 | Brain - Cerebellum |
| | | SPATS2 | | | | | | 1.70E-11 | -0.3 | Thyroid |
| | | | | | | | | 6.70E-05 | -0.18 | Esophagus - Mucosa |
| AN & Basal metabolic rate | rs11245450 | CTBP2 | chr10 | 124969506 | 10q26.13 | G | A | 6.20E-05 | 0.25 | Brain - Cerebellum |
| | | RP11-59C5.3 | | | | | | 8.20E-06 | 0.16 | Esophagus - Mucosa |
| | | | | | | | | 2.20E-05 | 0.16 | Colon - Transverse |
| AN & Urate (quantile) | rs11609805 | NEMP1 | chr12 | 57341262 | 12q13.3 | G | A | 4.70E-05 | 0.13 | Thyroid |
| | | TSFM | | | | | | 0.00015 | -0.13 | Esophagus - Muscularis |
| AN & Glucose (quantile) | rs11678685 | G6PC2 | chr2 | 168854097 | 2q31.1 | G | A | 4.20E-81 | 0.96 | Adipose - Subcutaneous |
| | | | | | | | | 1.90E-79 | 0.98 | Adipose - Visceral (Omentum) |
| | | | | | | | | 5.70E-55 | 0.79 | Thyroid |
| | | | | | | | | 9.60E-33 | 0.72 | Stomach |
| | | | | | | | | 1.70E-20 | 0.58 | Colon - Transverse |
| | | | | | | | | 8.10E-14 | 0.72 | Small Intestine - Terminal Ileum |
| | | | | | | | | 7.80E-12 | 0.44 | Colon - Sigmoid |
| | | NOSTRIN | | | | | | 1.30E-10 | -0.22 | Stomach |
| | | | | | | | | 5.90E-06 | -0.13 | Colon - Transverse |
| AN & Body mass index (BMI)<br>AN & Whole body fat mass | rs11689201 | AFF3 | chr2 | 100203969 | 2q11.2 | A | G | 3.60E-12 | 0.34 | Brain - Cerebellar Hemisphere |

| | | | | | | | | | | |
|---|---|---|---|---|---|---|---|---|---|---|
| | | | | | | | | 3.60E-09 | 0.32 | Brain - Frontal Cortex (BA9) |
| | | | | | | | | 2.10E-08 | 0.27 | Brain - Anterior cingulate cortex (BA24) |
| | | | | | | | | 2.20E-08 | 0.28 | Brain - Cerebellum |
| | | | | | | | | 7.00E-07 | 0.31 | Brain - Nucleus accumbens (basal ganglia) |
| | | | | | | | | 2.60E-06 | 0.15 | Adipose - Subcutaneous |
| | | | | | | | | 7.80E-06 | 0.25 | Brain - Cortex |
| | | | | | | | | 2.70E-05 | 0.18 | Adipose - Visceral (Omentum) |
| | | CHST10 | | | | | | 9.80E-07 | 0.17 | Thyroid |
| | | LONRF2 | | | | | | 3.90E-09 | 0.21 | Thyroid |
| AN & four psychiatric traits | rs11713763 | ALAS1 | chr3 | 53126344 | 3p21.1 | G | A | 0.0001 | 0.1 | Adipose - Subcutaneous |
| | | GLYCTK | | | | | | 4.70E-06 | -0.17 | Esophagus - Mucosa |
| | | | | | | | | 7.30E-06 | -0.15 | Thyroid |
| | | | | | | | | 1.80E-05 | -0.3 | Brain - Cerebellum |
| | | | | | | | | 0.00019 | -0.1 | Colon - Transverse |
| | | GNL3 | | | | | | 1.90E-08 | -0.37 | Brain - Cerebellum |
| | | | | | | | | 9.30E-06 | -0.13 | Esophagus - Muscularis |
| | | | | | | | | 2.70E-05 | -0.16 | Colon - Sigmoid |
| | | | | | | | | 4.00E-05 | -0.15 | Esophagus - Gastroesophageal Junction |
| | | | | | | | | 4.10E-05 | -0.3 | Brain - Cerebellar Hemisphere |
| | | | | | | | | 5.30E-05 | -0.084 | Colon - Transverse |
| | | ITIH4 | | | | | | 1.00E-09 | 0.23 | Thyroid |
| | | | | | | | | 1.00E-08 | 0.25 | Esophagus - Mucosa |
| | | | | | | | | 6.50E-08 | 0.16 | Adipose - Subcutaneous |
| | | | | | | | | 7.20E-08 | 0.22 | Adipose - Visceral (Omentum) |
| | | NEK4 | | | | | | 6.50E-08 | -0.16 | Thyroid |
| | | | | | | | | 5.90E-05 | -0.14 | Esophagus - Mucosa |
| | | | | | | | | 0.00015 | -0.15 | Adipose - Visceral (Omentum) |
| | | NT5DC2 | | | | | | 3.70E-09 | -0.24 | Colon - Transverse |
| | | | | | | | | 6.90E-06 | -0.24 | Colon - Sigmoid |
| | | PBRM1 | | | | | | 9.50E-13 | 0.22 | Thyroid |
| | | RFT1 | | | | | | 1.20E-05 | 0.21 | Stomach |
| | | RP5-966M1.7 | | | | | | 3.00E-05 | -0.34 | Brain - Cerebellum |
| | | SERBP1P3 | | | | | | 5.20E-05 | 0.21 | Esophagus - Gastroesophageal Junction |

| | | | | | | | | | | |
|---|---|---|---|---|---|---|---|---|---|---|
| | | | | | | | | 7.20E-05 | 0.21 | Thyroid |
| | | | | | | | | 0.0001 | 0.34 | Brain - Cortex |
| | | SFMBT1 | | | | | | 3.10E-06 | 0.25 | Colon - Sigmoid |
| | | | | | | | | 5.70E-06 | 0.32 | Brain - Cerebellum |
| | | TMEM110 | | | | | | 0.00021 | 0.11 | Adipose - Subcutaneous |
| AN & Urate (quantile) | rs12189774 | RP11-344J7.4 | chr6 | 43848389 | 6p21.1 | G | A | 1.20E-14 | -0.43 | Thyroid |
| | | VEGFA | | | | | | 5.00E-06 | 0.15 | Thyroid |
| AN & Body mass index (BMI) | rs12411523 | LOXL4 | chr10 | 98102974 | 10q24.2 | C | T | 1.80E-11 | 0.31 | Esophagus - Muscularis |
| | | | | | | | | 1.30E-09 | 0.22 | Colon - Sigmoid |
| | | | | | | | | 1.00E-07 | -0.2 | Thyroid |
| | | | | | | | | 8.70E-07 | 0.28 | Esophagus - Gastroesophageal Junction |
| | | | | | | | | 1.60E-05 | 0.14 | Esophagus - Mucosa |
| | | | | | | | | 0.00015 | 0.15 | Thyroid |
| | | | | | | | | 2.60E-10 | 0.23 | Esophagus - Mucosa |
| | | | | | | | | 7.90E-06 | 0.14 | Adipose - Subcutaneous |
| | | | | | | | | 3.20E-05 | 0.25 | Brain - Cerebellum |
| | | | | | | | | 0.00013 | 0.22 | Esophagus - Muscularis |
| AN & Body mass index (BMI)<br>AN & Whole body fat mass | rs12583517 | PCDH17 | chr13 | 58089323 | 13q21.1 | G | T | 2.80E-05 | -0.19 | Esophagus - Muscularis |
| AN & Glycated hemoglobin levels（HbA1c） | rs12938531 | KRBA2 | chr17 | 8222223 | 17p13.1 | A | G | 6.70E-07 | 0.2 | Esophagus - Muscularis |
| | | | | | | | | 2.20E-05 | 0.17 | Colon - Transverse |
| | | LINC00324 | | | | | | 1.00E-05 | 0.18 | Esophagus - Mucosa |
| | | PFAS | | | | | | 1.40E-05 | -0.1 | Adipose - Subcutaneous |
| | | | | | | | | 4.30E-05 | -0.11 | Esophagus - Muscularis |
| | | VAMP2 | | | | | | 2.40E-07 | 0.23 | Brain - Caudate (basal ganglia) |
| | | | | | | | | 4.90E-06 | 0.14 | Brain - Frontal Cortex (BA9) |
| | | | | | | | | 5.40E-06 | 0.19 | Brain - Nucleus accumbens (basal ganglia) |
| | | | | | | | | 5.50E-06 | 0.23 | Brain - Hypothalamus |
| | | | | | | | | 3.50E-05 | 0.24 | Brain - Cerebellum |
| AN & four psychiatric traits<br>AN & Major Depressive Disorder | rs12944041 | CTC1 | chr17 | 8169779 | 17p13.1 | G | A | 2.20E-06 | 0.18 | Colon - Transverse |
| | | | | | | | | 1.20E-05 | 0.12 | Adipose - Subcutaneous |
| | | KRBA2 | | | | | | 5.30E-06 | 0.18 | Esophagus - Muscularis |
| | | LINC00324 | | | | | | 0.00011 | 0.15 | Esophagus - Mucosa |

| | | | | | | | | | | |
|---|---|---|---|---|---|---|---|---|---|---|
| | | VAMP2 | | | | | | 9.40E-10 | 0.32 | Brain - Cerebellum |
| | | | | | | | | 1.20E-07 | 0.23 | Brain - Caudate (basal ganglia) |
| | | | | | | | | 1.50E-07 | 0.31 | Brain - Putamen (basal ganglia) |
| | | | | | | | | 1.10E-06 | 0.24 | Brain - Hypothalamus |
| | | | | | | | | 4.40E-06 | 0.14 | Brain - Frontal Cortex (BA9) |
| | | | | | | | | 1.10E-05 | 0.16 | Brain - Hippocampus |
| | | | | | | | | 2.20E-05 | 0.2 | Colon - Sigmoid |
| | | | | | | | | 4.90E-05 | 0.19 | Brain - Cerebellar Hemisphere |
| | | | | | | | | 4.90E-05 | 0.17 | Esophagus - Gastroesophageal Junction |
| AN & SHBG (quantile) | rs13133311 | HSD17B11 | chr4 | 87296071 | 4q22.1 | G | A | 3.20E-11 | 0.17 | Thyroid |
| | | | | | | | | 5.10E-08 | 0.13 | Esophagus - Muscularis |
| | | | | | | | | 2.00E-07 | 0.15 | Adipose - Visceral (Omentum) |
| | | | | | | | | 2.70E-06 | 0.13 | Adipose - Subcutaneous |
| | | | | | | | | 6.00E-06 | 0.12 | Esophagus - Gastroesophageal Junction |
| | | | | | | | | 6.40E-05 | 0.11 | Esophagus - Mucosa |
| | | | | | | | | 1.00E-17 | 0.4 | Adipose - Visceral (Omentum) |
| | | | | | | | | 1.10E-13 | 0.65 | Brain - Cortex |
| | | | | | | | | 9.90E-09 | 0.23 | Adipose - Subcutaneous |
| | | | | | | | | 3.80E-07 | 0.52 | Brain - Hypothalamus |
| | | | | | | | | 2.30E-06 | 0.4 | Brain - Cerebellum |
| | | | | | | | | 4.30E-06 | 0.42 | Brain - Cerebellar Hemisphere |
| | | | | | | | | 6.60E-06 | 0.23 | Esophagus - Muscularis |
| | | | | | | | | 3.40E-05 | 0.36 | Brain - Nucleus accumbens (basal ganglia) |
| | | | | | | | | 7.50E-05 | 0.4 | Brain - Frontal Cortex (BA9) |
| AN & Worrier / anxious feelings | rs13262595 | TSNARE1 | chr8 | 142235609 | 8q24.3 | A | G | 5.70E-06 | 0.16 | Adipose - Subcutaneous |
| AN & Sex hormone-binding globulin levels | rs1640267 | SGTA | chr19 | 2789339 | 19p13.3 | T | C | 5.30E-09 | 0.11 | Thyroid |
| | | | | | | | | 5.70E-06 | -0.11 | Adipose - Subcutaneous |
| | | | | | | | | 1.60E-05 | -0.09 | Esophagus - Muscularis |
| | | THOP1 | | | | | | 7.90E-74 | 0.55 | Thyroid |
| | | | | | | | | 3.40E-13 | 0.36 | Brain - Cerebellum |
| | | | | | | | | 1.40E-08 | 0.14 | Esophagus - Muscularis |
| | | | | | | | | 4.50E-08 | 0.17 | Esophagus - Mucosa |
| | | | | | | | | 4.00E-07 | 0.19 | Brain - Cerebellar Hemisphere |

| | | | | | | | | | | |
|---|---|---|---|---|---|---|---|---|---|---|
| | | | | | | | | 1.10E-06 | 0.42 | Brain - Spinal cord (cervical c-1) |
| | | ZNF554 | | | | | | 1.10E-38 | 0.5 | Esophagus - Mucosa |
| | | | | | | | | 9.10E-29 | 0.4 | Adipose - Subcutaneous |
| | | | | | | | | 3.60E-22 | 0.32 | Thyroid |
| | | | | | | | | 5.70E-14 | 0.31 | Esophagus - Muscularis |
| | | | | | | | | 2.90E-13 | 0.26 | Adipose - Visceral (Omentum) |
| | | | | | | | | 3.30E-12 | 0.42 | Brain - Cerebellum |
| | | | | | | | | 2.70E-11 | 0.34 | Stomach |
| | | | | | | | | 1.10E-06 | 0.3 | Brain - Cerebellar Hemisphere |
| | | | chr19 | | | | | 2.40E-06 | 0.23 | Esophagus - Gastroesophageal Junction |
| AN & High light scatter reticulocyte count<br>AN & High light scatter reticulocyte percentage<br>AN & Immature reticulocyte fraction<br>AN & Reticulocyte count<br>AN & Reticulocyte percentage<br>AN & Sex hormone-binding globulin levels | rs174723 | RFPL1S | chr22 | 29451337 | 22q12.2 | A | G | 1.70E-07 | 0.2 | Brain - Frontal Cortex (BA9) |
| | | RP3-394A18.1 | | | | | | 7.90E-06 | -0.1 | Thyroid |
| | | | | | | | | 4.00E-05 | -0.14 | Esophagus - Gastroesophageal Junction |
| | | THOC5 | | | | | | 1.00E-14 | -0.33 | Thyroid |
| | | | | | | | | 2.40E-12 | -0.34 | Adipose - Subcutaneous |
| | | | | | | | | 3.50E-10 | -0.38 | Esophagus - Muscularis |
| | | | | | | | | 8.40E-08 | -0.22 | Esophagus - Mucosa |
| | | | | | | | | 1.50E-07 | -0.29 | Adipose - Visceral (Omentum) |
| | | | | | | | | 1.20E-06 | -0.36 | Esophagus - Gastroesophageal Junction |
| | | | | | | | | 6.20E-05 | -0.27 | Colon - Sigmoid |
| AN & four psychiatric traits | rs17514846 | FES | chr15 | 90873320 | 15q26.1 | C | A | 4.20E-23 | -0.34 | Thyroid |
| | | | | | | | | 1.30E-08 | -0.2 | Adipose - Visceral (Omentum) |
| | | | | | | | | 6.50E-07 | -0.17 | Stomach |
| | | | | | | | | 2.30E-06 | -0.13 | Adipose - Subcutaneous |
| | | | | | | | | 3.00E-06 | -0.14 | Colon - Transverse |
| | | | | | | | | 0.0001 | -0.11 | Esophagus - Muscularis |
| | | FURIN | | | | | | 4.10E-14 | 0.21 | Esophagus - Mucosa |
| AN & four psychiatric traits | rs1814149 | CTC-498M16.4 | chr5 | 88097929 | 5q14.3 | A | G | 1.10E-10 | 0.64 | Brain - Caudate (basal ganglia) |
| | | | | | | | | 2.90E-07 | 0.49 | Brain - Cortex |
| | | | | | | | | 2.30E-06 | 0.42 | Brain - Nucleus accumbens (basal ganglia) |
| | | | | | | | | 4.10E-06 | 0.42 | Brain - Anterior cingulate cortex (BA24) |
| | | | | | | | | 3.30E-05 | 0.43 | Brain - Frontal Cortex (BA9) |
| HDL cholesterol (quantile) | rs2072113 | BSCL2 | chr11 | 61837495 | 11q12.2 | C | T | 4.50E-06 | 0.22 | Esophagus - Muscularis |

| | | | | | | | | | | |
|---|---|---|---|---|---|---|---|---|---|---|
| | | FADS1 | | | | | | 2.00E-11 | -0.35 | Esophagus - Mucosa |
| | | | | | | | | 1.70E-09 | -0.66 | Brain - Cerebellum |
| | | | | | | | | 2.00E-08 | -0.58 | Brain - Cerebellar Hemisphere |
| | | | | | | | | 1.10E-07 | -0.35 | Stomach |
| | | | | | | | | 2.90E-07 | -0.23 | Thyroid |
| | | | | | | | | 8.90E-07 | -0.29 | Adipose - Subcutaneous |
| | | | | | | | | 1.00E-06 | -0.31 | Brain - Cortex |
| | | | | | | | | 1.60E-06 | -0.29 | Colon - Sigmoid |
| | | | | | | | | 4.50E-05 | -0.35 | Brain - Putamen (basal ganglia) |
| | | | | | | | | 5.20E-05 | -0.26 | Esophagus - Gastroesophageal Junction |
| AN & four psychiatric traits | rs2081361 | AP000662.4 | chr11 | 57644269 | 11q12.1 | C | T | 1.50E-15 | -0.4 | Thyroid |
| | | MED19 | | | | | | 2.10E-11 | -0.31 | Esophagus - Mucosa |
| | | | | | | | | 1.60E-10 | -0.23 | Thyroid |
| | | | | | | | | 2.60E-09 | -0.25 | Adipose - Subcutaneous |
| | | | | | | | | 5.60E-07 | -0.24 | Esophagus - Muscularis |
| | | | | | | | | 0.00014 | -0.37 | Brain - Cerebellum |
| | | SELENOH | | | | | | 2.10E-05 | -0.096 | Thyroid |
| | | SLC43A1 | | | | | | 4.50E-05 | -0.4 | Brain - Cerebellum |
| | | TIMM10 | | | | | | 5.50E-15 | -0.39 | Esophagus - Muscularis |
| | | | | | | | | 2.40E-13 | -0.27 | Thyroid |
| | | | | | | | | 1.60E-12 | -0.32 | Esophagus - Mucosa |
| | | | | | | | | 8.00E-11 | -0.23 | Adipose - Visceral (Omentum) |
| | | | | | | | | 2.70E-10 | -0.35 | Stomach |
| | | | | | | | | 1.70E-09 | -0.2 | Adipose - Subcutaneous |
| | | | | | | | | 3.10E-08 | -0.27 | Colon - Transverse |
| | | | | | | | | 5.00E-08 | -0.31 | Esophagus - Gastroesophageal Junction |
| | | | | | | | | 9.20E-05 | -0.26 | Colon - Sigmoid |
| | | | | | | | | 0.00013 | -0.21 | Brain - Caudate (basal ganglia) |
| | | ZDHHC5 | | | | | | 5.80E-05 | 0.09 | Adipose - Subcutaneous |
| | | | | | | | | 0.00018 | 0.081 | Adipose - Visceral (Omentum) |
| AN & C-reactive protein (quantile) | rs2203474 | ACADS | chr12 | 120794597 | 12q24.31 | T | G | 8.50E-11 | 0.37 | Thyroid |
| | | | | | | | | 6.40E-05 | 0.26 | Esophagus - Muscularis |
| | | C12orf43 | | | | | | 9.60E-06 | -0.3 | Adipose - Subcutaneous |

| | | | | | | | | | | |
|---|---|---|---|---|---|---|---|---|---|---|
| | | SPPL3 | | | | | | 0.0001 | -0.23 | Esophagus - Muscularis |
| | | UNC119B | | | | | | 3.50E-06 | 0.23 | Esophagus - Muscularis |
| | | | | | | | | 5.50E-05 | 0.29 | Stomach |
| AN & Body mass index (BMI)<br>AN & Whole body fat mass | rs2466826 | APOBR | chr16 | 28337023 | 16p11.2 | G | A | 7.40E-06 | 0.12 | Thyroid |
| | | ATXN2L | | | | | | 3.00E-05 | -0.096 | Adipose - Subcutaneous |
| | | | | | | | | 5.60E-05 | -0.1 | Adipose - Visceral (Omentum) |
| | | CDC37P1 | | | | | | 3.50E-57 | 0.76 | Thyroid |
| | | | | | | | | 1.80E-37 | 0.66 | Adipose - Subcutaneous |
| | | | | | | | | 4.20E-32 | 0.75 | Colon - Transverse |
| | | | | | | | | 9.80E-21 | 0.61 | Stomach |
| | | | | | | | | 1.80E-14 | 0.69 | Small Intestine - Terminal Ileum |
| | | CDC37P2 | | | | | | 9.10E-08 | -0.22 | Thyroid |
| | | | | | | | | 1.50E-05 | -0.22 | Colon - Transverse |
| | | CLN3 | | | | | | 2.20E-07 | 0.12 | Thyroid |
| | | | | | | | | 1.50E-05 | 0.12 | Esophagus - Mucosa |
| | | | | | | | | 3.90E-05 | 0.11 | Stomach |
| | | EIF3C | | | | | | 6.10E-11 | 0.29 | Adipose - Subcutaneous |
| | | | | | | | | 4.20E-08 | 0.25 | Esophagus - Muscularis |
| | | | | | | | | 1.50E-05 | 0.33 | Brain - Nucleus accumbens (basal ganglia) |
| | | | | | | | | 1.70E-05 | 0.39 | Brain - Hypothalamus |
| | | | | | | | | 4.90E-05 | 0.32 | Brain - Hippocampus |
| | | | | | | | | 0.00014 | 0.13 | Esophagus - Mucosa |
| | | LAT | | | | | | 2.00E-09 | 0.14 | Esophagus - Muscularis |
| | | | | | | | | 1.60E-05 | 0.25 | Brain - Putamen (basal ganglia) |
| | | NFATC2IP | | | | | | 2.70E-05 | 0.14 | Esophagus - Mucosa |
| | | | | | | | | 5.50E-05 | 0.096 | Thyroid |
| | | | | | | | | 5.90E-05 | 0.11 | Colon - Transverse |
| | | NPIPB6 | | | | | | 4.20E-05 | 0.21 | Brain - Cerebellar Hemisphere |
| | | NPIPB7 | | | | | | 1.10E-13 | -0.7 | Brain - Cortex |
| | | | | | | | | 2.90E-13 | -0.57 | Brain - Cerebellum |
| | | | | | | | | 6.10E-12 | -0.55 | Brain - Nucleus accumbens (basal ganglia) |
| | | | | | | | | 1.20E-11 | -0.55 | Brain - Cerebellar Hemisphere |
| | | | | | | | | 3.10E-08 | -0.5 | Brain - Hypothalamus |

| | | | | | | | | | | |
|---|---|---|---|---|---|---|---|---|---|---|
| | | | | | | | | 2.70E-07 | -0.51 | Brain - Frontal Cortex (BA9) |
| | | | | | | | | 3.50E-07 | -0.56 | Brain - Hippocampus |
| | | | | | | | | 5.90E-07 | -0.57 | Brain - Anterior cingulate cortex (BA24) |
| | | | | | | | | 3.00E-06 | -0.49 | Brain - Putamen (basal ganglia) |
| | | NPIPB9 | | | | | | 5.20E-17 | -0.54 | Esophagus - Muscularis |
| | | | | | | | | 4.20E-09 | -0.43 | Colon - Sigmoid |
| | | | | | | | | 5.30E-09 | -0.46 | Esophagus - Gastroesophageal Junction |
| | | | | | | | | 1.40E-07 | -0.56 | Brain - Caudate (basal ganglia) |
| | | | | | | | | 4.00E-07 | -0.31 | Colon - Transverse |
| | | | | | | | | 5.70E-07 | -0.3 | Adipose - Visceral (Omentum) |
| | | | | | | | | 9.00E-07 | -0.24 | Thyroid |
| | | | | | | | | 4.20E-06 | -0.54 | Brain - Cerebellar Hemisphere |
| | | | | | | | | 6.60E-06 | -0.26 | Adipose - Subcutaneous |
| | | | | | | | | 9.20E-06 | -0.29 | Stomach |
| | | | | | | | | 1.00E-05 | -0.44 | Brain - Cerebellum |
| | | | | | | | | 1.10E-05 | -0.47 | Brain - Nucleus accumbens (basal ganglia) |
| | | | | | | | | 4.70E-05 | -0.43 | Brain - Cortex |
| | | NUPR1 | | | | | | 1.20E-13 | -0.25 | Esophagus - Muscularis |
| | | | | | | | | 9.00E-09 | 0.39 | Brain - Cerebellar Hemisphere |
| | | | | | | | | 2.20E-05 | 0.28 | Brain - Cerebellum |
| | | | | | | | | 7.00E-05 | -0.2 | Esophagus - Gastroesophageal Junction |
| | | RP11-1348G14.4 | | | | | | 2.80E-20 | -0.48 | Adipose - Subcutaneous |
| | | | | | | | | 4.70E-16 | -0.43 | Esophagus - Muscularis |
| | | | | | | | | 1.40E-14 | -0.32 | Thyroid |
| | | | | | | | | 4.40E-11 | -0.31 | Adipose - Visceral (Omentum) |
| | | | | | | | | 2.20E-09 | -0.37 | Colon - Transverse |
| | | | | | | | | 3.60E-09 | -0.52 | Brain - Nucleus accumbens (basal ganglia) |
| | | | | | | | | 4.00E-08 | -0.3 | Colon - Sigmoid |
| | | | | | | | | 1.40E-07 | -0.47 | Small Intestine - Terminal Ileum |
| | | | | | | | | 2.50E-07 | -0.56 | Brain - Hypothalamus |
| | | | | | | | | 3.30E-07 | -0.61 | Brain - Amygdala |
| | | | | | | | | 3.00E-06 | -0.3 | Stomach |
| | | | | | | | | 3.20E-06 | -0.36 | Brain - Cerebellum |

| | | | | | | | | | | |
|---|---|---|---|---|---|---|---|---|---|---|
| | | | | | | | | 4.20E-06 | -0.4 | Brain - Cerebellar Hemisphere |
| | | | | | | | | 5.70E-06 | -0.48 | Brain - Frontal Cortex (BA9) |
| | | | | | | | | 6.10E-06 | -0.47 | Brain - Hippocampus |
| | | | | | | | | 2.90E-05 | -0.27 | Esophagus - Gastroesophageal Junction |
| | | | | | | | | 3.60E-05 | -0.47 | Brain - Anterior cingulate cortex (BA24) |
| | | | | | | | | 4.20E-05 | -0.38 | Brain - Cortex |
| | | | | | | | | 0.00016 | -0.2 | Esophagus - Mucosa |
| | | RP11-1348G14.6 | | | | | | 4.00E-21 | 0.5 | Thyroid |
| | | | | | | | | 1.80E-06 | 0.49 | Brain - Nucleus accumbens (basal ganglia) |
| | | | | | | | | 3.40E-05 | 0.47 | Brain - Hippocampus |
| | | RP11-22P6.2 | | | | | | 2.50E-06 | -0.36 | Brain - Cerebellum |
| | | | | | | | | 8.90E-06 | -0.39 | Brain - Cerebellar Hemisphere |
| | | RP11-435I10.5 | | | | | | 8.40E-39 | -0.64 | Thyroid |
| | | RP11-666O2.4 | | | | | | 3.40E-07 | 0.57 | Brain - Cerebellar Hemisphere |
| | | SBK1 | | | | | | 3.60E-07 | -0.25 | Esophagus - Muscularis |
| | | | | | | | | 3.30E-06 | -0.2 | Colon - Transverse |
| | | SGF29 | | | | | | 6.40E-07 | -0.13 | Esophagus - Muscularis |
| | | | | | | | | 1.40E-05 | -0.14 | Colon - Sigmoid |
| | | | | | | | | 1.80E-05 | -0.13 | Colon - Transverse |
| | | | | | | | | 4.10E-05 | -0.13 | Stomach |
| | | SH2B1 | | | | | | 1.10E-12 | -0.15 | Adipose - Subcutaneous |
| | | | | | | | | 5.80E-10 | -0.11 | Esophagus - Muscularis |
| | | | | | | | | 4.20E-08 | -0.11 | Adipose - Visceral (Omentum) |
| | | | | | | | | 7.70E-07 | -0.23 | Brain - Cerebellar Hemisphere |
| | | | | | | | | 4.40E-06 | -0.13 | Esophagus - Gastroesophageal Junction |
| | | | | | | | | 2.00E-05 | -0.072 | Esophagus - Mucosa |
| | | | | | | | | 2.00E-05 | -0.12 | Stomach |
| | | SULT1A1 | | | | | | 4.00E-25 | -0.45 | Thyroid |
| | | | | | | | | 5.70E-18 | -0.39 | Esophagus - Mucosa |
| | | | | | | | | 1.50E-09 | -0.45 | Brain - Hypothalamus |
| | | | | | | | | 6.70E-09 | -0.55 | Brain - Anterior cingulate cortex (BA24) |
| | | | | | | | | 1.70E-08 | -0.26 | Stomach |
| | | | | | | | | 2.90E-08 | -0.31 | Esophagus - Gastroesophageal Junction |

| | | | | | | | | | | |
|---|---|---|---|---|---|---|---|---|---|---|
| | | | | | | | | 1.40E-07 | -0.44 | Brain - Cerebellum |
| | | | | | | | | 1.50E-07 | -0.28 | Colon - Sigmoid |
| | | | | | | | | 9.20E-07 | -0.48 | Brain - Amygdala |
| | | | | | | | | 2.60E-06 | -0.43 | Brain - Caudate (basal ganglia) |
| | | | | | | | | 9.30E-06 | -0.41 | Brain - Cortex |
| | | | | | | | | 1.10E-05 | -0.14 | Colon - Transverse |
| | | | | | | | | 1.30E-05 | -0.23 | Adipose - Visceral (Omentum) |
| | | | | | | | | 1.80E-05 | -0.19 | Esophagus - Muscularis |
| | | | | | | | | 2.20E-05 | -0.35 | Brain - Nucleus accumbens (basal ganglia) |
| | | | | | | | | 7.40E-05 | -0.4 | Brain - Cerebellar Hemisphere |
| | | SULT1A2 | | | | | | 2.30E-44 | 0.58 | Adipose - Subcutaneous |
| | | | | | | | | 7.40E-38 | 0.53 | Adipose - Visceral (Omentum) |
| | | | | | | | | 5.30E-32 | 0.63 | Esophagus - Mucosa |
| | | | | | | | | 3.90E-29 | 0.54 | Esophagus - Muscularis |
| | | | | | | | | 5.60E-23 | 0.53 | Esophagus - Gastroesophageal Junction |
| | | | | | | | | 1.40E-20 | 0.58 | Stomach |
| | | | | | | | | 3.00E-15 | 0.44 | Colon - Sigmoid |
| | | | | | | | | 2.20E-13 | 0.8 | Brain - Anterior cingulate cortex (BA24) |
| | | | | | | | | 4.60E-11 | 0.69 | Brain - Putamen (basal ganglia) |
| | | | | | | | | 7.70E-11 | 0.57 | Brain - Nucleus accumbens (basal ganglia) |
| | | | | | | | | 6.00E-10 | 0.63 | Brain - Cortex |
| | | | | | | | | 3.10E-09 | 0.6 | Brain - Caudate (basal ganglia) |
| | | | | | | | | 1.10E-08 | 0.56 | Brain - Hippocampus |
| | | | | | | | | 1.60E-08 | 0.59 | Brain - Hypothalamus |
| | | | | | | | | 9.10E-08 | 0.69 | Brain - Spinal cord (cervical c-1) |
| | | | | | | | | 2.00E-07 | 0.67 | Brain - Amygdala |
| | | | | | | | | 4.80E-07 | 0.13 | Colon - Transverse |
| | | | | | | | | 2.20E-06 | 0.51 | Brain - Frontal Cortex (BA9) |
| | | TUFM | | | | | | 7.60E-31 | 0.24 | Thyroid |
| | | | | | | | | 7.00E-18 | 0.24 | Esophagus - Muscularis |
| | | | | | | | | 9.40E-13 | 0.17 | Adipose - Subcutaneous |
| | | | | | | | | 2.90E-11 | 0.23 | Colon - Sigmoid |
| | | | | | | | | 9.10E-11 | 0.25 | Brain - Putamen (basal ganglia) |

| | | | | | | | | | | |
|---|---|---|---|---|---|---|---|---|---|---|
| | | | | | | | | 1.00E-10 | 0.27 | Brain - Nucleus accumbens (basal ganglia) |
| | | | | | | | | 5.10E-10 | 0.22 | Esophagus - Gastroesophageal Junction |
| | | | | | | | | 6.70E-09 | 0.12 | Esophagus - Mucosa |
| | | | | | | | | 3.60E-08 | 0.2 | Brain - Caudate (basal ganglia) |
| | | | | | | | | 7.70E-08 | 0.11 | Colon - Transverse |
| | | | | | | | | 3.70E-07 | 0.21 | Brain - Cortex |
| | | | | | | | | 3.80E-07 | 0.25 | Brain - Hippocampus |
| | | | | | | | | 4.70E-07 | 0.08 | Adipose - Visceral (Omentum) |
| | | | | | | | | 3.70E-06 | 0.1 | Stomach |
| | | | | | | | | 4.10E-06 | 0.27 | Brain - Amygdala |
| | | | | | | | | 5.00E-06 | 0.12 | Small Intestine - Terminal Ileum |
| | | | | | | | | 1.10E-05 | 0.22 | Brain - Frontal Cortex (BA9) |
| AN & C-reactive protein (quantile) | rs2701184 | ACADS | chr12 | 120915285 | 12q24.31 | T | G | 2.00E-21 | 0.29 | Thyroid |
| | | CABP1 | | | | | | 8.50E-05 | 0.21 | Brain - Cerebellum |
| | | SPPL3 | | | | | | 1.00E-17 | -0.25 | Adipose - Subcutaneous |
| | | | | | | | | 1.90E-14 | -0.26 | Adipose - Visceral (Omentum) |
| | | | | | | | | 4.10E-14 | -0.23 | Esophagus - Muscularis |
| | | | | | | | | 8.90E-13 | -0.35 | Brain - Cortex |
| | | | | | | | | 3.70E-12 | -0.19 | Colon - Transverse |
| | | | | | | | | 1.20E-10 | -0.28 | Colon - Sigmoid |
| | | | | | | | | 5.40E-10 | -0.34 | Brain - Cerebellum |
| | | | | | | | | 2.10E-08 | -0.22 | Esophagus - Gastroesophageal Junction |
| | | | | | | | | 2.10E-08 | -0.29 | Brain - Caudate (basal ganglia) |
| | | | | | | | | 3.30E-08 | -0.21 | Stomach |
| | | | | | | | | 1.60E-06 | -0.2 | Brain - Nucleus accumbens (basal ganglia) |
| | | | | | | | | 1.80E-06 | -0.27 | Brain - Hypothalamus |
| | | | | | | | | 7.00E-06 | -0.24 | Brain - Frontal Cortex (BA9) |
| | | | | | | | | 1.10E-05 | -0.24 | Brain - Putamen (basal ganglia) |
| | | | | | | | | 1.30E-05 | -0.24 | Brain - Cerebellar Hemisphere |
| | | UNC119B | | | | | | 2.50E-05 | 0.23 | Brain - Anterior cingulate cortex (BA24) |
| AN & Body mass index (BMI) | rs273882 | PTBP2 | chr1 | 96811132 | 1p21.3 | T | C | 5.00E-14 | 0.21 | Adipose - Subcutaneous |
| | | | | | | | | 2.30E-09 | 0.17 | Esophagus - Mucosa |
| | | | | | | | | 1.50E-07 | 0.11 | Thyroid |

| | | | | | | | | | | |
|---|---|---|---|---|---|---|---|---|---|---|
| | | | | | | | | 2.90E-06 | 0.13 | Adipose - Visceral (Omentum) |
| | | | | | | | | 0.0002 | 0.098 | Esophagus - Muscularis |
| AN & Basal metabolic rate | rs34298551 | CEP250 | chr20 | 35442798 | 20q11.22 | G | A | 1.30E-05 | 0.16 | Esophagus - Mucosa |
| | | CPNE1 | | | | | | 2.70E-14 | -0.4 | Adipose - Subcutaneous |
| | | | | | | | | 1.00E-11 | -0.38 | Esophagus - Mucosa |
| | | | | | | | | 3.30E-10 | -0.31 | Thyroid |
| | | | | | | | | 5.20E-09 | -0.36 | Adipose - Visceral (Omentum) |
| | | | | | | | | 1.30E-05 | -0.3 | Colon - Sigmoid |
| | | | | | | | | 1.60E-05 | -0.33 | Brain - Caudate (basal ganglia) |
| | | | | | | | | 2.40E-05 | -0.22 | Esophagus - Muscularis |
| | | | | | | | | 0.00016 | -0.29 | Esophagus - Gastroesophageal Junction |
| | | EDEM2 | | | | | | 0.00012 | -0.11 | Adipose - Subcutaneous |
| | | EPB41L1 | | | | | | 0.00011 | -0.15 | Esophagus - Mucosa |
| | | ERGIC3 | | | | | | 5.90E-06 | 0.28 | Brain - Hippocampus |
| | | | | | | | | 5.40E-05 | 0.077 | Thyroid |
| | | FER1L4 | | | | | | 0.00034 | -0.13 | Esophagus - Muscularis |
| | | GDF5 | | | | | | 3.70E-07 | 0.46 | Brain - Putamen (basal ganglia) |
| | | | | | | | | 7.20E-06 | 0.31 | Colon - Sigmoid |
| | | | | | | | | 4.90E-05 | 0.31 | Esophagus - Gastroesophageal Junction |
| | | | | | | | | 8.30E-05 | 0.24 | Esophagus - Muscularis |
| | | MMP24-AS1 | | | | | | 0.00031 | -0.11 | Adipose - Subcutaneous |
| | | PROCR | | | | | | 2.80E-05 | -0.16 | Adipose - Subcutaneous |
| | | RPL36P4 | | | | | | 1.90E-30 | 0.71 | Thyroid |
| | | | | | | | | 1.90E-27 | 0.68 | Esophagus - Mucosa |
| | | | | | | | | 4.20E-25 | 0.64 | Adipose - Subcutaneous |
| | | | | | | | | 1.60E-17 | 0.57 | Esophagus - Muscularis |
| | | | | | | | | 1.10E-16 | 0.71 | Colon - Sigmoid |
| | | | | | | | | 1.30E-15 | 0.57 | Adipose - Visceral (Omentum) |
| | | | | | | | | 3.30E-15 | 0.67 | Colon - Transverse |
| | | | | | | | | 3.00E-13 | 0.67 | Esophagus - Gastroesophageal Junction |
| | | | | | | | | 3.40E-13 | 0.67 | Stomach |
| | | | | | | | | 4.70E-09 | 0.55 | Brain - Nucleus accumbens (basal ganglia) |
| | | | | | | | | 1.40E-08 | 0.59 | Brain - Caudate (basal ganglia) |

| | | | | | | | | | | |
|---|---|---|---|---|---|---|---|---|---|---|
| | | | | | | | | 1.70E-08 | 0.69 | Brain - Cerebellum |
| | | | | | | | | 5.30E-08 | 0.67 | Small Intestine - Terminal Ileum |
| | | | | | | | | 1.40E-07 | 0.61 | Brain - Cortex |
| | | | | | | | | 1.80E-07 | 0.61 | Brain - Hippocampus |
| | | | | | | | | 2.00E-07 | 0.56 | Brain - Putamen (basal ganglia) |
| | | | | | | | | 3.10E-06 | 0.54 | Brain - Hypothalamus |
| | | | | | | | | 4.00E-06 | 0.54 | Brain - Anterior cingulate cortex (BA24) |
| | | | | | | | | 6.90E-06 | 0.53 | Brain - Frontal Cortex (BA9) |
| | | | | | | | | 1.20E-05 | 0.51 | Brain - Cerebellar Hemisphere |
| | | | | | | | | 3.30E-05 | 0.6 | Brain - Spinal cord (cervical c-1) |
| | | UQCC1 | | | | | | 2.30E-09 | -0.18 | Esophagus - Mucosa |
| | | | | | | | | 1.80E-08 | -0.18 | Adipose - Visceral (Omentum) |
| | | | | | | | | 1.40E-07 | -0.21 | Adipose - Subcutaneous |
| | | | | | | | | 2.10E-05 | -0.14 | Thyroid |
| | | | | | | | | 8.50E-05 | -0.16 | Stomach |
| | | | | | | | | 0.00014 | -0.14 | Colon - Sigmoid |
| AN & Sex hormone-binding globulin levels | rs34894639 | NCK1 | chr3 | 136079816 | 3q22.3 | C | T | 3.20E-05 | -0.16 | Esophagus - Mucosa |
| | | NCK1-AS1 | | | | | | 4.20E-07 | -0.33 | Esophagus - Mucosa |
| | | PCCB | | | | | | 3.80E-15 | -0.16 | Thyroid |
| | | | | | | | | 2.80E-09 | -0.17 | Esophagus - Muscularis |
| | | | | | | | | 3.40E-07 | -0.11 | Adipose - Visceral (Omentum) |
| | | | | | | | | 2.00E-06 | -0.15 | Stomach |
| | | | | | | | | 1.30E-05 | -0.18 | Esophagus - Gastroesophageal Junction |
| | | | | | | | | 2.10E-05 | -0.24 | Brain - Nucleus accumbens (basal ganglia) |
| | | | | | | | | 2.40E-05 | -0.32 | Brain - Frontal Cortex (BA9) |
| | | | | | | | | 5.70E-05 | -0.29 | Brain - Cortex |
| | | | | | | | | 0.00011 | -0.11 | Esophagus - Mucosa |
| | | RP11-102M11.2 | | | | | | 9.40E-06 | -0.39 | Brain - Hippocampus |
| | | RP11-731C17.2 | | | | | | 2.90E-06 | -0.45 | Brain - Hippocampus |
| | | | | | | | | 8.20E-05 | -0.31 | Brain - Cerebellar Hemisphere |
| AN & Alanine aminotransferase (quantile) | rs35614792 | BLOC1S2 | chr10 | 100174478 | 10q24.31 | C | T | 8.00E-10 | 0.2 | Adipose - Subcutaneous |
| | | | | | | | | 5.30E-08 | 0.18 | Thyroid |
| | | | | | | | | 4.00E-07 | 0.3 | Brain - Cerebellum |

| | | | | | | | | | | |
|---|---|---|---|---|---|---|---|---|---|---|
| | | | | | | | | 1.60E-05 | 0.26 | Small Intestine - Terminal Ileum |
| | | | | | | | | 5.30E-05 | 0.13 | Esophagus - Mucosa |
| | | CWF19L1 | | | | | | 7.30E-92 | 0.67 | Esophagus - Mucosa |
| | | | | | | | | 2.30E-81 | 0.59 | Thyroid |
| | | | | | | | | 5.30E-78 | 0.68 | Esophagus - Muscularis |
| | | | | | | | | 6.60E-56 | 0.54 | Adipose - Subcutaneous |
| | | | | | | | | 2.10E-50 | 0.65 | Adipose - Visceral (Omentum) |
| | | | | | | | | 5.80E-46 | 0.63 | Colon - Transverse |
| | | | | | | | | 4.90E-44 | 0.69 | Esophagus - Gastroesophageal Junction |
| | | | | | | | | 7.60E-41 | 0.71 | Stomach |
| | | | | | | | | 3.60E-38 | 0.77 | Brain - Cortex |
| | | | | | | | | 3.40E-35 | 0.61 | Colon - Sigmoid |
| | | | | | | | | 1.10E-31 | 0.79 | Brain - Frontal Cortex (BA9) |
| | | | | | | | | 1.10E-31 | 0.87 | Brain - Nucleus accumbens (basal ganglia) |
| | | | | | | | | 2.90E-28 | 0.68 | Brain - Cerebellum |
| | | | | | | | | 3.60E-28 | 0.81 | Brain - Caudate (basal ganglia) |
| | | | | | | | | 3.30E-27 | 0.76 | Brain - Putamen (basal ganglia) |
| | | | | | | | | 3.70E-26 | 0.76 | Brain - Anterior cingulate cortex (BA24) |
| | | | | | | | | 3.40E-24 | 0.87 | Brain - Hypothalamus |
| | | | | | | | | 5.60E-21 | 0.66 | Brain - Cerebellar Hemisphere |
| | | | | | | | | 9.40E-19 | 0.4 | Small Intestine - Terminal Ileum |
| | | | | | | | | 4.60E-17 | 0.66 | Brain - Spinal cord (cervical c-1) |
| | | | | | | | | 3.20E-14 | 0.67 | Brain - Hippocampus |
| | | | | | | | | 2.50E-12 | 0.75 | Brain - Amygdala |
| | | CWF19L1 | | | | | | 6.10E-11 | 0.73 | Brain - Substantia nigra |
| | | DNMBP | | | | | | 9.20E-08 | 0.25 | Thyroid |
| | | PHBP9 | | | | | | 3.20E-15 | 0.31 | Esophagus - Muscularis |
| | | | | | | | | 7.40E-13 | 0.29 | Adipose - Visceral (Omentum) |
| | | | | | | | | 2.20E-11 | 0.21 | Thyroid |
| | | | | | | | | 2.10E-10 | 0.21 | Adipose - Subcutaneous |
| | | | | | | | | 2.40E-09 | 0.26 | Esophagus - Mucosa |
| | | | | | | | | 1.70E-08 | 0.29 | Colon - Transverse |
| | | | | | | | | 4.60E-08 | 0.26 | Esophagus - Gastroesophageal Junction |

| | | | | | | | | | | |
|---|---|---|---|---|---|---|---|---|---|---|
| | | | | | | | | 8.90E-07 | 0.32 | Stomach |
| | | | | | | | | 2.00E-06 | 0.34 | Brain - Cortex |
| | | | | | | | | 4.70E-06 | 0.27 | Small Intestine - Terminal Ileum |
| | | | | | | | | 6.10E-06 | 0.24 | Colon - Sigmoid |
| AN & HDL cholesterol (quantile) | rs3768321 | BMP8A | chr1 | 39570256 | 1p34.3 | G | T | 8.50E-08 | 0.34 | Esophagus - Muscularis |
| | | | | | | | | 9.60E-06 | 0.44 | Brain - Hippocampus |
| | | | | | | | | 2.20E-05 | 0.28 | Adipose - Subcutaneous |
| | | | | | | | | 0.0002 | 0.13 | Thyroid |
| | | MACF1 | | | | | | 3.70E-08 | 0.21 | Thyroid |
| | | OXCT2P1 | | | | | | 1.20E-13 | 0.53 | Adipose - Visceral (Omentum) |
| | | | | | | | | 1.70E-13 | 0.54 | Esophagus - Muscularis |
| | | | | | | | | 7.90E-11 | 0.41 | Adipose - Subcutaneous |
| | | | | | | | | 7.00E-10 | 0.3 | Thyroid |
| | | | | | | | | 2.10E-08 | 0.5 | Colon - Sigmoid |
| | | | | | | | | 5.80E-08 | 0.38 | Esophagus - Mucosa |
| | | | | | | | | 3.80E-06 | 0.43 | Esophagus - Gastroesophageal Junction |
| | | | | | | | | 8.60E-06 | 0.54 | Small Intestine - Terminal Ileum |
| | | | | | | | | 2.10E-05 | 0.38 | Colon - Transverse |
| | | PABPC4 | | | | | | 1.10E-26 | -0.41 | Esophagus - Mucosa |
| | | | | | | | | 1.70E-21 | -0.28 | Adipose - Subcutaneous |
| | | | | | | | | 4.80E-12 | -0.23 | Thyroid |
| | | | | | | | | 1.20E-11 | -0.17 | Adipose - Visceral (Omentum) |
| | | | | | | | | 5.00E-10 | -0.24 | Colon - Transverse |
| | | | | | | | | 3.00E-08 | -0.21 | Esophagus - Gastroesophageal Junction |
| | | | | | | | | 6.40E-08 | -0.16 | Esophagus - Muscularis |
| | | | | | | | | 1.60E-05 | -0.18 | Colon - Sigmoid |
| | | | | | | | | 4.90E-05 | -0.13 | Stomach |
| | | RP11-69E11.4 | | | | | | 1.40E-10 | 0.35 | Adipose - Subcutaneous |
| | | | | | | | | 1.80E-08 | 0.29 | Adipose - Visceral (Omentum) |
| | | | | | | | | 2.80E-08 | 0.32 | Colon - Transverse |
| | | | | | | | | 3.70E-08 | 0.3 | Colon - Sigmoid |
| | | | | | | | | 1.60E-07 | 0.23 | Esophagus - Muscularis |
| | | | | | | | | 5.40E-07 | 0.24 | Thyroid |

| | | | | | | | | | | |
|---|---|---|---|---|---|---|---|---|---|---|
| | | | | | | | | 9.10E-07 | 0.22 | Esophagus - Mucosa |
| | | | | | | | | 9.60E-06 | 0.28 | Stomach |
| | | | | | | | | 1.30E-05 | 0.18 | Esophagus - Gastroesophageal Junction |
| | | | | | | | | 4.80E-05 | 0.3 | Brain - Nucleus accumbens (basal ganglia) |
| | | RP11-69E11.8 | | | | | | 2.20E-07 | -0.33 | Thyroid |
| | | | | | | | | 2.60E-06 | -0.31 | Esophagus - Mucosa |
| | | | | | | | | 6.90E-06 | -0.27 | Adipose - Subcutaneous |
| | | | | | | | | 5.10E-05 | -0.3 | Stomach |
| AN & HDL cholesterol (quantile) | rs3809114 | ARHGAP9 | chr12 | 57454856 | 12q13.3 | G | A | 4.10E-06 | 0.32 | Brain - Cerebellum |
| | | | | | | | | 3.90E-05 | 0.27 | Brain - Cerebellar Hemisphere |
| AN & Body mass index (BMI)<br>AN & Whole body fat mass | rs3927084 | APOBR | chr16 | 28627726 | 16p11.2 | A | C | 5.90E-05 | 0.11 | Thyroid |
| | | ATXN2L | | | | | | 6.80E-05 | -0.1 | Adipose - Visceral (Omentum) |
| | | | | | | | | 0.00015 | -0.085 | Adipose - Subcutaneous |
| | | CDC37P1 | | | | | | 1.50E-38 | 0.65 | Thyroid |
| | | | | | | | | 1.50E-33 | 0.62 | Adipose - Subcutaneous |
| | | | | | | | | 5.80E-24 | 0.66 | Colon - Transverse |
| | | | | | | | | 8.20E-15 | 0.53 | Stomach |
| | | | | | | | | 2.50E-11 | 0.63 | Small Intestine - Terminal Ileum |
| | | CDC37P2 | | | | | | 5.90E-06 | -0.19 | Thyroid |
| | | CLN3 | | | | | | 4.30E-06 | 0.11 | Thyroid |
| | | EIF3C | | | | | | 1.60E-10 | 0.28 | Adipose - Subcutaneous |
| | | | | | | | | 4.30E-07 | 0.23 | Esophagus - Muscularis |
| | | | | | | | | 2.10E-05 | 0.37 | Brain - Hypothalamus |
| | | | | | | | | 2.20E-05 | 0.3 | Brain - Hippocampus |
| | | | | | | | | 3.20E-05 | 0.3 | Brain - Nucleus accumbens (basal ganglia) |
| | | | | | | | | 7.50E-05 | 0.14 | Esophagus - Mucosa |
| | | LAT | | | | | | 1.90E-12 | 0.16 | Esophagus - Muscularis |
| | | | | | | | | 1.80E-05 | 0.23 | Brain - Putamen (basal ganglia) |
| | | | | | | | | 2.60E-05 | 0.23 | Brain - Cortex |
| | | NFATC2IP | | | | | | 2.90E-06 | 0.16 | Esophagus - Mucosa |
| | | | | | | | | 3.20E-05 | 0.2 | Brain - Caudate (basal ganglia) |
| | | | | | | | | 9.30E-05 | 0.12 | Adipose - Subcutaneous |
| | | NPIPB6 | | | | | | 0.0001 | 0.19 | Brain - Cerebellar Hemisphere |

| | | | | | | | | | | |
|---|---|---|---|---|---|---|---|---|---|---|
| | | NPIPB7 | | | | | | 1.30E-13 | -0.65 | Brain - Cortex |
| | | | | | | | | 1.50E-10 | -0.48 | Brain - Cerebellum |
| | | | | | | | | 1.80E-09 | -0.45 | Brain - Cerebellar Hemisphere |
| | | | | | | | | 9.70E-09 | -0.44 | Brain - Nucleus accumbens (basal ganglia) |
| | | | | | | | | 6.50E-08 | -0.46 | Brain - Hypothalamus |
| | | | | | | | | 1.90E-07 | -0.49 | Brain - Frontal Cortex (BA9) |
| | | | | | | | | 2.70E-07 | -0.54 | Brain - Anterior cingulate cortex (BA24) |
| | | | | | | | | 3.80E-06 | -0.47 | Brain - Hippocampus |
| | | | | | | | | 3.40E-05 | -0.41 | Brain - Putamen (basal ganglia) |
| | | | | | | | | 6.10E-05 | -0.23 | Colon - Transverse |
| | | NPIPB9 | | | | | | 2.20E-13 | -0.48 | Esophagus - Muscularis |
| | | | | | | | | 1.40E-09 | -0.45 | Colon - Sigmoid |
| | | | | | | | | 1.60E-07 | -0.43 | Esophagus - Gastroesophageal Junction |
| | | | | | | | | 5.90E-07 | -0.48 | Brain - Cortex |
| | | | | | | | | 2.50E-06 | -0.29 | Colon - Transverse |
| | | | | | | | | 2.90E-06 | -0.31 | Stomach |
| | | | | | | | | 3.30E-06 | -0.5 | Brain - Cerebellar Hemisphere |
| | | | | | | | | 6.50E-06 | -0.47 | Brain - Caudate (basal ganglia) |
| | | | | | | | | 9.20E-06 | -0.42 | Brain - Cerebellum |
| | | | | | | | | 1.00E-05 | -0.41 | Small Intestine - Terminal Ileum |
| | | | | | | | | 1.10E-05 | -0.27 | Adipose - Visceral (Omentum) |
| | | | | | | | | 2.80E-05 | -0.42 | Brain - Nucleus accumbens (basal ganglia) |
| | | | | | | | | 4.60E-05 | -0.2 | Thyroid |
| | | | | | | | | 8.80E-05 | -0.22 | Adipose - Subcutaneous |
| | | NUPR1 | | | | | | 4.50E-11 | -0.22 | Esophagus - Muscularis |
| | | | | | | | | 1.10E-06 | 0.31 | Brain - Cerebellar Hemisphere |
| | | | | | | | | 5.20E-05 | 0.12 | Thyroid |
| | | | | | | | | 5.20E-05 | 0.25 | Brain - Cerebellum |
| | | RP11-1348G14.4 | | | | | | 2.90E-28 | -0.55 | Adipose - Subcutaneous |
| | | | | | | | | 4.40E-21 | -0.49 | Esophagus - Muscularis |
| | | | | | | | | 1.10E-17 | -0.36 | Thyroid |
| | | | | | | | | 1.30E-14 | -0.37 | Adipose - Visceral (Omentum) |
| | | | | | | | | 3.30E-11 | -0.7 | Brain - Amygdala |

| | | | | | | | | | | |
|---|---|---|---|---|---|---|---|---|---|---|
| | | | | | | | | 1.10E-10 | -0.4 | Colon - Transverse |
| | | | | | | | | 9.40E-10 | -0.34 | Colon - Sigmoid |
| | | | | | | | | 1.10E-08 | -0.57 | Brain - Hypothalamus |
| | | | | | | | | 1.20E-08 | -0.47 | Brain - Nucleus accumbens (basal ganglia) |
| | | | | | | | | 4.20E-08 | -0.36 | Esophagus - Gastroesophageal Junction |
| | | | | | | | | 7.30E-08 | -0.39 | Brain - Cerebellum |
| | | | | | | | | 1.20E-07 | -0.34 | Stomach |
| | | | | | | | | 2.30E-07 | -0.49 | Brain - Hippocampus |
| | | | | | | | | 3.60E-07 | -0.46 | Small Intestine - Terminal Ileum |
| | | | | | | | | 1.00E-06 | -0.41 | Brain - Cortex |
| | | | | | | | | 1.10E-06 | -0.26 | Esophagus - Mucosa |
| | | | | | | | | 1.40E-06 | -0.38 | Brain - Cerebellar Hemisphere |
| | | | | | | | | 1.40E-05 | -0.45 | Brain - Anterior cingulate cortex (BA24) |
| | | | | | | | | 1.60E-05 | -0.43 | Brain - Frontal Cortex (BA9) |
| | | | | | | | | 3.60E-05 | -0.4 | Brain - Putamen (basal ganglia) |
| | | RP11-1348G14.6 | | | | | | 9.90E-20 | 0.49 | Thyroid |
| | | | | | | | | 8.70E-08 | 0.51 | Brain - Nucleus accumbens (basal ganglia) |
| | | RP11-22P6.2 | | | | | | 8.90E-08 | -0.42 | Brain - Cerebellar Hemisphere |
| | | | | | | | | 4.00E-07 | -0.36 | Brain - Cerebellum |
| | | RP11-426C22.5 | | | | | | 0.00014 | 0.15 | Adipose - Subcutaneous |
| | | RP11-435I10.5 | | | | | | 5.80E-30 | -0.58 | Thyroid |
| | | RP11-666O2.4 | | | | | | 7.50E-07 | 0.5 | Brain - Cerebellar Hemisphere |
| | | SBK1 | | | | | | 4.30E-06 | -0.23 | Esophagus - Muscularis |
| | | SGF29 | | | | | | 1.50E-06 | -0.15 | Colon - Sigmoid |
| | | | | | | | | 1.60E-06 | -0.13 | Esophagus - Muscularis |
| | | | | | | | | 2.70E-06 | -0.15 | Stomach |
| | | | | | | | | 6.00E-05 | -0.12 | Colon - Transverse |
| | | SH2B1 | | | | | | 3.50E-10 | -0.12 | Adipose - Visceral (Omentum) |
| | | | | | | | | 4.80E-10 | -0.13 | Adipose - Subcutaneous |
| | | | | | | | | 2.60E-09 | -0.1 | Esophagus - Muscularis |
| | | | | | | | | 3.50E-06 | -0.2 | Brain - Cerebellar Hemisphere |
| | | | | | | | | 1.20E-05 | -0.13 | Esophagus - Gastroesophageal Junction |

| | | | | | | | | | | |
|---|---|---|---|---|---|---|---|---|---|---|
| | | SPNS1 | | | | | | 1.70E-05 | 0.12 | Esophagus - Muscularis |
| | | SULT1A1 | | | | | | 2.80E-20 | -0.41 | Thyroid |
| | | | | | | | | 7.10E-19 | -0.4 | Esophagus - Mucosa |
| | | | | | | | | 3.50E-08 | -0.39 | Brain - Hypothalamus |
| | | | | | | | | 2.40E-07 | -0.46 | Brain - Anterior cingulate cortex (BA24) |
| | | | | | | | | 1.20E-06 | -0.28 | Esophagus - Gastroesophageal Junction |
| | | | | | | | | 1.20E-06 | -0.22 | Stomach |
| | | | | | | | | 1.80E-06 | -0.38 | Brain - Cerebellum |
| | | | | | | | | 3.30E-06 | -0.25 | Colon - Sigmoid |
| | | | | | | | | 5.00E-06 | -0.41 | Brain - Amygdala |
| | | | | | | | | 5.90E-06 | -0.4 | Brain - Caudate (basal ganglia) |
| | | | | | | | | 8.80E-06 | -0.39 | Brain - Cortex |
| | | | | | | | | 7.70E-05 | -0.37 | Brain - Cerebellar Hemisphere |
| | | SULT1A2 | | | | | | 9.70E-42 | 0.55 | Adipose - Subcutaneous |
| | | | | | | | | 5.70E-39 | 0.54 | Adipose - Visceral (Omentum) |
| | | | | | | | | 1.10E-27 | 0.6 | Esophagus - Mucosa |
| | | | | | | | | 3.20E-27 | 0.53 | Esophagus - Muscularis |
| | | | | | | | | 1.10E-21 | 0.53 | Esophagus - Gastroesophageal Junction |
| | | | | | | | | 3.80E-20 | 0.59 | Stomach |
| | | | | | | | | 3.10E-15 | 0.44 | Colon - Sigmoid |
| | | | | | | | | 2.60E-13 | 0.74 | Brain - Anterior cingulate cortex (BA24) |
| | | | | | | | | 5.40E-11 | 0.64 | Brain - Putamen (basal ganglia) |
| | | | | | | | | 3.70E-10 | 0.59 | Brain - Cortex |
| | | | | | | | | 1.00E-09 | 0.51 | Brain - Nucleus accumbens (basal ganglia) |
| | | | | | | | | 2.80E-09 | 0.59 | Brain - Caudate (basal ganglia) |
| | | | | | | | | 1.10E-08 | 0.57 | Brain - Hypothalamus |
| | | | | | | | | 1.50E-08 | 0.51 | Brain - Hippocampus |
| | | | | | | | | 2.80E-07 | 0.61 | Brain - Amygdala |
| | | | | | | | | 1.10E-06 | 0.5 | Brain - Frontal Cortex (BA9) |
| | | | | | | | | 2.00E-06 | 0.12 | Colon - Transverse |
| | | | | | | | | 2.10E-06 | 0.55 | Brain - Spinal cord (cervical c-1) |
| | | TUFM | | | | | | 1.50E-41 | 0.27 | Thyroid |
| | | | | | | | | 1.90E-23 | 0.27 | Esophagus - Muscularis |

| | | | | | | | | | | |
|---|---|---|---|---|---|---|---|---|---|---|
| | | | | | | | | 2.10E-20 | 0.21 | Adipose - Subcutaneous |
| | | | | | | | | 6.70E-18 | 0.29 | Colon - Sigmoid |
| | | | | | | | | 1.00E-16 | 0.3 | Esophagus - Gastroesophageal Junction |
| | | | | | | | | 8.00E-14 | 0.29 | Brain - Nucleus accumbens (basal ganglia) |
| | | | | | | | | 2.10E-13 | 0.15 | Esophagus - Mucosa |
| | | | | | | | | 1.10E-11 | 0.14 | Colon - Transverse |
| | | | | | | | | 1.10E-11 | 0.24 | Brain - Putamen (basal ganglia) |
| | | | | | | | | 2.50E-09 | 0.26 | Brain - Hippocampus |
| | | | | | | | | 5.60E-09 | 0.13 | Stomach |
| | | | | | | | | 7.70E-09 | 0.092 | Adipose - Visceral (Omentum) |
| | | | | | | | | 1.70E-08 | 0.3 | Brain - Amygdala |
| | | | | | | | | 3.20E-08 | 0.14 | Small Intestine - Terminal Ileum |
| | | | | | | | | 7.30E-08 | 0.19 | Brain - Caudate (basal ganglia) |
| | | | | | | | | 7.40E-08 | 0.2 | Brain - Cortex |
| | | | | | | | | 8.50E-07 | 0.23 | Brain - Frontal Cortex (BA9) |
| | | | | | | | | 2.50E-06 | 0.25 | Brain - Anterior cingulate cortex (BA24) |
| AN & Body mass index (BMI)<br>AN & Whole body fat mass | rs394608 | ADARB1 | chr21 | 45161883 | 21q22.3 | T | C | 9.30E-07 | 0.26 | Brain - Cerebellum |
| | | LINC00165 | | | | | | 1.20E-05 | 0.21 | Adipose - Visceral (Omentum) |
| | | PICSAR | | | | | | 2.20E-05 | 0.4 | Brain - Putamen (basal ganglia) |
| | | PICSAR | | | | | | 3.40E-05 | 0.2 | Adipose - Visceral (Omentum) |
| AN & four psychiatric traits<br>AN & Nervous feelings<br>AN & Schizophrenia<br>AN & Tense / 'highly strung' | rs4129585 | ADGRB1 | chr8 | 142231572 | 8q24.3 | A | C | 3.60E-05 | -0.18 | Esophagus - Mucosa |
| | | TSNARE1 | | 142231572 | | A | C | 5.70E-06 | 0.16 | Adipose - Subcutaneous |
| AN & Major Depressive Disorder<br>AN & Tense / 'highly strung' | rs4281434 | ADK | chr10 | 74483014 | 10q22.2 | A | G | 1.80E-10 | -0.38 | Colon - Transverse |
| | | | | | | | | 3.70E-09 | 0.13 | Esophagus - Mucosa |
| | | | | | | | | 1.50E-08 | -0.38 | Colon - Sigmoid |
| | | | | | | | | 8.70E-06 | -0.17 | Esophagus - Muscularis |
| | | | | | | | | 0.00035 | 0.11 | Thyroid |
| | | AP3M1 | | | | | | 7.60E-06 | -0.19 | Colon - Sigmoid |
| | | | | | | | | 0.00013 | -0.16 | Colon - Transverse |
| | | SEC24C | | | | | | 0.00016 | 0.095 | Esophagus - Mucosa |

| | | | | | | | | | | |
|---|---|---|---|---|---|---|---|---|---|---|
| AN & Whole body fat mass | rs4320241 | GCNT4 | chr5 | 75003319 | 5q13.3 | C | T | 2.90E-06 | -0.19 | Esophagus - Muscularis |
| | | LINC01336 | | | | | | 0.00011 | -0.23 | Esophagus - Muscularis |
| AN & Worrier / anxious feelings | rs4338470 | ADK | chr10 | 74473690 | 10q22.2 | G | A | 5.80E-10 | -0.35 | Colon - Transverse |
| | | | | | | | | 1.60E-07 | 0.11 | Esophagus - Mucosa |
| | | | | | | | | 1.10E-06 | -0.32 | Colon - Sigmoid |
| | | | | | | | | 3.40E-05 | -0.16 | Esophagus - Muscularis |
| | | AP3M1 | | | | | | 5.10E-06 | -0.19 | Colon - Sigmoid |
| | | SEC24C | | | | | | 4.00E-05 | 0.1 | Esophagus - Mucosa |
| AN & C-reactive protein (quantile) | rs439401 | APOC1 | chr19 | 44911194 | 19q13.32 | T | C | 6.60E-06 | -0.24 | Adipose - Visceral (Omentum) |
| | | APOC1P1 | | | | | | 4.00E-06 | -0.24 | Adipose - Subcutaneous |
| | | CTB-171A8.1 | | | | | | 2.70E-05 | 0.26 | Thyroid |
| AN & Urate (quantile) | rs4697893 | DRD5 | chr4 | 9824559 | 4p16.1 | T | C | 1.50E-05 | 0.96 | Brain - Anterior cingulate cortex (BA24) |
| | | ENPP7P11 | | | | | | 0.00011 | -0.82 | Adipose - Visceral (Omentum) |
| | | | | | | | | 0.00016 | -0.79 | Thyroid |
| | | UNC93B7 | | | | | | 0.00013 | -0.82 | Thyroid |
| AN & Basal metabolic rate<br>AN & Body mass index (BMI)<br>AN & Reticulocyte count | rs4751 | CDK11A | chr1 | 1754601 | 1p36.33 | G | T | 5.50E-05 | 0.19 | Thyroid |
| | | CFAP74 | | | | | | 4.80E-15 | -0.47 | Brain - Cortex |
| | | | | | | | | 2.80E-05 | -0.28 | Brain - Frontal Cortex (BA9) |
| | | LINC01770 | | | | | | 5.90E-06 | -0.3 | Small Intestine - Terminal Ileum |
| | | MMP23A | | | | | | 2.30E-07 | 0.29 | Thyroid |
| | | | | | | | | 9.00E-07 | 0.44 | Brain - Nucleus accumbens (basal ganglia) |
| | | | | | | | | 1.30E-05 | 0.24 | Colon - Transverse |
| | | | | | | | | 3.70E-05 | 0.28 | Adipose - Visceral (Omentum) |
| | | | | | | | | 9.60E-05 | 0.24 | Esophagus - Mucosa |
| | | NADK | | | | | | 5.60E-07 | 0.34 | Brain - Cerebellar Hemisphere |
| | | | | | | | | 5.70E-07 | 0.29 | Brain - Cerebellum |
| | | | | | | | | 4.80E-05 | 0.13 | Esophagus - Gastroesophageal Junction |
| | | | | | | | | 8.90E-05 | -0.12 | Esophagus - Mucosa |
| | | RP1-140A9.1 | | | | | | 1.50E-20 | -0.43 | Thyroid |
| | | | | | | | | 7.40E-14 | -0.46 | Stomach |
| | | | | | | | | 1.70E-12 | -0.37 | Esophagus - Mucosa |
| | | | | | | | | 1.80E-12 | -0.31 | Adipose - Subcutaneous |
| | | | | | | | | 5.10E-11 | -0.38 | Colon - Transverse |

| | | | | | | | | | | |
|---|---|---|---|---|---|---|---|---|---|---|
| | | | | | | | | 3.70E-08 | -0.42 | Brain - Caudate (basal ganglia) |
| | | | | | | | | 6.20E-08 | -0.48 | Brain - Cortex |
| | | | | | | | | 3.70E-07 | -0.37 | Brain - Cerebellar Hemisphere |
| | | | | | | | | 6.00E-07 | -0.45 | Brain - Nucleus accumbens (basal ganglia) |
| | | | | | | | | 1.70E-06 | -0.31 | Esophagus - Muscularis |
| | | | | | | | | 1.80E-06 | -0.46 | Brain - Frontal Cortex (BA9) |
| | | | | | | | | 5.40E-06 | -0.38 | Small Intestine - Terminal Ileum |
| | | | | | | | | 9.10E-06 | -0.34 | Esophagus - Gastroesophageal Junction |
| | | | | | | | | 1.20E-05 | -0.22 | Adipose - Visceral (Omentum) |
| | | | | | | | | 1.40E-05 | -0.43 | Brain - Hypothalamus |
| | | | | | | | | 1.60E-05 | -0.4 | Brain - Hippocampus |
| | | | | | | | | 1.90E-05 | -0.53 | Brain - Spinal cord (cervical c-1) |
| | | | | | | | | 2.90E-05 | -0.3 | Colon - Sigmoid |
| | | | | | | | | 6.00E-05 | -0.3 | Brain - Cerebellum |
| | | RP1-283E3.4 | | | | | | 9.20E-07 | 0.31 | Colon - Sigmoid |
| | | | | | | | | 1.80E-06 | 0.31 | Stomach |
| | | | | | | | | 2.90E-05 | 0.2 | Thyroid |
| | | | | | | | | 3.90E-05 | 0.3 | Esophagus - Gastroesophageal Junction |
| | | | | | | | | 4.00E-05 | 0.28 | Colon - Transverse |
| | | | | | | | | 5.80E-05 | 0.21 | Esophagus - Mucosa |
| | | SLC35E2 | | | | | | 6.30E-09 | 0.26 | Esophagus - Mucosa |
| | | | | | | | | 2.90E-08 | 0.25 | Adipose - Subcutaneous |
| | | | | | | | | 9.80E-08 | 0.35 | Stomach |
| | | | | | | | | 1.50E-05 | 0.19 | Thyroid |
| | | | | | | | | 2.50E-05 | 0.2 | Adipose - Visceral (Omentum) |
| | | | | | | | | 4.60E-05 | 0.22 | Colon - Sigmoid |
| | | TMEM52 | | | | | | 1.40E-28 | -0.53 | Esophagus - Mucosa |
| AN & C-reactive protein (quantile) | rs4845373 | IL6R | chr1 | 154445353 | 1q21.3 | C | T | 4.10E-15 | -0.33 | Colon - Transverse |
| | | | | | | | | 2.10E-06 | -0.32 | Colon - Sigmoid |
| | | | | | | | | 5.10E-06 | -0.17 | Esophagus - Muscularis |
| | | | | | | | | 8.20E-06 | -0.19 | Esophagus - Gastroesophageal Junction |
| | | | | | | | | 0.00012 | -0.16 | Esophagus - Mucosa |
| | | TDRD10 | | | | | | 3.10E-08 | 0.16 | Adipose - Visceral (Omentum) |

| | | | | | | | | | | |
|---|---|---|---|---|---|---|---|---|---|---|
| | | | | | | | | 4.30E-07 | 0.14 | Esophagus - Muscularis |
| | | | | | | | | 0.00029 | 0.11 | Thyroid |
| AN & Sex hormone-binding globulin levels adjusted for BMI | rs4858828 | AMT | chr3 | 48687378 | 3p21.31 | C | T | 2.80E-31 | 0.28 | Thyroid |
| | | | | | | | | 2.70E-27 | 0.24 | Adipose - Subcutaneous |
| | | | | | | | | 4.10E-24 | 0.2 | Esophagus - Muscularis |
| | | | | | | | | 7.20E-22 | 0.22 | Adipose - Visceral (Omentum) |
| | | | | | | | | 3.50E-19 | 0.19 | Esophagus - Mucosa |
| | | | | | | | | 2.10E-15 | 0.46 | Brain - Cerebellum |
| | | | | | | | | 7.30E-13 | 0.41 | Brain - Cerebellar Hemisphere |
| | | | | | | | | 8.70E-13 | 0.2 | Esophagus - Gastroesophageal Junction |
| | | | | | | | | 3.90E-11 | 0.21 | Colon - Sigmoid |
| | | | | | | | | 8.70E-10 | 0.38 | Brain - Cortex |
| | | | | | | | | 2.50E-08 | 0.18 | Colon - Transverse |
| | | | | | | | | 3.80E-08 | 0.32 | Brain - Caudate (basal ganglia) |
| | | | | | | | | 9.70E-08 | 0.27 | Brain - Nucleus accumbens (basal ganglia) |
| | | | | | | | | 1.20E-07 | 0.28 | Brain - Hippocampus |
| | | | | | | | | 1.80E-07 | 0.28 | Brain - Frontal Cortex (BA9) |
| | | | | | | | | 8.70E-07 | 0.18 | Stomach |
| | | | | | | | | 1.40E-05 | 0.3 | Brain - Spinal cord (cervical c-1) |
| | | | | | | | | 0.00014 | 0.19 | Brain - Anterior cingulate cortex (BA24) |
| | | ARIH2 | | | | | | 5.20E-20 | 0.27 | Thyroid |
| | | | | | | | | 1.30E-13 | 0.17 | Adipose - Subcutaneous |
| | | | | | | | | 9.10E-09 | 0.17 | Colon - Transverse |
| | | | | | | | | 1.40E-08 | 0.16 | Esophagus - Mucosa |
| | | | | | | | | 4.40E-08 | 0.19 | Stomach |
| | | | | | | | | 9.20E-06 | 0.11 | Esophagus - Gastroesophageal Junction |
| | | | | | | | | 8.10E-05 | 0.21 | Small Intestine - Terminal Ileum |
| | | ATRIP | | | | | | 3.40E-05 | -0.13 | Esophagus - Mucosa |
| | | BSN-AS2 | | | | | | 2.10E-05 | -0.38 | Brain - Putamen (basal ganglia) |
| | | | | | | | | 5.60E-05 | -0.32 | Brain - Cortex |
| | | C3orf62 | | | | | | 0.0002 | 0.12 | Adipose - Visceral (Omentum) |
| | | CCDC36 | | | | | | 4.30E-17 | 0.5 | Esophagus - Muscularis |
| | | | | | | | | 2.80E-16 | 0.44 | Adipose - Visceral (Omentum) |

| | | | | | | | | | | |
|---|---|---|---|---|---|---|---|---|---|---|
| | | | | | | | | 1.30E-15 | 0.5 | Colon - Sigmoid |
| | | | | | | | | 9.40E-15 | 0.5 | Esophagus - Gastroesophageal Junction |
| | | | | | | | | 1.80E-11 | 0.32 | Adipose - Subcutaneous |
| | | | | | | | | 3.60E-11 | 0.26 | Colon - Transverse |
| | | | | | | | | 1.40E-09 | 0.32 | Esophagus - Mucosa |
| | | | | | | | | 4.00E-06 | 0.17 | Thyroid |
| | | | | | | | | 5.80E-05 | 0.22 | Stomach |
| | | CCDC71 | | | | | | 4.40E-17 | -0.46 | Brain - Cerebellum |
| | | | | | | | | 1.30E-16 | -0.53 | Brain - Cerebellar Hemisphere |
| | | | | | | | | 9.00E-07 | -0.34 | Brain - Frontal Cortex (BA9) |
| | | | | | | | | 6.40E-06 | -0.089 | Thyroid |
| | | | | | | | | 3.40E-05 | -0.3 | Brain - Nucleus accumbens (basal ganglia) |
| | | | | | | | | 5.10E-05 | -0.27 | Brain - Hippocampus |
| | | | | | | | | 5.40E-05 | -0.2 | Brain - Caudate (basal ganglia) |
| | | | | | | | | 7.80E-05 | -0.27 | Brain - Putamen (basal ganglia) |
| | | CDC25A | | | | | | 0.00028 | 0.2 | Adipose - Subcutaneous |
| | | | | | | | | 0.00062 | 0.15 | Thyroid |
| | | CELSR3 | | | | | | 3.80E-07 | 0.18 | Adipose - Subcutaneous |
| | | DALRD3 | | | | | | 5.70E-18 | 0.25 | Adipose - Subcutaneous |
| | | | | | | | | 7.60E-10 | 0.2 | Esophagus - Mucosa |
| | | | | | | | | 1.70E-09 | 0.34 | Brain - Cerebellum |
| | | | | | | | | 3.90E-09 | 0.17 | Adipose - Visceral (Omentum) |
| | | | | | | | | 1.40E-08 | 0.27 | Brain - Cortex |
| | | | | | | | | 1.00E-07 | 0.31 | Brain - Cerebellar Hemisphere |
| | | | | | | | | 1.60E-06 | 0.11 | Thyroid |
| | | | | | | | | 7.10E-05 | 0.12 | Esophagus - Muscularis |
| | | | | | | | | 0.00014 | 0.17 | Brain - Frontal Cortex (BA9) |
| | | GPX1 | | | | | | 7.50E-14 | -0.16 | Thyroid |
| | | | | | | | | 9.50E-09 | -0.19 | Brain - Caudate (basal ganglia) |
| | | | | | | | | 5.90E-08 | -0.19 | Esophagus - Mucosa |
| | | | | | | | | 3.30E-06 | -0.21 | Brain - Nucleus accumbens (basal ganglia) |
| | | | | | | | | 1.50E-05 | -0.24 | Brain - Cortex |
| | | | | | | | | 8.10E-05 | -0.12 | Colon - Transverse |

| | | | | | | | | | | |
|---|---|---|---|---|---|---|---|---|---|---|
| | | | | | | | | 0.00025 | -0.18 | Brain - Cerebellum |
| | | | | | | | | 0.00026 | -0.072 | Adipose - Visceral (Omentum) |
| | | KLHDC8B | | | | | | 1.60E-07 | -0.14 | Thyroid |
| | | | | | | | | 8.50E-07 | -0.14 | Esophagus - Muscularis |
| | | | | | | | | 1.50E-05 | -0.22 | Brain - Cerebellum |
| | | MRPS18AP1 | | | | | | 1.80E-07 | 0.29 | Thyroid |
| | | | | | | | | 4.20E-07 | 0.29 | Adipose - Subcutaneous |
| | | | | | | | | 1.60E-05 | 0.28 | Esophagus - Muscularis |
| | | | | | | | | 4.60E-05 | 0.26 | Esophagus - Mucosa |
| | | | | | | | | 9.00E-05 | 0.31 | Esophagus - Gastroesophageal Junction |
| | | NCKIPSD | | | | | | 2.10E-48 | 0.54 | Esophagus - Muscularis |
| | | | | | | | | 4.20E-40 | 0.48 | Thyroid |
| | | | | | | | | 1.10E-39 | 0.46 | Esophagus - Mucosa |
| | | | | | | | | 1.40E-39 | 0.4 | Adipose - Subcutaneous |
| | | | | | | | | 6.30E-37 | 0.58 | Esophagus - Gastroesophageal Junction |
| | | | | | | | | 1.00E-35 | 0.5 | Colon - Sigmoid |
| | | | | | | | | 8.20E-35 | 0.42 | Adipose - Visceral (Omentum) |
| | | | | | | | | 6.30E-34 | 0.41 | Brain - Cortex |
| | | | | | | | | 4.90E-33 | 0.44 | Brain - Caudate (basal ganglia) |
| | | | | | | | | 2.60E-30 | 0.48 | Brain - Putamen (basal ganglia) |
| | | | | | | | | 1.40E-27 | 0.44 | Brain - Frontal Cortex (BA9) |
| | | | | | | | | 1.00E-25 | 0.42 | Brain - Anterior cingulate cortex (BA24) |
| | | | | | | | | 1.80E-25 | 0.41 | Colon - Transverse |
| | | | | | | | | 1.90E-24 | 0.46 | Brain - Cerebellum |
| | | | | | | | | 2.80E-23 | 0.38 | Stomach |
| | | | | | | | | 6.60E-23 | 0.54 | Brain - Cerebellar Hemisphere |
| | | | | | | | | 8.60E-22 | 0.33 | Brain - Nucleus accumbens (basal ganglia) |
| | | | | | | | | 1.30E-18 | 0.38 | Brain - Hippocampus |
| | | | | | | | | 8.40E-17 | 0.78 | Brain - Spinal cord (cervical c-1) |
| | | | | | | | | 1.40E-15 | 0.49 | Brain - Amygdala |
| | | | | | | | | 6.20E-15 | 0.36 | Brain - Hypothalamus |
| | | | | | | | | 1.80E-11 | 0.45 | Brain - Substantia nigra |
| | | | | | | | | 7.70E-06 | 0.31 | Small Intestine - Terminal Ileum |

| | | | | | | | | | | |
|---|---|---|---|---|---|---|---|---|---|---|
| | | NDUFAF3 | | | | | | 2.70E-05 | 0.11 | Esophagus - Muscularis |
| | | NICN1 | | | | | | 1.40E-20 | 0.26 | Thyroid |
| | | | | | | | | 3.20E-10 | 0.16 | Adipose - Subcutaneous |
| | | | | | | | | 1.00E-07 | 0.13 | Esophagus - Muscularis |
| | | | | | | | | 1.10E-06 | 0.14 | Adipose - Visceral (Omentum) |
| | | | | | | | | 1.60E-06 | 0.17 | Esophagus - Gastroesophageal Junction |
| | | | | | | | | 6.70E-05 | -0.17 | Brain - Nucleus accumbens (basal ganglia) |
| | | | | | | | | 0.00014 | 0.13 | Colon - Sigmoid |
| | | | | | | | | 0.00017 | -0.13 | Brain - Frontal Cortex (BA9) |
| | | NME6 | | | | | | 6.20E-05 | -0.12 | Adipose - Subcutaneous |
| | | | | | | | | 0.00014 | -0.15 | Esophagus - Muscularis |
| | | | | | | | | 0.00064 | -0.11 | Thyroid |
| | | P4HTM | | | | | | 1.40E-18 | 0.21 | Thyroid |
| | | | | | | | | 9.40E-11 | 0.43 | Brain - Spinal cord (cervical c-1) |
| | | | | | | | | 2.80E-10 | 0.19 | Brain - Frontal Cortex (BA9) |
| | | | | | | | | 2.90E-10 | 0.14 | Adipose - Subcutaneous |
| | | | | | | | | 5.80E-09 | 0.12 | Adipose - Visceral (Omentum) |
| | | | | | | | | 3.60E-08 | 0.14 | Esophagus - Muscularis |
| | | | | | | | | 6.50E-08 | 0.16 | Brain - Cortex |
| | | | | | | | | 9.20E-08 | 0.21 | Colon - Sigmoid |
| | | | | | | | | 1.70E-07 | 0.17 | Brain - Hypothalamus |
| | | | | | | | | 1.10E-06 | 0.24 | Brain - Cerebellum |
| | | | | | | | | 2.40E-06 | 0.12 | Colon - Transverse |
| | | | | | | | | 8.80E-06 | 0.11 | Brain - Nucleus accumbens (basal ganglia) |
| | | | | | | | | 4.30E-05 | 0.11 | Brain - Putamen (basal ganglia) |
| | | | | | | | | 4.90E-05 | 0.13 | Esophagus - Gastroesophageal Junction |
| | | | | | | | | 0.00017 | 0.19 | Brain - Cerebellar Hemisphere |
| | | PRKAR2A | | | | | | 0.00011 | -0.077 | Adipose - Subcutaneous |
| | | QRICH1 | | | | | | 6.20E-14 | -0.21 | Adipose - Subcutaneous |
| | | | | | | | | 1.60E-11 | -0.13 | Thyroid |
| | | | | | | | | 1.30E-08 | -0.15 | Esophagus - Muscularis |
| | | | | | | | | 1.00E-07 | -0.16 | Colon - Transverse |
| | | | | | | | | 6.10E-07 | -0.26 | Brain - Cerebellar Hemisphere |

| | | | | | | | | | | |
|---|---|---|---|---|---|---|---|---|---|---|
| | | | | | | | | 1.10E-06 | -0.22 | Brain - Cerebellum |
| | | | | | | | | 6.70E-05 | -0.12 | Adipose - Visceral (Omentum) |
| | | | | | | | | 0.00038 | -0.088 | Esophagus - Mucosa |
| | | RHOA-IT1 | | | | | | 0.00047 | -0.15 | Thyroid |
| | | RP11-3B7.1 | | | | | | 1.20E-13 | 0.41 | Adipose - Visceral (Omentum) |
| | | | | | | | | 1.20E-13 | 0.51 | Esophagus - Gastroesophageal Junction |
| | | | | | | | | 1.90E-13 | 0.46 | Esophagus - Muscularis |
| | | | | | | | | 1.70E-12 | 0.49 | Colon - Sigmoid |
| | | | | | | | | 7.10E-05 | 0.19 | Colon - Transverse |
| | | | | | | | | 0.00023 | 0.18 | Adipose - Subcutaneous |
| | | RP11-694I15.7 | | | | | | 4.80E-05 | 0.16 | Esophagus - Muscularis |
| | | RP13-131K19.6 | | | | | | 1.90E-05 | -0.22 | Esophagus - Muscularis |
| | | | | | | | | 3.20E-05 | -0.25 | Esophagus - Gastroesophageal Junction |
| | | | | | | | | 3.50E-05 | -0.23 | Colon - Sigmoid |
| | | SLC26A6 | | | | | | 3.70E-06 | -0.1 | Thyroid |
| | | | | | | | | 0.00033 | -0.11 | Adipose - Subcutaneous |
| | | TCTA | | | | | | 6.30E-07 | 0.19 | Esophagus - Gastroesophageal Junction |
| | | | | | | | | 4.90E-06 | 0.078 | Thyroid |
| | | | | | | | | 1.30E-05 | 0.12 | Adipose - Subcutaneous |
| | | | | | | | | 0.00034 | 0.11 | Colon - Transverse |
| | | | | | | | | 0.00046 | 0.11 | Esophagus - Muscularis |
| | | WDR6 | | | | | | 2.00E-102 | -0.67 | Adipose - Subcutaneous |
| | | | | | | | | 6.40E-56 | -0.61 | Esophagus - Mucosa |
| | | | | | | | | 2.50E-46 | -0.4 | Adipose - Visceral (Omentum) |
| | | | | | | | | 3.40E-23 | -0.42 | Brain - Cerebellum |
| | | | | | | | | 1.60E-22 | -0.31 | Colon - Transverse |
| | | | | | | | | 2.10E-21 | -0.4 | Stomach |
| | | | | | | | | 2.00E-20 | -0.22 | Esophagus - Muscularis |
| | | | | | | | | 5.10E-16 | -0.39 | Brain - Cerebellar Hemisphere |
| | | | | | | | | 9.00E-15 | -0.25 | Esophagus - Gastroesophageal Junction |
| | | | | | | | | 8.20E-12 | -0.34 | Brain - Frontal Cortex (BA9) |
| | | | | | | | | 3.10E-11 | -0.29 | Brain - Putamen (basal ganglia) |

| | | | | | | | | | | |
|---|---|---|---|---|---|---|---|---|---|---|
| | | | | | | | | 6.80E-11 | -0.3 | Brain - Nucleus accumbens (basal ganglia) |
| | | | | | | | | 1.10E-10 | -0.31 | Brain - Cortex |
| | | | | | | | | 6.00E-08 | -0.12 | Thyroid |
| | | | | | | | | 6.50E-08 | -0.25 | Brain - Hippocampus |
| | | | | | | | | 6.60E-08 | -0.21 | Brain - Caudate (basal ganglia) |
| | | | | | | | | 7.40E-07 | -0.3 | Brain - Substantia nigra |
| | | | | | | | | 9.80E-07 | -0.29 | Small Intestine - Terminal Ileum |
| | | | | | | | | 1.40E-06 | -0.35 | Brain - Spinal cord (cervical c-1) |
| | | | | | | | | 2.10E-05 | -0.26 | Brain - Amygdala |
| | | ZNF589 | | | | | | 3.90E-06 | 0.17 | Thyroid |
| | | ZNF589 | | | | | | 1.60E-05 | 0.21 | Esophagus - Mucosa |
| | | ZNF589 | | | | | | 2.10E-05 | 0.17 | Adipose - Subcutaneous |
| | | ZNF589 | | | | | | 4.70E-05 | 0.24 | Esophagus - Gastroesophageal Junction |
| | | ZNF589 | | | | | | 0.00021 | 0.17 | Esophagus - Muscularis |
| AN & C-reactive protein (quantile) | rs55709272 | IL1RN | chr2 | 113109711 | 2q13 | T | C | 2.30E-08 | -0.2 | Thyroid |
| AN & C-reactive protein (quantile) | rs55709272 | IL1RN | chr2 | 113109711 | 2q13 | T | C | 4.60E-06 | -0.13 | Adipose - Subcutaneous |
| AN & HDL cholesterol (quantile) | rs59781045 | AP000892.6 | chr11 | 117224567 | 11q23.3 | C | T | 5.90E-16 | -0.39 | Adipose - Visceral (Omentum) |
| AN & HDL cholesterol (quantile) | rs59781045 | AP000892.6 | chr11 | 117224567 | 11q23.3 | C | T | 2.20E-15 | -0.49 | Esophagus - Mucosa |
| AN & HDL cholesterol (quantile) | rs59781045 | AP000892.6 | chr11 | 117224567 | 11q23.3 | C | T | 3.30E-14 | -0.53 | Esophagus - Muscularis |
| AN & HDL cholesterol (quantile) | rs59781045 | AP000892.6 | chr11 | 117224567 | 11q23.3 | C | T | 5.40E-14 | -0.39 | Adipose - Subcutaneous |
| AN & HDL cholesterol (quantile) | rs59781045 | AP000892.6 | chr11 | 117224567 | 11q23.3 | C | T | 1.30E-09 | -0.32 | Thyroid |
| AN & HDL cholesterol (quantile) | rs59781045 | AP000892.6 | chr11 | 117224567 | 11q23.3 | C | T | 6.20E-07 | -0.43 | Esophagus - Gastroesophageal Junction |
| AN & HDL cholesterol (quantile) | rs59781045 | AP000892.6 | chr11 | 117224567 | 11q23.3 | C | T | 2.00E-06 | -0.22 | Colon - Transverse |
| AN & HDL cholesterol (quantile) | rs59781045 | AP000892.6 | chr11 | 117224567 | 11q23.3 | C | T | 2.20E-06 | -0.35 | Colon - Sigmoid |
| AN & HDL cholesterol (quantile) | rs59781045 | PAFAH1B2 | chr11 | 117224567 | 11q23.3 | C | T | 6.00E-05 | -0.25 | Brain - Cerebellum |
| AN & HDL cholesterol (quantile) | rs59781045 | RP11-109L13.1 | chr11 | 117224567 | 11q23.3 | C | T | 1.30E-20 | 0.89 | Esophagus - Mucosa |
| AN & HDL cholesterol (quantile) | rs59781045 | RP11-109L13.1 | chr11 | 117224567 | 11q23.3 | C | T | 1.60E-18 | 0.83 | Thyroid |
| AN & HDL cholesterol (quantile) | rs59781045 | RP11-109L13.1 | chr11 | 117224567 | 11q23.3 | C | T | 1.70E-18 | 0.84 | Adipose - Subcutaneous |
| AN & HDL cholesterol (quantile) | rs59781045 | RP11-109L13.1 | chr11 | 117224567 | 11q23.3 | C | T | 2.10E-18 | 0.86 | Adipose - Visceral (Omentum) |
| AN & HDL cholesterol (quantile) | rs59781045 | RP11-109L13.1 | chr11 | 117224567 | 11q23.3 | C | T | 2.70E-17 | 0.84 | Esophagus - Muscularis |
| AN & HDL cholesterol (quantile) | rs59781045 | RP11-109L13.1 | chr11 | 117224567 | 11q23.3 | C | T | 8.00E-15 | 0.96 | Esophagus - Gastroesophageal Junction |
| AN & HDL cholesterol (quantile) | rs59781045 | RP11-109L13.1 | chr11 | 117224567 | 11q23.3 | C | T | 1.20E-12 | 0.85 | Stomach |
| AN & HDL cholesterol (quantile) | rs59781045 | RP11-109L13.1 | chr11 | 117224567 | 11q23.3 | C | T | 1.20E-11 | 0.86 | Colon - Sigmoid |

| | | | | | | | | | | |
|---|---|---|---|---|---|---|---|---|---|---|
| | | | | | | | | 2.20E-09 | 0.64 | Colon - Transverse |
| | | | | | | | | 6.10E-07 | 0.9 | Brain - Cerebellar Hemisphere |
| | | | | | | | | 7.80E-06 | 0.78 | Small Intestine - Terminal Ileum |
| | | | | | | | | 1.20E-05 | 0.8 | Brain - Hypothalamus |
| | | | | | | | | 1.40E-05 | 0.77 | Brain - Cortex |
| | | | | | | | | 1.50E-05 | 0.79 | Brain - Hippocampus |
| | | | | | | | | 3.60E-05 | 0.69 | Brain - Cerebellum |
| | | SIDT2 | | | | | | 3.00E-09 | -0.33 | Colon - Transverse |
| | | | | | | | | 9.40E-08 | -0.23 | Adipose - Visceral (Omentum) |
| | | | | | | | | 0.0001 | -0.15 | Adipose - Subcutaneous |
| | | | | | | | | 0.00014 | -0.19 | Thyroid |
| AN & Gamma glutamyltransferase (quantile) | rs61946383 | ACADS | chr12 | 120918272 | 12q24.31 | A | G | 2.20E-36 | 0.39 | Thyroid |
| | | SPPL3 | | | | | | 9.90E-15 | -0.24 | Adipose - Subcutaneous |
| | | | | | | | | 6.30E-14 | -0.24 | Esophagus - Muscularis |
| | | | | | | | | 4.10E-12 | -0.35 | Brain - Cortex |
| | | | | | | | | 5.70E-12 | -0.2 | Colon - Transverse |
| | | | | | | | | 1.00E-11 | -0.25 | Adipose - Visceral (Omentum) |
| | | | | | | | | 8.50E-11 | -0.31 | Colon - Sigmoid |
| | | | | | | | | 1.20E-09 | -0.35 | Brain - Cerebellum |
| | | | | | | | | 2.50E-09 | -0.33 | Brain - Caudate (basal ganglia) |
| | | | | | | | | 5.70E-09 | -0.24 | Stomach |
| | | | | | | | | 6.90E-09 | -0.23 | Esophagus - Gastroesophageal Junction |
| | | | | | | | | 1.20E-08 | -0.26 | Brain - Nucleus accumbens (basal ganglia) |
| | | | | | | | | 2.20E-06 | -0.26 | Brain - Cerebellar Hemisphere |
| | | | | | | | | 1.10E-05 | -0.27 | Brain - Putamen (basal ganglia) |
| | | UNC119B | | | | | | 1.70E-05 | 0.1 | Thyroid |
| AN & Sex hormone-binding globulin levels | rs62062620 | RP11-1099M24.6 | chr17 | 7756514 | 17p13.1 | G | A | 3.10E-05 | -0.18 | Esophagus - Mucosa |
| | | | | | | | | 3.10E-05 | -0.18 | Esophagus - Mucosa |
| AN & Body mass index (BMI)<br>AN & Whole body fat mass | rs6265 | BDNF-AS | chr11 | 27658369 | 11p14.1 | C | T | 2.50E-06 | -0.25 | Brain - Nucleus accumbens (basal ganglia) |
| | | | | | | | | 3.80E-05 | -0.24 | Brain - Frontal Cortex (BA9) |
| | | | | | | | | 0.0001 | -0.12 | Thyroid |
| AN & C-reactive protein (quantile) | rs6588156 | LEPR | chr1 | 65694750 | 1p31.3 | A | G | 7.30E-05 | -0.14 | Adipose - Visceral (Omentum) |
| AN & C-reactive protein (quantile) | rs6859 | NECTIN2 | chr19 | 44878777 | 19q13.32 | A | G | 2.40E-05 | 0.1 | Esophagus - Muscularis |

| | | | | | | | | | | |
|---|---|---|---|---|---|---|---|---|---|---|
| | | | | | | | | 4.30E-05 | 0.075 | Thyroid |
| | | | | | | | | 4.80E-05 | 0.083 | Esophagus - Mucosa |
| AN & Mean corpuscular haemoglobin | rs6899500 | ALDH8A1 | chr6 | 134905951 | 6q23.3 | T | C | 1.50E-28 | 0.5 | Esophagus - Muscularis |
| | | | | | | | | 3.80E-25 | 0.55 | Thyroid |
| | | | | | | | | 1.40E-20 | 0.56 | Colon - Sigmoid |
| | | | | | | | | 1.00E-19 | 0.5 | Adipose - Subcutaneous |
| | | | | | | | | 1.90E-18 | 0.53 | Esophagus - Gastroesophageal Junction |
| | | | | | | | | 3.00E-16 | 0.44 | Esophagus - Mucosa |
| | | | | | | | | 5.10E-16 | 0.64 | Brain - Cerebellum |
| | | | | | | | | 1.10E-13 | 0.69 | Brain - Cerebellar Hemisphere |
| | | | | | | | | 1.40E-12 | 0.39 | Stomach |
| | | | | | | | | 3.50E-12 | 0.37 | Colon - Transverse |
| | | | | | | | | 4.10E-12 | 0.38 | Adipose - Visceral (Omentum) |
| | | | | | | | | 1.50E-09 | 0.51 | Brain - Caudate (basal ganglia) |
| | | | | | | | | 2.90E-09 | 0.43 | Brain - Nucleus accumbens (basal ganglia) |
| | | | | | | | | 1.10E-08 | 0.54 | Brain - Hypothalamus |
| | | | | | | | | 1.60E-08 | 0.46 | Brain - Hippocampus |
| | | | | | | | | 3.40E-07 | 0.46 | Brain - Putamen (basal ganglia) |
| | | | | | | | | 8.00E-07 | 0.56 | Brain - Spinal cord (cervical c-1) |
| | | | | | | | | 3.30E-06 | 0.36 | Brain - Cortex |
| | | | | | | | | 4.10E-06 | 0.47 | Brain - Substantia nigra |
| | | | | | | | | 2.40E-05 | 0.37 | Small Intestine - Terminal Ileum |
| | | | | | | | | 3.60E-05 | 0.37 | Brain - Anterior cingulate cortex (BA24) |
| | | | | | | | | 4.60E-05 | 0.33 | Brain - Frontal Cortex (BA9) |
| | | CTA-212D2.2 | | | | | | 1.30E-14 | 0.46 | Esophagus - Muscularis |
| | | | | | | | | 2.80E-12 | 0.38 | Thyroid |
| | | | | | | | | 7.80E-10 | 0.63 | Brain - Caudate (basal ganglia) |
| | | | | | | | | 5.40E-09 | 0.33 | Adipose - Subcutaneous |
| | | | | | | | | 8.20E-09 | 0.65 | Brain - Putamen (basal ganglia) |
| | | | | | | | | 1.10E-08 | 0.45 | Esophagus - Gastroesophageal Junction |
| | | | | | | | | 2.30E-08 | 0.56 | Brain - Nucleus accumbens (basal ganglia) |
| | | | | | | | | 4.30E-08 | 0.32 | Adipose - Visceral (Omentum) |
| | | | | | | | | 9.00E-07 | 0.58 | Brain - Cerebellar Hemisphere |

| | | | | | | | | | | |
|---|---|---|---|---|---|---|---|---|---|---|
| | | | | | | | | 5.40E-06 | 0.47 | Brain - Cortex |
| | | | | | | | | 1.10E-05 | 0.44 | Brain - Cerebellum |
| | | | | | | | | 1.30E-05 | 0.32 | Colon - Transverse |
| | | | | | | | | 1.40E-05 | 0.32 | Colon - Sigmoid |
| | | | | | | | | 3.80E-05 | 0.48 | Brain - Hippocampus |
| | | HBS1L | | | | | | 8.60E-19 | 0.25 | Adipose - Subcutaneous |
| | | | | | | | | 1.40E-12 | 0.14 | Esophagus - Mucosa |
| | | | | | | | | 4.30E-12 | 0.2 | Thyroid |
| | | | | | | | | 2.80E-10 | 0.12 | Esophagus - Muscularis |
| | | | | | | | | 1.40E-09 | 0.17 | Colon - Sigmoid |
| | | | | | | | | 7.20E-08 | 0.14 | Colon - Transverse |
| | | | | | | | | 1.90E-07 | 0.17 | Adipose - Visceral (Omentum) |
| | | | | | | | | 2.90E-07 | 0.38 | Brain - Putamen (basal ganglia) |
| | | | | | | | | 7.80E-07 | 0.46 | Brain - Substantia nigra |
| | | | | | | | | 2.20E-06 | 0.13 | Esophagus - Gastroesophageal Junction |
| | | | | | | | | 2.20E-06 | 0.4 | Brain - Spinal cord (cervical c-1) |
| | | | | | | | | 3.70E-06 | 0.15 | Stomach |
| | | | | | | | | 4.10E-06 | 0.2 | Brain - Nucleus accumbens (basal ganglia) |
| | | | | | | | | 4.90E-06 | 0.28 | Brain - Hippocampus |
| | | | | | | | | 5.40E-06 | 0.3 | Brain - Caudate (basal ganglia) |
| | | | | | | | | 2.40E-05 | 0.38 | Brain - Amygdala |
| AN & Whole body fat mass | rs719012 | ENO4 | chr10 | 116929684 | 10q25.3 | T | C | 7.80E-07 | -0.31 | Colon - Transverse |
| | | RP11-539I5.1 | | | | | | 3.90E-09 | -0.21 | Colon - Transverse |
| | | SHTN1 | | | | | | 1.80E-07 | 0.1 | Thyroid |
| | | | | | | | | 1.50E-06 | -0.12 | Adipose - Subcutaneous |
| AN & Sex hormone-binding globulin levels | rs7687658 | HSD17B11 | chr4 | 87299357 | 4q22.1 | A | G | 3.20E-11 | 0.17 | Thyroid |
| | | | | | | | | 5.10E-08 | 0.13 | Esophagus - Muscularis |
| | | | | | | | | 2.00E-07 | 0.15 | Adipose - Visceral (Omentum) |
| | | | | | | | | 2.70E-06 | 0.13 | Adipose - Subcutaneous |
| | | | | | | | | 6.00E-06 | 0.12 | Esophagus - Gastroesophageal Junction |
| | | | | | | | | 6.40E-05 | 0.11 | Esophagus - Mucosa |
| | | HSD17B13 | | | | | | 1.00E-17 | 0.4 | Adipose - Visceral (Omentum) |
| | | | | | | | | 1.10E-13 | 0.65 | Brain - Cortex |

| | | | | | | | | | | |
|---|---|---|---|---|---|---|---|---|---|---|
| | | | | | | | | 9.90E-09 | 0.23 | Adipose - Subcutaneous |
| | | | | | | | | 3.80E-07 | 0.52 | Brain - Hypothalamus |
| | | | | | | | | 2.30E-06 | 0.4 | Brain - Cerebellum |
| | | | | | | | | 4.30E-06 | 0.42 | Brain - Cerebellar Hemisphere |
| | | | | | | | | 6.60E-06 | 0.23 | Esophagus - Muscularis |
| | | | | | | | | 3.40E-05 | 0.36 | Brain - Nucleus accumbens (basal ganglia) |
| | | | | | | | | 7.50E-05 | 0.4 | Brain - Frontal Cortex (BA9) |
| AN & HDL cholesterol (quantile) | rs7814983 | LPL | chr8 | 19781854 | 8p21.3 | C | T | 1.90E-05 | 0.32 | Brain - Anterior cingulate cortex (BA24) |
| AN & Whole body fat mass | rs7934481 | AGBL2 | chr11 | 47861785 | 11p11.2 | C | T | 3.60E-05 | -0.095 | Thyroid |
| | | C1QTNF4 | | | | | | 4.80E-17 | 0.32 | Adipose - Subcutaneous |
| | | | | | | | | 4.10E-06 | 0.19 | Colon - Transverse |
| | | | | | | | | 9.60E-06 | 0.18 | Thyroid |
| | | | | | | | | 2.00E-05 | 0.2 | Adipose - Visceral (Omentum) |
| | | | | | | | | 0.00013 | 0.18 | Esophagus - Muscularis |
| | | CELF1 | | | | | | 0.00032 | -0.084 | Thyroid |
| | | FAM180B | | | | | | 0.00018 | -0.3 | Brain - Cerebellum |
| | | MADD | | | | | | 1.90E-05 | -0.15 | Esophagus - Muscularis |
| | | NDUFS3 | | | | | | 1.40E-06 | -0.19 | Brain - Cerebellar Hemisphere |
| | | PACSIN3 | | | | | | 0.00033 | -0.11 | Esophagus - Mucosa |
| | | PTPMT1 | | | | | | 1.20E-05 | -0.13 | Adipose - Subcutaneous |
| | | PTPRJ | | | | | | 0.00043 | -0.14 | Esophagus - Muscularis |
| | | RP11-750H9.5 | | | | | | 3.00E-05 | -0.32 | Brain - Cerebellar Hemisphere |
| | | | | | | | | 0.00012 | -0.27 | Brain - Cerebellum |
| | | SLC39A13 | | | | | | 0.00048 | -0.075 | Thyroid |
| AN & Sex hormone-binding globulin levels | rs79930761 | EFNB3 | chr17 | 7912394 | 17p13.1 | C | T | 7.10E-05 | 0.32 | Adipose - Subcutaneous |
| | | KCNAB3 | | | | | | 2.40E-05 | 0.3 | Thyroid |
| | | TNFSF12 | | | | | | 8.90E-05 | 0.19 | Esophagus - Muscularis |
| AN & Sex hormone-binding globulin levels | rs80201571 | IL20RB | chr3 | 136940937 | 3q22.3 | G | T | 2.70E-06 | 0.15 | Esophagus - Mucosa |
| | | NCK1 | | | | | | 6.90E-05 | -0.16 | Esophagus - Mucosa |
| | | NCK1-AS1 | | | | | | 1.50E-13 | -0.51 | Esophagus - Mucosa |
| | | | | | | | | 5.80E-06 | -0.34 | Colon - Sigmoid |
| | | | | | | | | 0.00011 | -0.2 | Adipose - Subcutaneous |

| | | | | | | | | | | |
|---|---|---|---|---|---|---|---|---|---|---|
| | | | | | | | | 0.00014 | -0.29 | Brain - Cerebellar Hemisphere |
| | | PCCB | | | | | | 9.00E-05 | -0.12 | Esophagus - Muscularis |
| | | | | | | | | 0.00024 | -0.084 | Thyroid |
| | | RP11-731C17.2 | | | | | | 7.10E-15 | -0.68 | Brain - Hippocampus |
| | | | | | | | | 2.80E-10 | -0.48 | Brain - Cerebellar Hemisphere |
| | | | | | | | | 8.50E-10 | -0.42 | Brain - Cerebellum |
| | | | | | | | | 9.70E-09 | -0.49 | Brain - Hypothalamus |
| | | | | | | | | 1.70E-07 | -0.48 | Brain - Caudate (basal ganglia) |
| | | | | | | | | 3.20E-07 | -0.48 | Brain - Nucleus accumbens (basal ganglia) |
| | | | | | | | | 4.00E-07 | -0.53 | Brain - Frontal Cortex (BA9) |
| | | | | | | | | 5.50E-07 | -0.57 | Brain - Spinal cord (cervical c-1) |
| | | | | | | | | 8.30E-07 | -0.48 | Brain - Cortex |
| | | | | | | | | 7.50E-06 | -0.48 | Brain - Anterior cingulate cortex (BA24) |
| | | | | | | | | 1.30E-05 | -0.46 | Brain - Amygdala |
| | | | | | | | | 2.70E-05 | -0.25 | Esophagus - Muscularis |
| AN & Gamma glutamyltransferase (quantile)<br>AN & Nervous feelings<br>AN & Neuroticism score<br>AN & Red blood cell (erythrocyte) count<br>AN & Tense / 'highly strung'<br>AN & Worrier / anxious feelings<br>AN & Worry too long after embarrassment | rs8070063 | CTC1 | chr17 | 8185294 | 17p13.1 | G | T | 5.40E-06 | 0.13 | Adipose - Subcutaneous |
| | | | | | | | | 2.40E-05 | 0.17 | Colon - Transverse |
| | | KRBA2 | | | | | | 9.70E-06 | 0.18 | Esophagus - Muscularis |
| | | VAMP2 | | | | | | 1.20E-09 | 0.32 | Brain - Cerebellum |
| | | | | | | | | 1.30E-07 | 0.31 | Brain - Putamen (basal ganglia) |
| | | | | | | | | 2.40E-07 | 0.23 | Brain - Caudate (basal ganglia) |
| | | | | | | | | 1.30E-06 | 0.24 | Brain - Hypothalamus |
| | | | | | | | | 1.50E-05 | 0.16 | Brain - Hippocampus |
| | | | | | | | | 1.80E-05 | 0.13 | Brain - Frontal Cortex (BA9) |
| | | | | | | | | 2.70E-05 | 0.2 | Colon - Sigmoid |
| | | | | | | | | 5.70E-05 | 0.19 | Brain - Cerebellar Hemisphere |
| AN & Sex hormone-binding globulin levels | rs833752 | IL20RB | chr3 | 136536327 | 3q22.3 | A | G | 5.70E-06 | -0.14 | Esophagus - Mucosa |
| | | NCK1 | | | | | | 0.00036 | 0.14 | Esophagus - Mucosa |
| | | NCK1-AS1 | | | | | | 1.10E-07 | 0.35 | Esophagus - Mucosa |
| | | PCCB | | | | | | 2.90E-13 | 0.15 | Thyroid |
| | | | | | | | | 5.00E-10 | 0.18 | Esophagus - Muscularis |
| | | | | | | | | 8.20E-07 | 0.15 | Stomach |
| | | | | | | | | 1.50E-06 | 0.14 | Esophagus - Mucosa |

| | | | | | | | | | | |
|---|---|---|---|---|---|---|---|---|---|---|
| | | | | | | | | 2.80E-06 | 0.1 | Adipose - Visceral (Omentum) |
| | | | | | | | | 1.10E-05 | 0.3 | Brain - Cortex |
| | | | | | | | | 4.00E-05 | 0.17 | Esophagus - Gastroesophageal Junction |
| | | | | | | | | 0.0001 | 0.17 | Brain - Cerebellar Hemisphere |
| | | PPP2R3A | | | | | | 1.90E-05 | -0.14 | Stomach |
| | | RP11-102M11.2 | | | | | | 6.20E-07 | 0.41 | Brain - Hippocampus |
| | | RP11-731C17.2 | | | | | | 3.20E-08 | 0.5 | Brain - Hippocampus |
| | | | | | | | | 2.70E-06 | 0.36 | Brain - Cerebellar Hemisphere |

**Table S10**. Two-trait MTAG of anorexia nervosa and psychiatric traits.

| Trait1 | Trait2 | SNP | CHR | POS | A1 | A2 | MTAG | | AN | | Risk factor | | Genes | Cytogenetic band | Annotation |
|---|---|---|---|---|---|---|---|---|---|---|---|---|---|---|---|
| | | | | | | | beta | p | beta | p | beta | p | | | |
| AN | Bipolar Disorder | rs10824069 | 10 | 75853796 | T | C | 0.032 | 3.60E-08 | 0.078 | 3.80E-07 | 0.037 | 3.90E-04 | AP3M1 | 10q22.2 | new |
| AN | Nervous feelings | rs10824069 | 10 | 75853796 | C | T | -0.031 | 3.39E-08 | -0.078 | 3.80E-07 | -0.003 | 0.004 | AP3M1 | 10q22.2 | new |
| AN | Tense / 'highly strung' | rs10824069 | 10 | 75853796 | C | T | -0.032 | 1.60E-08 | -0.078 | 3.80E-07 | -0.003 | 9.75E-04 | AP3M1 | 10q22.2 | new |
| AN | Worrier / anxious feelings | rs10824069 | 10 | 75853796 | C | T | -0.032 | 1.32E-08 | -0.078 | 3.80E-07 | -0.004 | 6.42E-04 | AP3M1 | 10q22.2 | new |
| AN | Worrier / anxious feelings | rs4338470 | 10 | 76233448 | G | A | -0.031 | 2.34E-08 | -0.077 | 3.92E-07 | -0.004 | 0.002 | ADK | 10q22.2 | new |
| AN | Major Depressive Disorder | rs4281434 | 10 | 76242772 | G | A | 0.032 | 3.63E-08 | 0.078 | 3.60E-07 | 0.016 | 9.45E-04 | ADK | 10q22.2 | new |
| AN | Tense / 'highly strung' | rs4281434 | 10 | 76242772 | A | G | -0.033 | 1.00E-08 | -0.078 | 3.60E-07 | -0.004 | 3.96E-04 | ADK | 10q22.2 | new |
| AN | Bipolar Disorder | rs11001053 | 10 | 76247105 | T | C | 0.032 | 3.24E-08 | 0.078 | 3.75E-07 | 0.038 | 2.99E-04 | ADK | 10q22.2 | new |
| AN | Nervous feelings | rs11001053 | 10 | 76247105 | C | T | -0.032 | 2.89E-08 | -0.078 | 3.75E-07 | -0.003 | 0.003 | ADK | 10q22.2 | new |
| AN | Neuroticism score | rs2008387 | 10 | 131448764 | G | A | 0.032 | 2.72E-09 | 0.082 | 1.73E-08 | 0.008 | 3.86E-03 | MGMT | 10q26.3 | known |
| AN | Worrier / anxious feelings | rs2008387 | 10 | 131448764 | G | A | 0.030 | 7.25E-09 | 0.082 | 1.73E-08 | 0.003 | 0.031 | MGMT | 10q26.3 | known |
| AN | Nervous feelings | rs900144 | 11 | 13294268 | C | T | -0.028 | 4.78E-08 | -0.060 | 1.07E-05 | -0.005 | 6.53E-07 | ARNTL | 11p15.2 | new |
| AN | Neuroticism score | rs900144 | 11 | 13294268 | C | T | -0.029 | 1.41E-08 | -0.060 | 1.07E-05 | -0.017 | 7.41E-11 | ARNTL | 11p15.2 | new |
| AN | Worry too long after embarrassment | rs900144 | 11 | 13294268 | C | T | -0.032 | 8.83E-10 | -0.060 | 1.07E-05 | -0.011 | 2.26E-20 | ARNTL | 11p15.2 | new |
| AN | Guilty feelings | rs900144 | 11 | 13294268 | C | T | -0.029 | 2.15E-08 | -0.060 | 1.07E-05 | -0.006 | 5.08E-09 | ARNTL | 11p15.2 | new |
| AN | Bipolar Disorder | rs750350 | 11 | 112885342 | T | G | -0.043 | 7.06E-09 | -0.111 | 9.42E-08 | -0.051 | 1.38E-04 | NCAM1 | 11q23.2 | new |
| AN | Nervous feelings | rs750350 | 11 | 112885342 | G | T | 0.041 | 3.18E-08 | 0.111 | 9.42E-08 | 0.003 | 0.034 | NCAM1 | 11q23.2 | new |
| AN | Schizophrenia | rs750350 | 11 | 112885342 | G | T | 0.043 | 7.23E-09 | 0.111 | 9.42E-08 | -0.048 | 1.48E-04 | NCAM1 | 11q23.2 | new |
| AN | Tense / 'highly strung' | rs750350 | 11 | 112885342 | G | T | 0.041 | 2.85E-08 | 0.111 | 9.42E-08 | 0.003 | 0.032 | NCAM1 | 11q23.2 | new |
| AN | Major Depressive Disorder | rs750350 | 11 | 113014620 | G | T | 0.041 | 3.95E-08 | 0.111 | 9.42E-08 | 0.015 | 0.023 | NCAM1 | 11q23.2 | new |
| AN | Professional informed about depression | rs17649730 | 11 | 115041429 | C | T | 0.048 | 3.70E-10 | 0.121 | 5.51E-10 | 0.008 | 4.68E-02 | CADM1 | 11q23.3 | known |
| AN | Worrier / anxious feelings | rs17649730 | 11 | 115041429 | C | T | 0.049 | 8.59E-12 | 0.121 | 5.51E-10 | 0.007 | 1.16E-04 | CADM1 | 11q23.3 | known |

| | | | | | | | | | | | | | | |
|---|---|---|---|---|---|---|---|---|---|---|---|---|---|---|
| AN | Neuroticism score | rs10790068 | 11 | 115101567 | C | T | -0.042 | 2.24E-10 | -0.105 | 8.11E-10 | -0.008 | 1.45E-02 | DHFR2 | 11q23.3 | known |
| AN | Nervous feelings | rs10502199 | 11 | 115120615 | C | T | 0.048 | 1.56E-11 | 0.121 | 2.70E-10 | 0.005 | 0.001 | CADM1 | 11q23.3 | known |
| AN | Sensitivity / hurt feelings | rs10502199 | 11 | 115120615 | C | T | 0.048 | 9.14E-11 | 0.121 | 2.70E-10 | 0.004 | 0.028 | CADM1 | 11q23.3 | known |
| AN | Tense / 'highly strung' | rs10502199 | 11 | 115120615 | C | T | 0.047 | 8.67E-11 | 0.121 | 2.70E-10 | 0.003 | 0.027 | CADM1 | 11q23.3 | known |
| AN | Worry too long after embarrassment | rs10502199 | 11 | 115120615 | C | T | 0.050 | 1.75E-11 | 0.121 | 2.70E-10 | 0.006 | 0.001 | CADM1 | 11q23.3 | known |
| AN | Guilty feelings | rs10502199 | 11 | 115120615 | C | T | 0.047 | 9.51E-11 | 0.121 | 2.70E-10 | 0.003 | 3.83E-02 | CADM1 | 11q23.3 | known |
| AN | Schizophrenia | rs12826213 | 12 | 24116781 | C | T | -0.028 | 2.70E-08 | -0.069 | 6.21E-07 | 0.036 | 2.73E-05 | SOX5 | 12p12.1 | new |
| AN | Worrier / anxious feelings | rs2884594 | 12 | 4505826 | A | G | 0.033 | 3.54E-08 | 0.087 | 9.37E-07 | 0.005 | 6.74E-04 | FGF23 | 12p13.32 | new |
| AN | Worrier / anxious feelings | rs11169059 | 12 | 49924832 | A | G | 0.037 | 2.71E-08 | 0.079 | 7.42E-06 | 0.007 | 1.66E-06 | KCNH3 | 12q13.12 | new |
| AN | Nervous feelings | rs8070063 | 17 | 8088612 | G | T | 0.029 | 5.67E-09 | 0.075 | 9.73E-08 | 0.003 | 0.002 | BORCS6 | 17p13.1 | new |
| AN | Neuroticism score | rs8070063 | 17 | 8088612 | G | T | 0.029 | 7.66E-09 | 0.075 | 9.73E-08 | 0.008 | 1.07E-03 | BORCS6 | 17p13.1 | new |
| AN | Tense / 'highly strung' | rs8070063 | 17 | 8088612 | G | T | 0.031 | 1.20E-09 | 0.075 | 9.73E-08 | 0.004 | 6.81E-05 | BORCS6 | 17p13.1 | new |
| AN | Worrier / anxious feelings | rs8070063 | 17 | 8088612 | G | T | 0.029 | 3.99E-09 | 0.075 | 9.73E-08 | 0.004 | 9.18E-04 | BORCS6 | 17p13.1 | new |
| AN | Worry too long after embarrassment | rs8070063 | 17 | 8088612 | G | T | 0.030 | 1.07E-08 | 0.075 | 9.73E-08 | 0.004 | 0.003 | BORCS6 | 17p13.1 | new |
| AN | Major Depressive Disorder | rs12944041 | 17 | 8169779 | G | A | 0.028 | 4.29E-08 | 0.073 | 2.72E-07 | 0.013 | 0.003 | VAMP2 | 17p13.1 | new |
| AN | Worrier / anxious feelings | rs77540208 | 18 | 53093367 | T | C | -0.029 | 2.30E-08 | -0.058 | 4.12E-05 | -0.007 | 2.06E-09 | TCF4 | 18q21.2 | new |
| AN | Nervous feelings | rs588660 | 1 | 96886604 | G | A | -0.029 | 2.08E-08 | -0.075 | 4.95E-08 | -0.002 | 0.047 | PTBP2 | 1p21.3 | known |
| AN | Major Depressive Disorder | rs1539725 | 1 | 193153682 | C | T | -0.029 | 3.29E-08 | -0.073 | 1.26E-07 | -0.012 | 0.009 | B3GALT2 | 1q31.2 | new |
| AN | Schizophrenia | rs1539725 | 1 | 193153682 | T | C | 0.028 | 4.93E-08 | 0.073 | 1.26E-07 | -0.023 | 1.12E-02 | B3GALT2 | 1q31.2 | new |
| AN | Bipolar Disorder | rs2287348 | 2 | 54039813 | T | C | -0.042 | 2.38E-09 | -0.104 | 5.62E-09 | -0.030 | 0.024 | ERLEC1 | 2p16.2 | known |
| AN | Professional informed about anxiety | rs2287348 | 2 | 54039813 | C | T | 0.043 | 3.87E-09 | 0.104 | 5.62E-09 | 0.010 | 4.77E-02 | ERLEC1 | 2p16.2 | known |
| AN | Schizophrenia | rs2287348 | 2 | 54039813 | C | T | 0.041 | 2.70E-09 | 0.104 | 5.62E-09 | -0.029 | 1.43E-02 | ERLEC1 | 2p16.2 | known |
| AN | Schizophrenia | rs73074378 | 3 | 48446237 | C | T | 0.056 | 3.17E-09 | 0.138 | 4.72E-08 | -0.065 | 9.88E-05 | PLXNB1 | 3p21.31 | known |
| AN | Professional informed about depression | rs2633958 | 3 | 48635432 | G | A | 0.055 | 4.49E-08 | 0.141 | 7.26E-09 | -0.010 | 3.52E-02 | COL7A1 | 3p21.31 | known |
| AN | Schizophrenia | rs9812977 | 3 | 48720303 | G | A | 0.068 | 2.78E-16 | 0.165 | 8.57E-15 | -0.062 | 9.90E-06 | NCKIPSD | 3p21.31 | known |

| | | | | | | | | | | | | | | | |
|---|---|---|---|---|---|---|---|---|---|---|---|---|---|---|---|
| AN | Professional informed about anxiety | rs1048940 | 3 | 48725707 | C | T | 0.060 | 2.28E-11 | 0.156 | 3.59E-12 | -0.014 | 2.69E-02 | IP6K2 | 3p21.31 | known |
| AN | Sensitivity / hurt feelings | rs73080361 | 3 | 48832146 | G | A | 0.066 | 6.84E-14 | 0.163 | 7.31E-13 | 0.006 | 0.001 | PRKAR2A | 3p21.31 | known |
| AN | Worry too long after embarrassment | rs73080361 | 3 | 48832146 | G | A | 0.065 | 2.05E-13 | 0.163 | 7.31E-13 | 0.005 | 0.019 | PRKAR2A | 3p21.31 | known |
| AN | Schizophrenia | rs73082362 | 3 | 49113729 | C | T | 0.071 | 3.13E-15 | 0.173 | 4.27E-14 | -0.062 | 5.57E-05 | QRICH1 | 3p21.31 | known |
| AN | Sensitivity / hurt feelings | rs73082362 | 3 | 49113729 | C | T | 0.070 | 4.76E-15 | 0.173 | 4.27E-14 | 0.006 | 0.002 | QRICH1 | 3p21.31 | known |
| AN | Worry too long after embarrassment | rs73082362 | 3 | 49113729 | C | T | 0.068 | 1.35E-14 | 0.173 | 4.27E-14 | 0.005 | 0.018 | QRICH1 | 3p21.31 | known |
| AN | Loneliness, isolation | rs4513485 | 3 | 49139966 | G | A | -0.032 | 1.72E-08 | -0.077 | 4.67E-08 | -0.002 | 1.08E-02 | QARS1 | 3p21.31 | known |
| AN | Tense / 'highly strung' | rs4513485 | 3 | 49139966 | G | A | -0.031 | 6.63E-09 | -0.077 | 4.67E-08 | -0.002 | 0.011 | QARS1 | 3p21.31 | known |
| AN | Major Depressive Disorder | rs1134043 | 3 | 49162704 | G | T | 0.043 | 1.13E-09 | 0.108 | 4.83E-09 | 0.016 | 0.007 | CCDC71 | 3p21.31 | known |
| AN | Neuroticism score | rs1134043 | 3 | 49200137 | G | T | 0.043 | 8.02E-10 | 0.108 | 4.83E-09 | 0.010 | 5.83E-03 | CCDC71 | 3p21.31 | known |
| AN | Schizophrenia | rs11716445 | 3 | 49406095 | G | A | 0.069 | 1.86E-14 | 0.165 | 3.10E-13 | -0.063 | 4.06E-05 | GPX1 | 3p21.31 | known |
| AN | Worry too long after embarrassment | rs11716445 | 3 | 49406095 | G | A | 0.066 | 8.48E-14 | 0.165 | 3.10E-13 | 0.005 | 0.015 | GPX1 | 3p21.31 | known |
| AN | Professional informed about depression | rs183270497 | 3 | 49479704 | C | T | 0.061 | 1.49E-11 | 0.171 | 1.32E-12 | -0.009 | 4.12E-02 | NICN1 | 3p21.31 | known |
| AN | Sensitivity / hurt feelings | rs73073015 | 3 | 49503166 | G | A | 0.068 | 2.59E-14 | 0.173 | 3.24E-13 | 0.007 | 8.82E-04 | DAG1 | 3p21.31 | known |
| AN | Professional informed about depression | rs55662945 | 3 | 49732699 | C | T | 0.055 | 6.71E-09 | 0.148 | 7.72E-10 | -0.011 | 1.44E-02 | RNF123 | 3p21.31 | known |
| AN | Schizophrenia | rs73077200 | 3 | 49826987 | C | T | 0.069 | 1.43E-14 | 0.169 | 2.77E-13 | -0.063 | 4.29E-05 | IP6K1 | 3p21.31 | known |
| AN | Sensitivity / hurt feelings | rs73077200 | 3 | 49826987 | C | T | 0.068 | 2.00E-14 | 0.169 | 2.77E-13 | 0.007 | 0.001 | IP6K1 | 3p21.31 | known |
| AN | Worry too long after embarrassment | rs73077200 | 3 | 49826987 | C | T | 0.066 | 6.22E-14 | 0.169 | 2.77E-13 | 0.005 | 0.015 | IP6K1 | 3p21.31 | known |
| AN | Guilty feelings | rs4688759 | 3 | 50008118 | T | C | 0.048 | 7.31E-09 | 0.146 | 3.17E-11 | -0.003 | 4.83E-02 | RBM6 | 3p21.31 | known |
| AN | Schizophrenia | rs73080977 | 3 | 50189531 | G | T | 0.070 | 4.15E-14 | 0.175 | 1.31E-12 | -0.070 | 1.23E-05 | SEMA3F | 3p21.31 | known |
| AN | Sensitivity / hurt feelings | rs73080977 | 3 | 50189531 | G | T | 0.068 | 2.43E-13 | 0.175 | 1.31E-12 | 0.006 | 0.008 | SEMA3F | 3p21.31 | known |
| AN | Worry too long after embarrassment | rs73080977 | 3 | 50189531 | G | T | 0.067 | 2.29E-13 | 0.175 | 1.31E-12 | 0.006 | 0.009 | SEMA3F | 3p21.31 | known |
| AN | Bipolar Disorder | rs7617915 | 3 | 50239012 | C | T | 0.051 | 3.50E-08 | 0.135 | 6.33E-08 | 0.034 | 0.049 | SLC38A3 | 3p21.31 | known |
| AN | Tense / 'highly strung' | rs6789500 | 3 | 20764696 | T | C | -0.035 | 4.51E-08 | -0.091 | 2.55E-07 | -0.003 | 0.016 | SGO1 | 3p24.3 | new |
| AN | Worrier / anxious feelings | rs6789500 | 3 | 20764696 | T | C | -0.035 | 3.21E-08 | -0.091 | 2.55E-07 | -0.004 | 0.007 | SGO1 | 3p24.3 | new |

| | | | | | | | | | | | | | | | |
|---|---|---|---|---|---|---|---|---|---|---|---|---|---|---|---|
| AN | Loneliness, isolation | rs13125932 | 4 | 58150175 | C | T | 0.029 | 2.77E-08 | 0.072 | 5.85E-08 | 0.002 | 1.65E-02 | IGFBP7 | 4q12 | new |
| AN | Major Depressive Disorder | rs13125932 | 4 | 58150175 | C | T | 0.028 | 2.89E-08 | 0.072 | 5.85E-08 | 0.009 | 0.028 | IGFBP7 | 4q12 | new |
| AN | Sensitivity / hurt feelings | rs13125932 | 4 | 58150175 | C | T | 0.029 | 2.20E-08 | 0.072 | 5.85E-08 | 0.002 | 0.033 | IGFBP7 | 4q12 | new |
| AN | Tense / 'highly strung' | rs13125932 | 4 | 58150175 | C | T | 0.028 | 1.95E-08 | 0.072 | 5.85E-08 | 0.002 | 0.034 | IGFBP7 | 4q12 | new |
| AN | Sensitivity / hurt feelings | rs6872919 | 5 | 25086440 | A | C | 0.031 | 3.16E-09 | 0.074 | 4.05E-08 | 0.004 | 8.57E-04 | CDH10 | 5p14.1 | known |
| AN | Worry too long after embarrassment | rs6872919 | 5 | 25086440 | A | C | 0.030 | 4.62E-09 | 0.074 | 4.05E-08 | 0.003 | 0.004 | CDH10 | 5p14.1 | known |
| AN | Schizophrenia | rs8192589 | 6 | 32187637 | G | T | -0.048 | 1.15E-10 | -0.089 | 2.50E-05 | 0.152 | 2.43E-29 | MHC region | 6p21.32 | |
| AN | Schizophrenia | rs9273463 | 6 | 32627934 | G | A | -0.044 | 4.53E-08 | -0.080 | 4.85E-04 | 0.137 | 3.08E-18 | MHC region | 6p21.32 | |
| AN | Nervous feelings | rs9274678 | 6 | 32636646 | A | G | -0.045 | 1.81E-08 | -0.098 | 8.27E-05 | -0.009 | 1.26E-10 | MHC region | 6p21.32 | |
| AN | Schizophrenia | rs75301392 | 6 | 32904051 | C | T | -0.057 | 1.68E-09 | -0.107 | 2.89E-05 | 0.163 | 1.45E-21 | MHC region | 6p21.32 | |
| AN | Bipolar Disorder | rs3132604 | 6 | 30741226 | T | C | 0.040 | 3.28E-08 | 0.086 | 1.71E-06 | 0.066 | 2.17E-07 | MHC region | 6p21.33 | |
| AN | Nervous feelings | rs3132604 | 6 | 30741226 | C | T | -0.035 | 2.15E-08 | -0.086 | 1.71E-06 | -0.005 | 2.74E-05 | MHC region | 6p21.33 | |
| AN | Schizophrenia | rs3132604 | 6 | 30741226 | C | T | -0.044 | 5.52E-11 | -0.086 | 1.71E-06 | 0.113 | 6.02E-21 | MHC region | 6p21.33 | |
| AN | Tense / 'highly strung' | rs3132604 | 6 | 30741226 | C | T | -0.036 | 6.71E-09 | -0.086 | 1.71E-06 | -0.005 | 1.72E-06 | MHC region | 6p21.33 | |
| AN | Guilty feelings | rs3132604 | 6 | 30741226 | C | T | -0.035 | 2.76E-08 | -0.086 | 1.71E-06 | -0.006 | 2.66E-05 | MHC region | 6p21.33 | |
| AN | Schizophrenia | rs3132510 | 6 | 31172151 | T | C | -0.049 | 2.24E-09 | -0.084 | 9.96E-05 | 0.155 | 9.50E-26 | MHC region | 6p21.33 | |
| AN | Schizophrenia | rs3130923 | 6 | 31462135 | G | A | -0.050 | 1.91E-10 | -0.092 | 1.25E-05 | 0.144 | 8.36E-25 | MHC region | 6p21.33 | |
| AN | Major Depressive Disorder | rs3130063 | 6 | 31590925 | C | T | -0.043 | 1.59E-08 | -0.094 | 9.83E-06 | -0.043 | 3.98E-11 | MHC region | 6p21.33 | |
| AN | Nervous feelings | rs3117581 | 6 | 31624864 | A | G | -0.041 | 1.65E-08 | -0.092 | 1.99E-05 | -0.009 | 9.76E-10 | MHC region | 6p21.33 | |
| AN | Schizophrenia | rs67682613 | 6 | 31826705 | G | A | -0.050 | 6.92E-12 | -0.093 | 1.81E-06 | 0.138 | 2.57E-27 | MHC region | 6p21.33 | |
| AN | Bipolar Disorder | rs532086 | 6 | 31881309 | C | T | 0.043 | 1.21E-08 | 0.090 | 2.83E-06 | 0.092 | 3.83E-11 | MHC region | 6p21.33 | |
| AN | Nervous feelings | rs532086 | 6 | 31881309 | T | C | -0.041 | 1.93E-09 | -0.090 | 2.83E-06 | -0.008 | 2.03E-09 | MHC region | 6p21.33 | |
| AN | Neuroticism score | rs532086 | 6 | 31881309 | T | C | -0.038 | 3.20E-08 | -0.090 | 2.83E-06 | -0.017 | 1.56E-06 | MHC region | 6p21.33 | |
| AN | Tense / 'highly strung' | rs532086 | 6 | 31881309 | T | C | -0.040 | 2.90E-09 | -0.090 | 2.83E-06 | -0.007 | 2.00E-08 | MHC region | 6p21.33 | |
| AN | Worrier / anxious feelings | rs532086 | 6 | 31881309 | T | C | -0.039 | 5.93E-09 | -0.090 | 2.83E-06 | -0.008 | 4.68E-07 | MHC region | 6p21.33 | |

| | | | | | | | | | | | | | | | |
|---|---|---|---|---|---|---|---|---|---|---|---|---|---|---|---|
| AN | Schizophrenia | rs17720293 | 6 | 28214698 | C | T | -0.047 | 8.46E-09 | -0.071 | 8.84E-04 | 0.173 | 1.06E-32 | MHC region | 6p22.1 | |
| AN | Schizophrenia | rs385816 | 6 | 29480224 | G | A | -0.045 | 4.52E-08 | -0.071 | 1.30E-03 | 0.166 | 2.08E-28 | MHC region | 6p22.1 | |
| AN | Schizophrenia | rs1117488 | 6 | 30170906 | G | T | -0.043 | 2.28E-11 | -0.081 | 2.20E-06 | 0.118 | 1.05E-24 | MHC region | 6p22.1 | |
| AN | Nervous feelings | rs971570 | 6 | 30172513 | A | C | -0.033 | 2.39E-08 | -0.081 | 2.24E-06 | -0.005 | 1.60E-05 | MHC region | 6p22.1 | |
| AN | Tense / 'highly strung' | rs971570 | 6 | 30172513 | A | C | -0.033 | 3.79E-08 | -0.081 | 2.24E-06 | -0.004 | 8.61E-05 | MHC region | 6p22.1 | |
| AN | Guilty feelings | rs971570 | 6 | 30172513 | A | C | -0.034 | 1.93E-08 | -0.081 | 2.24E-06 | -0.006 | 3.11E-06 | MHC region | 6p22.1 | |
| AN | Bipolar Disorder | rs1487445 | 6 | 98565211 | T | C | -0.029 | 3.12E-08 | -0.056 | 2.70E-05 | -0.074 | 1.48E-15 | POU3F2 | 6q16.1 | new |
| AN | Nervous feelings | rs4129585 | 8 | 143312933 | A | C | -0.029 | 6.84E-09 | -0.059 | 9.42E-06 | -0.006 | 9.44E-10 | TSNARE1 | 8q24.3 | new |
| AN | Schizophrenia | rs4129585 | 8 | 143312933 | A | C | -0.031 | 1.63E-09 | -0.059 | 9.42E-06 | 0.075 | 5.11E-18 | TSNARE1 | 8q24.3 | new |
| AN | Tense / 'highly strung' | rs4129585 | 8 | 143312933 | A | C | -0.029 | 6.74E-09 | -0.059 | 9.42E-06 | -0.005 | 2.30E-09 | TSNARE1 | 8q24.3 | new |
| AN | Worrier / anxious feelings | rs13262595 | 8 | 143316970 | A | G | -0.028 | 1.62E-08 | -0.059 | 1.01E-05 | -0.006 | 1.27E-07 | TSNARE1 | 8q24.3 | new |

MTAG section of the table presents the MTAG results specific to Trait 1. Trait 1, anorexia nervosa.

**Table S11.** Two-trait MTAG of anorexia nervosa and metabolic traits.

| Trait1 | Trait2 | SNP | CHR | POS | A1 | A2 | MTAG | | AN | | Risk factor | | Genes | Cytogenetic band | Annotation |
|---|---|---|---|---|---|---|---|---|---|---|---|---|---|---|---|
| | | | | | | | beta | p | beta | p | beta | p | | | |
| AN | Body mass index (BMI) | rs12411523 | 10 | 99862731 | C | T | 0.027 | 4.72E-08 | 0.051 | 4.06E-04 | -0.019 | 1.30E-14 | R3HCC1L | 10q24.2 | new |
| AN | Whole body fat mass | rs719012 | 10 | 118689195 | T | C | 0.029 | 3.01E-08 | 0.060 | 1.56E-04 | -0.019 | 3.75E-12 | HSPA12A | 10q25.3 | new |
| AN | Basal metabolic rate | rs11245450 | 10 | 126658075 | G | A | 0.029 | 4.57E-08 | 0.065 | 7.92E-06 | -0.010 | 4.54E-11 | ZRANB1 | 10q26.13 | new |
| AN | Body mass index (BMI) | rs4962725 | 10 | 126733321 | T | C | -0.026 | 1.93E-08 | -0.061 | 1.37E-05 | 0.014 | 2.08E-09 | ZRANB1 | 10q26.13 | new |
| AN | Whole body fat mass | rs4962725 | 10 | 126733321 | T | C | -0.026 | 1.66E-08 | -0.061 | 1.37E-05 | 0.013 | 5.85E-09 | ZRANB1 | 10q26.13 | new |
| AN | Fasting insulin | rs12251589 | 10 | 131417354 | C | A | 0.030 | 4.06E-08 | 0.078 | 4.31E-08 | -0.002 | 0.047 | MGMT | 10q26.3 | known |
| AN | Basal metabolic rate | rs2008387 | 10 | 131448764 | G | A | 0.031 | 4.12E-09 | 0.082 | 1.73E-08 | -0.005 | 9.41E-04 | MGMT | 10q26.3 | known |
| AN | Body mass index (BMI) | rs2008387 | 10 | 131448764 | G | A | 0.027 | 4.94E-08 | 0.082 | 1.73E-08 | -0.005 | 4.47E-02 | MGMT | 10q26.3 | known |
| AN | Whole body fat mass | rs2008387 | 10 | 131448764 | G | A | 0.029 | 1.16E-09 | 0.082 | 1.73E-08 | -0.010 | 9.11E-05 | MGMT | 10q26.3 | known |
| AN | Body mass index (BMI) | rs10769282 | 11 | 47626492 | T | G | 0.029 | 6.23E-09 | 0.055 | 2.39E-04 | -0.021 | 2.88E-17 | IFRD2 | 11p11.2 | known |
| AN | Whole body fat mass | rs10769282 | 11 | 47626492 | T | G | 0.030 | 1.09E-09 | 0.055 | 2.39E-04 | -0.022 | 5.66E-19 | IFRD2 | 11p11.2 | known |
| AN | Whole body fat mass | rs7934481 | 11 | 47883337 | C | T | 0.028 | 2.30E-08 | 0.049 | 8.23E-04 | -0.021 | 2.20E-16 | NUP160 | 11p11.2 | known |
| AN | Body mass index (BMI) | rs11030088 | 11 | 27646247 | G | A | -0.035 | 5.04E-11 | -0.051 | 1.17E-03 | 0.032 | 2.11E-33 | BDNF | 11p14.1 | new |
| AN | Whole body fat mass | rs11030088 | 11 | 27646247 | G | A | -0.035 | 1.54E-11 | -0.051 | 1.17E-03 | 0.032 | 7.57E-33 | BDNF | 11p14.1 | new |
| AN | Body mass index (BMI) | rs6265 | 11 | 27679916 | C | T | 0.037 | 3.67E-10 | 0.039 | 2.07E-02 | -0.041 | 3.54E-42 | BDNF | 11p14.1 | new |
| AN | Whole body fat mass | rs6265 | 11 | 27679916 | C | T | 0.034 | 7.69E-09 | 0.039 | 2.07E-02 | -0.033 | 4.25E-30 | BDNF | 11p14.1 | new |
| AN | Body mass index (BMI) | rs900144 | 11 | 13294268 | C | T | -0.028 | 2.63E-09 | -0.060 | 1.07E-05 | 0.016 | 4.95E-12 | ARNTL | 11p15.2 | new |
| AN | Whole body fat mass | rs900144 | 11 | 13294268 | C | T | -0.026 | 1.42E-08 | -0.060 | 1.07E-05 | 0.013 | 8.20E-09 | ARNTL | 11p15.2 | new |
| AN | HDL cholesterol (quantile) | rs2072113 | 11 | 61604967 | C | T | -0.047 | 2.79E-09 | -0.050 | 1.31E-02 | -0.061 | 1.72E-66 | FADS1 | 11q12.2 | new |
| AN | Body mass index (BMI) | rs10502199 | 11 | 115120615 | C | T | 0.043 | 4.56E-11 | 0.121 | 2.70E-10 | -0.012 | 2.26E-04 | CADM1 | 11q23.3 | known |
| AN | HDL cholesterol (quantile) | rs10502199 | 11 | 115120615 | C | T | 0.046 | 1.14E-10 | 0.121 | 2.70E-10 | 0.008 | 1.19E-02 | CADM1 | 11q23.3 | known |
| AN | Whole body fat mass | rs10502199 | 11 | 115120615 | C | T | 0.042 | 9.68E-11 | 0.121 | 2.70E-10 | -0.011 | 1.03E-03 | CADM1 | 11q23.3 | known |

| | | | | | | | | | | | | | | |
|---|---|---|---|---|---|---|---|---|---|---|---|---|---|---|
| AN | Triglycerides (quantile) | rs17649730 | 11 | 115041429 | C | T | 0.047 | 3.91E-10 | 0.121 | 5.51E-10 | -0.007 | 4.52E-02 | CADM1 | 11q23.3 | known |
| AN | Basal metabolic rate | rs45456599 | 11 | 115066675 | T | C | 0.042 | 1.18E-09 | 0.108 | 2.45E-09 | -0.006 | 7.79E-03 | CADM1 | 11q23.3 | known |
| AN | Triglycerides (quantile) | rs486394 | 11 | 116526322 | A | C | -0.035 | 6.75E-10 | -0.034 | 2.80E-02 | 0.072 | 1.53E-170 | BUD13 | 11q23.3 | new |
| AN | Body mass index (BMI) | rs4938181 | 11 | 115041163 | A | G | -0.027 | 3.90E-09 | -0.061 | 5.86E-06 | 0.015 | 1.65E-10 | CADM1 | 11q23.3 | known |
| AN | HDL cholesterol (quantile) | rs59781045 | 11 | 117095283 | C | T | 0.057 | 1.63E-08 | 0.058 | 3.04E-02 | 0.078 | 1.96E-66 | PCSK7 | 11q23.3 | new |
| AN | Basal metabolic rate | rs705696 | 12 | 56480648 | G | A | 0.029 | 3.88E-08 | 0.064 | 7.67E-06 | -0.011 | 1.35E-11 | ERBB3 | 12q13.2 | known |
| AN | Body mass index (BMI) | rs705696 | 12 | 56480648 | G | A | 0.030 | 1.09E-09 | 0.064 | 7.67E-06 | -0.018 | 7.66E-13 | ERBB3 | 12q13.2 | known |
| AN | Whole body fat mass | rs705696 | 12 | 56480648 | G | A | 0.027 | 1.98E-08 | 0.064 | 7.67E-06 | -0.013 | 6.28E-08 | ERBB3 | 12q13.2 | known |
| AN | HDL cholesterol (quantile) | rs3809114 | 12 | 57848639 | G | A | 0.030 | 3.17E-09 | 0.053 | 2.18E-04 | 0.027 | 1.39E-31 | INHBE | 12q13.3 | new |
| AN | Body mass index (BMI) | rs1869893 | 12 | 62262577 | T | G | -0.031 | 1.51E-08 | -0.079 | 1.38E-06 | 0.013 | 1.56E-06 | USP15 | 12q14.1 | new |
| AN | Whole body fat mass | rs1869893 | 12 | 62262577 | T | G | -0.030 | 4.40E-08 | -0.079 | 1.38E-06 | 0.011 | 4.51E-05 | USP15 | 12q14.1 | new |
| AN | Basal metabolic rate | rs7968902 | 12 | 66363070 | T | G | 0.031 | 1.40E-09 | 0.043 | 1.79E-03 | -0.025 | 1.15E-57 | HMGA2 | 12q14.3 | new |
| AN | Body mass index (BMI) | rs12583517 | 13 | 58663457 | G | T | 0.034 | 7.02E-10 | 0.066 | 3.87E-05 | -0.023 | 6.30E-17 | PCDH17 | 13q21.1 | new |
| AN | Whole body fat mass | rs12583517 | 13 | 58663457 | G | T | 0.030 | 3.79E-08 | 0.066 | 3.87E-05 | -0.016 | 2.24E-09 | PCDH17 | 13q21.1 | new |
| AN | Basal metabolic rate | rs9538138 | 13 | 59174424 | A | G | -0.032 | 4.53E-08 | -0.077 | 3.54E-06 | 0.011 | 6.23E-09 | PCDH17 | 13q21.1 | new |
| AN | Body mass index (BMI) | rs9538138 | 13 | 59174424 | A | G | -0.032 | 7.19E-09 | -0.077 | 3.54E-06 | 0.016 | 1.11E-08 | PCDH17 | 13q21.1 | new |
| AN | Whole body fat mass | rs9538138 | 13 | 59174424 | A | G | -0.030 | 2.32E-08 | -0.077 | 3.54E-06 | 0.013 | 1.01E-06 | PCDH17 | 13q21.1 | new |
| AN | HDL cholesterol (quantile) | rs62000865 | 15 | 58698662 | G | A | 0.069 | 1.00E-11 | 0.075 | 1.39E-02 | 0.092 | 3.45E-93 | LIPC | 15q21.3 | new |
| AN | HDL cholesterol (quantile) | rs6494005 | 15 | 58724524 | A | G | -0.036 | 1.18E-12 | -0.031 | 2.17E-02 | -0.054 | 3.17E-123 | LIPC | 15q21.3 | new |
| AN | Body mass index (BMI) | rs2466826 | 16 | 28348344 | G | A | -0.026 | 4.07E-08 | -0.035 | 2.17E-02 | 0.026 | 2.73E-28 | NPIPB6 | 16p11.2 | new |
| AN | Whole body fat mass | rs2466826 | 16 | 28348344 | G | A | -0.029 | 5.82E-10 | -0.035 | 2.17E-02 | 0.029 | 5.17E-36 | NPIPB6 | 16p11.2 | new |
| AN | Body mass index (BMI) | rs3927084 | 16 | 28639047 | A | C | -0.027 | 1.73E-08 | -0.035 | 1.82E-02 | 0.027 | 2.40E-29 | NPIPB8 | 16p11.2 | new |
| AN | Whole body fat mass | rs3927084 | 16 | 28639047 | A | C | -0.031 | 1.07E-10 | -0.035 | 1.82E-02 | 0.030 | 6.03E-39 | NPIPB8 | 16p11.2 | new |
| AN | Body mass index (BMI) | rs72805611 | 16 | 53831354 | C | T | -0.034 | 7.52E-13 | 0.028 | 3.88E-02 | 0.067 | 3.79E-177 | RPGRIP1L | 16q12.2 | new |
| AN | Whole body fat mass | rs72805611 | 16 | 53831354 | C | T | -0.029 | 2.23E-10 | 0.028 | 3.88E-02 | 0.056 | 1.48E-129 | RPGRIP1L | 16q12.2 | new |

| | | | | | | | | | | | | | | | |
|---|---|---|---|---|---|---|---|---|---|---|---|---|---|---|---|
| AN | HDL cholesterol (quantile) | rs59542880 | 16 | 56848871 | G | A | -0.074 | 1.79E-17 | -0.056 | 2.27E-02 | -0.119 | 2.22E-201 | SLC12A3 | 16q13 | new |
| AN | Basal metabolic rate | rs11165643 | 1 | 96924097 | C | T | -0.032 | 3.13E-10 | -0.075 | 3.47E-08 | 0.009 | 6.13E-09 | PTBP2 | 1p21.3 | known |
| AN | Body mass index (BMI) | rs11165643 | 1 | 96924097 | C | T | -0.034 | 3.95E-13 | -0.075 | 3.47E-08 | 0.019 | 1.53E-15 | PTBP2 | 1p21.3 | known |
| AN | HDL cholesterol (quantile) | rs11165643 | 1 | 96924097 | C | T | -0.029 | 1.86E-08 | -0.075 | 3.47E-08 | -0.005 | 3.44E-02 | PTBP2 | 1p21.3 | known |
| AN | Whole body fat mass | rs11165643 | 1 | 96924097 | C | T | -0.035 | 9.25E-14 | -0.075 | 3.47E-08 | 0.019 | 2.07E-16 | PTBP2 | 1p21.3 | known |
| AN | Body mass index (BMI) | rs17426874 | 1 | 97063163 | A | C | 0.053 | 4.17E-08 | 0.135 | 3.39E-06 | -0.023 | 2.14E-06 | PTBP2 | 1p21.3 | known |
| AN | Body mass index (BMI) | rs273882 | 1 | 97276688 | T | C | -0.027 | 4.69E-08 | -0.062 | 3.13E-05 | 0.015 | 2.07E-09 | PTBP2 | 1p21.3 | known |
| AN | HDL cholesterol (quantile) | rs3768321 | 1 | 40035928 | G | T | -0.035 | 4.46E-08 | -0.039 | 2.15E-02 | -0.045 | 3.08E-56 | PABPC4 | 1p34.3 | new |
| AN | Basal metabolic rate | rs4751 | 1 | 1686040 | G | T | 0.035 | 5.22E-09 | 0.082 | 1.16E-06 | -0.010 | 6.08E-10 | NADK | 1p36.33 | new |
| AN | Body mass index (BMI) | rs4751 | 1 | 1686040 | G | T | 0.032 | 4.47E-09 | 0.082 | 1.16E-06 | -0.013 | 1.01E-07 | NADK | 1p36.33 | new |
| AN | Body mass index (BMI) | rs10788797 | 1 | 151003071 | G | A | 0.041 | 1.41E-08 | 0.090 | 6.22E-06 | -0.021 | 9.75E-09 | BNIPL | 1q21.3 | new |
| AN | Whole body fat mass | rs10788797 | 1 | 151003071 | G | A | 0.039 | 2.67E-08 | 0.090 | 6.22E-06 | -0.018 | 2.27E-07 | BNIPL | 1q21.3 | new |
| AN | HDL cholesterol (quantile) | rs612577 | 1 | 230324364 | T | C | -0.047 | 6.24E-11 | -0.047 | 2.04E-02 | -0.065 | 1.43E-91 | GALNT2 | 1q42.13 | new |
| AN | Basal metabolic rate | rs34298551 | 20 | 34030606 | G | A | -0.034 | 3.74E-08 | -0.043 | 1.54E-02 | 0.031 | 1.17E-60 | GDF5 | 20q11.22 | new |
| AN | Body mass index (BMI) | rs394608 | 21 | 46581798 | T | C | -0.027 | 9.23E-09 | -0.056 | 5.82E-05 | 0.017 | 3.15E-13 | ADARB1 | 21q22.3 | new |
| AN | Whole body fat mass | rs394608 | 21 | 46581798 | T | C | -0.026 | 1.22E-08 | -0.056 | 5.82E-05 | 0.016 | 9.52E-12 | ADARB1 | 21q22.3 | new |
| AN | Body mass index (BMI) | rs4671328 | 2 | 58935282 | T | G | 0.027 | 6.41E-09 | 0.046 | 7.18E-04 | -0.022 | 4.20E-20 | FANCL | 2p16.1 | new |
| AN | Whole body fat mass | rs4671328 | 2 | 58935282 | T | G | 0.026 | 7.59E-09 | 0.046 | 7.18E-04 | -0.020 | 5.52E-18 | FANCL | 2p16.1 | new |
| AN | Body mass index (BMI) | rs12620667 | 2 | 54290997 | G | A | 0.044 | 4.91E-09 | 0.108 | 2.31E-07 | -0.017 | 7.20E-06 | PSME4 | 2p16.2 | known |
| AN | HDL cholesterol (quantile) | rs12620667 | 2 | 54290997 | G | A | 0.045 | 4.19E-08 | 0.108 | 2.31E-07 | 0.011 | 2.31E-03 | PSME4 | 2p16.2 | known |
| AN | Whole body fat mass | rs12620667 | 2 | 54290997 | G | A | 0.041 | 4.75E-08 | 0.108 | 2.31E-07 | -0.012 | 1.47E-03 | PSME4 | 2p16.2 | known |
| AN | Body mass index (BMI) | rs2287348 | 2 | 54039813 | C | T | 0.039 | 1.60E-09 | 0.104 | 5.62E-09 | -0.011 | 1.06E-03 | ERLEC1 | 2p16.2 | known |
| AN | Fasting insulin | rs2287348 | 2 | 54039813 | C | T | 0.040 | 3.02E-09 | 0.104 | 5.62E-09 | -0.005 | 0.047 | ERLEC1 | 2p16.2 | known |
| AN | HDL cholesterol (quantile) | rs2287348 | 2 | 54039813 | C | T | 0.042 | 2.58E-09 | 0.104 | 5.62E-09 | 0.007 | 1.82E-02 | ERLEC1 | 2p16.2 | known |
| AN | Whole body fat mass | rs2287348 | 2 | 54039813 | C | T | 0.036 | 9.65E-09 | 0.104 | 5.62E-09 | -0.007 | 1.98E-02 | ERLEC1 | 2p16.2 | known |

| | | | | | | | | | | | | | | |
|---|---|---|---|---|---|---|---|---|---|---|---|---|---|---|
| AN | Body mass index (BMI) | rs11689201 | 2 | 100820431 | A | G | 0.027 | 1.15E-08 | 0.052 | 1.33E-04 | -0.018 | 1.78E-14 | AFF3 | 2q11.2 | new |
| AN | Whole body fat mass | rs11689201 | 2 | 100820431 | A | G | 0.027 | 8.21E-09 | 0.052 | 1.33E-04 | -0.018 | 7.36E-14 | AFF3 | 2q11.2 | new |
| AN | Whole body fat mass | rs55893301 | 2 | 172909764 | A | C | -0.028 | 6.01E-09 | -0.075 | 2.92E-06 | 0.014 | 4.61E-08 | DLX1 | 2q31.1 | new |
| AN | Body mass index (BMI) | rs62183827 | 2 | 172908951 | A | G | -0.027 | 4.07E-08 | -0.072 | 4.70E-06 | 0.013 | 8.26E-07 | DLX1 | 2q31.1 | new |
| AN | T2D_2018 | rs1915304 | 3 | 89655243 | C | T | -0.035 | 4.26E-08 | -0.095 | 3.14E-07 | 0.068 | 1.54E-04 | EPHA3 | 3p11.1 | new |
| AN | Body mass index (BMI) | rs1915304 | 3 | 89704393 | C | T | -0.030 | 2.25E-08 | -0.095 | 3.14E-07 | 0.010 | 1.20E-04 | EPHA3 | 3p11.1 | new |
| AN | Whole body fat mass | rs1979837 | 3 | 89590902 | T | C | 0.032 | 2.75E-09 | 0.083 | 3.56E-06 | -0.014 | 5.71E-09 | EPHA3 | 3p11.1 | new |
| AN | Body mass index (BMI) | rs55931912 | 3 | 90038054 | G | A | -0.031 | 1.16E-08 | -0.087 | 2.84E-06 | 0.014 | 1.19E-07 | EPHA3 | 3p11.1 | new |
| AN | Whole body fat mass | rs55931912 | 3 | 90038054 | G | A | -0.032 | 2.31E-09 | -0.087 | 2.84E-06 | 0.015 | 4.13E-09 | EPHA3 | 3p11.1 | new |
| AN | Fasting blood insulin | rs9847154 | 3 | 70994006 | C | T | -0.031 | 1.16E-08 | -0.079 | 2.75E-08 | 0.009 | 0.021 | MDFIC2 | 3p13 | known |
| AN | Fasting insulin | rs9874207 | 3 | 71019750 | C | T | -0.030 | 7.36E-09 | -0.081 | 2.05E-08 | 0.005 | 0.034 | FOXP1 | 3p13 | known |
| AN | T2D_2018 | rs11709092 | 3 | 48849594 | C | A | -0.063 | 5.65E-12 | -0.165 | 4.55E-13 | -0.054 | 0.048 | PRKAR2A | 3p21.31 | known |
| AN | Body mass index (BMI) | rs11710277 | 3 | 50199747 | A | G | 0.050 | 6.06E-09 | 0.156 | 1.72E-09 | -0.009 | 3.93E-02 | SEMA3F | 3p21.31 | known |
| AN | Triglycerides (quantile) | rs11712982 | 3 | 48938762 | C | T | 0.041 | 9.11E-09 | 0.119 | 3.56E-10 | 0.007 | 3.54E-02 | SLC25A20 | 3p21.31 | known |
| AN | Body mass index (BMI) | rs11716445 | 3 | 49406095 | G | A | 0.057 | 9.73E-13 | 0.165 | 3.10E-13 | -0.011 | 4.78E-03 | GPX1 | 3p21.31 | known |
| AN | Whole body fat mass | rs11716445 | 3 | 49406095 | G | A | 0.059 | 3.86E-14 | 0.165 | 3.10E-13 | -0.016 | 3.85E-05 | GPX1 | 3p21.31 | known |
| AN | Whole body fat mass | rs11716961 | 3 | 49908338 | G | A | 0.059 | 2.51E-14 | 0.169 | 3.78E-13 | -0.017 | 1.69E-05 | CAMKV | 3p21.31 | known |
| AN | Fasting insulin | rs12493578 | 3 | 48692683 | C | T | 0.030 | 3.51E-08 | 0.076 | 7.77E-08 | -0.004 | 0.044 | NCKIPSD | 3p21.31 | known |
| AN | Basal metabolic rate | rs13090013 | 3 | 48742578 | G | A | 0.053 | 3.73E-12 | 0.140 | 1.72E-12 | -0.005 | 5.00E-02 | NCKIPSD | 3p21.31 | known |
| AN | HDL cholesterol (quantile) | rs199956414 | 3 | 50022089 | G | A | -0.031 | 7.03E-10 | -0.058 | 2.07E-05 | -0.024 | 3.37E-25 | RBM6 | 3p21.31 | known |
| AN | Whole body fat mass | rs2008877 | 3 | 50162291 | T | G | 0.033 | 9.81E-13 | 0.055 | 4.05E-05 | -0.025 | 9.38E-28 | SEMA3F | 3p21.31 | known |
| AN | Basal metabolic rate | rs2071206 | 3 | 50160109 | G | A | 0.030 | 3.21E-09 | 0.058 | 1.95E-05 | -0.015 | 2.95E-23 | SEMA3F | 3p21.31 | known |
| AN | Body mass index (BMI) | rs2071206 | 3 | 50160109 | G | A | 0.035 | 2.34E-14 | 0.058 | 1.95E-05 | -0.029 | 1.50E-36 | SEMA3F | 3p21.31 | known |
| AN | Whole body fat mass | rs2352974 | 3 | 49890613 | C | T | -0.032 | 1.27E-12 | -0.058 | 2.02E-05 | 0.024 | 2.24E-25 | TRAIP | 3p21.31 | known |
| AN | Basal metabolic rate | rs2681780 | 3 | 49897830 | C | T | -0.030 | 2.10E-09 | -0.059 | 1.52E-05 | 0.015 | 4.12E-23 | TRAIP | 3p21.31 | known |

| | | | | | | | | | | | | | | | |
|---|---|---|---|---|---|---|---|---|---|---|---|---|---|---|---|
| AN | Body mass index (BMI) | rs2681780 | 3 | 49897830 | C | T | -0.036 | 4.23E-15 | -0.059 | 1.52E-05 | 0.031 | 3.45E-39 | TRAIP | 3p21.31 | known |
| AN | HDL cholesterol (quantile) | rs4513485 | 3 | 49139966 | G | A | -0.031 | 1.14E-08 | -0.077 | 4.67E-08 | -0.007 | 5.84E-03 | QARS1 | 3p21.31 | known |
| AN | Basal metabolic rate | rs56116382 | 3 | 49606188 | A | C | 0.037 | 2.12E-08 | 0.092 | 7.13E-08 | -0.006 | 3.42E-03 | BSN | 3p21.31 | known |
| AN | HDL cholesterol (quantile) | rs56116382 | 3 | 49606188 | A | C | 0.037 | 1.77E-08 | 0.092 | 7.13E-08 | 0.008 | 6.25E-03 | BSN | 3p21.31 | known |
| AN | Fasting insulin | rs61583136 | 3 | 49189195 | C | T | -0.032 | 1.39E-08 | -0.077 | 4.52E-08 | 0.005 | 0.012 | DHFR2 | 3p21.31 | known |
| AN | Triglycerides (quantile) | rs61583136 | 3 | 49189195 | C | T | -0.031 | 1.84E-08 | -0.077 | 4.52E-08 | 0.006 | 1.43E-02 | DHFR2 | 3p21.31 | known |
| AN | Whole body fat mass | rs73074378 | 3 | 48446237 | C | T | 0.049 | 1.58E-08 | 0.138 | 4.72E-08 | -0.013 | 2.37E-03 | PLXNB1 | 3p21.31 | known |
| AN | Body mass index (BMI) | rs73077200 | 3 | 49826987 | C | T | 0.056 | 1.00E-12 | 0.169 | 2.77E-13 | -0.011 | 6.88E-03 | IP6K1 | 3p21.31 | known |
| AN | Whole body fat mass | rs73080977 | 3 | 50189531 | G | T | 0.060 | 1.75E-13 | 0.175 | 1.31E-12 | -0.016 | 7.25E-05 | SEMA3F | 3p21.31 | known |
| AN | Body mass index (BMI) | rs73082362 | 3 | 49113729 | C | T | 0.058 | 4.18E-13 | 0.173 | 4.27E-14 | -0.010 | 1.33E-02 | QRICH1 | 3p21.31 | known |
| AN | Whole body fat mass | rs73082362 | 3 | 49113729 | C | T | 0.060 | 1.53E-14 | 0.173 | 4.27E-14 | -0.015 | 1.41E-04 | QRICH1 | 3p21.31 | known |
| AN | Body mass index (BMI) | rs7645445 | 3 | 49502243 | C | T | -0.028 | 1.12E-09 | -0.066 | 1.06E-06 | 0.014 | 1.47E-09 | DAG1 | 3p21.31 | known |
| AN | Fasting insulin | rs7645445 | 3 | 49502243 | C | T | -0.029 | 4.69E-08 | -0.066 | 1.06E-06 | 0.011 | 3.82E-07 | DAG1 | 3p21.31 | known |
| AN | Whole body fat mass | rs7645445 | 3 | 49502243 | C | T | -0.030 | 7.58E-11 | -0.066 | 1.06E-06 | 0.016 | 1.73E-12 | DAG1 | 3p21.31 | known |
| AN | HDL cholesterol (quantile) | rs9812977 | 3 | 48720303 | G | A | 0.064 | 8.45E-15 | 0.165 | 8.57E-15 | 0.009 | 2.01E-02 | NCKIPSD | 3p21.31 | known |
| AN | Whole body fat mass | rs9812977 | 3 | 48720303 | G | A | 0.060 | 1.19E-15 | 0.165 | 8.57E-15 | -0.016 | 2.73E-05 | NCKIPSD | 3p21.31 | known |
| AN | Body mass index (BMI) | rs9873726 | 3 | 48712367 | C | T | 0.055 | 2.07E-14 | 0.156 | 7.88E-15 | -0.011 | 1.93E-03 | NCKIPSD | 3p21.31 | known |
| AN | Basal metabolic rate | rs9813198 | 3 | 41258048 | A | G | -0.028 | 2.22E-08 | -0.061 | 7.08E-06 | 0.011 | 4.20E-13 | CTNNB1 | 3p22.1 | known |
| AN | Whole body fat mass | rs9813198 | 3 | 41258048 | A | G | -0.027 | 2.22E-09 | -0.061 | 7.08E-06 | 0.015 | 7.83E-11 | CTNNB1 | 3p22.1 | known |
| AN | Body mass index (BMI) | rs1492014 | 3 | 94071481 | T | C | -0.028 | 1.43E-09 | -0.057 | 4.11E-05 | 0.019 | 1.00E-15 | DHFR2 | 3q11.2 | known |
| AN | Whole body fat mass | rs1492014 | 3 | 94071481 | T | C | -0.028 | 1.53E-09 | -0.057 | 4.11E-05 | 0.018 | 2.49E-14 | DHFR2 | 3q11.2 | known |
| AN | Body mass index (BMI) | rs4629334 | 3 | 94581146 | G | A | -0.027 | 7.53E-09 | -0.077 | 9.64E-08 | 0.009 | 1.41E-04 | DHFR2 | 3q11.2 | known |
| AN | Basal metabolic rate | rs7619675 | 3 | 94593959 | A | C | -0.028 | 1.80E-08 | -0.075 | 5.23E-08 | 0.004 | 4.03E-03 | DHFR2 | 3q11.2 | known |
| AN | HDL cholesterol (quantile) | rs7619675 | 3 | 94593959 | A | C | -0.028 | 2.55E-08 | -0.075 | 5.23E-08 | -0.005 | 2.45E-02 | DHFR2 | 3q11.2 | known |
| AN | Triglycerides (quantile) | rs7619675 | 3 | 94593959 | A | C | -0.029 | 1.59E-08 | -0.075 | 5.23E-08 | 0.007 | 5.20E-03 | DHFR2 | 3q11.2 | known |

| | | | | | | | | | | | | | | |
|---|---|---|---|---|---|---|---|---|---|---|---|---|---|---|
| AN | Whole body fat mass | rs7619675 | 3 | 94593959 | A | C | -0.028 | 1.56E-09 | -0.075 | 5.23E-08 | 0.010 | 2.47E-05 | DHFR2 | 3q11.2 | known |
| AN | Whole body fat mass | rs7647832 | 3 | 94284034 | C | T | -0.027 | 1.17E-08 | -0.076 | 6.66E-07 | 0.010 | 9.73E-06 | DHFR2 | 3q11.2 | known |
| AN | Basal metabolic rate | rs10939795 | 4 | 18495811 | A | G | -0.030 | 4.67E-08 | -0.067 | 6.54E-06 | 0.011 | 1.51E-10 | LCORL | 4p15.31 | new |
| AN | Body mass index (BMI) | rs11729200 | 4 | 18521591 | G | A | -0.030 | 1.11E-09 | -0.065 | 8.81E-06 | 0.018 | 4.84E-13 | LCORL | 4p15.31 | new |
| AN | Whole body fat mass | rs11735817 | 4 | 18454458 | C | T | -0.030 | 1.40E-09 | -0.064 | 8.31E-06 | 0.017 | 1.58E-11 | LCORL | 4p15.31 | new |
| AN | Fasting blood insulin | rs13125932 | 4 | 58150175 | C | T | 0.030 | 2.70E-08 | 0.072 | 5.85E-08 | -0.008 | 0.027 | IGFBP7 | 4q12 | new |
| AN | Whole body fat mass | rs4320241 | 5 | 74299144 | C | T | 0.031 | 1.71E-08 | 0.063 | 1.40E-04 | -0.020 | 7.34E-13 | GCNT4 | 5q13.3 | new |
| AN | Whole body fat mass | rs4472337 | 6 | 34769765 | C | T | -0.039 | 6.40E-10 | -0.039 | 2.81E-02 | 0.041 | 3.87E-38 | MHC region | 6p21.31 | |
| AN | Basal metabolic rate | rs3131296 | 6 | 32172993 | C | T | -0.045 | 8.67E-11 | -0.085 | 2.17E-05 | 0.027 | 1.14E-37 | MHC region | 6p21.32 | |
| AN | Whole body fat mass | rs3131296 | 6 | 32172993 | C | T | -0.037 | 6.94E-09 | -0.085 | 2.17E-05 | 0.021 | 6.55E-11 | MHC region | 6p21.32 | |
| AN | HDL cholesterol (quantile) | rs3134942 | 6 | 32168771 | G | T | -0.040 | 1.42E-08 | -0.085 | 2.05E-05 | -0.025 | 1.12E-15 | MHC region | 6p21.32 | |
| AN | Basal metabolic rate | rs9274678 | 6 | 32636646 | A | G | -0.050 | 2.41E-10 | -0.098 | 8.27E-05 | 0.026 | 8.22E-34 | MHC region | 6p21.32 | |
| AN | Basal metabolic rate | rs1052486 | 6 | 31610686 | A | G | 0.028 | 2.18E-08 | 0.045 | 8.24E-04 | -0.019 | 6.90E-36 | MHC region | 6p21.33 | |
| AN | HDL cholesterol (quantile) | rs2507989 | 6 | 31324415 | A | C | -0.037 | 2.28E-09 | -0.078 | 1.22E-05 | -0.025 | 3.90E-19 | MHC region | 6p21.33 | |
| AN | Whole body fat mass | rs3130047 | 6 | 31610479 | C | T | -0.035 | 2.16E-09 | -0.067 | 1.16E-04 | 0.023 | 5.68E-16 | MHC region | 6p21.33 | |
| AN | HDL cholesterol (quantile) | rs3131379 | 6 | 31721033 | G | A | -0.043 | 5.16E-09 | -0.093 | 1.44E-05 | -0.028 | 1.75E-17 | MHC region | 6p21.33 | |
| AN | Basal metabolic rate | rs3132604 | 6 | 30741226 | C | T | -0.037 | 2.90E-09 | -0.086 | 1.71E-06 | 0.014 | 5.82E-14 | MHC region | 6p21.33 | |
| AN | Body mass index (BMI) | rs3132604 | 6 | 30741226 | C | T | -0.032 | 2.62E-08 | -0.086 | 1.71E-06 | 0.013 | 3.46E-06 | MHC region | 6p21.33 | |
| AN | HDL cholesterol (quantile) | rs3132604 | 6 | 30741226 | C | T | -0.039 | 7.00E-10 | -0.086 | 1.71E-06 | -0.023 | 1.39E-15 | MHC region | 6p21.33 | |
| AN | Whole body fat mass | rs3132604 | 6 | 30741226 | C | T | -0.035 | 6.68E-10 | -0.086 | 1.71E-06 | 0.018 | 3.18E-10 | MHC region | 6p21.33 | |
| AN | Whole body fat mass | rs532086 | 6 | 31881309 | T | C | -0.034 | 2.77E-08 | -0.090 | 2.83E-06 | 0.015 | 2.61E-06 | MHC region | 6p21.33 | |
| AN | Basal metabolic rate | rs9265645 | 6 | 31300197 | G | A | -0.044 | 1.09E-09 | -0.092 | 3.36E-05 | 0.024 | 1.78E-28 | MHC region | 6p21.33 | |
| AN | Basal metabolic rate | rs9267549 | 6 | 31677391 | G | A | -0.048 | 6.99E-11 | -0.093 | 1.54E-05 | 0.029 | 9.36E-37 | MHC region | 6p21.33 | |
| AN | HDL cholesterol (quantile) | rs885912 | 6 | 30174633 | C | A | -0.035 | 6.70E-09 | -0.081 | 2.19E-06 | -0.017 | 8.00E-11 | MHC region | 6p22.1 | |
| AN | Body mass index (BMI) | rs9320913 | 6 | 98584733 | C | A | 0.026 | 2.55E-08 | 0.056 | 2.58E-05 | -0.015 | 4.88E-10 | POU3F2 | 6q16.1 | new |

| | | | | | | | | | | | | | | |
|---|---|---|---|---|---|---|---|---|---|---|---|---|---|---|
| AN | Whole body fat mass | rs9401593 | 6 | 98549801 | A | C | 0.028 | 1.55E-09 | 0.057 | 2.38E-05 | -0.017 | 2.79E-13 | POU3F2 | 6q16.1 | new |
| AN | HDL cholesterol (quantile) | rs10105606 | 8 | 19827848 | C | A | 0.058 | 3.68E-26 | 0.041 | 4.37E-03 | 0.092 | 0.00E+00 | LPL | 8p21.3 | new |
| AN | HDL cholesterol (quantile) | rs142084074 | 8 | 19768150 | G | A | 0.124 | 1.22E-08 | 0.186 | 2.81E-03 | 0.127 | 9.86E-40 | LPL | 8p21.3 | new |
| AN | HDL cholesterol (quantile) | rs17091237 | 8 | 19593584 | C | T | 0.030 | 4.64E-08 | 0.048 | 9.78E-04 | 0.028 | 1.44E-29 | CSGALNACT1 | 8p21.3 | new |
| AN | Triglycerides (quantile) | rs73600084 | 8 | 19849992 | T | C | 0.057 | 5.89E-15 | 0.056 | 3.02E-03 | -0.111 | 5.43E-253 | LPL | 8p21.3 | new |
| AN | Triglycerides (quantile) | rs77829308 | 8 | 19741186 | C | T | 0.044 | 7.97E-11 | 0.050 | 4.21E-03 | -0.078 | 1.54E-145 | LPL | 8p21.3 | new |
| AN | HDL cholesterol (quantile) | rs7814983 | 8 | 19639365 | C | T | 0.034 | 1.75E-09 | 0.030 | 4.26E-02 | 0.050 | 7.95E-86 | CSGALNACT1 | 8p21.3 | new |
| AN | HDL cholesterol (quantile) | rs2625674 | 8 | 116492221 | G | A | -0.031 | 9.24E-10 | -0.049 | 4.31E-04 | -0.031 | 3.55E-41 | TRPS1 | 8q23.3 | new |

MTAG section of the table presents the MTAG results specific to Trait 1. Trait 1, anorexia nervosa.

**Table S12**. Two-trait MTAG of anorexia nervosa and other traits.

| Trait1 | Trait2 | SNP | CHR | POS | A1 | A2 | MTAG | | AN | | Risk factor | | Genes | Cytogenetic band | Annotation |
|---|---|---|---|---|---|---|---|---|---|---|---|---|---|---|---|
| | | | | | | | beta | p | beta | p | beta | p | | | |
| AN | Snoring | rs725861 | 10 | 9063776 | A | G | -0.040 | 2.39E-09 | -0.088 | 4.86E-07 | -0.008 | 1.02E-08 | GATA3 | 10p14 | new |
| AN | SHBG (quantile) | rs11815832 | 10 | 65065137 | C | A | -0.037 | 3.63E-13 | -0.031 | 2.66E-02 | -0.053 | 1.44E-115 | NRBF2 | 10q21.3 | new |
| AN | Sex hormone-binding globulin levels | rs11815832 | 10 | 65065137 | C | A | -0.040 | 2.67E-15 | -0.031 | 2.66E-02 | 0.029 | 3.20E-221 | NRBF2 | 10q21.3 | new |
| AN | Alanine aminotransferase (quantile) | rs35614792 | 10 | 101934235 | C | T | 0.031 | 5.44E-09 | 0.031 | 2.08E-02 | -0.055 | 6.94E-132 | ERLIN1 | 10q24.31 | new |
| AN | Red blood cell (erythrocyte) distribution width | rs7079165 | 10 | 131430240 | G | A | 0.030 | 2.53E-08 | 0.076 | 3.66E-08 | -0.005 | 4.07E-02 | MGMT | 10q26.3 | known |
| AN | Diastolic blood pressure, automated reading | rs2008387 | 10 | 131448764 | G | A | 0.032 | 4.56E-09 | 0.082 | 1.73E-08 | -0.007 | 3.85E-03 | MGMT | 10q26.3 | known |
| AN | Creatinine (quantile) | rs2008387 | 10 | 131448764 | G | A | 0.033 | 2.79E-09 | 0.082 | 1.73E-08 | -0.014 | 1.70E-11 | MGMT | 10q26.3 | known |
| AN | Age completed full time education | rs2008387 | 10 | 131448764 | G | A | 0.032 | 1.01E-08 | 0.082 | 1.73E-08 | 0.005 | 4.56E-02 | MGMT | 10q26.3 | known |
| AN | Sex hormone-binding globulin levels | rs900144 | 11 | 13272721 | C | T | -0.030 | 4.86E-09 | -0.060 | 1.07E-05 | 0.008 | 1.00E-19 | ARNTL | 11p15.2 | new |
| AN | SHBG (quantile) | rs900144 | 11 | 13294268 | C | T | -0.029 | 1.16E-08 | -0.060 | 1.07E-05 | -0.016 | 2.01E-12 | ARNTL | 11p15.2 | new |
| AN | Urate (quantile) | rs2078267 | 11 | 64334114 | C | T | 0.038 | 2.26E-13 | 0.033 | 1.36E-02 | -0.054 | 7.15E-153 | NRXN2 | 11q13.1 | new |
| AN | Number of cigarettes previously smoked daily | rs750350 | 11 | 112885342 | G | T | 0.042 | 4.45E-08 | 0.111 | 9.42E-08 | -0.017 | 4.84E-03 | NCAM1 | 11q23.2 | new |
| AN | Ever taken cannabis | rs750350 | 11 | 112885342 | G | T | 0.043 | 2.32E-08 | 0.111 | 9.42E-08 | 0.012 | 2.72E-02 | NCAM1 | 11q23.2 | new |
| AN | Ever addicted to any substance or behaviour | rs750350 | 11 | 112885342 | G | T | 0.044 | 2.01E-08 | 0.111 | 9.42E-08 | 0.004 | 6.13E-03 | NCAM1 | 11q23.2 | new |
| AN | Apoliprotein B (quantile) | rs56182612 | 11 | 115032622 | A | G | 0.045 | 1.20E-08 | 0.116 | 1.31E-08 | -0.007 | 4.89E-02 | CADM1 | 11q23.3 | known |
| AN | Alanine aminotransferase (quantile) | rs4468381 | 11 | 115048420 | C | T | 0.039 | 2.33E-08 | 0.093 | 4.90E-08 | -0.006 | 4.85E-02 | CADM1 | 11q23.3 | known |
| AN | Mean corpuscular volume | rs1938736 | 11 | 115092997 | G | T | 0.039 | 1.20E-08 | 0.103 | 1.56E-09 | -0.006 | 4.69E-02 | CADM1 | 11q23.3 | known |
| AN | Trouble falling asleep | rs10790068 | 11 | 115101567 | C | T | -0.044 | 2.97E-10 | -0.105 | 8.11E-10 | 0.011 | 3.59E-03 | DHFR2 | 11q23.3 | known |
| AN | Amount of alcohol drunk on a typical drinking day | rs4409873 | 11 | 115104646 | A | G | -0.046 | 7.68E-10 | -0.111 | 1.63E-09 | 0.017 | 4.47E-03 | CADM1 | 11q23.3 | known |
| AN | Urate (quantile) | rs10502199 | 11 | 115120615 | C | T | 0.047 | 1.40E-10 | 0.121 | 2.70E-10 | -0.007 | 2.65E-02 | CADM1 | 11q23.3 | known |
| AN | Reticulocyte percentage | rs10502199 | 11 | 115120615 | C | T | 0.048 | 1.55E-10 | 0.121 | 2.70E-10 | -0.008 | 2.52E-02 | CADM1 | 11q23.3 | known |
| AN | Gamma glutamyltransferase (quantile) | rs10502199 | 11 | 115120615 | C | T | 0.048 | 1.26E-10 | 0.121 | 2.70E-10 | -0.008 | 1.33E-02 | CADM1 | 11q23.3 | known |
| AN | Eosinophill count | rs10502199 | 11 | 115120615 | C | T | 0.047 | 4.48E-10 | 0.121 | 2.70E-10 | -0.007 | 9.39E-03 | CADM1 | 11q23.3 | known |
| AN | C-reactive protein (quantile) | rs10502199 | 11 | 115120615 | C | T | 0.049 | 1.33E-11 | 0.121 | 2.70E-10 | -0.014 | 6.90E-05 | CADM1 | 11q23.3 | known |
| AN | Age started smoking in former smokers | rs10502199 | 11 | 115120615 | C | T | 0.050 | 2.17E-11 | 0.121 | 2.70E-10 | 0.023 | 6.09E-04 | CADM1 | 11q23.3 | known |

| | | | | | | | | | | | | | | | |
|---|---|---|---|---|---|---|---|---|---|---|---|---|---|---|---|
| AN | Snoring | rs72994087 | 11 | 115134635 | A | C | 0.054 | 3.26E-11 | 0.130 | 4.09E-10 | 0.006 | 1.47E-03 | CADM1 | 11q23.3 | known |
| AN | SHBG (quantile) | rs72994087 | 11 | 115134635 | A | C | 0.050 | 2.34E-10 | 0.130 | 4.09E-10 | 0.008 | 2.37E-02 | CADM1 | 11q23.3 | known |
| AN | Reticulocyte count | rs72994087 | 11 | 115134635 | A | C | 0.052 | 1.43E-10 | 0.130 | 4.09E-10 | -0.011 | 3.42E-03 | CADM1 | 11q23.3 | known |
| AN | High light scatter reticulocyte percentage | rs72994087 | 11 | 115134635 | A | C | 0.052 | 1.60E-10 | 0.130 | 4.09E-10 | -0.010 | 5.92E-03 | CADM1 | 11q23.3 | known |
| AN | High light scatter reticulocyte count | rs72994087 | 11 | 115134635 | A | C | 0.052 | 1.51E-10 | 0.130 | 4.09E-10 | -0.010 | 5.06E-03 | CADM1 | 11q23.3 | known |
| AN | Sex hormone-binding globulin levels adjusted for BMI | rs10458969 | 11 | 115232473 | A | G | -0.044 | 1.03E-09 | -0.110 | 1.48E-09 | 0.003 | 2.20E-02 | CADM1 | 11q23.3 | known |
| AN | Sex hormone-binding globulin levels | rs10502199 | 11 | 115249895 | C | T | 0.047 | 6.03E-11 | 0.121 | 2.70E-10 | -0.004 | 6.80E-04 | CADM1 | 11q23.3 | known |
| AN | Urate (quantile) | rs11609805 | 12 | 57735045 | G | A | 0.038 | 1.20E-10 | 0.044 | 5.90E-03 | -0.048 | 2.56E-90 | R3HDM2 | 12q13.3 | new |
| AN | C-reactive protein (quantile) | rs2203474 | 12 | 121232400 | T | G | 0.074 | 1.31E-11 | 0.065 | 4.73E-02 | -0.098 | 1.45E-98 | ACADS | 12q24.31 | new |
| AN | C-reactive protein (quantile) | rs2701184 | 12 | 121353088 | T | G | 0.051 | 1.22E-22 | 0.032 | 2.13E-02 | -0.091 | 3.01E-306 | SPPL3 | 12q24.31 | new |
| AN | Gamma glutamyltransferase (quantile) | rs61946383 | 12 | 121356075 | A | G | 0.032 | 6.47E-09 | 0.040 | 4.32E-03 | -0.063 | 3.89E-163 | SPPL3 | 12q24.31 | new |
| AN | SHBG (quantile) | rs62062620 | 17 | 7659832 | G | A | 0.029 | 5.98E-09 | 0.049 | 6.04E-04 | 0.026 | 1.18E-30 | DNAH2 | 17p13.1 | new |
| AN | Sex hormone-binding globulin levels | rs62062620 | 17 | 7659832 | G | A | 0.029 | 5.82E-09 | 0.049 | 6.04E-04 | -0.012 | 6.70E-40 | DNAH2 | 17p13.1 | new |
| AN | SHBG (quantile) | rs79930761 | 17 | 7815712 | C | T | 0.074 | 8.07E-12 | 0.074 | 1.46E-02 | 0.068 | 7.83E-63 | RNF227 | 17p13.1 | new |
| AN | Sex hormone-binding globulin levels | rs79930761 | 17 | 7815712 | C | T | 0.073 | 2.77E-11 | 0.074 | 1.46E-02 | -0.031 | 2.90E-76 | RNF227 | 17p13.1 | new |
| AN | Red blood cell (erythrocyte) count | rs8070063 | 17 | 8088612 | G | T | 0.030 | 2.02E-08 | 0.075 | 9.73E-08 | -0.010 | 3.97E-07 | BORCS6 | 17p13.1 | new |
| AN | Gamma glutamyltransferase (quantile) | rs8070063 | 17 | 8088612 | G | T | 0.030 | 8.40E-09 | 0.075 | 9.73E-08 | -0.011 | 4.40E-07 | BORCS6 | 17p13.1 | new |
| AN | Glycated hemoglobin levels（HbA1c） | rs12938531 | 17 | 8222223 | G | A | -0.029 | 2.11E-08 | -0.070 | 3.23E-07 | 0.005 | 4.36E-04 | AURKB | 17p13.1 | new |
| AN | Gamma glutamyltransferase (quantile) | rs10153315 | 17 | 79481772 | T | C | -0.034 | 3.43E-09 | -0.057 | 9.73E-05 | 0.039 | 1.60E-68 | ACTG1 | 17q25.3 | new |
| AN | SHBG (quantile) | rs1640267 | 19 | 2789337 | T | C | 0.031 | 4.82E-08 | 0.035 | 4.19E-02 | 0.038 | 1.34E-52 | THOP1 | 19p13.3 | new |
| AN | Sex hormone-binding globulin levels | rs1640267 | 19 | 2789337 | T | C | 0.032 | 1.72E-08 | 0.035 | 4.19E-02 | -0.019 | 4.90E-85 | THOP1 | 19p13.3 | new |
| AN | C-reactive protein (quantile) | rs6859 | 19 | 45382034 | A | G | -0.034 | 4.31E-11 | -0.029 | 3.13E-02 | 0.054 | 9.81E-111 | TOMM40 | 19q13.32 | new |
| AN | C-reactive protein (quantile) | rs439401 | 19 | 45414451 | T | C | 0.049 | 1.33E-20 | 0.030 | 3.26E-02 | -0.089 | 3.36E-279 | APOC1 | 19q13.32 | new |
| AN | Snoring | rs10747478 | 1 | 96901455 | T | G | -0.032 | 2.01E-09 | -0.076 | 3.13E-08 | -0.004 | 5.07E-04 | PTBP2 | 1p21.3 | known |
| AN | Glucose (quantile) | rs10747478 | 1 | 96901455 | T | G | -0.030 | 1.06E-08 | -0.076 | 3.13E-08 | 0.006 | 2.87E-02 | PTBP2 | 1p21.3 | known |
| AN | White blood cell (leukocyte) count | rs11165643 | 1 | 96924097 | C | T | -0.030 | 2.36E-08 | -0.075 | 3.47E-08 | 0.006 | 1.33E-02 | PTBP2 | 1p21.3 | known |
| AN | SHBG (quantile) | rs11165643 | 1 | 96924097 | C | T | -0.029 | 9.93E-09 | -0.075 | 3.47E-08 | -0.006 | 1.12E-02 | PTBP2 | 1p21.3 | known |
| AN | Sex hormone-binding globulin levels | rs11165643 | 1 | 96924097 | C | T | -0.030 | 3.39E-09 | -0.075 | 3.47E-08 | 0.004 | 9.40E-05 | PTBP2 | 1p21.3 | known |

| | | | | | | | | | | | | | | | |
|---|---|---|---|---|---|---|---|---|---|---|---|---|---|---|---|
| AN | Reticulocyte percentage | rs11165643 | 1 | 96924097 | C | T | -0.030 | 1.42E-08 | -0.075 | 3.47E-08 | 0.006 | 9.85E-03 | PTBP2 | 1p21.3 | known |
| AN | Reticulocyte count | rs11165643 | 1 | 96924097 | C | T | -0.030 | 1.51E-08 | -0.075 | 3.47E-08 | 0.006 | 1.45E-02 | PTBP2 | 1p21.3 | known |
| AN | Neutrophill count | rs11165643 | 1 | 96924097 | C | T | -0.030 | 2.18E-08 | -0.075 | 3.47E-08 | 0.005 | 3.55E-02 | PTBP2 | 1p21.3 | known |
| AN | Immature reticulocyte fraction | rs11165643 | 1 | 96924097 | C | T | -0.030 | 1.11E-08 | -0.075 | 3.47E-08 | 0.006 | 1.02E-02 | PTBP2 | 1p21.3 | known |
| AN | High light scatter reticulocyte percentage | rs11165643 | 1 | 96924097 | C | T | -0.030 | 9.21E-09 | -0.075 | 3.47E-08 | 0.008 | 2.03E-03 | PTBP2 | 1p21.3 | known |
| AN | High light scatter reticulocyte count | rs11165643 | 1 | 96924097 | C | T | -0.030 | 8.68E-09 | -0.075 | 3.47E-08 | 0.008 | 1.83E-03 | PTBP2 | 1p21.3 | known |
| AN | Gamma glutamyltransferase (quantile) | rs11165643 | 1 | 96924097 | C | T | -0.031 | 7.81E-09 | -0.075 | 3.47E-08 | 0.008 | 5.27E-04 | PTBP2 | 1p21.3 | known |
| AN | Diastolic blood pressure, automated reading | rs11165643 | 1 | 96924097 | C | T | -0.030 | 1.09E-08 | -0.075 | 3.47E-08 | 0.006 | 1.41E-02 | PTBP2 | 1p21.3 | known |
| AN | C-reactive protein (quantile) | rs11165643 | 1 | 96924097 | C | T | -0.031 | 2.53E-09 | -0.075 | 3.47E-08 | 0.009 | 2.36E-04 | PTBP2 | 1p21.3 | known |
| AN | circulating leptin levels | rs11165643 | 1 | 96924097 | C | T | -0.031 | 1.38E-08 | -0.075 | 3.47E-08 | 0.014 | 0.027 | PTBP2 | 1p21.3 | known |
| AN | Alanine aminotransferase (quantile) | rs11165643 | 1 | 96924097 | C | T | -0.031 | 6.75E-09 | -0.075 | 3.47E-08 | 0.007 | 3.27E-03 | PTBP2 | 1p21.3 | known |
| AN | C-reactive protein (quantile) | rs6588156 | 1 | 66160433 | A | G | -0.068 | 2.23E-26 | -0.051 | 1.59E-03 | 0.113 | 0.00E+00 | PDE4B | 1p31.3 | new |
| AN | C-reactive protein (quantile) | rs17448335 | 1 | 66195595 | A | G | -0.047 | 2.70E-18 | -0.033 | 2.87E-02 | 0.084 | 7.53E-234 | PDE4B | 1p31.3 | new |
| AN | C-reactive protein (quantile) | rs12058296 | 1 | 66402424 | C | A | -0.038 | 1.25E-12 | -0.057 | 8.66E-05 | 0.042 | 1.21E-62 | PDE4B | 1p31.3 | new |
| AN | Reticulocyte count | rs4751 | 1 | 1686040 | G | T | 0.035 | 3.52E-08 | 0.082 | 1.16E-06 | -0.014 | 5.23E-09 | NADK | 1p36.33 | new |
| AN | C-reactive protein (quantile) | rs4845373 | 1 | 154417829 | C | T | 0.029 | 1.21E-08 | -0.028 | 3.91E-02 | -0.091 | 7.00E-307 | IL6R | 1q21.3 | new |
| AN | C-reactive protein (quantile) | rs145663198 | 1 | 159475349 | A | G | 0.098 | 6.70E-09 | 0.150 | 2.11E-03 | -0.111 | 2.39E-44 | OR10J5 | 1q23.2 | new |
| AN | C-reactive protein (quantile) | rs4255379 | 1 | 159718312 | G | A | -0.033 | 1.48E-10 | -0.030 | 3.15E-02 | 0.053 | 1.43E-102 | DUSP23 | 1q23.2 | new |
| AN | C-reactive protein (quantile) | rs10754051 | 1 | 193133527 | T | C | 0.028 | 4.61E-08 | 0.073 | 1.51E-07 | -0.006 | 8.76E-03 | B3GALT2 | 1q31.2 | new |
| AN | Systolic blood pressure, automated reading | rs1539725 | 1 | 193153682 | T | C | 0.029 | 4.55E-08 | 0.073 | 1.26E-07 | -0.005 | 1.75E-02 | B3GALT2 | 1q31.2 | new |
| AN | SHBG (quantile) | rs1539725 | 1 | 193153682 | T | C | 0.028 | 4.37E-08 | 0.073 | 1.26E-07 | 0.006 | 1.46E-02 | B3GALT2 | 1q31.2 | new |
| AN | Sex hormone-binding globulin levels | rs1539725 | 1 | 193153682 | T | C | 0.029 | 2.22E-08 | 0.073 | 1.26E-07 | -0.003 | 1.10E-03 | B3GALT2 | 1q31.2 | new |
| AN | Diastolic blood pressure, automated reading | rs1539725 | 1 | 193153682 | T | C | 0.029 | 4.99E-08 | 0.073 | 1.26E-07 | -0.006 | 2.02E-02 | B3GALT2 | 1q31.2 | new |
| AN | Bioavailable testosterone levels | rs1539725 | 1 | 193153682 | T | C | 0.030 | 3.44E-08 | 0.073 | 1.26E-07 | 0.007 | 4.80E-04 | B3GALT2 | 1q31.2 | new |
| AN | Immature reticulocyte fraction | rs61855662 | 1 | 248010606 | C | T | 0.043 | 4.35E-08 | 0.062 | 6.93E-03 | -0.053 | 3.35E-68 | OR11L1 | 1q44 | new |
| AN | Alanine aminotransferase (quantile) | rs6092932 | 20 | 37349253 | G | A | -0.039 | 4.26E-08 | -0.095 | 3.50E-07 | 0.011 | 2.47E-04 | SLC32A1 | 20q11.23 | new |
| AN | Sex hormone-binding globulin levels | rs174723 | 22 | 29847326 | A | G | -0.028 | 4.66E-08 | -0.072 | 2.69E-07 | 0.003 | 2.90E-04 | RFPL1 | 22q12.2 | new |
| AN | Reticulocyte percentage | rs174723 | 22 | 29847326 | A | G | -0.029 | 4.66E-08 | -0.072 | 2.69E-07 | 0.010 | 2.35E-05 | RFPL1 | 22q12.2 | new |

| | | | | | | | | | | | | | | |
|---|---|---|---|---|---|---|---|---|---|---|---|---|---|---|
| AN | Reticulocyte count | rs174723 | 22 | 29847326 | A | G | -0.029 | 4.02E-08 | -0.072 | 2.69E-07 | 0.010 | 1.83E-05 | RFPL1 | 22q12.2 | new |
| AN | Immature reticulocyte fraction | rs174723 | 22 | 29847326 | A | G | -0.030 | 8.98E-09 | -0.072 | 2.69E-07 | 0.014 | 2.01E-08 | RFPL1 | 22q12.2 | new |
| AN | High light scatter reticulocyte percentage | rs174723 | 22 | 29847326 | A | G | -0.030 | 1.39E-08 | -0.072 | 2.69E-07 | 0.014 | 2.14E-08 | RFPL1 | 22q12.2 | new |
| AN | High light scatter reticulocyte count | rs174723 | 22 | 29847326 | A | G | -0.030 | 1.31E-08 | -0.072 | 2.69E-07 | 0.013 | 3.53E-08 | RFPL1 | 22q12.2 | new |
| AN | Red blood cell (erythrocyte) count | rs77732866 | 2 | 58979879 | G | A | -0.042 | 4.93E-08 | -0.099 | 4.99E-07 | 0.022 | 4.10E-13 | FANCL | 2p16.1 | new |
| AN | Mean corpuscular volume | rs77732866 | 2 | 58979879 | G | A | -0.044 | 1.01E-08 | -0.099 | 4.99E-07 | -0.029 | 1.58E-17 | FANCL | 2p16.1 | new |
| AN | Mean corpuscular haemoglobin | rs77732866 | 2 | 58979879 | G | A | -0.043 | 1.75E-08 | -0.099 | 4.99E-07 | -0.027 | 5.90E-15 | FANCL | 2p16.1 | new |
| AN | Glucose (quantile) | rs10187718 | 2 | 53971019 | A | C | 0.045 | 2.03E-08 | 0.107 | 6.35E-08 | -0.008 | 3.31E-02 | CHAC2 | 2p16.2 | known |
| AN | White blood cell (leukocyte) count | rs2287348 | 2 | 54039813 | C | T | 0.041 | 2.67E-08 | 0.104 | 5.62E-09 | 0.010 | 3.44E-03 | ERLEC1 | 2p16.2 | known |
| AN | Total protein (quantile) | rs2287348 | 2 | 54039813 | C | T | 0.042 | 8.83E-09 | 0.104 | 5.62E-09 | 0.009 | 1.14E-02 | ERLEC1 | 2p16.2 | known |
| AN | SHBG (quantile) | rs2287348 | 2 | 54039813 | C | T | 0.041 | 3.42E-09 | 0.104 | 5.62E-09 | 0.007 | 3.63E-02 | ERLEC1 | 2p16.2 | known |
| AN | Sex hormone-binding globulin levels | rs2287348 | 2 | 54039813 | C | T | 0.041 | 3.44E-09 | 0.104 | 5.62E-09 | -0.003 | 1.90E-02 | ERLEC1 | 2p16.2 | known |
| AN | Reticulocyte percentage | rs2287348 | 2 | 54039813 | C | T | 0.045 | 6.56E-10 | 0.104 | 5.62E-09 | -0.015 | 7.61E-06 | ERLEC1 | 2p16.2 | known |
| AN | Reticulocyte count | rs2287348 | 2 | 54039813 | C | T | 0.045 | 4.07E-10 | 0.104 | 5.62E-09 | -0.016 | 8.08E-07 | ERLEC1 | 2p16.2 | known |
| AN | Red blood cell (erythrocyte) distribution width | rs2287348 | 2 | 54039813 | C | T | 0.043 | 3.10E-09 | 0.104 | 5.62E-09 | -0.008 | 1.14E-02 | ERLEC1 | 2p16.2 | known |
| AN | Red blood cell (erythrocyte) count | rs2287348 | 2 | 54039813 | C | T | 0.043 | 3.73E-09 | 0.104 | 5.62E-09 | -0.007 | 1.59E-02 | ERLEC1 | 2p16.2 | known |
| AN | Mean reticulocyte volume | rs2287348 | 2 | 54039813 | C | T | 0.042 | 9.25E-09 | 0.104 | 5.62E-09 | -0.008 | 1.79E-02 | ERLEC1 | 2p16.2 | known |
| AN | Lymphocyte count | rs2287348 | 2 | 54039813 | C | T | 0.042 | 9.98E-09 | 0.104 | 5.62E-09 | 0.008 | 1.06E-02 | ERLEC1 | 2p16.2 | known |
| AN | Immature reticulocyte fraction | rs2287348 | 2 | 54039813 | C | T | 0.045 | 4.76E-10 | 0.104 | 5.62E-09 | -0.014 | 3.14E-05 | ERLEC1 | 2p16.2 | known |
| AN | High light scatter reticulocyte percentage | rs2287348 | 2 | 54039813 | C | T | 0.045 | 3.07E-10 | 0.104 | 5.62E-09 | -0.017 | 2.90E-07 | ERLEC1 | 2p16.2 | known |
| AN | High light scatter reticulocyte count | rs2287348 | 2 | 54039813 | C | T | 0.046 | 1.93E-10 | 0.104 | 5.62E-09 | -0.018 | 3.13E-08 | ERLEC1 | 2p16.2 | known |
| AN | Haemoglobin concentration | rs2287348 | 2 | 54039813 | C | T | 0.042 | 7.74E-09 | 0.104 | 5.62E-09 | -0.008 | 2.44E-03 | ERLEC1 | 2p16.2 | known |
| AN | Haematocrit percentage | rs2287348 | 2 | 54039813 | C | T | 0.043 | 6.29E-09 | 0.104 | 5.62E-09 | -0.010 | 2.03E-04 | ERLEC1 | 2p16.2 | known |
| AN | Gamma glutamyltransferase (quantile) | rs2287348 | 2 | 54039813 | C | T | 0.043 | 2.05E-09 | 0.104 | 5.62E-09 | -0.009 | 4.63E-03 | ERLEC1 | 2p16.2 | known |
| AN | Eosinophill count | rs3861570 | 2 | 54044560 | G | A | 0.044 | 4.95E-08 | 0.108 | 3.28E-08 | 0.007 | 1.10E-02 | ERLEC1 | 2p16.2 | known |
| AN | Glucose (quantile) | rs58851686 | 2 | 54284316 | G | T | 0.046 | 4.49E-08 | 0.107 | 2.40E-07 | -0.011 | 5.84E-03 | PSME4 | 2p16.2 | known |
| AN | Reticulocyte percentage | rs12620667 | 2 | 54290997 | G | A | 0.048 | 1.88E-08 | 0.108 | 2.31E-07 | -0.021 | 1.48E-07 | PSME4 | 2p16.2 | known |
| AN | Reticulocyte count | rs12620667 | 2 | 54290997 | G | A | 0.048 | 1.44E-08 | 0.108 | 2.31E-07 | -0.021 | 6.53E-08 | PSME4 | 2p16.2 | known |

| | | | | | | | | | | | | | | | |
|---|---|---|---|---|---|---|---|---|---|---|---|---|---|---|---|
| AN | Immature reticulocyte fraction | rs12620667 | 2 | 54290997 | G | A | 0.047 | 2.21E-08 | 0.108 | 2.31E-07 | -0.016 | 2.57E-05 | PSME4 | 2p16.2 | known |
| AN | High light scatter reticulocyte percentage | rs12620667 | 2 | 54290997 | G | A | 0.048 | 1.06E-08 | 0.108 | 2.31E-07 | -0.022 | 1.74E-08 | PSME4 | 2p16.2 | known |
| AN | High light scatter reticulocyte count | rs12620667 | 2 | 54290997 | G | A | 0.049 | 7.91E-09 | 0.108 | 2.31E-07 | -0.023 | 5.36E-09 | PSME4 | 2p16.2 | known |
| AN | C-reactive protein (quantile) | rs55709272 | 2 | 113867288 | T | C | -0.034 | 2.12E-11 | -0.044 | 1.05E-03 | 0.043 | 9.98E-71 | IL1RN | 2q13 | new |
| AN | Glucose (quantile) | rs11678685 | 2 | 169710607 | G | A | 0.034 | 1.08E-09 | 0.029 | 4.45E-02 | -0.066 | 1.14E-133 | G6PC2 | 2q31.1 | new |
| AN | Sex hormone-binding globulin levels | rs1915304 | 3 | 89655243 | C | T | -0.033 | 3.22E-08 | -0.095 | 3.14E-07 | 0.004 | 5.80E-05 | EPHA3 | 3p11.1 | new |
| AN | Urate (quantile) | rs1915304 | 3 | 89704393 | C | T | -0.034 | 2.21E-08 | -0.095 | 3.14E-07 | 0.009 | 1.29E-04 | EPHA3 | 3p11.1 | new |
| AN | Alanine aminotransferase (quantile) | rs9847154 | 3 | 70994006 | T | C | 0.031 | 5.86E-09 | 0.079 | 2.75E-08 | -0.007 | 2.52E-03 | MDFIC2 | 3p13 | known |
| AN | Systolic blood pressure, automated reading | rs9874207 | 3 | 71019750 | T | C | 0.030 | 9.47E-09 | 0.081 | 2.05E-08 | -0.005 | 3.60E-02 | FOXP1 | 3p13 | known |
| AN | SHBG (quantile) | rs9874207 | 3 | 71019750 | T | C | 0.029 | 9.33E-09 | 0.081 | 2.05E-08 | 0.005 | 2.25E-02 | FOXP1 | 3p13 | known |
| AN | Sex hormone-binding globulin levels adjusted for BMI | rs9874207 | 3 | 71019750 | T | C | 0.031 | 4.44E-09 | 0.081 | 2.05E-08 | -0.004 | 3.80E-07 | FOXP1 | 3p13 | known |
| AN | Sex hormone-binding globulin levels | rs9874207 | 3 | 71019750 | T | C | 0.030 | 1.76E-09 | 0.081 | 2.05E-08 | -0.004 | 1.40E-04 | FOXP1 | 3p13 | known |
| AN | Mean reticulocyte volume | rs9874207 | 3 | 71019750 | T | C | 0.029 | 3.38E-08 | 0.081 | 2.05E-08 | -0.006 | 1.45E-02 | FOXP1 | 3p13 | known |
| AN | Haemoglobin concentration | rs9874207 | 3 | 71019750 | T | C | 0.030 | 2.88E-08 | 0.081 | 2.05E-08 | -0.005 | 8.12E-03 | FOXP1 | 3p13 | known |
| AN | Haematocrit percentage | rs9874207 | 3 | 71019750 | T | C | 0.030 | 2.42E-08 | 0.081 | 2.05E-08 | -0.006 | 1.14E-03 | FOXP1 | 3p13 | known |
| AN | Diastolic blood pressure, automated reading | rs9874207 | 3 | 71019750 | T | C | 0.030 | 1.03E-08 | 0.081 | 2.05E-08 | -0.005 | 4.35E-02 | FOXP1 | 3p13 | known |
| AN | Bioavailable testosterone levels | rs9874207 | 3 | 71019750 | T | C | 0.030 | 9.28E-09 | 0.081 | 2.05E-08 | 0.005 | 1.30E-02 | FOXP1 | 3p13 | known |
| AN | Sleep duration | rs73074378 | 3 | 48446237 | C | T | 0.056 | 2.46E-08 | 0.138 | 4.72E-08 | -0.009 | 9.71E-03 | PLXNB1 | 3p21.31 | known |
| AN | Sex hormone-binding globulin levels | rs73074378 | 3 | 48446237 | C | T | 0.053 | 2.84E-08 | 0.138 | 4.72E-08 | -0.004 | 3.10E-02 | PLXNB1 | 3p21.31 | known |
| AN | Reticulocyte percentage | rs73074378 | 3 | 48446237 | C | T | 0.061 | 1.05E-09 | 0.138 | 4.72E-08 | -0.032 | 1.52E-12 | PLXNB1 | 3p21.31 | known |
| AN | Reticulocyte count | rs73074378 | 3 | 48446237 | C | T | 0.060 | 1.19E-09 | 0.138 | 4.72E-08 | -0.030 | 5.25E-11 | PLXNB1 | 3p21.31 | known |
| AN | Red blood cell (erythrocyte) distribution width | rs73074378 | 3 | 48446237 | C | T | 0.056 | 1.88E-08 | 0.138 | 4.72E-08 | -0.015 | 1.23E-03 | PLXNB1 | 3p21.31 | known |
| AN | Neutrophill count | rs73074378 | 3 | 48446237 | C | T | 0.055 | 3.19E-08 | 0.138 | 4.72E-08 | -0.010 | 2.55E-02 | PLXNB1 | 3p21.31 | known |
| AN | Immature reticulocyte fraction | rs73074378 | 3 | 48446237 | C | T | 0.059 | 2.42E-09 | 0.138 | 4.72E-08 | -0.022 | 8.77E-07 | PLXNB1 | 3p21.31 | known |
| AN | High light scatter reticulocyte percentage | rs73074378 | 3 | 48446237 | C | T | 0.062 | 4.75E-10 | 0.138 | 4.72E-08 | -0.034 | 8.26E-14 | PLXNB1 | 3p21.31 | known |
| AN | High light scatter reticulocyte count | rs73074378 | 3 | 48446237 | C | T | 0.062 | 4.91E-10 | 0.138 | 4.72E-08 | -0.033 | 6.81E-13 | PLXNB1 | 3p21.31 | known |
| AN | Fluid intelligence score | rs73074378 | 3 | 48446237 | C | T | 0.056 | 2.54E-08 | 0.138 | 4.72E-08 | 0.017 | 3.24E-02 | PLXNB1 | 3p21.31 | known |
| AN | Creatinine (quantile) | rs73074378 | 3 | 48446237 | C | T | 0.057 | 1.72E-08 | 0.138 | 4.72E-08 | -0.016 | 7.80E-06 | PLXNB1 | 3p21.31 | known |

| | | | | | | | | | | | | | | | |
|---|---|---|---|---|---|---|---|---|---|---|---|---|---|---|---|
| AN | C-reactive protein (quantile) | rs73074378 | 3 | 48446237 | C | T | 0.054 | 2.91E-08 | 0.138 | 4.72E-08 | -0.009 | 4.41E-02 | PLXNB1 | 3p21.31 | known |
| AN | Basophill count | rs73074378 | 3 | 48446237 | C | T | 0.056 | 3.89E-08 | 0.138 | 4.72E-08 | -0.008 | 2.65E-02 | PLXNB1 | 3p21.31 | known |
| AN | Sex hormone-binding globulin levels adjusted for BMI | rs4858828 | 3 | 48687378 | C | T | -0.032 | 7.54E-09 | -0.076 | 6.80E-08 | 0.006 | 3.60E-13 | NCKIPSD | 3p21.31 | known |
| AN | Total protein (quantile) | rs9873726 | 3 | 48712367 | C | T | 0.064 | 1.55E-14 | 0.156 | 7.88E-15 | -0.010 | 1.35E-02 | NCKIPSD | 3p21.31 | known |
| AN | Sleep duration | rs9873726 | 3 | 48712367 | C | T | 0.065 | 4.97E-15 | 0.156 | 7.88E-15 | -0.006 | 2.52E-02 | NCKIPSD | 3p21.31 | known |
| AN | Sex hormone-binding globulin levels | rs9873726 | 3 | 48712367 | C | T | 0.062 | 5.57E-15 | 0.156 | 7.88E-15 | -0.004 | 7.10E-03 | NCKIPSD | 3p21.31 | known |
| AN | Neutrophill count | rs9873726 | 3 | 48712367 | C | T | 0.065 | 3.97E-15 | 0.156 | 7.88E-15 | -0.011 | 4.58E-03 | NCKIPSD | 3p21.31 | known |
| AN | Haemoglobin concentration | rs9873726 | 3 | 48712367 | C | T | 0.064 | 1.49E-14 | 0.156 | 7.88E-15 | -0.007 | 2.82E-02 | NCKIPSD | 3p21.31 | known |
| AN | Glycated hemoglobin levels（HbA2c） | rs9873726 | 3 | 48712367 | C | T | 0.056 | 1.32E-12 | 0.156 | 7.88E-15 | 0.007 | 0.003 | NCKIPSD | 3p21.31 | known |
| AN | Fluid intelligence score | rs9873726 | 3 | 48712367 | C | T | 0.065 | 2.57E-15 | 0.156 | 7.88E-15 | 0.018 | 6.03E-03 | NCKIPSD | 3p21.31 | known |
| AN | Creatinine (quantile) | rs9873726 | 3 | 48712367 | C | T | 0.067 | 1.00E-15 | 0.156 | 7.88E-15 | -0.019 | 5.94E-10 | NCKIPSD | 3p21.31 | known |
| AN | C-reactive protein (quantile) | rs9873726 | 3 | 48712367 | C | T | 0.062 | 5.37E-15 | 0.156 | 7.88E-15 | -0.009 | 1.51E-02 | NCKIPSD | 3p21.31 | known |
| AN | circulating leptin levels adjusted for BMI | rs9873726 | 3 | 48712367 | C | T | 0.065 | 6.58E-15 | 0.156 | 7.88E-15 | -0.016 | 0.043 | NCKIPSD | 3p21.31 | known |
| AN | Bioavailable testosterone levels | rs9873726 | 3 | 48712367 | C | T | 0.066 | 2.41E-15 | 0.156 | 7.88E-15 | 0.009 | 3.00E-03 | NCKIPSD | 3p21.31 | known |
| AN | Basophill count | rs9873726 | 3 | 48712367 | C | T | 0.065 | 7.08E-15 | 0.156 | 7.88E-15 | -0.007 | 2.79E-02 | NCKIPSD | 3p21.31 | known |
| AN | White blood cell (leukocyte) count | rs9809222 | 3 | 48714411 | C | T | 0.064 | 9.59E-15 | 0.156 | 8.86E-15 | -0.007 | 4.67E-02 | NCKIPSD | 3p21.31 | known |
| AN | Reticulocyte percentage | rs9812977 | 3 | 48720303 | G | A | 0.074 | 5.78E-18 | 0.165 | 8.57E-15 | -0.038 | 8.41E-22 | NCKIPSD | 3p21.31 | known |
| AN | Reticulocyte count | rs9812977 | 3 | 48720303 | G | A | 0.074 | 5.63E-18 | 0.165 | 8.57E-15 | -0.036 | 5.02E-20 | NCKIPSD | 3p21.31 | known |
| AN | Red blood cell (erythrocyte) distribution width | rs9812977 | 3 | 48720303 | G | A | 0.068 | 2.06E-15 | 0.165 | 8.57E-15 | -0.015 | 1.20E-04 | NCKIPSD | 3p21.31 | known |
| AN | Number of cigarettes previously smoked daily | rs9812977 | 3 | 48720303 | G | A | 0.064 | 1.23E-13 | 0.165 | 8.57E-15 | 0.018 | 7.38E-03 | NCKIPSD | 3p21.31 | known |
| AN | Mean reticulocyte volume | rs9812977 | 3 | 48720303 | G | A | 0.067 | 1.54E-14 | 0.165 | 8.57E-15 | 0.009 | 1.95E-02 | NCKIPSD | 3p21.31 | known |
| AN | Mean corpuscular volume | rs9812977 | 3 | 48720303 | G | A | 0.063 | 3.51E-13 | 0.165 | 8.57E-15 | -0.012 | 2.28E-03 | NCKIPSD | 3p21.31 | known |
| AN | Mean corpuscular haemoglobin | rs9812977 | 3 | 48720303 | G | A | 0.062 | 4.91E-13 | 0.165 | 8.57E-15 | -0.014 | 2.93E-04 | NCKIPSD | 3p21.31 | known |
| AN | Immature reticulocyte fraction | rs9812977 | 3 | 48720303 | G | A | 0.072 | 5.58E-17 | 0.165 | 8.57E-15 | -0.024 | 9.51E-10 | NCKIPSD | 3p21.31 | known |
| AN | High light scatter reticulocyte percentage | rs9812977 | 3 | 48720303 | G | A | 0.075 | 1.88E-18 | 0.165 | 8.57E-15 | -0.039 | 1.02E-22 | NCKIPSD | 3p21.31 | known |
| AN | High light scatter reticulocyte count | rs9812977 | 3 | 48720303 | G | A | 0.075 | 1.80E-18 | 0.165 | 8.57E-15 | -0.038 | 1.48E-21 | NCKIPSD | 3p21.31 | known |
| AN | SHBG (quantile) | rs4858828 | 3 | 48724811 | C | T | -0.032 | 1.06E-09 | -0.076 | 6.80E-08 | -0.011 | 2.89E-06 | NCKIPSD | 3p21.31 | known |
| AN | Red blood cell (erythrocyte) count | rs4858828 | 3 | 48724811 | C | T | -0.030 | 4.02E-08 | -0.076 | 6.80E-08 | 0.006 | 9.31E-03 | NCKIPSD | 3p21.31 | known |

| | | | | | | | | | | | | | | | |
|---|---|---|---|---|---|---|---|---|---|---|---|---|---|---|---|
| AN | Systolic blood pressure, automated reading | rs11557629 | 3 | 48732223 | G | A | 0.052 | 6.12E-11 | 0.140 | 1.59E-12 | 0.007 | 4.97E-02 | NCKIPSD | 3p21.31 | known |
| AN | Haematocrit percentage | rs11719291 | 3 | 48735706 | A | G | 0.057 | 1.87E-12 | 0.141 | 1.31E-12 | -0.006 | 2.96E-02 | CAMKV | 3p21.31 | known |
| AN | High light scatter reticulocyte percentage | rs1961527 | 3 | 48764954 | A | G | -0.042 | 2.96E-08 | -0.063 | 2.49E-03 | 0.059 | 5.03E-77 | IP6K2 | 3p21.31 | known |
| AN | High light scatter reticulocyte count | rs1961527 | 3 | 48764954 | A | G | -0.043 | 1.39E-08 | -0.063 | 2.49E-03 | 0.059 | 7.91E-78 | IP6K2 | 3p21.31 | known |
| AN | Age completed full time education | rs56735597 | 3 | 48765168 | A | G | 0.056 | 5.85E-12 | 0.138 | 1.26E-11 | 0.008 | 2.67E-02 | IP6K2 | 3p21.31 | known |
| AN | Eosinophill count | rs13094900 | 3 | 48837732 | G | A | 0.060 | 3.24E-12 | 0.148 | 1.85E-12 | -0.006 | 3.74E-02 | PRKAR2A | 3p21.31 | known |
| AN | Age started smoking in former smokers | rs71324924 | 3 | 48865444 | G | A | 0.055 | 4.95E-11 | 0.148 | 1.50E-12 | -0.015 | 4.68E-02 | PRKAR2A | 3p21.31 | known |
| AN | Alanine aminotransferase (quantile) | rs67332931 | 3 | 48866548 | C | T | 0.055 | 5.37E-11 | 0.148 | 1.47E-12 | 0.007 | 4.81E-02 | PRKAR2A | 3p21.31 | known |
| AN | Urate (quantile) | rs55674449 | 3 | 48940875 | G | A | 0.040 | 9.75E-09 | 0.104 | 2.15E-08 | -0.006 | 2.96E-02 | SLC25A20 | 3p21.31 | known |
| AN | Sex hormone-binding globulin levels adjusted for BMI | rs4513485 | 3 | 49102533 | G | A | -0.033 | 5.17E-09 | -0.077 | 4.67E-08 | 0.006 | 4.80E-13 | QARS1 | 3p21.31 | known |
| AN | White blood cell (leukocyte) count | rs73082362 | 3 | 49113729 | C | T | 0.069 | 5.00E-14 | 0.173 | 4.27E-14 | -0.008 | 4.41E-02 | QRICH1 | 3p21.31 | known |
| AN | Total protein (quantile) | rs73082362 | 3 | 49113729 | C | T | 0.068 | 8.91E-14 | 0.173 | 4.27E-14 | -0.015 | 5.22E-04 | QRICH1 | 3p21.31 | known |
| AN | Sleep duration | rs73082362 | 3 | 49113729 | C | T | 0.070 | 2.15E-14 | 0.173 | 4.27E-14 | -0.009 | 6.08E-03 | QRICH1 | 3p21.31 | known |
| AN | Sex hormone-binding globulin levels | rs73082362 | 3 | 49113729 | C | T | 0.065 | 5.10E-14 | 0.173 | 4.27E-14 | -0.004 | 3.10E-02 | QRICH1 | 3p21.31 | known |
| AN | Reticulocyte percentage | rs73082362 | 3 | 49113729 | C | T | 0.077 | 1.87E-17 | 0.173 | 4.27E-14 | -0.043 | 4.63E-25 | QRICH1 | 3p21.31 | known |
| AN | Reticulocyte count | rs73082362 | 3 | 49113729 | C | T | 0.076 | 1.95E-17 | 0.173 | 4.27E-14 | -0.040 | 1.26E-22 | QRICH1 | 3p21.31 | known |
| AN | Red blood cell (erythrocyte) distribution width | rs73082362 | 3 | 49113729 | C | T | 0.071 | 6.11E-15 | 0.173 | 4.27E-14 | -0.020 | 2.04E-06 | QRICH1 | 3p21.31 | known |
| AN | Number of cigarettes previously smoked daily | rs73082362 | 3 | 49113729 | C | T | 0.065 | 5.99E-13 | 0.173 | 4.27E-14 | 0.018 | 1.02E-02 | QRICH1 | 3p21.31 | known |
| AN | Neutrophill count | rs73082362 | 3 | 49113729 | C | T | 0.069 | 2.41E-14 | 0.173 | 4.27E-14 | -0.012 | 4.80E-03 | QRICH1 | 3p21.31 | known |
| AN | Mean corpuscular volume | rs73082362 | 3 | 49113729 | C | T | 0.062 | 6.03E-12 | 0.173 | 4.27E-14 | -0.020 | 1.41E-06 | QRICH1 | 3p21.31 | known |
| AN | Mean corpuscular haemoglobin | rs73082362 | 3 | 49113729 | C | T | 0.062 | 7.01E-12 | 0.173 | 4.27E-14 | -0.021 | 2.02E-07 | QRICH1 | 3p21.31 | known |
| AN | Immature reticulocyte fraction | rs73082362 | 3 | 49113729 | C | T | 0.075 | 5.24E-17 | 0.173 | 4.27E-14 | -0.032 | 1.15E-14 | QRICH1 | 3p21.31 | known |
| AN | High light scatter reticulocyte percentage | rs73082362 | 3 | 49113729 | C | T | 0.078 | 3.09E-18 | 0.173 | 4.27E-14 | -0.046 | 1.56E-28 | QRICH1 | 3p21.31 | known |
| AN | High light scatter reticulocyte count | rs73082362 | 3 | 49113729 | C | T | 0.078 | 2.89E-18 | 0.173 | 4.27E-14 | -0.045 | 5.05E-27 | QRICH1 | 3p21.31 | known |
| AN | Haemoglobin concentration | rs73082362 | 3 | 49113729 | C | T | 0.068 | 8.75E-14 | 0.173 | 4.27E-14 | -0.007 | 2.55E-02 | QRICH1 | 3p21.31 | known |
| AN | Glycated hemoglobin levels（HbA3c） | rs73082362 | 3 | 49113729 | C | T | 0.060 | 2.12E-11 | 0.173 | 4.27E-14 | 0.010 | 4.59E-05 | QRICH1 | 3p21.31 | known |
| AN | Fluid intelligence score | rs73082362 | 3 | 49113729 | C | T | 0.069 | 1.65E-14 | 0.173 | 4.27E-14 | 0.019 | 7.26E-03 | QRICH1 | 3p21.31 | known |
| AN | Eosinophill count | rs73082362 | 3 | 49113729 | C | T | 0.068 | 9.88E-14 | 0.173 | 4.27E-14 | -0.006 | 4.83E-02 | QRICH1 | 3p21.31 | known |

| | | | | | | | | | | | | | | | |
|---|---|---|---|---|---|---|---|---|---|---|---|---|---|---|---|
| AN | Creatinine (quantile) | rs73082362 | 3 | 49113729 | C | T | 0.071 | 5.01E-15 | 0.173 | 4.27E-14 | -0.022 | 2.90E-11 | QRICH1 | 3p21.31 | known |
| AN | C-reactive protein (quantile) | rs73082362 | 3 | 49113729 | C | T | 0.068 | 7.68E-15 | 0.173 | 4.27E-14 | -0.014 | 1.02E-03 | QRICH1 | 3p21.31 | known |
| AN | Basophill count | rs73082362 | 3 | 49113729 | C | T | 0.069 | 3.20E-14 | 0.173 | 4.27E-14 | -0.010 | 4.04E-03 | QRICH1 | 3p21.31 | known |
| AN | Bioavailable testosterone levels | rs34326553 | 3 | 49134429 | A | G | 0.062 | 3.12E-13 | 0.152 | 4.04E-13 | 0.006 | 4.90E-02 | QRICH1 | 3p21.31 | known |
| AN | Alanine aminotransferase (quantile) | rs34326553 | 3 | 49134429 | A | G | 0.056 | 1.71E-11 | 0.152 | 4.04E-13 | 0.007 | 4.64E-02 | QRICH1 | 3p21.31 | known |
| AN | Testosterone (quantile) | rs4521268 | 3 | 49137904 | G | T | -0.031 | 3.77E-08 | -0.076 | 6.35E-08 | -0.003 | 3.38E-02 | QARS1 | 3p21.31 | known |
| AN | Age started smoking in former smokers | rs35673421 | 3 | 49139406 | G | A | 0.057 | 1.44E-11 | 0.151 | 4.50E-13 | -0.015 | 4.98E-02 | QARS1 | 3p21.31 | known |
| AN | Red blood cell (erythrocyte) count | rs4513485 | 3 | 49139966 | G | A | -0.031 | 2.39E-08 | -0.077 | 4.67E-08 | 0.006 | 4.40E-03 | QARS1 | 3p21.31 | known |
| AN | SHBG (quantile) | rs61583136 | 3 | 49189195 | C | T | -0.033 | 6.61E-10 | -0.077 | 4.52E-08 | -0.011 | 2.21E-06 | DHFR2 | 3p21.31 | known |
| AN | Urate (quantile) | rs1134043 | 3 | 49200137 | G | T | 0.042 | 1.78E-09 | 0.108 | 4.83E-09 | -0.007 | 1.63E-02 | CCDC71 | 3p21.31 | known |
| AN | Amount of alcohol drunk on a typical drinking day | rs112890859 | 3 | 49252583 | A | G | 0.067 | 1.49E-13 | 0.167 | 2.02E-13 | -0.015 | 4.53E-02 | IHO1 | 3p21.31 | known |
| AN | Gamma glutamyltransferase (quantile) | rs150429489 | 3 | 49319336 | C | T | 0.057 | 2.21E-11 | 0.149 | 1.12E-12 | 0.008 | 4.07E-02 | C3orf62 | 3p21.31 | known |
| AN | Systolic blood pressure, automated reading | rs9870030 | 3 | 49332709 | G | A | 0.055 | 1.17E-10 | 0.150 | 2.99E-12 | 0.007 | 4.91E-02 | C3orf62 | 3p21.31 | known |
| AN | Sex hormone-binding globulin levels | rs11716445 | 3 | 49368662 | G | A | 0.064 | 2.27E-13 | 0.165 | 3.10E-13 | -0.004 | 1.60E-02 | GPX1 | 3p21.31 | known |
| AN | Sex hormone-binding globulin levels adjusted for BMI | rs73088112 | 3 | 49375919 | T | C | 0.121 | 2.75E-08 | 0.331 | 2.87E-08 | -0.005 | 3.10E-02 | GPX1 | 3p21.31 | known |
| AN | White blood cell (leukocyte) count | rs11716445 | 3 | 49406095 | G | A | 0.066 | 3.22E-13 | 0.165 | 3.10E-13 | -0.009 | 2.91E-02 | GPX1 | 3p21.31 | known |
| AN | Total protein (quantile) | rs11716445 | 3 | 49406095 | G | A | 0.066 | 6.10E-13 | 0.165 | 3.10E-13 | -0.015 | 4.43E-04 | GPX1 | 3p21.31 | known |
| AN | Reticulocyte percentage | rs11716445 | 3 | 49406095 | G | A | 0.075 | 1.19E-16 | 0.165 | 3.10E-13 | -0.044 | 1.49E-26 | GPX1 | 3p21.31 | known |
| AN | Reticulocyte count | rs11716445 | 3 | 49406095 | G | A | 0.074 | 1.20E-16 | 0.165 | 3.10E-13 | -0.041 | 4.62E-24 | GPX1 | 3p21.31 | known |
| AN | Number of cigarettes previously smoked daily | rs11716445 | 3 | 49406095 | G | A | 0.063 | 3.52E-12 | 0.165 | 3.10E-13 | 0.017 | 1.52E-02 | GPX1 | 3p21.31 | known |
| AN | Neutrophill count | rs11716445 | 3 | 49406095 | G | A | 0.067 | 1.55E-13 | 0.165 | 3.10E-13 | -0.012 | 2.99E-03 | GPX1 | 3p21.31 | known |
| AN | Mean corpuscular volume | rs11716445 | 3 | 49406095 | G | A | 0.060 | 2.96E-11 | 0.165 | 3.10E-13 | -0.019 | 4.70E-06 | GPX1 | 3p21.31 | known |
| AN | Mean corpuscular haemoglobin | rs11716445 | 3 | 49406095 | G | A | 0.060 | 3.75E-11 | 0.165 | 3.10E-13 | -0.021 | 3.77E-07 | GPX1 | 3p21.31 | known |
| AN | Immature reticulocyte fraction | rs11716445 | 3 | 49406095 | G | A | 0.073 | 3.45E-16 | 0.165 | 3.10E-13 | -0.033 | 2.35E-15 | GPX1 | 3p21.31 | known |
| AN | High light scatter reticulocyte percentage | rs11716445 | 3 | 49406095 | G | A | 0.076 | 1.94E-17 | 0.165 | 3.10E-13 | -0.047 | 3.69E-30 | GPX1 | 3p21.31 | known |
| AN | High light scatter reticulocyte count | rs11716445 | 3 | 49406095 | G | A | 0.076 | 1.79E-17 | 0.165 | 3.10E-13 | -0.046 | 1.49E-28 | GPX1 | 3p21.31 | known |
| AN | Haemoglobin concentration | rs11716445 | 3 | 49406095 | G | A | 0.066 | 6.00E-13 | 0.165 | 3.10E-13 | -0.007 | 2.55E-02 | GPX1 | 3p21.31 | known |
| AN | Eosinophill count | rs11716445 | 3 | 49406095 | G | A | 0.066 | 6.76E-13 | 0.165 | 3.10E-13 | -0.006 | 4.28E-02 | GPX1 | 3p21.31 | known |

| | | | | | | | | | | | | | | | |
|---|---|---|---|---|---|---|---|---|---|---|---|---|---|---|---|
| AN | Creatinine (quantile) | rs11716445 | 3 | 49406095 | G | A | 0.069 | 3.32E-14 | 0.165 | 3.10E-13 | -0.023 | 5.55E-12 | GPX1 | 3p21.31 | known |
| AN | C-reactive protein (quantile) | rs11716445 | 3 | 49406095 | G | A | 0.066 | 4.57E-14 | 0.165 | 3.10E-13 | -0.014 | 7.55E-04 | GPX1 | 3p21.31 | known |
| AN | Basophill count | rs11716445 | 3 | 49406095 | G | A | 0.067 | 2.02E-13 | 0.165 | 3.10E-13 | -0.011 | 1.57E-03 | GPX1 | 3p21.31 | known |
| AN | Diastolic blood pressure, automated reading | rs6797765 | 3 | 49423976 | A | G | -0.029 | 3.43E-08 | -0.066 | 1.15E-06 | 0.013 | 2.78E-08 | TCTA | 3p21.31 | known |
| AN | Glucose (quantile) | rs183270497 | 3 | 49479704 | C | T | 0.058 | 7.19E-11 | 0.171 | 1.32E-12 | 0.009 | 4.15E-02 | NICN1 | 3p21.31 | known |
| AN | Urate (quantile) | rs7645445 | 3 | 49502243 | C | T | -0.029 | 1.92E-08 | -0.066 | 1.06E-06 | 0.011 | 1.81E-07 | DAG1 | 3p21.31 | known |
| AN | SHBG (quantile) | rs7645445 | 3 | 49502243 | C | T | -0.033 | 3.66E-11 | -0.066 | 1.06E-06 | -0.021 | 1.07E-19 | DAG1 | 3p21.31 | known |
| AN | Sex hormone-binding globulin levels adjusted for BMI | rs7645445 | 3 | 49502243 | C | T | -0.029 | 4.60E-08 | -0.066 | 1.06E-06 | 0.008 | 4.00E-21 | DAG1 | 3p21.31 | known |
| AN | Sex hormone-binding globulin levels | rs7645445 | 3 | 49502243 | C | T | -0.033 | 5.24E-11 | -0.066 | 1.06E-06 | 0.010 | 8.20E-26 | DAG1 | 3p21.31 | known |
| AN | Reticulocyte percentage | rs7645445 | 3 | 49502243 | C | T | -0.031 | 3.84E-09 | -0.066 | 1.06E-06 | 0.025 | 4.04E-24 | DAG1 | 3p21.31 | known |
| AN | Reticulocyte count | rs7645445 | 3 | 49502243 | C | T | -0.032 | 1.12E-09 | -0.066 | 1.06E-06 | 0.027 | 2.89E-29 | DAG1 | 3p21.31 | known |
| AN | Red blood cell (erythrocyte) distribution width | rs7645445 | 3 | 49502243 | C | T | -0.029 | 3.29E-08 | -0.066 | 1.06E-06 | 0.020 | 2.33E-16 | DAG1 | 3p21.31 | known |
| AN | Immature reticulocyte fraction | rs7645445 | 3 | 49502243 | C | T | -0.031 | 2.74E-09 | -0.066 | 1.06E-06 | 0.021 | 2.89E-18 | DAG1 | 3p21.31 | known |
| AN | High light scatter reticulocyte percentage | rs7645445 | 3 | 49502243 | C | T | -0.032 | 6.81E-10 | -0.066 | 1.06E-06 | 0.028 | 6.47E-30 | DAG1 | 3p21.31 | known |
| AN | High light scatter reticulocyte count | rs7645445 | 3 | 49502243 | C | T | -0.033 | 2.25E-10 | -0.066 | 1.06E-06 | 0.029 | 2.42E-34 | DAG1 | 3p21.31 | known |
| AN | C-reactive protein (quantile) | rs7645445 | 3 | 49502243 | C | T | -0.031 | 1.37E-09 | -0.066 | 1.06E-06 | 0.017 | 7.24E-13 | DAG1 | 3p21.31 | known |
| AN | Sleep duration | rs73073015 | 3 | 49503166 | G | A | 0.067 | 1.41E-13 | 0.173 | 3.24E-13 | -0.009 | 3.48E-03 | DAG1 | 3p21.31 | known |
| AN | Red blood cell (erythrocyte) distribution width | rs73073015 | 3 | 49503166 | G | A | 0.068 | 3.97E-14 | 0.173 | 3.24E-13 | -0.020 | 6.95E-07 | DAG1 | 3p21.31 | known |
| AN | Glycated hemoglobin levels（HbA4c） | rs73073015 | 3 | 49503166 | G | A | 0.057 | 6.21E-11 | 0.173 | 3.24E-13 | 0.009 | 3.73E-04 | DAG1 | 3p21.31 | known |
| AN | Fluid intelligence score | rs73073015 | 3 | 49503166 | G | A | 0.067 | 9.66E-14 | 0.173 | 3.24E-13 | 0.021 | 2.86E-03 | DAG1 | 3p21.31 | known |
| AN | Testosterone (quantile) | rs56116382 | 3 | 49606188 | A | C | 0.038 | 3.06E-08 | 0.092 | 7.13E-08 | 0.005 | 1.83E-02 | BSN | 3p21.31 | known |
| AN | Ever taken cannabis | rs56116382 | 3 | 49606188 | A | C | 0.039 | 1.11E-08 | 0.092 | 7.13E-08 | 0.012 | 1.23E-02 | BSN | 3p21.31 | known |
| AN | Diastolic blood pressure, automated reading | rs9836291 | 3 | 49697459 | G | A | 0.033 | 1.10E-08 | 0.072 | 6.37E-07 | -0.016 | 3.48E-10 | APEH | 3p21.31 | known |
| AN | Age completed full time education | rs9836291 | 3 | 49697459 | G | A | 0.033 | 1.12E-08 | 0.072 | 6.37E-07 | 0.018 | 2.40E-13 | APEH | 3p21.31 | known |
| AN | Urate (quantile) | rs34823813 | 3 | 49749976 | G | A | 0.059 | 3.49E-12 | 0.151 | 4.04E-12 | -0.007 | 5.00E-02 | AMIGO3 | 3p21.31 | known |
| AN | Systolic blood pressure, automated reading | rs34823813 | 3 | 49749976 | G | A | 0.055 | 1.59E-10 | 0.151 | 4.04E-12 | 0.008 | 4.10E-02 | AMIGO3 | 3p21.31 | known |
| AN | Glucose (quantile) | rs34823813 | 3 | 49749976 | G | A | 0.054 | 2.82E-10 | 0.151 | 4.04E-12 | 0.010 | 1.32E-02 | AMIGO3 | 3p21.31 | known |
| AN | Age started smoking in former smokers | rs34823813 | 3 | 49749976 | G | A | 0.056 | 1.12E-10 | 0.151 | 4.04E-12 | -0.016 | 4.16E-02 | AMIGO3 | 3p21.31 | known |

| | | | | | | | | | | | | | | | |
|---|---|---|---|---|---|---|---|---|---|---|---|---|---|---|---|
| AN | White blood cell (leukocyte) count | rs73077200 | 3 | 49826987 | C | T | 0.067 | 2.45E-13 | 0.169 | 2.77E-13 | -0.009 | 3.09E-02 | IP6K1 | 3p21.31 | known |
| AN | Total protein (quantile) | rs73077200 | 3 | 49826987 | C | T | 0.066 | 4.61E-13 | 0.169 | 2.77E-13 | -0.015 | 4.21E-04 | IP6K1 | 3p21.31 | known |
| AN | Sleep duration | rs73077200 | 3 | 49826987 | C | T | 0.067 | 1.20E-13 | 0.169 | 2.77E-13 | -0.008 | 7.40E-03 | IP6K1 | 3p21.31 | known |
| AN | Sex hormone-binding globulin levels | rs73077200 | 3 | 49826987 | C | T | 0.064 | 1.67E-13 | 0.169 | 2.77E-13 | -0.004 | 1.50E-02 | IP6K1 | 3p21.31 | known |
| AN | Reticulocyte percentage | rs73077200 | 3 | 49826987 | C | T | 0.075 | 8.38E-17 | 0.169 | 2.77E-13 | -0.044 | 1.04E-26 | IP6K1 | 3p21.31 | known |
| AN | Reticulocyte count | rs73077200 | 3 | 49826987 | C | T | 0.075 | 8.11E-17 | 0.169 | 2.77E-13 | -0.042 | 2.16E-24 | IP6K1 | 3p21.31 | known |
| AN | Red blood cell (erythrocyte) distribution width | rs73077200 | 3 | 49826987 | C | T | 0.068 | 3.42E-14 | 0.169 | 2.77E-13 | -0.019 | 2.41E-06 | IP6K1 | 3p21.31 | known |
| AN | Number of cigarettes previously smoked daily | rs73077200 | 3 | 49826987 | C | T | 0.063 | 2.62E-12 | 0.169 | 2.77E-13 | 0.017 | 1.71E-02 | IP6K1 | 3p21.31 | known |
| AN | Neutrophill count | rs73077200 | 3 | 49826987 | C | T | 0.067 | 1.25E-13 | 0.169 | 2.77E-13 | -0.012 | 4.24E-03 | IP6K1 | 3p21.31 | known |
| AN | Immature reticulocyte fraction | rs73077200 | 3 | 49826987 | C | T | 0.073 | 2.56E-16 | 0.169 | 2.77E-13 | -0.033 | 2.53E-15 | IP6K1 | 3p21.31 | known |
| AN | High light scatter reticulocyte percentage | rs73077200 | 3 | 49826987 | C | T | 0.076 | 1.36E-17 | 0.169 | 2.77E-13 | -0.047 | 2.70E-30 | IP6K1 | 3p21.31 | known |
| AN | High light scatter reticulocyte count | rs73077200 | 3 | 49826987 | C | T | 0.076 | 1.22E-17 | 0.169 | 2.77E-13 | -0.046 | 8.32E-29 | IP6K1 | 3p21.31 | known |
| AN | Haemoglobin concentration | rs73077200 | 3 | 49826987 | C | T | 0.066 | 4.53E-13 | 0.169 | 2.77E-13 | -0.007 | 2.39E-02 | IP6K1 | 3p21.31 | known |
| AN | Glycated hemoglobin levels（HbA5c） | rs73077200 | 3 | 49826987 | C | T | 0.058 | 6.68E-11 | 0.169 | 2.77E-13 | 0.011 | 1.13E-04 | IP6K1 | 3p21.31 | known |
| AN | Fluid intelligence score | rs73077200 | 3 | 49826987 | C | T | 0.068 | 6.64E-14 | 0.169 | 2.77E-13 | 0.021 | 2.29E-03 | IP6K1 | 3p21.31 | known |
| AN | Eosinophill count | rs73077200 | 3 | 49826987 | C | T | 0.066 | 5.11E-13 | 0.169 | 2.77E-13 | -0.006 | 3.89E-02 | IP6K1 | 3p21.31 | known |
| AN | Creatinine (quantile) | rs73077200 | 3 | 49826987 | C | T | 0.069 | 2.59E-14 | 0.169 | 2.77E-13 | -0.023 | 1.06E-11 | IP6K1 | 3p21.31 | known |
| AN | C-reactive protein (quantile) | rs73077200 | 3 | 49826987 | C | T | 0.066 | 4.33E-14 | 0.169 | 2.77E-13 | -0.013 | 1.26E-03 | IP6K1 | 3p21.31 | known |
| AN | Basophill count | rs73077200 | 3 | 49826987 | C | T | 0.067 | 1.51E-13 | 0.169 | 2.77E-13 | -0.011 | 1.54E-03 | IP6K1 | 3p21.31 | known |
| AN | SHBG (quantile) | rs34735338 | 3 | 49831698 | G | A | -0.033 | 1.22E-09 | -0.059 | 4.96E-05 | -0.024 | 1.03E-23 | CDHR4 | 3p21.31 | known |
| AN | Sex hormone-binding globulin levels | rs62260729 | 3 | 49852486 | T | C | -0.032 | 2.38E-09 | -0.061 | 3.67E-05 | 0.011 | 1.00E-28 | TRAIP | 3p21.31 | known |
| AN | C-reactive protein (quantile) | rs2681780 | 3 | 49897830 | C | T | -0.030 | 3.61E-09 | -0.059 | 1.52E-05 | 0.021 | 5.61E-19 | TRAIP | 3p21.31 | known |
| AN | SHBG (quantile) | rs11716961 | 3 | 49908338 | G | A | 0.062 | 3.69E-13 | 0.169 | 3.78E-13 | 0.008 | 3.15E-02 | CAMKV | 3p21.31 | known |
| AN | Mean corpuscular volume | rs11716961 | 3 | 49908338 | G | A | 0.060 | 1.84E-11 | 0.169 | 3.78E-13 | -0.015 | 1.45E-04 | CAMKV | 3p21.31 | known |
| AN | Mean corpuscular haemoglobin | rs11716961 | 3 | 49908338 | G | A | 0.060 | 2.35E-11 | 0.169 | 3.78E-13 | -0.017 | 1.80E-05 | CAMKV | 3p21.31 | known |
| AN | Age started smoking in former smokers | rs4688759 | 3 | 50008118 | T | C | 0.053 | 6.74E-10 | 0.146 | 3.17E-11 | -0.015 | 4.55E-02 | RBM6 | 3p21.31 | known |
| AN | Haemoglobin concentration | rs2023953 | 3 | 50103917 | G | A | 0.056 | 6.21E-11 | 0.146 | 4.08E-11 | -0.007 | 1.94E-02 | RBM5 | 3p21.31 | known |
| AN | SHBG (quantile) | rs2071206 | 3 | 50160109 | G | A | 0.029 | 8.73E-09 | 0.058 | 1.95E-05 | 0.018 | 4.59E-15 | SEMA3F | 3p21.31 | known |

| | | | | | | | | | | | | | | | |
|---|---|---|---|---|---|---|---|---|---|---|---|---|---|---|---|
| AN | C-reactive protein (quantile) | rs2008877 | 3 | 50162291 | T | G | 0.028 | 3.54E-08 | 0.055 | 4.05E-05 | -0.019 | 4.60E-16 | SEMA3F | 3p21.31 | known |
| AN | White blood cell (leukocyte) count | rs73080977 | 3 | 50189531 | G | T | 0.067 | 1.03E-12 | 0.175 | 1.31E-12 | -0.010 | 1.47E-02 | SEMA3F | 3p21.31 | known |
| AN | Total protein (quantile) | rs73080977 | 3 | 50189531 | G | T | 0.067 | 2.12E-12 | 0.175 | 1.31E-12 | -0.019 | 2.77E-05 | SEMA3F | 3p21.31 | known |
| AN | Sleep duration | rs73080977 | 3 | 50189531 | G | T | 0.068 | 5.69E-13 | 0.175 | 1.31E-12 | -0.009 | 7.03E-03 | SEMA3F | 3p21.31 | known |
| AN | Sex hormone-binding globulin levels | rs73080977 | 3 | 50189531 | G | T | 0.064 | 8.16E-13 | 0.175 | 1.31E-12 | -0.004 | 1.10E-02 | SEMA3F | 3p21.31 | known |
| AN | Reticulocyte percentage | rs73080977 | 3 | 50189531 | G | T | 0.076 | 4.48E-16 | 0.175 | 1.31E-12 | -0.046 | 4.88E-27 | SEMA3F | 3p21.31 | known |
| AN | Reticulocyte count | rs73080977 | 3 | 50189531 | G | T | 0.076 | 3.89E-16 | 0.175 | 1.31E-12 | -0.044 | 3.76E-25 | SEMA3F | 3p21.31 | known |
| AN | Red blood cell (erythrocyte) distribution width | rs73080977 | 3 | 50189531 | G | T | 0.069 | 2.90E-13 | 0.175 | 1.31E-12 | -0.017 | 8.35E-05 | SEMA3F | 3p21.31 | known |
| AN | Number of cigarettes previously smoked daily | rs73080977 | 3 | 50189531 | G | T | 0.064 | 9.90E-12 | 0.175 | 1.31E-12 | 0.016 | 3.42E-02 | SEMA3F | 3p21.31 | known |
| AN | Neutrophill count | rs73080977 | 3 | 50189531 | G | T | 0.068 | 6.26E-13 | 0.175 | 1.31E-12 | -0.012 | 5.42E-03 | SEMA3F | 3p21.31 | known |
| AN | Mean corpuscular volume | rs73080977 | 3 | 50189531 | G | T | 0.062 | 2.74E-11 | 0.175 | 1.31E-12 | -0.011 | 6.53E-03 | SEMA3F | 3p21.31 | known |
| AN | Mean corpuscular haemoglobin | rs73080977 | 3 | 50189531 | G | T | 0.062 | 3.86E-11 | 0.175 | 1.31E-12 | -0.014 | 8.59E-04 | SEMA3F | 3p21.31 | known |
| AN | Immature reticulocyte fraction | rs73080977 | 3 | 50189531 | G | T | 0.076 | 3.39E-16 | 0.175 | 1.31E-12 | -0.039 | 5.78E-20 | SEMA3F | 3p21.31 | known |
| AN | High light scatter reticulocyte percentage | rs73080977 | 3 | 50189531 | G | T | 0.078 | 4.87E-17 | 0.175 | 1.31E-12 | -0.051 | 1.17E-32 | SEMA3F | 3p21.31 | known |
| AN | High light scatter reticulocyte count | rs73080977 | 3 | 50189531 | G | T | 0.078 | 3.86E-17 | 0.175 | 1.31E-12 | -0.050 | 1.52E-31 | SEMA3F | 3p21.31 | known |
| AN | Glycated hemoglobin levels（HbA6c） | rs73080977 | 3 | 50189531 | G | T | 0.060 | 3.59E-10 | 0.175 | 1.31E-12 | 0.011 | 9.08E-06 | SEMA3F | 3p21.31 | known |
| AN | Fluid intelligence score | rs73080977 | 3 | 50189531 | G | T | 0.068 | 5.91E-13 | 0.175 | 1.31E-12 | 0.016 | 2.39E-02 | SEMA3F | 3p21.31 | known |
| AN | Eosinophill count | rs73080977 | 3 | 50189531 | G | T | 0.067 | 2.37E-12 | 0.175 | 1.31E-12 | -0.006 | 3.97E-02 | SEMA3F | 3p21.31 | known |
| AN | Creatinine (quantile) | rs73080977 | 3 | 50189531 | G | T | 0.070 | 1.43E-13 | 0.175 | 1.31E-12 | -0.023 | 4.49E-11 | SEMA3F | 3p21.31 | known |
| AN | C-reactive protein (quantile) | rs73080977 | 3 | 50189531 | G | T | 0.066 | 3.24E-13 | 0.175 | 1.31E-12 | -0.012 | 3.65E-03 | SEMA3F | 3p21.31 | known |
| AN | Basophill count | rs73080977 | 3 | 50189531 | G | T | 0.068 | 7.24E-13 | 0.175 | 1.31E-12 | -0.011 | 1.52E-03 | SEMA3F | 3p21.31 | known |
| AN | Testosterone (quantile) | rs7617915 | 3 | 50239012 | T | C | -0.052 | 2.56E-08 | -0.135 | 6.33E-08 | -0.007 | 1.13E-02 | SLC38A3 | 3p21.31 | known |
| AN | Reticulocyte percentage | rs1076872 | 3 | 50329826 | A | G | 0.035 | 1.82E-09 | 0.039 | 1.45E-02 | -0.081 | 4.32E-213 | IFRD2 | 3p21.31 | known |
| AN | Reticulocyte count | rs1076872 | 3 | 50329826 | A | G | 0.036 | 5.65E-10 | 0.039 | 1.45E-02 | -0.078 | 2.26E-206 | IFRD2 | 3p21.31 | known |
| AN | Immature reticulocyte fraction | rs1076872 | 3 | 50329826 | A | G | 0.039 | 2.31E-11 | 0.039 | 1.45E-02 | -0.077 | 2.21E-194 | IFRD2 | 3p21.31 | known |
| AN | High light scatter reticulocyte percentage | rs1076872 | 3 | 50329826 | A | G | 0.041 | 1.24E-12 | 0.039 | 1.45E-02 | -0.093 | 6.57E-283 | IFRD2 | 3p21.31 | known |
| AN | High light scatter reticulocyte count | rs1076872 | 3 | 50329826 | A | G | 0.042 | 3.08E-13 | 0.039 | 1.45E-02 | -0.092 | 1.91E-279 | IFRD2 | 3p21.31 | known |
| AN | Reticulocyte percentage | rs11715725 | 3 | 50519141 | T | C | 0.054 | 3.99E-09 | 0.121 | 1.06E-06 | -0.043 | 1.09E-23 | CACNA2D2 | 3p21.31 | known |

| | | | | | | | | | | | | | | | |
|---|---|---|---|---|---|---|---|---|---|---|---|---|---|---|---|
| AN | Reticulocyte count | rs11715725 | 3 | 50519141 | T | C | 0.054 | 3.70E-09 | 0.121 | 1.06E-06 | -0.040 | 9.29E-22 | CACNA2D2 | 3p21.31 | known |
| AN | Immature reticulocyte fraction | rs11715725 | 3 | 50519141 | T | C | 0.054 | 3.13E-09 | 0.121 | 1.06E-06 | -0.036 | 1.52E-17 | CACNA2D2 | 3p21.31 | known |
| AN | High light scatter reticulocyte percentage | rs11715725 | 3 | 50519141 | T | C | 0.056 | 7.95E-10 | 0.121 | 1.06E-06 | -0.047 | 8.64E-29 | CACNA2D2 | 3p21.31 | known |
| AN | High light scatter reticulocyte count | rs11715725 | 3 | 50519141 | T | C | 0.056 | 6.88E-10 | 0.121 | 1.06E-06 | -0.046 | 1.40E-27 | CACNA2D2 | 3p21.31 | known |
| AN | Fluid intelligence score | rs4629334 | 3 | 94581146 | G | A | -0.030 | 3.48E-08 | -0.077 | 9.64E-08 | -0.011 | 7.15E-03 | DHFR2 | 3q11.2 | known |
| AN | C-reactive protein (quantile) | rs4629334 | 3 | 94581146 | G | A | -0.029 | 4.72E-08 | -0.077 | 9.64E-08 | 0.005 | 3.36E-02 | DHFR2 | 3q11.2 | known |
| AN | Age completed full time education | rs4629334 | 3 | 94581146 | G | A | -0.030 | 3.24E-08 | -0.077 | 9.64E-08 | -0.006 | 8.49E-03 | DHFR2 | 3q11.2 | known |
| AN | Urate (quantile) | rs7619675 | 3 | 94593959 | A | C | -0.029 | 1.29E-08 | -0.075 | 5.23E-08 | 0.006 | 5.64E-03 | DHFR2 | 3q11.2 | known |
| AN | Systolic blood pressure, automated reading | rs7619675 | 3 | 94593959 | A | C | -0.029 | 2.34E-08 | -0.075 | 5.23E-08 | 0.005 | 3.54E-02 | DHFR2 | 3q11.2 | known |
| AN | Diastolic blood pressure, automated reading | rs7619675 | 3 | 94593959 | A | C | -0.030 | 1.05E-08 | -0.075 | 5.23E-08 | 0.007 | 1.91E-03 | DHFR2 | 3q11.2 | known |
| AN | Creatinine (quantile) | rs7619675 | 3 | 94593959 | A | C | -0.029 | 4.43E-08 | -0.075 | 5.23E-08 | 0.004 | 4.60E-02 | DHFR2 | 3q11.2 | known |
| AN | Age started smoking in former smokers | rs7619675 | 3 | 94593959 | A | C | -0.030 | 1.04E-08 | -0.075 | 5.23E-08 | -0.013 | 7.10E-03 | DHFR2 | 3q11.2 | known |
| AN | Sex hormone-binding globulin levels | rs7619675 | 3 | 94875115 | A | C | -0.028 | 3.37E-08 | -0.075 | 5.23E-08 | 0.002 | 2.90E-02 | DHFR2 | 3q11.2 | known |
| AN | Glucose (quantile) | rs35283189 | 3 | 49113668 | T | C | 0.056 | 3.37E-11 | 0.150 | 6.43E-13 | 0.008 | 4.97E-02 | QRICH1 | 3q22.3 | known |
| AN | SHBG (quantile) | rs34894639 | 3 | 135798658 | C | T | 0.042 | 1.65E-12 | 0.036 | 3.81E-02 | 0.061 | 7.48E-112 | PPP2R3A | 3q22.3 | new |
| AN | Sex hormone-binding globulin levels | rs34894639 | 3 | 135798658 | C | T | 0.043 | 5.65E-13 | 0.036 | 3.81E-02 | -0.030 | 2.20E-170 | PPP2R3A | 3q22.3 | new |
| AN | SHBG (quantile) | rs833752 | 3 | 136255169 | A | G | -0.038 | 2.06E-11 | -0.031 | 4.32E-02 | -0.055 | 5.05E-99 | STAG1 | 3q22.3 | new |
| AN | Sex hormone-binding globulin levels | rs833752 | 3 | 136255169 | A | G | -0.039 | 7.05E-12 | -0.031 | 4.32E-02 | 0.027 | 2.10E-154 | STAG1 | 3q22.3 | new |
| AN | Sex hormone-binding globulin levels | rs80201571 | 3 | 136940937 | G | T | 0.034 | 3.05E-08 | 0.034 | 4.07E-02 | -0.021 | 4.90E-83 | IL20RB | 3q22.3 | new |
| AN | Urate (quantile) | rs1543064 | 3 | 141756541 | C | T | -0.038 | 8.80E-09 | -0.071 | 2.33E-05 | 0.024 | 8.01E-20 | TFDP2 | 3q23 | new |
| AN | Urate (quantile) | rs4697893 | 4 | 9826183 | T | C | 0.167 | 2.81E-12 | 0.141 | 2.14E-02 | -0.238 | 2.45E-138 | DRD5 | 4p16.1 | new |
| AN | Number of cigarettes previously smoked daily | rs13125932 | 4 | 58150175 | C | T | 0.029 | 4.06E-08 | 0.072 | 5.85E-08 | -0.009 | 3.73E-02 | IGFBP7 | 4q12 | new |
| AN | Bioavailable testosterone levels | rs13125932 | 4 | 58150175 | C | T | 0.029 | 3.57E-08 | 0.072 | 5.85E-08 | 0.004 | 3.90E-02 | IGFBP7 | 4q12 | new |
| AN | Sex hormone-binding globulin levels | rs7687658 | 4 | 87299357 | A | G | -0.029 | 1.06E-08 | -0.027 | 4.80E-02 | 0.019 | 1.80E-94 | HSD17B13 | 4q22.1 | new |
| AN | SHBG (quantile) | rs13133311 | 4 | 88217223 | G | A | -0.029 | 6.59E-09 | -0.027 | 4.66E-02 | -0.040 | 2.34E-67 | HSD17B13 | 4q22.1 | new |
| AN | Glycated hemoglobin levels（HbA7c） | rs6872919 | 5 | 25086331 | C | A | -0.030 | 8.46E-09 | -0.074 | 4.05E-08 | 0.004 | 0.002 | CDH10 | 5p14.1 | known |
| AN | Gamma glutamyltransferase (quantile) | rs6872919 | 5 | 25086440 | A | C | 0.030 | 1.73E-08 | 0.074 | 4.05E-08 | -0.006 | 1.22E-02 | CDH10 | 5p14.1 | known |
| AN | Alanine aminotransferase (quantile) | rs6872919 | 5 | 25086440 | A | C | 0.030 | 1.25E-08 | 0.074 | 4.05E-08 | -0.005 | 1.51E-02 | CDH10 | 5p14.1 | known |

| | | | | | | | | | | | | | | |
|---|---|---|---|---|---|---|---|---|---|---|---|---|---|---|
| AN | Urate (quantile) | rs12189774 | 6 | 43816126 | G | A | 0.033 | 5.98E-09 | 0.049 | 9.99E-04 | -0.032 | 5.05E-46 | VEGFA | 6p21.1 | new |
| AN | Urate (quantile) | rs3132604 | 6 | 30741226 | C | T | -0.038 | 2.47E-09 | -0.086 | 1.71E-06 | 0.019 | 7.29E-14 | MHC region | 6p21.33 | |
| AN | Red blood cell (erythrocyte) distribution width | rs3132604 | 6 | 30741226 | C | T | -0.038 | 8.36E-09 | -0.086 | 1.71E-06 | 0.034 | 1.44E-31 | MHC region | 6p21.33 | |
| AN | Ever taken cannabis | rs3132604 | 6 | 30741226 | C | T | -0.037 | 1.87E-08 | -0.086 | 1.71E-06 | -0.024 | 2.83E-07 | MHC region | 6p21.33 | |
| AN | Red blood cell (erythrocyte) distribution width | rs3130063 | 6 | 31558702 | C | T | -0.042 | 2.59E-08 | -0.094 | 9.83E-06 | 0.046 | 2.26E-41 | MHC region | 6p21.33 | |
| AN | Red blood cell (erythrocyte) distribution width | rs532086 | 6 | 31881309 | T | C | -0.040 | 1.31E-08 | -0.090 | 2.83E-06 | 0.039 | 4.59E-33 | MHC region | 6p21.33 | |
| AN | Urate (quantile) | rs853682 | 6 | 28295875 | T | C | -0.040 | 7.30E-09 | -0.072 | 2.32E-04 | 0.033 | 2.09E-33 | MHC region | 6p22.1 | |
| AN | Urate (quantile) | rs4712944 | 6 | 25488265 | C | A | 0.029 | 3.68E-08 | 0.041 | 3.43E-03 | -0.031 | 4.02E-47 | MHC region | 6p22.2 | |
| AN | Urate (quantile) | rs1165215 | 6 | 25798932 | G | A | -0.047 | 7.13E-20 | -0.041 | 1.94E-03 | 0.067 | 8.62E-234 | DHFR2 | 6p22.2 | known |
| AN | Urate (quantile) | rs7748167 | 6 | 25904652 | A | C | -0.048 | 2.78E-10 | -0.079 | 3.33E-04 | 0.047 | 6.59E-53 | MHC region | 6p22.2 | |
| AN | Mean corpuscular haemoglobin | rs6899500 | 6 | 135227089 | T | C | -0.031 | 2.06E-08 | -0.032 | 2.68E-02 | -0.091 | 3.00E-285 | ALDH8A1 | 6q23.3 | new |
| AN | Immature reticulocyte fraction | rs666350 | 6 | 139843192 | C | T | 0.115 | 9.14E-15 | 0.095 | 2.62E-02 | -0.242 | 7.26E-304 | CITED2 | 6q24.1 | new |
| AN | High light scatter reticulocyte percentage | rs666350 | 6 | 139843192 | C | T | 0.103 | 3.12E-12 | 0.095 | 2.62E-02 | -0.231 | 9.91E-275 | CITED2 | 6q24.1 | new |
| AN | High light scatter reticulocyte count | rs666350 | 6 | 139843192 | C | T | 0.101 | 7.42E-12 | 0.095 | 2.62E-02 | -0.213 | 1.08E-238 | CITED2 | 6q24.1 | new |
| AN | SHBG (quantile) | rs572909 | 8 | 81346743 | T | G | -0.040 | 2.59E-08 | -0.050 | 2.57E-02 | -0.034 | 2.41E-36 | ZBTB10 | 8q21.13 | new |
| AN | Alanine aminotransferase (quantile) | rs10733608 | 9 | 117148430 | G | T | 0.035 | 3.05E-11 | 0.049 | 2.18E-04 | -0.047 | 2.66E-96 | AKNA | 9q32 | new |

MTAG section of the table presents the MTAG results specific to Trait 1. Trait 1, anorexia nervosa.

**Table S13**. TWAS-identified genes associated with anorexia nervosa in brain tissues.

| gene_name | pvalue | n | n_indep | p_i_best | t_i_best | eigen_max | eigen_min | eigen_min_kept | z_min | z_max | z_mean | z_sd | tmi |
|---|---|---|---|---|---|---|---|---|---|---|---|---|---|
| CELSR3 | 3.61E-13 | 3 | 3 | 9.25E-14 | Frontal_Cortex_BA9 | 1.86 | 1.87E-01 | 0.19 | -7.45 | -1.05 | -5.29 | 3.67 | 3 |
| QRICH1 | 5.39E-13 | 8 | 2 | 1.46E-06 | Hippocampus | 7.63 | 2.68E-18 | 0.35 | -4.82 | -0.18 | -0.79 | 1.63 | 2 |
| DALRD3 | 5.65E-13 | 9 | 2 | 2.46E-06 | Nucleus_accumbens_basal_ganglia | 7.97 | 2.23E-17 | 0.97 | 0.02 | 4.71 | 2.61 | 2.46 | 2 |
| P4HTM | 6.82E-13 | 12 | 2 | 2.72E-06 | Anterior_cingulate_cortex_BA24 | 10.83 | 2.29E-18 | 1.16 | 0.04 | 4.69 | 2.58 | 2.29 | 2 |
| NDUFAF3 | 1.03E-12 | 6 | 2 | 2.46E-06 | Cerebellum | 5.43 | 1.09E-17 | 0.57 | 0.09 | 4.71 | 1.64 | 2.38 | 2 |
| PRKAR2A | 1.18E-12 | 13 | 2 | 4.65E-13 | Spinal_cord_cervical_c-1 | 11.53 | 2.63E-18 | 1.47 | 3.89 | 7.24 | 6.16 | 1.43 | 2 |
| CAMKV | 1.42E-12 | 10 | 2 | 3.78E-13 | Hippocampus | 9.05 | 4.76E-18 | 0.94 | -3.16 | 7.26 | 2.49 | 3.23 | 2 |
| RP11-3B7.1 | 1.45E-12 | 4 | 2 | 1.78E-12 | Cortex | 2.85 | 1.26E-17 | 1.15 | 4.98 | 7.05 | 6.02 | 1.19 | 2 |
| RP11-572O6.1 | 2.04E-12 | 13 | 3 | 4.57E-14 | Caudate_basal_ganglia | 6.81 | 2.40E-18 | 1.84 | -1.00 | 7.54 | 3.66 | 3.65 | 3 |
| KLHDC8B | 6.68E-12 | 13 | 2 | 9.73E-13 | Hypothalamus | 11.45 | 8.45E-17 | 1.55 | -7.13 | -0.04 | -6.00 | 1.90 | 2 |
| C3orf62 | 7.20E-12 | 13 | 1 | 7.20E-12 | Amygdala | 13.00 | 3.63E-33 | 13.00 | 6.85 | 6.85 | 6.85 | 0.00 | 1 |
| WDR6 | 1.57E-11 | 13 | 2 | 3.52E-11 | Cortex | 12.48 | 2.87E-17 | 0.52 | -6.62 | -4.68 | -5.90 | 0.86 | 2 |
| NICN1 | 2.50E-11 | 11 | 4 | 8.31E-12 | Substantia_nigra | 6.36 | 1.70E-17 | 0.45 | -6.83 | -0.76 | -3.72 | 1.79 | 4 |
| SEMA3F | 1.33E-10 | 7 | 3 | 6.74E-10 | Substantia_nigra | 5.32 | 5.74E-17 | 0.47 | -6.17 | -2.25 | -3.65 | 1.77 | 3 |
| SLC26A6 | 2.12E-10 | 3 | 1 | 1.21E-10 | Putamen_basal_ganglia | 2.95 | 1.83E-17 | 2.95 | -6.44 | -6.24 | -6.30 | 0.12 | 1 |
| CCDC71 | 3.97E-08 | 12 | 1 | 3.85E-12 | Cerebellum | 11.37 | 6.51E-19 | 11.37 | -6.94 | -4.84 | -5.36 | 0.73 | 1 |
| UCN2 | 2.15E-07 | 13 | 3 | 2.52E-05 | Cerebellum | 8.02 | 1.57E-18 | 0.43 | 0.10 | 4.21 | 0.89 | 1.04 | 3 |
| NCKIPSD | 2.36E-07 | 13 | 1 | 6.08E-08 | Cerebellum | 12.72 | 2.88E-18 | 12.72 | 3.65 | 5.42 | 5.11 | 0.47 | 1 |
| LINC00324 | 3.72E-07 | 1 | 1 | 3.72E-07 | Cerebellum | 1.00 | 1.00E+00 | 1.00 | -5.08 | -5.08 | -5.08 | NA | 1 |
| USP4 | 4.84E-07 | 12 | 2 | 4.48E-07 | Frontal_Cortex_BA9 | 8.04 | 1.68E-18 | 3.96 | -5.05 | 0.89 | -2.22 | 2.36 | 2 |
| CCDC36 | 5.23E-07 | 9 | 1 | 4.91E-07 | Caudate_basal_ganglia | 8.99 | 5.87E-20 | 8.99 | 4.98 | 5.03 | 5.01 | 0.02 | 1 |
| IP6K2 | 6.16E-07 | 5 | 3 | 3.48E-05 | Cerebellar_Hemisphere | 2.50 | 7.28E-17 | 0.61 | -1.00 | 4.14 | 1.12 | 2.30 | 3 |
| BSN | 9.62E-07 | 2 | 2 | 8.88E-06 | Cortex | 1.39 | 6.05E-01 | 0.61 | 0.84 | 4.44 | 2.64 | 2.55 | 2 |
| GPX1 | 1.18E-06 | 13 | 2 | 1.01E-05 | Hippocampus | 11.77 | 2.40E-17 | 1.23 | -0.86 | 4.42 | 1.68 | 1.53 | 2 |
| DAG1 | 1.28E-06 | 4 | 2 | 2.36E-07 | Caudate_basal_ganglia | 2.60 | 1.30E-17 | 1.40 | -5.17 | -0.86 | -3.82 | 2.00 | 2 |
| RHOA | 1.41E-06 | 11 | 2 | 1.50E-06 | Anterior_cingulate_cortex_BA24 | 7.93 | 3.11E-17 | 3.06 | -0.89 | 4.81 | 4.10 | 1.67 | 2 |
| BSN-AS2 | 1.45E-06 | 13 | 2 | 1.29E-06 | Anterior_cingulate_cortex_BA24 | 12.16 | 8.22E-18 | 0.83 | -4.84 | -0.84 | -4.43 | 1.12 | 2 |
| ZMYND10 | 1.76E-06 | 2 | 2 | 2.73E-07 | Cerebellar_Hemisphere | 1.14 | 8.59E-01 | 0.86 | -0.98 | 5.14 | 2.08 | 4.33 | 2 |
| ARIH2OS | 2.01E-06 | 7 | 1 | 1.88E-06 | Anterior_cingulate_cortex_BA24 | 7.00 | 4.91E-17 | 7.00 | -4.77 | -4.74 | -4.75 | 0.01 | 1 |
| ARIH2 | 2.07E-06 | 5 | 1 | 1.88E-06 | Cerebellum | 5.00 | 3.89E-68 | 5.00 | 4.74 | 4.77 | 4.75 | 0.01 | 1 |
| TCTA | 2.19E-06 | 13 | 3 | 2.30E-05 | Caudate_basal_ganglia | 9.63 | 1.73E-18 | 0.52 | -3.68 | 4.23 | 0.39 | 2.23 | 3 |
| APEH | 2.89E-06 | 9 | 2 | 1.00E-06 | Putamen_basal_ganglia | 7.24 | 6.57E-17 | 1.76 | -4.89 | 0.31 | -3.86 | 1.59 | 2 |
| ACYP2 | 2.89E-06 | 13 | 4 | 1.30E-06 | Spinal_cord_cervical_c-1 | 6.81 | 4.81E-17 | 0.94 | -1.95 | 4.84 | 2.51 | 1.87 | 4 |
| BORCS6 | 3.39E-06 | 6 | 4 | 8.01E-05 | Frontal_Cortex_BA9 | 3.08 | 7.35E-19 | 0.30 | -2.65 | 3.94 | 0.57 | 2.32 | 4 |

| | | | | | | | | | | | | | |
|---|---|---|---|---|---|---|---|---|---|---|---|---|---|
| CYB561D2 | 4.21E-06 | 13 | 3 | 2.73E-07 | Caudate_basal_ganglia | 7.96 | 1.91E-18 | 0.77 | -5.14 | -0.51 | -3.18 | 2.26 | 3 |
| FAM212A | 4.51E-06 | 9 | 2 | 5.65E-05 | Anterior_cingulate_cortex_BA24 | 7.08 | 7.38E-18 | 1.92 | -4.03 | 2.16 | -0.17 | 2.85 | 2 |
| MST1 | 4.79E-06 | 7 | 5 | 1.93E-05 | Hypothalamus | 3.62 | 5.91E-17 | 0.25 | -4.27 | 1.99 | 0.30 | 2.19 | 5 |
| CAAP1 | 9.84E-06 | 7 | 5 | 0.002285 | Hypothalamus | 3.08 | 1.01E-17 | 0.12 | -3.05 | 1.03 | -0.90 | 1.19 | 5 |
| ACTR5 | 1.18E-05 | 12 | 4 | 1.70E-06 | Putamen_basal_ganglia | 8.53 | 1.16E-17 | 0.91 | -4.79 | 0.70 | -0.27 | 1.57 | 4 |
| SUOX | 1.50E-05 | 12 | 1 | 9.13E-06 | Hypothalamus | 11.56 | 4.67E-17 | 11.56 | 4.15 | 4.44 | 4.25 | 0.08 | 1 |
| CNPY4 | 1.53E-05 | 8 | 5 | 0.0007864 | Spinal_cord_cervical_c-1 | 5.48 | 8.51E-18 | 0.19 | -1.87 | 3.36 | -0.19 | 2.20 | 5 |
| CTNNB1 | 1.76E-05 | 12 | 1 | 8.43E-06 | Spinal_cord_cervical_c-1 | 11.82 | 2.15E-18 | 11.82 | 4.24 | 4.45 | 4.26 | 0.06 | 1 |
| SCUBE1 | 1.95E-05 | 11 | 5 | 0.0004049 | Spinal_cord_cervical_c-1 | 7.54 | 8.53E-17 | 0.47 | -3.54 | -0.16 | -2.10 | 0.87 | 5 |
| CHAC2 | 2.38E-05 | 10 | 4 | 5.45E-07 | Amygdala | 3.47 | 6.59E-33 | 0.52 | -5.01 | 2.96 | 0.15 | 2.45 | 4 |
| APOM | 2.39E-05 | 10 | 1 | 2.31E-05 | Amygdala | 9.93 | 3.60E-17 | 9.93 | -4.23 | -4.20 | -4.21 | 0.01 | 1 |
| ERBB3 | 2.60E-05 | 8 | 1 | 2.60E-05 | Hippocampus | 8.00 | 1.34E-17 | 8.00 | 4.21 | 4.21 | 4.21 | 0.00 | 1 |
| GPR75 | 2.60E-05 | 10 | 3 | 1.80E-06 | Amygdala | 7.09 | 4.83E-18 | 0.26 | -4.77 | -1.78 | -3.13 | 1.46 | 3 |
| ERLEC1 | 2.78E-05 | 8 | 3 | 0.05238 | Cerebellar_Hemisphere | 6.19 | 6.84E-17 | 0.21 | -1.94 | 1.69 | -1.37 | 1.28 | 3 |
| CALML6 | 3.09E-05 | 13 | 2 | 6.68E-06 | Spinal_cord_cervical_c-1 | 11.21 | 1.78E-17 | 1.58 | 0.55 | 4.50 | 2.58 | 1.01 | 2 |
| ASB3 | 3.23E-05 | 13 | 4 | 5.89E-07 | Amygdala | 8.32 | 9.86E-18 | 0.34 | -4.99 | -0.68 | -3.58 | 1.83 | 4 |
| RBM6 | 3.24E-05 | 12 | 1 | 3.24E-05 | Nucleus_accumbens_basal_ganglia | 12.00 | 2.53E-34 | 12.00 | -4.16 | -4.16 | -4.16 | 0.00 | 1 |
| VWA7 | 3.27E-05 | 13 | 7 | 2.23E-05 | Substantia_nigra | 4.67 | 1.49E-17 | 0.18 | -4.24 | 1.52 | -0.48 | 2.11 | 7 |
| C6orf48 | 3.42E-05 | 13 | 3 | 9.96E-06 | Cerebellum | 9.61 | 6.06E-18 | 0.58 | -4.42 | 0.02 | -1.76 | 1.60 | 3 |
| KCNH3 | 3.54E-05 | 3 | 2 | 1.06E-05 | Substantia_nigra | 2.51 | 6.52E-02 | 0.43 | 3.70 | 4.40 | 4.11 | 0.36 | 2 |
| TRAIP | 3.77E-05 | 7 | 1 | 3.77E-05 | Amygdala | 7.00 | 5.79E-17 | 7.00 | 4.12 | 4.12 | 4.12 | 0.00 | 1 |
| ABHD16A | 3.96E-05 | 13 | 3 | 2.19E-05 | Amygdala | 6.00 | 6.61E-18 | 1.68 | -4.24 | 0.91 | -1.90 | 2.03 | 3 |
| C4A | 4.14E-05 | 13 | 1 | 2.76E-05 | Caudate_basal_ganglia | 12.88 | 1.13E-17 | 12.88 | -4.19 | -3.86 | -4.08 | 0.09 | 1 |
| MSH5 | 4.26E-05 | 12 | 3 | 5.04E-06 | Caudate_basal_ganglia | 6.67 | 3.25E-17 | 0.97 | -4.56 | -1.75 | -3.23 | 1.27 | 3 |
| RP11-477N3.1 | 4.58E-05 | 13 | 4 | 1.75E-06 | Cerebellar_Hemisphere | 10.00 | 3.62E-17 | 0.46 | -4.78 | -1.01 | -3.37 | 1.10 | 4 |
| PFAS | 4.68E-05 | 12 | 5 | 6.03E-06 | Cerebellum | 5.70 | 6.04E-19 | 0.23 | -1.18 | 4.53 | 0.34 | 1.82 | 5 |
| GBE1 | 4.77E-05 | 11 | 3 | 3.28E-06 | Anterior_cingulate_cortex_BA24 | 6.11 | 4.81E-18 | 1.69 | -4.65 | 4.43 | -0.75 | 3.22 | 3 |
| CLIC1 | 4.88E-05 | 12 | 2 | 6.48E-06 | Cortex | 8.10 | 1.21E-17 | 3.72 | -4.51 | 0.77 | -1.19 | 2.40 | 2 |
| LY6G6C | 5.04E-05 | 8 | 3 | 1.66E-05 | Cortex | 5.16 | 1.24E-17 | 0.39 | -4.31 | 3.80 | 0.24 | 3.02 | 3 |
| ZBTB12 | 5.45E-05 | 2 | 2 | 3.27E-05 | Cortex | 1.02 | 9.76E-01 | 0.98 | -4.15 | 1.64 | -1.26 | 4.10 | 2 |
| DNAJC22 | 5.47E-05 | 13 | 2 | 1.70E-05 | Spinal_cord_cervical_c-1 | 11.72 | 7.28E-20 | 0.90 | 1.87 | 4.30 | 3.03 | 0.88 | 2 |
| FBXO27 | 5.72E-05 | 13 | 5 | 3.85E-05 | Putamen_basal_ganglia | 7.06 | 2.48E-17 | 0.33 | -1.94 | 4.12 | 1.45 | 2.04 | 5 |
| VAMP2 | 6.24E-05 | 9 | 3 | 0.0001549 | Putamen_basal_ganglia | 6.91 | 1.98E-17 | 0.96 | -3.78 | 1.17 | -2.98 | 1.65 | 3 |
| MST1R | 7.61E-05 | 13 | 1 | 1.59E-05 | Caudate_basal_ganglia | 12.52 | 8.21E-18 | 12.52 | 3.71 | 4.32 | 3.88 | 0.14 | 1 |
| HLA-DMA | 7.68E-05 | 13 | 1 | 4.34E-05 | Anterior_cingulate_cortex_BA24 | 12.88 | 5.74E-19 | 12.88 | 3.89 | 4.09 | 3.94 | 0.05 | 1 |
| ACTG1 | 7.89E-05 | 5 | 1 | 7.89E-05 | Caudate_basal_ganglia | 5.00 | 5.19E-33 | 5.00 | -3.95 | 3.95 | 0.79 | 4.32 | 1 |
| RNF123 | 7.96E-05 | 13 | 1 | 7.75E-05 | Spinal_cord_cervical_c-1 | 12.99 | 2.88E-19 | 12.99 | 3.93 | 3.95 | 3.94 | 0.01 | 1 |

| | | | | | | | | | | | | | |
|---|---|---|---|---|---|---|---|---|---|---|---|---|---|
| RBM5 | 8.44E-05 | 3 | 1 | 8.44E-05 | Cerebellar_Hemisphere | 3.00 | 5.35E-35 | 3.00 | -3.93 | -3.93 | -3.93 | 0.00 | 1 |
| CCDC93 | 8.58E-05 | 13 | 3 | 0.0006369 | Cerebellum | 11.55 | 2.15E-17 | 0.39 | -3.42 | -0.49 | -2.38 | 0.87 | 3 |
| PSME4 | 8.99E-05 | 13 | 4 | 6.88E-05 | Hypothalamus | 8.35 | 6.56E-18 | 0.93 | -2.45 | 3.98 | -0.49 | 2.35 | 4 |
| PRKCSH | 9.53E-05 | 9 | 2 | 2.30E-05 | Cerebellum | 8.31 | 1.27E-17 | 0.44 | -4.23 | -2.55 | -3.88 | 0.51 | 2 |
| SLC25A35 | 9.88E-05 | 13 | 5 | 4.44E-05 | Cortex | 7.00 | 9.74E-18 | 0.49 | -2.13 | 4.08 | 1.48 | 2.27 | 5 |
| MGMT | 1.09E-04 | 13 | 3 | 4.98E-06 | Substantia_nigra | 11.22 | 6.23E-19 | 0.38 | 2.79 | 4.57 | 4.13 | 0.45 | 3 |
| DHFR2 | 1.14E-04 | 8 | 1 | 0.0001137 | Frontal_Cortex_BA9 | 8.00 | 6.54E-18 | 8.00 | -3.86 | -3.86 | -3.86 | 0.00 | 1 |
| STAC3 | 1.18E-04 | 10 | 2 | 2.04E-05 | Cortex | 9.48 | 2.68E-18 | 0.52 | -4.26 | -3.56 | -3.76 | 0.33 | 2 |
| GDNF | 1.18E-04 | 4 | 3 | 0.0003971 | Frontal_Cortex_BA9 | 2.17 | 2.51E-34 | 0.18 | -3.54 | 2.27 | -0.66 | 3.33 | 3 |
| RAB5B | 1.21E-04 | 9 | 2 | 1.69E-05 | Frontal_Cortex_BA9 | 7.37 | 8.63E-17 | 1.44 | 0.38 | 4.30 | 1.17 | 1.53 | 2 |
| ZRANB1 | 1.24E-04 | 9 | 3 | 5.57E-06 | Hippocampus | 7.10 | 1.26E-22 | 0.56 | -1.10 | 4.54 | -0.17 | 1.98 | 3 |
| SOX5 | 1.32E-04 | 2 | 2 | 6.36E-05 | Frontal_Cortex_BA9 | 1.73 | 2.67E-01 | 0.27 | -4.00 | -3.86 | -3.93 | 0.10 | 2 |
| DDX18 | 1.39E-04 | 12 | 6 | 0.0006882 | Anterior_cingulate_cortex_BA24 | 4.32 | 3.20E-17 | 0.23 | -1.19 | 3.39 | 0.79 | 1.58 | 6 |
| STX19 | 1.44E-04 | 10 | 1 | 0.0001007 | Substantia_nigra | 9.99 | 4.36E-18 | 9.99 | 3.79 | 3.89 | 3.80 | 0.03 | 1 |
| TRPV1 | 1.49E-04 | 12 | 6 | 0.001453 | Anterior_cingulate_cortex_BA24 | 4.97 | 9.56E-17 | 0.24 | -3.18 | 2.47 | -0.91 | 2.19 | 6 |
| ZNF3 | 1.53E-04 | 6 | 3 | 8.29E-06 | Anterior_cingulate_cortex_BA24 | 3.03 | 6.20E-33 | 0.97 | -4.46 | 0.75 | -1.20 | 2.57 | 3 |
| IKZF4 | 1.57E-04 | 5 | 2 | 4.68E-05 | Frontal_Cortex_BA9 | 3.79 | 8.51E-17 | 1.21 | -4.07 | -0.43 | -2.90 | 1.68 | 2 |
| NR6A1 | 1.69E-04 | 1 | 1 | 0.0001685 | Nucleus_accumbens_basal_ganglia | 1.00 | 1.00E+00 | 1.00 | 3.76 | 3.76 | 3.76 | NA | 1 |
| SLC12A5 | 1.82E-04 | 12 | 6 | 2.93E-05 | Nucleus_accumbens_basal_ganglia | 5.74 | 3.49E-17 | 0.28 | -3.23 | 4.18 | 0.26 | 2.76 | 6 |
| TMEM114 | 1.91E-04 | 1 | 1 | 0.0001911 | Cortex | 1.00 | 1.00E+00 | 1.00 | 3.73 | 3.73 | 3.73 | NA | 1 |
| RNF2 | 1.95E-04 | 1 | 1 | 0.0001948 | Cerebellar_Hemisphere | 1.00 | 1.00E+00 | 1.00 | -3.73 | -3.73 | -3.73 | NA | 1 |
| SYNCRIP | 1.99E-04 | 6 | 1 | 0.0001829 | Cortex | 5.99 | 6.67E-17 | 5.99 | -3.71 | 3.74 | 2.48 | 3.03 | 1 |
| CELSR2 | 2.01E-04 | 6 | 2 | 0.0042 | Cortex | 5.32 | 3.89E-17 | 0.65 | -2.86 | 2.86 | 0.16 | 2.75 | 2 |
| TRIM26 | 2.20E-04 | 12 | 4 | 4.52E-05 | Spinal_cord_cervical_c-1 | 7.58 | 8.51E-20 | 0.31 | 1.38 | 4.08 | 2.56 | 0.90 | 4 |
| SELENOH | 2.28E-04 | 7 | 1 | 0.0002283 | Cerebellar_Hemisphere | 7.00 | 9.45E-49 | 7.00 | 3.69 | 3.69 | 3.69 | 0.00 | 1 |
| TMEM52 | 2.43E-04 | 10 | 3 | 0.0001274 | Caudate_basal_ganglia | 7.93 | 1.78E-17 | 0.34 | -0.74 | 3.83 | 1.64 | 1.71 | 3 |
| PTH2 | 2.76E-04 | 1 | 1 | 0.0002763 | Cerebellar_Hemisphere | 1.00 | 1.00E+00 | 1.00 | 3.64 | 3.64 | 3.64 | NA | 1 |
| FOXA2 | 2.80E-04 | 1 | 1 | 0.00028 | Substantia_nigra | 1.00 | 1.00E+00 | 1.00 | -3.63 | -3.63 | -3.63 | NA | 1 |
| ARMS2 | 3.06E-04 | 10 | 2 | 0.0003991 | Anterior_cingulate_cortex_BA24 | 8.81 | 9.58E-17 | 1.00 | -1.70 | 3.54 | -0.43 | 1.45 | 2 |
| RP1-153G14.4 | 3.08E-04 | 8 | 2 | 0.002554 | Anterior_cingulate_cortex_BA24 | 6.96 | 1.18E-17 | 1.00 | 0.31 | 3.02 | 1.37 | 1.36 | 2 |
| ZSCAN23 | 3.18E-04 | 13 | 1 | 0.000142 | Substantia_nigra | 12.56 | 5.60E-17 | 12.56 | 3.35 | 3.80 | 3.54 | 0.10 | 1 |
| C1orf64 | 3.22E-04 | 11 | 3 | 0.0002929 | Anterior_cingulate_cortex_BA24 | 8.40 | 1.45E-17 | 0.43 | -3.62 | 0.54 | -0.63 | 1.51 | 3 |
| RGPD8 | 3.62E-04 | 12 | 1 | 0.000179 | Caudate_basal_ganglia | 11.65 | 7.49E-18 | 11.65 | -3.75 | -2.55 | -3.51 | 0.41 | 1 |
| KNOP1 | 3.64E-04 | 12 | 1 | 0.000261 | Cerebellar_Hemisphere | 11.84 | 3.74E-17 | 11.84 | -3.65 | -3.49 | -3.54 | 0.05 | 1 |
| PHF2 | 3.76E-04 | 2 | 1 | 0.0003755 | Amygdala | 2.00 | 3.53E-16 | 2.00 | 3.56 | 3.56 | 3.56 | 0.00 | 1 |
| PRR7 | 3.82E-04 | 3 | 1 | 0.0003821 | Cerebellar_Hemisphere | 3.00 | 5.35E-35 | 3.00 | -3.55 | 3.55 | -1.18 | 4.10 | 1 |
| C1QL4 | 3.85E-04 | 7 | 2 | 0.001599 | Caudate_basal_ganglia | 5.95 | 3.67E-19 | 1.00 | -3.16 | 2.51 | -2.34 | 2.14 | 2 |

| | | | | | | | | | | | | | |
|---|---|---|---|---|---|---|---|---|---|---|---|---|---|
| CELF1 | 3.98E-04 | 13 | 3 | 0.002956 | Cerebellar_Hemisphere | 11.13 | 2.88E-17 | 0.52 | -2.24 | 2.97 | -1.01 | 1.64 | 3 |
| CDC73 | 4.25E-04 | 10 | 1 | 0.0004251 | Frontal_Cortex_BA9 | 10.00 | 3.02E-18 | 10.00 | -3.52 | -3.52 | -3.52 | 0.00 | 1 |
| SMARCAL1 | 4.40E-04 | 12 | 2 | 5.66E-05 | Hippocampus | 10.37 | 8.70E-17 | 1.41 | -4.03 | 1.59 | -3.33 | 1.56 | 2 |
| PXDNL | 4.47E-04 | 10 | 3 | 5.36E-05 | Anterior_cingulate_cortex_BA24 | 7.85 | 3.14E-17 | 0.35 | -4.04 | 4.04 | 0.84 | 3.48 | 3 |
| ZBTB1 | 4.47E-04 | 10 | 1 | 0.0004036 | Frontal_Cortex_BA9 | 9.63 | 5.19E-19 | 9.63 | 2.99 | 3.54 | 3.44 | 0.16 | 1 |
| VCL | 4.63E-04 | 2 | 1 | 0.0004628 | Cortex | 2.00 | 3.35E-17 | 2.00 | 3.50 | 3.50 | 3.50 | 0.00 | 1 |
| MYO5A | 4.71E-04 | 11 | 4 | 0.0001405 | Caudate_basal_ganglia | 7.58 | 2.48E-17 | 0.26 | -3.81 | 1.75 | -2.25 | 1.72 | 4 |
| RP11-104L21.3 | 4.75E-04 | 1 | 1 | 0.0004748 | Hypothalamus | 1.00 | 1.00E+00 | 1.00 | 3.49 | 3.49 | 3.49 | NA | 1 |
| ABHD8 | 4.89E-04 | 7 | 3 | 0.05162 | Anterior_cingulate_cortex_BA24 | 5.68 | 3.99E-17 | 0.40 | -1.95 | 0.93 | -1.13 | 1.34 | 3 |
| PIAS2 | 4.98E-04 | 11 | 1 | 0.0005163 | Anterior_cingulate_cortex_BA24 | 10.56 | 4.12E-20 | 10.56 | -3.47 | -3.01 | -3.41 | 0.15 | 1 |
| DDAH2 | 5.01E-04 | 12 | 1 | 0.000418 | Cerebellar_Hemisphere | 11.90 | 2.17E-17 | 11.90 | -3.53 | -2.88 | -3.47 | 0.19 | 1 |
| RCCD1 | 5.07E-04 | 12 | 5 | 0.01109 | Frontal_Cortex_BA9 | 6.99 | 1.06E-17 | 0.26 | -2.54 | 1.79 | -0.92 | 1.48 | 5 |
| ZNF595 | 5.36E-04 | 2 | 2 | 0.001085 | Substantia_nigra | 1.16 | 8.45E-01 | 0.84 | -1.56 | 3.27 | 0.85 | 3.41 | 2 |
| ALDH16A1 | 5.47E-04 | 13 | 2 | 0.0006018 | Caudate_basal_ganglia | 12.26 | 7.50E-17 | 0.51 | -3.43 | -1.94 | -2.98 | 0.56 | 2 |
| CFAP74 | 5.77E-04 | 6 | 4 | 1.49E-05 | Amygdala | 3.52 | 1.22E-02 | 0.39 | -1.31 | 4.33 | 1.77 | 2.04 | 4 |
| AP003774.4 | 5.80E-04 | 6 | 2 | 0.0092145 | Cerebellum | 5.19 | 9.81E-18 | 0.81 | -1.59 | 2.60 | -0.36 | 1.93 | 2 |
| ZFP91 | 5.81E-04 | 4 | 1 | 0.0005809 | Amygdala | 4.00 | 2.97E-17 | 4.00 | -3.44 | -3.44 | -3.44 | 0.00 | 1 |
| FNBP4 | 5.86E-04 | 6 | 1 | 0.0005856 | Hippocampus | 6.00 | 2.93E-34 | 6.00 | 3.44 | 3.44 | 3.44 | 0.00 | 1 |
| AC068831.16 | 5.90E-04 | 10 | 4 | 0.0051637 | Frontal_Cortex_BA9 | 7.53 | 4.67E-17 | 0.32 | -2.80 | 2.22 | 1.10 | 1.65 | 4 |
| NIPSNAP1 | 5.91E-04 | 6 | 3 | 0.002305 | Hippocampus | 3.15 | 2.19E-32 | 0.84 | -3.05 | 1.69 | 0.11 | 1.87 | 3 |
| SECISBP2 | 5.94E-04 | 13 | 1 | 0.0005109 | Cerebellar_Hemisphere | 12.64 | 3.03E-17 | 12.64 | 2.78 | 3.47 | 3.38 | 0.19 | 1 |
| GCSAM | 5.94E-04 | 3 | 2 | 0.0001289 | Caudate_basal_ganglia | 1.97 | 5.74E-04 | 1.03 | -0.50 | 3.83 | 2.37 | 2.48 | 2 |
| P2RX4 | 6.14E-04 | 12 | 3 | 0.0359366 | Cerebellum | 9.75 | 1.81E-18 | 0.49 | -0.45 | 2.10 | 0.63 | 0.90 | 3 |
| PIH1D1 | 6.33E-04 | 13 | 3 | 0.0008765 | Amygdala | 10.04 | 6.98E-18 | 0.40 | -3.33 | 1.91 | -0.55 | 1.84 | 3 |
| USP45 | 6.66E-04 | 6 | 5 | 0.0003523 | Cerebellar_Hemisphere | 2.76 | 1.68E-17 | 0.15 | -3.57 | -0.14 | -1.94 | 1.13 | 5 |
| B3GALT2 | 6.70E-04 | 3 | 2 | 0.0002029 | Putamen_basal_ganglia | 2.78 | 1.65E-03 | 0.22 | -3.61 | 3.72 | 1.23 | 4.19 | 2 |
| FCN1 | 6.79E-04 | 12 | 3 | 0.0008463 | Cerebellum | 7.81 | 5.37E-17 | 1.27 | -3.14 | 3.34 | 1.41 | 2.18 | 3 |
| PCMTD1 | 6.80E-04 | 13 | 4 | 1.83E-05 | Hypothalamus | 9.22 | 9.98E-18 | 0.37 | 0.40 | 4.28 | 3.27 | 1.29 | 4 |
| SEMA6D | 6.87E-04 | 1 | 1 | 0.000687 | Cerebellum | 1.00 | 1.00E+00 | 1.00 | -3.39 | -3.39 | -3.39 | NA | 1 |
| NAT6 | 6.89E-04 | 13 | 2 | 0.02323 | Amygdala | 11.91 | 1.97E-17 | 0.83 | -2.27 | 0.21 | -0.76 | 1.02 | 2 |
| DVL2 | 6.93E-04 | 4 | 1 | 0.0006928 | Caudate_basal_ganglia | 4.00 | 1.00E-32 | 4.00 | -3.39 | -3.39 | -3.39 | 0.00 | 1 |
| EPB41 | 7.34E-04 | 11 | 2 | 0.0005398 | Anterior_cingulate_cortex_BA24 | 9.20 | 3.76E-20 | 1.80 | -3.46 | 3.46 | 1.10 | 3.24 | 2 |
| MSANTD4 | 7.43E-04 | 11 | 2 | 0.00748 | Amygdala | 10.06 | 4.15E-18 | 0.73 | -2.67 | 0.63 | 0.16 | 1.05 | 2 |
| HLA-DMB | 7.44E-04 | 10 | 4 | 2.89E-05 | Frontal_Cortex_BA9 | 6.75 | 3.26E-18 | 0.23 | -0.79 | 4.18 | 2.72 | 2.02 | 4 |
| FGF14-AS2 | 7.58E-04 | 13 | 2 | 5.62E-05 | Hypothalamus | 12.36 | 6.92E-17 | 0.46 | 3.47 | 4.03 | 3.66 | 0.20 | 2 |
| JRK | 7.71E-04 | 11 | 5 | 0.001073 | Hippocampus | 6.78 | 2.83E-18 | 0.24 | -3.27 | 3.14 | 1.23 | 1.82 | 5 |
| FSCN2 | 7.71E-04 | 8 | 3 | 0.0001439 | Amygdala | 5.73 | 2.87E-17 | 0.52 | 0.59 | 3.80 | 2.74 | 1.08 | 3 |

| | | | | | | | | | | | | | |
|---|---|---|---|---|---|---|---|---|---|---|---|---|---|
| IP6K3 | 7.76E-04 | 13 | 5 | 0.0012866 | Hypothalamus | 8.32 | 7.37E-17 | 0.74 | -3.22 | 3.07 | -1.47 | 1.89 | 5 |
| ZDHHC5 | 8.05E-04 | 13 | 2 | 0.0002235 | Cortex | 10.82 | 2.42E-17 | 2.18 | -3.53 | 3.69 | 2.57 | 1.85 | 2 |
| RP11-290F5.2 | 8.07E-04 | 4 | 1 | 0.0006895 | Substantia_nigra | 3.98 | 3.14E-17 | 3.98 | -3.39 | -3.27 | -3.34 | 0.06 | 1 |
| EFTUD2 | 8.18E-04 | 13 | 3 | 0.0011557 | Hypothalamus | 9.79 | 7.91E-18 | 0.70 | -3.25 | 0.38 | -1.55 | 0.89 | 3 |
| ZNF653 | 8.21E-04 | 13 | 1 | 0.0008214 | Cortex | 13.00 | 1.86E-18 | 13.00 | -3.35 | 3.35 | 2.83 | 1.86 | 1 |
| ACP7 | 8.22E-04 | 10 | 4 | 2.36E-05 | Spinal_cord_cervical_c-1 | 6.56 | 2.92E-17 | 0.31 | 1.03 | 4.23 | 2.41 | 1.14 | 4 |
| GOLGA6L10 | 8.41E-04 | 3 | 3 | 0.0003395 | Putamen_basal_ganglia | 2.24 | 1.45E-01 | 0.14 | -3.58 | -1.77 | -2.46 | 0.98 | 3 |
| SLC25A12 | 8.60E-04 | 8 | 3 | 6.21E-05 | Nucleus_accumbens_basal_ganglia | 6.28 | 1.12E-16 | 0.47 | -4.00 | -0.76 | -3.13 | 1.01 | 3 |
| CCDC88C | 8.67E-04 | 13 | 5 | 0.0027687 | Cortex | 6.40 | 6.70E-18 | 0.26 | -2.99 | 2.95 | -0.83 | 1.87 | 5 |
| AGFG2 | 8.78E-04 | 13 | 3 | 0.0179509 | Cerebellum | 10.53 | 7.98E-20 | 0.61 | -1.81 | 2.37 | -1.01 | 1.55 | 3 |
| TYMP | 9.02E-04 | 10 | 5 | 0.0001533 | Hypothalamus | 6.39 | 4.97E-17 | 0.39 | -1.49 | 3.79 | 1.48 | 1.49 | 5 |
| NEGR1 | 9.03E-04 | 13 | 1 | 0.0006247 | Nucleus_accumbens_basal_ganglia | 12.48 | 1.04E-20 | 12.48 | -3.42 | -3.03 | -3.25 | 0.11 | 1 |
| CNTLN | 9.32E-04 | 11 | 5 | 0.0002565 | Putamen_basal_ganglia | 5.91 | 1.27E-17 | 0.24 | -3.66 | 2.32 | -0.99 | 2.06 | 5 |
| LGR4 | 9.43E-04 | 13 | 5 | 0.0001313 | Cerebellum | 7.17 | 3.11E-17 | 0.32 | -3.82 | 3.82 | 1.27 | 2.10 | 5 |
| ZKSCAN8 | 9.44E-04 | 13 | 1 | 0.0006957 | Frontal_Cortex_BA9 | 12.73 | 2.51E-17 | 12.73 | -3.39 | -2.79 | -3.27 | 0.20 | 1 |
| BNIPL | 9.64E-04 | 11 | 3 | 8.25E-05 | Hippocampus | 8.14 | 4.32E-18 | 1.03 | 1.89 | 3.94 | 3.33 | 0.68 | 3 |
| PGM3 | 9.71E-04 | 11 | 3 | 0.0004732 | Anterior_cingulate_cortex_BA24 | 9.19 | 2.18E-17 | 0.56 | 2.44 | 3.50 | 2.61 | 0.32 | 3 |
| KBTBD4 | 9.88E-04 | 9 | 1 | 0.0009879 | Hypothalamus | 9.00 | 0.00E+00 | 9.00 | -3.29 | 3.29 | 0.37 | 3.47 | 1 |
| YPEL4 | 9.89E-04 | 2 | 2 | 0.001388 | Anterior_cingulate_cortex_BA24 | 1.04 | 9.60E-01 | 0.96 | -3.20 | 1.77 | -0.71 | 3.51 | 2 |
| HORMAD2 | 1.02E-03 | 10 | 2 | 0.001014 | Cerebellum | 9.17 | 4.12E-33 | 0.83 | -3.29 | 3.29 | -0.04 | 3.42 | 2 |
| SLC17A7 | 1.04E-03 | 8 | 1 | 0.001017 | Anterior_cingulate_cortex_BA24 | 7.95 | 3.65E-17 | 7.95 | -3.29 | 3.29 | -1.66 | 3.02 | 1 |
| P2RY1 | 1.04E-03 | 13 | 2 | 0.0001307 | Substantia_nigra | 9.30 | 8.12E-18 | 3.49 | -2.04 | 3.83 | 1.66 | 1.89 | 2 |
| SRP9 | 1.04E-03 | 13 | 4 | 0.0014173 | Amygdala | 6.98 | 9.09E-18 | 0.69 | -3.19 | 0.78 | -1.67 | 1.46 | 4 |
| IL1RN | 1.05E-03 | 1 | 1 | 0.001048 | Hippocampus | 1.00 | 1.00E+00 | 1.00 | 3.28 | 3.28 | 3.28 | NA | 1 |
| MED7 | 1.05E-03 | 8 | 2 | 0.0003317 | Cerebellum | 6.20 | 2.79E-18 | 1.80 | -3.59 | 2.79 | 0.49 | 2.68 | 2 |
| ANO5 | 1.10E-03 | 8 | 2 | 0.001615 | Putamen_basal_ganglia | 7.01 | 3.73E-17 | 0.99 | -1.57 | 3.15 | -0.98 | 1.67 | 2 |
| ZNF335 | 1.11E-03 | 12 | 3 | 0.0007008 | Cerebellum | 8.19 | 3.29E-17 | 0.45 | -1.72 | 3.39 | -0.18 | 2.31 | 3 |
| C14orf132 | 1.16E-03 | 12 | 4 | 0.0052173 | Nucleus_accumbens_basal_ganglia | 7.74 | 3.77E-18 | 0.62 | -2.79 | 1.40 | -0.68 | 1.21 | 4 |
| MPIG6B | 1.16E-03 | 12 | 3 | 0.0006838 | Caudate_basal_ganglia | 8.90 | 3.50E-17 | 0.47 | -2.57 | 3.40 | 0.10 | 1.73 | 3 |
| PWWP2A | 1.16E-03 | 13 | 4 | 0.0004346 | Cerebellar_Hemisphere | 6.38 | 1.57E-18 | 0.43 | -3.52 | 1.06 | -1.51 | 2.14 | 4 |
| SFMBT1 | 1.17E-03 | 13 | 5 | 0.0334205 | Hypothalamus | 6.72 | 8.95E-17 | 0.36 | -1.03 | 2.13 | 0.44 | 1.23 | 5 |
| RP11-154H23.3 | 1.17E-03 | 2 | 2 | 0.0003113 | Spinal_cord_cervical_c-1 | 1.02 | 9.82E-01 | 0.98 | 0.64 | 3.61 | 2.12 | 2.10 | 2 |
| MTMR3 | 1.17E-03 | 10 | 3 | 0.0009011 | Cortex | 7.51 | 1.70E-16 | 0.90 | -3.32 | 1.13 | -0.04 | 1.30 | 3 |
| SAMD5 | 1.18E-03 | 8 | 5 | 0.0001794 | Hypothalamus | 3.54 | 6.41E-17 | 0.27 | -3.75 | 1.71 | -0.60 | 2.10 | 5 |
| DDIT3 | 1.18E-03 | 8 | 3 | 0.0004254 | Frontal_Cortex_BA9 | 6.15 | 4.95E-18 | 0.35 | -3.52 | 2.16 | 0.64 | 1.75 | 3 |
| PNISR | 1.18E-03 | 10 | 2 | 0.000213 | Spinal_cord_cervical_c-1 | 8.36 | 6.13E-18 | 1.64 | -3.14 | 3.70 | 2.68 | 2.14 | 2 |
| C4BPA | 1.19E-03 | 4 | 2 | 0.0003015 | Cortex | 3.02 | 2.32E-63 | 0.98 | -3.61 | -0.24 | -1.08 | 1.69 | 2 |

| | | | | | | | | | | | | | |
|---|---|---|---|---|---|---|---|---|---|---|---|---|---|
| DLC1 | 1.23E-03 | 6 | 4 | 0.004294 | Spinal_cord_cervical_c-1 | 3.53 | 1.79E-17 | 0.48 | -2.72 | 2.86 | -0.11 | 1.79 | 4 |
| TENM4 | 1.25E-03 | 13 | 3 | 0.0060065 | Caudate_basal_ganglia | 9.24 | 2.96E-20 | 0.81 | -1.55 | 2.75 | 0.80 | 1.41 | 3 |
| ZCCHC4 | 1.25E-03 | 2 | 2 | 0.000298 | Nucleus_accumbens_basal_ganglia | 1.92 | 8.45E-02 | 0.08 | 3.10 | 3.62 | 3.36 | 0.37 | 2 |
| ZNF80 | 1.25E-03 | 1 | 1 | 0.001251 | Spinal_cord_cervical_c-1 | 1.00 | 1.00E+00 | 1.00 | -3.23 | -3.23 | -3.23 | NA | 1 |
| RUNDC3B | 1.27E-03 | 4 | 4 | 7.73E-05 | Cerebellar_Hemisphere | 2.19 | 1.96E-01 | 0.20 | -3.95 | 1.30 | -1.99 | 2.31 | 4 |
| SNX14 | 1.27E-03 | 12 | 4 | 0.000536 | Amygdala | 5.38 | 9.91E-19 | 0.85 | -2.44 | 3.46 | -0.75 | 1.87 | 4 |
| LOXL4 | 1.28E-03 | 3 | 2 | 0.0002697 | Spinal_cord_cervical_c-1 | 2.21 | 1.42E-16 | 0.79 | -1.17 | 3.64 | 0.48 | 2.74 | 2 |
| RP11-139I14.2 | 1.30E-03 | 2 | 2 | 0.0003747 | Cerebellum | 1.20 | 8.02E-01 | 0.80 | -3.56 | 0.08 | -1.74 | 2.57 | 2 |
| GAS1RR | 1.30E-03 | 11 | 5 | 0.0002616 | Hippocampus | 6.19 | 4.65E-03 | 0.36 | -3.65 | 1.18 | -2.30 | 1.65 | 5 |
| TMEM107 | 1.30E-03 | 12 | 3 | 0.0001082 | Cerebellum | 8.08 | 4.09E-17 | 1.69 | -3.87 | 0.58 | -1.23 | 1.37 | 3 |
| RARG | 1.31E-03 | 1 | 1 | 0.001308 | Hypothalamus | 1.00 | 1.00E+00 | 1.00 | 3.21 | 3.21 | 3.21 | NA | 1 |
| DBN1 | 1.31E-03 | 6 | 2 | 0.0003084 | Cortex | 5.04 | 2.13E-17 | 0.96 | 1.87 | 3.61 | 3.03 | 0.90 | 2 |
| TRIP12 | 1.32E-03 | 11 | 3 | 0.01645 | Caudate_basal_ganglia | 8.09 | 5.36E-19 | 0.81 | -2.26 | 2.40 | -1.07 | 1.88 | 3 |
| WDR31 | 1.36E-03 | 9 | 5 | 0.0045948 | Cortex | 5.23 | 8.27E-17 | 0.18 | -0.74 | 2.83 | 1.15 | 1.04 | 5 |
| KIF18B | 1.36E-03 | 9 | 3 | 0.03032 | Putamen_basal_ganglia | 5.67 | 9.89E-18 | 1.01 | -2.17 | 2.17 | 0.77 | 1.83 | 3 |
| ADAM22 | 1.38E-03 | 7 | 3 | 0.0004892 | Putamen_basal_ganglia | 5.22 | 3.87E-17 | 0.65 | -3.49 | 1.33 | -1.89 | 1.50 | 3 |
| WDR35 | 1.40E-03 | 13 | 4 | 0.0001329 | Hippocampus | 8.11 | 1.76E-17 | 0.48 | -3.82 | -0.79 | -2.79 | 0.98 | 4 |
| ALOX15 | 1.41E-03 | 8 | 2 | 0.0841392 | Substantia_nigra | 7.41 | 8.55E-18 | 0.29 | -0.78 | 1.73 | 0.99 | 0.77 | 2 |
| CEP164 | 1.42E-03 | 13 | 3 | 0.0066423 | Caudate_basal_ganglia | 8.98 | 5.81E-18 | 0.66 | -1.09 | 2.71 | 1.11 | 1.32 | 3 |
| DNAH14 | 1.42E-03 | 3 | 3 | 0.0001037 | Anterior_cingulate_cortex_BA24 | 2.09 | 2.61E-01 | 0.26 | -3.88 | -0.99 | -2.34 | 1.45 | 3 |
| ECSIT | 1.42E-03 | 12 | 1 | 0.0009904 | Anterior_cingulate_cortex_BA24 | 11.65 | 9.38E-18 | 11.65 | 3.14 | 3.29 | 3.15 | 0.05 | 1 |
| ZSCAN26 | 1.42E-03 | 13 | 3 | 0.0002106 | Hippocampus | 11.52 | 7.63E-18 | 0.42 | -3.71 | -0.60 | -3.11 | 0.82 | 3 |
| DDX5 | 1.44E-03 | 9 | 1 | 0.001438 | Frontal_Cortex_BA9 | 9.00 | 1.64E-34 | 9.00 | -3.19 | -3.19 | -3.19 | 0.00 | 1 |
| CEP95 | 1.44E-03 | 10 | 1 | 0.001438 | Frontal_Cortex_BA9 | 10.00 | 1.37E-17 | 10.00 | 3.19 | 3.19 | 3.19 | 0.00 | 1 |
| SLFNL1 | 1.44E-03 | 13 | 4 | 0.0003338 | Cerebellar_Hemisphere | 9.82 | 1.91E-17 | 0.63 | -2.14 | 3.59 | 2.43 | 1.86 | 4 |
| CHPF | 1.45E-03 | 8 | 3 | 0.0004182 | Substantia_nigra | 5.69 | 1.64E-17 | 1.00 | -1.30 | 3.53 | 1.50 | 1.64 | 3 |
| ADARB1 | 1.48E-03 | 12 | 6 | 9.43E-05 | Caudate_basal_ganglia | 6.45 | 4.00E-17 | 0.44 | -2.10 | 3.90 | 0.68 | 1.70 | 6 |
| ITK | 1.50E-03 | 9 | 2 | 0.0004358 | Substantia_nigra | 8.01 | 4.62E-17 | 0.99 | -3.52 | 3.52 | 1.68 | 3.06 | 2 |
| SCMH1 | 1.50E-03 | 12 | 3 | 0.0003094 | Cerebellum | 5.55 | 4.26E-18 | 2.09 | -3.61 | 2.13 | -1.20 | 2.01 | 3 |
| APBB1 | 1.54E-03 | 11 | 5 | 0.0001487 | Caudate_basal_ganglia | 5.41 | 3.31E-17 | 0.61 | -3.79 | 2.16 | -0.92 | 2.53 | 5 |
| SUPT16H | 1.56E-03 | 5 | 2 | 0.006896 | Cortex | 4.00 | 3.13E-23 | 1.00 | -2.49 | 2.70 | -1.45 | 2.32 | 2 |
| SYCP3 | 1.57E-03 | 12 | 3 | 0.0007237 | Anterior_cingulate_cortex_BA24 | 8.72 | 4.83E-18 | 1.18 | -0.70 | 3.38 | 0.67 | 1.54 | 3 |
| ZC3H8 | 1.62E-03 | 9 | 5 | 4.92E-05 | Caudate_basal_ganglia | 5.98 | 4.72E-17 | 0.41 | -0.41 | 4.06 | 2.72 | 1.63 | 5 |
| RP11-301G7.1 | 1.62E-03 | 7 | 2 | 0.0003399 | Amygdala | 6.02 | 3.25E-17 | 0.98 | 0.07 | 3.58 | 3.06 | 1.32 | 2 |
| COX6B1 | 1.63E-03 | 12 | 4 | 0.001236 | Amygdala | 8.76 | 3.45E-17 | 0.60 | -3.23 | 1.47 | -1.52 | 1.13 | 4 |
| CDKN2AIP | 1.65E-03 | 4 | 2 | 0.001413 | Amygdala | 3.10 | 1.12E-16 | 0.90 | -0.74 | 3.19 | 2.21 | 1.97 | 2 |
| SNX27 | 1.66E-03 | 12 | 3 | 9.41E-05 | Cerebellum | 10.64 | 1.86E-17 | 0.44 | -3.91 | -2.47 | -3.54 | 0.50 | 3 |

| | | | | | | | | | | | | | |
|---|---|---|---|---|---|---|---|---|---|---|---|---|---|
| ODF3B | 1.67E-03 | 7 | 5 | 0.0008918 | Hypothalamus | 3.09 | 3.07E-04 | 0.14 | -2.72 | 3.32 | 0.71 | 2.54 | 5 |
| MED19 | 1.69E-03 | 8 | 3 | 0.0002299 | Cerebellum | 4.98 | 6.83E-19 | 0.87 | -0.19 | 3.68 | 0.90 | 1.27 | 3 |
| CYP21A2 | 1.70E-03 | 4 | 3 | 0.001841 | Cerebellar_Hemisphere | 2.07 | 6.51E-02 | 0.44 | -2.73 | 3.11 | 0.60 | 2.73 | 3 |
| HSPA1A | 1.70E-03 | 3 | 1 | 0.001704 | Cerebellar_Hemisphere | 3.00 | 6.09E-17 | 3.00 | -3.14 | -3.14 | -3.14 | 0.00 | 1 |
| CDC27 | 1.71E-03 | 12 | 2 | 0.0004104 | Cerebellum | 7.06 | 3.34E-33 | 4.94 | -3.53 | 3.53 | -1.01 | 2.62 | 2 |
| GPANK1 | 1.71E-03 | 9 | 4 | 0.0001304 | Cortex | 3.97 | 1.96E-18 | 1.06 | -3.83 | 1.43 | -0.91 | 1.69 | 4 |
| ZSCAN31 | 1.72E-03 | 13 | 2 | 4.78E-05 | Spinal_cord_cervical_c-1 | 10.83 | 3.24E-17 | 1.79 | 1.87 | 4.07 | 3.20 | 0.64 | 2 |
| NDUFS3 | 1.72E-03 | 12 | 3 | 0.0009762 | Cortex | 6.69 | 4.27E-18 | 0.43 | 0.89 | 3.30 | 2.46 | 0.76 | 3 |
| GMDS | 1.72E-03 | 7 | 3 | 0.001983 | Caudate_basal_ganglia | 5.02 | 1.17E-30 | 0.93 | -3.09 | -0.59 | -2.57 | 0.97 | 3 |
| CTD-2534I21.8 | 1.73E-03 | 4 | 1 | 0.001735 | Caudate_basal_ganglia | 4.00 | 2.97E-17 | 4.00 | -3.13 | -3.13 | -3.13 | 0.00 | 1 |
| SUFU | 1.75E-03 | 10 | 2 | 0.000374 | Cortex | 8.67 | 2.77E-17 | 1.11 | 2.88 | 3.56 | 3.24 | 0.34 | 2 |
| SLX4 | 1.75E-03 | 11 | 2 | 0.0003494 | Frontal_Cortex_BA9 | 8.08 | 3.75E-19 | 2.85 | -0.21 | 3.58 | 1.03 | 1.60 | 2 |
| AMD1 | 1.76E-03 | 9 | 2 | 0.0022 | Anterior_cingulate_cortex_BA24 | 6.06 | 1.54E-18 | 2.94 | -3.06 | 3.06 | 1.18 | 2.51 | 2 |
| CBR3 | 1.78E-03 | 13 | 2 | 8.75E-05 | Cortex | 10.92 | 1.29E-17 | 1.56 | 1.53 | 3.92 | 3.08 | 0.68 | 2 |
| ZKSCAN3 | 1.78E-03 | 13 | 4 | 0.0007432 | Hippocampus | 9.04 | 9.22E-18 | 0.33 | -3.37 | 0.83 | -1.73 | 1.50 | 4 |
| STK35 | 1.80E-03 | 12 | 4 | 0.0004438 | Cerebellum | 7.65 | 1.86E-17 | 0.72 | -1.88 | 3.51 | 0.61 | 1.45 | 4 |
| RP11-275H4.1 | 1.81E-03 | 13 | 1 | 0.001092 | Cerebellar_Hemisphere | 12.79 | 7.75E-18 | 12.79 | 2.88 | 3.27 | 3.09 | 0.10 | 1 |
| SH2B3 | 1.83E-03 | 13 | 1 | 0.001834 | Amygdala | 13.00 | 2.36E-17 | 13.00 | 3.12 | 3.12 | 3.12 | 0.00 | 1 |
| SPEN | 1.85E-03 | 4 | 2 | 0.0004568 | Amygdala | 3.53 | 5.22E-49 | 0.47 | 1.94 | 3.50 | 2.33 | 0.78 | 2 |
| ADK | 1.87E-03 | 11 | 3 | 1.44E-05 | Cortex | 8.06 | 2.48E-17 | 0.96 | -4.34 | 3.82 | 0.72 | 2.77 | 3 |
| CTC1 | 1.88E-03 | 9 | 4 | 0.0105269 | Spinal_cord_cervical_c-1 | 5.77 | 1.18E-16 | 0.33 | -2.56 | 1.77 | 0.01 | 1.42 | 4 |
| KRBA2 | 1.90E-03 | 12 | 2 | 0.0004863 | Hippocampus | 9.88 | 2.92E-18 | 2.02 | -3.49 | 0.23 | -1.02 | 1.32 | 2 |
| AJUBA | 1.90E-03 | 13 | 2 | 0.001003 | Nucleus_accumbens_basal_ganglia | 11.68 | 1.38E-17 | 1.14 | -2.87 | 3.29 | 1.64 | 1.77 | 2 |
| PUM1 | 1.90E-03 | 9 | 1 | 0.001903 | Anterior_cingulate_cortex_BA24 | 9.00 | 6.23E-33 | 9.00 | 3.10 | 3.10 | 3.10 | 0.00 | 1 |
| CCL25 | 1.94E-03 | 8 | 4 | 0.0014379 | Cerebellar_Hemisphere | 4.60 | 8.45E-17 | 0.91 | -3.19 | -1.56 | -2.61 | 0.53 | 4 |
| TLR5 | 1.94E-03 | 8 | 2 | 0.006257 | Spinal_cord_cervical_c-1 | 7.71 | 4.09E-17 | 0.29 | -2.73 | -1.01 | -2.17 | 0.58 | 2 |
| SLC39A13 | 1.94E-03 | 12 | 2 | 0.001686 | Cerebellar_Hemisphere | 10.98 | 2.34E-17 | 0.91 | 1.17 | 3.14 | 1.66 | 0.79 | 2 |
| IQCK | 1.96E-03 | 6 | 2 | 0.0004161 | Hippocampus | 5.12 | 5.23E-17 | 0.88 | -3.53 | 3.53 | 0.76 | 3.46 | 2 |
| CKMT1A | 1.96E-03 | 9 | 1 | 0.001897 | Anterior_cingulate_cortex_BA24 | 8.72 | 6.76E-18 | 8.72 | -3.11 | 2.96 | -2.39 | 2.01 | 1 |
| CXCL5 | 2.00E-03 | 11 | 1 | 0.002021 | Anterior_cingulate_cortex_BA24 | 10.81 | 1.38E-17 | 10.81 | -3.09 | -3.04 | -3.06 | 0.02 | 1 |
| NADK | 2.00E-03 | 11 | 4 | 0.0007189 | Cortex | 7.07 | 2.22E-16 | 0.41 | -3.38 | 1.15 | -1.73 | 1.65 | 4 |
| PSMC3 | 2.02E-03 | 11 | 2 | 0.008984 | Caudate_basal_ganglia | 9.33 | 4.20E-18 | 1.50 | -2.61 | 1.17 | 0.14 | 1.40 | 2 |
| RAD51 | 2.03E-03 | 7 | 3 | 0.0004889 | Cortex | 4.73 | 6.75E-17 | 1.08 | -3.49 | 0.82 | -0.79 | 1.50 | 3 |
| SKIV2L | 2.04E-03 | 12 | 2 | 0.0005655 | Cerebellum | 10.81 | 4.36E-17 | 1.13 | -3.45 | 0.05 | -0.91 | 1.02 | 2 |
| NKIRAS1 | 2.04E-03 | 12 | 3 | 0.0002788 | Putamen_basal_ganglia | 9.53 | 9.82E-17 | 0.80 | 1.62 | 3.63 | 3.06 | 0.57 | 3 |
| PLTP | 2.06E-03 | 13 | 5 | 4.41E-05 | Frontal_Cortex_BA9 | 7.84 | 8.08E-17 | 0.27 | 0.50 | 4.09 | 3.02 | 1.18 | 5 |
| TLE6 | 2.09E-03 | 10 | 4 | 0.0041712 | Anterior_cingulate_cortex_BA24 | 6.91 | 1.48E-16 | 0.28 | -2.46 | 2.86 | 1.23 | 1.54 | 4 |

| | | | | | | | | | | | | | |
|---|---|---|---|---|---|---|---|---|---|---|---|---|---|
| FAM180B | 2.10E-03 | 13 | 4 | 0.0001754 | Hypothalamus | 8.41 | 2.72E-17 | 0.34 | -2.88 | 3.75 | 1.56 | 2.14 | 4 |
| TRIM33 | 2.15E-03 | 13 | 4 | 6.29E-05 | Putamen_basal_ganglia | 8.59 | 5.05E-17 | 0.33 | -1.87 | 4.00 | 1.47 | 1.31 | 4 |
| RP11-449D8.5 | 2.16E-03 | 2 | 1 | 0.001941 | Cerebellum | 2.00 | 3.58E-03 | 2.00 | 3.03 | 3.10 | 3.07 | 0.05 | 1 |
| FAM20B | 2.16E-03 | 13 | 3 | 0.00274 | Cerebellum | 11.23 | 3.21E-17 | 0.59 | -3.00 | 0.64 | -1.15 | 0.84 | 3 |
| ZKSCAN4 | 2.18E-03 | 11 | 3 | 0.0004499 | Amygdala | 9.71 | 5.37E-17 | 0.40 | 2.96 | 3.51 | 3.45 | 0.16 | 3 |
| CACFD1 | 2.20E-03 | 12 | 4 | 0.003322 | Hippocampus | 7.02 | 1.15E-17 | 0.73 | -1.77 | 2.94 | 0.91 | 1.71 | 4 |
| GMDS-AS1 | 2.22E-03 | 12 | 4 | 0.001983 | Anterior_cingulate_cortex_BA24 | 6.10 | 2.14E-17 | 0.34 | -3.09 | 3.09 | -1.52 | 1.89 | 4 |
| MMP21 | 2.22E-03 | 6 | 2 | 0.0003133 | Hippocampus | 4.79 | 1.64E-17 | 0.94 | 0.45 | 3.60 | 1.49 | 1.23 | 2 |
| RHBDD3 | 2.23E-03 | 13 | 4 | 0.000383 | Frontal_Cortex_BA9 | 10.16 | 5.82E-17 | 0.44 | -0.29 | 3.55 | 0.51 | 0.93 | 4 |
| CEP290 | 2.23E-03 | 13 | 3 | 0.0004038 | Nucleus_accumbens_basal_ganglia | 10.77 | 4.34E-17 | 1.03 | 1.26 | 3.54 | 2.70 | 0.63 | 3 |
| ATXN2 | 2.25E-03 | 8 | 1 | 0.002245 | Amygdala | 8.00 | 1.64E-17 | 8.00 | 3.06 | 3.06 | 3.06 | 0.00 | 1 |
| CHPT1 | 2.29E-03 | 13 | 2 | 0.0052453 | Anterior_cingulate_cortex_BA24 | 11.64 | 9.48E-07 | 0.68 | -2.79 | -0.70 | -1.89 | 0.73 | 2 |
| C11orf58 | 2.30E-03 | 12 | 3 | 0.0016216 | Hypothalamus | 8.36 | 1.98E-17 | 0.87 | 1.63 | 3.15 | 2.25 | 0.56 | 3 |
| RBPMS | 2.30E-03 | 8 | 3 | 0.001209 | Cerebellum | 6.17 | 1.83E-17 | 0.77 | -1.97 | 3.24 | -0.72 | 2.03 | 3 |
| TBC1D14 | 2.31E-03 | 2 | 2 | 0.005289 | Putamen_basal_ganglia | 1.02 | 9.79E-01 | 0.98 | -2.03 | 2.79 | 0.38 | 3.41 | 2 |
| PSMC6 | 2.33E-03 | 10 | 1 | 0.002333 | Cerebellum | 10.00 | 1.32E-32 | 10.00 | -3.04 | -3.04 | -3.04 | 0.00 | 1 |
| LINC01336 | 2.34E-03 | 3 | 3 | 0.0003502 | Putamen_basal_ganglia | 1.60 | 3.81E-01 | 0.38 | 1.18 | 3.58 | 2.40 | 1.20 | 3 |
| LINC01556 | 2.35E-03 | 8 | 1 | 0.002335 | Nucleus_accumbens_basal_ganglia | 8.00 | 5.14E-18 | 8.00 | 3.04 | 3.04 | 3.04 | 0.00 | 1 |
| RP11-392O17.2 | 2.36E-03 | 11 | 3 | 0.00522 | Anterior_cingulate_cortex_BA24 | 6.71 | 3.18E-18 | 1.39 | -2.79 | 2.40 | -0.15 | 1.85 | 3 |
| EHMT2 | 2.38E-03 | 5 | 3 | 0.000361 | Caudate_basal_ganglia | 3.27 | 2.87E-18 | 0.68 | -3.57 | 2.81 | -1.63 | 2.84 | 3 |
| RP11-638L3.1 | 2.38E-03 | 1 | 1 | 0.002383 | Nucleus_accumbens_basal_ganglia | 1.00 | 1.00E+00 | 1.00 | -3.04 | -3.04 | -3.04 | NA | 1 |
| CCR7 | 2.39E-03 | 2 | 1 | 0.002385 | Amygdala | 2.00 | 3.35E-17 | 2.00 | -3.04 | 3.04 | 0.00 | 4.30 | 1 |
| RP11-890B15.3 | 2.40E-03 | 12 | 4 | 0.0078678 | Amygdala | 8.20 | 7.05E-17 | 0.58 | -2.66 | -0.06 | -1.36 | 0.91 | 4 |
| IGSF3 | 2.42E-03 | 1 | 1 | 0.002421 | Frontal_Cortex_BA9 | 1.00 | 1.00E+00 | 1.00 | -3.03 | -3.03 | -3.03 | NA | 1 |
| CTD-2325M2.1 | 2.43E-03 | 13 | 2 | 0.0005325 | Nucleus_accumbens_basal_ganglia | 10.78 | 1.07E-17 | 2.03 | -3.46 | -2.79 | -3.15 | 0.26 | 2 |
| ANKRD63 | 2.44E-03 | 2 | 1 | 0.00224 | Cerebellum | 1.96 | 4.41E-02 | 1.96 | -3.06 | -2.94 | -3.00 | 0.08 | 1 |
| FLRT1 | 2.45E-03 | 11 | 4 | 0.01747 | Cerebellum | 7.94 | 7.78E-17 | 0.74 | -2.26 | 2.38 | 1.77 | 1.36 | 4 |
| VEZF1 | 2.49E-03 | 10 | 3 | 0.0002117 | Hypothalamus | 5.14 | 2.90E-33 | 0.71 | -1.94 | 3.70 | 0.00 | 2.09 | 3 |
| RSAD1 | 2.49E-03 | 12 | 4 | 0.002359 | Substantia_nigra | 8.34 | 1.55E-17 | 0.31 | 1.41 | 3.04 | 2.50 | 0.54 | 4 |
| TSTD3 | 2.50E-03 | 10 | 3 | 0.0198391 | Anterior_cingulate_cortex_BA24 | 5.78 | 1.97E-18 | 0.64 | -2.33 | 1.60 | -1.16 | 1.25 | 3 |
| KLHL24 | 2.51E-03 | 8 | 1 | 0.002134 | Amygdala | 7.92 | 1.66E-16 | 7.92 | -3.07 | -2.92 | -3.01 | 0.07 | 1 |
| FAM57B | 2.58E-03 | 13 | 1 | 0.002575 | Putamen_basal_ganglia | 13.00 | 1.50E-17 | 13.00 | 3.01 | 3.01 | 3.01 | 0.00 | 1 |
| ZBTB17 | 2.58E-03 | 5 | 2 | 0.003146 | Cortex | 4.69 | 4.23E-17 | 0.31 | -2.95 | -1.28 | -2.62 | 0.75 | 2 |
| FAM174A | 2.60E-03 | 4 | 1 | 0.002534 | Cortex | 4.00 | 6.32E-17 | 4.00 | 3.01 | 3.02 | 3.01 | 0.01 | 1 |
| NR1D2 | 2.63E-03 | 7 | 1 | 0.002629 | Putamen_basal_ganglia | 7.00 | 6.98E-17 | 7.00 | -3.01 | -3.01 | -3.01 | 0.00 | 1 |
| DYNC1I2 | 2.63E-03 | 8 | 3 | 0.0004442 | Cortex | 6.17 | 7.92E-17 | 0.70 | -1.46 | 3.51 | 2.30 | 2.13 | 3 |
| VASH2 | 2.65E-03 | 8 | 2 | 0.002842 | Nucleus_accumbens_basal_ganglia | 7.02 | 4.31E-17 | 0.98 | -1.34 | 2.98 | -0.13 | 1.75 | 2 |

| | | | | | | | | | | | | | |
|---|---|---|---|---|---|---|---|---|---|---|---|---|---|
| RP11-227D13.1 | 2.67E-03 | 6 | 3 | 0.0005994 | Frontal_Cortex_BA9 | 4.63 | 9.84E-17 | 0.19 | -3.43 | -2.63 | -3.10 | 0.37 | 3 |
| LGALSL | 2.67E-03 | 9 | 2 | 0.00109 | Anterior_cingulate_cortex_BA24 | 6.33 | 1.95E-17 | 2.67 | -3.27 | 3.27 | -0.52 | 3.00 | 2 |
| RP4-718N17.2 | 2.68E-03 | 1 | 1 | 0.002676 | Nucleus_accumbens_basal_ganglia | 1.00 | 1.00E+00 | 1.00 | -3.00 | -3.00 | -3.00 | NA | 1 |
| MMP9 | 2.68E-03 | 12 | 2 | 0.000331 | Cerebellar_Hemisphere | 10.11 | 1.41E-17 | 1.82 | -3.59 | 3.00 | 0.01 | 1.41 | 2 |
| SPAG9 | 2.70E-03 | 11 | 4 | 0.07425 | Cerebellum | 8.25 | 5.17E-17 | 0.31 | -1.79 | 0.34 | -0.61 | 0.63 | 4 |
| R3HCC1L | 2.71E-03 | 9 | 4 | 0.0005105 | Hippocampus | 6.64 | 2.69E-17 | 0.26 | -3.48 | -1.17 | -2.49 | 1.06 | 4 |
| RGS16 | 2.71E-03 | 10 | 1 | 0.002713 | Amygdala | 10.00 | 3.02E-33 | 10.00 | -3.00 | -3.00 | -3.00 | 0.00 | 1 |
| LINC01750 | 2.71E-03 | 13 | 1 | 0.0018245 | Cortex | 12.69 | 2.21E-17 | 12.69 | -3.12 | -2.86 | -2.96 | 0.09 | 1 |
| IKZF5 | 2.73E-03 | 11 | 4 | 5.85E-05 | Cortex | 5.18 | 2.80E-18 | 0.55 | 0.50 | 4.02 | 2.07 | 1.65 | 4 |
| MMP16 | 2.75E-03 | 10 | 4 | 0.002272 | Cerebellar_Hemisphere | 6.73 | 5.24E-18 | 0.46 | -1.41 | 3.05 | -0.51 | 1.53 | 4 |
| MFAP1 | 2.76E-03 | 13 | 1 | 0.002763 | Amygdala | 13.00 | 2.98E-33 | 13.00 | -2.99 | -2.99 | -2.99 | 0.00 | 1 |
| ZBTB25 | 2.77E-03 | 13 | 3 | 0.0002057 | Hypothalamus | 11.25 | 2.44E-17 | 0.52 | 0.38 | 3.71 | 3.13 | 0.90 | 3 |
| BHLHE22 | 2.80E-03 | 7 | 3 | 0.007793 | Anterior_cingulate_cortex_BA24 | 4.67 | 1.88E-17 | 0.95 | -2.66 | 2.23 | -1.16 | 1.76 | 3 |
| EIF6 | 2.83E-03 | 12 | 3 | 0.04309 | Amygdala | 8.29 | 4.17E-17 | 0.41 | -2.02 | 0.44 | -0.57 | 0.75 | 3 |
| CNN1 | 2.83E-03 | 8 | 2 | 0.0005618 | Hypothalamus | 6.51 | 5.69E-17 | 1.46 | -3.34 | 3.45 | 1.28 | 3.00 | 2 |
| ELAVL1 | 2.83E-03 | 7 | 2 | 0.00417 | Cerebellum | 6.14 | 9.97E-18 | 0.86 | -2.87 | 2.87 | 1.21 | 2.79 | 2 |
| SMARCC2 | 2.85E-03 | 13 | 1 | 0.00213 | Caudate_basal_ganglia | 12.64 | 6.84E-17 | 12.64 | -3.07 | 3.07 | -2.00 | 2.26 | 1 |
| MBOAT4 | 2.88E-03 | 10 | 2 | 0.0008953 | Amygdala | 9.00 | 9.70E-31 | 1.00 | -3.32 | 3.32 | -1.09 | 3.13 | 2 |
| ITGA5 | 2.93E-03 | 6 | 2 | 0.01738 | Cortex | 5.76 | 2.94E-33 | 0.22 | -2.38 | -1.25 | -1.79 | 0.60 | 2 |
| DAXX | 2.95E-03 | 11 | 3 | 0.0074606 | Cerebellar_Hemisphere | 9.07 | 2.70E-18 | 0.77 | -1.66 | 2.68 | -0.65 | 1.72 | 3 |
| PTPMT1 | 2.96E-03 | 13 | 2 | 0.002956 | Cortex | 11.96 | 1.60E-17 | 1.04 | -2.09 | 2.97 | -1.45 | 1.34 | 2 |
| NLRC3 | 2.99E-03 | 13 | 2 | 0.0022127 | Cerebellar_Hemisphere | 11.87 | 2.13E-17 | 0.64 | 1.26 | 3.06 | 1.77 | 0.61 | 2 |
| MYCBPAP | 3.00E-03 | 13 | 3 | 0.001443 | Cortex | 8.83 | 4.75E-18 | 0.62 | -0.59 | 3.19 | 1.49 | 1.39 | 3 |
| WDR76 | 3.06E-03 | 9 | 1 | 0.003061 | Cortex | 9.00 | 6.78E-34 | 9.00 | -2.96 | -2.96 | -2.96 | 0.00 | 1 |
| HAUS4 | 3.08E-03 | 12 | 1 | 0.001628 | Putamen_basal_ganglia | 11.57 | 9.68E-17 | 11.57 | 2.74 | 3.15 | 2.91 | 0.11 | 1 |
| ACSF2 | 3.11E-03 | 12 | 3 | 0.0006983 | Nucleus_accumbens_basal_ganglia | 6.15 | 8.33E-18 | 2.65 | -3.39 | 2.22 | 0.41 | 2.14 | 3 |
| SHPK | 3.12E-03 | 13 | 1 | 0.0008127 | Caudate_basal_ganglia | 12.50 | 1.29E-17 | 12.50 | 2.37 | 3.35 | 2.90 | 0.31 | 1 |
| ASTN1 | 3.14E-03 | 12 | 2 | 0.0006855 | Nucleus_accumbens_basal_ganglia | 10.00 | 4.19E-24 | 2.00 | -3.40 | 0.01 | -0.58 | 1.32 | 2 |
| WBP2NL | 3.14E-03 | 13 | 2 | 0.02483 | Cerebellum | 11.78 | 6.54E-17 | 0.87 | -0.78 | 2.24 | 1.28 | 0.91 | 2 |
| TOMM34 | 3.17E-03 | 8 | 2 | 0.0008928 | Cerebellar_Hemisphere | 5.75 | 2.56E-17 | 2.25 | -3.32 | 3.32 | 0.64 | 2.75 | 2 |
| RP11-418I22.2 | 3.18E-03 | 1 | 1 | 0.003176 | Putamen_basal_ganglia | 1.00 | 1.00E+00 | 1.00 | 2.95 | 2.95 | 2.95 | NA | 1 |
| CARMIL3 | 3.20E-03 | 13 | 4 | 0.0006102 | Hippocampus | 5.76 | 1.35E-17 | 0.70 | -1.20 | 3.43 | 1.69 | 1.78 | 4 |
| TSPAN14 | 3.21E-03 | 9 | 6 | 0.0010764 | Nucleus_accumbens_basal_ganglia | 4.03 | 5.52E-17 | 0.19 | -3.27 | 1.21 | -1.08 | 1.65 | 6 |
| PENK | 3.24E-03 | 4 | 3 | 0.0009671 | Caudate_basal_ganglia | 2.07 | 1.76E-16 | 0.91 | -1.81 | 3.30 | 1.33 | 2.47 | 3 |
| ZNF184 | 3.26E-03 | 11 | 2 | 0.007826 | Putamen_basal_ganglia | 10.60 | 4.00E-17 | 0.40 | -2.66 | -1.53 | -2.35 | 0.52 | 2 |
| CLP1 | 3.27E-03 | 3 | 3 | 0.001388 | Spinal_cord_cervical_c-1 | 2.30 | 8.56E-02 | 0.09 | -3.20 | -0.07 | -2.11 | 1.77 | 3 |
| NELL2 | 3.27E-03 | 11 | 3 | 0.03828 | Hypothalamus | 9.35 | 8.27E-18 | 0.55 | -2.07 | 0.92 | -1.75 | 0.90 | 3 |

| | | | | | | | | | | | | | |
|---|---|---|---|---|---|---|---|---|---|---|---|---|---|
| CRTAC1 | 3.27E-03 | 10 | 1 | 0.00312 | Cortex | 9.99 | 8.77E-18 | 9.99 | -2.96 | -2.80 | -2.94 | 0.05 | 1 |
| RILPL2 | 3.30E-03 | 9 | 3 | 0.0137 | Anterior_cingulate_cortex_BA24 | 7.27 | 1.75E-17 | 0.40 | -2.47 | 1.29 | 0.25 | 1.51 | 3 |
| NKAPL | 3.35E-03 | 10 | 4 | 0.003085 | Anterior_cingulate_cortex_BA24 | 5.67 | 1.32E-17 | 0.49 | -0.25 | 2.96 | 0.76 | 1.15 | 4 |
| CTDNEP1 | 3.37E-03 | 12 | 1 | 0.0008003 | Caudate_basal_ganglia | 11.72 | 1.51E-17 | 11.72 | 2.33 | 3.35 | 2.90 | 0.31 | 1 |
| CUL3 | 3.37E-03 | 9 | 4 | 0.0001601 | Cortex | 4.83 | 1.08E-17 | 0.18 | -3.77 | 1.85 | -1.04 | 1.85 | 4 |
| RPL24 | 3.37E-03 | 8 | 1 | 0.003373 | Caudate_basal_ganglia | 8.00 | 3.68E-17 | 8.00 | -2.93 | -2.93 | -2.93 | 0.00 | 1 |
| HSPB7 | 3.40E-03 | 13 | 6 | 0.0001586 | Substantia_nigra | 5.54 | 6.80E-17 | 0.20 | -2.65 | 3.78 | 0.51 | 1.89 | 6 |
| PLXND1 | 3.40E-03 | 10 | 3 | 0.0002851 | Frontal_Cortex_BA9 | 6.78 | 1.69E-32 | 1.31 | -3.63 | -0.16 | -2.48 | 0.95 | 3 |
| TESK2 | 3.40E-03 | 13 | 3 | 0.0002587 | Cerebellar_Hemisphere | 9.33 | 3.15E-17 | 0.68 | -1.04 | 3.65 | 0.71 | 1.49 | 3 |
| PCNP | 3.42E-03 | 5 | 3 | 0.001686 | Frontal_Cortex_BA9 | 2.64 | 1.42E-17 | 0.63 | -0.18 | 3.14 | 1.78 | 1.79 | 3 |
| ORC5 | 3.42E-03 | 8 | 2 | 0.005992 | Cerebellar_Hemisphere | 6.45 | 6.21E-48 | 1.55 | -1.02 | 2.75 | 0.09 | 1.59 | 2 |
| RHEB | 3.43E-03 | 5 | 2 | 0.0007878 | Hypothalamus | 4.26 | 6.12E-17 | 0.74 | 1.79 | 3.36 | 2.10 | 0.70 | 2 |
| INTS12 | 3.45E-03 | 8 | 3 | 0.006892 | Caudate_basal_ganglia | 4.17 | 5.22E-33 | 0.93 | -2.70 | 1.47 | -1.50 | 1.88 | 3 |
| IBTK | 3.47E-03 | 13 | 5 | 0.0006495 | Hypothalamus | 7.42 | 4.75E-17 | 0.45 | -2.73 | 3.41 | 1.43 | 2.12 | 5 |
| ZBED9 | 3.53E-03 | 11 | 3 | 0.0004599 | Caudate_basal_ganglia | 9.32 | 2.19E-17 | 0.63 | -3.50 | -0.89 | -1.22 | 0.76 | 3 |
| GLT1D1 | 3.54E-03 | 13 | 3 | 0.004431 | Cerebellar_Hemisphere | 9.37 | 8.31E-17 | 0.76 | -2.03 | 2.85 | -0.17 | 1.31 | 3 |
| SLC9A8 | 3.57E-03 | 13 | 3 | 0.0012771 | Spinal_cord_cervical_c-1 | 11.32 | 2.68E-17 | 0.38 | -3.22 | -1.33 | -2.32 | 0.58 | 3 |
| RAB26 | 3.57E-03 | 12 | 2 | 0.001163 | Caudate_basal_ganglia | 8.46 | 5.73E-18 | 3.50 | -3.25 | 3.25 | 0.10 | 2.36 | 2 |
| FOXP1 | 3.61E-03 | 11 | 2 | 0.001012 | Caudate_basal_ganglia | 10.04 | 2.00E-18 | 0.96 | -0.05 | 3.29 | 0.33 | 0.98 | 2 |
| CEP97 | 3.64E-03 | 3 | 3 | 0.001764 | Substantia_nigra | 1.88 | 3.71E-01 | 0.37 | 0.15 | 3.13 | 1.67 | 1.49 | 3 |
| FAM181A | 3.65E-03 | 6 | 2 | 0.002199 | Amygdala | 3.67 | 5.20E-38 | 2.26 | 2.35 | 3.06 | 2.59 | 0.36 | 2 |
| SYTL1 | 3.68E-03 | 13 | 2 | 0.0007463 | Anterior_cingulate_cortex_BA24 | 10.68 | 6.21E-17 | 2.32 | 1.73 | 3.37 | 2.85 | 0.54 | 2 |
| IMMT | 3.74E-03 | 11 | 2 | 0.0009324 | Hippocampus | 8.46 | 2.54E-19 | 2.17 | -0.17 | 3.31 | 1.34 | 1.08 | 2 |
| EVI2B | 3.76E-03 | 13 | 1 | 0.003757 | Anterior_cingulate_cortex_BA24 | 13.00 | 3.82E-32 | 13.00 | -2.90 | -2.90 | -2.90 | 0.00 | 1 |
| NDEL1 | 3.77E-03 | 9 | 5 | 0.0002145 | Putamen_basal_ganglia | 4.98 | 8.19E-17 | 0.25 | -3.70 | 2.18 | 0.46 | 1.73 | 5 |
| THBS4 | 3.77E-03 | 10 | 4 | 0.0004604 | Cerebellum | 7.23 | 1.46E-17 | 0.27 | -3.50 | 1.17 | -0.28 | 1.28 | 4 |
| RP11-51J9.5 | 3.81E-03 | 11 | 2 | 0.0009146 | Cerebellar_Hemisphere | 10.49 | 5.63E-18 | 0.51 | -3.32 | -2.55 | -2.62 | 0.23 | 2 |
| HPX | 3.86E-03 | 13 | 5 | 0.010015 | Cortex | 7.66 | 3.66E-17 | 0.75 | -1.48 | 2.58 | 0.75 | 1.29 | 5 |
| HSPH1 | 3.88E-03 | 12 | 2 | 0.04018 | Amygdala | 10.00 | 1.05E-18 | 1.97 | -0.55 | 2.05 | 1.39 | 1.13 | 2 |
| AK5 | 3.88E-03 | 9 | 3 | 0.001887 | Nucleus_accumbens_basal_ganglia | 6.27 | 4.70E-18 | 0.75 | -3.11 | 1.17 | -0.28 | 1.80 | 3 |
| DYNLL1 | 3.90E-03 | 10 | 5 | 0.03631 | Putamen_basal_ganglia | 6.35 | 6.86E-18 | 0.22 | 0.37 | 2.09 | 1.57 | 0.58 | 5 |
| TMEM14A | 3.91E-03 | 6 | 3 | 0.002784 | Caudate_basal_ganglia | 4.01 | 5.26E-17 | 0.55 | 0.49 | 2.99 | 1.58 | 1.24 | 3 |
| ZNF804A | 3.97E-03 | 11 | 3 | 0.0098012 | Cortex | 6.25 | 1.49E-18 | 1.31 | -1.71 | 2.58 | 0.77 | 1.58 | 3 |
| ERC1 | 4.04E-03 | 8 | 6 | 0.0023105 | Anterior_cingulate_cortex_BA24 | 2.55 | 2.64E-17 | 0.25 | -0.62 | 3.05 | 1.10 | 1.18 | 6 |
| BCL2A1 | 4.07E-03 | 3 | 1 | 0.004068 | Frontal_Cortex_BA9 | 3.00 | 4.80E-17 | 3.00 | -2.87 | -2.87 | -2.87 | 0.00 | 1 |
| ZNF800 | 4.08E-03 | 13 | 2 | 0.0009736 | Hippocampus | 8.94 | 1.02E-18 | 3.96 | -3.30 | 0.39 | -1.03 | 1.60 | 2 |
| GLRX2 | 4.10E-03 | 13 | 3 | 0.0006519 | Cerebellar_Hemisphere | 9.08 | 1.87E-17 | 0.72 | -3.41 | 0.79 | -0.81 | 1.52 | 3 |

| | | | | | | | | | | | | | |
|---|---|---|---|---|---|---|---|---|---|---|---|---|---|
| RP11-142A22.4 | 4.11E-03 | 2 | 1 | 0.004109 | Substantia_nigra | 2.00 | 3.35E-17 | 2.00 | -2.87 | -2.87 | -2.87 | 0.00 | 1 |
| PLPP2 | 4.12E-03 | 13 | 2 | 0.0010464 | Frontal_Cortex_BA9 | 11.89 | 2.90E-17 | 0.76 | 2.33 | 3.28 | 3.12 | 0.27 | 2 |
| SEC14L1 | 4.12E-03 | 3 | 3 | 0.08428 | Nucleus_accumbens_basal_ganglia | 1.99 | 1.01E-01 | 0.10 | -1.73 | -0.18 | -0.98 | 0.78 | 3 |
| MTX3 | 4.15E-03 | 12 | 5 | 0.0004604 | Spinal_cord_cervical_c-1 | 8.03 | 3.00E-17 | 0.40 | -0.35 | 3.50 | 1.88 | 1.18 | 5 |
| GABARAP | 4.16E-03 | 9 | 3 | 0.0007871 | Cerebellum | 7.21 | 1.11E-17 | 0.86 | -3.36 | 3.36 | 0.80 | 3.17 | 3 |
| FAM120AOS | 4.21E-03 | 13 | 4 | 0.0102768 | Cerebellum | 7.50 | 1.92E-17 | 0.60 | -2.57 | 1.89 | 0.96 | 1.25 | 4 |
| EVI2A | 4.21E-03 | 13 | 1 | 0.0036226 | Nucleus_accumbens_basal_ganglia | 12.94 | 1.23E-17 | 12.94 | -2.91 | -2.85 | -2.86 | 0.02 | 1 |
| PCOLCE | 4.22E-03 | 12 | 3 | 0.1387497 | Cortex | 9.90 | 3.71E-17 | 0.47 | -0.85 | 1.48 | -0.07 | 0.76 | 3 |
| GNPTAB | 4.23E-03 | 12 | 3 | 0.0006244 | Cortex | 7.05 | 9.07E-18 | 1.04 | -3.42 | 0.23 | -2.01 | 1.24 | 3 |
| CSNK2B | 4.23E-03 | 11 | 5 | 0.005532 | Nucleus_accumbens_basal_ganglia | 5.01 | 3.15E-52 | 0.36 | -0.22 | 2.77 | 1.20 | 0.93 | 5 |
| CC2D2B | 4.25E-03 | 13 | 4 | 0.0078138 | Cerebellum | 7.76 | 5.91E-17 | 0.29 | 0.29 | 2.66 | 1.25 | 0.68 | 4 |
| GPR1 | 4.26E-03 | 8 | 3 | 0.1128 | Cerebellar_Hemisphere | 6.57 | 5.34E-17 | 0.58 | 1.24 | 1.59 | 1.53 | 0.12 | 3 |
| ZNF787 | 4.26E-03 | 3 | 2 | 0.009737 | Cortex | 2.60 | 2.81E-17 | 0.40 | 0.31 | 2.59 | 1.83 | 1.32 | 2 |
| FAM120A | 4.27E-03 | 7 | 3 | 0.00992 | Amygdala | 5.57 | 3.71E-17 | 0.50 | -1.31 | 2.58 | 1.09 | 1.19 | 3 |
| SLC22A3 | 4.27E-03 | 11 | 4 | 0.01869 | Nucleus_accumbens_basal_ganglia | 8.59 | 2.99E-17 | 0.32 | -1.32 | 2.35 | 1.37 | 1.15 | 4 |
| FGB | 4.27E-03 | 10 | 2 | 0.0141535 | Caudate_basal_ganglia | 7.12 | 2.31E-17 | 2.87 | -0.93 | 2.45 | 1.34 | 1.22 | 2 |
| PCSK1 | 4.28E-03 | 2 | 2 | 0.001234 | Caudate_basal_ganglia | 1.06 | 9.38E-01 | 0.94 | -3.23 | 0.48 | -1.37 | 2.63 | 2 |
| PGBD1 | 4.30E-03 | 12 | 4 | 0.0001635 | Spinal_cord_cervical_c-1 | 5.23 | 4.42E-17 | 0.43 | -2.00 | 3.77 | 1.00 | 2.04 | 4 |
| CYP3A5 | 4.31E-03 | 13 | 2 | 0.00197 | Caudate_basal_ganglia | 11.28 | 1.67E-18 | 1.62 | 1.52 | 3.09 | 2.94 | 0.43 | 2 |
| RP11-26J3.1 | 4.31E-03 | 11 | 2 | 0.004781 | Amygdala | 9.65 | 3.68E-20 | 1.04 | -1.96 | 2.82 | 2.34 | 1.43 | 2 |
| RP11-13A1.1 | 4.36E-03 | 9 | 3 | 0.003802 | Frontal_Cortex_BA9 | 7.45 | 1.13E-17 | 0.56 | -2.89 | 1.77 | -2.05 | 1.49 | 3 |
| MAP3K20 | 4.37E-03 | 12 | 5 | 0.000123 | Hippocampus | 6.02 | 4.92E-18 | 0.39 | -3.84 | 1.37 | -0.97 | 2.19 | 5 |
| MAPK8IP2 | 4.39E-03 | 11 | 5 | 5.74E-05 | Cerebellar_Hemisphere | 4.97 | 7.65E-18 | 0.35 | -4.02 | 2.92 | -0.98 | 1.96 | 5 |
| OR7C1 | 4.41E-03 | 6 | 3 | 0.02598 | Caudate_basal_ganglia | 4.32 | 3.58E-17 | 0.67 | -0.64 | 2.23 | 0.23 | 1.36 | 3 |
| OTUB2 | 4.42E-03 | 6 | 4 | 0.2098 | Hippocampus | 4.22 | 4.36E-17 | 0.21 | -1.12 | 1.25 | 0.24 | 0.90 | 4 |
| ZSCAN12 | 4.45E-03 | 13 | 5 | 9.46E-05 | Frontal_Cortex_BA9 | 6.12 | 1.17E-17 | 0.43 | -3.75 | 3.90 | -0.53 | 2.22 | 5 |
| CTNS | 4.46E-03 | 10 | 4 | 0.004037 | Cerebellar_Hemisphere | 8.04 | 2.77E-17 | 0.33 | -1.51 | 2.88 | -0.33 | 1.30 | 4 |
| ZNF536 | 4.48E-03 | 4 | 2 | 0.006717 | Cerebellum | 3.80 | 1.11E-16 | 0.20 | 1.40 | 2.71 | 2.38 | 0.66 | 2 |
| APOH | 4.48E-03 | 1 | 1 | 0.004481 | Hippocampus | 1.00 | 1.00E+00 | 1.00 | -2.84 | -2.84 | -2.84 | NA | 1 |
| KMT2B | 4.48E-03 | 3 | 3 | 0.0004303 | Caudate_basal_ganglia | 1.54 | 5.71E-01 | 0.57 | -1.40 | 3.52 | 0.33 | 2.76 | 3 |
| MTMR12 | 4.50E-03 | 9 | 3 | 0.001684 | Cerebellar_Hemisphere | 5.83 | 7.67E-17 | 0.93 | -3.14 | 1.61 | -1.74 | 1.35 | 3 |
| TMEM87B | 4.54E-03 | 13 | 5 | 0.0004765 | Cortex | 5.62 | 3.44E-17 | 0.53 | -1.55 | 3.49 | 0.21 | 1.32 | 5 |
| TF | 4.55E-03 | 7 | 3 | 0.003663 | Cerebellum | 4.61 | 6.74E-17 | 0.71 | 1.75 | 2.91 | 2.28 | 0.53 | 3 |
| AP000473.5 | 4.64E-03 | 10 | 4 | 0.0059478 | Anterior_cingulate_cortex_BA24 | 5.87 | 3.28E-17 | 0.47 | -0.65 | 2.75 | 0.79 | 0.93 | 4 |
| GGNBP2 | 4.68E-03 | 12 | 2 | 0.0006154 | Amygdala | 10.47 | 3.42E-17 | 1.18 | -3.42 | -0.12 | -2.94 | 0.91 | 2 |
| GPC6 | 4.68E-03 | 12 | 3 | 0.0085881 | Caudate_basal_ganglia | 7.17 | 7.00E-18 | 1.42 | -2.63 | 1.55 | 0.23 | 1.34 | 3 |
| RP11-463I20.3 | 4.68E-03 | 1 | 1 | 0.004685 | Nucleus_accumbens_basal_ganglia | 1.00 | 1.00E+00 | 1.00 | -2.83 | -2.83 | -2.83 | NA | 1 |

| | | | | | | | | | | | | | |
|---|---|---|---|---|---|---|---|---|---|---|---|---|---|
| LIN37 | 4.70E-03 | 12 | 2 | 0.001517 | Cortex | 11.38 | 3.94E-17 | 0.62 | 2.54 | 3.17 | 3.12 | 0.18 | 2 |
| ADAMTS3 | 4.70E-03 | 8 | 3 | 0.0005736 | Putamen_basal_ganglia | 5.79 | 1.63E-18 | 0.22 | 0.69 | 3.44 | 1.42 | 1.25 | 3 |
| NDUFS1 | 4.71E-03 | 13 | 1 | 0.002118 | Frontal_Cortex_BA9 | 12.36 | 2.40E-17 | 12.36 | 2.38 | 3.07 | 2.75 | 0.21 | 1 |
| PRKCA | 4.73E-03 | 8 | 6 | 0.005818 | Spinal_cord_cervical_c-1 | 3.45 | 2.12E-16 | 0.46 | -0.94 | 2.76 | 0.20 | 1.44 | 6 |
| CTD-2035E11.5 | 4.77E-03 | 4 | 2 | 0.004747 | Cerebellar_Hemisphere | 3.08 | 2.18E-18 | 0.92 | -2.82 | 2.82 | -1.27 | 2.74 | 2 |
| CCSER1 | 4.80E-03 | 3 | 2 | 0.04884 | Caudate_basal_ganglia | 2.31 | 1.87E-16 | 0.69 | -1.97 | 1.43 | -0.84 | 1.96 | 2 |
| RP5-978I12.1 | 4.85E-03 | 3 | 2 | 0.002723 | Frontal_Cortex_BA9 | 2.35 | 1.56E-16 | 0.65 | -3.00 | -0.33 | -1.22 | 1.54 | 2 |
| OR2H2 | 4.86E-03 | 12 | 7 | 0.04709 | Hippocampus | 5.00 | 3.23E-18 | 0.27 | -1.99 | 1.53 | 0.11 | 1.20 | 7 |
| GNB1 | 4.86E-03 | 11 | 3 | 0.0005449 | Substantia_nigra | 7.80 | 7.11E-17 | 1.01 | -2.16 | 3.46 | 0.88 | 1.75 | 3 |
| FAM53A | 4.88E-03 | 13 | 4 | 0.0001794 | Hypothalamus | 9.64 | 1.30E-17 | 0.43 | -3.75 | -0.92 | -2.94 | 0.91 | 4 |
| DCTN6 | 4.88E-03 | 12 | 2 | 0.0009146 | Frontal_Cortex_BA9 | 11.45 | 4.66E-17 | 0.53 | 2.48 | 3.32 | 2.67 | 0.22 | 2 |
| TMEM51 | 4.91E-03 | 8 | 3 | 0.0043692 | Cerebellar_Hemisphere | 5.36 | 3.40E-17 | 0.45 | -2.85 | 0.68 | -2.10 | 1.14 | 3 |
| CATSPER2 | 5.03E-03 | 1 | 1 | 0.005026 | Hippocampus | 1.00 | 1.00E+00 | 1.00 | -2.81 | -2.81 | -2.81 | NA | 1 |
| FBN1 | 5.04E-03 | 7 | 2 | 0.00443 | Putamen_basal_ganglia | 5.88 | 4.86E-17 | 1.05 | -2.85 | -0.32 | -2.15 | 1.18 | 2 |
| TMEM117 | 5.05E-03 | 12 | 3 | 0.0004411 | Amygdala | 10.20 | 6.95E-18 | 0.77 | -3.51 | 0.48 | -2.94 | 1.23 | 3 |
| C18orf21 | 5.06E-03 | 9 | 1 | 0.005064 | Caudate_basal_ganglia | 9.00 | 7.35E-33 | 9.00 | -2.80 | 2.80 | -2.18 | 1.87 | 1 |
| HEY1 | 5.07E-03 | 1 | 1 | 0.005066 | Hypothalamus | 1.00 | 1.00E+00 | 1.00 | -2.80 | -2.80 | -2.80 | NA | 1 |
| SGCZ | 5.09E-03 | 3 | 1 | 0.001532 | Cerebellar_Hemisphere | 2.92 | 2.25E-16 | 2.92 | -3.17 | -2.57 | -2.77 | 0.35 | 1 |
| LEMD2 | 5.10E-03 | 12 | 3 | 0.0900299 | Cortex | 9.80 | 7.44E-17 | 0.67 | -1.31 | 1.70 | 0.34 | 1.11 | 3 |
| HSDL2 | 5.11E-03 | 7 | 3 | 0.0008045 | Substantia_nigra | 5.26 | 6.45E-17 | 0.28 | -1.90 | 3.35 | -0.69 | 2.14 | 3 |
| AP003774.6 | 5.12E-03 | 13 | 2 | 0.0013049 | Hypothalamus | 12.05 | 2.06E-17 | 0.95 | -3.21 | -1.97 | -3.10 | 0.34 | 2 |
| FADS1 | 5.13E-03 | 12 | 3 | 0.01305 | Nucleus_accumbens_basal_ganglia | 10.68 | 1.71E-17 | 0.41 | -2.48 | -0.16 | -1.17 | 0.66 | 3 |
| AC007405.6 | 5.13E-03 | 10 | 2 | 0.0005395 | Anterior_cingulate_cortex_BA24 | 8.99 | 1.13E-16 | 0.62 | -3.46 | -2.64 | -2.90 | 0.28 | 2 |
| SPATA2 | 5.13E-03 | 11 | 2 | 0.0325842 | Substantia_nigra | 9.82 | 4.62E-17 | 1.00 | -1.48 | 2.14 | 0.46 | 1.07 | 2 |
| SPTY2D1-AS1 | 5.18E-03 | 3 | 1 | 0.005176 | Substantia_nigra | 3.00 | 1.11E-16 | 3.00 | 2.80 | 2.80 | 2.80 | 0.00 | 1 |
| RP11-159D12.2 | 5.18E-03 | 6 | 2 | 0.001185 | Anterior_cingulate_cortex_BA24 | 5.35 | 3.71E-17 | 0.65 | -3.24 | -1.91 | -3.00 | 0.53 | 2 |
| FIGN | 5.19E-03 | 9 | 3 | 0.004248 | Hypothalamus | 5.16 | 1.22E-18 | 0.95 | -2.86 | 0.94 | -2.19 | 1.23 | 3 |
| U2AF1L4 | 5.22E-03 | 13 | 2 | 0.0013828 | Spinal_cord_cervical_c-1 | 11.22 | 3.47E-17 | 1.16 | -0.46 | 3.20 | 2.57 | 1.01 | 2 |
| CRISP2 | 5.25E-03 | 3 | 2 | 0.0119379 | Nucleus_accumbens_basal_ganglia | 2.62 | 4.06E-17 | 0.38 | -0.34 | 2.51 | 0.61 | 1.65 | 2 |
| PIK3R3 | 5.25E-03 | 13 | 2 | 0.0009705 | Anterior_cingulate_cortex_BA24 | 9.13 | 9.11E-18 | 3.86 | -3.30 | 2.51 | -1.78 | 2.00 | 2 |
| GAD1 | 5.28E-03 | 13 | 4 | 0.0191871 | Putamen_basal_ganglia | 9.20 | 4.31E-18 | 0.49 | -2.34 | 1.81 | -0.72 | 1.13 | 4 |
| C1orf109 | 5.28E-03 | 13 | 3 | 0.0117596 | Cortex | 11.68 | 5.31E-17 | 0.42 | -2.52 | -0.51 | -1.29 | 0.71 | 3 |
| TGFBR1 | 5.30E-03 | 3 | 2 | 0.001231 | Substantia_nigra | 2.70 | 3.42E-33 | 0.30 | -3.23 | 2.62 | -1.28 | 3.38 | 2 |
| C4B | 5.31E-03 | 8 | 5 | 5.00E-05 | Cortex | 2.90 | 2.15E-16 | 0.22 | 0.42 | 4.06 | 1.46 | 1.33 | 5 |
| SELENON | 5.33E-03 | 8 | 2 | 0.001582 | Caudate_basal_ganglia | 6.93 | 7.04E-18 | 1.00 | -3.16 | 1.21 | -2.50 | 1.50 | 2 |
| PTRH1 | 5.35E-03 | 10 | 2 | 0.1358 | Frontal_Cortex_BA9 | 9.55 | 7.01E-17 | 0.41 | -1.49 | -0.01 | -0.52 | 0.68 | 2 |
| GOLIM4 | 5.36E-03 | 10 | 3 | 0.0005591 | Putamen_basal_ganglia | 7.97 | 1.16E-17 | 0.71 | -3.45 | -0.34 | -1.91 | 0.79 | 3 |

| | | | | | | | | | | | | | |
|---|---|---|---|---|---|---|---|---|---|---|---|---|---|
| C6orf226 | 5.39E-03 | 13 | 1 | 0.005391 | Putamen_basal_ganglia | 13.00 | 7.06E-17 | 13.00 | 2.78 | 2.78 | 2.78 | 0.00 | 1 |
| CD40 | 5.42E-03 | 13 | 3 | 0.03464 | Cerebellum | 11.00 | 2.54E-17 | 0.63 | -2.11 | 0.79 | -0.41 | 0.80 | 3 |
| PRC1 | 5.42E-03 | 12 | 4 | 0.0001728 | Nucleus_accumbens_basal_ganglia | 7.28 | 6.33E-18 | 0.73 | -1.25 | 3.76 | 0.45 | 2.02 | 4 |
| TMEM198 | 5.48E-03 | 9 | 4 | 0.0004182 | Cerebellar_Hemisphere | 5.40 | 8.53E-17 | 0.99 | -3.53 | 1.95 | 0.29 | 1.84 | 4 |
| LINC00354 | 5.51E-03 | 2 | 1 | 0.005507 | Caudate_basal_ganglia | 2.00 | 3.35E-17 | 2.00 | -2.78 | -2.78 | -2.78 | 0.00 | 1 |
| RNF139 | 5.55E-03 | 6 | 2 | 0.0106 | Cerebellar_Hemisphere | 5.01 | 2.70E-17 | 0.99 | 2.17 | 2.56 | 2.23 | 0.16 | 2 |
| ZNHIT3 | 5.61E-03 | 11 | 2 | 0.001259 | Anterior_cingulate_cortex_BA24 | 10.19 | 4.46E-17 | 0.67 | -3.23 | 3.23 | 1.91 | 2.49 | 2 |
| SLITRK5 | 5.62E-03 | 6 | 2 | 0.002195 | Cerebellum | 5.00 | 3.71E-86 | 1.00 | -1.02 | 3.06 | -0.34 | 1.67 | 2 |
| ENTPD5 | 5.63E-03 | 13 | 5 | 0.0023566 | Cerebellar_Hemisphere | 6.14 | 4.51E-34 | 0.23 | -3.04 | 1.82 | 0.86 | 1.39 | 5 |
| ARPP19 | 5.63E-03 | 5 | 3 | 0.0008479 | Substantia_nigra | 3.28 | 3.74E-17 | 0.68 | -3.34 | 0.09 | -2.49 | 1.48 | 3 |
| CTD-2162K18.3 | 5.64E-03 | 5 | 2 | 0.001795 | Hypothalamus | 4.73 | 1.19E-52 | 0.27 | 3.02 | 3.12 | 3.10 | 0.05 | 2 |
| SDK1 | 5.65E-03 | 6 | 3 | 0.1131748 | Putamen_basal_ganglia | 4.18 | 1.19E-16 | 0.24 | -0.43 | 1.58 | 1.10 | 0.77 | 3 |
| SLC38A6 | 5.67E-03 | 13 | 3 | 0.001635 | Amygdala | 8.26 | 8.42E-18 | 0.94 | -3.15 | 3.15 | -0.67 | 2.57 | 3 |
| ENPP4 | 5.69E-03 | 13 | 2 | 0.0012077 | Substantia_nigra | 11.53 | 2.88E-17 | 1.27 | -3.24 | 3.09 | -2.15 | 2.18 | 2 |
| SLC12A8 | 5.71E-03 | 8 | 5 | 0.0007118 | Anterior_cingulate_cortex_BA24 | 2.46 | 2.78E-17 | 0.86 | -1.94 | 3.38 | 0.13 | 2.13 | 5 |
| STYX | 5.77E-03 | 13 | 2 | 0.001359 | Cerebellar_Hemisphere | 12.20 | 5.03E-19 | 0.67 | -3.20 | -2.68 | -3.11 | 0.15 | 2 |
| ELP5 | 5.80E-03 | 11 | 3 | 0.0004289 | Caudate_basal_ganglia | 9.19 | 2.63E-18 | 0.42 | -3.52 | 0.20 | -3.01 | 1.08 | 3 |
| LINC02175 | 5.81E-03 | 4 | 4 | 0.001864 | Frontal_Cortex_BA9 | 2.71 | 1.04E-01 | 0.10 | -3.11 | -2.38 | -2.78 | 0.32 | 4 |
| SP5 | 5.88E-03 | 11 | 2 | 0.007779 | Amygdala | 8.85 | 1.15E-17 | 2.12 | -2.66 | 2.21 | -0.97 | 1.67 | 2 |
| YWHAE | 5.89E-03 | 12 | 4 | 0.0059731 | Cerebellum | 6.03 | 2.73E-17 | 0.63 | -2.75 | 0.29 | -1.28 | 1.07 | 4 |
| ZFR | 5.89E-03 | 13 | 3 | 0.0742618 | Cerebellar_Hemisphere | 10.01 | 1.63E-17 | 0.55 | -1.78 | 1.60 | 0.76 | 1.35 | 3 |
| IRF2BPL | 5.90E-03 | 9 | 2 | 0.08967 | Cerebellar_Hemisphere | 7.48 | 1.28E-32 | 1.52 | -0.93 | 1.70 | 0.44 | 1.33 | 2 |
| PAMR1 | 5.90E-03 | 10 | 6 | 0.0001473 | Cerebellar_Hemisphere | 3.79 | 1.49E-17 | 0.22 | -3.80 | 0.02 | -1.11 | 1.42 | 6 |
| AC005753.1 | 5.93E-03 | 13 | 5 | 0.1558557 | Hypothalamus | 8.17 | 6.75E-17 | 0.32 | -0.98 | 1.42 | -0.46 | 0.66 | 5 |
| SIRPB1 | 5.94E-03 | 13 | 2 | 0.0016963 | Putamen_basal_ganglia | 7.66 | 2.19E-18 | 5.21 | -3.14 | -1.52 | -2.31 | 0.77 | 2 |
| RPRD1A | 5.94E-03 | 10 | 2 | 0.002672 | Nucleus_accumbens_basal_ganglia | 8.62 | 7.35E-18 | 1.19 | -2.05 | 3.00 | 2.35 | 1.56 | 2 |
| SPATA21 | 5.95E-03 | 3 | 3 | 0.0029377 | Cerebellum | 1.96 | 1.04E-01 | 0.10 | -1.17 | 2.97 | 1.29 | 2.17 | 3 |
| WDR4 | 5.99E-03 | 12 | 5 | 0.00762 | Amygdala | 5.88 | 1.48E-17 | 0.54 | -0.80 | 2.67 | 1.37 | 1.32 | 5 |
| RP11-265E18.1 | 6.06E-03 | 9 | 3 | 0.02578 | Nucleus_accumbens_basal_ganglia | 7.43 | 5.73E-17 | 0.68 | -2.23 | 1.77 | -1.17 | 1.13 | 3 |
| UNKL | 6.07E-03 | 2 | 2 | 0.001619 | Cerebellar_Hemisphere | 1.43 | 5.73E-01 | 0.57 | -3.15 | -0.87 | -2.01 | 1.61 | 2 |
| KIF5C | 6.07E-03 | 2 | 2 | 0.001639 | Cerebellar_Hemisphere | 1.92 | 8.32E-02 | 0.08 | -3.15 | 3.10 | -0.02 | 4.42 | 2 |
| MAPK11 | 6.12E-03 | 11 | 6 | 0.001179 | Cortex | 5.54 | 3.60E-17 | 0.22 | -3.24 | 3.24 | 0.42 | 2.11 | 6 |
| PAQR7 | 6.13E-03 | 5 | 1 | 0.006131 | Cerebellar_Hemisphere | 5.00 | 8.36E-84 | 5.00 | -2.74 | -2.74 | -2.74 | 0.00 | 1 |
| ARF6 | 6.15E-03 | 4 | 1 | 0.005093 | Caudate_basal_ganglia | 3.98 | 3.88E-17 | 3.98 | 2.66 | 2.80 | 2.73 | 0.08 | 1 |
| RP1-265C24.8 | 6.16E-03 | 4 | 2 | 0.001439 | Cerebellum | 2.29 | 3.33E-17 | 1.71 | 0.30 | 3.19 | 1.74 | 1.67 | 2 |
| MGAT4B | 6.18E-03 | 9 | 3 | 0.004396 | Hippocampus | 6.45 | 6.19E-33 | 0.87 | -2.85 | 2.11 | -1.61 | 2.00 | 3 |
| AKAP5 | 6.19E-03 | 2 | 2 | 0.3897 | Spinal_cord_cervical_c-1 | 1.93 | 6.76E-02 | 0.07 | -0.31 | 0.86 | 0.28 | 0.83 | 2 |

| | | | | | | | | | | | | | |
|---|---|---|---|---|---|---|---|---|---|---|---|---|---|
| UEVLD | 6.20E-03 | 4 | 2 | 0.005176 | Cerebellum | 3.33 | 8.44E-17 | 0.67 | 0.10 | 2.80 | 2.12 | 1.35 | 2 |
| ENOX1-AS1 | 6.23E-03 | 8 | 2 | 0.001722 | Caudate_basal_ganglia | 7.54 | 4.62E-17 | 0.25 | 2.41 | 3.13 | 3.02 | 0.25 | 2 |
| RP11-147L13.14 | 6.24E-03 | 13 | 2 | 0.00524 | Amygdala | 12.09 | 7.68E-17 | 0.61 | 1.36 | 2.79 | 1.90 | 0.54 | 2 |
| C1orf132 | 6.26E-03 | 9 | 2 | 0.0014461 | Caudate_basal_ganglia | 8.10 | 7.70E-17 | 0.90 | 0.92 | 3.19 | 1.17 | 0.76 | 2 |
| PPP1R35 | 6.28E-03 | 1 | 1 | 0.006276 | Spinal_cord_cervical_c-1 | 1.00 | 1.00E+00 | 1.00 | 2.73 | 2.73 | 2.73 | NA | 1 |
| NEK1 | 6.30E-03 | 13 | 2 | 0.001642 | Amygdala | 12.23 | 1.04E-17 | 0.67 | -3.15 | -1.34 | -1.68 | 0.48 | 2 |
| PSMB7 | 6.31E-03 | 12 | 1 | 0.004597 | Cortex | 11.61 | 1.90E-17 | 11.61 | -2.83 | -2.56 | -2.69 | 0.12 | 1 |
| SELENBP1 | 6.34E-03 | 5 | 3 | 0.0005343 | Hippocampus | 3.82 | 7.18E-18 | 0.19 | 2.67 | 3.46 | 2.96 | 0.35 | 3 |
| RP11-579E24.2 | 6.34E-03 | 1 | 1 | 0.00634 | Anterior_cingulate_cortex_BA24 | 1.00 | 1.00E+00 | 1.00 | -2.73 | -2.73 | -2.73 | NA | 1 |
| RP11-61L19.2 | 6.34E-03 | 12 | 4 | 0.002519 | Amygdala | 8.15 | 2.01E-17 | 0.66 | -0.64 | 3.02 | 0.15 | 1.22 | 4 |
| DOPEY2 | 6.34E-03 | 11 | 3 | 0.0004703 | Caudate_basal_ganglia | 8.07 | 4.26E-17 | 0.98 | 0.44 | 3.50 | 2.44 | 1.07 | 3 |
| TLE4 | 6.36E-03 | 7 | 2 | 0.001531 | Amygdala | 5.08 | 9.31E-33 | 1.92 | -3.17 | 3.17 | -1.36 | 2.50 | 2 |
| EPHA3 | 6.38E-03 | 8 | 3 | 0.0007368 | Cerebellum | 4.06 | 7.04E-33 | 0.94 | -3.38 | 1.55 | 0.33 | 1.73 | 3 |
| ASPSCR1 | 6.38E-03 | 7 | 1 | 0.006385 | Amygdala | 7.00 | 4.99E-50 | 7.00 | 2.73 | 2.73 | 2.73 | 0.00 | 1 |
| OS9 | 6.39E-03 | 1 | 1 | 0.006388 | Hippocampus | 1.00 | 1.00E+00 | 1.00 | -2.73 | -2.73 | -2.73 | NA | 1 |
| CCND1 | 6.40E-03 | 5 | 1 | 0.006399 | Amygdala | 5.00 | 8.51E-33 | 5.00 | -2.73 | -2.73 | -2.73 | 0.00 | 1 |
| RP11-676J15.1 | 6.41E-03 | 7 | 5 | 0.0002369 | Anterior_cingulate_cortex_BA24 | 3.53 | 3.09E-17 | 0.28 | 1.11 | 3.68 | 1.89 | 0.89 | 5 |
| LINC01606 | 6.45E-03 | 4 | 2 | 0.0106647 | Cerebellum | 2.34 | 9.73E-17 | 1.66 | -2.55 | -2.28 | -2.43 | 0.11 | 2 |
| BUB1B | 6.48E-03 | 10 | 3 | 0.001902 | Cerebellar_Hemisphere | 6.31 | 8.01E-19 | 1.67 | -3.11 | 0.12 | -1.65 | 0.92 | 3 |
| CTD-2201E18.5 | 6.48E-03 | 11 | 2 | 0.001886 | Amygdala | 9.08 | 3.89E-17 | 1.92 | -3.11 | -1.51 | -2.39 | 0.83 | 2 |
| RP1-288L1.5 | 6.55E-03 | 3 | 2 | 0.01004 | Anterior_cingulate_cortex_BA24 | 2.16 | 5.63E-32 | 0.84 | 2.56 | 2.57 | 2.56 | 0.01 | 2 |
| AOAH | 6.55E-03 | 10 | 3 | 0.005964 | Cerebellum | 7.67 | 1.94E-19 | 0.57 | -2.75 | 2.68 | 0.46 | 1.47 | 3 |
| PLAGL1 | 6.58E-03 | 5 | 4 | 0.0006534 | Cerebellar_Hemisphere | 2.56 | 5.13E-18 | 0.29 | -3.41 | 3.41 | -1.14 | 2.67 | 4 |
| GABPB2 | 6.63E-03 | 13 | 3 | 0.0086667 | Cerebellar_Hemisphere | 7.47 | 6.20E-19 | 1.14 | -1.75 | 2.62 | -0.45 | 1.17 | 3 |
| LINC01623 | 6.64E-03 | 1 | 1 | 0.006638 | Hypothalamus | 1.00 | 1.00E+00 | 1.00 | 2.71 | 2.71 | 2.71 | NA | 1 |
| PARK2 | 6.65E-03 | 12 | 3 | 0.002883 | Amygdala | 6.42 | 7.10E-18 | 1.50 | -2.98 | -0.92 | -2.33 | 0.78 | 3 |
| ZFP82 | 6.68E-03 | 5 | 3 | 0.001789 | Cerebellar_Hemisphere | 3.58 | 7.61E-18 | 0.48 | -3.12 | -0.33 | -0.94 | 1.22 | 3 |
| CHMP4B | 6.69E-03 | 9 | 3 | 0.00632 | Cerebellar_Hemisphere | 7.49 | 4.50E-17 | 0.68 | -2.73 | 0.25 | -0.74 | 0.80 | 3 |
| RP11-367F23.1 | 6.73E-03 | 4 | 2 | 0.0015275 | Frontal_Cortex_BA9 | 3.81 | 1.45E-16 | 0.19 | -3.17 | -3.04 | -3.08 | 0.06 | 2 |
| ZNF236 | 6.75E-03 | 13 | 4 | 0.001839 | Caudate_basal_ganglia | 9.06 | 3.47E-17 | 0.54 | 1.02 | 3.12 | 1.76 | 0.82 | 4 |
| ACTR1A | 6.75E-03 | 12 | 3 | 0.0003722 | Amygdala | 7.86 | 2.29E-18 | 0.87 | -0.37 | 3.56 | 2.27 | 1.63 | 3 |
| PTCH1 | 6.78E-03 | 4 | 2 | 0.005004 | Frontal_Cortex_BA9 | 2.34 | 2.41E-17 | 1.66 | -2.81 | 2.81 | 0.48 | 2.36 | 2 |
| HSPG2 | 6.78E-03 | 7 | 3 | 0.001471 | Cerebellum | 4.05 | 6.84E-18 | 0.73 | -3.18 | -0.25 | -0.96 | 1.01 | 3 |
| PAK6 | 6.78E-03 | 3 | 3 | 0.0008286 | Cerebellum | 1.87 | 1.31E-01 | 0.13 | -3.34 | -0.98 | -2.49 | 1.31 | 3 |
| DRAM2 | 6.78E-03 | 13 | 1 | 0.006483 | Anterior_cingulate_cortex_BA24 | 12.96 | 5.35E-18 | 12.96 | 2.67 | 2.72 | 2.70 | 0.02 | 1 |
| PSD | 6.82E-03 | 12 | 3 | 0.0013691 | Cerebellum | 10.08 | 1.55E-17 | 0.49 | -0.53 | 3.20 | 0.69 | 0.85 | 3 |
| DDX31 | 6.85E-03 | 13 | 3 | 0.0005354 | Cerebellar_Hemisphere | 8.73 | 1.16E-17 | 0.77 | -3.46 | 2.30 | -0.39 | 1.68 | 3 |

| | | | | | | | | | | | | | |
|---|---|---|---|---|---|---|---|---|---|---|---|---|---|
| CYB5R4 | 6.86E-03 | 13 | 1 | 0.006863 | Amygdala | 13.00 | 1.84E-33 | 13.00 | 2.70 | 2.70 | 2.70 | 0.00 | 1 |
| RNF34 | 6.88E-03 | 7 | 2 | 0.005643 | Caudate_basal_ganglia | 6.48 | 1.11E-17 | 0.52 | -2.77 | -1.37 | -2.37 | 0.68 | 2 |
| MCPH1 | 6.91E-03 | 8 | 3 | 0.02441 | Nucleus_accumbens_basal_ganglia | 6.41 | 6.56E-18 | 0.41 | -1.41 | 2.25 | 1.05 | 1.24 | 3 |
| JARID2 | 6.92E-03 | 7 | 3 | 0.003272 | Caudate_basal_ganglia | 4.85 | 1.59E-17 | 0.94 | -2.94 | 0.30 | -0.98 | 1.09 | 3 |
| RNF222 | 6.96E-03 | 1 | 1 | 0.006956 | Cerebellar_Hemisphere | 1.00 | 1.00E+00 | 1.00 | -2.70 | -2.70 | -2.70 | NA | 1 |
| RBM23 | 6.96E-03 | 13 | 2 | 0.001585 | Spinal_cord_cervical_c-1 | 11.54 | 7.74E-17 | 1.14 | -3.16 | -0.90 | -1.69 | 0.74 | 2 |
| HDAC11 | 7.02E-03 | 13 | 6 | 0.0132159 | Hippocampus | 7.12 | 8.30E-18 | 0.24 | -0.24 | 2.48 | 0.99 | 0.81 | 6 |
| TICRR | 7.03E-03 | 12 | 3 | 0.0013661 | Spinal_cord_cervical_c-1 | 9.73 | 1.55E-17 | 0.51 | 1.86 | 3.20 | 2.40 | 0.42 | 3 |
| WARS2 | 7.05E-03 | 13 | 2 | 0.0152096 | Putamen_basal_ganglia | 11.89 | 3.55E-17 | 0.77 | -2.43 | 0.40 | -1.10 | 0.77 | 2 |
| KLHL20 | 7.06E-03 | 11 | 2 | 0.0133846 | Nucleus_accumbens_basal_ganglia | 10.64 | 2.26E-67 | 0.36 | -2.47 | 0.73 | 0.04 | 1.07 | 2 |
| NF2 | 7.09E-03 | 13 | 4 | 0.000515 | Cerebellum | 7.84 | 1.51E-17 | 1.04 | 1.57 | 3.47 | 2.30 | 0.64 | 4 |
| RP11-94C24.13 | 7.10E-03 | 6 | 2 | 0.002446 | Nucleus_accumbens_basal_ganglia | 4.95 | 8.43E-17 | 0.90 | -3.03 | -0.50 | -2.34 | 0.96 | 2 |
| RAB40C | 7.13E-03 | 13 | 3 | 0.0047256 | Cerebellar_Hemisphere | 8.26 | 4.98E-17 | 1.32 | -2.83 | 0.41 | -1.34 | 1.03 | 3 |
| CIART | 7.13E-03 | 1 | 1 | 0.007135 | Anterior_cingulate_cortex_BA24 | 1.00 | 1.00E+00 | 1.00 | -2.69 | -2.69 | -2.69 | NA | 1 |
| GLDC | 7.15E-03 | 12 | 4 | 0.0004761 | Cerebellum | 7.35 | 1.21E-17 | 0.99 | -3.49 | 0.41 | -0.44 | 1.10 | 4 |
| MRPL35 | 7.18E-03 | 13 | 3 | 0.0009378 | Substantia_nigra | 9.51 | 6.08E-17 | 1.43 | 1.09 | 3.31 | 2.09 | 0.71 | 3 |
| EEF1B2 | 7.22E-03 | 5 | 1 | 0.00722 | Caudate_basal_ganglia | 5.00 | 1.25E-32 | 5.00 | -2.69 | -2.69 | -2.69 | 0.00 | 1 |
| NR1H3 | 7.22E-03 | 13 | 3 | 0.0069369 | Spinal_cord_cervical_c-1 | 8.99 | 1.28E-17 | 0.55 | 1.47 | 2.70 | 2.20 | 0.44 | 3 |
| RP11-700H6.1 | 7.23E-03 | 4 | 2 | 0.008551 | Nucleus_accumbens_basal_ganglia | 3.55 | 5.92E-03 | 0.43 | 0.46 | 2.63 | 1.88 | 1.02 | 2 |
| RP11-480A16.1 | 7.31E-03 | 9 | 6 | 0.0241639 | Nucleus_accumbens_basal_ganglia | 3.87 | 1.23E-16 | 0.29 | -1.65 | 2.25 | -0.13 | 1.31 | 6 |
| DHRS11 | 7.33E-03 | 10 | 1 | 0.007299 | Anterior_cingulate_cortex_BA24 | 9.78 | 5.41E-18 | 9.78 | 2.56 | 2.68 | 2.65 | 0.05 | 1 |
| CTBP2 | 7.34E-03 | 9 | 3 | 0.000653 | Cerebellum | 5.01 | 1.46E-17 | 0.99 | -3.41 | 0.78 | -0.42 | 1.29 | 3 |
| HTR1E | 7.38E-03 | 11 | 3 | 0.01326 | Caudate_basal_ganglia | 6.20 | 3.68E-18 | 0.46 | -2.26 | 2.48 | 1.06 | 1.49 | 3 |
| TPK1 | 7.38E-03 | 13 | 3 | 0.001006 | Putamen_basal_ganglia | 10.22 | 2.93E-18 | 1.06 | -3.29 | -1.72 | -2.48 | 0.48 | 3 |
| PPP1R14B | 7.40E-03 | 11 | 2 | 0.0106554 | Amygdala | 10.32 | 3.00E-18 | 0.60 | -2.55 | -0.07 | -0.33 | 0.74 | 2 |
| RP11-44N21.1 | 7.41E-03 | 13 | 4 | 0.0002435 | Hypothalamus | 8.42 | 5.15E-18 | 0.46 | -3.67 | 0.01 | -1.42 | 1.15 | 4 |
| CLIC5 | 7.42E-03 | 8 | 3 | 0.0008515 | Amygdala | 4.67 | 1.87E-18 | 1.40 | -3.34 | -0.58 | -1.41 | 1.16 | 3 |
| LINC00251 | 7.45E-03 | 1 | 1 | 0.007447 | Amygdala | 1.00 | 1.00E+00 | 1.00 | 2.68 | 2.68 | 2.68 | NA | 1 |
| TRIM66 | 7.47E-03 | 13 | 3 | 0.005085 | Cerebellar_Hemisphere | 10.23 | 1.14E-17 | 0.83 | -2.80 | 0.97 | -1.10 | 1.08 | 3 |
| CDKN1A | 7.48E-03 | 5 | 2 | 0.0019054 | Amygdala | 2.91 | 7.01E-34 | 2.09 | -3.10 | 0.34 | -1.83 | 1.69 | 2 |
| LINC01694 | 7.49E-03 | 6 | 3 | 0.01467 | Putamen_basal_ganglia | 4.73 | 4.85E-16 | 0.17 | -0.75 | 2.44 | 0.48 | 1.15 | 3 |
| ABCC9 | 7.54E-03 | 12 | 5 | 0.0412749 | Anterior_cingulate_cortex_BA24 | 6.59 | 5.76E-17 | 0.22 | -1.73 | 2.04 | 0.27 | 1.22 | 5 |
| SPATA3 | 7.59E-03 | 7 | 2 | 0.00158 | Substantia_nigra | 5.93 | 2.55E-17 | 0.98 | -3.16 | -1.04 | -2.75 | 0.76 | 2 |
| TMOD4 | 7.59E-03 | 2 | 2 | 0.004471 | Cortex | 1.13 | 8.66E-01 | 0.87 | -2.84 | -1.66 | -2.25 | 0.83 | 2 |
| FOXH1 | 7.61E-03 | 7 | 1 | 0.007607 | Hippocampus | 7.00 | 4.72E-32 | 7.00 | 2.67 | 2.67 | 2.67 | 0.00 | 1 |
| BICDL1 | 7.61E-03 | 10 | 2 | 0.005096 | Hypothalamus | 8.52 | 1.03E-33 | 1.47 | -2.80 | 1.14 | -2.15 | 1.27 | 2 |
| GOLGA1 | 7.62E-03 | 5 | 2 | 0.02728 | Cerebellum | 4.26 | 9.31E-18 | 0.66 | -0.77 | 2.21 | 1.56 | 1.30 | 2 |

| | | | | | | | | | | | | | |
|---|---|---|---|---|---|---|---|---|---|---|---|---|---|
| CMTM3 | 7.65E-03 | 7 | 2 | 0.004544 | Spinal_cord_cervical_c-1 | 5.56 | 7.20E-33 | 1.44 | -2.84 | -2.70 | -2.76 | 0.07 | 2 |
| SMCP | 7.67E-03 | 3 | 1 | 0.007356 | Cerebellum | 3.00 | 6.09E-17 | 3.00 | 2.64 | 2.68 | 2.67 | 0.02 | 1 |
| LZTS1 | 7.70E-03 | 8 | 3 | 0.00224 | Amygdala | 5.01 | 1.14E-18 | 0.98 | -3.06 | 1.23 | -1.47 | 2.19 | 3 |
| RP11-274B21.9 | 7.77E-03 | 13 | 3 | 0.0038625 | Cerebellum | 11.01 | 9.27E-17 | 0.37 | 1.33 | 2.89 | 2.01 | 0.55 | 3 |
| ADRB2 | 7.78E-03 | 9 | 2 | 0.004859 | Hypothalamus | 8.02 | 6.51E-18 | 0.77 | -2.82 | -0.02 | -2.38 | 0.91 | 2 |
| NIPAL1 | 7.82E-03 | 5 | 3 | 0.01168 | Caudate_basal_ganglia | 3.22 | 4.01E-17 | 0.64 | -0.79 | 2.52 | 1.02 | 1.69 | 3 |
| ANGEL2 | 7.85E-03 | 11 | 2 | 0.006154 | Putamen_basal_ganglia | 10.27 | 2.22E-18 | 0.54 | 1.30 | 2.74 | 1.96 | 0.58 | 2 |
| ACADVL | 7.85E-03 | 7 | 3 | 0.0006928 | Nucleus_accumbens_basal_ganglia | 5.26 | 3.64E-17 | 0.27 | -3.39 | 0.75 | -2.45 | 1.51 | 3 |
| KIAA0141 | 7.91E-03 | 11 | 4 | 0.0319068 | Hypothalamus | 8.22 | 1.29E-18 | 0.34 | -0.88 | 2.15 | 0.02 | 0.83 | 4 |
| SMCR8 | 7.92E-03 | 10 | 2 | 0.0057903 | Hypothalamus | 5.06 | 8.13E-19 | 4.90 | 0.71 | 2.76 | 2.05 | 0.79 | 2 |
| RAB3IL1 | 7.94E-03 | 8 | 2 | 0.0017175 | Putamen_basal_ganglia | 7.35 | 1.12E-18 | 0.59 | 2.02 | 3.14 | 2.95 | 0.38 | 2 |
| VPS33B | 7.97E-03 | 11 | 5 | 0.0005205 | Anterior_cingulate_cortex_BA24 | 5.49 | 9.32E-18 | 0.39 | -1.48 | 3.47 | 1.52 | 1.48 | 5 |
| LRP1B | 7.98E-03 | 13 | 4 | 0.0004129 | Cerebellar_Hemisphere | 9.81 | 3.96E-19 | 0.62 | 0.63 | 3.53 | 1.69 | 0.69 | 4 |
| FAM131C | 8.00E-03 | 11 | 4 | 0.0003133 | Caudate_basal_ganglia | 7.12 | 2.47E-17 | 0.37 | -2.31 | 3.60 | 1.48 | 1.64 | 4 |
| FOXF1 | 8.03E-03 | 1 | 1 | 0.008029 | Frontal_Cortex_BA9 | 1.00 | 1.00E+00 | 1.00 | -2.65 | -2.65 | -2.65 | NA | 1 |
| ANAPC4 | 8.08E-03 | 13 | 3 | 0.0012732 | Frontal_Cortex_BA9 | 10.55 | 3.97E-17 | 0.40 | 0.06 | 3.22 | 2.42 | 0.90 | 3 |
| AK2 | 8.12E-03 | 7 | 1 | 0.00109 | Frontal_Cortex_BA9 | 6.81 | 2.04E-17 | 6.81 | 2.51 | 3.27 | 2.62 | 0.29 | 1 |
| KATNAL2 | 8.13E-03 | 13 | 4 | 0.0008794 | Hypothalamus | 7.14 | 1.21E-17 | 0.53 | -3.33 | 3.06 | 0.47 | 1.77 | 4 |
| EEF1E1 | 8.13E-03 | 8 | 3 | 0.0012786 | Spinal_cord_cervical_c-1 | 5.06 | 3.11E-17 | 0.48 | 0.75 | 3.22 | 2.27 | 0.83 | 3 |
| SLC26A10 | 8.14E-03 | 11 | 6 | 0.0202856 | Cortex | 4.15 | 9.61E-18 | 0.26 | -2.16 | 2.32 | 0.03 | 1.69 | 6 |
| EPHA10 | 8.16E-03 | 7 | 4 | 0.0012121 | Frontal_Cortex_BA9 | 4.41 | 7.41E-20 | 0.28 | -3.24 | 1.72 | -0.89 | 1.71 | 4 |
| PRMT6 | 8.19E-03 | 13 | 1 | 0.0043867 | Cortex | 12.84 | 2.31E-17 | 12.84 | -2.85 | -2.51 | -2.63 | 0.08 | 1 |
| SYN2 | 8.20E-03 | 12 | 7 | 6.24E-05 | Putamen_basal_ganglia | 5.20 | 1.86E-17 | 0.32 | -3.83 | 4.00 | -0.02 | 2.56 | 7 |
| CMYA5 | 8.23E-03 | 3 | 2 | 0.002097 | Anterior_cingulate_cortex_BA24 | 2.00 | 1.56E-32 | 1.00 | -3.08 | 0.41 | -0.75 | 2.01 | 2 |
| RP11-61G23.2 | 8.26E-03 | 1 | 1 | 0.008256 | Frontal_Cortex_BA9 | 1.00 | 1.00E+00 | 1.00 | -2.64 | -2.64 | -2.64 | NA | 1 |
| RGS13 | 8.30E-03 | 1 | 1 | 0.008304 | Caudate_basal_ganglia | 1.00 | 1.00E+00 | 1.00 | 2.64 | 2.64 | 2.64 | NA | 1 |
| AKT3 | 8.34E-03 | 6 | 1 | 0.008337 | Anterior_cingulate_cortex_BA24 | 6.00 | 3.91E-48 | 6.00 | 2.64 | 2.64 | 2.64 | 0.00 | 1 |
| TOR1AIP1 | 8.34E-03 | 3 | 2 | 0.05971 | Cerebellar_Hemisphere | 2.33 | 1.64E-16 | 0.67 | -1.88 | 1.88 | -0.43 | 2.03 | 2 |
| LZTR1 | 8.35E-03 | 10 | 6 | 0.0078171 | Hippocampus | 5.37 | 2.37E-17 | 0.25 | -2.66 | 1.19 | -1.38 | 1.17 | 6 |
| CFAP157 | 8.36E-03 | 3 | 3 | 0.1358 | Cerebellum | 1.99 | 1.73E-01 | 0.17 | -1.49 | 0.93 | -0.04 | 1.28 | 3 |
| NTM | 8.38E-03 | 2 | 2 | 0.003473 | Spinal_cord_cervical_c-1 | 1.76 | 2.43E-01 | 0.24 | -2.92 | -2.87 | -2.90 | 0.03 | 2 |
| RP11-44N11.2 | 8.42E-03 | 10 | 2 | 0.006798 | Hippocampus | 8.19 | 3.11E-17 | 1.76 | -2.71 | 2.20 | 1.23 | 2.05 | 2 |
| ALG1L | 8.52E-03 | 12 | 4 | 0.01877 | Frontal_Cortex_BA9 | 7.04 | 6.10E-18 | 0.58 | -1.78 | 2.35 | 0.76 | 1.69 | 4 |
| SPI1 | 8.54E-03 | 12 | 2 | 0.0014391 | Cerebellar_Hemisphere | 9.61 | 3.24E-18 | 2.21 | -3.07 | 3.19 | -0.76 | 1.55 | 2 |
| MPPED1 | 8.59E-03 | 10 | 6 | 0.0012492 | Substantia_nigra | 4.19 | 7.92E-18 | 0.44 | -1.58 | 3.23 | 1.24 | 1.64 | 6 |
| RP11-321M21.4 | 8.62E-03 | 2 | 1 | 0.008621 | Cortex | 2.00 | 6.12E-17 | 2.00 | 2.63 | 2.63 | 2.63 | 0.00 | 1 |
| LMO4 | 8.63E-03 | 7 | 2 | 0.005713 | Cerebellar_Hemisphere | 6.00 | 5.72E-38 | 1.00 | 1.19 | 2.76 | 1.41 | 0.60 | 2 |

| | | | | | | | | | | | | | |
|---|---|---|---|---|---|---|---|---|---|---|---|---|---|
| SLC6A5 | 8.65E-03 | 3 | 1 | 0.008647 | Cerebellum | 3.00 | 5.35E-35 | 3.00 | -2.63 | -2.63 | -2.63 | 0.00 | 1 |
| GRIK5 | 8.65E-03 | 3 | 2 | 0.002538 | Caudate_basal_ganglia | 2.68 | 4.63E-02 | 0.27 | 2.68 | 3.02 | 2.90 | 0.20 | 2 |
| LINC00320 | 8.66E-03 | 8 | 3 | 0.0008652 | Cortex | 6.91 | 1.08E-16 | 0.38 | 2.63 | 3.33 | 3.02 | 0.28 | 3 |
| BNIP3 | 8.71E-03 | 11 | 2 | 0.005278 | Cortex | 8.92 | 4.34E-18 | 2.08 | -2.79 | 0.76 | -0.03 | 1.44 | 2 |
| GCNT4 | 8.85E-03 | 12 | 2 | 0.06811 | Cerebellum | 10.76 | 1.29E-17 | 1.07 | -0.92 | 1.82 | 1.31 | 0.99 | 2 |
| MADD | 8.92E-03 | 13 | 2 | 0.0017619 | Cerebellum | 11.70 | 2.54E-17 | 1.15 | -3.13 | -0.80 | -1.93 | 0.61 | 2 |
| HSPA1B | 9.06E-03 | 11 | 2 | 0.01637 | Hippocampus | 8.01 | 2.19E-17 | 2.99 | -2.40 | 2.15 | 0.82 | 2.07 | 2 |
| VIPR1 | 9.06E-03 | 4 | 2 | 0.004706 | Amygdala | 3.40 | 5.30E-17 | 0.50 | -2.83 | -0.82 | -2.18 | 0.95 | 2 |
| FURIN | 9.07E-03 | 12 | 4 | 0.001752 | Anterior_cingulate_cortex_BA24 | 8.38 | 8.23E-17 | 0.51 | 0.97 | 3.13 | 2.81 | 0.65 | 4 |
| BTBD3 | 9.08E-03 | 12 | 4 | 0.001244 | Cortex | 6.27 | 4.74E-19 | 0.77 | -3.23 | -1.46 | -2.24 | 0.68 | 4 |
| CMTM1 | 9.10E-03 | 13 | 2 | 0.0029451 | Hippocampus | 12.33 | 1.05E-17 | 0.56 | -2.97 | -2.40 | -2.69 | 0.14 | 2 |
| CCDC192 | 9.12E-03 | 3 | 1 | 0.009124 | Caudate_basal_ganglia | 3.00 | 5.35E-35 | 3.00 | -2.61 | 2.61 | -0.87 | 3.01 | 1 |
| MFSD1 | 9.13E-03 | 5 | 3 | 0.0343561 | Cortex | 3.14 | 1.13E-17 | 0.72 | -2.12 | 1.96 | -0.23 | 1.45 | 3 |
| WDR46 | 9.19E-03 | 12 | 3 | 0.01334 | Amygdala | 8.06 | 4.90E-18 | 1.02 | -2.47 | 0.96 | -0.70 | 1.40 | 3 |
| MYL6 | 9.20E-03 | 3 | 1 | 0.009201 | Frontal_Cortex_BA9 | 3.00 | 4.80E-17 | 3.00 | -2.60 | -2.60 | -2.60 | 0.00 | 1 |
| MRPS21 | 9.20E-03 | 13 | 2 | 0.002228 | Cerebellar_Hemisphere | 12.15 | 5.64E-17 | 0.69 | 2.22 | 3.06 | 2.80 | 0.28 | 2 |
| ALDH1A1 | 9.23E-03 | 5 | 1 | 0.009226 | Amygdala | 5.00 | 8.76E-33 | 5.00 | -2.60 | -2.60 | -2.60 | 0.00 | 1 |
| PPP1R16A | 9.23E-03 | 10 | 1 | 0.008347 | Substantia_nigra | 9.98 | 5.41E-18 | 9.98 | -2.64 | 2.64 | -0.52 | 2.69 | 1 |
| DDAH1 | 9.23E-03 | 2 | 2 | 0.002519 | Cerebellar_Hemisphere | 1.46 | 5.36E-01 | 0.54 | -3.02 | -1.84 | -2.43 | 0.84 | 2 |
| AKAP13 | 9.24E-03 | 11 | 3 | 0.01051 | Cerebellar_Hemisphere | 8.00 | 2.98E-18 | 1.08 | -2.56 | 2.56 | 0.87 | 1.26 | 3 |
| SPOCK1 | 9.26E-03 | 5 | 2 | 0.006973 | Amygdala | 3.61 | 3.71E-17 | 1.35 | -2.70 | 0.58 | -1.84 | 1.38 | 2 |
| ZNF469 | 9.32E-03 | 2 | 1 | 0.009322 | Caudate_basal_ganglia | 2.00 | 3.35E-17 | 2.00 | -2.60 | -2.60 | -2.60 | 0.00 | 1 |
| SOCS5 | 9.33E-03 | 6 | 2 | 0.005986 | Cerebellar_Hemisphere | 4.85 | 4.51E-19 | 1.15 | 0.00 | 2.75 | 1.60 | 1.28 | 2 |
| LA16c-385E7.1 | 9.33E-03 | 10 | 5 | 0.0245707 | Spinal_cord_cervical_c-1 | 3.41 | 8.72E-17 | 0.81 | -2.25 | 2.15 | -0.46 | 1.76 | 5 |
| PFDN6 | 9.34E-03 | 1 | 1 | 0.009339 | Cerebellar_Hemisphere | 1.00 | 1.00E+00 | 1.00 | -2.60 | -2.60 | -2.60 | NA | 1 |
| RP1-288L1.4 | 9.36E-03 | 1 | 1 | 0.009355 | Anterior_cingulate_cortex_BA24 | 1.00 | 1.00E+00 | 1.00 | 2.60 | 2.60 | 2.60 | NA | 1 |
| C4orf47 | 9.37E-03 | 12 | 6 | 0.00204 | Amygdala | 5.70 | 2.84E-17 | 0.32 | -3.08 | 2.12 | 0.33 | 1.50 | 6 |
| NT5E | 9.38E-03 | 12 | 4 | 0.01953 | Substantia_nigra | 9.48 | 3.93E-18 | 0.41 | -1.97 | 2.34 | 0.28 | 0.97 | 4 |
| EPC1 | 9.38E-03 | 8 | 1 | 0.009382 | Cerebellum | 8.00 | 1.65E-33 | 8.00 | 2.60 | 2.60 | 2.60 | 0.00 | 1 |
| HYPK | 9.41E-03 | 5 | 2 | 0.002416 | Putamen_basal_ganglia | 3.16 | 5.86E-49 | 1.84 | -3.03 | -0.18 | -1.89 | 1.56 | 2 |
| RP11-333I13.1 | 9.43E-03 | 9 | 5 | 0.001688 | Cerebellar_Hemisphere | 5.99 | 1.71E-18 | 0.25 | -3.14 | 3.03 | 0.98 | 2.35 | 5 |
| SOX6 | 9.44E-03 | 13 | 2 | 0.005326 | Cerebellum | 11.18 | 2.08E-18 | 1.67 | -2.79 | 1.37 | -0.51 | 1.15 | 2 |
| DKFZp779M0652 | 9.49E-03 | 13 | 3 | 0.0017074 | Substantia_nigra | 10.70 | 3.43E-17 | 0.56 | -3.14 | -2.17 | -2.63 | 0.31 | 3 |
| ZSCAN16 | 9.50E-03 | 11 | 3 | 0.0014012 | Substantia_nigra | 7.95 | 1.92E-17 | 0.65 | -3.19 | 3.02 | -1.29 | 2.50 | 3 |
| COMMD10 | 9.55E-03 | 13 | 4 | 0.0010317 | Cerebellum | 8.95 | 3.11E-17 | 0.71 | -3.28 | -0.38 | -1.48 | 0.90 | 4 |
| SLFN12 | 9.58E-03 | 12 | 2 | 0.01728 | Caudate_basal_ganglia | 10.77 | 1.17E-36 | 0.96 | 1.97 | 2.38 | 2.31 | 0.15 | 2 |
| ACTN2 | 9.59E-03 | 13 | 5 | 0.0101 | Spinal_cord_cervical_c-1 | 7.06 | 2.20E-17 | 0.28 | 0.20 | 2.57 | 0.92 | 0.74 | 5 |

| | | | | | | | | | | | | | |
|---|---|---|---|---|---|---|---|---|---|---|---|---|---|
| RP11-65M17.3 | 9.59E-03 | 12 | 3 | 0.003228 | Caudate_basal_ganglia | 8.79 | 3.81E-17 | 0.33 | -2.95 | -2.41 | -2.76 | 0.16 | 3 |
| CEBPZ | 9.60E-03 | 12 | 2 | 0.0008525 | Cerebellar_Hemisphere | 10.78 | 3.71E-17 | 0.59 | -3.34 | -1.81 | -2.87 | 0.37 | 2 |
| DUT | 9.64E-03 | 3 | 1 | 0.009638 | Amygdala | 3.00 | 6.09E-17 | 3.00 | -2.59 | 2.59 | 0.86 | 2.99 | 1 |
| CTD-2287O16.4 | 9.64E-03 | 10 | 3 | 0.001438 | Anterior_cingulate_cortex_BA24 | 6.67 | 1.27E-18 | 0.47 | -1.10 | 3.19 | 0.47 | 1.61 | 3 |
| EIF3M | 9.70E-03 | 5 | 2 | 0.003877 | Cerebellar_Hemisphere | 4.44 | 3.95E-17 | 0.43 | 2.07 | 2.89 | 2.39 | 0.44 | 2 |
| CDCA2 | 9.78E-03 | 1 | 1 | 0.009778 | Substantia_nigra | 1.00 | 1.00E+00 | 1.00 | 2.58 | 2.58 | 2.58 | NA | 1 |
| BAG6 | 9.84E-03 | 13 | 2 | 0.0005125 | Anterior_cingulate_cortex_BA24 | 11.42 | 1.81E-17 | 0.84 | -3.47 | -2.16 | -2.76 | 0.42 | 2 |
| RBM17 | 9.84E-03 | 13 | 6 | 0.0148272 | Frontal_Cortex_BA9 | 6.64 | 1.45E-16 | 0.25 | -2.44 | 1.24 | -0.83 | 1.01 | 6 |
| LINC00533 | 9.88E-03 | 2 | 2 | 0.2129 | Cerebellum | 1.86 | 1.35E-01 | 0.14 | -1.25 | 0.31 | -0.47 | 1.10 | 2 |
| CSNK2A2 | 9.89E-03 | 8 | 2 | 0.003795 | Cerebellum | 7.26 | 1.04E-19 | 0.74 | -2.21 | 2.89 | -1.57 | 1.80 | 2 |
| PIP4K2C | 9.91E-03 | 11 | 3 | 0.004079 | Cerebellum | 5.68 | 3.36E-18 | 1.53 | -2.87 | 0.48 | -0.92 | 1.40 | 3 |
| TRPT1 | 9.92E-03 | 13 | 1 | 0.0021773 | Cortex | 12.33 | 9.32E-18 | 12.33 | -3.06 | -2.17 | -2.51 | 0.22 | 1 |
| MAPK7 | 9.98E-03 | 2 | 1 | 0.009975 | Frontal_Cortex_BA9 | 2.00 | 3.53E-16 | 2.00 | -2.58 | -2.58 | -2.58 | 0.00 | 1 |
| WWTR1 | 9.98E-03 | 1 | 1 | 0.009982 | Caudate_basal_ganglia | 1.00 | 1.00E+00 | 1.00 | 2.58 | 2.58 | 2.58 | NA | 1 |

Pvalue, significance p-value of S-MultiXcan association; n, number of "tissues" available for this gene; n_indep, number of independent components of variation kept among the tissues' predictions. (Synthetic independent tissues); p_i_best, best p-value of single-tissue S-PrediXcan association; t_i_best, name of best single-tissue S-PrediXcan association; eigen_max, In the SVD decomposition of predicted expression correlation: eigenvalue (variance explained) of the top independent component; eigen_min, In the SVD decomposition of predicted expression correlation: eigenvalue (variance explained) of the last independent component; eigen_min_kept, In the SVD decomposition of predicted expression correlation: eigenvalue (variance explained) of the smalles independent component that was kept; z_min, minimum z-score among single-tissue S-Predican associations; z_max, maximum z-score among single-tissue S-Predican associations; z_mean, mean z-score among single-tissue S-Predican associations; z_sd, standard deviation of the mean z-score among single-tissue S-Predican associations; tmi, trace of T * T', where Tis correlation of predicted expression levels for different tissues multiplied by its SVD pseudo-inverse.

**Table S14**. TWAS-identified genes associated with anorexia nervosa in adipose tissues.

| gene_name | pvalue | n | n_indep | p_i_best | t_i_best | eigen_max | eigen_min | eigen_min_kept | z_min | z_max | z_mean | z_sd | tmi |
|---|---|---|---|---|---|---|---|---|---|---|---|---|---|
| CELSR3 | 5.76E-14 | 2 | 2 | 9.25E-14 | Adipose_Visceral_Omentum | 1.42 | 0.58 | 0.58 | 5.27 | 7.45 | 6.36 | 1.54 | 2 |
| KLHDC8B | 1.35E-12 | 2 | 2 | 1.50E-05 | Adipose_Visceral_Omentum | 1.81 | 0.19 | 0.19 | -4.33 | 0.03 | -2.15 | 3.08 | 2 |
| C3orf62 | 7.20E-12 | 1 | 1 | 7.20E-12 | Adipose_Visceral_Omentum | 1.00 | 1.00 | 1.00 | 6.85 | 6.85 | 6.85 | NA | 1 |
| WDR6 | 8.76E-11 | 2 | 1 | 5.55E-11 | Adipose_Subcutaneous | 2.00 | 0.00 | 2.00 | -6.56 | -6.41 | -6.48 | 0.10 | 1 |
| SLC26A6 | 3.00E-10 | 1 | 1 | 3.00E-10 | Adipose_Subcutaneous | 1.00 | 1.00 | 1.00 | -6.30 | -6.30 | -6.30 | NA | 1 |
| NCKIPSD | 1.05E-07 | 2 | 1 | 6.20E-08 | Adipose_Visceral_Omentum | 1.98 | 0.02 | 1.98 | 5.16 | 5.41 | 5.29 | 0.18 | 1 |
| USP4 | 4.20E-07 | 2 | 2 | 1.37E-06 | Adipose_Visceral_Omentum | 1.45 | 0.55 | 0.55 | -4.83 | -4.37 | -4.60 | 0.33 | 2 |
| CCDC71 | 4.75E-07 | 1 | 1 | 4.75E-07 | Adipose_Visceral_Omentum | 1.00 | 1.00 | 1.00 | -5.04 | -5.04 | -5.04 | NA | 1 |
| CCDC36 | 5.43E-07 | 2 | 1 | 4.91E-07 | Adipose_Visceral_Omentum | 2.00 | 0.00 | 2.00 | 4.98 | 5.03 | 5.01 | 0.03 | 1 |
| RP11-3B7.1 | 6.21E-07 | 2 | 1 | 6.21E-07 | Adipose_Subcutaneous | 2.00 | 0.00 | 2.00 | 4.98 | 4.98 | 4.98 | 0.00 | 1 |
| P4HTM | 1.06E-06 | 2 | 1 | 5.27E-07 | Adipose_Subcutaneous | 1.96 | 0.04 | 1.96 | 4.66 | 5.02 | 4.84 | 0.25 | 1 |
| ARIH2 | 1.19E-06 | 2 | 1 | 8.38E-07 | Adipose_Subcutaneous | 1.99 | 0.01 | 1.99 | 4.77 | 4.93 | 4.85 | 0.11 | 1 |
| DAG1 | 1.23E-06 | 2 | 2 | 1.35E-06 | Adipose_Subcutaneous | 1.59 | 0.41 | 0.41 | -4.83 | -4.43 | -4.63 | 0.28 | 2 |
| ARIH2OS | 1.88E-06 | 1 | 1 | 1.88E-06 | Adipose_Subcutaneous | 1.00 | 1.00 | 1.00 | -4.77 | -4.77 | -4.77 | NA | 1 |
| APEH | 2.55E-06 | 2 | 2 | 4.45E-07 | Adipose_Visceral_Omentum | 1.93 | 0.07 | 0.07 | -5.05 | -4.89 | -4.97 | 0.11 | 2 |
| DALRD3 | 2.55E-06 | 2 | 1 | 2.46E-06 | Adipose_Subcutaneous | 1.99 | 0.01 | 1.99 | 4.68 | 4.71 | 4.70 | 0.02 | 1 |
| FAM212A | 5.44E-06 | 2 | 2 | 3.83E-05 | Adipose_Visceral_Omentum | 1.13 | 0.87 | 0.87 | -4.12 | 2.16 | -0.98 | 4.44 | 2 |
| RHOA | 1.15E-05 | 2 | 1 | 1.15E-05 | Adipose_Subcutaneous | 2.00 | 0.00 | 2.00 | 4.39 | 4.39 | 4.39 | 0.00 | 1 |
| SUOX | 1.50E-05 | 2 | 1 | 1.41E-05 | Adipose_Visceral_Omentum | 1.97 | 0.03 | 1.97 | 4.24 | 4.34 | 4.29 | 0.07 | 1 |
| CLIC1 | 1.69E-05 | 2 | 1 | 1.69E-05 | Adipose_Visceral_Omentum | 2.00 | 0.00 | 2.00 | -4.30 | -4.30 | -4.30 | 0.00 | 1 |
| CTNNB1 | 2.24E-05 | 2 | 1 | 2.24E-05 | Adipose_Subcutaneous | 2.00 | 0.00 | 2.00 | 4.24 | 4.24 | 4.24 | 0.00 | 1 |
| APOM | 2.57E-05 | 2 | 1 | 2.57E-05 | Adipose_Visceral_Omentum | 2.00 | 0.00 | 2.00 | -4.21 | -4.21 | -4.21 | 0.00 | 1 |
| MSH5 | 2.78E-05 | 2 | 2 | 7.78E-06 | Adipose_Subcutaneous | 1.79 | 0.21 | 0.21 | -4.47 | -4.13 | -4.30 | 0.24 | 2 |
| PRKCSH | 3.04E-05 | 2 | 1 | 3.04E-05 | Adipose_Subcutaneous | 2.00 | 0.00 | 2.00 | -4.17 | -4.17 | -4.17 | 0.00 | 1 |
| VWA7 | 3.27E-05 | 2 | 2 | 1.27E-05 | Adipose_Subcutaneous | 1.26 | 0.74 | 0.74 | -4.37 | -2.34 | -3.35 | 1.43 | 2 |
| C4A | 3.55E-05 | 2 | 1 | 1.75E-05 | Adipose_Subcutaneous | 1.96 | 0.04 | 1.96 | -4.30 | -3.89 | -4.09 | 0.29 | 1 |
| RBM6 | 3.72E-05 | 2 | 1 | 3.72E-05 | Adipose_Subcutaneous | 2.00 | 0.00 | 2.00 | -4.12 | -4.12 | -4.12 | 0.00 | 1 |
| TRAIP | 3.77E-05 | 2 | 1 | 3.77E-05 | Adipose_Visceral_Omentum | 2.00 | 0.00 | 2.00 | 4.12 | 4.12 | 4.12 | 0.00 | 1 |
| ACADSB | 6.21E-05 | 1 | 1 | 6.21E-05 | Adipose_Visceral_Omentum | 1.00 | 1.00 | 1.00 | 4.00 | 4.00 | 4.00 | NA | 1 |
| ERBB3 | 6.35E-05 | 2 | 2 | 2.01E-05 | Adipose_Subcutaneous | 1.86 | 0.14 | 0.14 | -4.26 | 4.21 | -0.03 | 5.99 | 2 |

| | | | | | | | | | | | | | |
|---|---|---|---|---|---|---|---|---|---|---|---|---|---|
| IKZF4 | 7.67E-05 | 2 | 2 | 1.41E-05 | Adipose_Subcutaneous | 1.91 | 0.09 | 0.09 | -4.34 | -4.07 | -4.21 | 0.19 | 2 |
| RNF123 | 7.99E-05 | 2 | 1 | 7.75E-05 | Adipose_Visceral_Omentum | 2.00 | 0.00 | 2.00 | 3.93 | 3.95 | 3.94 | 0.01 | 1 |
| BORCS6 | 8.01E-05 | 1 | 1 | 8.01E-05 | Adipose_Visceral_Omentum | 1.00 | 1.00 | 1.00 | 3.94 | 3.94 | 3.94 | NA | 1 |
| DNAJC22 | 8.79E-05 | 1 | 1 | 8.79E-05 | Adipose_Visceral_Omentum | 1.00 | 1.00 | 1.00 | 3.92 | 3.92 | 3.92 | NA | 1 |
| SYN2 | 8.81E-05 | 2 | 1 | 8.81E-05 | Adipose_Visceral_Omentum | 2.00 | 0.00 | 2.00 | -3.92 | -3.92 | -3.92 | 0.00 | 1 |
| LGR4 | 1.03E-04 | 2 | 2 | 0.015645196 | Adipose_Subcutaneous | 1.74 | 0.26 | 0.26 | -0.60 | 2.42 | 0.91 | 2.13 | 2 |
| MST1R | 1.05E-04 | 2 | 1 | 0.0001055 | Adipose_Visceral_Omentum | 2.00 | 0.00 | 2.00 | 3.88 | 3.88 | 3.88 | 0.00 | 1 |
| NAT6 | 1.13E-04 | 2 | 2 | 0.023230001 | Adipose_Subcutaneous | 1.82 | 0.18 | 0.18 | -2.27 | 0.21 | -1.03 | 1.76 | 2 |
| VAMP2 | 1.13E-04 | 2 | 1 | 4.68E-05 | Adipose_Subcutaneous | 1.95 | 0.05 | 1.95 | -4.07 | -3.55 | -3.81 | 0.37 | 1 |
| PPP1R18 | 1.18E-04 | 2 | 2 | 2.55E-05 | Adipose_Visceral_Omentum | 1.82 | 0.18 | 0.18 | -4.21 | -3.10 | -3.65 | 0.79 | 2 |
| SHTN1 | 1.19E-04 | 2 | 2 | 0.007564378 | Adipose_Subcutaneous | 1.90 | 0.10 | 0.10 | 0.92 | 2.67 | 1.80 | 1.24 | 2 |
| MAP3K20 | 1.23E-04 | 1 | 1 | 0.000123 | Adipose_Subcutaneous | 1.00 | 1.00 | 1.00 | -3.84 | -3.84 | -3.84 | NA | 1 |
| DHFR2 | 1.37E-04 | 2 | 1 | 0.0001137 | Adipose_Subcutaneous | 2.00 | 0.00 | 2.00 | -3.86 | -3.76 | -3.81 | 0.07 | 1 |
| FSCN2 | 1.44E-04 | 1 | 1 | 0.0001439 | Adipose_Visceral_Omentum | 1.00 | 1.00 | 1.00 | 3.80 | 3.80 | 3.80 | NA | 1 |
| ZC3H8 | 1.51E-04 | 2 | 2 | 4.09E-05 | Adipose_Visceral_Omentum | 1.90 | 0.10 | 0.10 | 4.08 | 4.10 | 4.09 | 0.02 | 2 |
| STX19 | 1.51E-04 | 2 | 1 | 0.0001511 | Adipose_Visceral_Omentum | 2.00 | 0.00 | 2.00 | 3.79 | 3.79 | 3.79 | 0.00 | 1 |
| CNPY4 | 1.56E-04 | 2 | 2 | 2.83E-05 | Adipose_Visceral_Omentum | 1.91 | 0.09 | 0.09 | 3.81 | 4.19 | 4.00 | 0.26 | 2 |
| GPANK1 | 1.62E-04 | 2 | 1 | 0.000172448 | Adipose_Subcutaneous | 1.96 | 0.04 | 1.96 | -3.76 | -3.71 | -3.73 | 0.04 | 1 |
| NEU1 | 1.66E-04 | 2 | 2 | 0.043205987 | Adipose_Visceral_Omentum | 1.90 | 0.10 | 0.10 | -2.02 | -0.21 | -1.11 | 1.28 | 2 |
| SPINT2 | 1.68E-04 | 2 | 2 | 9.48E-05 | Adipose_Visceral_Omentum | 1.70 | 0.30 | 0.30 | -3.90 | -1.67 | -2.78 | 1.58 | 2 |
| BNIPL | 1.68E-04 | 2 | 1 | 0.000159459 | Adipose_Subcutaneous | 2.00 | 0.00 | 2.00 | 3.75 | 3.78 | 3.76 | 0.02 | 1 |
| NR6A1 | 1.69E-04 | 1 | 1 | 0.0001685 | Adipose_Visceral_Omentum | 1.00 | 1.00 | 1.00 | 3.76 | 3.76 | 3.76 | NA | 1 |
| DCTN2 | 1.79E-04 | 1 | 1 | 0.0001795 | Adipose_Visceral_Omentum | 1.00 | 1.00 | 1.00 | -3.75 | -3.75 | -3.75 | NA | 1 |
| CNTLN | 2.01E-04 | 2 | 2 | 5.03E-05 | Adipose_Subcutaneous | 1.72 | 0.28 | 0.28 | -4.05 | -3.46 | -3.76 | 0.42 | 2 |
| B3GALT2 | 2.03E-04 | 1 | 1 | 0.0002029 | Adipose_Visceral_Omentum | 1.00 | 1.00 | 1.00 | -3.72 | -3.72 | -3.72 | NA | 1 |
| SYNCRIP | 2.07E-04 | 1 | 1 | 0.0002067 | Adipose_Subcutaneous | 1.00 | 1.00 | 1.00 | -3.71 | -3.71 | -3.71 | NA | 1 |
| HLA-B | 2.08E-04 | 2 | 2 | 0.016112899 | Adipose_Visceral_Omentum | 1.74 | 0.26 | 0.26 | -0.45 | 2.41 | 0.98 | 2.02 | 2 |
| R3HDM2 | 2.18E-04 | 1 | 1 | 0.0002184 | Adipose_Subcutaneous | 1.00 | 1.00 | 1.00 | -3.70 | -3.70 | -3.70 | NA | 1 |
| ZSCAN23 | 2.25E-04 | 2 | 1 | 0.000151291 | Adipose_Subcutaneous | 1.95 | 0.05 | 1.95 | 3.49 | 3.79 | 3.64 | 0.21 | 1 |
| PNISR | 2.26E-04 | 1 | 1 | 0.0002263 | Adipose_Subcutaneous | 1.00 | 1.00 | 1.00 | 3.69 | 3.69 | 3.69 | NA | 1 |
| SELENOH | 2.28E-04 | 2 | 1 | 0.0002283 | Adipose_Subcutaneous | 2.00 | 0.00 | 2.00 | 3.69 | 3.69 | 3.69 | 0.00 | 1 |
| RGPD8 | 2.31E-04 | 2 | 2 | 0.000247443 | Adipose_Subcutaneous | 1.92 | 0.08 | 0.08 | -3.66 | -2.63 | -3.14 | 0.74 | 2 |
| METAP1D | 2.38E-04 | 2 | 1 | 0.0002338 | Adipose_Visceral_Omentum | 2.00 | 0.00 | 2.00 | 3.67 | 3.68 | 3.68 | 0.01 | 1 |

| | | | | | | | | | | | | | |
|---|---|---|---|---|---|---|---|---|---|---|---|---|---|
| TMEM89 | 2.43E-04 | 1 | 1 | 0.0002427 | Adipose_Subcutaneous | 1.00 | 1.00 | 1.00 | -3.67 | -3.67 | -3.67 | NA | 1 |
| TMEM14A | 2.43E-04 | 1 | 1 | 0.0002432 | Adipose_Visceral_Omentum | 1.00 | 1.00 | 1.00 | 3.67 | 3.67 | 3.67 | NA | 1 |
| FOSL2 | 2.61E-04 | 2 | 2 | 9.79E-05 | Adipose_Visceral_Omentum | 1.20 | 0.80 | 0.80 | -3.90 | 1.92 | -0.99 | 4.11 | 2 |
| SFTA2 | 2.88E-04 | 2 | 2 | 0.036609991 | Adipose_Visceral_Omentum | 1.54 | 0.46 | 0.46 | -1.80 | 2.09 | 0.15 | 2.75 | 2 |
| DBN1 | 3.08E-04 | 1 | 1 | 0.0003084 | Adipose_Visceral_Omentum | 1.00 | 1.00 | 1.00 | -3.61 | -3.61 | -3.61 | NA | 1 |
| IKZF5 | 3.39E-04 | 2 | 2 | 0.001125739 | Adipose_Subcutaneous | 1.90 | 0.10 | 0.10 | 1.93 | 3.26 | 2.59 | 0.94 | 2 |
| LINC01336 | 3.50E-04 | 1 | 1 | 0.0003502 | Adipose_Subcutaneous | 1.00 | 1.00 | 1.00 | 3.58 | 3.58 | 3.58 | NA | 1 |
| STAC3 | 3.75E-04 | 2 | 1 | 0.0003749 | Adipose_Subcutaneous | 2.00 | 0.00 | 2.00 | -3.56 | -3.56 | -3.56 | 0.00 | 1 |
| ACTG1 | 3.86E-04 | 2 | 2 | 7.89E-05 | Adipose_Subcutaneous | 1.73 | 0.27 | 0.27 | 2.64 | 3.95 | 3.29 | 0.93 | 2 |
| CDC27 | 4.10E-04 | 1 | 1 | 0.0004104 | Adipose_Visceral_Omentum | 1.00 | 1.00 | 1.00 | -3.53 | -3.53 | -3.53 | NA | 1 |
| HACD4 | 4.17E-04 | 2 | 2 | 9.66E-05 | Adipose_Subcutaneous | 1.65 | 0.35 | 0.35 | 2.08 | 3.90 | 2.99 | 1.28 | 2 |
| KNOP1 | 4.18E-04 | 2 | 1 | 0.0003759 | Adipose_Visceral_Omentum | 1.99 | 0.01 | 1.99 | -3.56 | -3.49 | -3.52 | 0.05 | 1 |
| CDC73 | 4.25E-04 | 1 | 1 | 0.0004251 | Adipose_Visceral_Omentum | 1.00 | 1.00 | 1.00 | -3.52 | -3.52 | -3.52 | NA | 1 |
| ITK | 4.36E-04 | 2 | 1 | 0.0004358 | Adipose_Visceral_Omentum | 2.00 | 0.00 | 2.00 | -3.52 | -3.52 | -3.52 | 0.00 | 1 |
| HLA-C | 4.70E-04 | 2 | 2 | 0.125286782 | Adipose_Visceral_Omentum | 1.83 | 0.17 | 0.17 | -0.74 | 1.53 | 0.40 | 1.61 | 2 |
| GAS1RR | 4.82E-04 | 2 | 1 | 0.000278234 | Adipose_Subcutaneous | 1.98 | 0.02 | 1.98 | -3.63 | -3.30 | -3.47 | 0.23 | 1 |
| ADAM22 | 4.89E-04 | 1 | 1 | 0.0004892 | Adipose_Visceral_Omentum | 1.00 | 1.00 | 1.00 | 3.49 | 3.49 | 3.49 | NA | 1 |
| ZSCAN12 | 4.94E-04 | 1 | 1 | 0.000494418 | Adipose_Visceral_Omentum | 1.00 | 1.00 | 1.00 | 3.48 | 3.48 | 3.48 | NA | 1 |
| SNX14 | 4.95E-04 | 2 | 2 | 0.000129992 | Adipose_Subcutaneous | 1.78 | 0.22 | 0.22 | 3.46 | 3.83 | 3.64 | 0.26 | 2 |
| NDUFS3 | 4.96E-04 | 2 | 2 | 0.002955999 | Adipose_Visceral_Omentum | 1.07 | 0.93 | 0.93 | 2.32 | 2.97 | 2.65 | 0.46 | 2 |
| SLC17A7 | 5.12E-04 | 2 | 2 | 0.001017 | Adipose_Visceral_Omentum | 1.23 | 0.77 | 0.77 | 2.80 | 3.29 | 3.04 | 0.34 | 2 |
| PIAS2 | 5.16E-04 | 2 | 1 | 0.0005163 | Adipose_Visceral_Omentum | 2.00 | 0.00 | 2.00 | -3.47 | -3.47 | -3.47 | 0.00 | 1 |
| UTP18 | 5.20E-04 | 2 | 2 | 0.001684 | Adipose_Subcutaneous | 1.74 | 0.26 | 0.26 | -3.14 | -0.79 | -1.96 | 1.67 | 2 |
| FAM98C | 5.32E-04 | 2 | 2 | 0.000109522 | Adipose_Visceral_Omentum | 1.49 | 0.51 | 0.51 | -3.87 | -1.60 | -2.73 | 1.61 | 2 |
| ZKSCAN3 | 5.46E-04 | 2 | 1 | 0.0004429 | Adipose_Visceral_Omentum | 1.98 | 0.02 | 1.98 | -3.51 | -3.37 | -3.44 | 0.10 | 1 |
| CUL3 | 5.59E-04 | 2 | 2 | 0.0001762 | Adipose_Subcutaneous | 1.42 | 0.58 | 0.58 | -3.75 | -0.72 | -2.24 | 2.14 | 2 |
| UQCC2 | 5.64E-04 | 2 | 2 | 0.004281724 | Adipose_Visceral_Omentum | 1.26 | 0.74 | 0.74 | -2.86 | 1.76 | -0.55 | 3.26 | 2 |
| ZFP91 | 5.81E-04 | 1 | 1 | 0.0005809 | Adipose_Subcutaneous | 1.00 | 1.00 | 1.00 | -3.44 | -3.44 | -3.44 | NA | 1 |
| ZSCAN26 | 5.85E-04 | 2 | 1 | 0.000556177 | Adipose_Visceral_Omentum | 2.00 | 0.00 | 2.00 | -3.45 | -3.42 | -3.44 | 0.02 | 1 |
| FNBP4 | 5.86E-04 | 1 | 1 | 0.0005856 | Adipose_Subcutaneous | 1.00 | 1.00 | 1.00 | 3.44 | 3.44 | 3.44 | NA | 1 |
| JRK | 5.97E-04 | 2 | 1 | 0.0004819 | Adipose_Subcutaneous | 1.95 | 0.05 | 1.95 | -3.49 | -3.29 | -3.39 | 0.14 | 1 |
| RP11-227D13.1 | 5.99E-04 | 2 | 1 | 0.0005994 | Adipose_Subcutaneous | 2.00 | 0.00 | 2.00 | -3.43 | -3.43 | -3.43 | 0.00 | 1 |
| TRIM33 | 6.09E-04 | 1 | 1 | 0.0006089 | Adipose_Visceral_Omentum | 1.00 | 1.00 | 1.00 | -3.43 | -3.43 | -3.43 | NA | 1 |

| | | | | | | | | | | | | | |
|---|---|---|---|---|---|---|---|---|---|---|---|---|---|
| IQCK | 6.17E-04 | 1 | 1 | 0.0006175 | Adipose_Visceral_Omentum | 1.00 | 1.00 | 1.00 | 3.42 | 3.42 | 3.42 | NA | 1 |
| MYO5A | 6.50E-04 | 2 | 2 | 0.000199003 | Adipose_Visceral_Omentum | 1.85 | 0.15 | 0.15 | 3.65 | 3.72 | 3.68 | 0.05 | 2 |
| RP11-6O2.2 | 6.75E-04 | 1 | 1 | 0.0006754 | Adipose_Subcutaneous | 1.00 | 1.00 | 1.00 | -3.40 | -3.40 | -3.40 | NA | 1 |
| OLFML2A | 6.79E-04 | 2 | 1 | 0.0004138 | Adipose_Subcutaneous | 1.99 | 0.01 | 1.99 | -3.53 | -3.25 | -3.39 | 0.20 | 1 |
| CCL15 | 6.86E-04 | 2 | 2 | 0.000159459 | Adipose_Visceral_Omentum | 1.74 | 0.26 | 0.26 | 3.18 | 3.78 | 3.48 | 0.42 | 2 |
| DVL2 | 6.93E-04 | 1 | 1 | 0.0006928 | Adipose_Visceral_Omentum | 1.00 | 1.00 | 1.00 | -3.39 | -3.39 | -3.39 | NA | 1 |
| P2RY1 | 6.94E-04 | 2 | 2 | 0.000405572 | Adipose_Visceral_Omentum | 1.46 | 0.54 | 0.54 | 2.89 | 3.54 | 3.21 | 0.46 | 2 |
| SPATA2 | 6.96E-04 | 2 | 2 | 0.000298471 | Adipose_Visceral_Omentum | 1.86 | 0.14 | 0.14 | 2.49 | 3.62 | 3.05 | 0.80 | 2 |
| ZKSCAN4 | 7.03E-04 | 2 | 2 | 0.000330459 | Adipose_Visceral_Omentum | 1.74 | 0.26 | 0.26 | 3.51 | 3.59 | 3.55 | 0.06 | 2 |
| NMT2 | 7.12E-04 | 2 | 2 | 0.004460769 | Adipose_Subcutaneous | 1.60 | 0.40 | 0.40 | -2.84 | 0.32 | -1.26 | 2.24 | 2 |
| DYNC1I2 | 7.33E-04 | 1 | 1 | 0.0007328 | Adipose_Subcutaneous | 1.00 | 1.00 | 1.00 | 3.38 | 3.38 | 3.38 | NA | 1 |
| RP11-76C10.5 | 7.35E-04 | 2 | 2 | 0.0001455 | Adipose_Subcutaneous | 1.37 | 0.63 | 0.63 | -3.80 | -1.38 | -2.59 | 1.71 | 2 |
| MRO | 7.43E-04 | 2 | 2 | 0.000154638 | Adipose_Visceral_Omentum | 1.13 | 0.87 | 0.87 | -3.78 | -0.80 | -2.29 | 2.11 | 2 |
| PNMA1 | 7.67E-04 | 2 | 1 | 0.000899032 | Adipose_Subcutaneous | 1.94 | 0.06 | 1.94 | -3.32 | -3.30 | -3.31 | 0.01 | 1 |
| GABARAP | 7.87E-04 | 1 | 1 | 0.0007871 | Adipose_Subcutaneous | 1.00 | 1.00 | 1.00 | 3.36 | 3.36 | 3.36 | NA | 1 |
| SWT1 | 7.96E-04 | 2 | 2 | 0.002820591 | Adipose_Subcutaneous | 1.63 | 0.37 | 0.37 | 0.10 | 2.99 | 1.54 | 2.04 | 2 |
| CHAC2 | 7.99E-04 | 2 | 2 | 0.000902122 | Adipose_Subcutaneous | 1.39 | 0.61 | 0.61 | -2.96 | 3.32 | 0.18 | 4.44 | 2 |
| CEP290 | 8.08E-04 | 2 | 1 | 0.000704565 | Adipose_Subcutaneous | 2.00 | 0.00 | 2.00 | 3.31 | 3.39 | 3.35 | 0.05 | 1 |
| ZNF653 | 8.21E-04 | 2 | 1 | 0.0008214 | Adipose_Visceral_Omentum | 2.00 | 0.00 | 2.00 | -3.35 | -3.35 | -3.35 | 0.00 | 1 |
| NF2 | 8.24E-04 | 2 | 2 | 0.000960254 | Adipose_Visceral_Omentum | 1.09 | 0.91 | 0.91 | 2.12 | 3.30 | 2.71 | 0.84 | 2 |
| FGF14-AS2 | 8.38E-04 | 2 | 2 | 0.000319119 | Adipose_Subcutaneous | 1.84 | 0.16 | 0.16 | 2.42 | 3.60 | 3.01 | 0.83 | 2 |
| CTD-2325M2.1 | 8.60E-04 | 2 | 1 | 0.00092463 | Adipose_Visceral_Omentum | 1.97 | 0.03 | 1.97 | -3.31 | -3.31 | -3.31 | 0.00 | 1 |
| OPRD1 | 8.98E-04 | 1 | 1 | 0.000898455 | Adipose_Visceral_Omentum | 1.00 | 1.00 | 1.00 | 3.32 | 3.32 | 3.32 | NA | 1 |
| RAD51 | 9.17E-04 | 2 | 2 | 0.0004889 | Adipose_Subcutaneous | 1.15 | 0.85 | 0.85 | -3.49 | 0.82 | -1.33 | 3.05 | 2 |
| ARPP19 | 9.72E-04 | 1 | 1 | 0.000972067 | Adipose_Visceral_Omentum | 1.00 | 1.00 | 1.00 | -3.30 | -3.30 | -3.30 | NA | 1 |
| KBTBD4 | 9.88E-04 | 2 | 1 | 0.0009879 | Adipose_Subcutaneous | 2.00 | 0.00 | 2.00 | -3.29 | -3.29 | -3.29 | 0.00 | 1 |
| LOXL4 | 1.01E-03 | 2 | 2 | 0.024850005 | Adipose_Visceral_Omentum | 1.36 | 0.64 | 0.64 | -2.24 | 1.94 | -0.15 | 2.96 | 2 |
| HORMAD2 | 1.01E-03 | 1 | 1 | 0.001014 | Adipose_Subcutaneous | 1.00 | 1.00 | 1.00 | 3.29 | 3.29 | 3.29 | NA | 1 |
| SMARCAL1 | 1.02E-03 | 2 | 1 | 0.00092357 | Adipose_Subcutaneous | 2.00 | 0.00 | 2.00 | -3.31 | -3.25 | -3.28 | 0.04 | 1 |
| PWWP2A | 1.02E-03 | 2 | 2 | 0.000207012 | Adipose_Subcutaneous | 1.71 | 0.29 | 0.29 | -3.71 | -2.65 | -3.18 | 0.75 | 2 |
| ACSF2 | 1.04E-03 | 2 | 2 | 0.000898088 | Adipose_Visceral_Omentum | 1.60 | 0.40 | 0.40 | 3.30 | 3.32 | 3.31 | 0.01 | 2 |
| IL1RN | 1.05E-03 | 1 | 1 | 0.001048 | Adipose_Subcutaneous | 1.00 | 1.00 | 1.00 | -3.28 | -3.28 | -3.28 | NA | 1 |
| NLRP1 | 1.06E-03 | 2 | 2 | 0.000225879 | Adipose_Subcutaneous | 1.67 | 0.33 | 0.33 | -3.69 | -2.72 | -3.20 | 0.68 | 2 |

| | | | | | | | | | | | | | |
|---|---|---|---|---|---|---|---|---|---|---|---|---|---|
| ZBTB25 | 1.06E-03 | 2 | 2 | 0.000215395 | Adipose_Subcutaneous | 1.88 | 0.12 | 0.12 | 3.19 | 3.70 | 3.45 | 0.36 | 2 |
| FCN2 | 1.07E-03 | 2 | 2 | 0.003198001 | Adipose_Subcutaneous | 1.01 | 0.99 | 0.99 | -2.95 | 2.26 | -0.34 | 3.68 | 2 |
| R3HCC1L | 1.07E-03 | 2 | 2 | 0.000858003 | Adipose_Subcutaneous | 1.39 | 0.61 | 0.61 | -3.33 | -2.78 | -3.06 | 0.39 | 2 |
| LIN37 | 1.09E-03 | 2 | 1 | 0.000989442 | Adipose_Visceral_Omentum | 1.96 | 0.04 | 1.96 | 3.17 | 3.29 | 3.23 | 0.09 | 1 |
| KHDRBS3 | 1.09E-03 | 2 | 2 | 0.000226876 | Adipose_Visceral_Omentum | 1.00 | 1.00 | 1.00 | 0.22 | 3.69 | 1.95 | 2.45 | 2 |
| ACAT1 | 1.10E-03 | 2 | 1 | 0.001103 | Adipose_Subcutaneous | 2.00 | 0.00 | 2.00 | -3.26 | -3.26 | -3.26 | 0.00 | 1 |
| YBX2 | 1.11E-03 | 2 | 1 | 0.001113 | Adipose_Visceral_Omentum | 2.00 | 0.00 | 2.00 | 3.26 | 3.26 | 3.26 | 0.00 | 1 |
| PRR7 | 1.13E-03 | 2 | 1 | 0.001129 | Adipose_Subcutaneous | 2.00 | 0.00 | 2.00 | 3.26 | 3.26 | 3.26 | 0.00 | 1 |
| ZNF80 | 1.15E-03 | 2 | 1 | 0.001265 | Adipose_Visceral_Omentum | 1.95 | 0.05 | 1.95 | -3.22 | -3.20 | -3.21 | 0.01 | 1 |
| KCNV2 | 1.16E-03 | 2 | 2 | 0.0004202 | Adipose_Subcutaneous | 1.15 | 0.85 | 0.85 | -3.53 | -1.56 | -2.54 | 1.39 | 2 |
| MGMT | 1.16E-03 | 2 | 1 | 0.00115105 | Adipose_Subcutaneous | 2.00 | 0.00 | 2.00 | 3.25 | 3.25 | 3.25 | 0.00 | 1 |
| WDR35 | 1.17E-03 | 2 | 2 | 0.000254642 | Adipose_Visceral_Omentum | 1.91 | 0.09 | 0.09 | -3.66 | -3.20 | -3.43 | 0.32 | 2 |
| EFTUD2 | 1.18E-03 | 2 | 2 | 0.000286947 | Adipose_Subcutaneous | 1.59 | 0.41 | 0.41 | 2.61 | 3.63 | 3.12 | 0.72 | 2 |
| FCN1 | 1.20E-03 | 2 | 1 | 0.0008463 | Adipose_Visceral_Omentum | 1.97 | 0.03 | 1.97 | 3.09 | 3.34 | 3.22 | 0.17 | 1 |
| SLC9A8 | 1.23E-03 | 2 | 1 | 0.001412207 | Adipose_Subcutaneous | 1.94 | 0.06 | 1.94 | -3.19 | -3.18 | -3.19 | 0.01 | 1 |
| RP11-392O17.2 | 1.27E-03 | 2 | 2 | 0.005219999 | Adipose_Visceral_Omentum | 1.11 | 0.89 | 0.89 | -2.79 | 2.04 | -0.38 | 3.42 | 2 |
| MED19 | 1.27E-03 | 2 | 2 | 0.001277 | Adipose_Visceral_Omentum | 1.06 | 0.94 | 0.94 | -1.91 | 3.22 | 0.65 | 3.63 | 2 |
| DDX5 | 1.28E-03 | 2 | 1 | 0.001438 | Adipose_Visceral_Omentum | 1.95 | 0.05 | 1.95 | -3.19 | -3.18 | -3.18 | 0.01 | 1 |
| DNASE1 | 1.29E-03 | 2 | 2 | 0.0002639 | Adipose_Visceral_Omentum | 1.12 | 0.88 | 0.88 | -0.40 | 3.65 | 1.63 | 2.86 | 2 |
| STYX | 1.30E-03 | 2 | 1 | 0.001201831 | Adipose_Subcutaneous | 1.99 | 0.01 | 1.99 | -3.24 | -3.18 | -3.21 | 0.04 | 1 |
| ZBTB32 | 1.31E-03 | 2 | 2 | 0.001881737 | Adipose_Subcutaneous | 1.90 | 0.10 | 0.10 | 1.94 | 3.11 | 2.52 | 0.83 | 2 |
| TMEM198 | 1.33E-03 | 2 | 2 | 0.0004182 | Adipose_Visceral_Omentum | 1.33 | 0.67 | 0.67 | -3.53 | -1.99 | -2.76 | 1.08 | 2 |
| RP11-333I13.1 | 1.36E-03 | 1 | 1 | 0.001357534 | Adipose_Visceral_Omentum | 1.00 | 1.00 | 1.00 | 3.20 | 3.20 | 3.20 | NA | 1 |
| SIGLEC5 | 1.36E-03 | 2 | 2 | 0.035006413 | Adipose_Subcutaneous | 1.52 | 0.48 | 0.48 | -1.41 | 2.11 | 0.35 | 2.49 | 2 |
| BRD3 | 1.37E-03 | 2 | 2 | 0.009003001 | Adipose_Visceral_Omentum | 1.90 | 0.10 | 0.10 | 1.25 | 2.61 | 1.93 | 0.97 | 2 |
| HSDL2 | 1.39E-03 | 2 | 2 | 0.00042674 | Adipose_Visceral_Omentum | 1.71 | 0.29 | 0.29 | -3.52 | -1.90 | -2.71 | 1.15 | 2 |
| USP53 | 1.42E-03 | 2 | 2 | 0.000321689 | Adipose_Subcutaneous | 1.93 | 0.07 | 0.07 | 3.21 | 3.60 | 3.41 | 0.27 | 2 |
| C11orf58 | 1.43E-03 | 2 | 2 | 0.013419996 | Adipose_Subcutaneous | 1.21 | 0.79 | 0.79 | 2.07 | 2.47 | 2.27 | 0.28 | 2 |
| RAB5B | 1.44E-03 | 2 | 2 | 0.0003517 | Adipose_Visceral_Omentum | 1.26 | 0.74 | 0.74 | -3.57 | -0.37 | -1.97 | 2.27 | 2 |
| CEP95 | 1.44E-03 | 2 | 1 | 0.001438 | Adipose_Visceral_Omentum | 2.00 | 0.00 | 2.00 | 3.19 | 3.19 | 3.19 | 0.00 | 1 |
| RP1-265C24.8 | 1.44E-03 | 1 | 1 | 0.001439 | Adipose_Visceral_Omentum | 1.00 | 1.00 | 1.00 | 3.19 | 3.19 | 3.19 | NA | 1 |
| AFF3 | 1.45E-03 | 1 | 1 | 0.001452 | Adipose_Subcutaneous | 1.00 | 1.00 | 1.00 | -3.18 | -3.18 | -3.18 | NA | 1 |
| GLRB | 1.45E-03 | 1 | 1 | 0.001452 | Adipose_Visceral_Omentum | 1.00 | 1.00 | 1.00 | -3.18 | -3.18 | -3.18 | NA | 1 |

| | | | | | | | | | | | | | |
|---|---|---|---|---|---|---|---|---|---|---|---|---|---|
| C14orf93 | 1.48E-03 | 2 | 2 | 0.0005112 | Adipose_Visceral_Omentum | 1.76 | 0.24 | 0.24 | 2.03 | 3.47 | 2.75 | 1.02 | 2 |
| GLRX2 | 1.49E-03 | 2 | 2 | 0.0006561 | Adipose_Visceral_Omentum | 1.89 | 0.11 | 0.11 | -3.41 | -2.49 | -2.95 | 0.65 | 2 |
| ZSCAN31 | 1.50E-03 | 2 | 2 | 0.000359356 | Adipose_Subcutaneous | 1.85 | 0.15 | 0.15 | 2.75 | 3.57 | 3.16 | 0.58 | 2 |
| RNF5 | 1.52E-03 | 2 | 1 | 0.001517 | Adipose_Subcutaneous | 2.00 | 0.00 | 2.00 | -3.17 | -3.17 | -3.17 | 0.00 | 1 |
| GAREM2 | 1.54E-03 | 2 | 2 | 0.243952636 | Adipose_Visceral_Omentum | 1.87 | 0.13 | 0.13 | -1.17 | 0.65 | -0.26 | 1.28 | 2 |
| REPS1 | 1.54E-03 | 2 | 1 | 0.001522 | Adipose_Visceral_Omentum | 1.99 | 0.01 | 1.99 | 3.15 | 3.17 | 3.16 | 0.02 | 1 |
| EFL1 | 1.54E-03 | 2 | 2 | 0.108046958 | Adipose_Visceral_Omentum | 1.80 | 0.20 | 0.20 | -0.64 | 1.61 | 0.49 | 1.59 | 2 |
| YIF1B | 1.56E-03 | 2 | 2 | 0.001712001 | Adipose_Subcutaneous | 1.07 | 0.93 | 0.93 | 1.96 | 3.14 | 2.55 | 0.83 | 2 |
| MINOS1 | 1.58E-03 | 2 | 2 | 0.000372071 | Adipose_Subcutaneous | 1.92 | 0.08 | 0.08 | -3.56 | -3.47 | -3.51 | 0.06 | 2 |
| AC007405.6 | 1.58E-03 | 1 | 1 | 0.001584122 | Adipose_Visceral_Omentum | 1.00 | 1.00 | 1.00 | -3.16 | -3.16 | -3.16 | NA | 1 |
| CEBPZ | 1.62E-03 | 2 | 1 | 0.001139711 | Adipose_Visceral_Omentum | 1.98 | 0.02 | 1.98 | -3.25 | -3.03 | -3.14 | 0.16 | 1 |
| SEC11C | 1.67E-03 | 2 | 1 | 0.001371 | Adipose_Subcutaneous | 1.99 | 0.01 | 1.99 | 3.07 | 3.20 | 3.14 | 0.09 | 1 |
| KRBA2 | 1.69E-03 | 2 | 2 | 0.083827479 | Adipose_Visceral_Omentum | 1.91 | 0.09 | 0.09 | -1.73 | -0.31 | -1.02 | 1.00 | 2 |
| LRP1B | 1.74E-03 | 2 | 2 | 0.0004129 | Adipose_Visceral_Omentum | 1.76 | 0.24 | 0.24 | 3.00 | 3.53 | 3.27 | 0.38 | 2 |
| CEP97 | 1.76E-03 | 1 | 1 | 0.001764 | Adipose_Visceral_Omentum | 1.00 | 1.00 | 1.00 | 3.13 | 3.13 | 3.13 | NA | 1 |
| MDH2 | 1.79E-03 | 2 | 1 | 0.0009106 | Adipose_Subcutaneous | 1.95 | 0.05 | 1.95 | -3.32 | -2.86 | -3.09 | 0.33 | 1 |
| ZNF165 | 1.80E-03 | 2 | 1 | 0.001803 | Adipose_Subcutaneous | 2.00 | 0.00 | 2.00 | -3.12 | -3.12 | -3.12 | 0.00 | 1 |
| HSPG2 | 1.81E-03 | 1 | 1 | 0.001811058 | Adipose_Subcutaneous | 1.00 | 1.00 | 1.00 | -3.12 | -3.12 | -3.12 | NA | 1 |
| DDX31 | 1.83E-03 | 2 | 1 | 0.001441119 | Adipose_Subcutaneous | 1.99 | 0.01 | 1.99 | -3.19 | -3.04 | -3.11 | 0.11 | 1 |
| SH2B3 | 1.83E-03 | 1 | 1 | 0.001834 | Adipose_Subcutaneous | 1.00 | 1.00 | 1.00 | -3.12 | -3.12 | -3.12 | NA | 1 |
| OXER1 | 1.87E-03 | 2 | 1 | 0.001696294 | Adipose_Visceral_Omentum | 1.99 | 0.01 | 1.99 | -3.14 | -3.06 | -3.10 | 0.05 | 1 |
| SMARCC2 | 1.88E-03 | 2 | 1 | 0.002130001 | Adipose_Subcutaneous | 1.95 | 0.05 | 1.95 | -3.07 | -3.07 | -3.07 | 0.00 | 1 |
| SCMH1 | 1.89E-03 | 2 | 2 | 0.002339922 | Adipose_Subcutaneous | 1.36 | 0.64 | 0.64 | -3.04 | 0.60 | -1.22 | 2.58 | 2 |
| CKMT1A | 1.90E-03 | 1 | 1 | 0.001897001 | Adipose_Visceral_Omentum | 1.00 | 1.00 | 1.00 | 3.11 | 3.11 | 3.11 | NA | 1 |
| SLC38A6 | 1.92E-03 | 2 | 1 | 0.001931284 | Adipose_Visceral_Omentum | 2.00 | 0.00 | 2.00 | -3.10 | -3.10 | -3.10 | 0.00 | 1 |
| SCUBE1 | 1.92E-03 | 2 | 2 | 0.0004049 | Adipose_Subcutaneous | 1.04 | 0.96 | 0.96 | -3.54 | -0.16 | -1.85 | 2.38 | 2 |
| CHAD | 1.93E-03 | 2 | 2 | 0.002193 | Adipose_Subcutaneous | 1.89 | 0.11 | 0.11 | 1.94 | 3.06 | 2.50 | 0.79 | 2 |
| LHPP | 1.95E-03 | 2 | 2 | 0.001344277 | Adipose_Visceral_Omentum | 1.62 | 0.38 | 0.38 | 3.14 | 3.21 | 3.18 | 0.04 | 2 |
| TMEM117 | 1.95E-03 | 2 | 2 | 0.0004411 | Adipose_Visceral_Omentum | 1.82 | 0.18 | 0.18 | -3.51 | -2.67 | -3.09 | 0.59 | 2 |
| CYP3A5 | 1.97E-03 | 2 | 1 | 0.00197 | Adipose_Subcutaneous | 2.00 | 0.00 | 2.00 | 3.09 | 3.09 | 3.09 | 0.00 | 1 |
| DDAH2 | 1.97E-03 | 2 | 2 | 0.000418 | Adipose_Subcutaneous | 1.37 | 0.63 | 0.63 | -3.53 | -1.36 | -2.45 | 1.53 | 2 |
| CYP1A1 | 1.98E-03 | 2 | 2 | 0.010310003 | Adipose_Subcutaneous | 1.93 | 0.07 | 0.07 | -2.57 | -1.52 | -2.04 | 0.74 | 2 |
| ANAPC4 | 2.00E-03 | 2 | 1 | 0.001881733 | Adipose_Subcutaneous | 2.00 | 0.00 | 2.00 | 3.07 | 3.11 | 3.09 | 0.03 | 1 |

| | | | | | | | | | | | | | |
|---|---|---|---|---|---|---|---|---|---|---|---|---|---|
| GPR1 | 2.00E-03 | 2 | 2 | 0.024149998 | Adipose_Visceral_Omentum | 1.40 | 0.60 | 0.60 | -1.59 | 2.25 | 0.33 | 2.72 | 2 |
| PXDNL | 2.01E-03 | 2 | 2 | 0.000472031 | Adipose_Visceral_Omentum | 1.91 | 0.09 | 0.09 | 3.38 | 3.50 | 3.44 | 0.09 | 2 |
| SPATA25 | 2.02E-03 | 2 | 2 | 0.017994536 | Adipose_Visceral_Omentum | 1.53 | 0.47 | 0.47 | -0.94 | 2.37 | 0.71 | 2.34 | 2 |
| SLC25A35 | 2.03E-03 | 2 | 1 | 0.001793211 | Adipose_Visceral_Omentum | 2.00 | 0.00 | 2.00 | 3.05 | 3.12 | 3.09 | 0.05 | 1 |
| ELP5 | 2.04E-03 | 2 | 2 | 0.000492928 | Adipose_Subcutaneous | 1.88 | 0.12 | 0.12 | -3.48 | -3.30 | -3.39 | 0.13 | 2 |
| TMC1 | 2.08E-03 | 2 | 1 | 0.002084 | Adipose_Subcutaneous | 2.00 | 0.00 | 2.00 | 3.08 | 3.08 | 3.08 | 0.00 | 1 |
| SEMA3F | 2.13E-03 | 2 | 1 | 0.002132 | Adipose_Subcutaneous | 2.00 | 0.00 | 2.00 | -3.07 | -3.07 | -3.07 | 0.00 | 1 |
| CSNK2B | 2.13E-03 | 2 | 2 | 0.005532001 | Adipose_Visceral_Omentum | 1.13 | 0.87 | 0.87 | -1.76 | 2.77 | 0.51 | 3.20 | 2 |
| CLP1 | 2.14E-03 | 1 | 1 | 0.002137001 | Adipose_Visceral_Omentum | 1.00 | 1.00 | 1.00 | -3.07 | -3.07 | -3.07 | NA | 1 |
| RP11-486M23.1 | 2.14E-03 | 1 | 1 | 0.002142 | Adipose_Subcutaneous | 1.00 | 1.00 | 1.00 | -3.07 | -3.07 | -3.07 | NA | 1 |
| GGNBP2 | 2.15E-03 | 2 | 1 | 0.001991542 | Adipose_Visceral_Omentum | 1.99 | 0.01 | 1.99 | -3.09 | -3.03 | -3.06 | 0.05 | 1 |
| SMAD3 | 2.17E-03 | 1 | 1 | 0.002173346 | Adipose_Subcutaneous | 1.00 | 1.00 | 1.00 | 3.07 | 3.07 | 3.07 | NA | 1 |
| TMEM200B | 2.23E-03 | 2 | 1 | 0.002050001 | Adipose_Subcutaneous | 1.96 | 0.04 | 1.96 | -3.08 | -2.98 | -3.03 | 0.07 | 1 |
| FAM124B | 2.23E-03 | 2 | 2 | 0.000882257 | Adipose_Subcutaneous | 1.32 | 0.68 | 0.68 | -3.33 | -0.05 | -1.69 | 2.32 | 2 |
| CXCL5 | 2.23E-03 | 2 | 1 | 0.002234001 | Adipose_Subcutaneous | 2.00 | 0.00 | 2.00 | -3.06 | -3.06 | -3.06 | 0.00 | 1 |
| LZTS1 | 2.24E-03 | 1 | 1 | 0.00224 | Adipose_Subcutaneous | 1.00 | 1.00 | 1.00 | -3.06 | -3.06 | -3.06 | NA | 1 |
| ATXN2 | 2.25E-03 | 2 | 1 | 0.002245 | Adipose_Visceral_Omentum | 2.00 | 0.00 | 2.00 | 3.06 | 3.06 | 3.06 | 0.00 | 1 |
| ZNF850 | 2.28E-03 | 1 | 1 | 0.002284999 | Adipose_Subcutaneous | 1.00 | 1.00 | 1.00 | 3.05 | 3.05 | 3.05 | NA | 1 |
| CLCNKB | 2.32E-03 | 2 | 2 | 0.008982999 | Adipose_Subcutaneous | 1.10 | 0.90 | 0.90 | -2.61 | -2.54 | -2.58 | 0.05 | 2 |
| RP11-9M16.2 | 2.33E-03 | 2 | 2 | 0.002304534 | Adipose_Subcutaneous | 1.93 | 0.07 | 0.07 | 2.23 | 3.05 | 2.64 | 0.58 | 2 |
| PSMC6 | 2.33E-03 | 2 | 1 | 0.002333 | Adipose_Visceral_Omentum | 2.00 | 0.00 | 2.00 | 3.04 | 3.04 | 3.04 | 0.00 | 1 |
| CACTIN | 2.35E-03 | 2 | 2 | 0.062264708 | Adipose_Subcutaneous | 1.92 | 0.08 | 0.08 | 0.58 | 1.86 | 1.22 | 0.91 | 2 |
| PARD3 | 2.35E-03 | 2 | 1 | 0.002261995 | Adipose_Subcutaneous | 1.95 | 0.05 | 1.95 | 2.95 | 3.05 | 3.00 | 0.07 | 1 |
| RACK1 | 2.40E-03 | 2 | 2 | 0.05759837 | Adipose_Subcutaneous | 1.92 | 0.08 | 0.08 | -1.90 | -0.57 | -1.23 | 0.94 | 2 |
| MICB | 2.40E-03 | 2 | 1 | 0.001807145 | Adipose_Subcutaneous | 1.96 | 0.04 | 1.96 | 2.89 | 3.12 | 3.01 | 0.16 | 1 |
| LRFN5 | 2.41E-03 | 2 | 1 | 0.002405001 | Adipose_Subcutaneous | 2.00 | 0.00 | 2.00 | 3.04 | 3.04 | 3.04 | 0.00 | 1 |
| ZNHIT3 | 2.47E-03 | 1 | 1 | 0.002467947 | Adipose_Visceral_Omentum | 1.00 | 1.00 | 1.00 | 3.03 | 3.03 | 3.03 | NA | 1 |
| YPEL4 | 2.48E-03 | 2 | 1 | 0.002482 | Adipose_Subcutaneous | 2.00 | 0.00 | 2.00 | -3.03 | -3.03 | -3.03 | 0.00 | 1 |
| DTNB | 2.57E-03 | 2 | 1 | 0.002409 | Adipose_Visceral_Omentum | 1.96 | 0.04 | 1.96 | 2.94 | 3.03 | 2.99 | 0.07 | 1 |
| CDC42EP5 | 2.58E-03 | 2 | 2 | 0.000591953 | Adipose_Visceral_Omentum | 1.64 | 0.36 | 0.36 | -3.44 | -2.47 | -2.95 | 0.69 | 2 |
| PCMTD1 | 2.58E-03 | 2 | 2 | 0.000580527 | Adipose_Visceral_Omentum | 1.77 | 0.23 | 0.23 | 2.85 | 3.44 | 3.14 | 0.42 | 2 |
| MEIOC | 2.62E-03 | 1 | 1 | 0.002621076 | Adipose_Subcutaneous | 1.00 | 1.00 | 1.00 | -3.01 | -3.01 | -3.01 | NA | 1 |
| KIAA0141 | 2.66E-03 | 2 | 2 | 0.006651541 | Adipose_Visceral_Omentum | 1.54 | 0.46 | 0.46 | -0.34 | 2.71 | 1.19 | 2.16 | 2 |

| | | | | | | | | | | | | | |
|---|---|---|---|---|---|---|---|---|---|---|---|---|---|
| LAMA2 | 2.68E-03 | 1 | 1 | 0.002682999 | Adipose_Subcutaneous | 1.00 | 1.00 | 1.00 | 3.00 | 3.00 | 3.00 | NA | 1 |
| FURIN | 2.69E-03 | 2 | 2 | 0.001752001 | Adipose_Visceral_Omentum | 1.40 | 0.60 | 0.60 | 2.55 | 3.13 | 2.84 | 0.41 | 2 |
| KLHL24 | 2.74E-03 | 2 | 1 | 0.002406001 | Adipose_Visceral_Omentum | 1.99 | 0.01 | 1.99 | -3.03 | -2.94 | -2.99 | 0.06 | 1 |
| MFAP1 | 2.76E-03 | 2 | 1 | 0.002763 | Adipose_Subcutaneous | 2.00 | 0.00 | 2.00 | -2.99 | -2.99 | -2.99 | 0.00 | 1 |
| RP11-290F5.2 | 2.80E-03 | 2 | 2 | 0.0006895 | Adipose_Visceral_Omentum | 1.06 | 0.94 | 0.94 | -3.39 | 0.29 | -1.55 | 2.60 | 2 |
| ARV1 | 2.81E-03 | 2 | 1 | 0.002241346 | Adipose_Visceral_Omentum | 1.95 | 0.05 | 1.95 | 2.84 | 3.06 | 2.95 | 0.15 | 1 |
| TSC2 | 3.04E-03 | 2 | 2 | 0.000747932 | Adipose_Subcutaneous | 1.47 | 0.53 | 0.53 | 2.01 | 3.37 | 2.69 | 0.96 | 2 |
| WDR76 | 3.06E-03 | 2 | 1 | 0.003061 | Adipose_Visceral_Omentum | 2.00 | 0.00 | 2.00 | -2.96 | -2.96 | -2.96 | 0.00 | 1 |
| BACE1 | 3.08E-03 | 2 | 2 | 0.0006764 | Adipose_Subcutaneous | 1.90 | 0.10 | 0.10 | 3.01 | 3.40 | 3.20 | 0.27 | 2 |
| LINC01750 | 3.10E-03 | 2 | 1 | 0.002781138 | Adipose_Subcutaneous | 1.98 | 0.02 | 1.98 | -2.99 | -2.90 | -2.94 | 0.07 | 1 |
| Z83851.4 | 3.12E-03 | 1 | 1 | 0.003121662 | Adipose_Subcutaneous | 1.00 | 1.00 | 1.00 | -2.96 | -2.96 | -2.96 | NA | 1 |
| SYCP3 | 3.14E-03 | 2 | 2 | 0.0007237 | Adipose_Subcutaneous | 1.14 | 0.86 | 0.86 | 0.77 | 3.38 | 2.08 | 1.85 | 2 |
| TIMM10 | 3.16E-03 | 2 | 1 | 0.003479115 | Adipose_Subcutaneous | 1.95 | 0.05 | 1.95 | 2.91 | 2.92 | 2.91 | 0.01 | 1 |
| TOMM34 | 3.17E-03 | 2 | 1 | 0.00280568 | Adipose_Visceral_Omentum | 2.00 | 0.00 | 2.00 | -2.99 | -2.91 | -2.95 | 0.05 | 1 |
| ZNF133 | 3.19E-03 | 1 | 1 | 0.003190004 | Adipose_Subcutaneous | 1.00 | 1.00 | 1.00 | -2.95 | -2.95 | -2.95 | NA | 1 |
| ALDH18A1 | 3.33E-03 | 2 | 2 | 0.0007505 | Adipose_Visceral_Omentum | 1.81 | 0.19 | 0.19 | 2.87 | 3.37 | 3.12 | 0.35 | 2 |
| DICER1 | 3.37E-03 | 1 | 1 | 0.003372999 | Adipose_Subcutaneous | 1.00 | 1.00 | 1.00 | 2.93 | 2.93 | 2.93 | NA | 1 |
| RPL24 | 3.37E-03 | 1 | 1 | 0.003372999 | Adipose_Visceral_Omentum | 1.00 | 1.00 | 1.00 | -2.93 | -2.93 | -2.93 | NA | 1 |
| HAUS4 | 3.38E-03 | 2 | 1 | 0.003325895 | Adipose_Subcutaneous | 2.00 | 0.00 | 2.00 | 2.92 | 2.94 | 2.93 | 0.01 | 1 |
| ALDH2 | 3.42E-03 | 2 | 2 | 0.018540004 | Adipose_Visceral_Omentum | 1.88 | 0.12 | 0.12 | -2.35 | -0.95 | -1.65 | 0.99 | 2 |
| RAB26 | 3.45E-03 | 2 | 2 | 0.001163 | Adipose_Visceral_Omentum | 1.07 | 0.93 | 0.93 | 1.12 | 3.25 | 2.18 | 1.51 | 2 |
| PGBD1 | 3.49E-03 | 2 | 2 | 0.000776154 | Adipose_Subcutaneous | 1.67 | 0.33 | 0.33 | 2.14 | 3.36 | 2.75 | 0.86 | 2 |
| COL11A2 | 3.50E-03 | 2 | 1 | 0.003200925 | Adipose_Subcutaneous | 1.98 | 0.02 | 1.98 | 2.87 | 2.95 | 2.91 | 0.06 | 1 |
| ZDHHC5 | 3.61E-03 | 1 | 1 | 0.003609999 | Adipose_Visceral_Omentum | 1.00 | 1.00 | 1.00 | -2.91 | -2.91 | -2.91 | NA | 1 |
| TSPEAR | 3.67E-03 | 1 | 1 | 0.003671628 | Adipose_Subcutaneous | 1.00 | 1.00 | 1.00 | 2.91 | 2.91 | 2.91 | NA | 1 |
| DKFZp779M0652 | 3.69E-03 | 2 | 1 | 0.003155003 | Adipose_Visceral_Omentum | 1.96 | 0.04 | 1.96 | -2.95 | -2.79 | -2.87 | 0.11 | 1 |
| EPN3 | 3.69E-03 | 2 | 2 | 0.000830744 | Adipose_Visceral_Omentum | 1.82 | 0.18 | 0.18 | 2.85 | 3.34 | 3.09 | 0.35 | 2 |
| GPC6 | 3.73E-03 | 2 | 2 | 0.245131918 | Adipose_Subcutaneous | 1.83 | 0.17 | 0.17 | -1.16 | 0.77 | -0.20 | 1.37 | 2 |
| PPA2 | 3.73E-03 | 2 | 2 | 0.00082727 | Adipose_Subcutaneous | 1.82 | 0.18 | 0.18 | 2.73 | 3.34 | 3.04 | 0.44 | 2 |
| EVI2B | 3.76E-03 | 1 | 1 | 0.003756999 | Adipose_Visceral_Omentum | 1.00 | 1.00 | 1.00 | 2.90 | 2.90 | 2.90 | NA | 1 |
| CBR3 | 3.76E-03 | 2 | 2 | 0.003384156 | Adipose_Subcutaneous | 1.91 | 0.09 | 0.09 | 2.00 | 2.93 | 2.47 | 0.66 | 2 |
| TTC39B | 3.78E-03 | 2 | 2 | 0.109025361 | Adipose_Subcutaneous | 1.78 | 0.22 | 0.22 | -1.60 | 0.58 | -0.51 | 1.54 | 2 |
| NADK | 3.84E-03 | 2 | 2 | 0.000854062 | Adipose_Visceral_Omentum | 1.22 | 0.78 | 0.78 | -3.33 | -0.80 | -2.07 | 1.79 | 2 |

| | | | | | | | | | | | | | |
|---|---|---|---|---|---|---|---|---|---|---|---|---|---|
| PSMG3 | 3.95E-03 | 2 | 2 | 0.012212497 | Adipose_Visceral_Omentum | 1.29 | 0.71 | 0.71 | -2.51 | -1.35 | -1.93 | 0.81 | 2 |
| ODF3B | 3.96E-03 | 2 | 2 | 0.00231994 | Adipose_Subcutaneous | 1.80 | 0.20 | 0.20 | 1.62 | 3.05 | 2.33 | 1.01 | 2 |
| SCGB2A2 | 3.97E-03 | 1 | 1 | 0.003967999 | Adipose_Subcutaneous | 1.00 | 1.00 | 1.00 | 2.88 | 2.88 | 2.88 | NA | 1 |
| KIF3B | 3.97E-03 | 2 | 2 | 0.183099996 | Adipose_Subcutaneous | 1.83 | 0.17 | 0.17 | 0.57 | 1.33 | 0.95 | 0.54 | 2 |
| SUFU | 3.99E-03 | 2 | 1 | 0.003985001 | Adipose_Subcutaneous | 2.00 | 0.00 | 2.00 | 2.88 | 2.88 | 2.88 | 0.00 | 1 |
| FAM180B | 3.99E-03 | 2 | 2 | 0.004007999 | Adipose_Visceral_Omentum | 1.07 | 0.93 | 0.93 | -2.88 | 1.86 | -0.51 | 3.35 | 2 |
| SKAP2 | 3.99E-03 | 1 | 1 | 0.003988 | Adipose_Visceral_Omentum | 1.00 | 1.00 | 1.00 | 2.88 | 2.88 | 2.88 | NA | 1 |
| NDUFAF7 | 3.99E-03 | 2 | 2 | 0.00190537 | Adipose_Subcutaneous | 1.04 | 0.96 | 0.96 | 1.31 | 3.10 | 2.21 | 1.27 | 2 |
| RP11-433J20.1 | 4.04E-03 | 2 | 2 | 0.001421133 | Adipose_Subcutaneous | 1.91 | 0.09 | 0.09 | 2.53 | 3.19 | 2.86 | 0.46 | 2 |
| PCOLCE | 4.05E-03 | 2 | 2 | 0.088339177 | Adipose_Subcutaneous | 1.70 | 0.30 | 0.30 | -0.85 | 1.70 | 0.43 | 1.81 | 2 |
| ZNF536 | 4.05E-03 | 2 | 2 | 0.006717002 | Adipose_Subcutaneous | 1.83 | 0.17 | 0.17 | 1.17 | 2.71 | 1.94 | 1.09 | 2 |
| EEF1E1 | 4.09E-03 | 2 | 1 | 0.004094999 | Adipose_Subcutaneous | 2.00 | 0.00 | 2.00 | 2.87 | 2.87 | 2.87 | 0.00 | 1 |
| RP11-142A22.4 | 4.11E-03 | 1 | 1 | 0.004108999 | Adipose_Visceral_Omentum | 1.00 | 1.00 | 1.00 | 2.87 | 2.87 | 2.87 | NA | 1 |
| GRB10 | 4.14E-03 | 2 | 1 | 0.003187999 | Adipose_Visceral_Omentum | 1.96 | 0.04 | 1.96 | 2.73 | 2.95 | 2.84 | 0.15 | 1 |
| FBXO18 | 4.15E-03 | 2 | 2 | 0.000979568 | Adipose_Subcutaneous | 1.89 | 0.11 | 0.11 | -3.30 | -3.08 | -3.19 | 0.15 | 2 |
| COL1A2 | 4.16E-03 | 2 | 1 | 0.004157998 | Adipose_Subcutaneous | 2.00 | 0.00 | 2.00 | 2.87 | 2.87 | 2.87 | 0.00 | 1 |
| CTDNEP1 | 4.18E-03 | 2 | 1 | 0.003537 | Adipose_Visceral_Omentum | 1.99 | 0.01 | 1.99 | 2.80 | 2.92 | 2.86 | 0.08 | 1 |
| RP3-473L9.4 | 4.30E-03 | 2 | 2 | 0.003179081 | Adipose_Subcutaneous | 1.04 | 0.96 | 0.96 | -1.35 | 2.95 | 0.80 | 3.04 | 2 |
| MYO19 | 4.30E-03 | 2 | 2 | 0.001283151 | Adipose_Subcutaneous | 1.93 | 0.07 | 0.07 | 2.71 | 3.22 | 2.96 | 0.36 | 2 |
| CYP26C1 | 4.36E-03 | 2 | 2 | 0.065760007 | Adipose_Visceral_Omentum | 1.82 | 0.18 | 0.18 | -1.84 | 0.03 | -0.90 | 1.33 | 2 |
| EVI2A | 4.36E-03 | 2 | 1 | 0.004362999 | Adipose_Visceral_Omentum | 2.00 | 0.00 | 2.00 | -2.85 | -2.85 | -2.85 | 0.00 | 1 |
| NCLN | 4.40E-03 | 2 | 1 | 0.002839859 | Adipose_Visceral_Omentum | 1.95 | 0.05 | 1.95 | 2.64 | 2.98 | 2.81 | 0.24 | 1 |
| TRPV1 | 4.40E-03 | 2 | 1 | 0.004037 | Adipose_Visceral_Omentum | 1.98 | 0.02 | 1.98 | -2.88 | -2.80 | -2.84 | 0.06 | 1 |
| FBN1 | 4.43E-03 | 1 | 1 | 0.004429999 | Adipose_Subcutaneous | 1.00 | 1.00 | 1.00 | -2.85 | -2.85 | -2.85 | NA | 1 |
| IFNE | 4.44E-03 | 1 | 1 | 0.004444999 | Adipose_Visceral_Omentum | 1.00 | 1.00 | 1.00 | -2.84 | -2.84 | -2.84 | NA | 1 |
| SNX27 | 4.47E-03 | 1 | 1 | 0.004471001 | Adipose_Subcutaneous | 1.00 | 1.00 | 1.00 | -2.84 | -2.84 | -2.84 | NA | 1 |
| KLF9 | 4.73E-03 | 1 | 1 | 0.004730999 | Adipose_Subcutaneous | 1.00 | 1.00 | 1.00 | -2.82 | -2.82 | -2.82 | NA | 1 |
| PIK3R5 | 4.73E-03 | 2 | 1 | 0.004730999 | Adipose_Visceral_Omentum | 2.00 | 0.00 | 2.00 | 2.82 | 2.82 | 2.82 | 0.00 | 1 |
| NDUFS1 | 4.76E-03 | 2 | 1 | 0.004748784 | Adipose_Visceral_Omentum | 2.00 | 0.00 | 2.00 | 2.82 | 2.82 | 2.82 | 0.00 | 1 |
| RGS16 | 4.77E-03 | 2 | 2 | 0.002713 | Adipose_Visceral_Omentum | 1.40 | 0.60 | 0.60 | -3.00 | -2.39 | -2.69 | 0.43 | 2 |
| PROCR | 4.79E-03 | 2 | 2 | 0.001842001 | Adipose_Visceral_Omentum | 1.18 | 0.82 | 0.82 | 0.42 | 3.11 | 1.77 | 1.91 | 2 |
| TPK1 | 4.93E-03 | 2 | 2 | 0.002901985 | Adipose_Visceral_Omentum | 1.89 | 0.11 | 0.11 | -2.98 | -2.05 | -2.51 | 0.66 | 2 |
| PTCH1 | 5.00E-03 | 2 | 1 | 0.005004002 | Adipose_Subcutaneous | 2.00 | 0.00 | 2.00 | -2.81 | -2.81 | -2.81 | 0.00 | 1 |

| | | | | | | | | | | | | | |
|---|---|---|---|---|---|---|---|---|---|---|---|---|---|
| C18orf21 | 5.06E-03 | 2 | 1 | 0.005064 | Adipose_Subcutaneous | 2.00 | 0.00 | 2.00 | -2.80 | -2.80 | -2.80 | 0.00 | 1 |
| FLOT1 | 5.07E-03 | 2 | 2 | 0.001703343 | Adipose_Visceral_Omentum | 1.75 | 0.25 | 0.25 | 1.78 | 3.14 | 2.46 | 0.96 | 2 |
| ARF6 | 5.09E-03 | 2 | 1 | 0.005093001 | Adipose_Subcutaneous | 2.00 | 0.00 | 2.00 | 2.80 | 2.80 | 2.80 | 0.00 | 1 |
| BICDL1 | 5.10E-03 | 1 | 1 | 0.005096001 | Adipose_Subcutaneous | 1.00 | 1.00 | 1.00 | -2.80 | -2.80 | -2.80 | NA | 1 |
| TAPBP | 5.10E-03 | 2 | 1 | 0.005100999 | Adipose_Visceral_Omentum | 2.00 | 0.00 | 2.00 | 2.80 | 2.80 | 2.80 | 0.00 | 1 |
| CMSS1 | 5.11E-03 | 2 | 2 | 0.005036366 | Adipose_Subcutaneous | 1.77 | 0.23 | 0.23 | 1.11 | 2.80 | 1.96 | 1.20 | 2 |
| ZSCAN16 | 5.15E-03 | 2 | 2 | 0.00159356 | Adipose_Subcutaneous | 1.13 | 0.87 | 0.87 | -3.16 | 0.32 | -1.42 | 2.46 | 2 |
| RP11-47A8.5 | 5.16E-03 | 2 | 1 | 0.004939 | Adipose_Visceral_Omentum | 2.00 | 0.00 | 2.00 | -2.81 | -2.78 | -2.80 | 0.02 | 1 |
| LINC01094 | 5.17E-03 | 1 | 1 | 0.005165 | Adipose_Visceral_Omentum | 1.00 | 1.00 | 1.00 | -2.80 | -2.80 | -2.80 | NA | 1 |
| AP4M1 | 5.26E-03 | 2 | 1 | 0.005107239 | Adipose_Subcutaneous | 2.00 | 0.00 | 2.00 | 2.78 | 2.80 | 2.79 | 0.01 | 1 |
| CHCHD5 | 5.28E-03 | 2 | 1 | 0.005256355 | Adipose_Visceral_Omentum | 2.00 | 0.00 | 2.00 | -2.79 | -2.79 | -2.79 | 0.00 | 1 |
| ZSWIM8 | 5.29E-03 | 2 | 1 | 0.005173464 | Adipose_Subcutaneous | 2.00 | 0.00 | 2.00 | -2.80 | -2.78 | -2.79 | 0.01 | 1 |
| ZSWIM1 | 5.31E-03 | 2 | 2 | 0.001378388 | Adipose_Visceral_Omentum | 1.29 | 0.71 | 0.71 | 0.46 | 3.20 | 1.83 | 1.94 | 2 |
| C6orf226 | 5.38E-03 | 2 | 1 | 0.005364458 | Adipose_Subcutaneous | 2.00 | 0.00 | 2.00 | 2.78 | 2.78 | 2.78 | 0.00 | 1 |
| ARID3B | 5.40E-03 | 1 | 1 | 0.005401999 | Adipose_Subcutaneous | 1.00 | 1.00 | 1.00 | 2.78 | 2.78 | 2.78 | NA | 1 |
| TRIM8 | 5.43E-03 | 2 | 1 | 0.005425001 | Adipose_Subcutaneous | 2.00 | 0.00 | 2.00 | -2.78 | -2.78 | -2.78 | 0.00 | 1 |
| GINM1 | 5.45E-03 | 1 | 1 | 0.005446 | Adipose_Subcutaneous | 1.00 | 1.00 | 1.00 | 2.78 | 2.78 | 2.78 | NA | 1 |
| CDKN2AIP | 5.46E-03 | 2 | 2 | 0.001413 | Adipose_Visceral_Omentum | 1.56 | 0.44 | 0.44 | 1.39 | 3.19 | 2.29 | 1.28 | 2 |
| ERCC6L2 | 5.50E-03 | 2 | 2 | 0.146421787 | Adipose_Visceral_Omentum | 1.90 | 0.10 | 0.10 | 0.02 | 1.45 | 0.74 | 1.01 | 2 |
| RSAD1 | 5.51E-03 | 2 | 2 | 0.001527 | Adipose_Subcutaneous | 1.51 | 0.49 | 0.49 | 1.10 | 3.17 | 2.13 | 1.46 | 2 |
| CHPF | 5.52E-03 | 2 | 2 | 0.021633372 | Adipose_Visceral_Omentum | 1.88 | 0.12 | 0.12 | 0.96 | 2.30 | 1.63 | 0.94 | 2 |
| SOX6 | 5.52E-03 | 2 | 1 | 0.005326001 | Adipose_Subcutaneous | 1.99 | 0.01 | 1.99 | -2.79 | -2.75 | -2.77 | 0.02 | 1 |
| FLII | 5.52E-03 | 2 | 2 | 0.032939993 | Adipose_Visceral_Omentum | 1.82 | 0.18 | 0.18 | -2.13 | -0.35 | -1.24 | 1.26 | 2 |
| PPP1R15B | 5.53E-03 | 1 | 1 | 0.005530686 | Adipose_Subcutaneous | 1.00 | 1.00 | 1.00 | -2.77 | -2.77 | -2.77 | NA | 1 |
| XXbac-BPG13B8.10 | 5.56E-03 | 2 | 2 | 0.001687342 | Adipose_Visceral_Omentum | 1.28 | 0.72 | 0.72 | 1.58 | 3.14 | 2.36 | 1.11 | 2 |
| WASL | 5.57E-03 | 2 | 1 | 0.005569001 | Adipose_Visceral_Omentum | 2.00 | 0.00 | 2.00 | 2.77 | 2.77 | 2.77 | 0.00 | 1 |
| LINC00377 | 5.64E-03 | 2 | 2 | 0.015849998 | Adipose_Subcutaneous | 1.92 | 0.08 | 0.08 | -2.41 | -1.41 | -1.91 | 0.71 | 2 |
| RP11-554D14.7 | 5.68E-03 | 2 | 2 | 0.00133 | Adipose_Subcutaneous | 1.08 | 0.92 | 0.92 | -0.45 | 3.21 | 1.38 | 2.59 | 2 |
| HSPA2 | 5.84E-03 | 1 | 1 | 0.005836001 | Adipose_Subcutaneous | 1.00 | 1.00 | 1.00 | -2.76 | -2.76 | -2.76 | NA | 1 |
| EML4 | 5.85E-03 | 2 | 1 | 0.005757117 | Adipose_Subcutaneous | 2.00 | 0.00 | 2.00 | -2.76 | -2.75 | -2.76 | 0.01 | 1 |
| PIP4K2C | 5.90E-03 | 2 | 2 | 0.004078999 | Adipose_Visceral_Omentum | 1.14 | 0.86 | 0.86 | -2.87 | -1.01 | -1.94 | 1.31 | 2 |
| FAM117A | 5.92E-03 | 1 | 1 | 0.005918 | Adipose_Subcutaneous | 1.00 | 1.00 | 1.00 | 2.75 | 2.75 | 2.75 | NA | 1 |
| HIGD1B | 5.94E-03 | 1 | 1 | 0.005944999 | Adipose_Visceral_Omentum | 1.00 | 1.00 | 1.00 | -2.75 | -2.75 | -2.75 | NA | 1 |

| | | | | | | | | | | | | | |
|---|---|---|---|---|---|---|---|---|---|---|---|---|---|
| POU6F2 | 5.96E-03 | 2 | 1 | 0.005872073 | Adipose_Subcutaneous | 2.00 | 0.00 | 2.00 | 2.75 | 2.75 | 2.75 | 0.01 | 1 |
| PCDH9 | 5.98E-03 | 2 | 1 | 0.005976998 | Adipose_Subcutaneous | 2.00 | 0.00 | 2.00 | -2.75 | -2.75 | -2.75 | 0.00 | 1 |
| CTNNAL1 | 6.05E-03 | 2 | 1 | 0.003887743 | Adipose_Subcutaneous | 1.98 | 0.02 | 1.98 | -2.89 | -2.58 | -2.73 | 0.22 | 1 |
| MBOAT4 | 6.10E-03 | 2 | 1 | 0.004707665 | Adipose_Subcutaneous | 1.99 | 0.01 | 1.99 | -2.83 | -2.64 | -2.73 | 0.13 | 1 |
| CSRNP3 | 6.13E-03 | 2 | 2 | 0.003385001 | Adipose_Subcutaneous | 1.25 | 0.75 | 0.75 | -2.93 | -1.97 | -2.45 | 0.68 | 2 |
| TAOK1 | 6.13E-03 | 2 | 2 | 0.001468 | Adipose_Visceral_Omentum | 1.90 | 0.10 | 0.10 | 2.74 | 3.18 | 2.96 | 0.31 | 2 |
| ZBTB1 | 6.14E-03 | 2 | 2 | 0.001770429 | Adipose_Visceral_Omentum | 1.85 | 0.15 | 0.15 | 2.99 | 3.13 | 3.06 | 0.09 | 2 |
| RP11-774O3.3 | 6.15E-03 | 2 | 2 | 0.004258097 | Adipose_Subcutaneous | 1.85 | 0.15 | 0.15 | 1.68 | 2.86 | 2.27 | 0.83 | 2 |
| RHCG | 6.26E-03 | 1 | 1 | 0.006258002 | Adipose_Visceral_Omentum | 1.00 | 1.00 | 1.00 | 2.73 | 2.73 | 2.73 | NA | 1 |
| TAF6 | 6.30E-03 | 2 | 2 | 0.004590002 | Adipose_Visceral_Omentum | 1.90 | 0.10 | 0.10 | 1.95 | 2.83 | 2.39 | 0.63 | 2 |
| CHMP4B | 6.32E-03 | 2 | 1 | 0.006320002 | Adipose_Subcutaneous | 2.00 | 0.00 | 2.00 | 2.73 | 2.73 | 2.73 | 0.00 | 1 |
| OS9 | 6.39E-03 | 1 | 1 | 0.006387998 | Adipose_Visceral_Omentum | 1.00 | 1.00 | 1.00 | -2.73 | -2.73 | -2.73 | NA | 1 |
| PIK3R3 | 6.42E-03 | 2 | 2 | 0.013239097 | Adipose_Subcutaneous | 1.68 | 0.32 | 0.32 | 0.21 | 2.48 | 1.34 | 1.61 | 2 |
| SLC37A2 | 6.46E-03 | 2 | 1 | 0.005004002 | Adipose_Visceral_Omentum | 1.99 | 0.01 | 1.99 | 2.63 | 2.81 | 2.72 | 0.13 | 1 |
| SEL1L | 6.46E-03 | 1 | 1 | 0.006459002 | Adipose_Subcutaneous | 1.00 | 1.00 | 1.00 | -2.72 | -2.72 | -2.72 | NA | 1 |
| RGMB | 6.46E-03 | 2 | 1 | 0.006462 | Adipose_Subcutaneous | 2.00 | 0.00 | 2.00 | 2.72 | 2.72 | 2.72 | 0.00 | 1 |
| CCL23 | 6.56E-03 | 1 | 1 | 0.006563002 | Adipose_Visceral_Omentum | 1.00 | 1.00 | 1.00 | 2.72 | 2.72 | 2.72 | NA | 1 |
| TTC7B | 6.57E-03 | 2 | 2 | 0.02048667 | Adipose_Subcutaneous | 1.68 | 0.32 | 0.32 | -2.32 | 0.01 | -1.15 | 1.64 | 2 |
| SRP9 | 6.62E-03 | 2 | 2 | 0.014270001 | Adipose_Subcutaneous | 1.79 | 0.21 | 0.21 | -2.45 | -0.70 | -1.58 | 1.24 | 2 |
| CMTM1 | 6.65E-03 | 2 | 1 | 0.006564998 | Adipose_Visceral_Omentum | 2.00 | 0.00 | 2.00 | -2.72 | -2.71 | -2.71 | 0.01 | 1 |
| IFFO1 | 6.67E-03 | 2 | 2 | 0.237427636 | Adipose_Subcutaneous | 1.80 | 0.20 | 0.20 | -0.80 | 1.18 | 0.19 | 1.40 | 2 |
| DCUN1D2 | 6.69E-03 | 1 | 1 | 0.006690998 | Adipose_Visceral_Omentum | 1.00 | 1.00 | 1.00 | -2.71 | -2.71 | -2.71 | NA | 1 |
| COMMD2 | 6.77E-03 | 2 | 1 | 0.004897001 | Adipose_Subcutaneous | 1.99 | 0.01 | 1.99 | 2.58 | 2.81 | 2.70 | 0.16 | 1 |
| UBE2E1 | 6.77E-03 | 2 | 1 | 0.006706621 | Adipose_Visceral_Omentum | 1.98 | 0.02 | 1.98 | -2.71 | -2.68 | -2.69 | 0.02 | 1 |
| C22orf31 | 6.77E-03 | 1 | 1 | 0.006771999 | Adipose_Visceral_Omentum | 1.00 | 1.00 | 1.00 | -2.71 | -2.71 | -2.71 | NA | 1 |
| RPS15A | 6.82E-03 | 2 | 2 | 0.02582837 | Adipose_Subcutaneous | 1.12 | 0.88 | 0.88 | -1.97 | 2.23 | 0.13 | 2.97 | 2 |
| CYB5R4 | 6.86E-03 | 2 | 1 | 0.006862998 | Adipose_Subcutaneous | 2.00 | 0.00 | 2.00 | 2.70 | 2.70 | 2.70 | 0.00 | 1 |
| KSR1 | 6.90E-03 | 2 | 2 | 0.008908001 | Adipose_Subcutaneous | 1.23 | 0.77 | 0.77 | -2.62 | 1.12 | -0.75 | 2.64 | 2 |
| FADS1 | 6.91E-03 | 1 | 1 | 0.006906001 | Adipose_Visceral_Omentum | 1.00 | 1.00 | 1.00 | -2.70 | -2.70 | -2.70 | NA | 1 |
| SEMA7A | 6.91E-03 | 2 | 1 | 0.006910999 | Adipose_Subcutaneous | 2.00 | 0.00 | 2.00 | 2.70 | 2.70 | 2.70 | 0.00 | 1 |
| MAP2K3 | 6.95E-03 | 1 | 1 | 0.006952001 | Adipose_Visceral_Omentum | 1.00 | 1.00 | 1.00 | -2.70 | -2.70 | -2.70 | NA | 1 |
| RNF222 | 6.96E-03 | 1 | 1 | 0.006956001 | Adipose_Subcutaneous | 1.00 | 1.00 | 1.00 | 2.70 | 2.70 | 2.70 | NA | 1 |
| CTD-2298J14.2 | 6.98E-03 | 1 | 1 | 0.006983076 | Adipose_Visceral_Omentum | 1.00 | 1.00 | 1.00 | 2.70 | 2.70 | 2.70 | NA | 1 |

| | | | | | | | | | | | | | |
|---|---|---|---|---|---|---|---|---|---|---|---|---|---|
| C1orf54 | 7.00E-03 | 2 | 1 | 0.007003999 | Adipose_Visceral_Omentum | 2.00 | 0.00 | 2.00 | -2.70 | -2.70 | -2.70 | 0.00 | 1 |
| ZNF302 | 7.01E-03 | 2 | 1 | 0.00709032 | Adipose_Visceral_Omentum | 1.97 | 0.03 | 1.97 | -2.69 | -2.66 | -2.68 | 0.02 | 1 |
| NEGR1 | 7.01E-03 | 1 | 1 | 0.007014193 | Adipose_Visceral_Omentum | 1.00 | 1.00 | 1.00 | 2.70 | 2.70 | 2.70 | NA | 1 |
| LINC01021 | 7.03E-03 | 1 | 1 | 0.00703 | Adipose_Subcutaneous | 1.00 | 1.00 | 1.00 | 2.70 | 2.70 | 2.70 | NA | 1 |
| U2AF1L4 | 7.09E-03 | 2 | 1 | 0.006374908 | Adipose_Visceral_Omentum | 2.00 | 0.00 | 2.00 | 2.66 | 2.73 | 2.69 | 0.05 | 1 |
| C4B | 7.10E-03 | 2 | 2 | 0.003910107 | Adipose_Subcutaneous | 1.73 | 0.27 | 0.27 | 1.24 | 2.89 | 2.06 | 1.16 | 2 |
| RER1 | 7.12E-03 | 1 | 1 | 0.00711561 | Adipose_Subcutaneous | 1.00 | 1.00 | 1.00 | 2.69 | 2.69 | 2.69 | NA | 1 |
| ZDBF2 | 7.12E-03 | 2 | 2 | 0.002205 | Adipose_Visceral_Omentum | 1.62 | 0.38 | 0.38 | -3.06 | -2.46 | -2.76 | 0.43 | 2 |
| CIB4 | 7.13E-03 | 2 | 1 | 0.007131012 | Adipose_Subcutaneous | 2.00 | 0.00 | 2.00 | -2.69 | -2.69 | -2.69 | 0.00 | 1 |
| LIN52 | 7.14E-03 | 2 | 1 | 0.006476338 | Adipose_Visceral_Omentum | 1.99 | 0.01 | 1.99 | -2.72 | -2.65 | -2.69 | 0.05 | 1 |
| CSNK1E | 7.20E-03 | 1 | 1 | 0.007202 | Adipose_Subcutaneous | 1.00 | 1.00 | 1.00 | 2.69 | 2.69 | 2.69 | NA | 1 |
| EEF1B2 | 7.22E-03 | 1 | 1 | 0.007219999 | Adipose_Visceral_Omentum | 1.00 | 1.00 | 1.00 | 2.69 | 2.69 | 2.69 | NA | 1 |
| LINC01655 | 7.25E-03 | 1 | 1 | 0.007253003 | Adipose_Subcutaneous | 1.00 | 1.00 | 1.00 | 2.68 | 2.68 | 2.68 | NA | 1 |
| JMJD8 | 7.28E-03 | 2 | 1 | 0.007282999 | Adipose_Subcutaneous | 2.00 | 0.00 | 2.00 | -2.68 | -2.68 | -2.68 | 0.00 | 1 |
| CASC20 | 7.30E-03 | 1 | 1 | 0.007295998 | Adipose_Visceral_Omentum | 1.00 | 1.00 | 1.00 | 2.68 | 2.68 | 2.68 | NA | 1 |
| TESK2 | 7.34E-03 | 2 | 1 | 0.004165403 | Adipose_Subcutaneous | 1.97 | 0.03 | 1.97 | 2.45 | 2.87 | 2.66 | 0.29 | 1 |
| CDCA2 | 7.35E-03 | 2 | 1 | 0.007317002 | Adipose_Subcutaneous | 2.00 | 0.00 | 2.00 | 2.68 | 2.68 | 2.68 | 0.00 | 1 |
| KCTD9 | 7.37E-03 | 1 | 1 | 0.007374 | Adipose_Subcutaneous | 1.00 | 1.00 | 1.00 | 2.68 | 2.68 | 2.68 | NA | 1 |
| PRPF31 | 7.47E-03 | 2 | 2 | 0.04543 | Adipose_Subcutaneous | 1.63 | 0.37 | 0.37 | -2.00 | 0.60 | -0.70 | 1.84 | 2 |
| CEBPZOS | 7.49E-03 | 2 | 2 | 0.001829 | Adipose_Visceral_Omentum | 1.45 | 0.55 | 0.55 | -3.12 | -1.66 | -2.39 | 1.03 | 2 |
| LIN7C | 7.50E-03 | 2 | 2 | 0.014967448 | Adipose_Subcutaneous | 1.93 | 0.07 | 0.07 | 1.52 | 2.43 | 1.98 | 0.64 | 2 |
| SMIM15 | 7.55E-03 | 1 | 1 | 0.007554001 | Adipose_Subcutaneous | 1.00 | 1.00 | 1.00 | -2.67 | -2.67 | -2.67 | NA | 1 |
| GRIK5 | 7.60E-03 | 1 | 1 | 0.007595999 | Adipose_Visceral_Omentum | 1.00 | 1.00 | 1.00 | 2.67 | 2.67 | 2.67 | NA | 1 |
| MIER2 | 7.65E-03 | 2 | 2 | 0.002086 | Adipose_Visceral_Omentum | 1.20 | 0.80 | 0.80 | -3.08 | -1.13 | -2.10 | 1.38 | 2 |
| PRRC2A | 7.71E-03 | 1 | 1 | 0.007708002 | Adipose_Subcutaneous | 1.00 | 1.00 | 1.00 | -2.66 | -2.66 | -2.66 | NA | 1 |
| TMEM161B | 7.71E-03 | 1 | 1 | 0.007712489 | Adipose_Subcutaneous | 1.00 | 1.00 | 1.00 | -2.66 | -2.66 | -2.66 | NA | 1 |
| SLC2A4 | 7.73E-03 | 2 | 2 | 0.018123295 | Adipose_Visceral_Omentum | 1.93 | 0.07 | 0.07 | 1.46 | 2.36 | 1.91 | 0.64 | 2 |
| LINC01124 | 7.78E-03 | 1 | 1 | 0.007778999 | Adipose_Subcutaneous | 1.00 | 1.00 | 1.00 | 2.66 | 2.66 | 2.66 | NA | 1 |
| RP11-147L13.14 | 7.86E-03 | 2 | 2 | 0.005239999 | Adipose_Subcutaneous | 1.87 | 0.13 | 0.13 | 1.74 | 2.79 | 2.27 | 0.74 | 2 |
| ZNF184 | 7.86E-03 | 2 | 1 | 0.007862998 | Adipose_Visceral_Omentum | 2.00 | 0.00 | 2.00 | -2.66 | -2.66 | -2.66 | 0.00 | 1 |
| STAG3 | 7.90E-03 | 2 | 2 | 0.005942001 | Adipose_Visceral_Omentum | 1.26 | 0.74 | 0.74 | -2.75 | -2.12 | -2.43 | 0.45 | 2 |
| FADS2 | 7.91E-03 | 2 | 1 | 0.007914002 | Adipose_Subcutaneous | 2.00 | 0.00 | 2.00 | -2.66 | -2.66 | -2.66 | 0.00 | 1 |
| NDUFV3 | 7.93E-03 | 2 | 1 | 0.006012179 | Adipose_Subcutaneous | 2.00 | 0.00 | 2.00 | 2.56 | 2.75 | 2.65 | 0.13 | 1 |

| | | | | | | | | | | | | | |
|---|---|---|---|---|---|---|---|---|---|---|---|---|---|
| GMDS | 7.97E-03 | 2 | 2 | 0.001983001 | Adipose_Visceral_Omentum | 1.21 | 0.79 | 0.79 | -3.09 | -0.94 | -2.02 | 1.52 | 2 |
| 6-Mar | 8.02E-03 | 2 | 2 | 0.096630011 | Adipose_Subcutaneous | 1.81 | 0.19 | 0.19 | -1.66 | 0.22 | -0.72 | 1.33 | 2 |
| MGA | 8.03E-03 | 2 | 2 | 0.002158 | Adipose_Subcutaneous | 1.50 | 0.50 | 0.50 | -3.07 | -1.95 | -2.51 | 0.79 | 2 |
| THOC5 | 8.03E-03 | 2 | 2 | 0.001921641 | Adipose_Subcutaneous | 1.84 | 0.16 | 0.16 | -3.10 | -2.53 | -2.82 | 0.40 | 2 |
| OR10Q1 | 8.05E-03 | 2 | 2 | 0.001911999 | Adipose_Subcutaneous | 1.37 | 0.63 | 0.63 | -3.10 | 1.25 | -0.92 | 3.08 | 2 |
| FAM208B | 8.09E-03 | 2 | 2 | 0.002130213 | Adipose_Subcutaneous | 1.37 | 0.63 | 0.63 | 0.72 | 3.07 | 1.90 | 1.66 | 2 |
| LYL1 | 8.10E-03 | 1 | 1 | 0.008095999 | Adipose_Subcutaneous | 1.00 | 1.00 | 1.00 | -2.65 | -2.65 | -2.65 | NA | 1 |
| RBM5 | 8.13E-03 | 1 | 1 | 0.008130711 | Adipose_Visceral_Omentum | 1.00 | 1.00 | 1.00 | -2.65 | -2.65 | -2.65 | NA | 1 |
| GNG12 | 8.17E-03 | 2 | 2 | 0.002298859 | Adipose_Visceral_Omentum | 1.85 | 0.15 | 0.15 | -3.05 | -2.90 | -2.97 | 0.11 | 2 |
| BHLHE22 | 8.29E-03 | 2 | 2 | 0.001962001 | Adipose_Visceral_Omentum | 1.73 | 0.27 | 0.27 | -2.27 | 3.10 | 0.41 | 3.80 | 2 |
| UHRF1BP1L | 8.30E-03 | 2 | 1 | 0.007462357 | Adipose_Visceral_Omentum | 1.97 | 0.03 | 1.97 | -2.68 | -2.57 | -2.62 | 0.08 | 1 |
| DR1 | 8.32E-03 | 2 | 1 | 0.006610594 | Adipose_Subcutaneous | 1.95 | 0.05 | 1.95 | -2.72 | -2.50 | -2.61 | 0.15 | 1 |
| AKT3 | 8.34E-03 | 2 | 1 | 0.008337002 | Adipose_Subcutaneous | 2.00 | 0.00 | 2.00 | 2.64 | 2.64 | 2.64 | 0.00 | 1 |
| APOLD1 | 8.35E-03 | 1 | 1 | 0.008353999 | Adipose_Visceral_Omentum | 1.00 | 1.00 | 1.00 | -2.64 | -2.64 | -2.64 | NA | 1 |
| AC012442.6 | 8.38E-03 | 2 | 1 | 0.008380002 | Adipose_Subcutaneous | 2.00 | 0.00 | 2.00 | -2.64 | -2.64 | -2.64 | 0.00 | 1 |
| INTS8 | 8.40E-03 | 2 | 2 | 0.002490046 | Adipose_Subcutaneous | 1.79 | 0.21 | 0.21 | -3.02 | -2.77 | -2.90 | 0.18 | 2 |
| MNT | 8.40E-03 | 2 | 2 | 0.011859998 | Adipose_Subcutaneous | 1.15 | 0.85 | 0.85 | -1.41 | 2.52 | 0.55 | 2.78 | 2 |
| XXbac-B135H6.18 | 8.42E-03 | 2 | 2 | 0.01062 | Adipose_Subcutaneous | 1.08 | 0.92 | 0.92 | -2.55 | -1.95 | -2.25 | 0.43 | 2 |
| RP11-345F18.1 | 8.43E-03 | 1 | 1 | 0.008431844 | Adipose_Visceral_Omentum | 1.00 | 1.00 | 1.00 | 2.63 | 2.63 | 2.63 | NA | 1 |
| TMEM245 | 8.43E-03 | 2 | 2 | 0.089163409 | Adipose_Subcutaneous | 1.82 | 0.18 | 0.18 | -1.70 | 0.08 | -0.81 | 1.26 | 2 |
| LINC00895 | 8.44E-03 | 2 | 1 | 0.008444 | Adipose_Subcutaneous | 2.00 | 0.00 | 2.00 | 2.63 | 2.63 | 2.63 | 0.00 | 1 |
| SPTBN2 | 8.46E-03 | 2 | 1 | 0.008456999 | Adipose_Subcutaneous | 2.00 | 0.00 | 2.00 | -2.63 | -2.63 | -2.63 | 0.00 | 1 |
| POLG | 8.46E-03 | 2 | 1 | 0.007444002 | Adipose_Subcutaneous | 1.99 | 0.01 | 1.99 | -2.68 | -2.57 | -2.62 | 0.07 | 1 |
| ZNF304 | 8.51E-03 | 2 | 1 | 0.006176409 | Adipose_Visceral_Omentum | 1.94 | 0.06 | 1.94 | 2.44 | 2.74 | 2.59 | 0.21 | 1 |
| MARK1 | 8.52E-03 | 1 | 1 | 0.008519998 | Adipose_Visceral_Omentum | 1.00 | 1.00 | 1.00 | -2.63 | -2.63 | -2.63 | NA | 1 |
| PENK | 8.53E-03 | 2 | 1 | 0.008526001 | Adipose_Subcutaneous | 2.00 | 0.00 | 2.00 | 2.63 | 2.63 | 2.63 | 0.00 | 1 |
| GATS | 8.54E-03 | 2 | 2 | 0.005942001 | Adipose_Subcutaneous | 1.27 | 0.73 | 0.73 | 2.08 | 2.75 | 2.42 | 0.47 | 2 |
| ZCCHC2 | 8.60E-03 | 2 | 2 | 0.004453 | Adipose_Visceral_Omentum | 1.08 | 0.92 | 0.92 | -0.97 | 2.84 | 0.94 | 2.70 | 2 |
| ZNF200 | 8.61E-03 | 2 | 2 | 0.052258501 | Adipose_Visceral_Omentum | 1.86 | 0.14 | 0.14 | 0.47 | 1.94 | 1.21 | 1.04 | 2 |
| SFT2D2 | 8.64E-03 | 1 | 1 | 0.008635999 | Adipose_Subcutaneous | 1.00 | 1.00 | 1.00 | -2.63 | -2.63 | -2.63 | NA | 1 |
| CELF1 | 8.65E-03 | 2 | 2 | 0.002955999 | Adipose_Visceral_Omentum | 1.10 | 0.90 | 0.90 | -1.12 | 2.97 | 0.92 | 2.90 | 2 |
| MAP3K14 | 8.68E-03 | 1 | 1 | 0.008683997 | Adipose_Subcutaneous | 1.00 | 1.00 | 1.00 | -2.62 | -2.62 | -2.62 | NA | 1 |
| IFT74 | 8.74E-03 | 2 | 1 | 0.008515187 | Adipose_Subcutaneous | 1.99 | 0.01 | 1.99 | -2.63 | -2.60 | -2.61 | 0.02 | 1 |

| | | | | | | | | | | | | | |
|---|---|---|---|---|---|---|---|---|---|---|---|---|---|
| FES | 8.76E-03 | 2 | 2 | 0.142594456 | Adipose_Subcutaneous | 1.78 | 0.22 | 0.22 | -1.47 | 0.57 | -0.45 | 1.44 | 2 |
| PRRC2C | 8.80E-03 | 2 | 2 | 0.002398066 | Adipose_Visceral_Omentum | 1.93 | 0.07 | 0.07 | 2.62 | 3.04 | 2.83 | 0.29 | 2 |
| PABPC1 | 8.82E-03 | 2 | 1 | 0.008824998 | Adipose_Subcutaneous | 2.00 | 0.00 | 2.00 | 2.62 | 2.62 | 2.62 | 0.00 | 1 |
| HS2ST1 | 8.86E-03 | 2 | 1 | 0.007220998 | Adipose_Visceral_Omentum | 1.94 | 0.06 | 1.94 | -2.69 | -2.48 | -2.58 | 0.15 | 1 |
| CRMP1 | 8.86E-03 | 2 | 2 | 0.027110008 | Adipose_Visceral_Omentum | 1.30 | 0.70 | 0.70 | -2.21 | 1.37 | -0.42 | 2.53 | 2 |
| ID4 | 8.89E-03 | 1 | 1 | 0.008893999 | Adipose_Subcutaneous | 1.00 | 1.00 | 1.00 | 2.62 | 2.62 | 2.62 | NA | 1 |
| ZC3H6 | 8.89E-03 | 2 | 2 | 0.002169001 | Adipose_Subcutaneous | 1.77 | 0.23 | 0.23 | 2.23 | 3.07 | 2.65 | 0.59 | 2 |
| PRMT6 | 8.94E-03 | 2 | 1 | 0.008939002 | Adipose_Visceral_Omentum | 2.00 | 0.00 | 2.00 | -2.61 | -2.61 | -2.61 | 0.00 | 1 |
| BUD31 | 8.98E-03 | 2 | 2 | 0.101599993 | Adipose_Visceral_Omentum | 1.74 | 0.26 | 0.26 | -1.64 | 0.52 | -0.56 | 1.53 | 2 |
| ME2 | 9.13E-03 | 2 | 2 | 0.003442393 | Adipose_Visceral_Omentum | 1.85 | 0.15 | 0.15 | -2.93 | -2.00 | -2.46 | 0.66 | 2 |
| KIF7 | 9.16E-03 | 1 | 1 | 0.009158002 | Adipose_Subcutaneous | 1.00 | 1.00 | 1.00 | 2.61 | 2.61 | 2.61 | NA | 1 |
| RORB | 9.16E-03 | 2 | 2 | 0.003118221 | Adipose_Visceral_Omentum | 1.37 | 0.63 | 0.63 | -1.85 | 2.96 | 0.55 | 3.40 | 2 |
| RDH10 | 9.18E-03 | 1 | 1 | 0.009178 | Adipose_Visceral_Omentum | 1.00 | 1.00 | 1.00 | 2.61 | 2.61 | 2.61 | NA | 1 |
| ALKBH5 | 9.18E-03 | 2 | 2 | 0.002456269 | Adipose_Visceral_Omentum | 1.15 | 0.85 | 0.85 | 0.92 | 3.03 | 1.97 | 1.49 | 2 |
| AMD1 | 9.21E-03 | 2 | 2 | 0.002200001 | Adipose_Visceral_Omentum | 1.19 | 0.81 | 0.81 | 0.58 | 3.06 | 1.82 | 1.76 | 2 |
| ALDH1A1 | 9.23E-03 | 1 | 1 | 0.009225997 | Adipose_Subcutaneous | 1.00 | 1.00 | 1.00 | -2.60 | -2.60 | -2.60 | NA | 1 |
| CCHCR1 | 9.32E-03 | 2 | 2 | 0.002357747 | Adipose_Visceral_Omentum | 1.76 | 0.24 | 0.24 | 2.52 | 3.04 | 2.78 | 0.37 | 2 |
| RP11-162A12.4 | 9.33E-03 | 2 | 2 | 0.01488085 | Adipose_Visceral_Omentum | 1.74 | 0.26 | 0.26 | 0.55 | 2.44 | 1.49 | 1.33 | 2 |
| PC | 9.36E-03 | 2 | 1 | 0.009356002 | Adipose_Subcutaneous | 2.00 | 0.00 | 2.00 | 2.60 | 2.60 | 2.60 | 0.00 | 1 |
| PFDN6 | 9.38E-03 | 1 | 1 | 0.009381003 | Adipose_Subcutaneous | 1.00 | 1.00 | 1.00 | -2.60 | -2.60 | -2.60 | NA | 1 |
| EPC1 | 9.38E-03 | 1 | 1 | 0.009381999 | Adipose_Visceral_Omentum | 1.00 | 1.00 | 1.00 | -2.60 | -2.60 | -2.60 | NA | 1 |
| HSPB6 | 9.41E-03 | 1 | 1 | 0.009413889 | Adipose_Subcutaneous | 1.00 | 1.00 | 1.00 | 2.60 | 2.60 | 2.60 | NA | 1 |
| MMP24-AS1 | 9.50E-03 | 2 | 2 | 0.165224036 | Adipose_Subcutaneous | 1.93 | 0.07 | 0.07 | 0.30 | 1.39 | 0.84 | 0.77 | 2 |
| DUT | 9.64E-03 | 1 | 1 | 0.009637999 | Adipose_Subcutaneous | 1.00 | 1.00 | 1.00 | 2.59 | 2.59 | 2.59 | NA | 1 |
| CCM2 | 9.65E-03 | 2 | 2 | 0.08901001 | Adipose_Subcutaneous | 1.83 | 0.17 | 0.17 | -1.70 | 0.00 | -0.85 | 1.20 | 2 |
| SUDS3 | 9.68E-03 | 2 | 2 | 0.002980571 | Adipose_Subcutaneous | 1.85 | 0.15 | 0.15 | -2.97 | -2.88 | -2.93 | 0.06 | 2 |
| FKBP2 | 9.70E-03 | 1 | 1 | 0.009696248 | Adipose_Visceral_Omentum | 1.00 | 1.00 | 1.00 | -2.59 | -2.59 | -2.59 | NA | 1 |
| RP11-477N3.1 | 9.71E-03 | 2 | 2 | 0.003053821 | Adipose_Subcutaneous | 1.84 | 0.16 | 0.16 | -2.96 | -2.88 | -2.92 | 0.06 | 2 |
| LPGAT1 | 9.78E-03 | 2 | 2 | 0.025526938 | Adipose_Subcutaneous | 1.11 | 0.89 | 0.89 | -1.80 | 2.23 | 0.21 | 2.86 | 2 |
| ZNF391 | 9.80E-03 | 2 | 1 | 0.007066019 | Adipose_Visceral_Omentum | 1.97 | 0.03 | 1.97 | 2.44 | 2.69 | 2.56 | 0.18 | 1 |
| ZNF524 | 9.84E-03 | 2 | 1 | 0.009837999 | Adipose_Subcutaneous | 2.00 | 0.00 | 2.00 | -2.58 | -2.58 | -2.58 | 0.00 | 1 |
| TIMM9 | 9.85E-03 | 1 | 1 | 0.009847943 | Adipose_Visceral_Omentum | 1.00 | 1.00 | 1.00 | -2.58 | -2.58 | -2.58 | NA | 1 |
| RP11-276H19.2 | 9.85E-03 | 2 | 1 | 0.003726001 | Adipose_Subcutaneous | 1.95 | 0.05 | 1.95 | 2.20 | 2.90 | 2.55 | 0.50 | 1 |

| | | | | | | | | | | | | | |
|---|---|---|---|---|---|---|---|---|---|---|---|---|---|
| GOLGA6L10 | 9.86E-03 | 1 | 1 | 0.009856001 | Adipose_Visceral_Omentum | 1.00 | 1.00 | 1.00 | -2.58 | -2.58 | -2.58 | NA | 1 |
| CCDC86 | 9.87E-03 | 2 | 2 | 0.104177946 | Adipose_Visceral_Omentum | 1.83 | 0.17 | 0.17 | -1.62 | 0.10 | -0.76 | 1.22 | 2 |
| FANCI | 9.90E-03 | 1 | 1 | 0.009899081 | Adipose_Visceral_Omentum | 1.00 | 1.00 | 1.00 | -2.58 | -2.58 | -2.58 | NA | 1 |
| EPB41 | 9.94E-03 | 2 | 2 | 0.121600003 | Adipose_Subcutaneous | 1.60 | 0.40 | 0.40 | -1.55 | 1.17 | -0.19 | 1.92 | 2 |
| B9D1 | 9.98E-03 | 2 | 1 | 0.009975 | Adipose_Visceral_Omentum | 2.00 | 0.00 | 2.00 | 2.58 | 2.58 | 2.58 | 0.00 | 1 |

Pvalue, significance p-value of S-MultiXcan association; n, number of "tissues" available for this gene; n_indep, number of independent components of variation kept among the tissues' predictions. (Synthetic independent tissues); p_i_best, best p-value of single-tissue S-PrediXcan association; t_i_best, name of best single-tissue S-PrediXcan association; eigen_max, In the SVD decomposition of predicted expression correlation: eigenvalue (variance explained) of the top independent component; eigen_min, In the SVD decomposition of predicted expression correlation: eigenvalue (variance explained) of the last independent component; eigen_min_kept, In the SVD decomposition of predicted expression correlation: eigenvalue (variance explained) of the smalles independent component that was kept; z_min, minimum z-score among single-tissue S-Predican associations; z_max, maximum z-score among single-tissue S-Predican associations; z_mean, mean z-score among single-tissue S-Predican associations; z_sd, standard deviation of the mean z-score among single-tissue S-Predican associations; tmi, trace of T * T', where Tis correlation of predicted expression levels for different tissues multiplied by its SVD pseudo-inverse.

**Table S15**. TWAS-identified genes associated with anorexia nervosa in the thyroid.

| gene_name | zscore | pvalue | var_g | n_snps_used | n_snps_in_cov | n_snps_in_model | best_gwas_p | largest_weight |
|---|---|---|---|---|---|---|---|---|
| RP11-572O6.1 | 7.534 | 4.925E-14 | 0.000238181 | 2 | 2 | 2 | 4.56199E-14 | 0.04 |
| PRKAR2A | 7.235 | 4.648E-13 | 0.000215477 | 1 | 1 | 1 | 4.648E-13 | 0.04 |
| RP11-3B7.1 | 7.051 | 1.78E-12 | 0.006986973 | 1 | 1 | 1 | 1.78E-12 | 0.20 |
| C3orf62 | 6.854 | 7.202E-12 | 0.002149916 | 1 | 1 | 1 | 7.202E-12 | 0.11 |
| WDR6 | -6.823 | 8.9285E-12 | 0.01588644 | 2 | 2 | 2 | 1.932E-12 | 0.18 |
| USP19 | -5.429 | 5.662E-08 | 0.000124274 | 1 | 1 | 1 | 5.662E-08 | 0.02 |
| NCKIPSD | 5.413 | 6.199E-08 | 0.121283823 | 1 | 1 | 1 | 6.199E-08 | 0.51 |
| CCDC71 | -5.036 | 4.75E-07 | 0.004024042 | 1 | 1 | 1 | 4.75E-07 | 0.09 |
| APEH | -4.891 | 1.004E-06 | 0.000488055 | 1 | 1 | 1 | 1.004E-06 | 0.03 |
| DAG1 | -4.826 | 1.39E-06 | 0.002578667 | 1 | 1 | 1 | 1.39E-06 | 0.07 |
| BSN-AS2 | -4.814 | 1.476E-06 | 0.02599515 | 1 | 1 | 1 | 1.476E-06 | 0.23 |
| KLHDC8B | -4.785 | 1.706E-06 | 0.017968169 | 1 | 1 | 1 | 1.706E-06 | 0.20 |
| ARIH2 | 4.766 | 1.879E-06 | 0.044417279 | 1 | 1 | 1 | 1.879E-06 | 0.31 |
| ARIH2OS | -4.741 | 2.123E-06 | 0.000205131 | 1 | 1 | 1 | 2.123E-06 | 0.02 |
| P4HTM | 4.680 | 2.87E-06 | 0.029681814 | 1 | 1 | 1 | 2.87E-06 | 0.26 |
| DALRD3 | 4.678 | 2.898E-06 | 0.009962355 | 1 | 1 | 1 | 2.898E-06 | 0.15 |
| CHAC2 | 4.630 | 3.6599E-06 | 0.005530317 | 2 | 2 | 2 | 5.454E-07 | 0.14 |
| ASB3 | 4.430 | 9.408E-06 | 0.001421996 | 1 | 1 | 1 | 9.40799E-06 | 0.08 |
| BSN | -4.430 | 9.441E-06 | 0.000622772 | 1 | 1 | 1 | 9.441E-06 | 0.04 |
| MGMT | 4.271 | 1.9431E-05 | 0.814800269 | 3 | 4 | 4 | 1.268E-06 | 0.77 |
| SUOX | 4.238 | 2.256E-05 | 0.190143238 | 1 | 4 | 4 | 2.256E-05 | 0.62 |
| PRKCSH | -4.171 | 3.039E-05 | 0.007933395 | 1 | 2 | 2 | 3.039E-05 | 0.23 |
| FAM180B | 4.155 | 3.2562E-05 | 0.001963893 | 3 | 3 | 3 | 0.001956 | 0.11 |
| RBM6 | -4.124 | 3.716E-05 | 0.141080055 | 1 | 1 | 1 | 3.716E-05 | 0.52 |
| MSH5 | -4.103 | 4.08E-05 | 0.005741465 | 3 | 3 | 3 | 1.687E-05 | 0.17 |
| SLC17A7 | 4.093 | 4.2617E-05 | 0.004807267 | 2 | 2 | 2 | 0.0001649 | 0.06 |
| HLA-DMB | -4.084 | 4.4324E-05 | 0.016149282 | 2 | 2 | 2 | 2.893E-05 | 0.35 |
| GBE1 | -4.016 | 5.9242E-05 | 0.004696509 | 2 | 3 | 3 | 9.295E-06 | 0.07 |
| PTBP2 | 3.993 | 6.536E-05 | 0.005488571 | 1 | 1 | 1 | 6.536E-05 | 0.11 |
| CYP7B1 | 3.985 | 6.7526E-05 | 0.013043655 | 2 | 3 | 3 | 0.0003999 | 0.20 |
| RNF123 | 3.935 | 8.326E-05 | 0.030610553 | 1 | 2 | 2 | 8.326E-05 | 0.24 |
| HLA-DMA | 3.899 | 9.6549E-05 | 0.786734511 | 2 | 3 | 3 | 8.529E-05 | 1.27 |
| WDR35 | -3.894 | 9.8616E-05 | 0.086418568 | 3 | 3 | 3 | 0.005462998 | 0.38 |
| FBXO27 | -3.882 | 0.0001035 | 0.031098657 | 1 | 1 | 1 | 0.0001035 | 0.30 |

| | | | | | | | | |
|---|---|---|---|---|---|---|---|---|
| MST1R | 3.878 | 0.0001055 | 0.027125608 | 1 | 2 | 2 | 0.0001055 | 0.23 |
| LOXL4 | 3.860 | 0.0001132 | 0.034952821 | 1 | 2 | 2 | 0.0001132 | 0.26 |
| DHFR2 | 3.859 | 0.0001137 | 3.22632E-05 | 1 | 2 | 2 | 0.0001137 | 0.01 |
| ZKSCAN3 | -3.806 | 0.00014117 | 0.041179328 | 2 | 3 | 3 | 0.000857 | 0.30 |
| ARHGEF15 | 3.796 | 0.0001472 | 0.002964039 | 1 | 1 | 1 | 0.0001472 | 0.08 |
| STX19 | 3.789 | 0.0001511 | 0.094468962 | 1 | 1 | 1 | 0.0001511 | 0.52 |
| RNF2 | -3.726 | 0.0001948 | 0.004976491 | 1 | 1 | 1 | 0.0001948 | 0.11 |
| SHTN1 | -3.715 | 0.0002033 | 0.007735088 | 1 | 1 | 1 | 0.0002033 | 0.14 |
| C4A | -3.686 | 0.00022802 | 0.159014537 | 2 | 2 | 2 | 5.001E-05 | 0.93 |
| SELENOH | 3.685 | 0.0002283 | 0.000871805 | 1 | 1 | 1 | 0.0002283 | 0.04 |
| RGPD8 | -3.666 | 0.0002466 | 0.415186996 | 1 | 3 | 3 | 0.0002466 | 1.22 |
| LY6G6F | -3.655 | 0.0002569 | 0.000135812 | 1 | 1 | 1 | 0.0002569 | 0.02 |
| ZSCAN26 | -3.655 | 0.00025744 | 0.154787691 | 2 | 2 | 2 | 0.0008678 | 0.43 |
| TRIM33 | -3.646 | 0.00026611 | 0.001134493 | 2 | 2 | 2 | 6.288E-05 | 0.05 |
| FOXA2 | 3.633 | 0.00028 | 0.000415097 | 1 | 1 | 1 | 0.00028 | 0.07 |
| AKNA | 3.633 | 0.00028008 | 0.002181057 | 2 | 2 | 2 | 0.000244 | 0.06 |
| TMEM89 | -3.630 | 0.00028358 | 0.078411124 | 2 | 2 | 2 | 0.0002427 | 0.38 |
| MICB | 3.624 | 0.0002898 | 0.278954076 | 5 | 6 | 6 | 1.245E-05 | 0.82 |
| PRUNE1 | 3.623 | 0.00029067 | 0.00444897 | 2 | 4 | 4 | 0.0003027 | 0.14 |
| ZSCAN23 | 3.613 | 0.00030305 | 0.369314004 | 4 | 4 | 4 | 0.0004094 | 0.50 |
| SLFNL1 | 3.611 | 0.000305 | 0.014447832 | 1 | 1 | 1 | 0.000305 | 0.37 |
| ZSCAN31 | 3.578 | 0.00034571 | 1.211620074 | 4 | 4 | 4 | 0.0004956 | 1.01 |
| RAB5B | -3.574 | 0.0003517 | 0.006298281 | 1 | 1 | 1 | 0.0003517 | 0.11 |
| THOC5 | -3.574 | 0.00035221 | 0.674580069 | 3 | 4 | 4 | 4.19E-05 | 0.79 |
| ACTR1A | 3.559 | 0.0003722 | 0.001545173 | 1 | 1 | 1 | 0.0003722 | 0.06 |
| PRR7 | 3.552 | 0.0003821 | 0.003519856 | 1 | 1 | 1 | 0.0003821 | 0.08 |
| VAMP2 | -3.552 | 0.0003829 | 0.011102436 | 1 | 1 | 1 | 0.0003829 | 0.16 |
| IQCK | -3.530 | 0.0004161 | 0.012474758 | 1 | 1 | 1 | 0.0004161 | 0.21 |
| CDC73 | -3.524 | 0.0004251 | 0.001228909 | 1 | 2 | 2 | 0.0004251 | 0.05 |
| ITK | 3.517 | 0.0004358 | 5.02564E-07 | 1 | 1 | 1 | 0.0004358 | 0.00 |
| DYNC1I2 | 3.512 | 0.0004442 | 0.001746323 | 1 | 3 | 3 | 0.0004442 | 0.06 |
| TMEM198 | -3.505 | 0.00045727 | 0.011983983 | 2 | 2 | 2 | 0.0004182 | 0.18 |
| LIN37 | 3.489 | 0.0004848 | 0.013721741 | 1 | 1 | 1 | 0.0004848 | 0.21 |
| ZBTB1 | 3.488 | 0.0004858 | 0.021464131 | 1 | 1 | 1 | 0.0004858 | 0.21 |
| RAD51 | 3.487 | 0.0004889 | 0.000191881 | 1 | 1 | 1 | 0.0004889 | 0.06 |
| RP11-129M6.1 | 3.479 | 0.0005033 | 0.038574957 | 1 | 1 | 1 | 0.0005033 | 0.33 |
| ZCCHC2 | 3.472 | 0.00051717 | 0.025952733 | 3 | 3 | 3 | 0.0005277 | 0.14 |

| | | | | | | | | |
|---|---|---|---|---|---|---|---|---|
| NAA60 | -3.460 | 0.0005411 | 0.005059771 | 1 | 1 | 1 | 0.0005411 | 0.14 |
| SLC25A35 | 3.454 | 0.00055155 | 0.009845687 | 2 | 2 | 2 | 9.354E-05 | 0.16 |
| CEP290 | 3.452 | 0.00055668 | 0.052966224 | 3 | 3 | 3 | 0.0004047 | 0.57 |
| CCDC36 | 3.443 | 0.00057514 | 0.030643868 | 2 | 3 | 3 | 6.254E-07 | 0.16 |
| ZFP91 | -3.440 | 0.0005809 | 0.001990987 | 1 | 1 | 1 | 0.0005809 | 0.07 |
| NMT2 | 3.434 | 0.00059557 | 0.10526323 | 3 | 3 | 3 | 0.003155 | 0.29 |
| RP11-227D13.1 | -3.432 | 0.0005994 | 0.029251946 | 1 | 2 | 2 | 0.0005994 | 0.32 |
| SYN2 | -3.420 | 0.00062541 | 0.33544346 | 2 | 2 | 2 | 8.813E-05 | 0.70 |
| CBR3 | 3.416 | 0.0006365 | 0.018881144 | 1 | 1 | 1 | 0.0006365 | 0.21 |
| GLRX2 | -3.409 | 0.0006519 | 0.003227037 | 1 | 2 | 2 | 0.0006519 | 0.09 |
| MYO19 | 3.403 | 0.0006671 | 0.021641779 | 1 | 2 | 2 | 0.0006671 | 0.21 |
| NDUFS3 | 3.396 | 0.00068303 | 0.00112877 | 2 | 2 | 2 | 0.001911999 | 0.04 |
| CCNE1 | -3.381 | 0.0007234 | 0.093127396 | 1 | 1 | 1 | 0.0007234 | 0.46 |
| FGF14-AS2 | 3.376 | 0.00073423 | 0.040317638 | 2 | 2 | 2 | 0.0004745 | 0.32 |
| DNASE1 | 3.369 | 0.000754 | 0.003246146 | 1 | 1 | 1 | 0.000754 | 0.09 |
| SRRM5 | -3.358 | 0.0007852 | 0.006569565 | 1 | 2 | 2 | 0.0007852 | 0.14 |
| IKZF5 | 3.354 | 0.00079774 | 0.019677491 | 2 | 2 | 2 | 6.13401E-05 | 0.12 |
| SYTL1 | 3.350 | 0.00080841 | 0.006033459 | 2 | 3 | 3 | 0.004571001 | 0.26 |
| TMEM51 | 3.347 | 0.0008174 | 0.013071966 | 1 | 2 | 2 | 0.0008174 | 0.21 |
| ZNF653 | -3.345 | 0.0008214 | 2.9887E-06 | 1 | 2 | 2 | 0.0008214 | 0.01 |
| RPL22 | 3.340 | 0.00083839 | 0.000118897 | 2 | 2 | 2 | 0.0006481 | 0.02 |
| EPB41 | 3.336 | 0.0008507 | 0.001242656 | 1 | 2 | 2 | 0.0008507 | 0.09 |
| CTD-2325M2.1 | -3.322 | 0.00089255 | 0.50941832 | 4 | 4 | 4 | 0.001712001 | 0.64 |
| TOMM34 | -3.322 | 0.0008928 | 0.047141499 | 1 | 2 | 2 | 0.0008928 | 0.36 |
| MST1 | -3.317 | 0.00091047 | 0.176605247 | 4 | 6 | 6 | 1.146E-06 | 0.47 |
| PRADC1 | -3.316 | 0.0009121 | 0.00031793 | 1 | 1 | 1 | 0.0009121 | 0.09 |
| RP11-51J9.5 | -3.316 | 0.0009146 | 0.007209862 | 1 | 1 | 1 | 0.0009146 | 0.27 |
| SFMBT2 | -3.313 | 0.00092156 | 0.007417798 | 2 | 2 | 2 | 0.0009833 | 0.13 |
| KBTBD4 | -3.294 | 0.0009879 | 0.000280949 | 1 | 1 | 1 | 0.0009879 | 0.03 |
| IGFLR1 | 3.294 | 0.00098853 | 0.048579256 | 3 | 3 | 3 | 0.001219 | 0.38 |
| IL1RN | -3.277 | 0.001048 | 0.027402877 | 1 | 2 | 2 | 0.001048 | 0.23 |
| ZBTB22 | -3.275 | 0.001056 | 0.006095132 | 1 | 1 | 1 | 0.001056 | 0.11 |
| LY75 | 3.270 | 0.001075 | 0.048396625 | 1 | 3 | 3 | 0.001075 | 0.37 |
| KIF7 | 3.254 | 0.001136 | 0.01578322 | 1 | 1 | 1 | 0.001136 | 0.17 |
| BNIPL | 3.253 | 0.00114221 | 0.500261898 | 3 | 3 | 3 | 0.0002907 | 0.86 |
| ZNF766 | 3.251 | 0.00115052 | 0.0045934 | 2 | 2 | 2 | 0.00106 | 0.14 |
| RAB26 | -3.248 | 0.001163 | 2.31426E-05 | 1 | 1 | 1 | 0.001163 | 0.01 |

| | | | | | | | | |
|---|---|---|---|---|---|---|---|---|
| RWDD3 | 3.238 | 0.00120179 | 0.012306278 | 2 | 5 | 5 | 0.001004 | 0.50 |
| GCSAM | 3.233 | 0.00122567 | 0.001741 | 2 | 2 | 2 | 0.0001287 | 0.09 |
| BTBD3 | -3.229 | 0.001244 | 0.005438504 | 1 | 1 | 1 | 0.001244 | 0.11 |
| PWWP2A | -3.216 | 0.00129843 | 0.007499213 | 2 | 2 | 2 | 0.001142 | 0.13 |
| CADM1 | 3.203 | 0.001359 | 0.349482786 | 1 | 1 | 1 | 0.001358999 | 0.85 |
| CDKN2AIP | -3.192 | 0.001413 | 0.002620927 | 1 | 1 | 1 | 0.001413 | 0.08 |
| CPNE6 | -3.192 | 0.0014151 | 0.041639149 | 2 | 2 | 2 | 0.001249 | 0.27 |
| DDX5 | -3.187 | 0.001438 | 0.014964189 | 1 | 2 | 2 | 0.001438 | 0.19 |
| AFF3 | -3.184 | 0.001452 | 0.002793518 | 1 | 1 | 1 | 0.001452 | 0.09 |
| CEBPZ | -3.182 | 0.00146359 | 0.039692274 | 3 | 3 | 3 | 0.001676 | 0.28 |
| HAUS4 | 3.182 | 0.00146464 | 0.83477628 | 3 | 3 | 3 | 0.003663 | 0.59 |
| U2AF1L4 | 3.181 | 0.00146784 | 0.429227124 | 2 | 2 | 2 | 0.001465999 | 1.00 |
| RP11-244E17.1 | -3.173 | 0.00150622 | 0.033932974 | 3 | 3 | 3 | 0.005550002 | 0.20 |
| STYX | -3.173 | 0.00150781 | 0.08909405 | 2 | 3 | 3 | 0.001686 | 0.25 |
| GLRB | -3.168 | 0.001533 | 0.004902883 | 1 | 1 | 1 | 0.001533001 | 0.11 |
| TMEM245 | -3.165 | 0.00155297 | 0.028087743 | 2 | 2 | 2 | 0.005082002 | 0.15 |
| USP53 | 3.161 | 0.00157224 | 0.023263167 | 2 | 2 | 2 | 0.004385002 | 0.22 |
| STAT6 | 3.150 | 0.00163387 | 0.000652033 | 2 | 2 | 2 | 0.008720999 | 0.07 |
| SLC44A4 | 3.149 | 0.00163676 | 0.033826998 | 3 | 3 | 3 | 0.001712001 | 0.22 |
| MTCL1 | -3.143 | 0.00166976 | 0.089136711 | 2 | 2 | 2 | 0.0004886 | 0.33 |
| NDUFAF7 | 3.142 | 0.001676 | 0.016360021 | 1 | 1 | 1 | 0.001676 | 0.19 |
| CTA-989H11.1 | 3.141 | 0.001684 | 0.014989192 | 1 | 2 | 2 | 0.001684 | 0.19 |
| DRAM1 | 3.138 | 0.00169841 | 0.030269511 | 3 | 3 | 3 | 9.629E-05 | 0.68 |
| HSPB6 | -3.126 | 0.001772 | 0.001547004 | 1 | 1 | 1 | 0.001772 | 0.08 |
| SPEN | 3.125 | 0.00177609 | 0.013090362 | 2 | 2 | 2 | 0.000673 | 0.12 |
| TRPV1 | -3.123 | 0.00178841 | 0.021126392 | 3 | 4 | 4 | 0.005183001 | 0.25 |
| SH2B3 | -3.116 | 0.001834 | 3.58976E-05 | 1 | 1 | 1 | 0.001834 | 0.01 |
| PPP1R18 | -3.115 | 0.00183659 | 0.007755474 | 2 | 3 | 3 | 0.001939 | 0.22 |
| PIGW | 3.113 | 0.00185085 | 0.005369394 | 2 | 2 | 2 | 0.002639 | 0.13 |
| MKX | 3.109 | 0.001878 | 0.000383254 | 1 | 2 | 2 | 0.001878001 | 0.03 |
| PTBP3 | 3.108 | 0.00188241 | 0.009290053 | 2 | 2 | 2 | 0.004474001 | 0.13 |
| ADGRE2 | 3.107 | 0.00188816 | 0.013935385 | 2 | 2 | 2 | 0.020430005 | 0.14 |
| CYP3A5 | 3.095 | 0.00197 | 0.015439699 | 1 | 1 | 1 | 0.00197 | 0.43 |
| CXCL5 | -3.087 | 0.002021 | 0.003112127 | 1 | 1 | 1 | 0.002021 | 0.12 |
| REPS1 | 3.076 | 0.00209636 | 0.001209369 | 2 | 3 | 3 | 0.001522 | 0.06 |
| CDHR4 | 3.076 | 0.0021 | 0.005674614 | 1 | 1 | 1 | 0.002099999 | 0.11 |
| DDX31 | -3.074 | 0.00211098 | 0.193828356 | 2 | 2 | 2 | 0.002395 | 0.71 |

| | | | | | | | | |
|---|---|---|---|---|---|---|---|---|
| SMARCC2 | -3.071 | 0.00213 | 0.0007439 | 1 | 1 | 1 | 0.002130001 | 0.07 |
| PSMG3-AS1 | 3.069 | 0.00214808 | 0.271752926 | 7 | 7 | 7 | 0.017130005 | 0.61 |
| SPI1 | -3.068 | 0.002155 | 2.47835E-06 | 1 | 1 | 1 | 0.002155 | 0.00 |
| ZC3H6 | 3.066 | 0.002169 | 0.001579712 | 1 | 3 | 3 | 0.002169001 | 0.06 |
| SEMA6D | 3.060 | 0.002213 | 0.004904217 | 1 | 1 | 1 | 0.002213 | 0.10 |
| HSPG2 | -3.060 | 0.002217 | 0.03880776 | 1 | 2 | 2 | 0.002216999 | 0.60 |
| SPHKAP | -3.057 | 0.002235 | 0.000335082 | 1 | 2 | 2 | 0.002235 | 0.03 |
| PIK3R3 | -3.045 | 0.00232828 | 0.003236284 | 2 | 2 | 2 | 0.0009705 | 0.05 |
| RP11-16E12.1 | 3.035 | 0.002403 | 0.007246627 | 1 | 2 | 2 | 0.002402999 | 0.20 |
| CDKN1A | -3.026 | 0.002477 | 5.29767E-05 | 1 | 1 | 1 | 0.002477 | 0.02 |
| SCIMP | 3.024 | 0.00249151 | 0.100769164 | 2 | 2 | 2 | 0.008067 | 0.55 |
| DNAJC22 | 3.017 | 0.00255299 | 0.061619269 | 3 | 3 | 3 | 7.88699E-06 | 0.57 |
| LINC00324 | -3.014 | 0.00257498 | 0.013485678 | 2 | 3 | 3 | 3.724E-07 | 0.47 |
| SOAT1 | 3.011 | 0.00260657 | 0.003526765 | 2 | 2 | 2 | 0.033060001 | 0.22 |
| ZKSCAN8 | 3.009 | 0.00262057 | 0.002175555 | 2 | 2 | 2 | 0.001867999 | 0.08 |
| ADAM22 | 3.002 | 0.00268341 | 0.033203767 | 3 | 4 | 4 | 0.0004892 | 0.21 |
| DLG4 | 3.000 | 0.00270173 | 0.001712063 | 2 | 2 | 2 | 0.001028 | 0.06 |
| SLC9A8 | -2.998 | 0.00271985 | 0.046201193 | 4 | 4 | 4 | 0.008363 | 0.91 |
| TPK1 | -2.995 | 0.00274022 | 0.066845969 | 3 | 3 | 3 | 0.007818002 | 0.34 |
| MFAP1 | -2.993 | 0.002763 | 8.75893E-05 | 1 | 1 | 1 | 0.002763 | 0.02 |
| GLI1 | 2.992 | 0.002773 | 9.53768E-05 | 1 | 2 | 2 | 0.002772999 | 0.01 |
| PLCL2 | -2.991 | 0.002778 | 0.0020258 | 1 | 1 | 1 | 0.002777999 | 0.07 |
| CCL15 | 2.989 | 0.0028021 | 0.069050019 | 2 | 2 | 2 | 0.006401998 | 0.59 |
| CNKSR1 | 2.978 | 0.00290566 | 0.018295247 | 2 | 4 | 4 | 0.010120003 | 0.31 |
| CHCHD5 | -2.977 | 0.00290909 | 0.019050687 | 3 | 3 | 3 | 0.010439998 | 0.23 |
| CAMK2N1 | -2.968 | 0.00299314 | 0.039806177 | 2 | 3 | 3 | 0.003248001 | 0.36 |
| ZNF80 | -2.965 | 0.0030242 | 0.014005811 | 3 | 3 | 3 | 0.001265 | 0.13 |
| ZC3H8 | 2.965 | 0.00302739 | 0.038675778 | 3 | 3 | 3 | 5.156E-05 | 0.33 |
| CKMT1A | 2.964 | 0.003041 | 0.06059601 | 1 | 3 | 3 | 0.003041001 | 0.54 |
| GALC | -2.959 | 0.00309131 | 0.053104648 | 2 | 2 | 2 | 0.003543001 | 0.43 |
| TRPT1 | -2.954 | 0.00313209 | 0.029844923 | 3 | 4 | 4 | 0.015580002 | 0.33 |
| EPN3 | 2.947 | 0.00321346 | 0.145194208 | 2 | 3 | 3 | 0.002982 | 0.54 |
| ADD2 | -2.946 | 0.003222 | 0.136142605 | 1 | 2 | 2 | 0.003222 | 0.87 |
| RP11-65M17.3 | 2.945 | 0.003228 | 0.010098653 | 1 | 1 | 1 | 0.003228 | 0.18 |
| DTNB | 2.938 | 0.003303 | 0.003100715 | 1 | 2 | 2 | 0.003303 | 0.08 |
| CNTLN | -2.935 | 0.00333829 | 0.087422764 | 3 | 3 | 3 | 0.003369001 | 0.30 |
| MRPL35 | 2.934 | 0.00335143 | 0.021736259 | 2 | 2 | 2 | 0.0006144 | 0.34 |

| | | | | | | | | |
|---|---|---|---|---|---|---|---|---|
| ARC | -2.932 | 0.00336961 | 0.071650758 | 2 | 2 | 2 | 0.003379 | 0.24 |
| ARMC4 | 2.931 | 0.00337351 | 0.081246031 | 2 | 2 | 2 | 0.001878001 | 0.35 |
| TBC1D4 | 2.930 | 0.003392 | 0.087221932 | 1 | 1 | 1 | 0.003392 | 0.42 |
| LRFN5 | 2.924 | 0.003454 | 0.021271186 | 1 | 1 | 1 | 0.003454001 | 0.21 |
| ALPK3 | -2.923 | 0.00346775 | 0.025234884 | 2 | 2 | 2 | 0.00214 | 0.17 |
| ECSIT | 2.913 | 0.00357638 | 0.014115603 | 3 | 3 | 3 | 0.001229 | 0.30 |
| PTGR2 | 2.908 | 0.00363408 | 0.436090593 | 6 | 6 | 6 | 0.01373 | 0.70 |
| CALM3 | -2.903 | 0.00370072 | 0.005729245 | 2 | 2 | 2 | 0.013650004 | 0.14 |
| LINC00923 | -2.899 | 0.00374 | 0.029039293 | 1 | 5 | 5 | 0.003739999 | 0.25 |
| LINC01750 | -2.895 | 0.003786 | 0.020180029 | 1 | 2 | 2 | 0.003786 | 0.31 |
| SEMA3D | 2.895 | 0.003792 | 0.001023168 | 1 | 2 | 2 | 0.003792 | 0.05 |
| CSNK2A2 | 2.895 | 0.003795 | 0.007055126 | 1 | 1 | 1 | 0.003795 | 0.14 |
| TK2 | -2.895 | 0.0037971 | 0.023081154 | 2 | 2 | 2 | 0.003996 | 0.34 |
| RP11-274B21.9 | 2.894 | 0.00380832 | 0.152526939 | 2 | 2 | 2 | 0.004108999 | 0.51 |
| PIP5K1A | -2.892 | 0.00382921 | 0.013080957 | 2 | 2 | 2 | 0.03345 | 0.40 |
| SMURF1 | -2.891 | 0.003835 | 0.003940512 | 1 | 1 | 1 | 0.003835001 | 0.12 |
| C2orf49 | 2.888 | 0.00387797 | 0.004059123 | 2 | 2 | 2 | 0.008856001 | 0.08 |
| CLDN7 | 2.886 | 0.00390595 | 0.027106109 | 4 | 4 | 4 | 0.0009086 | 0.18 |
| B4GALT6 | 2.880 | 0.00398123 | 0.044836936 | 3 | 3 | 3 | 0.032600002 | 0.63 |
| SYT9 | 2.879 | 0.00398874 | 0.076334992 | 2 | 2 | 2 | 0.004646999 | 0.42 |
| B3GLCT | 2.878 | 0.00399974 | 0.168302755 | 2 | 2 | 2 | 0.004052 | 0.73 |
| ZNF565 | 2.872 | 0.00407558 | 0.252633825 | 2 | 2 | 2 | 0.007238002 | 0.51 |
| ELAVL1 | 2.865 | 0.00417 | 6.80433E-11 | 1 | 1 | 1 | 0.004170001 | 0.00 |
| STAG3 | -2.859 | 0.00424326 | 0.039361055 | 3 | 3 | 3 | 0.005998999 | 0.33 |
| AC012370.3 | 2.855 | 0.00430136 | 0.791594172 | 2 | 2 | 2 | 0.00306 | 0.76 |
| EVI2A | -2.851 | 0.004363 | 0.006706366 | 1 | 1 | 1 | 0.004362999 | 0.12 |
| ANGEL2 | 2.840 | 0.00450714 | 0.104708484 | 2 | 3 | 3 | 0.004389999 | 0.34 |
| TAF6 | 2.834 | 0.00459 | 0.029473975 | 1 | 1 | 1 | 0.004590002 | 0.24 |
| KIFC3 | 2.832 | 0.004623 | 0.002600476 | 1 | 1 | 1 | 0.004622998 | 0.08 |
| RFX5 | 2.831 | 0.004647 | 0.004415879 | 1 | 1 | 1 | 0.004646999 | 0.10 |
| C1orf194 | -2.822 | 0.00476681 | 0.038818047 | 2 | 2 | 2 | 0.011460001 | 0.30 |
| IFNE | -2.821 | 0.00479 | 0.00425691 | 1 | 1 | 1 | 0.004789999 | 0.19 |
| LOXL1 | -2.817 | 0.00484738 | 0.003673513 | 3 | 3 | 3 | 0.023540002 | 0.11 |
| ELOVL7 | 2.815 | 0.00487432 | 0.035227305 | 3 | 3 | 3 | 0.007474 | 0.26 |
| TRIM41 | 2.813 | 0.004912 | 0.011838478 | 1 | 2 | 2 | 0.004911999 | 0.18 |
| CYHR1 | -2.811 | 0.00493318 | 0.012259714 | 2 | 3 | 3 | 0.005222 | 0.16 |
| CMTM2 | -2.809 | 0.004977 | 0.010814918 | 1 | 1 | 1 | 0.004977 | 0.26 |

| | | | | | | | | |
|---|---|---|---|---|---|---|---|---|
| WISP1 | -2.807 | 0.005002 | 0.001794123 | 1 | 1 | 1 | 0.005001999 | 0.06 |
| PTCH1 | -2.807 | 0.005004 | 0.004555144 | 1 | 1 | 1 | 0.005004002 | 0.18 |
| GRAMD1B | 2.804 | 0.00505 | 0.000232872 | 1 | 1 | 1 | 0.005049999 | 0.08 |
| SEPT7-AS1 | -2.804 | 0.005051 | 0.002651188 | 1 | 2 | 2 | 0.005050999 | 0.11 |
| C18orf21 | -2.803 | 0.005064 | 0.002321609 | 1 | 1 | 1 | 0.005064 | 0.08 |
| ARF6 | 2.801 | 0.005093 | 0.001977693 | 1 | 1 | 1 | 0.005093001 | 0.10 |
| TAPBP | 2.800 | 0.005103 | 0.033329654 | 1 | 2 | 2 | 0.005103 | 0.29 |
| CTDNEP1 | 2.798 | 0.005147 | 0.005484782 | 1 | 1 | 1 | 0.005146999 | 0.11 |
| LINC01094 | -2.797 | 0.005165 | 0.010671497 | 1 | 1 | 1 | 0.005165 | 0.17 |
| MYO5A | 2.795 | 0.00519117 | 0.046499276 | 3 | 3 | 3 | 0.0004341 | 0.58 |
| ICE1 | 2.792 | 0.005236 | 9.50455E-06 | 1 | 1 | 1 | 0.005236 | 0.00 |
| ZNF532 | 2.789 | 0.005289 | 0.004635339 | 1 | 1 | 1 | 0.005289002 | 0.10 |
| SLC39A13 | 2.786 | 0.00533706 | 0.010722375 | 2 | 2 | 2 | 0.0002479 | 0.09 |
| C6orf226 | 2.783 | 0.005391 | 0.027426267 | 1 | 2 | 2 | 0.005391 | 0.72 |
| FBXO9 | -2.782 | 0.00540663 | 0.005437008 | 2 | 2 | 2 | 0.026329995 | 0.13 |
| RP11-47A8.5 | -2.781 | 0.005425 | 0.004359742 | 1 | 1 | 1 | 0.005425001 | 0.09 |
| GPBP1L1 | -2.780 | 0.00543486 | 0.0005666 | 2 | 2 | 2 | 0.005339 | 0.03 |
| PPARG | 2.777 | 0.00548477 | 0.012744552 | 2 | 2 | 2 | 0.001244 | 0.20 |
| ZSWIM8 | -2.777 | 0.00549042 | 0.005750993 | 2 | 2 | 2 | 0.012179999 | 0.09 |
| AC007405.4 | 2.776 | 0.005496 | 0.012658393 | 1 | 1 | 1 | 0.005496001 | 0.17 |
| WASL | 2.772 | 0.005569 | 0.000827019 | 1 | 1 | 1 | 0.005569001 | 0.15 |
| GOLIM4 | -2.770 | 0.00560301 | 0.010263568 | 3 | 3 | 3 | 0.001281 | 0.22 |
| DR1 | -2.768 | 0.005648 | 0.012581757 | 1 | 1 | 1 | 0.005648 | 0.17 |
| GAS1RR | -2.767 | 0.00566196 | 0.066202955 | 2 | 2 | 2 | 0.0009517 | 0.39 |
| SUSD4 | -2.763 | 0.005729 | 0.00981313 | 3 | 3 | 3 | 0.020909999 | 0.19 |
| ODF3B | -2.762 | 0.00574836 | 0.019603943 | 2 | 2 | 2 | 0.002617 | 0.35 |
| INHBE | 2.760 | 0.005773 | 0.010607585 | 1 | 1 | 1 | 0.005772999 | 0.15 |
| LINC01224 | -2.760 | 0.00577511 | 0.055393995 | 2 | 2 | 2 | 0.005658002 | 0.21 |
| RBMXL1 | 2.760 | 0.00577608 | 0.094673069 | 2 | 3 | 3 | 0.002707001 | 0.27 |
| CDCP1 | 2.760 | 0.0057765 | 0.018226608 | 2 | 2 | 2 | 0.074550005 | 0.18 |
| ONECUT2 | 2.756 | 0.005852 | 0.00010743 | 1 | 1 | 1 | 0.005851998 | 0.03 |
| IQCH | -2.756 | 0.00585959 | 0.117050317 | 2 | 2 | 2 | 0.004435999 | 0.34 |
| NCLN | 2.750 | 0.00595142 | 0.19763916 | 2 | 2 | 2 | 0.007146998 | 0.38 |
| MDH2 | -2.750 | 0.0059537 | 0.006172797 | 2 | 2 | 2 | 0.0009106 | 0.07 |
| ZNF304 | 2.749 | 0.0059733 | 0.159663569 | 2 | 2 | 2 | 0.005536999 | 0.68 |
| SRGAP3 | 2.747 | 0.00601374 | 0.014589787 | 2 | 2 | 2 | 0.043279997 | 0.37 |
| ACSF2 | 2.741 | 0.00613223 | 0.002289772 | 2 | 2 | 2 | 0.006142002 | 0.11 |

| | | | | | | | | |
|---|---|---|---|---|---|---|---|---|
| GPANK1 | -2.737 | 0.00620048 | 0.044298239 | 4 | 5 | 5 | 2.048E-05 | 0.31 |
| RP11-555G19.1 | 2.736 | 0.00622301 | 0.081826984 | 2 | 3 | 3 | 0.006186 | 0.55 |
| CLTB | 2.735 | 0.00623303 | 0.001314711 | 2 | 2 | 2 | 0.001697 | 0.06 |
| CTNS | 2.734 | 0.006253 | 0.009271296 | 1 | 1 | 1 | 0.006253001 | 0.30 |
| ARHGAP12 | -2.729 | 0.00635044 | 0.002513811 | 2 | 2 | 2 | 0.007933998 | 0.07 |
| HILPDA | -2.728 | 0.006378 | 8.82603E-05 | 1 | 2 | 2 | 0.006377998 | 0.05 |
| CEPT1 | 2.722 | 0.006483 | 0.000664562 | 1 | 1 | 1 | 0.006483 | 0.04 |
| DRAM2 | 2.722 | 0.006483 | 0.12724556 | 1 | 1 | 1 | 0.006483 | 0.55 |
| UQCC2 | 2.718 | 0.00655789 | 0.122434496 | 3 | 3 | 3 | 3.564E-05 | 0.44 |
| CMTM1 | -2.718 | 0.006565 | 0.006149516 | 1 | 1 | 1 | 0.006564998 | 0.17 |
| VPS53 | -2.718 | 0.00657001 | 0.581419201 | 3 | 3 | 3 | 0.015050003 | 0.74 |
| MOCOS | 2.717 | 0.00658674 | 0.207448814 | 4 | 4 | 4 | 0.005658999 | 0.77 |
| BBOF1 | -2.712 | 0.006686 | 0.035581441 | 1 | 1 | 1 | 0.006686 | 0.28 |
| PRTG | 2.706 | 0.006801 | 0.066169392 | 1 | 4 | 4 | 0.006801 | 0.43 |
| COL11A2 | -2.705 | 0.00683498 | 0.112054499 | 2 | 2 | 2 | 0.008053999 | 0.46 |
| ZNF391 | 2.704 | 0.00686086 | 0.161001924 | 4 | 5 | 5 | 0.002966001 | 0.56 |
| CYB5R4 | 2.703 | 0.006863 | 0.002079798 | 1 | 1 | 1 | 0.006862998 | 0.18 |
| INTS12 | -2.702 | 0.006892 | 0.000123717 | 1 | 3 | 3 | 0.006891998 | 0.03 |
| SEMA7A | 2.701 | 0.006911 | 7.23122E-05 | 1 | 1 | 1 | 0.006910999 | 0.03 |
| NDUFAF4 | -2.699 | 0.00695907 | 0.030120415 | 2 | 2 | 2 | 0.015500001 | 0.33 |
| RP11-184E9.2 | 2.697 | 0.00700715 | 1.44093774 | 3 | 3 | 3 | 9.80699E-06 | 1.18 |
| GGNBP2 | -2.695 | 0.0070357 | 0.016359558 | 2 | 2 | 2 | 0.007237 | 0.18 |
| SCN11A | -2.694 | 0.00706 | 0.012280879 | 1 | 1 | 1 | 0.007060001 | 0.34 |
| TMED5 | 2.689 | 0.007176 | 0.009855226 | 1 | 1 | 1 | 0.007176001 | 0.15 |
| CEP95 | 2.688 | 0.00718969 | 0.005636335 | 2 | 2 | 2 | 0.001438 | 0.11 |
| JMJD8 | -2.684 | 0.007283 | 0.029419537 | 1 | 1 | 1 | 0.007282999 | 0.32 |
| PALD1 | -2.682 | 0.007308 | 0.005191766 | 1 | 2 | 2 | 0.007308001 | 0.13 |
| CDCA2 | 2.682 | 0.007317 | 0.012595569 | 1 | 1 | 1 | 0.007317002 | 0.17 |
| EPHB2 | -2.680 | 0.00737 | 0.018743569 | 1 | 1 | 1 | 0.00737 | 0.19 |
| SGTB | 2.678 | 0.007417 | 0.00525409 | 1 | 1 | 1 | 0.007417 | 0.12 |
| PECR | -2.676 | 0.007457 | 0.017042822 | 1 | 2 | 2 | 0.007456998 | 0.19 |
| CFAP74 | -2.676 | 0.00745833 | 0.0297502 | 2 | 2 | 2 | 0.007893002 | 0.28 |
| ERICH2 | 2.673 | 0.007517 | 0.017959909 | 1 | 1 | 1 | 0.007516998 | 0.20 |
| ZNF574 | 2.670 | 0.007596 | 0.001636993 | 1 | 1 | 1 | 0.007595999 | 0.08 |
| LHPP | 2.669 | 0.00760667 | 0.120119415 | 5 | 5 | 5 | 0.047329994 | 0.38 |
| SRRM1 | 2.666 | 0.007682 | 0.002089813 | 1 | 1 | 1 | 0.007682 | 0.06 |
| MRPL37 | -2.665 | 0.00770134 | 0.005311998 | 2 | 2 | 2 | 0.008147998 | 0.15 |

| | | | | | | | | |
|---|---|---|---|---|---|---|---|---|
| SP5 | 2.662 | 0.007779 | 0.001045724 | 1 | 1 | 1 | 0.007778999 | 0.05 |
| PPARGC1B | 2.661 | 0.007796 | 0.003715692 | 1 | 1 | 1 | 0.007796 | 0.10 |
| CXCL14 | 2.660 | 0.007822 | 0.011190368 | 1 | 1 | 1 | 0.007821999 | 0.18 |
| ZNF184 | -2.659 | 0.00784 | 0.010149137 | 1 | 1 | 1 | 0.00784 | 0.16 |
| FADS2 | -2.656 | 0.007914 | 0.130014423 | 1 | 1 | 1 | 0.007914002 | 0.70 |
| Z83851.4 | -2.654 | 0.00796033 | 0.037572936 | 2 | 2 | 2 | 0.001183 | 0.20 |
| CCS | 2.654 | 0.007966 | 0.003973834 | 1 | 2 | 2 | 0.007966001 | 0.11 |
| TNIP2 | -2.653 | 0.00797 | 0.01270721 | 1 | 2 | 2 | 0.007969997 | 0.41 |
| REEP1 | -2.653 | 0.00797072 | 0.004912603 | 2 | 2 | 2 | 0.019010002 | 0.22 |
| LRRN3 | 2.652 | 0.00801211 | 0.017304791 | 2 | 2 | 2 | 0.023889996 | 0.24 |
| PML | 2.651 | 0.00802771 | 0.000526945 | 2 | 2 | 2 | 0.010240003 | 0.05 |
| TOX2 | 2.649 | 0.00806592 | 0.051126343 | 2 | 2 | 2 | 0.005354999 | 0.29 |
| RP11-539G18.3 | 2.649 | 0.008073 | 0.003001918 | 1 | 1 | 1 | 0.008073001 | 0.08 |
| TMOD3 | -2.647 | 0.008121 | 0.000905782 | 1 | 1 | 1 | 0.008121002 | 0.09 |
| KAT8 | 2.646 | 0.00815301 | 0.146485912 | 2 | 2 | 2 | 0.005280998 | 0.34 |
| SCG3 | -2.641 | 0.008258 | 0.030235447 | 1 | 1 | 1 | 0.008258002 | 0.26 |
| AKT3 | 2.638 | 0.008337 | 1.55609E-06 | 1 | 1 | 1 | 0.008337002 | 0.01 |
| MTSS1 | 2.638 | 0.00834111 | 0.04696218 | 2 | 2 | 2 | 0.025650005 | 0.23 |
| DKFZp779M0652 | -2.637 | 0.00835901 | 0.23401825 | 4 | 4 | 4 | 0.004261999 | 0.61 |
| C11orf58 | 2.637 | 0.00837058 | 0.00034588 | 2 | 2 | 2 | 0.013419996 | 0.03 |
| AC012442.6 | -2.636 | 0.00838 | 0.015292696 | 1 | 1 | 1 | 0.008380002 | 0.23 |
| KBTBD3 | 2.636 | 0.00839542 | 0.033465166 | 2 | 2 | 2 | 0.006749999 | 0.19 |
| CEBPZOS | -2.634 | 0.00842809 | 0.144870941 | 3 | 4 | 4 | 0.001829 | 0.33 |
| TP53BP1 | 2.634 | 0.008431 | 0.005579803 | 1 | 1 | 1 | 0.008431001 | 0.16 |
| MAP1A | -2.634 | 0.008431 | 0.004282659 | 1 | 1 | 1 | 0.008431001 | 0.14 |
| ZNF90 | -2.632 | 0.008491 | 0.054889519 | 1 | 3 | 3 | 0.008491002 | 0.53 |
| ABRACL | 2.630 | 0.00852646 | 0.053079305 | 4 | 4 | 4 | 0.024579997 | 0.46 |
| RP11-700H6.1 | 2.630 | 0.008551 | 0.07447022 | 1 | 1 | 1 | 0.008551001 | 0.44 |
| ACTN2 | 2.628 | 0.00857845 | 0.016744296 | 2 | 2 | 2 | 0.0101 | 0.22 |
| REEP5 | 2.627 | 0.008607 | 0.004216791 | 1 | 2 | 2 | 0.008607 | 0.09 |
| CNPPD1 | -2.626 | 0.008633 | 0.001019115 | 1 | 3 | 3 | 0.008632999 | 0.13 |
| TGFBI | -2.625 | 0.008669 | 4.53337E-06 | 1 | 1 | 1 | 0.008668999 | 0.01 |
| RNF13 | -2.623 | 0.00871917 | 0.00520049 | 2 | 2 | 2 | 0.027940004 | 0.10 |
| RNF5 | -2.622 | 0.00873997 | 0.03668114 | 2 | 2 | 2 | 0.005100999 | 0.31 |
| GPR63 | 2.621 | 0.00875503 | 0.000120768 | 2 | 3 | 3 | 0.018730002 | 0.03 |
| ST6GALNAC4 | 2.620 | 0.00879 | 0.003819412 | 1 | 2 | 2 | 0.008789999 | 0.15 |
| PABPC1 | 2.619 | 0.008825 | 0.001861053 | 1 | 1 | 1 | 0.008824998 | 0.08 |

| | | | | | | | | |
|---|---|---|---|---|---|---|---|---|
| JKAMP | -2.616 | 0.00890858 | 0.012290948 | 2 | 2 | 2 | 0.011560002 | 0.13 |
| LCMT2 | 2.615 | 0.00893342 | 0.014575961 | 2 | 2 | 2 | 0.010400003 | 0.13 |
| ATF6B | -2.611 | 0.00903 | 0.027546273 | 1 | 1 | 1 | 0.009029997 | 0.26 |
| TATDN1 | 2.610 | 0.009044 | 0.001486448 | 1 | 1 | 1 | 0.009044001 | 0.06 |
| LINC00242 | 2.608 | 0.009095 | 0.01110207 | 1 | 1 | 1 | 0.009095002 | 0.19 |
| LDHAL6A | 2.608 | 0.00910212 | 0.075380894 | 2 | 2 | 2 | 0.027320005 | 0.44 |
| CTD-3162L10.1 | 2.607 | 0.009125 | 0.016966685 | 1 | 2 | 2 | 0.009125001 | 0.19 |
| NLN | 2.605 | 0.00919082 | 0.013651677 | 2 | 2 | 2 | 0.007317002 | 0.17 |
| ALDH1A1 | -2.604 | 0.009226 | 0.001295782 | 1 | 1 | 1 | 0.009225997 | 0.05 |
| MANEA | -2.601 | 0.00929317 | 0.028361668 | 3 | 3 | 3 | 0.038689993 | 0.36 |
| AGBL2 | 2.600 | 0.009312 | 0.004280214 | 1 | 1 | 1 | 0.009312001 | 0.13 |
| ME1 | -2.600 | 0.009316 | 4.19574E-06 | 1 | 1 | 1 | 0.009316001 | 0.01 |
| ZNF518A | 2.600 | 0.00931991 | 0.005352297 | 3 | 3 | 3 | 0.002966001 | 0.09 |
| ZNF469 | -2.600 | 0.009322 | 3.99256E-07 | 1 | 2 | 2 | 0.009322003 | 0.00 |
| ACAD9 | -2.599 | 0.009343 | 0.011236872 | 1 | 3 | 3 | 0.009343002 | 0.22 |
| PC | 2.599 | 0.009356 | 6.64498E-06 | 1 | 1 | 1 | 0.009356002 | 0.01 |
| ABHD8 | 2.596 | 0.00943753 | 0.028311703 | 2 | 2 | 2 | 0.025530002 | 0.17 |
| KANK3 | 2.595 | 0.009452 | 0.006339059 | 1 | 1 | 1 | 0.009452 | 0.12 |
| CD320 | 2.594 | 0.009486 | 0.004460141 | 1 | 2 | 2 | 0.009486001 | 0.10 |
| ANO2 | -2.593 | 0.00952612 | 0.072344563 | 3 | 3 | 3 | 0.028660002 | 0.30 |
| PRRC2A | -2.586 | 0.00969962 | 0.013262548 | 2 | 2 | 2 | 0.007708002 | 0.09 |
| GABPB2 | 2.586 | 0.00970319 | 0.028914404 | 2 | 2 | 2 | 0.003304001 | 0.19 |
| EGLN1 | -2.586 | 0.009714 | 0.000307569 | 1 | 1 | 1 | 0.009714 | 0.04 |
| NDC80 | 2.585 | 0.00972867 | 0.014222938 | 2 | 2 | 2 | 0.011679998 | 0.10 |
| RP1-60O19.2 | 2.584 | 0.00976018 | 0.180747637 | 3 | 4 | 4 | 0.020080003 | 0.47 |
| ZNF576 | -2.583 | 0.009797 | 0.000977944 | 1 | 1 | 1 | 0.009796998 | 0.05 |
| ZNHIT3 | 2.582 | 0.009814 | 0.013326526 | 1 | 1 | 1 | 0.009813997 | 0.17 |
| ABHD17C | 2.582 | 0.009826 | 0.015910494 | 1 | 1 | 1 | 0.009825999 | 0.19 |
| ARSK | 2.580 | 0.00988466 | 0.005635342 | 2 | 2 | 2 | 0.009647999 | 0.09 |
| BRD3 | 2.579 | 0.00990097 | 0.052237082 | 2 | 2 | 2 | 0.007484999 | 0.23 |
| FAM120A | 2.579 | 0.00992 | 6.04515E-05 | 1 | 1 | 1 | 0.009920003 | 0.01 |
| B9D1 | -2.577 | 0.009975 | 3.26591E-05 | 1 | 1 | 1 | 0.009975 | 0.01 |
| RUVBL1 | 2.577 | 0.00997717 | 0.005894205 | 2 | 2 | 2 | 0.029699995 | 0.14 |
| SLC37A2 | 2.576 | 0.009987 | 0.005262671 | 1 | 2 | 2 | 0.009986998 | 0.12 |

zscore: S-PrediXcan's association result for the gene, typically HUGO for a gene; pvalue, significance p-value of S-PrediXcan association; var_g: variance of the gene expression, calculated as W' * G * W (where W is the vector of SNP weights in a gene's model, W' is its transpose, and G is the covariance matrix); n_snps_used: number of snps from GWAS that got used in S-PrediXcan analysis; n_snps_in_cov, number of snps in the covariance matrix; n_snps_in_model, number of snps in the model; best_gwas_p, the highest p-value from GWAS snps used in this model; largest_weight: the largest (absolute value) weight in this model.

**Table S16**. TWAS-identified genes associated with anorexia nervosa in gastrointestinal tissues.

| gene_name | pvalue | n | n_indep | p_i_best | t_i_best | eigen_max | eigen_min | eigen_min_kept | z_min | z_max | z_mean | z_sd | tmi |
|---|---|---|---|---|---|---|---|---|---|---|---|---|---|
| CELSR3 | 4.74E-14 | 23 | 2 | 9.25E-14 | Colon_Sigmoid | 3.61 | 6.05E-17 | 0.39 | -7.45 | 7.47 | 3.73 | 5.76 | 2 |
| USP19 | 4.26E-13 | 10 | 2 | 7.55E-13 | Colon_Sigmoid | 1.47 | 5.34E-01 | 0.53 | -7.17 | 7.17 | 3.61 | 5.30 | 2 |
| DALRD3 | 6.67E-13 | 36 | 2 | 2.72E-06 | Esophagus_Mucosa | 3.68 | 3.19E-18 | 0.32 | -1.51 | 7.17 | 2.40 | 2.52 | 2 |
| KLHDC8B | 7.33E-13 | 49 | 2 | 3.95E-10 | Esophagus_Mucosa | 5.42 | 5.10E-17 | 1.41 | -7.13 | 7.16 | -3.37 | 3.95 | 2 |
| CAMKV | 1.35E-12 | 16 | 2 | 3.78E-13 | Esophagus_Muscularis | 3.10 | 6.01E-17 | 0.90 | -3.16 | 7.26 | 3.09 | 2.78 | 2 |
| PRKAR2A | 1.36E-12 | 49 | 2 | 4.65E-13 | Colon_Transverse | 5.44 | 1.94E-17 | 1.52 | -7.24 | 7.24 | 4.92 | 3.73 | 2 |
| RP11-3B7.1 | 1.88E-12 | 31 | 2 | 3.78E-12 | Colon_Transverse | 6.64 | 2.14E-17 | 0.36 | 4.32 | 7.07 | 5.58 | 0.94 | 2 |
| RP11-572O6.1 | 2.13E-12 | 42 | 3 | 3.79E-14 | Stomach | 3.74 | 1.34E-16 | 0.31 | -1.11 | 7.61 | 3.23 | 3.60 | 3 |
| P4HTM | 5.07E-12 | 41 | 2 | 2.72E-06 | Esophagus_Muscularis | 4.65 | 3.72E-17 | 0.35 | 0.04 | 5.09 | 2.88 | 2.08 | 2 |
| C3orf62 | 7.20E-12 | 46 | 1 | 7.20E-12 | Esophagus_Gastroesophageal_Junction | 7.00 | 7.33E-17 | 7.00 | 0.71 | 6.85 | 6.43 | 1.36 | 1 |
| WDR6 | 2.26E-10 | 49 | 1 | 1.41E-11 | Esophagus_Mucosa | 6.83 | 8.33E-17 | 6.83 | -7.03 | -4.68 | -6.26 | 0.73 | 1 |
| NME6 | 1.38E-09 | 48 | 2 | 1.01E-05 | Esophagus_Mucosa | 6.68 | 1.96E-17 | 0.29 | -4.42 | 1.00 | -1.03 | 0.58 | 2 |
| NCKIPSD | 2.89E-07 | 49 | 1 | 2.55E-08 | Colon_Transverse | 6.79 | 1.30E-17 | 6.79 | 3.34 | 5.60 | 5.12 | 0.57 | 1 |
| APEH | 3.10E-07 | 39 | 3 | 1.00E-06 | Esophagus_Gastroesophageal_Junction | 4.59 | 1.09E-16 | 0.35 | -5.05 | 0.31 | -3.50 | 1.67 | 3 |
| DAG1 | 1.22E-06 | 25 | 2 | 1.35E-06 | Colon_Transverse | 4.76 | 1.76E-17 | 1.23 | -5.17 | 0.86 | -4.10 | 1.69 | 2 |
| BSN-AS2 | 1.31E-06 | 32 | 2 | 1.48E-06 | Esophagus_Muscularis | 2.47 | 8.98E-33 | 0.53 | -5.20 | -0.84 | -4.46 | 1.00 | 2 |
| CCDC36 | 1.63E-06 | 39 | 1 | 4.91E-07 | Esophagus_Gastroesophageal_Junction | 6.94 | 2.18E-19 | 6.94 | 0.57 | 5.03 | 4.61 | 0.87 | 1 |
| ZMYND10 | 1.76E-06 | 11 | 2 | 2.73E-07 | Small_Intestine_Terminal_Ileum | 1.14 | 8.59E-01 | 0.86 | -5.14 | 5.14 | 1.89 | 3.56 | 2 |
| RHOA | 1.85E-06 | 38 | 2 | 1.50E-06 | Colon_Sigmoid | 4.52 | 1.71E-17 | 1.48 | -4.81 | 4.81 | 0.73 | 4.12 | 2 |
| FBXO27 | 1.93E-06 | 49 | 4 | 5.25E-05 | Small_Intestine_Terminal_Ileum | 3.26 | 4.73E-03 | 0.12 | -4.04 | 4.12 | -0.80 | 2.11 | 4 |
| ARIH2OS | 2.00E-06 | 28 | 1 | 1.88E-06 | Colon_Sigmoid | 6.00 | 2.94E-18 | 6.00 | -4.77 | -4.74 | -4.75 | 0.01 | 1 |
| ARIH2 | 2.05E-06 | 37 | 1 | 1.88E-06 | Colon_Transverse | 7.00 | 2.27E-49 | 7.00 | 4.74 | 4.93 | 4.76 | 0.03 | 1 |
| ACYP2 | 2.78E-06 | 49 | 4 | 6.84E-05 | Colon_Sigmoid | 4.73 | 3.23E-03 | 0.18 | -3.41 | 4.84 | 1.31 | 2.06 | 4 |
| LINC00324 | 3.37E-06 | 28 | 3 | 3.72E-07 | Esophagus_Mucosa | 3.36 | 6.96E-17 | 0.15 | -5.08 | -0.19 | -2.70 | 2.20 | 3 |
| TCTA | 3.50E-06 | 49 | 2 | 1.19E-05 | Esophagus_Gastroesophageal_Junction | 6.44 | 4.44E-18 | 0.56 | -4.39 | 4.46 | 0.83 | 1.68 | 2 |
| FAM212A | 3.64E-06 | 43 | 3 | 4.96E-05 | Stomach | 3.67 | 7.35E-19 | 0.34 | -4.12 | 4.59 | -0.25 | 2.82 | 3 |
| CAAP1 | 3.90E-06 | 34 | 4 | 2.28E-03 | Colon_Sigmoid | 4.18 | 2.09E-03 | 0.16 | -3.16 | 1.03 | -1.09 | 1.03 | 4 |
| PSME4 | 5.37E-06 | 48 | 5 | 6.88E-05 | Stomach | 3.67 | 1.92E-16 | 0.17 | -3.43 | 3.98 | 0.11 | 2.50 | 5 |

| | | | | | | | | | | | | | |
|---|---|---|---|---|---|---|---|---|---|---|---|---|---|
| PTBP2 | 5.42E-06 | 49 | 2 | 6.54E-05 | Esophagus_Muscularis | 6.35 | 1.01E-16 | 0.49 | -0.29 | 4.68 | 1.61 | 1.18 | 2 |
| CNPY4 | 1.15E-05 | 43 | 3 | 6.63E-07 | Esophagus_Muscularis | 6.01 | 6.71E-17 | 0.27 | -1.87 | 5.05 | 2.67 | 2.22 | 3 |
| CTNNB1 | 1.17E-05 | 42 | 1 | 5.77E-06 | Colon_Sigmoid | 5.79 | 1.18E-18 | 5.79 | -4.24 | 4.53 | 3.72 | 2.01 | 1 |
| ZEB2 | 1.18E-05 | 1 | 1 | 1.18E-05 | Stomach | 1.00 | 1.00E+00 | 1.00 | 4.38 | 4.38 | 4.38 | NA | 1 |
| SUOX | 1.85E-05 | 48 | 1 | 5.80E-06 | Esophagus_Gastroesophageal_Junction | 6.96 | 6.78E-17 | 6.96 | 4.03 | 4.68 | 4.25 | 0.12 | 1 |
| ASB3 | 1.92E-05 | 47 | 3 | 9.41E-06 | Esophagus_Gastroesophageal_Junction | 5.20 | 1.65E-17 | 0.18 | -4.99 | 4.95 | -0.11 | 3.32 | 3 |
| MGMT | 2.30E-05 | 48 | 2 | 9.07E-06 | Colon_Transverse | 6.40 | 5.12E-10 | 0.26 | -0.63 | 4.68 | 3.80 | 0.88 | 2 |
| ERBB3 | 2.60E-05 | 22 | 1 | 2.60E-05 | Colon_Transverse | 3.00 | 2.11E-48 | 3.00 | -4.26 | 4.21 | 2.29 | 3.61 | 1 |
| ENPP5 | 2.69E-05 | 43 | 4 | 1.51E-03 | Esophagus_Muscularis | 2.90 | 6.71E-03 | 0.27 | -3.42 | 2.60 | -1.38 | 1.45 | 4 |
| VWA7 | 3.53E-05 | 48 | 4 | 1.18E-05 | Stomach | 3.57 | 2.08E-16 | 0.20 | -4.38 | 1.95 | -0.64 | 2.10 | 4 |
| RP11-477N3.1 | 3.58E-05 | 48 | 4 | 8.59E-04 | Colon_Sigmoid | 5.45 | 2.06E-03 | 0.32 | -4.78 | 1.23 | -2.69 | 1.25 | 4 |
| MON1A | 3.76E-05 | 9 | 1 | 3.76E-05 | Colon_Sigmoid | 2.00 | 3.35E-17 | 2.00 | -4.12 | 0.78 | -3.06 | 1.83 | 1 |
| TRAIP | 3.77E-05 | 33 | 1 | 3.77E-05 | Esophagus_Gastroesophageal_Junction | 4.00 | 3.93E-80 | 4.00 | -4.12 | 4.12 | 3.87 | 1.43 | 1 |
| RBM5 | 3.79E-05 | 24 | 2 | 8.44E-05 | Colon_Sigmoid | 4.73 | 3.77E-17 | 1.27 | -3.93 | -0.05 | -3.39 | 1.32 | 2 |
| RP11-396B14.2 | 4.07E-05 | 43 | 4 | 3.25E-04 | Colon_Sigmoid | 3.48 | 1.77E-16 | 0.14 | -3.59 | 2.07 | -0.69 | 1.30 | 4 |
| IP6K1 | 4.41E-05 | 19 | 2 | 5.30E-05 | Esophagus_Mucosa | 4.28 | 2.18E-17 | 0.71 | 0.23 | 4.04 | 1.40 | 1.68 | 2 |
| RBM6 | 4.55E-05 | 48 | 1 | 3.72E-05 | Colon_Sigmoid | 6.96 | 4.88E-19 | 6.96 | -4.16 | -3.71 | -4.13 | 0.06 | 1 |
| GPR75 | 4.60E-05 | 45 | 4 | 1.31E-04 | Colon_Transverse | 4.72 | 4.64E-18 | 0.24 | -4.77 | 5.53 | -0.65 | 2.55 | 4 |
| ZC3H8 | 5.29E-05 | 41 | 1 | 5.16E-05 | Colon_Transverse | 5.86 | 5.84E-17 | 5.86 | -4.12 | 4.10 | 2.98 | 2.02 | 1 |
| PXDNL | 5.36E-05 | 32 | 1 | 5.36E-05 | Colon_Transverse | 3.00 | 3.85E-17 | 3.00 | -4.04 | 4.04 | 2.18 | 2.53 | 1 |
| C4A | 5.68E-05 | 49 | 1 | 2.25E-05 | Esophagus_Gastroesophageal_Junction | 6.75 | 1.29E-17 | 6.75 | -4.32 | -2.76 | -4.01 | 0.30 | 1 |
| KCNH3 | 5.75E-05 | 13 | 2 | 1.06E-05 | Esophagus_Muscularis | 2.75 | 5.96E-35 | 0.25 | -3.46 | 4.40 | 2.18 | 3.13 | 2 |
| CALML6 | 5.93E-05 | 49 | 3 | 4.90E-03 | Esophagus_Gastroesophageal_Junction | 5.20 | 3.52E-09 | 0.21 | -3.36 | 4.50 | 0.79 | 2.25 | 3 |
| SOX5 | 6.36E-05 | 13 | 1 | 6.36E-05 | Colon_Transverse | 1.00 | 1.00E+00 | 1.00 | -4.00 | 3.81 | -2.50 | 2.66 | 1 |
| PTPMT1 | 6.42E-05 | 49 | 3 | 1.20E-04 | Small_Intestine_Terminal_Ileum | 5.03 | 1.05E-16 | 0.50 | -2.17 | 3.85 | -1.15 | 1.66 | 3 |
| RNF123 | 7.93E-05 | 41 | 1 | 7.75E-05 | Colon_Transverse | 5.00 | 1.84E-19 | 5.00 | 3.92 | 3.97 | 3.95 | 0.01 | 1 |
| PDE4B | 8.01E-05 | 30 | 3 | 4.94E-05 | Stomach | 1.76 | 3.25E-01 | 0.32 | -4.06 | 2.31 | -1.31 | 1.54 | 3 |
| ACTN2 | 8.07E-05 | 49 | 5 | 3.97E-06 | Stomach | 3.96 | 5.61E-18 | 0.24 | -1.59 | 4.61 | 1.18 | 1.40 | 5 |
| STAC3 | 8.52E-05 | 41 | 3 | 3.49E-04 | Colon_Transverse | 4.56 | 1.23E-17 | 0.49 | -4.26 | 1.12 | -3.09 | 1.27 | 3 |
| HLA-DMA | 8.98E-05 | 48 | 1 | 8.53E-05 | Stomach | 5.87 | 5.56E-05 | 5.87 | -3.93 | 4.33 | 3.33 | 1.61 | 1 |
| TMEM52 | 9.16E-05 | 45 | 2 | 1.49E-05 | Colon_Sigmoid | 5.04 | 4.02E-17 | 1.92 | -4.33 | 4.35 | 1.44 | 2.09 | 2 |
| SLC12A5 | 1.01E-04 | 40 | 2 | 2.33E-05 | Esophagus_Gastroesophageal_Junction | 5.14 | 1.24E-17 | 0.78 | -4.23 | 4.18 | -1.13 | 2.58 | 2 |
| PRUNE1 | 1.05E-04 | 43 | 3 | 6.22E-06 | Colon_Sigmoid | 3.79 | 7.46E-05 | 0.99 | -1.24 | 4.53 | 1.57 | 1.87 | 3 |
| MST1R | 1.05E-04 | 44 | 1 | 1.05E-04 | Colon_Sigmoid | 6.00 | 3.70E-33 | 6.00 | -3.88 | 4.32 | 3.56 | 1.52 | 1 |

| | | | | | | | | | | | | | |
|---|---|---|---|---|---|---|---|---|---|---|---|---|---|
| DHFR2 | 1.13E-04 | 33 | 1 | 1.14E-04 | Colon_Sigmoid | 6.98 | 4.14E-17 | 6.98 | -3.86 | 3.86 | -2.92 | 2.56 | 1 |
| NAT6 | 1.13E-04 | 48 | 2 | 2.32E-02 | Esophagus_Gastroesophageal_Junction | 5.56 | 7.73E-18 | 0.44 | -2.27 | 0.21 | -0.76 | 0.99 | 2 |
| MSH5 | 1.15E-04 | 46 | 5 | 7.33E-06 | Esophagus_Muscularis | 3.80 | 2.49E-16 | 0.22 | -4.73 | 0.77 | -3.11 | 1.51 | 5 |
| RP11-373D23.3 | 1.27E-04 | 45 | 2 | 7.56E-05 | Colon_Sigmoid | 4.72 | 1.96E-03 | 1.08 | -3.96 | 2.26 | 0.19 | 1.34 | 2 |
| MMP16 | 1.29E-04 | 42 | 2 | 1.94E-01 | Esophagus_Mucosa | 6.69 | 9.70E-18 | 0.22 | -1.41 | 3.05 | -0.01 | 1.02 | 2 |
| PRKCSH | 1.43E-04 | 33 | 2 | 2.25E-05 | Esophagus_Mucosa | 4.72 | 2.52E-17 | 0.27 | -4.31 | -0.38 | -3.91 | 0.70 | 2 |
| CTSA | 1.46E-04 | 36 | 2 | 4.61E-05 | Esophagus_Mucosa | 2.44 | 2.05E-16 | 0.56 | -4.07 | 3.31 | 0.81 | 1.51 | 2 |
| ARHGEF15 | 1.47E-04 | 13 | 1 | 1.47E-04 | Stomach | 1.00 | 1.00E+00 | 1.00 | 0.05 | 4.45 | 2.67 | 1.20 | 1 |
| APOM | 1.47E-04 | 43 | 2 | 2.57E-05 | Stomach | 6.01 | 1.34E-17 | 0.98 | -4.25 | 0.89 | -3.69 | 1.34 | 2 |
| SNX27 | 1.49E-04 | 33 | 1 | 9.41E-05 | Colon_Transverse | 2.93 | 6.23E-18 | 2.93 | -3.91 | -0.36 | -3.39 | 0.81 | 1 |
| STX19 | 1.51E-04 | 33 | 1 | 1.51E-04 | Colon_Sigmoid | 5.00 | 1.92E-17 | 5.00 | 2.60 | 3.89 | 3.76 | 0.21 | 1 |
| ERLEC1 | 1.55E-04 | 34 | 4 | 1.58E-02 | Esophagus_Muscularis | 3.11 | 4.71E-18 | 0.21 | -1.94 | 4.42 | 0.42 | 1.91 | 4 |
| ZFP91 | 1.57E-04 | 31 | 2 | 5.81E-04 | Colon_Sigmoid | 6.09 | 1.11E-16 | 0.91 | -3.44 | 3.44 | -2.69 | 1.59 | 2 |
| PFAS | 1.57E-04 | 37 | 4 | 8.12E-05 | Esophagus_Muscularis | 3.85 | 1.17E-16 | 0.20 | -1.18 | 4.53 | 0.75 | 1.88 | 4 |
| TOPBP1 | 1.64E-04 | 45 | 6 | 8.08E-05 | Esophagus_Gastroesophageal_Junction | 2.26 | 3.02E-02 | 0.22 | -2.43 | 3.94 | 0.97 | 1.76 | 6 |
| CHAC2 | 1.65E-04 | 39 | 4 | 3.07E-03 | Esophagus_Gastroesophageal_Junction | 3.71 | 4.80E-19 | 0.14 | -5.01 | 5.01 | 0.58 | 2.48 | 4 |
| CDHR4 | 1.74E-04 | 10 | 2 | 7.73E-05 | Esophagus_Mucosa | 1.42 | 5.79E-01 | 0.58 | -4.03 | 4.11 | 1.35 | 3.01 | 2 |
| BORCS6 | 1.75E-04 | 26 | 3 | 3.04E-05 | Esophagus_Muscularis | 3.13 | 1.84E-17 | 0.62 | -2.65 | 4.49 | 1.73 | 1.80 | 3 |
| ENO4 | 1.76E-04 | 2 | 1 | 1.76E-04 | Colon_Transverse | 1.00 | 1.00E+00 | 1.00 | -1.19 | 3.75 | 1.28 | 3.49 | 1 |
| SYN2 | 1.88E-04 | 45 | 2 | 3.45E-05 | Esophagus_Muscularis | 5.66 | 2.43E-18 | 0.28 | -4.14 | 4.00 | -2.51 | 2.20 | 2 |
| IBTK | 1.88E-04 | 48 | 3 | 8.48E-02 | Esophagus_Mucosa | 4.88 | 1.17E-16 | 0.17 | -2.73 | 3.41 | -0.44 | 1.83 | 3 |
| SELENOH | 1.89E-04 | 29 | 1 | 1.92E-04 | Esophagus_Muscularis | 3.92 | 3.11E-19 | 3.92 | 3.69 | 3.79 | 3.69 | 0.02 | 1 |
| DNAJC22 | 2.03E-04 | 43 | 2 | 1.22E-04 | Esophagus_Muscularis | 5.29 | 3.37E-05 | 0.59 | -1.87 | 4.51 | 2.80 | 1.39 | 2 |
| SYNCRIP | 2.07E-04 | 25 | 1 | 2.07E-04 | Colon_Transverse | 5.00 | 4.66E-34 | 5.00 | -3.71 | 3.74 | -1.01 | 3.62 | 1 |
| RNF2 | 2.09E-04 | 18 | 2 | 7.22E-05 | Esophagus_Mucosa | 1.62 | 3.77E-01 | 0.38 | -3.97 | 0.08 | -3.18 | 1.10 | 2 |
| CCDC93 | 2.16E-04 | 49 | 4 | 9.81E-03 | Stomach | 4.81 | 2.17E-17 | 0.22 | -3.42 | 2.40 | -1.07 | 1.47 | 4 |
| ENPP4 | 2.21E-04 | 49 | 3 | 1.48E-03 | Esophagus_Muscularis | 5.85 | 6.22E-17 | 0.28 | -3.24 | 3.09 | -1.39 | 1.95 | 3 |
| SCUBE1 | 2.48E-04 | 34 | 4 | 4.05E-04 | Esophagus_Muscularis | 2.06 | 3.12E-02 | 0.51 | -3.54 | 2.30 | -1.35 | 1.81 | 4 |
| USP45 | 2.61E-04 | 37 | 4 | 3.52E-04 | Esophagus_Muscularis | 3.21 | 1.22E-17 | 0.58 | -3.57 | 2.35 | -1.00 | 1.80 | 4 |
| RGPD8 | 2.63E-04 | 47 | 2 | 2.47E-04 | Small_Intestine_Terminal_Ileum | 6.62 | 7.68E-17 | 0.30 | -3.75 | -2.36 | -3.25 | 0.57 | 2 |
| JRK | 2.66E-04 | 44 | 5 | 3.22E-05 | Esophagus_Mucosa | 2.85 | 8.10E-17 | 0.24 | -4.31 | 3.14 | -1.48 | 2.23 | 5 |
| BNIPL | 2.73E-04 | 45 | 2 | 7.73E-05 | Colon_Sigmoid | 6.11 | 9.72E-17 | 0.73 | 1.89 | 4.06 | 3.34 | 0.62 | 2 |
| FAM180B | 2.73E-04 | 45 | 3 | 1.03E-03 | Small_Intestine_Terminal_Ileum | 5.03 | 2.27E-17 | 0.53 | -3.35 | 4.24 | 0.37 | 2.43 | 3 |
| ALDH16A1 | 2.77E-04 | 48 | 2 | 1.60E-04 | Esophagus_Mucosa | 6.32 | 1.40E-16 | 0.52 | -4.06 | -0.26 | -2.70 | 0.83 | 2 |
| FOXA2 | 2.80E-04 | 13 | 1 | 2.80E-04 | Colon_Sigmoid | 4.00 | 3.28E-33 | 4.00 | -3.63 | 3.63 | -3.07 | 2.02 | 1 |
| AURKB | 2.95E-04 | 16 | 2 | 2.63E-04 | Esophagus_Muscularis | 1.18 | 8.19E-01 | 0.82 | -4.57 | 1.03 | -0.60 | 2.25 | 2 |

| | | | | | | | | | | | | | |
|---|---|---|---|---|---|---|---|---|---|---|---|---|---|
| ABHD8 | 3.00E-04 | 35 | 4 | 5.16E-02 | Colon_Sigmoid | 4.54 | 8.22E-17 | 0.28 | -1.95 | 2.65 | 0.75 | 1.49 | 4 |
| AP3M1 | 3.11E-04 | 46 | 2 | 1.50E-05 | Esophagus_Muscularis | 5.19 | 8.64E-18 | 0.68 | -4.33 | 2.55 | 0.62 | 1.85 | 2 |
| CCDC34 | 3.12E-04 | 10 | 1 | 3.12E-04 | Stomach | 1.00 | 1.00E+00 | 1.00 | 0.15 | 3.60 | 1.39 | 1.65 | 1 |
| KNOP1 | 3.48E-04 | 44 | 1 | 3.65E-04 | Colon_Transverse | 3.95 | 1.50E-18 | 3.95 | -3.65 | -3.46 | -3.52 | 0.04 | 1 |
| CDC73 | 3.51E-04 | 35 | 1 | 2.33E-04 | Colon_Transverse | 4.94 | 1.11E-79 | 4.94 | -5.15 | -0.05 | -3.49 | 0.66 | 1 |
| C6orf48 | 3.51E-04 | 49 | 3 | 1.74E-02 | Esophagus_Muscularis | 5.78 | 4.93E-02 | 0.25 | -4.42 | 1.62 | -0.96 | 1.45 | 3 |
| IP6K2 | 3.55E-04 | 26 | 3 | 1.46E-04 | Esophagus_Muscularis | 1.51 | 5.36E-01 | 0.54 | -1.05 | 6.77 | 0.70 | 2.32 | 3 |
| CDC27 | 3.91E-04 | 43 | 4 | 4.08E-05 | Stomach | 2.94 | 1.83E-17 | 0.50 | -4.10 | 3.53 | -1.30 | 2.21 | 4 |
| GDNF | 3.97E-04 | 19 | 1 | 3.97E-04 | Esophagus_Gastroesophageal_Junction | 2.00 | 3.35E-17 | 2.00 | -3.54 | 3.54 | -0.33 | 2.95 | 1 |
| ZNF335 | 4.01E-04 | 43 | 4 | 1.68E-04 | Stomach | 2.65 | 6.62E-17 | 0.94 | -3.79 | 3.76 | -0.68 | 1.88 | 4 |
| EBF3 | 4.10E-04 | 29 | 2 | 7.84E-05 | Esophagus_Mucosa | 1.05 | 9.48E-01 | 0.95 | 0.16 | 3.95 | 0.73 | 0.91 | 2 |
| IQCK | 4.16E-04 | 24 | 1 | 4.16E-04 | Colon_Transverse | 1.00 | 1.00E+00 | 1.00 | -3.53 | 3.53 | -2.03 | 2.76 | 1 |
| BANK1 | 4.17E-04 | 5 | 1 | 4.17E-04 | Esophagus_Muscularis | 1.00 | 1.00E+00 | 1.00 | -3.53 | 1.12 | -0.85 | 1.68 | 1 |
| PHF2 | 4.19E-04 | 21 | 2 | 3.76E-04 | Stomach | 5.02 | 9.38E-19 | 1.83 | -2.94 | 3.56 | 2.34 | 2.11 | 2 |
| TMEM107 | 4.29E-04 | 44 | 3 | 8.27E-04 | Esophagus_Mucosa | 4.43 | 2.19E-17 | 0.79 | -3.87 | 1.20 | -1.09 | 1.69 | 3 |
| KMT2B | 4.30E-04 | 17 | 1 | 4.30E-04 | Esophagus_Muscularis | 1.00 | 1.00E+00 | 1.00 | -1.40 | 3.52 | 1.17 | 2.06 | 1 |
| SNX14 | 4.35E-04 | 44 | 4 | 9.86E-05 | Esophagus_Gastroesophageal_Junction | 2.75 | 4.69E-17 | 0.58 | -2.44 | 3.89 | 0.23 | 1.87 | 4 |
| RP11-436K8.1 | 4.47E-04 | 10 | 2 | 1.23E-04 | Small_Intestine_Terminal_Ileum | 1.34 | 6.61E-01 | 0.66 | -3.84 | 1.62 | -1.37 | 1.68 | 2 |
| PRR7 | 4.49E-04 | 26 | 1 | 3.82E-04 | Colon_Transverse | 5.91 | 1.99E-17 | 5.91 | -3.55 | 3.55 | 1.86 | 3.00 | 1 |
| PIH1D1 | 4.71E-04 | 39 | 3 | 3.78E-04 | Stomach | 5.42 | 7.47E-18 | 0.25 | -3.94 | 3.59 | -1.01 | 2.37 | 3 |
| ZBTB1 | 4.86E-04 | 45 | 1 | 4.86E-04 | Colon_Transverse | 6.00 | 2.43E-33 | 6.00 | -2.73 | 3.60 | 3.22 | 1.03 | 1 |
| RP11-227D13.1 | 4.91E-04 | 35 | 1 | 2.35E-04 | Colon_Sigmoid | 6.92 | 2.32E-18 | 6.92 | -3.68 | -0.40 | -3.30 | 0.54 | 1 |
| PLTP | 4.99E-04 | 46 | 5 | 9.33E-04 | Stomach | 3.21 | 1.84E-02 | 0.17 | -3.49 | 4.09 | 1.28 | 2.18 | 5 |
| SMARCAL1 | 5.01E-04 | 47 | 2 | 1.14E-04 | Esophagus_Muscularis | 5.91 | 4.73E-17 | 0.88 | -4.03 | 1.59 | -2.80 | 1.34 | 2 |
| ADK | 5.02E-04 | 46 | 3 | 4.01E-05 | Colon_Sigmoid | 4.51 | 7.01E-17 | 0.45 | -4.44 | 3.94 | -0.54 | 2.66 | 3 |
| PIAS2 | 5.03E-04 | 38 | 1 | 5.16E-04 | Esophagus_Gastroesophageal_Junction | 5.85 | 3.12E-33 | 5.85 | -3.47 | 0.25 | -2.98 | 0.93 | 1 |
| SLC9A8 | 5.07E-04 | 48 | 4 | 2.28E-05 | Colon_Transverse | 4.18 | 7.11E-04 | 0.19 | -4.24 | 0.42 | -2.60 | 0.97 | 4 |
| LGR4 | 5.13E-04 | 46 | 2 | 1.25E-04 | Small_Intestine_Terminal_Ileum | 4.82 | 3.19E-17 | 2.03 | -3.84 | 3.82 | 0.91 | 2.36 | 2 |
| NDUFS3 | 5.23E-04 | 43 | 2 | 2.96E-03 | Colon_Transverse | 4.32 | 5.57E-18 | 1.68 | -1.54 | 3.51 | 2.45 | 1.10 | 2 |
| ACTG1 | 5.26E-04 | 28 | 2 | 7.89E-05 | Colon_Transverse | 3.69 | 2.13E-17 | 1.21 | -3.95 | 3.95 | 2.28 | 2.48 | 2 |
| DYNC1I2 | 5.30E-04 | 39 | 1 | 4.44E-04 | Esophagus_Gastroesophageal_Junction | 6.96 | 4.91E-17 | 6.96 | -3.51 | 3.63 | 2.54 | 1.86 | 1 |
| GPANK1 | 5.44E-04 | 41 | 6 | 2.05E-05 | Esophagus_Muscularis | 3.82 | 7.84E-17 | 0.18 | -4.26 | 1.43 | -1.77 | 1.69 | 6 |
| IKZF4 | 5.51E-04 | 32 | 3 | 4.68E-05 | Stomach | 3.25 | 3.26E-18 | 0.50 | -4.34 | -0.09 | -2.48 | 1.62 | 3 |
| STK35 | 5.64E-04 | 47 | 5 | 2.30E-05 | Colon_Transverse | 2.58 | 1.23E-02 | 0.34 | -4.23 | 3.51 | 0.57 | 1.40 | 5 |
| WDR35 | 5.73E-04 | 49 | 3 | 9.01E-05 | Colon_Sigmoid | 5.65 | 2.00E-04 | 0.27 | -4.12 | -0.79 | -3.04 | 0.87 | 3 |

| | | | | | | | | | | | | | |
|---|---|---|---|---|---|---|---|---|---|---|---|---|---|
| HLA-DMB | 5.81E-04 | 34 | 3 | 2.98E-05 | Stomach | 3.86 | 5.26E-17 | 0.94 | -4.08 | 4.18 | 2.00 | 2.21 | 3 |
| RP11-11C20.3 | 5.81E-04 | 5 | 1 | 5.81E-04 | Colon_Transverse | 1.00 | 1.00E+00 | 1.00 | -3.44 | 0.43 | -2.64 | 1.71 | 1 |
| CNTLN | 5.84E-04 | 41 | 5 | 1.53E-05 | Esophagus_Mucosa | 4.15 | 2.97E-04 | 0.16 | -4.32 | 2.32 | -2.43 | 1.76 | 5 |
| SLC17A7 | 5.97E-04 | 35 | 3 | 4.20E-05 | Esophagus_Muscularis | 3.89 | 9.76E-18 | 0.39 | -3.29 | 4.10 | 1.74 | 2.76 | 3 |
| TRIM26 | 6.00E-04 | 46 | 4 | 1.00E-02 | Esophagus_Gastroesophageal_Junction | 3.61 | 2.51E-16 | 0.18 | -2.61 | 4.08 | 1.38 | 1.52 | 4 |
| TFDP2 | 6.16E-04 | 18 | 2 | 1.21E-04 | Esophagus_Mucosa | 1.14 | 8.61E-01 | 0.86 | -3.84 | 3.84 | -0.07 | 1.97 | 2 |
| PCMTD1 | 6.35E-04 | 49 | 4 | 1.63E-03 | Colon_Transverse | 4.73 | 8.29E-18 | 0.28 | -1.01 | 4.28 | 2.36 | 1.48 | 4 |
| MYO5A | 6.48E-04 | 45 | 4 | 1.56E-03 | Small_Intestine_Terminal_Ileum | 3.25 | 1.09E-17 | 0.24 | -3.81 | 3.72 | 0.48 | 2.20 | 4 |
| FAM57B | 6.63E-04 | 36 | 2 | 2.58E-03 | Colon_Sigmoid | 4.01 | 1.02E-16 | 0.88 | -2.19 | 3.01 | 1.02 | 2.20 | 2 |
| CDKN2AIP | 6.78E-04 | 25 | 3 | 2.28E-04 | Stomach | 2.98 | 8.14E-02 | 0.31 | -3.19 | 3.69 | 2.70 | 1.54 | 3 |
| MMP21 | 6.85E-04 | 19 | 2 | 3.13E-04 | Colon_Sigmoid | 1.53 | 4.72E-01 | 0.47 | -1.72 | 3.79 | 1.55 | 1.42 | 2 |
| NADK | 7.01E-04 | 44 | 5 | 6.33E-04 | Esophagus_Mucosa | 3.03 | 2.20E-17 | 0.33 | -3.38 | 3.42 | -0.55 | 2.02 | 5 |
| LY6G6C | 7.24E-04 | 36 | 2 | 1.92E-04 | Esophagus_Mucosa | 4.20 | 6.52E-33 | 0.80 | -4.31 | 3.80 | 0.57 | 1.56 | 2 |
| APBB1 | 7.32E-04 | 44 | 2 | 1.50E-04 | Colon_Transverse | 6.27 | 6.67E-18 | 0.71 | -3.79 | 2.16 | -2.02 | 1.64 | 2 |
| VAMP2 | 7.52E-04 | 42 | 2 | 1.39E-04 | Colon_Transverse | 6.61 | 6.00E-17 | 0.38 | -4.57 | 1.32 | -3.12 | 1.44 | 2 |
| SLC39A13 | 7.59E-04 | 47 | 2 | 2.48E-04 | Small_Intestine_Terminal_Ileum | 5.44 | 1.38E-17 | 1.56 | -3.66 | 3.68 | 1.22 | 1.67 | 2 |
| REPIN1 | 7.73E-04 | 41 | 2 | 1.77E-04 | Esophagus_Mucosa | 5.75 | 9.07E-18 | 1.10 | -1.41 | 3.75 | 1.29 | 0.71 | 2 |
| TMEM89 | 7.89E-04 | 21 | 2 | 2.43E-04 | Small_Intestine_Terminal_Ileum | 3.48 | 1.58E-16 | 0.52 | -6.57 | 3.67 | -3.24 | 1.80 | 2 |
| RP11-849F2.9 | 8.02E-04 | 43 | 5 | 2.49E-02 | Esophagus_Mucosa | 1.99 | 5.50E-02 | 0.29 | -2.78 | 2.95 | 0.71 | 1.44 | 5 |
| RAB5B | 8.10E-04 | 35 | 2 | 2.20E-04 | Esophagus_Mucosa | 4.04 | 8.97E-20 | 0.96 | -4.15 | 4.32 | -0.28 | 2.59 | 2 |
| SYCP3 | 8.10E-04 | 39 | 3 | 1.93E-03 | Esophagus_Muscularis | 5.52 | 1.55E-18 | 0.36 | -0.77 | 3.38 | 0.49 | 1.30 | 3 |
| ZMAT5 | 8.21E-04 | 34 | 3 | 3.69E-04 | Small_Intestine_Terminal_Ileum | 3.28 | 4.18E-18 | 0.81 | -3.56 | 0.82 | -0.55 | 1.01 | 3 |
| ZNF653 | 8.21E-04 | 42 | 1 | 8.21E-04 | Colon_Sigmoid | 5.00 | 8.84E-17 | 5.00 | -3.35 | 3.35 | 0.14 | 3.36 | 1 |
| AIF1 | 8.38E-04 | 44 | 4 | 2.33E-02 | Colon_Transverse | 3.04 | 1.64E-16 | 0.28 | -2.87 | 1.21 | -0.36 | 1.02 | 4 |
| GCSAM | 8.56E-04 | 38 | 2 | 1.29E-04 | Small_Intestine_Terminal_Ileum | 4.79 | 1.04E-16 | 2.19 | -0.54 | 3.83 | 1.94 | 1.66 | 2 |
| PNISR | 8.64E-04 | 39 | 2 | 2.26E-04 | Stomach | 3.83 | 1.45E-16 | 0.16 | -3.14 | 3.70 | 2.87 | 1.23 | 2 |
| AJUBA | 8.90E-04 | 46 | 2 | 1.00E-03 | Colon_Sigmoid | 5.34 | 1.96E-17 | 0.66 | -3.32 | 3.43 | 1.15 | 1.87 | 2 |
| TOMM34 | 8.93E-04 | 41 | 1 | 8.93E-04 | Colon_Sigmoid | 7.00 | 1.77E-17 | 7.00 | -3.36 | 3.32 | -2.40 | 2.00 | 1 |
| FAM163A | 8.98E-04 | 29 | 3 | 2.72E-03 | Stomach | 1.34 | 6.58E-01 | 0.66 | -3.00 | 3.00 | 0.99 | 1.71 | 3 |
| ZSCAN12 | 8.99E-04 | 44 | 3 | 8.61E-05 | Small_Intestine_Terminal_Ileum | 4.62 | 4.08E-17 | 0.25 | -3.75 | 3.93 | 1.25 | 2.57 | 3 |
| CLIC1 | 9.21E-04 | 46 | 5 | 1.69E-05 | Colon_Transverse | 2.04 | 1.07E-16 | 0.40 | -4.59 | 0.78 | -2.29 | 2.12 | 5 |
| COL11A2 | 9.28E-04 | 49 | 4 | 5.11E-03 | Small_Intestine_Terminal_Ileum | 5.38 | 1.15E-17 | 0.21 | -2.71 | 2.95 | 0.82 | 1.63 | 4 |
| AK2 | 9.41E-04 | 30 | 2 | 3.27E-04 | Colon_Sigmoid | 5.76 | 1.88E-17 | 0.24 | 2.51 | 3.59 | 2.72 | 0.36 | 2 |
| PENK | 9.65E-04 | 25 | 1 | 9.58E-04 | Esophagus_Gastroesophageal_Junction | 5.00 | 6.43E-17 | 5.00 | -1.81 | 3.30 | 2.74 | 1.19 | 1 |
| MSANTD4 | 9.72E-04 | 45 | 3 | 5.82E-03 | Esophagus_Gastroesophageal_Junction | 2.18 | 1.38E-18 | 0.86 | -2.67 | 2.76 | 0.74 | 1.20 | 3 |
| IKZF5 | 1.05E-03 | 45 | 3 | 6.13E-05 | Colon_Transverse | 4.98 | 5.61E-17 | 0.81 | -0.50 | 4.02 | 2.37 | 1.45 | 3 |

| | | | | | | | | | | | | | |
|---|---|---|---|---|---|---|---|---|---|---|---|---|---|
| NKAPL | 1.08E-03 | 40 | 3 | 3.09E-03 | Small_Intestine_Terminal_Ileum | 4.19 | 9.96E-17 | 0.41 | -0.43 | 2.96 | 0.47 | 0.90 | 3 |
| ITK | 1.09E-03 | 42 | 2 | 4.36E-04 | Esophagus_Mucosa | 6.00 | 2.43E-32 | 1.00 | -3.52 | 3.52 | 0.32 | 3.40 | 2 |
| LOXL4 | 1.10E-03 | 29 | 2 | 2.70E-04 | Esophagus_Muscularis | 5.33 | 3.62E-17 | 0.65 | -3.64 | 3.86 | -0.76 | 2.89 | 2 |
| ZKSCAN4 | 1.15E-03 | 42 | 2 | 3.58E-04 | Colon_Sigmoid | 6.69 | 4.22E-65 | 0.31 | -1.86 | 3.87 | 3.27 | 0.98 | 2 |
| GLRX2 | 1.17E-03 | 47 | 3 | 1.42E-02 | Colon_Sigmoid | 5.25 | 1.02E-18 | 0.20 | -4.22 | 3.41 | -1.35 | 1.72 | 3 |
| AP003774.6 | 1.18E-03 | 21 | 1 | 1.18E-03 | Small_Intestine_Terminal_Ileum | 1.00 | 1.00E+00 | 1.00 | -3.24 | 3.21 | -2.09 | 2.05 | 1 |
| ZBED9 | 1.19E-03 | 35 | 2 | 4.60E-04 | Esophagus_Mucosa | 3.38 | 1.93E-16 | 0.62 | -3.50 | 3.50 | -0.51 | 1.48 | 2 |
| ZBTB12 | 1.23E-03 | 22 | 3 | 2.65E-04 | Esophagus_Gastroesophageal_Junction | 4.00 | 8.82E-17 | 0.93 | -4.15 | 4.15 | 0.06 | 2.43 | 3 |
| ZNF80 | 1.23E-03 | 22 | 1 | 1.17E-03 | Small_Intestine_Terminal_Ileum | 3.00 | 1.19E-16 | 3.00 | -3.25 | 3.23 | -2.38 | 1.89 | 1 |
| RAPSN | 1.26E-03 | 45 | 3 | 2.98E-03 | Esophagus_Mucosa | 5.47 | 3.50E-16 | 0.41 | -2.42 | 2.97 | 1.04 | 1.90 | 3 |
| GATS | 1.28E-03 | 34 | 2 | 1.10E-03 | Esophagus_Gastroesophageal_Junction | 5.03 | 1.05E-16 | 0.97 | -2.12 | 3.26 | 0.89 | 1.90 | 2 |
| MAP3K20 | 1.33E-03 | 41 | 4 | 1.23E-04 | Esophagus_Gastroesophageal_Junction | 3.42 | 1.55E-17 | 0.33 | -3.84 | 3.84 | -0.69 | 2.13 | 4 |
| USHBP1 | 1.34E-03 | 27 | 3 | 8.00E-03 | Colon_Sigmoid | 3.72 | 8.86E-19 | 0.38 | -3.48 | 2.01 | -0.07 | 1.28 | 3 |
| KCNE3 | 1.35E-03 | 47 | 2 | 1.00E-03 | Esophagus_Mucosa | 6.04 | 3.59E-17 | 0.68 | -1.01 | 3.29 | 1.18 | 0.90 | 2 |
| TRIP12 | 1.38E-03 | 34 | 3 | 1.65E-02 | Colon_Transverse | 2.64 | 5.92E-18 | 0.14 | -2.26 | 2.40 | -0.63 | 1.84 | 3 |
| TOP1 | 1.39E-03 | 45 | 2 | 8.23E-03 | Esophagus_Mucosa | 6.00 | 1.61E-31 | 1.00 | -2.32 | 2.64 | -0.94 | 2.04 | 2 |
| P2RY1 | 1.41E-03 | 43 | 3 | 1.18E-04 | Stomach | 3.67 | 3.59E-17 | 0.34 | -2.89 | 3.85 | 1.90 | 2.10 | 3 |
| C1orf127 | 1.41E-03 | 25 | 1 | 1.41E-03 | Colon_Sigmoid | 3.00 | 2.11E-48 | 3.00 | -3.19 | 3.19 | -1.18 | 1.99 | 1 |
| DDX5 | 1.44E-03 | 39 | 1 | 1.44E-03 | Colon_Transverse | 7.00 | 2.47E-17 | 7.00 | -3.19 | -0.85 | -3.13 | 0.37 | 1 |
| LINC01336 | 1.44E-03 | 18 | 2 | 3.50E-04 | Colon_Sigmoid | 2.43 | 2.95E-16 | 0.57 | -2.06 | 3.90 | 2.67 | 1.45 | 2 |
| LINC01725 | 1.50E-03 | 18 | 2 | 4.28E-04 | Stomach | 1.13 | 8.66E-01 | 0.87 | -3.52 | 0.15 | -1.02 | 1.34 | 2 |
| FAM109A | 1.52E-03 | 46 | 2 | 2.18E-02 | Small_Intestine_Terminal_Ileum | 5.15 | 2.60E-18 | 0.85 | -2.29 | 2.87 | -0.33 | 1.80 | 2 |
| CEP290 | 1.53E-03 | 49 | 2 | 3.18E-04 | Colon_Sigmoid | 6.51 | 5.29E-17 | 0.47 | 1.26 | 3.62 | 2.95 | 0.59 | 2 |
| ZBTB25 | 1.53E-03 | 47 | 2 | 3.60E-04 | Colon_Transverse | 6.15 | 6.05E-17 | 0.75 | -3.57 | 3.71 | 2.62 | 1.54 | 2 |
| RP11-290F5.1 | 1.58E-03 | 44 | 4 | 4.22E-03 | Esophagus_Gastroesophageal_Junction | 3.67 | 1.01E-02 | 0.17 | -0.68 | 2.86 | 1.91 | 0.67 | 4 |
| ZRANB1 | 1.59E-03 | 40 | 3 | 1.93E-02 | Colon_Transverse | 5.31 | 5.85E-18 | 0.66 | -3.52 | 4.54 | -0.81 | 1.27 | 3 |
| C1QL4 | 1.60E-03 | 20 | 1 | 1.60E-03 | Small_Intestine_Terminal_Ileum | 1.00 | 1.00E+00 | 1.00 | -3.16 | 3.07 | -2.27 | 1.77 | 1 |
| HLA-DPB1 | 1.60E-03 | 37 | 3 | 7.31E-03 | Esophagus_Muscularis | 2.00 | 1.88E-01 | 0.19 | -1.78 | 2.68 | 0.35 | 1.16 | 3 |
| ZSCAN23 | 1.60E-03 | 47 | 2 | 3.89E-04 | Colon_Sigmoid | 6.73 | 8.17E-17 | 0.26 | 3.26 | 3.80 | 3.52 | 0.10 | 2 |
| ZSCAN26 | 1.60E-03 | 49 | 2 | 3.24E-04 | Esophagus_Gastroesophageal_Junction | 6.51 | 1.02E-16 | 0.41 | -3.71 | -0.60 | -3.30 | 0.50 | 2 |
| MRO | 1.63E-03 | 43 | 3 | 7.69E-05 | Colon_Sigmoid | 2.25 | 9.83E-18 | 0.80 | -3.95 | 2.59 | -0.98 | 1.57 | 3 |
| FAM131C | 1.66E-03 | 38 | 4 | 1.32E-02 | Stomach | 2.83 | 2.33E-05 | 0.15 | -2.96 | 3.60 | -0.28 | 2.01 | 4 |
| MTCH2 | 1.68E-03 | 28 | 2 | 3.56E-04 | Esophagus_Gastroesophageal_Junction | 3.30 | 8.97E-17 | 0.70 | -1.65 | 3.57 | 1.65 | 0.90 | 2 |
| NCK2 | 1.68E-03 | 43 | 4 | 7.72E-04 | Esophagus_Mucosa | 4.91 | 1.45E-17 | 0.30 | -1.68 | 3.70 | 1.51 | 1.14 | 4 |

| | | | | | | | | | | | | | |
|---|---|---|---|---|---|---|---|---|---|---|---|---|---|
| ENOX1-AS1 | 1.72E-03 | 9 | 1 | 1.72E-03 | Colon_Sigmoid | 1.00 | 1.00E+00 | 1.00 | 2.41 | 3.13 | 3.04 | 0.24 | 1 |
| GAS1RR | 1.73E-03 | 39 | 3 | 4.26E-04 | Colon_Sigmoid | 2.79 | 1.11E-17 | 0.17 | -3.65 | 1.18 | -2.54 | 1.33 | 3 |
| RAB40C | 1.74E-03 | 49 | 3 | 2.28E-04 | Esophagus_Mucosa | 5.75 | 2.53E-03 | 0.24 | -3.69 | 3.15 | -0.40 | 1.73 | 3 |
| ZKSCAN3 | 1.77E-03 | 45 | 3 | 4.43E-04 | Colon_Transverse | 4.28 | 2.24E-16 | 0.18 | -4.00 | 1.55 | -2.29 | 1.54 | 3 |
| SUFU | 1.77E-03 | 39 | 2 | 3.74E-04 | Esophagus_Gastroesophageal_Junction | 5.70 | 6.77E-17 | 0.30 | -3.56 | 3.56 | 2.44 | 2.08 | 2 |
| RP11-129M6.1 | 1.78E-03 | 4 | 2 | 5.03E-04 | Esophagus_Mucosa | 1.33 | 6.73E-01 | 0.67 | 1.84 | 3.48 | 3.07 | 0.82 | 2 |
| TMEM120A | 1.78E-03 | 46 | 2 | 2.34E-02 | Stomach | 5.77 | 5.21E-17 | 1.09 | -2.73 | 2.27 | -0.78 | 1.50 | 2 |
| FAM129A | 1.79E-03 | 47 | 5 | 2.14E-02 | Esophagus_Mucosa | 2.59 | 2.49E-16 | 0.35 | -2.30 | 1.04 | -0.29 | 0.75 | 5 |
| SH2B3 | 1.83E-03 | 41 | 1 | 1.83E-03 | Colon_Sigmoid | 5.00 | 7.08E-33 | 5.00 | -3.12 | 3.12 | 1.35 | 2.80 | 1 |
| FTL | 1.86E-03 | 5 | 1 | 1.86E-03 | Esophagus_Mucosa | 1.00 | 1.00E+00 | 1.00 | 0.73 | 3.11 | 1.36 | 0.98 | 1 |
| CTD-2201E18.5 | 1.89E-03 | 33 | 1 | 1.89E-03 | Esophagus_Gastroesophageal_Junction | 4.00 | 2.97E-17 | 4.00 | -3.11 | 3.11 | -2.34 | 1.25 | 1 |
| PUM1 | 1.90E-03 | 33 | 1 | 1.90E-03 | Stomach | 5.00 | 1.38E-33 | 5.00 | -2.29 | 3.10 | 2.39 | 1.60 | 1 |
| Z83851.4 | 1.91E-03 | 23 | 3 | 1.12E-03 | Esophagus_Mucosa | 2.16 | 7.39E-03 | 0.86 | -3.26 | 1.24 | -2.36 | 1.20 | 3 |
| ZCCHC4 | 1.97E-03 | 21 | 3 | 1.33E-03 | Esophagus_Gastroesophageal_Junction | 3.85 | 2.21E-18 | 0.32 | 1.56 | 3.76 | 3.01 | 0.57 | 3 |
| HSD17B8 | 1.99E-03 | 49 | 2 | 1.53E-01 | Colon_Sigmoid | 6.38 | 9.85E-06 | 0.41 | -1.57 | 2.36 | 0.15 | 0.96 | 2 |
| MPIG6B | 2.01E-03 | 39 | 2 | 9.69E-04 | Esophagus_Mucosa | 4.12 | 2.06E-20 | 0.88 | -2.57 | 3.40 | 0.33 | 1.26 | 2 |
| METAP1D | 2.01E-03 | 45 | 3 | 1.89E-04 | Stomach | 3.77 | 2.34E-17 | 0.71 | -2.03 | 3.74 | 1.46 | 1.75 | 3 |
| CEBPZ | 2.02E-03 | 47 | 1 | 1.60E-03 | Esophagus_Mucosa | 6.86 | 1.28E-16 | 6.86 | -3.36 | 1.03 | -2.88 | 0.73 | 1 |
| ANKRD16 | 2.02E-03 | 32 | 3 | 7.21E-04 | Esophagus_Muscularis | 3.64 | 2.22E-17 | 0.37 | -3.38 | 3.51 | -1.55 | 2.01 | 3 |
| GMDS | 2.03E-03 | 34 | 1 | 1.98E-03 | Esophagus_Muscularis | 4.00 | 4.92E-17 | 4.00 | -3.09 | 1.12 | -2.57 | 1.11 | 1 |
| RAB5A | 2.06E-03 | 48 | 3 | 1.35E-03 | Esophagus_Gastroesophageal_Junction | 4.14 | 3.04E-17 | 0.70 | -3.25 | 3.22 | 0.58 | 1.87 | 3 |
| FNBP4 | 2.06E-03 | 20 | 2 | 5.86E-04 | Colon_Sigmoid | 4.14 | 5.85E-21 | 0.86 | -3.44 | 3.44 | 2.61 | 1.79 | 2 |
| CARMIL3 | 2.08E-03 | 46 | 3 | 1.91E-03 | Esophagus_Mucosa | 4.75 | 1.30E-07 | 0.27 | -3.10 | 3.43 | 0.26 | 1.83 | 3 |
| C1orf56 | 2.10E-03 | 38 | 3 | 1.62E-02 | Esophagus_Mucosa | 4.29 | 4.34E-17 | 0.35 | -3.29 | 2.28 | 0.32 | 1.78 | 3 |
| HACD4 | 2.11E-03 | 48 | 4 | 1.87E-04 | Esophagus_Mucosa | 4.60 | 1.32E-04 | 0.16 | -3.07 | 4.18 | 0.53 | 2.52 | 4 |
| RAB26 | 2.12E-03 | 45 | 2 | 1.16E-03 | Esophagus_Gastroesophageal_Junction | 5.00 | 4.36E-17 | 1.00 | -3.25 | 3.25 | -0.27 | 2.62 | 2 |
| CKMT1A | 2.13E-03 | 19 | 1 | 1.90E-03 | Stomach | 2.97 | 2.06E-18 | 2.97 | -3.11 | 3.11 | -0.13 | 3.07 | 1 |
| CXCL5 | 2.14E-03 | 45 | 1 | 2.02E-03 | Esophagus_Mucosa | 5.94 | 5.59E-20 | 5.94 | -4.30 | 3.09 | -2.97 | 0.95 | 1 |
| SCMH1 | 2.15E-03 | 43 | 4 | 2.24E-04 | Esophagus_Gastroesophageal_Junction | 3.57 | 9.93E-17 | 0.20 | -3.69 | 2.13 | -1.42 | 1.73 | 4 |
| TMEM198 | 2.16E-03 | 37 | 2 | 1.10E-03 | Esophagus_Mucosa | 4.92 | 1.55E-19 | 0.98 | -3.84 | 1.95 | -1.10 | 2.03 | 2 |
| R3HCC1L | 2.17E-03 | 31 | 2 | 5.10E-04 | Esophagus_Gastroesophageal_Junction | 4.06 | 7.48E-18 | 0.92 | -3.88 | 0.17 | -2.58 | 1.06 | 2 |
| RPL4 | 2.20E-03 | 45 | 2 | 1.29E-03 | Esophagus_Muscularis | 4.39 | 7.24E-17 | 2.61 | -1.29 | 3.23 | 1.55 | 1.46 | 2 |
| MTX3 | 2.22E-03 | 47 | 3 | 1.44E-04 | Esophagus_Muscularis | 4.97 | 1.16E-16 | 0.37 | -2.49 | 3.80 | 1.35 | 1.44 | 3 |
| KYAT3 | 2.22E-03 | 49 | 2 | 4.59E-04 | Colon_Sigmoid | 6.44 | 8.24E-04 | 0.32 | 1.41 | 3.66 | 2.81 | 0.58 | 2 |

| | | | | | | | | | | | | | |
|---|---|---|---|---|---|---|---|---|---|---|---|---|---|
| CUL3 | 2.23E-03 | 34 | 2 | 4.45E-02 | Esophagus_Gastroesophageal_Junction | 2.78 | 1.16E-17 | 1.22 | -3.77 | 1.85 | -0.59 | 1.85 | 2 |
| ATXN2 | 2.25E-03 | 23 | 1 | 2.25E-03 | Esophagus_Mucosa | 2.00 | 2.86E-16 | 2.00 | 1.06 | 3.06 | 2.97 | 0.42 | 1 |
| MDH2 | 2.26E-03 | 48 | 2 | 6.34E-04 | Small_Intestine_Terminal_Ileum | 6.68 | 5.83E-17 | 0.28 | -3.43 | 3.15 | -1.94 | 1.64 | 2 |
| NMT2 | 2.26E-03 | 48 | 3 | 5.38E-04 | Esophagus_Muscularis | 5.25 | 1.74E-02 | 0.38 | -2.84 | 3.46 | 2.32 | 1.36 | 3 |
| AZGP1 | 2.27E-03 | 29 | 2 | 1.73E-01 | Esophagus_Gastroesophageal_Junction | 4.81 | 1.60E-18 | 0.19 | -1.36 | 2.66 | -0.85 | 1.09 | 2 |
| RAD51 | 2.27E-03 | 30 | 3 | 4.89E-04 | Colon_Sigmoid | 3.09 | 6.77E-18 | 0.84 | -3.61 | 3.49 | -0.87 | 1.85 | 3 |
| INTS12 | 2.28E-03 | 35 | 2 | 4.96E-03 | Small_Intestine_Terminal_Ileum | 4.96 | 1.29E-16 | 1.96 | -2.82 | 1.47 | -2.12 | 1.00 | 2 |
| ZNF536 | 2.28E-03 | 22 | 1 | 2.28E-03 | Small_Intestine_Terminal_Ileum | 1.00 | 1.00E+00 | 1.00 | -2.95 | 3.14 | 1.32 | 2.11 | 1 |
| CSNK2B | 2.29E-03 | 45 | 4 | 5.24E-03 | Esophagus_Gastroesophageal_Junction | 3.76 | 6.10E-33 | 0.39 | -1.76 | 3.70 | 1.57 | 1.36 | 4 |
| DAXX | 2.30E-03 | 47 | 4 | 5.88E-03 | Stomach | 3.07 | 1.25E-16 | 0.42 | -1.70 | 2.79 | 0.96 | 2.06 | 4 |
| ZSCAN31 | 2.33E-03 | 46 | 2 | 2.52E-04 | Colon_Sigmoid | 6.55 | 1.09E-03 | 0.23 | -0.18 | 4.07 | 3.09 | 0.81 | 2 |
| PSMC6 | 2.33E-03 | 31 | 1 | 2.33E-03 | Colon_Transverse | 3.00 | 2.11E-48 | 3.00 | -3.04 | 3.04 | 0.40 | 3.01 | 1 |
| PWWP2A | 2.34E-03 | 46 | 4 | 3.65E-04 | Colon_Transverse | 3.26 | 1.01E-17 | 0.60 | -3.84 | 1.06 | -1.38 | 2.04 | 4 |
| RP11-9M16.2 | 2.35E-03 | 28 | 2 | 6.01E-04 | Esophagus_Mucosa | 5.80 | 5.73E-17 | 1.20 | -3.43 | 3.43 | 1.60 | 2.10 | 2 |
| DVL2 | 2.38E-03 | 24 | 2 | 6.93E-04 | Esophagus_Mucosa | 1.29 | 7.08E-01 | 0.71 | -3.39 | 3.39 | -1.32 | 3.11 | 2 |
| SLC38A6 | 2.38E-03 | 46 | 2 | 1.63E-03 | Colon_Sigmoid | 5.37 | 7.14E-18 | 0.49 | -3.15 | 3.15 | -2.22 | 1.78 | 2 |
| TLR3 | 2.40E-03 | 33 | 4 | 1.63E-02 | Esophagus_Mucosa | 4.48 | 1.31E-16 | 0.16 | -2.52 | 2.40 | 0.56 | 1.50 | 4 |
| CRTAC1 | 2.42E-03 | 42 | 2 | 6.96E-04 | Colon_Sigmoid | 5.16 | 3.68E-17 | 1.83 | -3.39 | 1.46 | -1.65 | 1.62 | 2 |
| LINC00243 | 2.42E-03 | 31 | 5 | 1.10E-03 | Small_Intestine_Terminal_Ileum | 2.86 | 1.42E-16 | 0.15 | -2.05 | 3.26 | 1.37 | 1.60 | 5 |
| FGF14-AS2 | 2.44E-03 | 46 | 3 | 4.75E-04 | Colon_Transverse | 4.31 | 2.01E-17 | 1.22 | 0.15 | 4.03 | 3.18 | 0.83 | 3 |
| SYT1 | 2.45E-03 | 24 | 2 | 4.57E-02 | Small_Intestine_Terminal_Ileum | 2.63 | 4.31E-02 | 0.32 | -2.35 | 2.43 | 1.24 | 1.32 | 2 |
| SPATA2 | 2.45E-03 | 43 | 2 | 5.35E-04 | Colon_Sigmoid | 4.06 | 1.71E-18 | 1.94 | -2.61 | 3.62 | 0.81 | 1.64 | 2 |
| CCL15 | 2.48E-03 | 26 | 2 | 1.77E-03 | Colon_Sigmoid | 5.26 | 2.21E-16 | 1.47 | -2.73 | 3.78 | 1.69 | 1.99 | 2 |
| 10-Mar | 2.52E-03 | 24 | 2 | 2.47E-03 | Esophagus_Muscularis | 1.78 | 2.22E-01 | 0.22 | -3.11 | 0.72 | -0.76 | 0.85 | 2 |
| DDX31 | 2.52E-03 | 47 | 1 | 2.40E-03 | Stomach | 6.99 | 1.49E-33 | 6.99 | -3.46 | 2.30 | -2.22 | 1.49 | 1 |
| PLPP2 | 2.52E-03 | 46 | 2 | 5.72E-04 | Stomach | 6.36 | 1.85E-02 | 0.34 | 1.76 | 3.58 | 3.00 | 0.45 | 2 |
| RP11-274B21.9 | 2.55E-03 | 49 | 2 | 1.28E-03 | Esophagus_Muscularis | 6.31 | 1.34E-05 | 0.38 | 0.65 | 3.28 | 2.22 | 0.61 | 2 |
| KIF13A | 2.57E-03 | 46 | 5 | 1.29E-02 | Esophagus_Mucosa | 3.82 | 2.38E-17 | 0.29 | -2.96 | 3.00 | 0.20 | 1.55 | 5 |
| TESK2 | 2.58E-03 | 49 | 3 | 2.69E-03 | Stomach | 4.09 | 8.24E-17 | 0.31 | -1.04 | 3.65 | 1.53 | 1.18 | 3 |
| FANCI | 2.62E-03 | 37 | 3 | 6.62E-03 | Esophagus_Mucosa | 4.99 | 4.87E-18 | 0.22 | -2.82 | 2.32 | -1.28 | 0.94 | 3 |
| EPB41 | 2.64E-03 | 45 | 2 | 5.40E-04 | Esophagus_Muscularis | 6.65 | 5.16E-17 | 0.35 | -3.46 | 3.46 | 2.45 | 2.02 | 2 |
| RARG | 2.66E-03 | 3 | 1 | 2.66E-03 | Esophagus_Mucosa | 1.00 | 1.00E+00 | 1.00 | 1.54 | 3.21 | 2.59 | 0.91 | 1 |
| RTN4RL2 | 2.67E-03 | 16 | 2 | 7.11E-04 | Colon_Transverse | 1.31 | 6.93E-01 | 0.69 | -3.39 | 1.35 | -1.71 | 1.18 | 2 |
| LGALSL | 2.67E-03 | 38 | 2 | 1.09E-03 | Esophagus_Mucosa | 4.22 | 1.77E-33 | 1.78 | -3.27 | 3.27 | 1.14 | 1.91 | 2 |
| ACTR5 | 2.69E-03 | 46 | 3 | 2.52E-03 | Colon_Transverse | 4.32 | 1.66E-18 | 1.25 | -4.79 | 1.65 | -0.31 | 1.56 | 3 |
| HSDL2 | 2.71E-03 | 41 | 3 | 2.11E-04 | Colon_Transverse | 4.92 | 1.29E-18 | 0.23 | -3.71 | 3.35 | -2.20 | 1.58 | 3 |

| | | | | | | | | | | | | | |
|---|---|---|---|---|---|---|---|---|---|---|---|---|---|
| WDR4 | 2.73E-03 | 42 | 3 | 7.62E-03 | Esophagus_Muscularis | 3.02 | 1.47E-16 | 0.96 | -0.89 | 2.67 | 1.13 | 1.27 | 3 |
| NEURL2 | 2.74E-03 | 29 | 7 | 1.81E-04 | Colon_Transverse | 3.08 | 1.42E-01 | 0.14 | -1.90 | 3.74 | 0.36 | 1.49 | 7 |
| MFAP1 | 2.76E-03 | 48 | 1 | 2.76E-03 | Colon_Sigmoid | 7.00 | 3.15E-17 | 7.00 | -2.99 | -2.99 | -2.99 | 0.00 | 1 |
| CNN1 | 2.78E-03 | 35 | 2 | 8.44E-04 | Colon_Transverse | 3.48 | 6.28E-34 | 2.48 | -3.39 | 3.45 | 0.78 | 2.53 | 2 |
| CTNND1 | 2.78E-03 | 21 | 1 | 2.78E-03 | Esophagus_Muscularis | 3.00 | 6.09E-17 | 3.00 | -2.99 | 3.71 | 2.10 | 2.13 | 1 |
| OXER1 | 2.80E-03 | 44 | 3 | 2.19E-03 | Esophagus_Gastroesophageal_Junction | 4.57 | 2.20E-18 | 0.24 | -3.14 | 3.06 | -1.46 | 1.45 | 3 |
| IP6K3 | 2.80E-03 | 49 | 4 | 1.80E-03 | Esophagus_Gastroesophageal_Junction | 4.90 | 9.30E-03 | 0.22 | -3.44 | 3.12 | -0.36 | 1.70 | 4 |
| CTD-2325M2.1 | 2.80E-03 | 47 | 2 | 6.30E-04 | Stomach | 6.34 | 2.04E-17 | 0.66 | -3.47 | -2.59 | -3.15 | 0.25 | 2 |
| HAUS4 | 2.84E-03 | 47 | 1 | 2.31E-03 | Colon_Transverse | 5.92 | 4.96E-17 | 5.92 | 2.74 | 3.30 | 2.94 | 0.11 | 1 |
| ACSF2 | 2.85E-03 | 47 | 4 | 4.65E-04 | Esophagus_Muscularis | 4.18 | 8.61E-18 | 0.60 | -3.39 | 3.86 | 1.67 | 1.71 | 4 |
| NKIRAS1 | 2.86E-03 | 43 | 3 | 5.79E-04 | Small_Intestine_Terminal_Ileum | 5.18 | 1.96E-16 | 0.68 | -3.64 | 3.63 | -0.07 | 2.96 | 3 |
| PLEKHG3 | 2.88E-03 | 35 | 3 | 1.44E-02 | Small_Intestine_Terminal_Ileum | 5.03 | 1.98E-16 | 0.23 | -3.12 | 2.92 | -0.06 | 1.23 | 3 |
| GGNBP2 | 2.89E-03 | 47 | 1 | 1.22E-03 | Colon_Sigmoid | 6.73 | 8.25E-06 | 6.73 | -3.42 | -0.12 | -2.96 | 0.51 | 1 |
| XXbac-BPG13B8.10 | 2.91E-03 | 18 | 4 | 1.74E-03 | Colon_Transverse | 3.85 | 2.12E-17 | 0.43 | -1.37 | 3.14 | 1.55 | 1.90 | 4 |
| PRMT5 | 2.92E-03 | 48 | 3 | 8.73E-03 | Small_Intestine_Terminal_Ileum | 5.57 | 1.07E-16 | 0.38 | -2.64 | 1.67 | -0.05 | 1.27 | 3 |
| TMEM51 | 2.93E-03 | 38 | 4 | 8.39E-03 | Colon_Transverse | 1.85 | 2.63E-01 | 0.26 | -2.85 | 3.42 | 0.73 | 2.06 | 4 |
| TMEM117 | 2.94E-03 | 45 | 3 | 4.41E-04 | Stomach | 5.64 | 1.35E-17 | 0.55 | -3.51 | 3.51 | -1.87 | 2.38 | 3 |
| TRIM33 | 2.94E-03 | 44 | 4 | 6.29E-05 | Small_Intestine_Terminal_Ileum | 2.75 | 2.20E-17 | 0.29 | -4.00 | 4.00 | 0.13 | 2.46 | 4 |
| CTC1 | 3.00E-03 | 42 | 3 | 7.26E-04 | Colon_Transverse | 5.16 | 2.04E-02 | 0.23 | -4.13 | 1.77 | -1.61 | 1.66 | 3 |
| ZNF165 | 3.00E-03 | 38 | 2 | 2.58E-04 | Stomach | 5.26 | 1.16E-18 | 0.53 | -3.65 | 2.88 | -2.52 | 1.73 | 2 |
| KLHL24 | 3.02E-03 | 28 | 1 | 2.14E-03 | Stomach | 3.98 | 9.78E-18 | 3.98 | -3.07 | -1.68 | -2.87 | 0.32 | 1 |
| AMIGO1 | 3.05E-03 | 49 | 2 | 2.09E-01 | Colon_Transverse | 6.65 | 4.12E-19 | 0.30 | -1.76 | 1.31 | -0.68 | 0.57 | 2 |
| SEMA6C | 3.06E-03 | 48 | 3 | 2.19E-01 | Esophagus_Gastroesophageal_Junction | 5.90 | 2.48E-17 | 0.24 | -1.35 | 2.09 | -0.41 | 0.78 | 3 |
| WDR76 | 3.06E-03 | 32 | 1 | 3.06E-03 | Colon_Transverse | 4.00 | 1.79E-16 | 4.00 | -2.96 | -1.01 | -2.90 | 0.35 | 1 |
| PRC1 | 3.07E-03 | 46 | 4 | 1.62E-04 | Small_Intestine_Terminal_Ileum | 3.02 | 5.90E-18 | 0.39 | -1.25 | 3.89 | 1.27 | 1.75 | 4 |
| ALKBH5 | 3.10E-03 | 49 | 4 | 3.18E-03 | Small_Intestine_Terminal_Ileum | 4.16 | 2.06E-04 | 0.17 | -2.05 | 3.03 | 1.17 | 0.92 | 4 |
| ZBTB17 | 3.15E-03 | 23 | 1 | 3.15E-03 | Colon_Sigmoid | 3.00 | 1.11E-16 | 3.00 | -2.95 | 2.95 | -1.45 | 2.21 | 1 |
| PROSER3 | 3.16E-03 | 48 | 2 | 5.95E-02 | Esophagus_Muscularis | 6.55 | 1.41E-07 | 0.23 | -2.22 | 1.25 | -0.99 | 1.01 | 2 |
| OGFOD3 | 3.19E-03 | 49 | 2 | 8.25E-02 | Colon_Transverse | 6.47 | 2.35E-08 | 0.36 | -0.55 | 3.19 | 0.38 | 0.80 | 2 |
| CHMP4B | 3.24E-03 | 41 | 2 | 6.32E-03 | Colon_Transverse | 6.51 | 2.87E-18 | 0.38 | -2.73 | 2.73 | 1.77 | 1.52 | 2 |
| CHAD | 3.24E-03 | 42 | 3 | 5.74E-04 | Esophagus_Muscularis | 5.37 | 1.61E-17 | 0.28 | -3.06 | 3.44 | -0.35 | 2.13 | 3 |
| CCDC88C | 3.24E-03 | 47 | 6 | 1.17E-03 | Colon_Transverse | 2.49 | 3.39E-16 | 0.20 | -3.41 | 3.77 | -0.38 | 1.99 | 6 |
| GBE1 | 3.27E-03 | 43 | 2 | 1.07E-02 | Stomach | 3.02 | 2.57E-20 | 0.98 | -4.65 | 4.43 | -1.39 | 2.51 | 2 |
| PCOLCE | 3.30E-03 | 44 | 3 | 1.19E-01 | Small_Intestine_Terminal_Ileum | 4.70 | 1.20E-16 | 0.44 | -0.85 | 2.52 | 0.04 | 0.92 | 3 |
| RPL24 | 3.37E-03 | 28 | 1 | 3.37E-03 | Colon_Sigmoid | 4.00 | 8.27E-17 | 4.00 | -2.93 | 1.60 | -2.77 | 0.86 | 1 |

| | | | | | | | | | | | | | |
|---|---|---|---|---|---|---|---|---|---|---|---|---|---|
| BAG6 | 3.38E-03 | 46 | 2 | 1.13E-04 | Colon_Sigmoid | 6.11 | 2.81E-04 | 0.57 | -4.21 | 4.21 | -2.19 | 1.33 | 2 |
| EEF1E1 | 3.45E-03 | 41 | 1 | 1.13E-03 | Esophagus_Mucosa | 6.85 | 3.93E-17 | 6.85 | 0.75 | 3.27 | 2.59 | 0.66 | 1 |
| ZNF574 | 3.53E-03 | 12 | 1 | 2.54E-03 | Esophagus_Mucosa | 2.97 | 1.96E-34 | 2.97 | -2.67 | 3.02 | 2.28 | 1.57 | 1 |
| FOXA1 | 3.55E-03 | 26 | 3 | 1.44E-02 | Esophagus_Mucosa | 1.62 | 4.07E-01 | 0.41 | -2.30 | 2.92 | 0.20 | 1.30 | 3 |
| VPS33B | 3.56E-03 | 44 | 4 | 1.67E-03 | Small_Intestine_Terminal_Ileum | 3.58 | 7.11E-17 | 0.54 | -2.17 | 3.78 | 1.56 | 1.70 | 4 |
| STAT6 | 3.59E-03 | 47 | 3 | 8.72E-03 | Stomach | 5.26 | 1.36E-16 | 0.71 | -2.62 | 3.15 | 0.94 | 1.87 | 3 |
| ZKSCAN8 | 3.65E-03 | 48 | 5 | 7.05E-04 | Stomach | 3.18 | 8.35E-17 | 0.19 | -3.40 | 3.27 | -1.60 | 1.66 | 5 |
| MTMR12 | 3.65E-03 | 38 | 4 | 3.41E-03 | Stomach | 3.44 | 1.78E-17 | 0.28 | -3.45 | 1.98 | -1.95 | 1.24 | 4 |
| NUP160 | 3.69E-03 | 19 | 2 | 8.23E-04 | Colon_Sigmoid | 2.31 | 5.75E-18 | 0.69 | -1.51 | 3.53 | 1.46 | 1.46 | 2 |
| ARPP19 | 3.72E-03 | 35 | 2 | 8.48E-04 | Colon_Transverse | 3.99 | 2.17E-17 | 1.89 | -3.57 | 0.09 | -2.69 | 1.27 | 2 |
| TBC1D4 | 3.74E-03 | 48 | 1 | 1.70E-03 | Esophagus_Muscularis | 6.84 | 1.71E-03 | 6.84 | 0.21 | 3.85 | 2.35 | 1.00 | 1 |
| CDH13 | 3.75E-03 | 24 | 2 | 1.58E-03 | Esophagus_Mucosa | 2.02 | 2.34E-17 | 0.98 | -3.16 | 2.69 | -0.67 | 1.56 | 2 |
| LINC01948 | 3.76E-03 | 12 | 2 | 8.52E-04 | Colon_Transverse | 2.00 | 4.27E-32 | 1.00 | -0.57 | 3.34 | 0.35 | 1.00 | 2 |
| ZBTB32 | 3.77E-03 | 44 | 3 | 2.91E-03 | Colon_Sigmoid | 5.64 | 1.39E-03 | 0.21 | -2.33 | 3.11 | 1.60 | 1.17 | 3 |
| C4orf47 | 3.84E-03 | 37 | 5 | 2.67E-03 | Esophagus_Muscularis | 2.92 | 6.60E-17 | 0.14 | -3.08 | 2.12 | 0.59 | 1.25 | 5 |
| MBOAT4 | 3.87E-03 | 42 | 2 | 8.95E-04 | Esophagus_Muscularis | 6.37 | 1.76E-17 | 0.58 | -3.32 | 3.32 | -2.27 | 1.96 | 2 |
| KRBA2 | 3.88E-03 | 48 | 2 | 1.38E-02 | Small_Intestine_Terminal_Ileum | 6.36 | 6.82E-17 | 0.57 | -3.49 | 1.63 | -0.80 | 1.09 | 2 |
| ZNF391 | 3.88E-03 | 46 | 1 | 2.97E-03 | Stomach | 6.94 | 4.83E-18 | 6.94 | -0.78 | 3.35 | 2.77 | 0.58 | 1 |
| UTP18 | 3.90E-03 | 47 | 4 | 4.96E-02 | Esophagus_Gastroesophageal_Junction | 4.18 | 5.21E-17 | 0.35 | -3.70 | 1.96 | 0.11 | 1.44 | 4 |
| ZNHIT3 | 3.90E-03 | 44 | 2 | 1.26E-03 | Colon_Sigmoid | 6.44 | 6.57E-17 | 0.46 | -3.23 | 3.23 | 1.84 | 2.42 | 2 |
| PTCH1 | 3.90E-03 | 29 | 2 | 1.11E-03 | Esophagus_Muscularis | 3.00 | 5.86E-17 | 0.96 | -3.26 | 2.81 | -1.61 | 1.87 | 2 |
| FLOT1 | 3.93E-03 | 49 | 2 | 8.36E-03 | Small_Intestine_Terminal_Ileum | 6.26 | 1.73E-03 | 0.34 | -1.34 | 3.28 | 1.61 | 1.37 | 2 |
| PCNP | 3.98E-03 | 25 | 2 | 1.69E-03 | Colon_Sigmoid | 2.13 | 3.25E-16 | 0.87 | -0.18 | 3.14 | 2.13 | 1.47 | 2 |
| RP11-434D9.1 | 4.06E-03 | 4 | 1 | 4.06E-03 | Esophagus_Muscularis | 1.00 | 1.00E+00 | 1.00 | -0.27 | 2.87 | 0.84 | 1.40 | 1 |
| CKMT1B | 4.07E-03 | 1 | 1 | 4.07E-03 | Esophagus_Mucosa | 1.00 | 1.00E+00 | 1.00 | 2.87 | 2.87 | 2.87 | NA | 1 |
| BCL2A1 | 4.07E-03 | 8 | 1 | 4.07E-03 | Esophagus_Mucosa | 2.00 | 3.35E-17 | 2.00 | -2.87 | 2.87 | -2.15 | 2.03 | 1 |
| CDH16 | 4.12E-03 | 6 | 2 | 1.52E-03 | Colon_Transverse | 1.79 | 2.10E-01 | 0.21 | 0.30 | 3.17 | 2.12 | 1.07 | 2 |
| DHRS11 | 4.13E-03 | 36 | 2 | 6.41E-04 | Esophagus_Mucosa | 5.58 | 1.08E-16 | 0.31 | -2.56 | 3.41 | 2.29 | 1.40 | 2 |
| COL1A2 | 4.16E-03 | 14 | 1 | 4.16E-03 | Esophagus_Gastroesophageal_Junction | 1.00 | 1.00E+00 | 1.00 | 0.37 | 2.87 | 2.52 | 0.89 | 1 |
| SRSF4 | 4.20E-03 | 5 | 1 | 4.20E-03 | Esophagus_Mucosa | 1.00 | 1.00E+00 | 1.00 | -2.86 | 2.86 | 0.02 | 2.03 | 1 |
| SMARCC2 | 4.20E-03 | 47 | 1 | 2.13E-03 | Esophagus_Mucosa | 6.84 | 2.43E-17 | 6.84 | -3.07 | 3.07 | -2.51 | 1.49 | 1 |
| CAND2 | 4.24E-03 | 47 | 3 | 8.96E-02 | Esophagus_Muscularis | 4.62 | 1.65E-17 | 0.52 | -1.70 | 1.74 | 0.98 | 0.66 | 3 |
| LCE2A | 4.25E-03 | 4 | 1 | 4.25E-03 | Small_Intestine_Terminal_Ileum | 1.00 | 1.00E+00 | 1.00 | 2.86 | 2.86 | 2.86 | 0.00 | 1 |
| EPN3 | 4.27E-03 | 43 | 3 | 1.87E-04 | Colon_Transverse | 5.28 | 2.48E-17 | 0.21 | -3.42 | 3.74 | 2.50 | 1.89 | 3 |
| RP11-147L13.14 | 4.30E-03 | 47 | 2 | 4.10E-03 | Small_Intestine_Terminal_Ileum | 6.52 | 3.87E-06 | 0.37 | -0.94 | 2.88 | 1.86 | 0.74 | 2 |
| GABARAP | 4.30E-03 | 33 | 3 | 7.87E-04 | Esophagus_Muscularis | 4.16 | 1.55E-18 | 0.68 | -3.36 | 3.49 | 1.32 | 2.69 | 3 |

| | | | | | | | | | | | | | |
|---|---|---|---|---|---|---|---|---|---|---|---|---|---|
| BCL2L11 | 4.32E-03 | 41 | 3 | 2.45E-03 | Colon_Sigmoid | 2.25 | 1.68E-17 | 0.69 | -3.03 | 3.37 | 1.19 | 1.15 | 3 |
| TRIM61 | 4.34E-03 | 42 | 2 | 1.71E-02 | Stomach | 5.59 | 2.15E-18 | 0.26 | 0.72 | 2.39 | 2.09 | 0.54 | 2 |
| KBTBD4 | 4.39E-03 | 39 | 2 | 9.88E-04 | Colon_Sigmoid | 5.24 | 9.54E-34 | 0.76 | -3.29 | 3.29 | -0.95 | 3.14 | 2 |
| ACADSB | 4.40E-03 | 40 | 5 | 6.14E-05 | Stomach | 2.33 | 1.54E-16 | 0.11 | -3.98 | 4.04 | 1.43 | 1.78 | 5 |
| FBN1 | 4.43E-03 | 16 | 1 | 4.43E-03 | Esophagus_Gastroesophageal_Junction | 1.00 | 1.00E+00 | 1.00 | -2.85 | 2.85 | -1.03 | 2.21 | 1 |
| CCDC152 | 4.44E-03 | 43 | 3 | 7.90E-03 | Esophagus_Mucosa | 5.02 | 4.41E-17 | 0.40 | -2.05 | 2.66 | 0.26 | 1.23 | 3 |
| PNMA1 | 4.46E-03 | 42 | 2 | 5.65E-04 | Colon_Transverse | 4.55 | 4.33E-03 | 0.29 | -3.45 | 1.16 | -2.01 | 1.47 | 2 |
| FCN1 | 4.47E-03 | 44 | 2 | 8.46E-04 | Colon_Sigmoid | 5.93 | 3.52E-17 | 0.95 | -3.14 | 3.41 | 1.94 | 1.76 | 2 |
| GPR1 | 4.47E-03 | 39 | 3 | 1.13E-01 | Colon_Sigmoid | 5.32 | 4.05E-17 | 0.68 | -1.82 | 2.25 | -0.19 | 1.45 | 3 |
| BRD3 | 4.49E-03 | 47 | 3 | 7.48E-03 | Colon_Transverse | 5.90 | 3.40E-18 | 0.38 | -1.12 | 2.80 | 1.85 | 0.90 | 3 |
| AKNA | 4.54E-03 | 47 | 4 | 1.73E-04 | Colon_Sigmoid | 4.98 | 3.52E-17 | 0.24 | -1.30 | 3.76 | 2.24 | 1.25 | 4 |
| GNPNAT1 | 4.56E-03 | 7 | 1 | 4.56E-03 | Colon_Transverse | 3.00 | 1.92E-33 | 3.00 | -3.08 | 2.84 | 0.24 | 3.11 | 1 |
| OS9 | 4.59E-03 | 15 | 1 | 8.70E-04 | Esophagus_Gastroesophageal_Junction | 5.81 | 1.46E-16 | 5.81 | -3.47 | -2.65 | -2.83 | 0.26 | 1 |
| PPP1R18 | 4.60E-03 | 46 | 2 | 7.79E-04 | Esophagus_Gastroesophageal_Junction | 3.94 | 3.81E-31 | 2.97 | -4.21 | 0.61 | -1.61 | 1.65 | 2 |
| LINC01750 | 4.62E-03 | 45 | 1 | 3.70E-03 | Esophagus_Muscularis | 5.94 | 7.14E-20 | 5.94 | -3.12 | 1.26 | -2.65 | 0.80 | 1 |
| COL8A1 | 4.65E-03 | 47 | 4 | 9.45E-02 | Colon_Sigmoid | 4.48 | 1.53E-16 | 0.31 | -2.30 | 1.92 | -0.26 | 1.43 | 4 |
| SPINT2 | 4.66E-03 | 37 | 2 | 3.10E-03 | Small_Intestine_Terminal_Ileum | 1.83 | 1.68E-01 | 0.17 | -3.90 | 2.20 | -1.25 | 1.59 | 2 |
| PPARGC1B | 4.72E-03 | 28 | 4 | 7.80E-03 | Esophagus_Muscularis | 1.69 | 1.55E-01 | 0.16 | -1.99 | 2.66 | 1.16 | 1.24 | 4 |
| PIP5K1A | 4.72E-03 | 43 | 2 | 1.28E-03 | Stomach | 4.57 | 5.04E-18 | 0.41 | -3.22 | -0.55 | -2.18 | 0.39 | 2 |
| FBXO18 | 4.73E-03 | 49 | 3 | 1.27E-03 | Esophagus_Muscularis | 5.32 | 2.42E-03 | 0.30 | -3.41 | 0.87 | -2.10 | 1.37 | 3 |
| SRP19 | 4.74E-03 | 46 | 1 | 4.61E-03 | Stomach | 5.00 | 5.63E-17 | 5.00 | -2.83 | 2.82 | -2.35 | 1.13 | 1 |
| RP11-1263C18.2 | 4.74E-03 | 15 | 2 | 1.37E-03 | Esophagus_Mucosa | 1.63 | 3.69E-01 | 0.37 | -3.20 | 2.55 | -2.05 | 1.40 | 2 |
| LIN37 | 4.77E-03 | 47 | 2 | 1.47E-03 | Esophagus_Muscularis | 6.26 | 2.52E-17 | 0.74 | -3.17 | 3.49 | 2.86 | 1.30 | 2 |
| ACADVL | 4.77E-03 | 23 | 2 | 3.95E-02 | Colon_Sigmoid | 2.50 | 2.30E-17 | 0.50 | -3.39 | 3.28 | -1.16 | 2.12 | 2 |
| PSMG3-AS1 | 4.81E-03 | 49 | 3 | 1.82E-04 | Colon_Transverse | 5.96 | 4.22E-02 | 0.27 | 0.12 | 3.93 | 2.61 | 0.75 | 3 |
| ARSK | 4.82E-03 | 31 | 3 | 9.65E-03 | Colon_Sigmoid | 4.07 | 8.89E-17 | 0.92 | -2.59 | 2.62 | -1.15 | 1.97 | 3 |
| FGB | 4.90E-03 | 37 | 3 | 3.23E-02 | Stomach | 2.62 | 1.05E-16 | 0.98 | -1.77 | 2.45 | 1.02 | 1.63 | 3 |
| PIP4K2C | 4.91E-03 | 44 | 3 | 4.08E-03 | Esophagus_Mucosa | 3.74 | 5.63E-18 | 1.22 | -3.20 | 0.48 | -1.27 | 1.38 | 3 |
| KRTDAP | 4.92E-03 | 14 | 2 | 4.22E-03 | Esophagus_Muscularis | 2.00 | 3.44E-32 | 1.00 | -2.86 | 2.86 | -1.26 | 2.20 | 2 |
| PPBP | 4.94E-03 | 3 | 2 | 2.28E-03 | Esophagus_Muscularis | 1.45 | 5.51E-01 | 0.55 | -3.05 | -2.39 | -2.77 | 0.34 | 2 |
| EVI2A | 4.95E-03 | 46 | 1 | 4.36E-03 | Colon_Sigmoid | 6.90 | 2.13E-20 | 6.90 | -2.95 | -2.09 | -2.82 | 0.13 | 1 |
| HTR1B | 4.95E-03 | 38 | 2 | 8.35E-03 | Small_Intestine_Terminal_Ileum | 3.99 | 1.66E-17 | 1.98 | -2.95 | 1.83 | -1.60 | 1.52 | 2 |
| ELAVL2 | 4.99E-03 | 13 | 3 | 1.03E-03 | Esophagus_Muscularis | 1.34 | 6.84E-01 | 0.68 | -0.02 | 3.28 | 1.87 | 1.00 | 3 |
| FLRT1 | 4.99E-03 | 45 | 4 | 1.75E-02 | Colon_Sigmoid | 3.86 | 1.42E-16 | 0.55 | -2.42 | 2.38 | -0.44 | 1.97 | 4 |
| USPL1 | 4.99E-03 | 5 | 1 | 4.99E-03 | Esophagus_Mucosa | 1.00 | 1.00E+00 | 1.00 | 0.70 | 2.81 | 1.14 | 0.93 | 1 |
| C18orf21 | 5.06E-03 | 36 | 1 | 5.06E-03 | Colon_Sigmoid | 5.00 | 7.49E-33 | 5.00 | -2.80 | 2.80 | -2.59 | 0.97 | 1 |

| | | | | | | | | | | | | | |
|---|---|---|---|---|---|---|---|---|---|---|---|---|---|
| ABCA7 | 5.07E-03 | 49 | 2 | 8.08E-02 | Colon_Transverse | 6.12 | 3.93E-03 | 0.46 | -2.43 | 3.27 | -0.54 | 1.26 | 2 |
| TCF20 | 5.08E-03 | 10 | 2 | 1.75E-03 | Colon_Transverse | 1.04 | 9.60E-01 | 0.96 | -3.11 | 3.13 | -0.65 | 1.60 | 2 |
| ADRB2 | 5.08E-03 | 31 | 1 | 4.86E-03 | Colon_Transverse | 3.99 | 1.09E-16 | 3.99 | -2.82 | 2.82 | -1.38 | 1.77 | 1 |
| CCNE1 | 5.12E-03 | 26 | 3 | 7.22E-04 | Small_Intestine_Terminal_Ileum | 4.24 | 1.59E-16 | 0.71 | -3.38 | 2.62 | -1.70 | 1.82 | 3 |
| MED19 | 5.12E-03 | 37 | 4 | 2.95E-04 | Stomach | 3.35 | 7.95E-17 | 0.43 | -1.91 | 3.68 | 0.77 | 1.75 | 4 |
| CYHR1 | 5.14E-03 | 17 | 1 | 3.62E-03 | Colon_Sigmoid | 2.93 | 1.84E-02 | 2.93 | -2.91 | 2.64 | -1.62 | 1.55 | 1 |
| UEVLD | 5.18E-03 | 20 | 1 | 5.18E-03 | Esophagus_Mucosa | 3.00 | 1.41E-16 | 3.00 | -0.42 | 2.80 | 1.80 | 1.32 | 1 |
| SPTY2D1-AS1 | 5.18E-03 | 18 | 1 | 5.18E-03 | Colon_Transverse | 5.00 | 5.57E-17 | 5.00 | -0.81 | 2.80 | 2.32 | 0.98 | 1 |
| AMPH | 5.22E-03 | 23 | 2 | 1.29E-03 | Small_Intestine_Terminal_Ileum | 1.01 | 9.92E-01 | 0.99 | -3.22 | 0.19 | -0.51 | 0.76 | 2 |
| MYO19 | 5.22E-03 | 46 | 3 | 4.68E-04 | Colon_Sigmoid | 5.64 | 3.74E-17 | 0.33 | -0.80 | 3.50 | 2.74 | 1.01 | 3 |
| IQCH | 5.29E-03 | 47 | 1 | 4.81E-03 | Stomach | 6.99 | 4.11E-17 | 6.99 | -3.07 | -1.63 | -2.59 | 0.34 | 1 |
| MSANTD1 | 5.32E-03 | 32 | 2 | 1.50E-03 | Esophagus_Mucosa | 4.96 | 9.10E-33 | 0.96 | -0.57 | 3.18 | 0.90 | 0.67 | 2 |
| ANGEL2 | 5.36E-03 | 46 | 2 | 4.15E-03 | Esophagus_Gastroesophageal_Junction | 6.50 | 7.82E-17 | 0.27 | 1.30 | 2.89 | 2.14 | 0.59 | 2 |
| TK2 | 5.49E-03 | 49 | 1 | 4.00E-03 | Colon_Sigmoid | 6.78 | 3.16E-17 | 6.78 | -2.90 | 2.75 | -1.22 | 1.86 | 1 |
| IVNS1ABP | 5.53E-03 | 49 | 3 | 1.07E-03 | Small_Intestine_Terminal_Ileum | 5.12 | 1.07E-16 | 0.84 | -1.55 | 3.29 | 0.37 | 0.90 | 3 |
| SCO2 | 5.54E-03 | 48 | 2 | 6.97E-02 | Colon_Transverse | 6.70 | 4.99E-17 | 0.28 | -2.32 | 1.99 | -0.57 | 1.15 | 2 |
| DYNLRB2 | 5.54E-03 | 21 | 2 | 1.56E-03 | Colon_Sigmoid | 2.06 | 2.66E-17 | 1.94 | -3.16 | 1.84 | -0.85 | 1.24 | 2 |
| AK6 | 5.57E-03 | 48 | 3 | 6.22E-02 | Small_Intestine_Terminal_Ileum | 3.37 | 9.26E-17 | 0.68 | -1.70 | 1.86 | 0.80 | 1.07 | 3 |
| HLA-G | 5.58E-03 | 47 | 4 | 5.79E-03 | Esophagus_Mucosa | 5.23 | 4.24E-02 | 0.24 | -3.38 | 0.81 | -1.44 | 1.14 | 4 |
| ARF6 | 5.62E-03 | 27 | 2 | 1.29E-03 | Esophagus_Mucosa | 4.80 | 2.05E-17 | 0.18 | 2.66 | 3.22 | 2.77 | 0.14 | 2 |
| ORC5 | 5.62E-03 | 29 | 2 | 3.06E-01 | Colon_Sigmoid | 2.78 | 1.80E-17 | 0.22 | -3.41 | 2.75 | -0.97 | 1.69 | 2 |
| GINM1 | 5.63E-03 | 31 | 1 | 5.45E-03 | Esophagus_Gastroesophageal_Junction | 6.68 | 2.72E-17 | 6.68 | -0.16 | 2.80 | 2.56 | 0.57 | 1 |
| PGBD1 | 5.63E-03 | 39 | 4 | 1.71E-03 | Colon_Transverse | 3.80 | 1.24E-16 | 0.27 | -2.20 | 3.77 | 1.51 | 1.85 | 4 |
| MIEF2 | 5.65E-03 | 42 | 3 | 2.35E-02 | Stomach | 4.58 | 1.90E-16 | 0.17 | -3.01 | 2.37 | 0.50 | 1.69 | 3 |
| LINC01224 | 5.66E-03 | 18 | 1 | 5.66E-03 | Esophagus_Mucosa | 2.00 | 3.35E-17 | 2.00 | -2.77 | 2.77 | -1.84 | 1.60 | 1 |
| TWF1 | 5.77E-03 | 39 | 4 | 3.27E-02 | Esophagus_Gastroesophageal_Junction | 4.46 | 2.43E-16 | 0.16 | -1.00 | 2.14 | -0.09 | 0.58 | 4 |
| FAM120AOS | 5.79E-03 | 49 | 4 | 9.92E-03 | Esophagus_Mucosa | 4.00 | 3.03E-16 | 0.46 | -2.58 | 2.29 | 0.60 | 1.46 | 4 |
| ZBTB22 | 5.79E-03 | 33 | 3 | 1.06E-03 | Colon_Transverse | 4.99 | 3.22E-18 | 0.18 | -3.36 | 2.75 | -2.18 | 1.46 | 3 |
| KANK3 | 5.83E-03 | 30 | 3 | 8.35E-03 | Small_Intestine_Terminal_Ileum | 4.66 | 9.43E-18 | 0.55 | -1.76 | 2.64 | 0.91 | 1.37 | 3 |
| P2RX4 | 5.85E-03 | 48 | 3 | 6.15E-02 | Esophagus_Mucosa | 5.99 | 1.95E-16 | 0.38 | -0.45 | 2.78 | 1.14 | 0.95 | 3 |
| PRKCA | 5.88E-03 | 36 | 4 | 1.27E-02 | Colon_Sigmoid | 2.39 | 1.23E-16 | 0.61 | -2.49 | 3.45 | 0.88 | 1.52 | 4 |
| CTDNEP1 | 5.91E-03 | 46 | 1 | 5.15E-03 | Esophagus_Gastroesophageal_Junction | 5.87 | 1.59E-33 | 5.87 | -3.33 | 3.39 | 2.72 | 0.93 | 1 |
| PAMR1 | 5.92E-03 | 35 | 6 | 1.47E-04 | Colon_Sigmoid | 2.93 | 2.98E-16 | 0.10 | -3.80 | 3.80 | 0.85 | 2.18 | 6 |
| EFTUD2 | 5.92E-03 | 48 | 4 | 2.51E-04 | Esophagus_Gastroesophageal_Junction | 4.11 | 2.41E-16 | 0.18 | -3.68 | 3.70 | 0.29 | 2.41 | 4 |
| SECISBP2 | 5.92E-03 | 47 | 3 | 5.91E-04 | Colon_Transverse | 3.96 | 1.72E-33 | 0.64 | -0.49 | 3.47 | 2.69 | 1.16 | 3 |

| | | | | | | | | | | | | | |
|---|---|---|---|---|---|---|---|---|---|---|---|---|---|
| SLC37A2 | 5.93E-03 | 41 | 4 | 1.53E-02 | Esophagus_Mucosa | 2.55 | 9.52E-18 | 0.95 | -1.68 | 2.97 | 0.68 | 1.65 | 4 |
| STX3 | 5.94E-03 | 45 | 3 | 1.20E-01 | Colon_Transverse | 5.61 | 3.19E-17 | 0.22 | -2.53 | 1.59 | 0.08 | 1.37 | 3 |
| TPK1 | 5.96E-03 | 49 | 2 | 5.13E-04 | Small_Intestine_Terminal_Ileum | 6.00 | 2.59E-17 | 0.81 | -3.49 | 1.13 | -2.50 | 0.99 | 2 |
| SP5 | 6.02E-03 | 34 | 2 | 7.78E-03 | Esophagus_Gastroesophageal_Junction | 4.39 | 2.27E-17 | 0.61 | -2.66 | 2.66 | 0.35 | 1.97 | 2 |
| SAYSD1 | 6.02E-03 | 45 | 4 | 3.09E-03 | Esophagus_Muscularis | 3.24 | 1.12E-16 | 0.74 | -2.51 | 2.96 | -0.48 | 1.40 | 4 |
| TOP3A | 6.02E-03 | 49 | 3 | 2.12E-02 | Colon_Transverse | 5.59 | 8.20E-17 | 0.29 | -0.87 | 2.54 | 1.45 | 0.87 | 3 |
| FAF2 | 6.04E-03 | 4 | 1 | 6.04E-03 | Esophagus_Mucosa | 1.00 | 1.00E+00 | 1.00 | -2.75 | 2.75 | -0.27 | 2.27 | 1 |
| SPATS2 | 6.05E-03 | 29 | 2 | 1.41E-03 | Esophagus_Mucosa | 2.06 | 1.23E-32 | 0.94 | -3.19 | 3.42 | 0.38 | 1.48 | 2 |
| AFF3 | 6.07E-03 | 31 | 2 | 1.45E-03 | Esophagus_Gastroesophageal_Junction | 2.98 | 2.67E-18 | 0.99 | -3.49 | 2.85 | -1.84 | 2.10 | 2 |
| SLC25A12 | 6.12E-03 | 39 | 4 | 5.31E-04 | Colon_Transverse | 3.84 | 1.03E-03 | 0.21 | -4.00 | 3.32 | -1.61 | 1.90 | 4 |
| HYAL1 | 6.12E-03 | 11 | 1 | 6.12E-03 | Colon_Transverse | 1.00 | 1.00E+00 | 1.00 | -1.02 | 2.74 | 0.29 | 1.29 | 1 |
| PAQR7 | 6.13E-03 | 13 | 1 | 6.13E-03 | Esophagus_Gastroesophageal_Junction | 2.00 | 6.12E-17 | 2.00 | -2.83 | 0.43 | -2.24 | 1.00 | 1 |
| STYX | 6.13E-03 | 49 | 2 | 1.22E-03 | Colon_Sigmoid | 6.53 | 1.63E-17 | 0.42 | -3.25 | 2.97 | -2.74 | 1.15 | 2 |
| SOCS5 | 6.15E-03 | 27 | 1 | 5.99E-03 | Esophagus_Gastroesophageal_Junction | 1.99 | 6.42E-03 | 1.99 | -2.49 | 2.81 | 1.15 | 1.53 | 1 |
| RP1-265C24.8 | 6.16E-03 | 20 | 2 | 1.44E-03 | Stomach | 3.03 | 2.90E-18 | 0.97 | -0.23 | 3.19 | 2.34 | 1.32 | 2 |
| SYTL1 | 6.16E-03 | 45 | 2 | 5.80E-03 | Esophagus_Mucosa | 3.07 | 8.93E-17 | 1.91 | -2.84 | 3.53 | 2.07 | 1.63 | 2 |
| DDAH2 | 6.18E-03 | 44 | 5 | 4.18E-04 | Esophagus_Gastroesophageal_Junction | 2.95 | 2.60E-16 | 0.15 | -3.53 | -0.02 | -2.59 | 1.18 | 5 |
| GUCA1C | 6.27E-03 | 11 | 1 | 6.27E-03 | Stomach | 1.00 | 1.00E+00 | 1.00 | -1.96 | 2.73 | 0.37 | 1.46 | 1 |
| MAN2A1 | 6.28E-03 | 41 | 5 | 2.62E-02 | Small_Intestine_Terminal_Ileum | 2.02 | 6.37E-17 | 0.65 | -2.22 | 2.91 | 0.03 | 1.63 | 5 |
| PARK2 | 6.33E-03 | 42 | 3 | 1.02E-03 | Stomach | 3.08 | 2.51E-17 | 0.84 | -3.36 | 0.92 | -2.00 | 1.14 | 3 |
| RP11-579E24.2 | 6.34E-03 | 3 | 1 | 6.34E-03 | Colon_Sigmoid | 2.00 | 3.35E-17 | 2.00 | -2.73 | -2.73 | -2.73 | 0.00 | 1 |
| PIK3R3 | 6.38E-03 | 45 | 2 | 1.59E-03 | Colon_Sigmoid | 3.26 | 1.02E-16 | 2.74 | -3.30 | 2.51 | -1.70 | 1.73 | 2 |
| CCND1 | 6.40E-03 | 18 | 1 | 6.40E-03 | Colon_Transverse | 3.00 | 3.85E-17 | 3.00 | -2.73 | 2.73 | -2.18 | 1.41 | 1 |
| REPS1 | 6.40E-03 | 33 | 2 | 1.52E-03 | Colon_Transverse | 3.05 | 5.17E-17 | 1.95 | -1.92 | 3.17 | 1.61 | 1.46 | 2 |
| OLA1 | 6.46E-03 | 31 | 4 | 6.80E-02 | Esophagus_Muscularis | 2.40 | 1.12E-01 | 0.11 | -2.55 | 1.54 | -0.72 | 0.85 | 4 |
| GAREM2 | 6.47E-03 | 49 | 4 | 1.26E-01 | Colon_Transverse | 5.74 | 3.31E-03 | 0.22 | -1.81 | 2.06 | 0.04 | 0.92 | 4 |
| DLEU1 | 6.55E-03 | 48 | 2 | 1.21E-02 | Colon_Sigmoid | 5.84 | 2.24E-17 | 1.05 | -2.22 | 2.51 | -1.06 | 1.69 | 2 |
| U2AF1L4 | 6.57E-03 | 49 | 1 | 4.17E-03 | Esophagus_Mucosa | 6.96 | 6.07E-17 | 6.96 | -0.46 | 3.20 | 2.70 | 0.62 | 1 |
| KMT2E | 6.62E-03 | 24 | 2 | 2.80E-03 | Esophagus_Muscularis | 2.54 | 5.11E-17 | 1.46 | -2.24 | 2.99 | 0.27 | 1.58 | 2 |
| ID4 | 6.65E-03 | 14 | 4 | 4.56E-04 | Stomach | 3.32 | 4.76E-18 | 0.18 | -3.51 | 3.51 | 1.75 | 2.36 | 4 |
| SPI1 | 6.65E-03 | 45 | 2 | 1.99E-03 | Esophagus_Muscularis | 4.16 | 3.29E-17 | 1.84 | -3.09 | 3.19 | -1.09 | 1.41 | 2 |
| ECSIT | 6.67E-03 | 47 | 2 | 1.71E-03 | Small_Intestine_Terminal_Ileum | 5.93 | 2.69E-17 | 0.87 | 0.02 | 3.40 | 2.78 | 0.90 | 2 |
| TLR5 | 6.71E-03 | 41 | 4 | 6.26E-03 | Small_Intestine_Terminal_Ileum | 4.78 | 9.92E-18 | 0.23 | -2.73 | 0.93 | -1.81 | 0.96 | 4 |
| C1orf132 | 6.77E-03 | 32 | 2 | 1.88E-03 | Small_Intestine_Terminal_Ileum | 4.02 | 1.78E-19 | 0.98 | -3.11 | 3.19 | 0.89 | 1.20 | 2 |
| GPC6 | 6.80E-03 | 45 | 4 | 1.43E-03 | Esophagus_Muscularis | 3.74 | 4.89E-17 | 0.18 | -2.63 | 4.67 | 1.01 | 1.62 | 4 |

| | | | | | | | | | | | | | |
|---|---|---|---|---|---|---|---|---|---|---|---|---|---|
| LZTS1 | 6.81E-03 | 28 | 2 | 2.24E-03 | Colon_Sigmoid | 2.24 | 3.38E-17 | 0.76 | -3.08 | 1.23 | -1.93 | 1.65 | 2 |
| CMTM1 | 6.85E-03 | 49 | 1 | 6.56E-03 | Colon_Transverse | 6.86 | 1.43E-19 | 6.86 | -3.12 | -1.17 | -2.65 | 0.26 | 1 |
| YPEL4 | 6.87E-03 | 28 | 3 | 2.14E-03 | Colon_Transverse | 4.95 | 6.53E-17 | 0.19 | -3.20 | 1.77 | -2.27 | 1.38 | 3 |
| VPS41 | 6.90E-03 | 44 | 4 | 1.50E-03 | Colon_Sigmoid | 4.09 | 4.75E-02 | 0.27 | -3.17 | 1.21 | -0.61 | 0.89 | 4 |
| NCLN | 6.90E-03 | 48 | 2 | 1.07E-03 | Colon_Sigmoid | 5.82 | 2.77E-05 | 0.90 | 0.36 | 3.27 | 2.74 | 0.41 | 2 |
| STMN1 | 6.92E-03 | 12 | 2 | 2.08E-03 | Esophagus_Mucosa | 1.76 | 2.38E-01 | 0.24 | -2.95 | 3.08 | -1.64 | 2.05 | 2 |
| GMDS-AS1 | 6.93E-03 | 46 | 3 | 1.98E-03 | Small_Intestine_Terminal_Ileum | 4.34 | 1.86E-17 | 0.68 | -3.09 | 3.09 | -1.06 | 1.63 | 3 |
| MAP2K3 | 6.95E-03 | 26 | 1 | 6.95E-03 | Colon_Transverse | 2.00 | 3.35E-17 | 2.00 | -2.70 | 2.70 | -1.70 | 1.96 | 1 |
| RNF222 | 6.96E-03 | 14 | 1 | 6.96E-03 | Esophagus_Mucosa | 1.00 | 1.00E+00 | 1.00 | -3.12 | 2.70 | -0.44 | 2.37 | 1 |
| RP11-367F23.1 | 6.97E-03 | 13 | 2 | 2.17E-03 | Small_Intestine_Terminal_Ileum | 2.84 | 1.29E-16 | 0.16 | -3.34 | -3.04 | -3.08 | 0.09 | 2 |
| FLJ22447 | 7.02E-03 | 21 | 3 | 4.12E-03 | Stomach | 2.25 | 2.14E-01 | 0.21 | -2.87 | 1.02 | -0.99 | 1.00 | 3 |
| LINC01021 | 7.03E-03 | 8 | 1 | 7.03E-03 | Esophagus_Muscularis | 2.00 | 3.53E-16 | 2.00 | -2.70 | 3.04 | -0.63 | 2.85 | 1 |
| HSPA1A | 7.07E-03 | 9 | 2 | 1.70E-03 | Esophagus_Mucosa | 1.22 | 7.83E-01 | 0.78 | -3.14 | 0.92 | -2.28 | 1.71 | 2 |
| TMEM258 | 7.10E-03 | 49 | 2 | 5.07E-02 | Stomach | 6.63 | 7.42E-17 | 0.37 | -3.02 | 0.22 | -1.55 | 0.73 | 2 |
| CIART | 7.13E-03 | 8 | 1 | 7.13E-03 | Colon_Transverse | 3.00 | 2.11E-48 | 3.00 | -2.69 | -2.69 | -2.69 | 0.00 | 1 |
| PARP11 | 7.20E-03 | 49 | 2 | 2.67E-02 | Colon_Sigmoid | 6.49 | 1.34E-16 | 0.23 | -2.22 | 0.99 | -0.91 | 0.83 | 2 |
| RP11-686D22.4 | 7.22E-03 | 46 | 2 | 1.97E-02 | Colon_Sigmoid | 6.46 | 7.89E-03 | 0.31 | -0.56 | 2.38 | 0.78 | 0.81 | 2 |
| EEF1B2 | 7.22E-03 | 20 | 1 | 7.22E-03 | Colon_Sigmoid | 3.00 | 3.85E-17 | 3.00 | -2.69 | 2.69 | -1.34 | 2.38 | 1 |
| NRM | 7.24E-03 | 39 | 3 | 1.94E-03 | Colon_Sigmoid | 2.17 | 8.47E-19 | 1.83 | -3.18 | 1.53 | -0.27 | 1.58 | 3 |
| ACAT1 | 7.28E-03 | 45 | 3 | 1.10E-03 | Esophagus_Muscularis | 3.76 | 7.41E-17 | 0.21 | -3.38 | 1.89 | -1.01 | 1.89 | 3 |
| LRFN5 | 7.34E-03 | 33 | 2 | 3.46E-03 | Esophagus_Gastroesophageal_Junctio n | 4.64 | 1.62E-17 | 0.28 | -3.04 | 3.04 | 0.59 | 2.78 | 2 |
| NEK1 | 7.37E-03 | 49 | 3 | 1.64E-03 | Small_Intestine_Terminal_Ileum | 4.80 | 6.06E-17 | 0.85 | -3.15 | 3.16 | -1.46 | 1.14 | 3 |
| PPP1R36 | 7.38E-03 | 47 | 5 | 1.30E-02 | Esophagus_Muscularis | 3.22 | 1.16E-16 | 0.22 | -1.26 | 2.57 | 0.07 | 0.91 | 5 |
| PTBP3 | 7.44E-03 | 39 | 2 | 4.18E-03 | Colon_Transverse | 5.45 | 4.40E-17 | 0.55 | -2.88 | 3.11 | 2.50 | 1.29 | 2 |
| CNTN1 | 7.45E-03 | 39 | 5 | 1.26E-03 | Esophagus_Mucosa | 2.50 | 1.95E-01 | 0.19 | -2.80 | 3.34 | 0.26 | 1.52 | 5 |
| SLFNL1 | 7.58E-03 | 48 | 2 | 8.27E-04 | Esophagus_Gastroesophageal_Junctio n | 4.49 | 1.08E-16 | 1.42 | -2.14 | 3.61 | 2.33 | 1.62 | 2 |
| FOXH1 | 7.61E-03 | 13 | 1 | 7.61E-03 | Colon_Transverse | 1.00 | 1.00E+00 | 1.00 | 2.67 | 2.67 | 2.67 | 0.00 | 1 |
| HTR1E | 7.63E-03 | 18 | 3 | 1.33E-02 | Colon_Sigmoid | 1.66 | 2.99E-01 | 0.30 | -2.48 | 2.48 | 0.34 | 1.84 | 3 |
| PPP1R3A | 7.63E-03 | 6 | 1 | 7.63E-03 | Esophagus_Muscularis | 1.00 | 1.00E+00 | 1.00 | -2.67 | 0.74 | -1.56 | 1.38 | 1 |
| UPF2 | 7.65E-03 | 40 | 4 | 1.93E-03 | Small_Intestine_Terminal_Ileum | 4.62 | 2.12E-16 | 0.21 | -1.33 | 3.10 | 1.20 | 1.15 | 4 |
| FES | 7.65E-03 | 46 | 2 | 2.14E-02 | Esophagus_Mucosa | 5.96 | 3.13E-16 | 0.77 | -3.15 | 0.78 | -1.03 | 1.32 | 2 |
| AP1B1 | 7.66E-03 | 30 | 3 | 1.17E-03 | Esophagus_Muscularis | 4.02 | 5.42E-17 | 0.41 | -3.25 | 2.38 | 1.00 | 2.13 | 3 |
| MUC22 | 7.67E-03 | 2 | 1 | 7.67E-03 | Esophagus_Mucosa | 1.00 | 1.00E+00 | 1.00 | -2.67 | 0.47 | -1.10 | 2.22 | 1 |
| ABHD17C | 7.69E-03 | 27 | 2 | 2.40E-03 | Stomach | 1.04 | 9.60E-01 | 0.96 | -1.31 | 3.04 | 0.88 | 1.49 | 2 |
| CEP95 | 7.69E-03 | 43 | 2 | 1.44E-03 | Esophagus_Gastroesophageal_Junctio n | 6.38 | 5.81E-17 | 0.55 | -1.79 | 3.48 | 2.74 | 1.19 | 2 |
| GLRB | 7.70E-03 | 37 | 4 | 1.17E-03 | Colon_Sigmoid | 3.82 | 7.97E-18 | 0.18 | -3.25 | 3.18 | -0.22 | 2.46 | 4 |

| | | | | | | | | | | | | | |
|---|---|---|---|---|---|---|---|---|---|---|---|---|---|
| CYP3A5 | 7.75E-03 | 46 | 2 | 1.97E-03 | Colon_Sigmoid | 6.01 | 4.69E-17 | 0.99 | -3.09 | 3.24 | 2.53 | 1.45 | 2 |
| CPAMD8 | 7.76E-03 | 11 | 1 | 7.76E-03 | Esophagus_Muscularis | 1.00 | 1.00E+00 | 1.00 | -0.75 | 2.66 | 0.57 | 1.01 | 1 |
| ZNF200 | 7.76E-03 | 43 | 3 | 1.05E-02 | Esophagus_Muscularis | 4.82 | 1.04E-05 | 0.18 | -1.14 | 2.56 | 1.09 | 0.91 | 3 |
| ATP11A | 7.77E-03 | 15 | 1 | 5.25E-03 | Esophagus_Muscularis | 3.88 | 3.83E-03 | 3.88 | -2.11 | 2.79 | 1.64 | 1.54 | 1 |
| PIK3R5 | 7.78E-03 | 35 | 2 | 4.73E-03 | Colon_Sigmoid | 2.08 | 1.29E-16 | 0.92 | -2.82 | 2.82 | 0.96 | 1.48 | 2 |
| STX12 | 7.80E-03 | 1 | 1 | 7.80E-03 | Colon_Sigmoid | 1.00 | 1.00E+00 | 1.00 | 2.66 | 2.66 | 2.66 | NA | 1 |
| FURIN | 7.88E-03 | 39 | 4 | 1.49E-03 | Esophagus_Muscularis | 2.00 | 1.34E-01 | 0.13 | -3.77 | 3.33 | 1.83 | 2.05 | 4 |
| ASIC4 | 7.91E-03 | 23 | 3 | 2.36E-03 | Small_Intestine_Terminal_Ileum | 3.11 | 3.31E-17 | 0.14 | -3.24 | 3.24 | -1.03 | 1.53 | 3 |
| HEXDC | 7.92E-03 | 47 | 2 | 3.98E-02 | Colon_Sigmoid | 6.38 | 2.22E-16 | 0.44 | -3.58 | 0.17 | -0.94 | 0.92 | 2 |
| MDFIC | 7.94E-03 | 37 | 3 | 3.65E-02 | Esophagus_Mucosa | 4.72 | 7.43E-18 | 0.35 | -2.09 | 3.43 | 0.63 | 1.03 | 3 |
| ZNF184 | 7.94E-03 | 40 | 1 | 7.83E-03 | Colon_Transverse | 6.00 | 3.92E-38 | 6.00 | -2.79 | 2.66 | -2.33 | 1.12 | 1 |
| PRMT6 | 7.95E-03 | 48 | 1 | 7.33E-03 | Stomach | 6.81 | 4.09E-17 | 6.81 | -2.90 | 2.68 | -2.48 | 0.78 | 1 |
| ZNF304 | 8.02E-03 | 49 | 1 | 5.69E-03 | Colon_Sigmoid | 6.76 | 3.70E-17 | 6.76 | -0.06 | 2.77 | 2.37 | 0.49 | 1 |
| HFE | 8.08E-03 | 40 | 2 | 1.77E-01 | Small_Intestine_Terminal_Ileum | 4.96 | 2.30E-17 | 0.83 | -1.36 | 1.35 | -0.40 | 0.94 | 2 |
| ZNF726 | 8.08E-03 | 45 | 4 | 4.26E-03 | Small_Intestine_Terminal_Ileum | 4.08 | 1.35E-16 | 0.16 | -2.86 | 2.26 | -0.08 | 0.99 | 4 |
| LYL1 | 8.10E-03 | 16 | 1 | 8.10E-03 | Colon_Transverse | 1.00 | 1.00E+00 | 1.00 | -2.65 | 2.65 | -1.73 | 1.79 | 1 |
| HSPG2 | 8.10E-03 | 33 | 3 | 1.47E-03 | Colon_Transverse | 3.99 | 2.22E-16 | 0.17 | -3.23 | 0.25 | -1.94 | 1.28 | 3 |
| DNASE1 | 8.12E-03 | 45 | 3 | 2.99E-02 | Esophagus_Muscularis | 5.10 | 3.95E-17 | 0.20 | -1.95 | 3.65 | 0.44 | 1.83 | 3 |
| CDCA7 | 8.12E-03 | 41 | 5 | 8.49E-04 | Esophagus_Muscularis | 2.94 | 1.05E-01 | 0.10 | -3.34 | 1.11 | -0.14 | 1.25 | 5 |
| EGFL8 | 8.12E-03 | 45 | 3 | 3.01E-02 | Esophagus_Gastroesophageal_Junction | 4.88 | 2.11E-16 | 0.96 | -0.97 | 2.17 | 0.93 | 0.81 | 3 |
| CEBPZOS | 8.16E-03 | 48 | 2 | 9.93E-04 | Esophagus_Mucosa | 6.05 | 3.05E-02 | 0.56 | -3.30 | 1.55 | -2.21 | 0.88 | 2 |
| CTBP2 | 8.17E-03 | 35 | 5 | 5.42E-03 | Stomach | 3.09 | 6.31E-17 | 0.14 | -3.41 | 2.79 | 0.14 | 1.15 | 5 |
| RBM23 | 8.17E-03 | 49 | 3 | 1.13E-02 | Stomach | 5.86 | 8.03E-07 | 0.35 | -3.16 | 2.20 | -0.87 | 1.49 | 3 |
| ZNF800 | 8.19E-03 | 41 | 3 | 9.74E-04 | Stomach | 3.00 | 1.19E-17 | 0.94 | -3.30 | 0.59 | -1.54 | 1.53 | 3 |
| LINC01146 | 8.19E-03 | 18 | 3 | 1.55E-03 | Stomach | 1.74 | 2.95E-01 | 0.29 | -2.49 | 3.16 | 1.03 | 1.29 | 3 |
| MRPL35 | 8.30E-03 | 49 | 3 | 2.42E-03 | Esophagus_Mucosa | 5.20 | 5.60E-03 | 0.44 | 0.01 | 3.31 | 2.09 | 0.77 | 3 |
| APOLD1 | 8.35E-03 | 20 | 1 | 8.35E-03 | Colon_Transverse | 3.00 | 2.11E-48 | 3.00 | -2.66 | -1.39 | -2.36 | 0.46 | 1 |
| TGFBR1 | 8.38E-03 | 11 | 1 | 8.38E-03 | Stomach | 1.00 | 1.00E+00 | 1.00 | -3.40 | 3.40 | -0.75 | 2.86 | 1 |
| CDCA2 | 8.38E-03 | 23 | 1 | 7.32E-03 | Esophagus_Mucosa | 4.00 | 1.18E-21 | 4.00 | -2.59 | 2.68 | 2.31 | 1.19 | 1 |
| AC012442.6 | 8.38E-03 | 21 | 1 | 8.38E-03 | Esophagus_Muscularis | 1.00 | 1.00E+00 | 1.00 | -2.84 | 1.51 | -1.81 | 1.50 | 1 |
| CLEC2L | 8.40E-03 | 3 | 1 | 8.40E-03 | Esophagus_Muscularis | 1.00 | 1.00E+00 | 1.00 | -0.79 | 2.64 | 0.57 | 1.82 | 1 |
| BBS9 | 8.41E-03 | 46 | 2 | 2.59E-03 | Small_Intestine_Terminal_Ileum | 4.55 | 5.41E-17 | 2.41 | -2.08 | 3.05 | 0.97 | 1.39 | 2 |
| MEIOC | 8.42E-03 | 24 | 2 | 5.34E-03 | Esophagus_Gastroesophageal_Junction | 3.12 | 3.68E-33 | 0.78 | -3.06 | -1.51 | -2.55 | 0.44 | 2 |
| TSTD3 | 8.46E-03 | 37 | 4 | 2.34E-02 | Esophagus_Mucosa | 2.20 | 1.01E-01 | 0.10 | -2.41 | 2.12 | -0.65 | 1.46 | 4 |
| SFMBT2 | 8.53E-03 | 41 | 4 | 3.21E-03 | Colon_Transverse | 3.39 | 8.52E-18 | 0.40 | -3.31 | 2.95 | -0.16 | 1.38 | 4 |
| PROCR | 8.55E-03 | 29 | 3 | 1.84E-03 | Colon_Sigmoid | 2.30 | 1.56E-02 | 0.49 | -3.11 | 3.11 | 0.02 | 1.86 | 3 |

| | | | | | | | | | | | | | |
|---|---|---|---|---|---|---|---|---|---|---|---|---|---|
| ASGR1 | 8.56E-03 | 18 | 2 | 4.47E-03 | Esophagus_Muscularis | 1.15 | 8.50E-01 | 0.85 | -2.84 | 0.76 | -0.09 | 0.75 | 2 |
| CTD-2306A12.1 | 8.58E-03 | 32 | 1 | 5.89E-03 | Esophagus_Gastroesophageal_Junction | 6.80 | 2.90E-17 | 6.80 | -2.83 | -1.58 | -2.52 | 0.26 | 1 |
| RNF114 | 8.58E-03 | 42 | 2 | 4.87E-03 | Stomach | 3.02 | 3.10E-16 | 1.96 | -1.25 | 3.34 | 1.88 | 1.12 | 2 |
| RNF5 | 8.64E-03 | 45 | 2 | 1.50E-03 | Colon_Sigmoid | 5.16 | 4.01E-16 | 0.60 | -3.22 | 0.66 | -2.26 | 1.11 | 2 |
| CMTM3 | 8.65E-03 | 39 | 2 | 2.29E-03 | Esophagus_Mucosa | 5.08 | 1.66E-17 | 0.84 | -3.11 | -0.58 | -2.27 | 0.64 | 2 |
| ADTRP | 8.72E-03 | 38 | 5 | 1.27E-02 | Esophagus_Gastroesophageal_Junction | 2.76 | 2.23E-17 | 0.24 | -2.49 | 2.03 | -0.71 | 1.59 | 5 |
| CCL23 | 8.72E-03 | 25 | 2 | 6.56E-03 | Esophagus_Muscularis | 2.24 | 4.39E-17 | 0.76 | 0.29 | 2.72 | 2.12 | 1.00 | 2 |
| CTB-31O20.2 | 8.75E-03 | 32 | 2 | 7.12E-02 | Stomach | 5.64 | 5.06E-17 | 0.27 | -2.29 | 0.89 | -0.80 | 0.92 | 2 |
| ZNRD1 | 8.79E-03 | 47 | 5 | 1.89E-02 | Colon_Transverse | 4.38 | 5.44E-02 | 0.15 | -2.35 | 2.74 | 0.16 | 1.19 | 5 |
| C6orf226 | 8.79E-03 | 49 | 1 | 5.34E-03 | Esophagus_Muscularis | 6.88 | 8.54E-18 | 6.88 | 1.37 | 2.79 | 2.72 | 0.28 | 1 |
| ZNF594 | 8.79E-03 | 33 | 3 | 5.06E-02 | Colon_Sigmoid | 3.21 | 2.43E-16 | 0.27 | -1.95 | 1.35 | 0.24 | 0.88 | 3 |
| HTRA1 | 8.82E-03 | 37 | 3 | 4.48E-03 | Esophagus_Gastroesophageal_Junction | 5.46 | 4.27E-19 | 0.33 | -2.84 | 2.12 | -1.50 | 1.49 | 3 |
| ZNF766 | 8.87E-03 | 40 | 4 | 1.57E-03 | Esophagus_Mucosa | 2.15 | 1.43E-01 | 0.14 | -1.30 | 3.42 | 0.92 | 1.86 | 4 |
| CTA-989H11.1 | 8.92E-03 | 26 | 2 | 1.42E-03 | Colon_Sigmoid | 5.42 | 2.45E-19 | 0.54 | -1.45 | 3.31 | 2.68 | 1.28 | 2 |
| OTUB2 | 9.03E-03 | 16 | 3 | 6.61E-03 | Colon_Sigmoid | 1.98 | 1.21E-01 | 0.12 | -2.72 | 1.25 | -0.38 | 1.43 | 3 |
| GLT1D1 | 9.04E-03 | 43 | 5 | 4.24E-02 | Colon_Transverse | 3.51 | 5.32E-17 | 0.21 | -2.03 | 2.85 | 0.39 | 1.21 | 5 |
| DLC1 | 9.11E-03 | 24 | 2 | 2.43E-03 | Esophagus_Gastroesophageal_Junction | 3.10 | 7.75E-17 | 0.90 | -3.06 | 2.86 | -0.13 | 1.41 | 2 |
| DBN1 | 9.13E-03 | 29 | 4 | 3.08E-04 | Esophagus_Mucosa | 2.74 | 1.36E-01 | 0.14 | -3.61 | 3.61 | -0.35 | 2.96 | 4 |
| AMD1 | 9.21E-03 | 37 | 2 | 2.20E-03 | Colon_Sigmoid | 4.05 | 2.17E-18 | 2.95 | -3.06 | 3.53 | 1.80 | 2.01 | 2 |
| HMX1 | 9.22E-03 | 16 | 2 | 5.77E-03 | Colon_Sigmoid | 1.08 | 9.22E-01 | 0.92 | -3.10 | -0.06 | -2.08 | 1.07 | 2 |
| ALDH1A1 | 9.23E-03 | 29 | 1 | 9.23E-03 | Colon_Sigmoid | 5.00 | 2.06E-67 | 5.00 | -3.26 | 2.60 | -2.18 | 1.38 | 1 |
| LYSMD1 | 9.31E-03 | 32 | 4 | 5.10E-03 | Small_Intestine_Terminal_Ileum | 4.05 | 1.04E-17 | 0.18 | -3.36 | 3.36 | 1.26 | 1.72 | 4 |
| CHCHD5 | 9.36E-03 | 48 | 3 | 1.16E-04 | Stomach | 5.48 | 1.48E-16 | 0.58 | -3.86 | -0.84 | -2.53 | 0.56 | 3 |
| PFDN6 | 9.38E-03 | 9 | 1 | 9.38E-03 | Colon_Transverse | 2.00 | 3.35E-17 | 2.00 | -2.60 | -2.60 | -2.60 | 0.00 | 1 |
| SOX1 | 9.39E-03 | 4 | 1 | 9.39E-03 | Esophagus_Gastroesophageal_Junction | 1.00 | 1.00E+00 | 1.00 | -1.82 | 2.60 | -0.37 | 2.01 | 1 |
| MOCOS | 9.39E-03 | 48 | 3 | 5.40E-03 | Esophagus_Muscularis | 5.82 | 8.16E-03 | 0.33 | -1.89 | 3.26 | 1.79 | 0.94 | 3 |
| CD34 | 9.43E-03 | 43 | 3 | 8.92E-04 | Colon_Sigmoid | 5.02 | 2.80E-33 | 0.40 | -3.32 | 0.29 | -0.44 | 0.57 | 3 |
| TIMM10 | 9.45E-03 | 49 | 2 | 2.72E-03 | Esophagus_Mucosa | 6.52 | 2.01E-04 | 0.23 | -0.72 | 3.00 | 2.48 | 0.63 | 2 |
| AC007405.6 | 9.47E-03 | 24 | 2 | 7.35E-03 | Stomach | 1.52 | 4.80E-01 | 0.48 | -3.46 | 0.14 | -2.26 | 1.11 | 2 |
| TAOK1 | 9.47E-03 | 26 | 2 | 3.04E-03 | Esophagus_Mucosa | 2.04 | 7.85E-17 | 0.96 | -3.18 | 3.18 | 0.59 | 1.80 | 2 |
| RP11-51J9.5 | 9.47E-03 | 43 | 1 | 8.27E-03 | Colon_Transverse | 5.86 | 1.03E-16 | 5.86 | -3.32 | -1.15 | -2.57 | 0.30 | 1 |
| ERICH2 | 9.48E-03 | 37 | 3 | 7.52E-03 | Colon_Sigmoid | 2.97 | 1.25E-16 | 0.80 | -3.08 | 2.88 | -0.10 | 1.58 | 3 |
| RP11-718O11.1 | 9.53E-03 | 3 | 1 | 9.53E-03 | Colon_Sigmoid | 1.00 | 1.00E+00 | 1.00 | -2.59 | -0.16 | -0.99 | 1.38 | 1 |
| WARS2 | 9.54E-03 | 49 | 3 | 6.34E-03 | Colon_Transverse | 5.51 | 3.59E-03 | 0.29 | -2.73 | 1.19 | -0.84 | 0.89 | 3 |
| SIRPB1 | 9.55E-03 | 49 | 2 | 1.94E-03 | Colon_Transverse | 6.00 | 3.82E-17 | 0.95 | -3.35 | 0.93 | -2.43 | 0.86 | 2 |

| | | | | | | | | | | | | | |
|---|---|---|---|---|---|---|---|---|---|---|---|---|---|
| MCM7 | 9.59E-03 | 41 | 3 | 3.90E-02 | Esophagus_Mucosa | 4.71 | 1.10E-17 | 0.22 | -2.76 | 1.74 | -1.33 | 0.84 | 3 |
| CPSF6 | 9.61E-03 | 37 | 1 | 9.61E-03 | Colon_Sigmoid | 7.00 | 1.14E-65 | 7.00 | 0.28 | 2.59 | 2.31 | 0.71 | 1 |
| AOAH | 9.62E-03 | 45 | 3 | 1.15E-02 | Esophagus_Gastroesophageal_Junction | 5.33 | 8.68E-05 | 0.34 | -2.75 | 2.68 | -0.47 | 1.25 | 3 |
| RERE | 9.68E-03 | 20 | 2 | 2.33E-03 | Esophagus_Mucosa | 1.27 | 7.29E-01 | 0.73 | 0.75 | 3.04 | 2.24 | 0.72 | 2 |
| ITPR3 | 9.72E-03 | 47 | 4 | 8.21E-03 | Esophagus_Gastroesophageal_Junction | 5.40 | 4.92E-02 | 0.20 | -2.26 | 2.69 | -0.12 | 1.07 | 4 |
| LPGAT1 | 9.73E-03 | 49 | 2 | 3.09E-02 | Small_Intestine_Terminal_Ileum | 4.44 | 1.43E-17 | 2.56 | -3.01 | 2.68 | 0.67 | 1.75 | 2 |
| ACTR1A | 9.73E-03 | 46 | 4 | 3.72E-04 | Esophagus_Mucosa | 3.68 | 8.63E-19 | 0.31 | -3.56 | 3.56 | 0.30 | 2.47 | 4 |
| PKNOX1 | 9.79E-03 | 38 | 3 | 2.12E-02 | Colon_Transverse | 4.23 | 1.84E-18 | 0.24 | -2.82 | 2.30 | 0.15 | 1.08 | 3 |
| SLC25A16 | 9.81E-03 | 46 | 3 | 5.66E-03 | Esophagus_Mucosa | 5.93 | 1.42E-16 | 0.20 | -3.05 | -0.62 | -2.30 | 0.60 | 3 |
| SULT1C2 | 9.83E-03 | 47 | 2 | 3.09E-03 | Stomach | 6.23 | 4.61E-03 | 0.45 | -0.42 | 2.96 | 1.85 | 0.56 | 2 |
| ZNF565 | 9.86E-03 | 40 | 2 | 4.29E-03 | Esophagus_Muscularis | 5.99 | 4.07E-17 | 0.97 | -0.60 | 2.87 | 1.68 | 1.14 | 2 |
| RBMXL1 | 9.89E-03 | 45 | 2 | 2.29E-03 | Colon_Sigmoid | 5.62 | 5.94E-18 | 1.28 | 1.41 | 3.05 | 2.34 | 0.65 | 2 |
| RHPN1 | 9.89E-03 | 45 | 3 | 8.03E-02 | Colon_Transverse | 6.06 | 7.39E-03 | 0.24 | -0.99 | 3.22 | 0.65 | 0.99 | 3 |
| IL17RC | 9.90E-03 | 49 | 3 | 1.71E-02 | Colon_Sigmoid | 5.38 | 2.52E-02 | 0.28 | -1.40 | 2.49 | -0.18 | 0.95 | 3 |
| RP11-94C24.13 | 9.92E-03 | 28 | 2 | 3.68E-03 | Small_Intestine_Terminal_Ileum | 3.28 | 1.88E-18 | 0.72 | -3.04 | 3.27 | -1.15 | 1.67 | 2 |
| AC005546.2 | 9.95E-03 | 1 | 1 | 9.95E-03 | Esophagus_Mucosa | 1.00 | 1.00E+00 | 1.00 | -2.58 | -2.58 | -2.58 | NA | 1 |
| ELP2 | 9.96E-03 | 38 | 4 | 1.25E-02 | Colon_Sigmoid | 3.13 | 8.94E-17 | 0.43 | -2.50 | 2.51 | 0.05 | 1.70 | 4 |
| RP11-700H6.1 | 9.97E-03 | 23 | 1 | 4.52E-03 | Esophagus_Muscularis | 5.82 | 1.10E-18 | 5.82 | 0.46 | 2.84 | 2.35 | 0.54 | 1 |
| RP5-994D16.3 | 9.97E-03 | 46 | 1 | 1.01E-02 | Esophagus_Mucosa | 5.82 | 5.30E-22 | 5.82 | -2.59 | 0.24 | -2.47 | 0.41 | 1 |

Pvalue, significance p-value of S-MultiXcan association; n, number of "tissues" available for this gene; n_indep, number of independent components of variation kept among the tissues' predictions. (Synthetic independent tissues); p_i_best, best p-value of single-tissue S-PrediXcan association; t_i_best, name of best single-tissue S-PrediXcan association; eigen_max, In the SVD decomposition of predicted expression correlation: eigenvalue (variance explained) of the top independent component; eigen_min, In the SVD decomposition of predicted expression correlation: eigenvalue (variance explained) of the last independent component; eigen_min_kept, In the SVD decomposition of predicted expression correlation: eigenvalue (variance explained) of the smalles independent component that was kept; z_min, minimum z-score among single-tissue S-Predican associations; z_max, maximum z-score among single-tissue S-Predican associations; z_mean, mean z-score among single-tissue S-Predican associations; z_sd, standard deviation of the mean z-score among single-tissue S-Predican associations; tmi, trace of T * T', where Tis correlation of predicted expression levels for different tissues multiplied by its SVD pseudo-inverse.

**Table S17**. Gene co-expression modules enriched with TWAS-identified genes across multiple tissues.

| Tissue (Spredixcan) | GeneNum (p<0.01) | ModuleIndex(wgcna) | p (fisher) | GeneNum | Gost |
|---|---|---|---|---|---|
| Brain | 640 | Brain - Amygdala _ M10 | 0.031 | 462 | Metabolic module |
| | | Brain - Amygdala _ M12 | 0.008 | 364 | Metabolic module |
| | | Brain - Cerebellar Hemisphere _ M5 | 0.035 | 480 | Neural module |
| | | Brain - Frontal Cortex (BA9) _ M5 | 0.022 | 1181 | Neural module |
| | | Brain - Hippocampus _M3 | 0.021 | 1551 | Metabolic/Neural module |
| | | Brain - Hippocampus _M16 | 0.034 | 182 | Metabolic/Neural module |
| Adipose | 464 | Adipose - Subcutaneous _M10 | 0.002 | 636 | Metabolic/Neural module |
| | | Adipose - Subcutaneous _M12 | 0.027 | 549 | Neural module |
| | | Adipose - Visceral (Omentum) _ M3 | 0.013 | 2479 | Metabolic module |
| | | Adipose - Visceral (Omentum) _M12 | 0.035 | 165 | Neural module |
| Thyroid | 364 | No significant results | | | |
| Gastrointestinal tissues | 576 | Esophagus - Gastroesophageal Junction _ M2 | 0.009 | 4094 | Metabolic/Neural module |
| | | Esophagus - Mucosa _M2 | 0.008 | 3237 | Metabolic/Neural module |
| | | Esophagus - Muscularis _M9 | 0.024 | 297 | Neural module |
| | | Esophagus - Muscularis _M14 | 0.016 | 154 | Neural module |

GeneNum ($p < 0.01$): number of TWAS-identified genes associated with anorexia nervosa in the corresponding tissues at a significance threshold of $p < 0.01$; ModuleIndex (wgcna): gene modules significantly enriched with TWAS-identified genes; p (Fisher): p-value from Fisher's exact test; GeneNum: number of genes in the gene module; Gost: summary of the ontology analysis.

**Table S18**. Enriched GO and KEGG Pathways for co-expression modules identified in each tissue.

| Tissue | Module | Category | ID | Description | P value |
|---|---|---|---|---|---|
| Brain | Brain - Amygdala _ M10 | GO:BP | GO:0019219 | regulation of nucleobase-containing compound metabolic process | 4.24E-31 |
| | | GO:BP | GO:0051252 | regulation of RNA metabolic process | 8.85E-27 |
| | | GO:BP | GO:0080090 | regulation of primary metabolic process | 1.64E-21 |
| | | GO:BP | GO:0051171 | regulation of nitrogen compound metabolic process | 1.74E-21 |
| | | GO:BP | GO:0031323 | regulation of cellular metabolic process | 6.35E-15 |
| | | GO:BP | GO:0060255 | regulation of macromolecule metabolic process | 1.48E-14 |
| | | GO:BP | GO:0019222 | regulation of metabolic process | 8.18E-13 |
| | | GO:BP | GO:0045935 | positive regulation of nucleobase-containing compound metabolic process | 1.30E-12 |
| | | GO:BP | GO:0034641 | cellular nitrogen compound metabolic process | 4.36E-12 |
| | | GO:BP | GO:0006139 | nucleobase-containing compound metabolic process | 8.01E-12 |
| | | GO:BP | GO:0006807 | nitrogen compound metabolic process | 1.20E-11 |
| | | GO:BP | GO:0051254 | positive regulation of RNA metabolic process | 2.04E-11 |
| | | GO:BP | GO:0090304 | nucleic acid metabolic process | 3.89E-11 |
| | | GO:BP | GO:0046483 | heterocycle metabolic process | 5.06E-11 |
| | | GO:BP | GO:0006725 | cellular aromatic compound metabolic process | 5.09E-11 |
| | | GO:BP | GO:0044238 | primary metabolic process | 7.45E-11 |
| | | GO:BP | GO:0045934 | negative regulation of nucleobase-containing compound metabolic process | 3.78E-10 |
| | | GO:BP | GO:1901360 | organic cyclic compound metabolic process | 1.71E-09 |
| | | GO:BP | GO:0051172 | negative regulation of nitrogen compound metabolic process | 6.41E-09 |
| | | GO:BP | GO:0051173 | positive regulation of nitrogen compound metabolic process | 6.67E-09 |
| | | GO:BP | GO:0016070 | RNA metabolic process | 1.56E-08 |
| | | GO:BP | GO:0051253 | negative regulation of RNA metabolic process | 1.49E-07 |
| | | GO:BP | GO:0044237 | cellular metabolic process | 2.33E-07 |
| | | GO:BP | GO:0010604 | positive regulation of macromolecule metabolic process | 3.38E-07 |
| | | GO:BP | GO:0043170 | macromolecule metabolic process | 1.09E-06 |
| | | GO:BP | GO:0008152 | metabolic process | 1.66E-06 |
| | | GO:BP | GO:0009893 | positive regulation of metabolic process | 1.80E-06 |
| | | GO:BP | GO:0031325 | positive regulation of cellular metabolic process | 1.82E-06 |
| | | GO:BP | GO:0071704 | organic substance metabolic process | 3.14E-06 |
| | Brain - Amygdala _ M12 | GO:CC | GO:0005739 | mitochondrion | 5.09E-04 |

| | | | | | |
|---|---|---|---|---|---|
| | | GO:MF | GO:0008757 | S-adenosylmethionine-dependent methyltransferase activity | 0.002 |
| | | GO:CC | GO:0005740 | mitochondrial envelope | 0.002 |
| | | GO:BP | GO:1901564 | organonitrogen compound metabolic process | 0.002 |
| | | GO:CC | GO:0031966 | mitochondrial membrane | 0.002 |
| | Brain - Cerebellar Hemisphere _ M5 | GO:BP | GO:0007399 | nervous system development | 9.45E-16 |
| | | GO:CC | GO:0045202 | synapse | 1.07E-12 |
| | | GO:BP | GO:0022008 | neurogenesis | 9.47E-12 |
| | | GO:BP | GO:0030182 | neuron differentiation | 2.81E-11 |
| | | GO:BP | GO:0048699 | generation of neurons | 4.70E-11 |
| | | GO:CC | GO:0043005 | neuron projection | 1.61E-10 |
| | | GO:BP | GO:0007417 | central nervous system development | 6.27E-08 |
| | | GO:BP | GO:0099536 | synaptic signaling | 5.34E-07 |
| | | GO:BP | GO:0007420 | brain development | 1.57E-06 |
| | | GO:CC | GO:0097060 | synaptic membrane | 2.74E-06 |
| | | GO:BP | GO:0031175 | neuron projection development | 2.99E-05 |
| | | GO:CC | GO:0098978 | glutamatergic synapse | 5.79E-05 |
| | | GO:CC | GO:0098794 | postsynapse | 7.74E-05 |
| | | GO:CC | GO:0045211 | postsynaptic membrane | 9.67E-05 |
| | | GO:BP | GO:0001764 | neuron migration | 5.50E-04 |
| | | GO:CC | GO:0098984 | neuron to neuron synapse | 8.31E-04 |
| | | GO:CC | GO:0014069 | postsynaptic density | 0.001 |
| | | GO:BP | GO:0048167 | regulation of synaptic plasticity | 0.001 |
| | | GO:CC | GO:0032279 | asymmetric synapse | 0.003 |
| | | GO:BP | GO:0048812 | neuron projection morphogenesis | 0.003 |
| | | GO:CC | GO:0099572 | postsynaptic specialization | 0.004 |
| | | GO:BP | GO:0035249 | synaptic transmission, glutamatergic | 0.014 |
| | Brain - Frontal Cortex (BA9) _ M5 | GO:BP | GO:0022008 | neurogenesis | 1.55E-10 |
| | | GO:BP | GO:0007399 | nervous system development | 8.95E-10 |
| | | GO:BP | GO:0008366 | axon ensheathment | 7.97E-09 |
| | | GO:BP | GO:0007272 | ensheathment of neurons | 7.97E-09 |
| | | GO:BP | GO:0048699 | generation of neurons | 3.67E-07 |
| | | GO:BP | GO:0010001 | glial cell differentiation | 4.54E-06 |
| | | GO:BP | GO:0032291 | axon ensheathment in central nervous system | 7.63E-05 |

| | | | | | |
|---|---|---|---|---|---|
| | | GO:BP | GO:0022010 | central nervous system myelination | 7.63E-05 |
| | Brain - Hippocampus _M3 | GO:BP | GO:0006119 | oxidative phosphorylation | 8.13E-45 |
| | | GO:CC | GO:0070469 | respirasome | 1.11E-39 |
| | | GO:CC | GO:0098803 | respiratory chain complex | 4.04E-39 |
| | | GO:BP | GO:0009060 | aerobic respiration | 1.01E-38 |
| | | GO:BP | GO:0042773 | ATP synthesis coupled electron transport | 1.43E-38 |
| | | GO:BP | GO:0022904 | respiratory electron transport chain | 4.66E-35 |
| | | GO:BP | GO:0006091 | generation of precursor metabolites and energy | 7.23E-26 |
| | | GO:CC | GO:0045271 | respiratory chain complex I | 3.06E-22 |
| | | GO:BP | GO:0008152 | metabolic process | 1.60E-20 |
| | | GO:BP | GO:0044238 | primary metabolic process | 6.15E-20 |
| | | GO:BP | GO:0046034 | ATP metabolic process | 2.77E-19 |
| | | KEGG | KEGG:05010 | Alzheimer disease | 1.83E-16 |
| | | GO:BP | GO:0019637 | organophosphate metabolic process | 3.33E-07 |
| | | KEGG | KEGG:05012 | Parkinson disease | 5.17E-27 |
| | | KEGG | KEGG:05016 | Huntington disease | 1.24E-25 |
| | | KEGG | KEGG:05022 | Pathways of neurodegeneration - multiple diseases | 4.71E-14 |
| | Brain - Hippocampus _M16 | KEGG | KEGG:00190 | Oxidative phosphorylation | 4.42E-12 |
| | | GO:MF | GO:0015453 | oxidoreduction-driven active transmembrane transporter activity | 4.30E-11 |
| | | GO:CC | GO:0098803 | respiratory chain complex | 1.80E-10 |
| | | GO:CC | GO:0005746 | mitochondrial respirasome | 2.57E-10 |
| | | GO:BP | GO:0042773 | ATP synthesis coupled electron transport | 4.67E-09 |
| | | GO:BP | GO:0006754 | ATP biosynthetic process | 1.57E-07 |
| | | GO:BP | GO:0046034 | ATP metabolic process | 3.14E-06 |
| | | KEGG | KEGG:05012 | Parkinson disease | 3.06E-10 |
| | | KEGG | KEGG:05022 | Pathways of neurodegeneration - multiple diseases | 2.27E-08 |
| | | KEGG | KEGG:05010 | Alzheimer disease | 7.01E-08 |
| Adipose | Adipose - Subcutaneous _M10 | GO:BP | GO:0006518 | peptide metabolic process | 8.14E-52 |
| | | GO:BP | GO:0043603 | amide metabolic process | 4.83E-43 |
| | | GO:BP | GO:0019538 | protein metabolic process | 1.47E-21 |
| | | GO:BP | GO:1901564 | organonitrogen compound metabolic process | 5.13E-17 |
| | | GO:BP | GO:0044238 | primary metabolic process | 2.56E-09 |
| | | GO:CC | GO:0045202 | synapse | 2.40E-07 |

| | | | | | |
|---|---|---|---|---|---|
| | | GO:BP | GO:0008152 | metabolic process | 2.25E-05 |
| | | GO:BP | GO:0043170 | macromolecule metabolic process | 2.26E-05 |
| | | GO:BP | GO:0071704 | organic substance metabolic process | 9.03E-05 |
| | Adipose - Subcutaneous _M12 | GO:BP | GO:0007399 | nervous system development | 3.53E-22 |
| | | GO:BP | GO:0022008 | neurogenesis | 9.57E-16 |
| | | GO:CC | GO:0045202 | synapse | 4.86E-15 |
| | | GO:BP | GO:0048699 | generation of neurons | 1.08E-14 |
| | | GO:BP | GO:0030182 | neuron differentiation | 3.87E-12 |
| | | GO:BP | GO:0031175 | neuron projection development | 1.18E-11 |
| | | GO:CC | GO:0030424 | axon | 1.08E-10 |
| | | GO:CC | GO:0043005 | neuron projection | 4.55E-10 |
| | | GO:BP | GO:0061564 | axon development | 1.12E-09 |
| | | GO:BP | GO:0048666 | neuron development | 2.46E-09 |
| | | GO:BP | GO:0050808 | synapse organization | 2.53E-08 |
| | | GO:BP | GO:0007411 | axon guidance | 1.06E-07 |
| | | GO:BP | GO:0097485 | neuron projection guidance | 1.06E-07 |
| | | GO:BP | GO:0007409 | axonogenesis | 1.34E-07 |
| | | GO:BP | GO:0099536 | synaptic signaling | 2.43E-07 |
| | | GO:BP | GO:0048812 | neuron projection morphogenesis | 3.31E-07 |
| | | GO:BP | GO:0008366 | axon ensheathment | 4.95E-07 |
| | | GO:BP | GO:0007272 | ensheathment of neurons | 4.95E-07 |
| | | GO:CC | GO:0043025 | neuronal cell body | 5.78E-07 |
| | | GO:CC | GO:0098984 | neuron to neuron synapse | 1.03E-06 |
| | | GO:CC | GO:0097060 | synaptic membrane | 1.86E-06 |
| | | GO:BP | GO:0099537 | trans-synaptic signaling | 2.78E-06 |
| | | GO:CC | GO:0098978 | glutamatergic synapse | 7.41E-06 |
| | | GO:CC | GO:0032279 | asymmetric synapse | 9.76E-06 |
| | | GO:CC | GO:0098793 | presynapse | 1.29E-04 |
| | Adipose - Visceral (Omentum) _ M3 | GO:BP | GO:0019219 | regulation of nucleobase-containing compound metabolic process | 9.68E-67 |
| | | GO:BP | GO:0051252 | regulation of RNA metabolic process | 1.50E-65 |
| | | GO:BP | GO:0080090 | regulation of primary metabolic process | 1.23E-60 |
| | | GO:BP | GO:0051171 | regulation of nitrogen compound metabolic process | 5.21E-59 |
| | | GO:BP | GO:0060255 | regulation of macromolecule metabolic process | 2.67E-44 |

| | | | | | |
|---|---|---|---|---|---|
| | | GO:BP | GO:0019222 | regulation of metabolic process | 5.24E-42 |
| | | GO:BP | GO:0051172 | negative regulation of nitrogen compound metabolic process | 1.20E-16 |
| | | GO:BP | GO:0051173 | positive regulation of nitrogen compound metabolic process | 1.58E-13 |
| | | GO:BP | GO:0010604 | positive regulation of macromolecule metabolic process | 1.13E-12 |
| | | GO:BP | GO:0006139 | nucleobase-containing compound metabolic process | 1.40E-12 |
| | | GO:BP | GO:0009893 | positive regulation of metabolic process | 1.66E-11 |
| | Adipose - Visceral (Omentum) _M12 | GO:BP | GO:0048699 | generation of neurons | 3.01E-04 |
| | | GO:BP | GO:0022008 | neurogenesis | 5.96E-04 |
| | | GO:BP | GO:0030182 | neuron differentiation | 0.001 |
| | | GO:BP | GO:0061564 | axon development | 0.008 |
| | | GO:BP | GO:0031175 | neuron projection development | 0.014 |
| | | GO:BP | GO:0007399 | nervous system development | 0.015 |
| Gastrointestinal tissues | Esophagus - Gastroesophageal Junction _ M2 | GO:BP | GO:0051171 | regulation of nitrogen compound metabolic process | 4.35E-55 |
| | | GO:BP | GO:0080090 | regulation of primary metabolic process | 1.31E-53 |
| | | GO:BP | GO:1901564 | organonitrogen compound metabolic process | 1.26E-38 |
| | | GO:BP | GO:0019219 | regulation of nucleobase-containing compound metabolic process | 6.87E-36 |
| | | GO:BP | GO:0019538 | protein metabolic process | 1.76E-33 |
| | | GO:BP | GO:0019222 | regulation of metabolic process | 7.17E-33 |
| | | GO:BP | GO:0051252 | regulation of RNA metabolic process | 3.16E-31 |
| | | GO:BP | GO:0031323 | regulation of cellular metabolic process | 8.03E-30 |
| | | GO:BP | GO:0060255 | regulation of macromolecule metabolic process | 3.48E-29 |
| | | GO:BP | GO:0051173 | positive regulation of nitrogen compound metabolic process | 1.24E-28 |
| | | GO:BP | GO:0009893 | positive regulation of metabolic process | 1.14E-27 |
| | | GO:BP | GO:0007399 | nervous system development | 4.07E-13 |
| | | GO:BP | GO:0031175 | neuron projection development | 1.76E-07 |
| | | GO:BP | GO:0044238 | primary metabolic process | 3.28E-07 |
| | | GO:CC | GO:0030424 | axon | 9.00E-06 |
| | | GO:CC | GO:0045202 | synapse | 0.002 |
| | Esophagus - Mucosa _M2 | GO:BP | GO:1901564 | organonitrogen compound metabolic process | 9.57E-30 |
| | | GO:BP | GO:0019538 | protein metabolic process | 1.21E-27 |
| | | GO:BP | GO:0006793 | phosphorus metabolic process | 1.51E-17 |
| | | GO:BP | GO:0006796 | phosphate-containing compound metabolic process | 5.52E-17 |
| | | GO:BP | GO:0051246 | regulation of protein metabolic process | 1.89E-10 |

| | | | | | |
|---|---|---|---|---|---|
| | | GO:BP | GO:0080090 | regulation of primary metabolic process | 2.95E-10 |
| | | GO:BP | GO:0006629 | lipid metabolic process | 3.65E-10 |
| | | GO:BP | GO:0009893 | positive regulation of metabolic process | 8.37E-10 |
| | | GO:BP | GO:0051171 | regulation of nitrogen compound metabolic process | 2.51E-09 |
| | | GO:BP | GO:0051172 | negative regulation of nitrogen compound metabolic process | 4.04E-07 |
| | | GO:BP | GO:0010604 | positive regulation of macromolecule metabolic process | 5.39E-07 |
| | | GO:BP | GO:0044255 | cellular lipid metabolic process | 3.15E-06 |
| | | GO:BP | GO:0031325 | positive regulation of cellular metabolic process | 5.84E-06 |
| | | GO:BP | GO:0051247 | positive regulation of protein metabolic process | 3.12E-05 |
| | | GO:BP | GO:0006672 | ceramide metabolic process | 0.015 |
| | | GO:CC | GO:0098978 | glutamatergic synapse | 0.019 |
| | | GO:BP | GO:0007399 | nervous system development | 0.022 |
| | | GO:CC | GO:0098793 | presynapse | 0.028 |
| | | GO:BP | GO:0060255 | regulation of macromolecule metabolic process | 0.038 |
| | | GO:BP | GO:0022008 | neurogenesis | 0.039 |
| | | GO:BP | GO:0031323 | regulation of cellular metabolic process | 0.040 |
| | | GO:BP | GO:0006665 | sphingolipid metabolic process | 0.049 |
| | Esophagus - Muscularis _M9 | GO:BP | GO:0007399 | nervous system development | 7.31E-07 |
| | | GO:BP | GO:0022008 | neurogenesis | 7.22E-05 |
| | | GO:BP | GO:0048699 | generation of neurons | 0.003 |
| | Esophagus - Muscularis _M14 | GO:BP | GO:0007399 | nervous system development | 0.002 |
| | | GO:BP | GO:0022008 | neurogenesis | 0.003 |

**Figure S1.** Genetic correlation network between anorexia nervosa and risk factors in males.

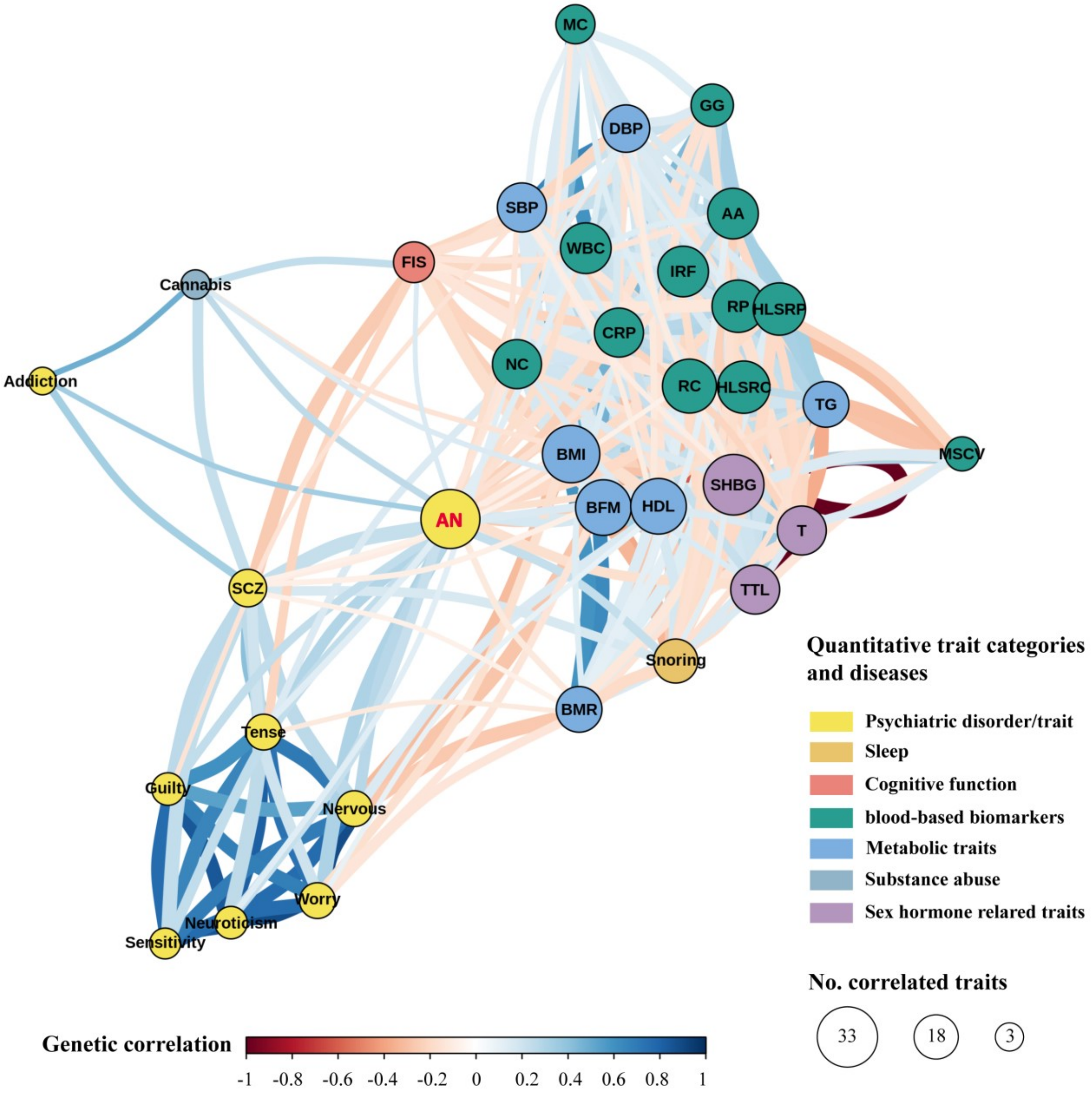

Genetically correlated traits and diseases are clustered close together. Each circle represents a trait, and each edge represents a significant genetic correlation ($P < 0.001$). Positive and negative genetic correlations are indicated by color according to the key. AN, anorexia nervosa; SCZ, schizophrenia; Guilty, guilty feelings; Addiction, ever addicted to any substance or behaviour; Sensitivity, sensitivity / hurt feelings; Nervous, nervous feelings; Worry, worrier / anxious feelings; Tense, Tense / 'highly strung'; Neuroticism, neuroticism score; FIS, Fluid intelligence score; Cannabis, ever taken cannabis; Snoring, snoring; SHBG, Sex hormone-binding globulin levels; BMI, body mass index; BFM, whole body fat mass; BMR, basal metabolic rate; SBP, systolic blood pressure; DBP, diastolic blood pressure; HDL, HDL cholesterol; TG, Triglycerides; WBC, white blood cell (leukocyte) count; MC, monocyte count; NC, Neutrophill count; RP, reticulocyte percentage; RC, reticulocyte count; IRF, immature reticulocyte fraction; HLSRP, high light scatter reticulocyte percentage; HLSRC, high light scatter reticulocyte count; AA, Alanine aminotransferase (quantile); CRP, C-reactive protein (quantile); GG, gamma glutamyltransferase (quantile).

**Figure S2.** Genetic correlation network between anorexia nervosa and risk factors in females.

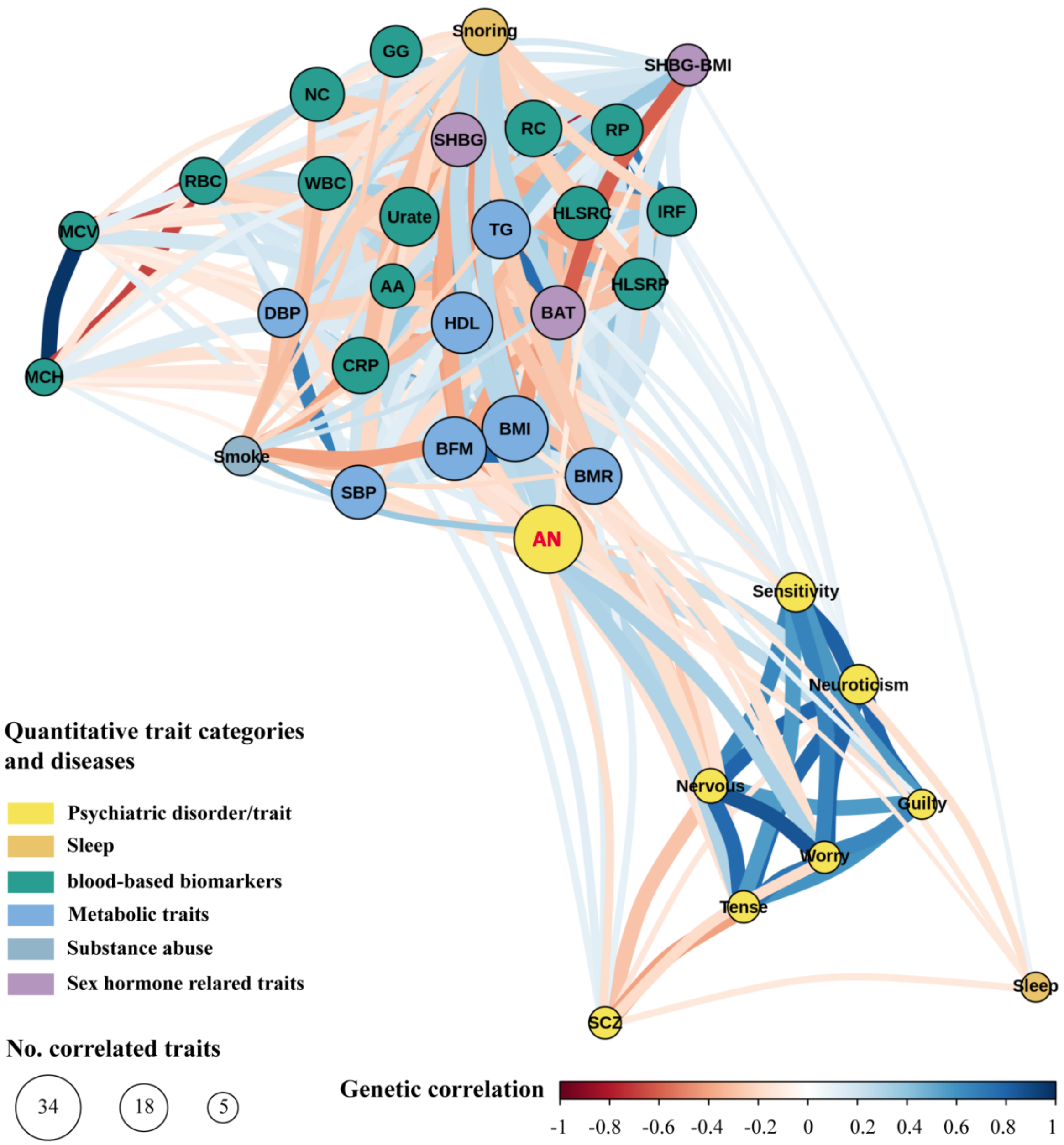

Genetically correlated traits and diseases are clustered close together. Each circle represents a trait, and each edge represents a significant genetic correlation ($P < 0.001$). Positive and negative genetic correlations are indicated by color according to the key. AN, anorexia nervosa; SCZ, schizophrenia; Guilty, guilty feelings; Sensitivity, sensitivity / hurt feelings; Nervous, nervous feelings; Worry, worrier / anxious feelings; Tense, Tense / 'highly strung'; Neuroticism, neuroticism score; Smoke, age started smoking in former smokers; Snoring, snoring; BAT, Bioavailable testosterone levels; SHBG, Sex hormone-binding globulin levels; SHBG-BMI, Sex hormone-binding globulin levels adjusted for BMI; BMI, body mass index; BFM, whole body fat mass; BMR, basal metabolic rate; SBP, systolic blood pressure; DBP, diastolic blood pressure; HDL, HDL cholesterol; TG, Triglycerides; WBC, white blood cell (leukocyte) count; RBC, red blood cell (erythrocyte) count; MCV, mean corpuscular volume; MCH, mean corpuscular haemoglobin; NC, Neutrophill count; RP, reticulocyte percentage; RC, reticulocyte count; IRF, immature reticulocyte fraction; HLSRP, high light scatter reticulocyte percentage; HLSRC, high light scatter reticulocyte count; AA, Alanine aminotransferase (quantile); CRP, C-reactive protein (quantile); GG, gamma glutamyltransferase (quantile); Urate, urate (quantile).

**Figure S3.** MR plots for the relationship between anorexia nervosa and risk factors. a, exposure: anorexia nervosa; outcome: risk factors; b, exposure: risk factors; outcome: anorexia nervosa. The effect estimates represent the odds ratio for outcome per one-point increment in exposure. Data are presented as mean values ± s.e.m. The width of the lines extending from the center point represent the 95% CI. Two-sided unadjusted association P values from five models are given.

**a**

| Exposure | Outcome Trait | MR method | nsnp | OR | pval |
|---|---|---|---|---|---|
| Anorexia Nervosa | Body mass index (BMI) | MR Egger | 7 | 0.997(0.829−1.198) | 0.974 |
| | | Weighted median | 7 | 0.935(0.906−0.966) | 4.52e−05 |
| | | Inverse variance weighted | 7 | 0.933(0.874−0.996) | 0.036 |
| | | Simple mode | 7 | 0.919(0.876−0.964) | 0.014 |
| | | Weighted mode | 7 | 0.922(0.877−0.970) | 0.02 |
| | Mean corpuscular volume | MR Egger | 7 | 0.916(0.864−0.970) | 0.031 |
| | | Weighted median | 7 | 0.951(0.923−0.979) | 0.001 |
| | | Inverse variance weighted | 7 | 0.956(0.934−0.979) | 0.000206 |
| | | Simple mode | 7 | 0.950(0.903−0.999) | 0.091 |
| | | Weighted mode | 7 | 0.949(0.903−0.998) | 0.086 |
| | Reticulocyte count | MR Egger | 7 | 0.789(0.699−0.890) | 0.012 |
| | | Weighted median | 7 | 0.936(0.899−0.974) | 0.001 |
| | | Inverse variance weighted | 7 | 0.896(0.846−0.949) | 0.000166 |
| | | Simple mode | 7 | 0.942(0.881−1.008) | 0.133 |
| | | Weighted mode | 7 | 0.949(0.905−0.994) | 0.071 |
| | High light scatter reticulocyte count | MR Egger | 7 | 0.784(0.686−0.896) | 0.016 |
| | | Weighted median | 7 | 0.931(0.893−0.971) | 0.000773 |
| | | Inverse variance weighted | 7 | 0.888(0.836−0.943) | 9.43e−05 |
| | | Simple mode | 7 | 0.939(0.856−1.030) | 0.231 |
| | | Weighted mode | 7 | 0.947(0.905−0.991) | 0.058 |
| | Red blood cell (erythrocyte) distribution width | MR Egger | 7 | 0.916(0.866−0.970) | 0.03 |
| | | Weighted median | 7 | 0.944(0.916−0.973) | 0.000159 |
| | | Inverse variance weighted | 7 | 0.951(0.931−0.972) | 4.74e−06 |
| | | Simple mode | 7 | 0.933(0.888−0.979) | 0.03 |
| | | Weighted mode | 7 | 0.932(0.887−0.979) | 0.032 |

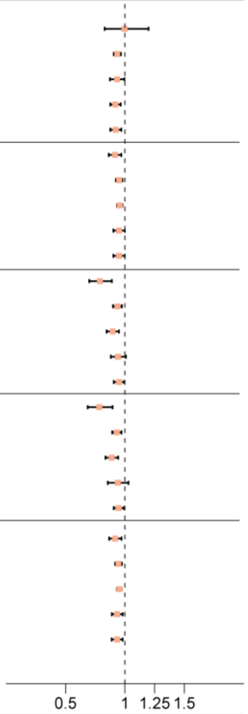

**b**

| Exposure | Outcome Trait | MR method | Nsnp | OR | Pval |
|---|---|---|---|---|---|
| Neuroticism score | Anorexia Nervosa | MR Egger | 92 | 0.814(0.245−2.705) | 0.738 |
| | | Weighted median | | 1.451(1.086−1.938) | 0.012 |
| | | Inverse variance weighted | | 1.603(1.251−2.054) | 0.000191 |
| | | Simple mode | | 1.468(0.721−2.987) | 0.292 |
| | | Weighted mode | | 1.468(0.771−2.796) | 0.246 |
| Basal metabolic rate | | MR Egger | 1236 | 1.086(0.859−1.372) | 0.49 |
| | | Weighted median | | 0.885(0.790−0.992) | 0.035 |
| | | Inverse variance weighted | | 0.802(0.738−0.873) | 3.16e−07 |
| | | Simple mode | | 1.040(0.635−1.705) | 0.875 |
| | | Weighted mode | | 0.913(0.613−1.359) | 0.653 |
| White blood cell (leukocyte) count | | MR Egger | 880 | 1.237(1.096−1.396) | 0.000596 |
| | | Weighted median | | 1.173(1.085−1.269) | 6.37e−05 |
| | | Inverse variance weighted | | 1.114(1.056−1.176) | 8.54e−05 |
| | | Simple mode | | 1.210(0.897−1.634) | 0.212 |
| | | Weighted mode | | 1.190(0.991−1.430) | 0.063 |
| Lymphocyte count | | MR Egger | 925 | 1.282(1.139−1.441) | 3.87e−05 |
| | | Weighted median | | 1.163(1.077−1.255) | 0.00011 |
| | | Inverse variance weighted | | 1.148(1.090−1.209) | 2.16e−07 |
| | | Simple mode | | 0.874(0.638−1.198) | 0.404 |
| | | Weighted mode | | 1.128(0.926−1.373) | 0.231 |
| Eosinophill count | | MR Egger | 911 | 1.202(1.016−1.422) | 0.032 |
| | | Weighted median | | 1.140(1.025−1.268) | 0.016 |
| | | Inverse variance weighted | | 1.164(1.082−1.252) | 4.28e−05 |
| | | Simple mode | | 1.121(0.780−1.612) | 0.536 |
| | | Weighted mode | | 1.096(0.880−1.363) | 0.413 |
| Mean reticulocyte volume | | MR Egger | 1047 | 0.929(0.855−1.009) | 0.082 |
| | | Weighted median | | 0.882(0.825−0.942) | 0.000198 |
| | | Inverse variance weighted | | 0.927(0.888−0.966) | 0.000386 |
| | | Simple mode | | 0.861(0.712−1.042) | 0.125 |
| | | Weighted mode | | 0.877(0.795−0.968) | 0.009 |
| C−reactive protein (quantile) | | MR Egger | 621 | 0.893(0.819−0.973) | 0.01 |
| | | Weighted median | | 0.868(0.798−0.943) | 0.001 |
| | | Inverse variance weighted | | 0.891(0.846−0.939) | 1.64e−05 |
| | | Simple mode | | 0.918(0.747−1.129) | 0.418 |
| | | Weighted mode | | 0.865(0.796−0.941) | 0.001 |

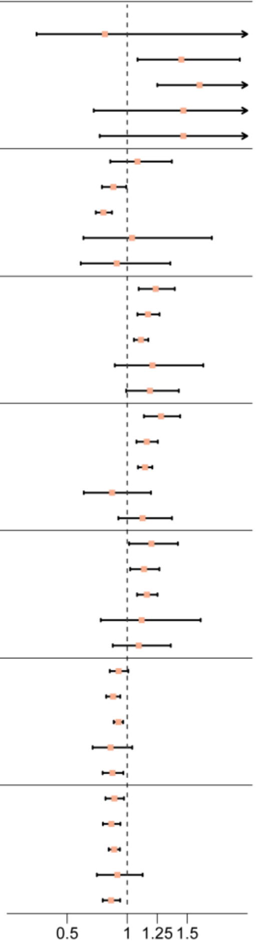

**Figure S4**. Local genetic correlation between anorexia nervosa and schizophrenia. AN, anorexia nervosa; SCZ, schizophrenia.

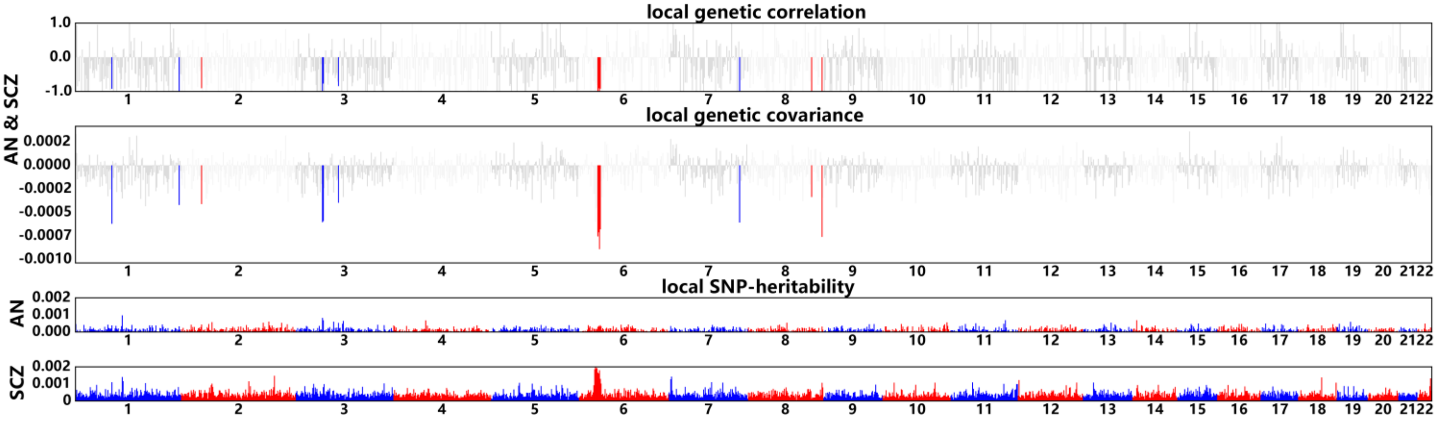

**Figure S5**. Local genetic correlation between anorexia nervosa and metabolic traits. (a) Manhattan plot showing the estimates of local genetic correlation, genetic covariance, and SNP heritability between anorexia nervosa and BMI. (b) Manhattan plot showing the estimates of local genetic correlation, genetic covariance, and SNP heritability between anorexia nervosa and whole body fat mass. (c) Manhattan plot showing the estimates of local genetic correlation, genetic covariance, and SNP heritability between anorexia nervosa and basal metabolic rate. (d) Manhattan plot showing the estimates of local genetic correlation, genetic covariance, and SNP heritability between anorexia nervosa and HDL cholesterol.

**a**

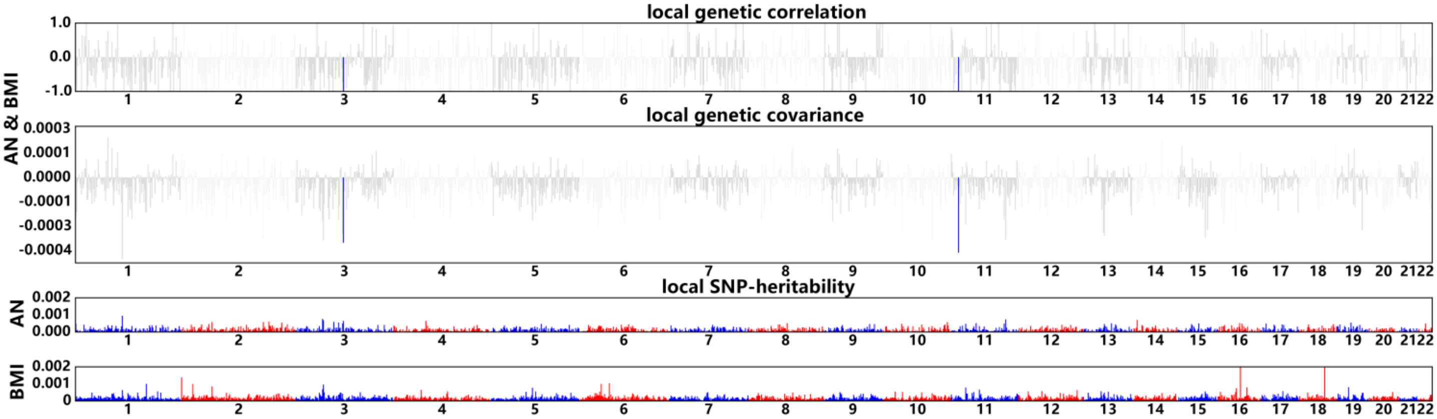

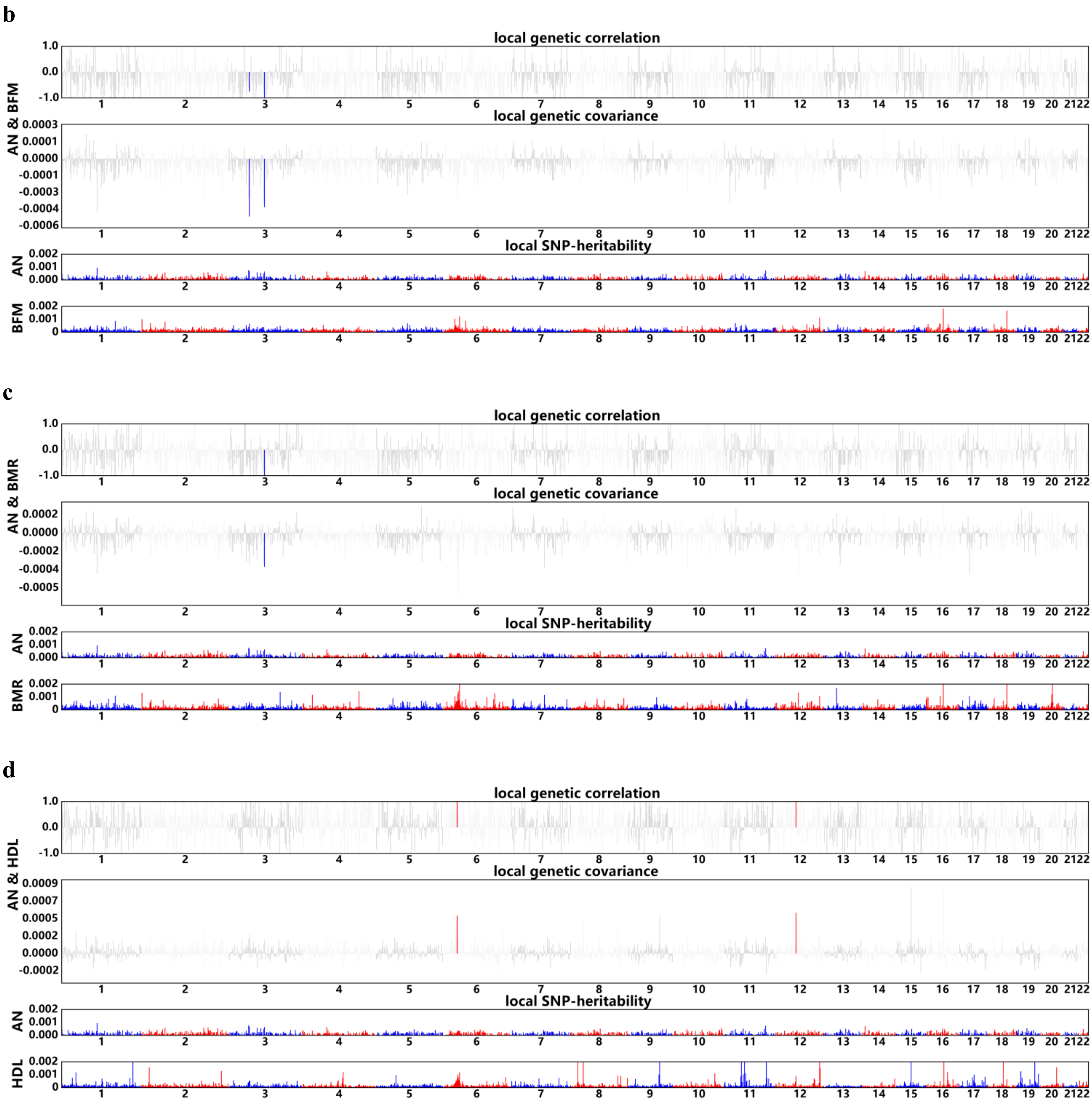

**Figure S6**. Local genetic correlation between anorexia nervosa and sex hormone related traits. (a) Manhattan plot showing the estimates of local genetic correlation, genetic covariance, and SNP heritability between anorexia nervosa and sex hormone-binding globulin levels. (b) Manhattan plot showing the estimates of local genetic correlation, genetic covariance, and SNP heritability between anorexia nervosa and sex hormone-binding globulin levels in female.

**a**

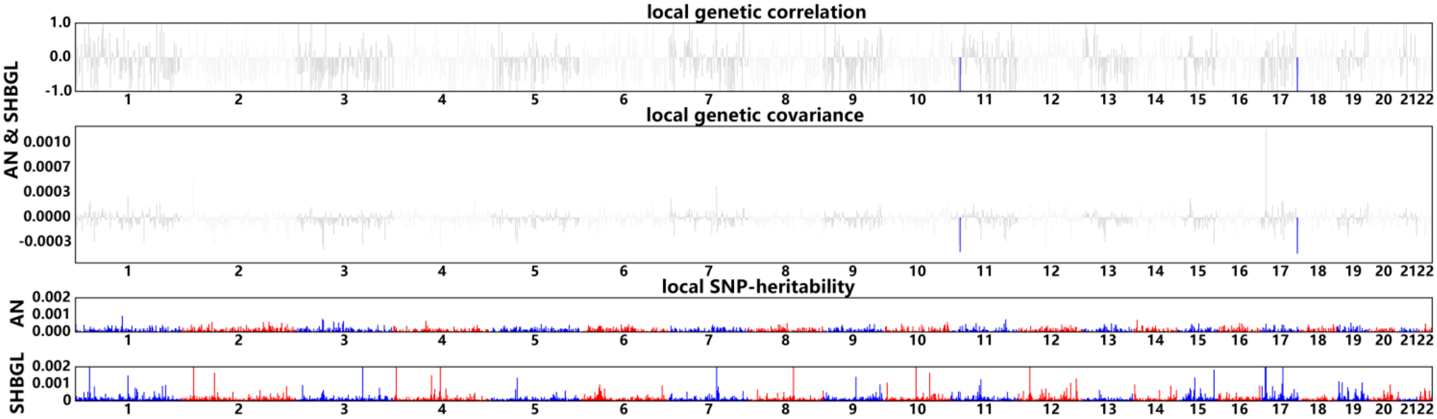

**b**

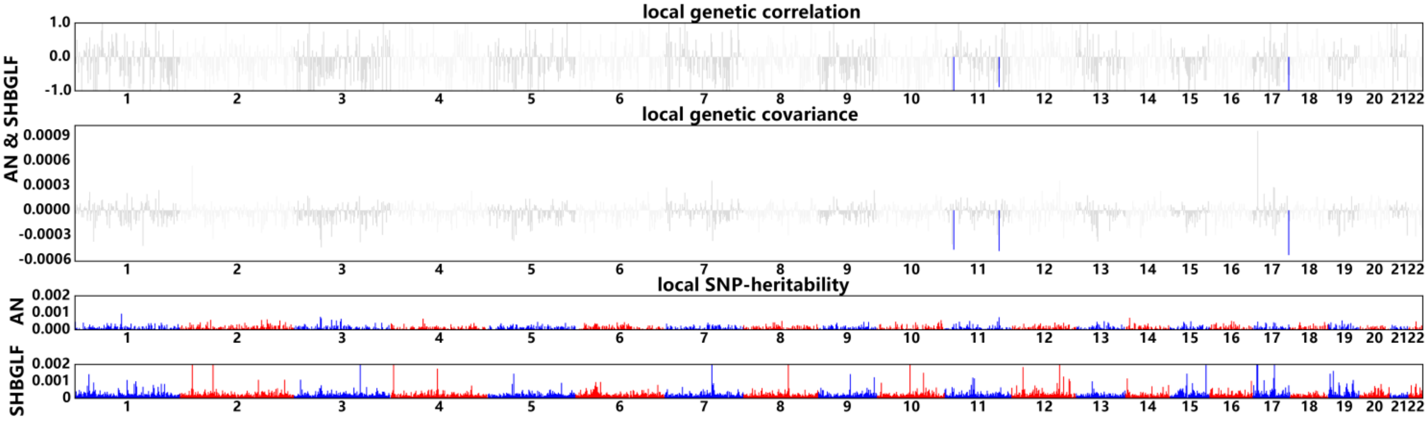

**Figure S7**. Local genetic correlation between anorexia nervosa and blood-based biomarkers. (a) Manhattan plot showing the estimates of local genetic correlation, genetic covariance, and SNP heritability between anorexia nervosa and white blood cell (leukocyte) count (WBC). (b) Manhattan plot showing the estimates of local genetic correlation, genetic covariance, and SNP heritability between anorexia nervosa and reticulocyte percentage (RP). (c) Manhattan plot showing the estimates of local genetic correlation, genetic covariance, and SNP heritability between anorexia nervosa and Reticulocyte count (RC). (d) Manhattan plot showing the estimates of local genetic correlation, genetic covariance, and SNP heritability between anorexia nervosa and high light scatter reticulocyte percentage (HLSRP). (e) Manhattan plot showing the estimates of local genetic correlation, genetic covariance, and SNP heritability between anorexia nervosa and high light scatter reticulocyte count (HLSRC). (f) Manhattan plot showing the estimates of local genetic correlation, genetic covariance, and SNP heritability between anorexia nervosa and urate.

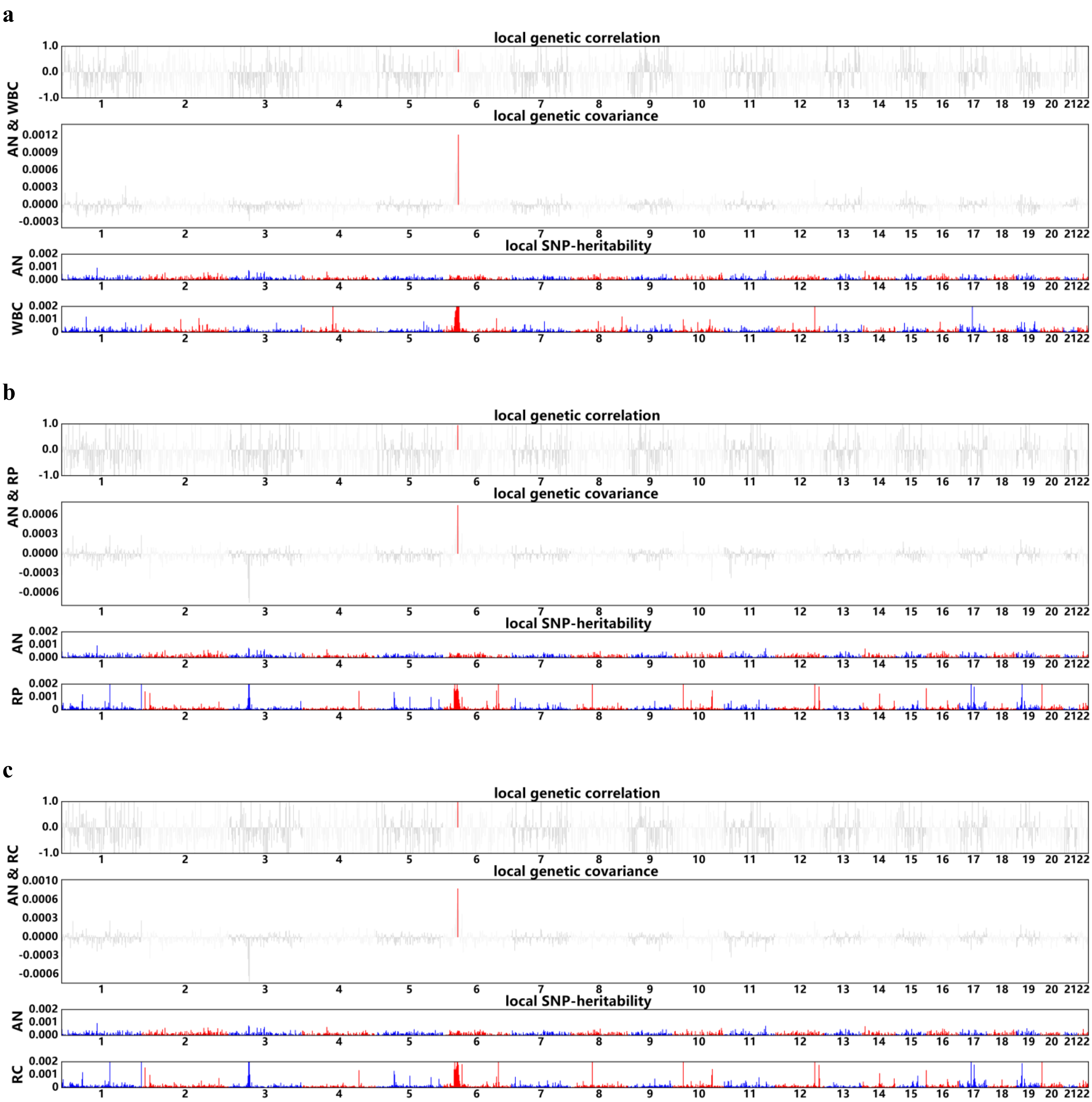
a
AN & WBC
local genetic correlation
local genetic covariance
local SNP-heritability
AN
WBC
b
AN & RP
local genetic correlation
local genetic covariance
local SNP-heritability
AN
RP
c
AN & RC
local genetic correlation
local genetic covariance
local SNP-heritability
AN
RC

d

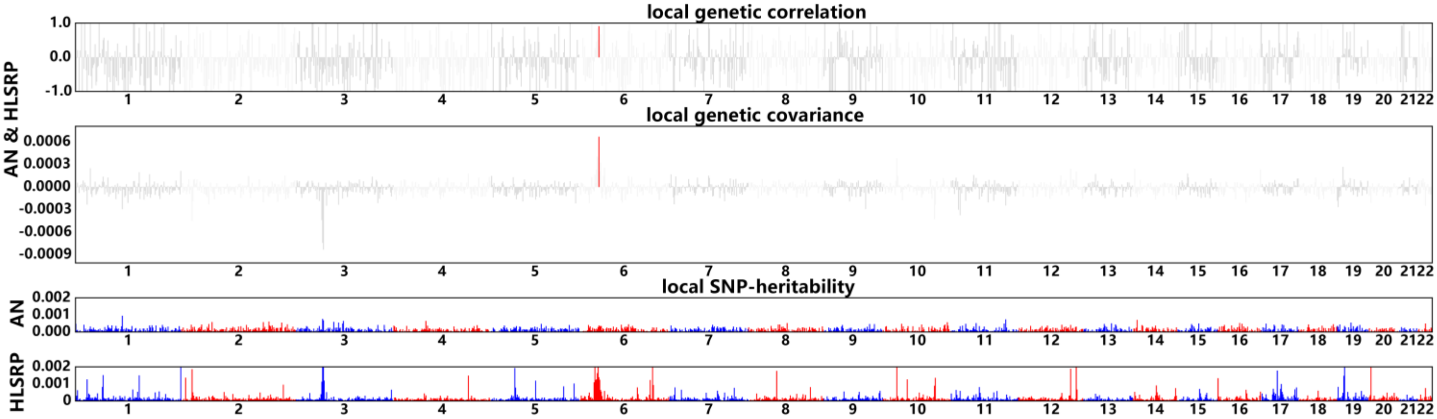
local genetic correlation
local genetic covariance
local SNP-heritability
AN & HLSRP
AN
HLSRP
1.0
0.0
-1.0
0.0006
0.0003
0.0000
-0.0003
-0.0006
-0.0009
0.002
0.001
0.000
0
1 2 3 4 5 6 7 8 9 10 11 12 13 14 15 16 17 18 19 20 2122

e

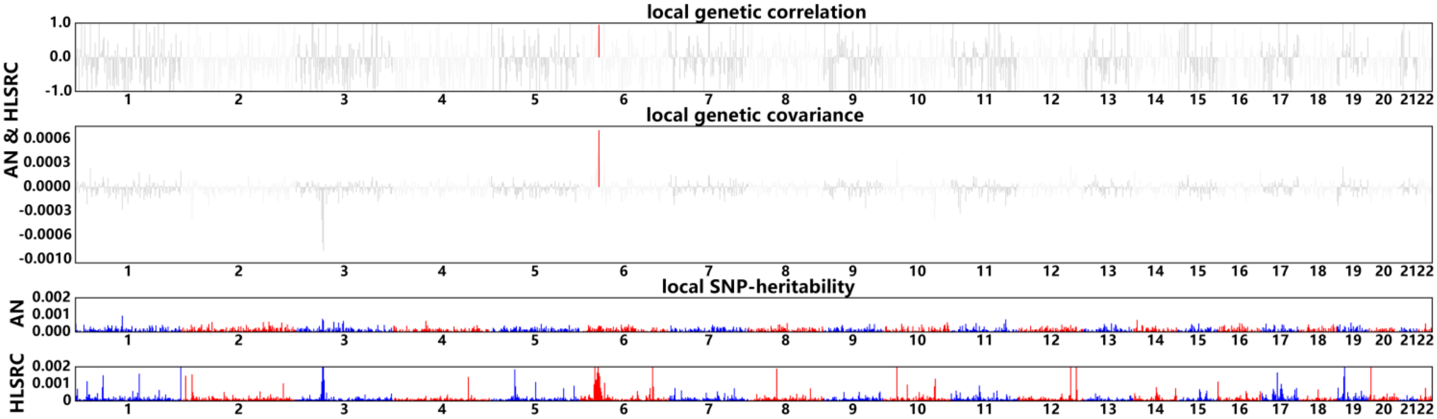
local genetic correlation
local genetic covariance
local SNP-heritability
AN & HLSRC
AN
HLSRC
1.0
0.0
-1.0
0.0006
0.0003
0.0000
-0.0003
-0.0006
-0.0010
0.002
0.001
0.000
0
1 2 3 4 5 6 7 8 9 10 11 12 13 14 15 16 17 18 19 20 2122

f

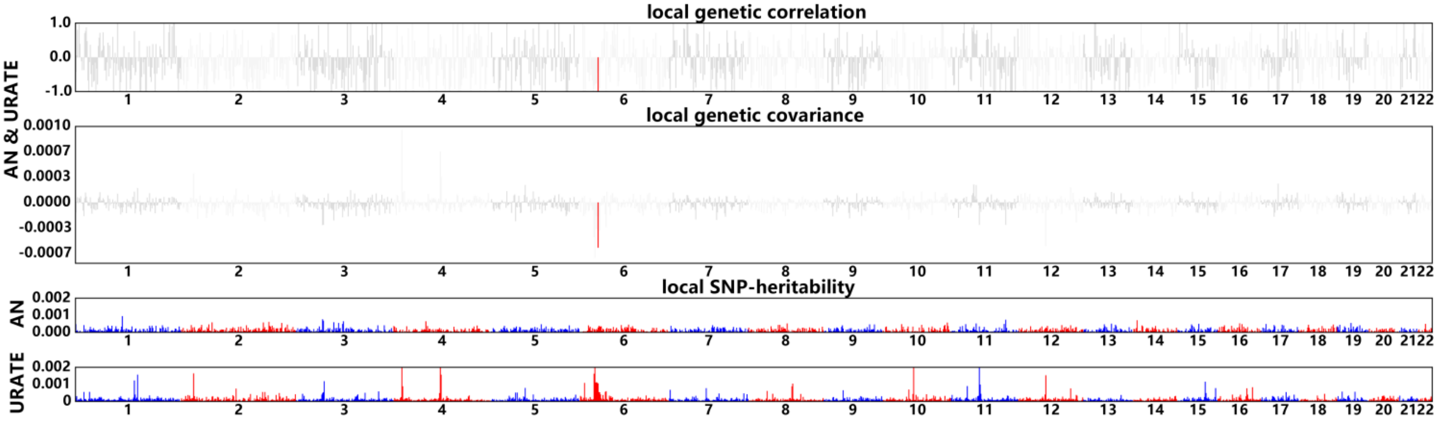
local genetic correlation
local genetic covariance
local SNP-heritability
AN & URATE
AN
URATE
1.0
0.0
-1.0
0.0010
0.0007
0.0003
0.0000
-0.0003
-0.0007
0.002
0.001
0.000
0
1 2 3 4 5 6 7 8 9 10 11 12 13 14 15 16 17 18 19 20 2122

**Figure S8.** Heat maps showing patterns of cell-type-specific enrichments of SNP-heritability for genetically associated traits across 88 cell types- or tissues-specific annotations. Each checkered rectangle reflects the z-score, scaled by traits. Red indicates enrichment, blue indicates depletion. Deeper color represents stronger magnitude of effects. Asterisks represent statistical significance. The category of cell-types is color coded to the left. (a) DNase (DNase I hypersensitive sites), (b) H3K27ac, (c) H3K36me3, (d) H3K4me1, (e) H3K4me3, and (f) H3K9ac. AN, anorexia nervosa; OCD, obsessive-compulsive disorder; MDD, major depressive disorder; SCZ, schizophrenia; BD, bipolar disorder; Guilty, guilty feelings; Addiction, ever addicted to any substance or behaviour; Sensitivity, sensitivity / hurt feelings; Nervous, nervous feelings; Worry, worrier / anxious feelings; Tense, Tense / 'highly strung'; Neuroticism, neuroticism score; EA, educational attainment (age completed full time education); FIS, Fluid intelligence score; Cannabis, ever taken cannabis; Smoke, age started smoking in former smokers; Snoring, snoring; BAT, Bioavailable testosterone levels; SHBG, Sex hormone-binding globulin levels; SHBG-BMI, Sex hormone-binding globulin levels adjusted for BMI; BMI, body mass index; BFM, whole body fat mass; T2D, type 2 diabetes; BMR, basal metabolic rate; SBP, systolic blood pressure; DBP, diastolic blood pressure; Insulin, fasting blood insulin; HDL, HDL cholesterol; TG, Triglycerides; Glucose, glucose; WBC, white blood cell (leukocyte) count; RBC, red blood cell (erythrocyte) count; MCV, mean corpuscular volume; LC, lymphocyte count; MC, monocyte count; NC, Neutrophill count; RP, reticulocyte percentage; RC, reticulocyte count; IRF, immature reticulocyte fraction; HLSRP, high light scatter reticulocyte percentage; HLSRC, high light scatter reticulocyte count; AA, Alanine aminotransferase (quantile); CRP, C-reactive protein (quantile); GG, gamma glutamyltransferase (quantile); Urate, urate (quantile).

### a DNase

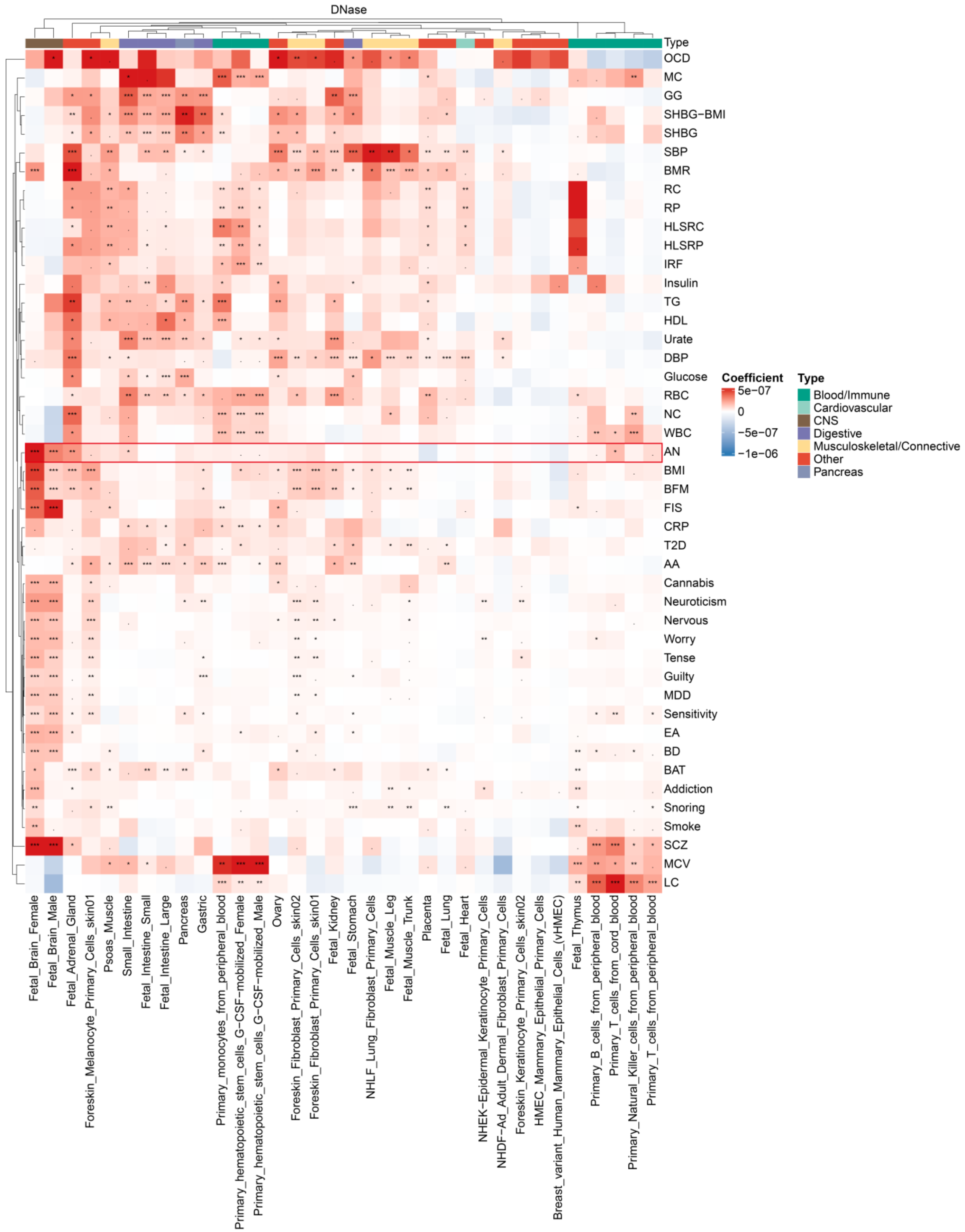

DNase
Type
OCD
MC
GG
SHBG−BMI
SHBG
SBP
BMR
RC
RP
HLSRC
HLSRP
IRF
Insulin
TG
HDL
Urate
DBP
Glucose
RBC
NC
WBC
AN
BMI
BFM
FIS
CRP
T2D
AA
Cannabis
Neuroticism
Nervous
Worry
Tense
Guilty
MDD
Sensitivity
EA
BD
BAT
Addiction
Snoring
Smoke
SCZ
MCV
LC
Coefficient
5e−07
0
−5e−07
−1e−06
Type
Blood/Immune
Cardiovascular
CNS
Digestive
Musculoskeletal/Connective
Other
Pancreas
Fetal_Brain_Female
Fetal_Brain_Male
Fetal_Adrenal_Gland
Foreskin_Melanocyte_Primary_Cells_skin01
Psoas_Muscle
Small_Intestine
Fetal_Intestine_Small
Fetal_Intestine_Large
Pancreas
Gastric
Primary_monocytes_from_peripheral_blood
Primary_hematopoietic_stem_cells_G−CSF−mobilized_Female
Primary_hematopoietic_stem_cells_G−CSF−mobilized_Male
Ovary
Foreskin_Fibroblast_Primary_Cells_skin02
Foreskin_Fibroblast_Primary_Cells_skin01
Fetal_Kidney
Fetal_Stomach
NHLF_Lung_Fibroblast_Primary_Cells
Fetal_Muscle_Leg
Fetal_Muscle_Trunk
Placenta
Fetal_Lung
Fetal_Heart
NHEK−Epidermal_Keratinocyte_Primary_Cells
NHDF−Ad_Adult_Dermal_Fibroblast_Primary_Cells
Foreskin_Keratinocyte_Primary_Cells_skin02
HMEC_Mammary_Epithelial_Primary_Cells
Breast_variant_Human_Mammary_Epithelial_Cells_(vHMEC)
Fetal_Thymus
Primary_B_cells_from_peripheral_blood
Primary_T_cells_from_cord_blood
Primary_Natural_Killer_cells_from_peripheral_blood
Primary_T_cells_from_peripheral_blood

## b H3K27ac

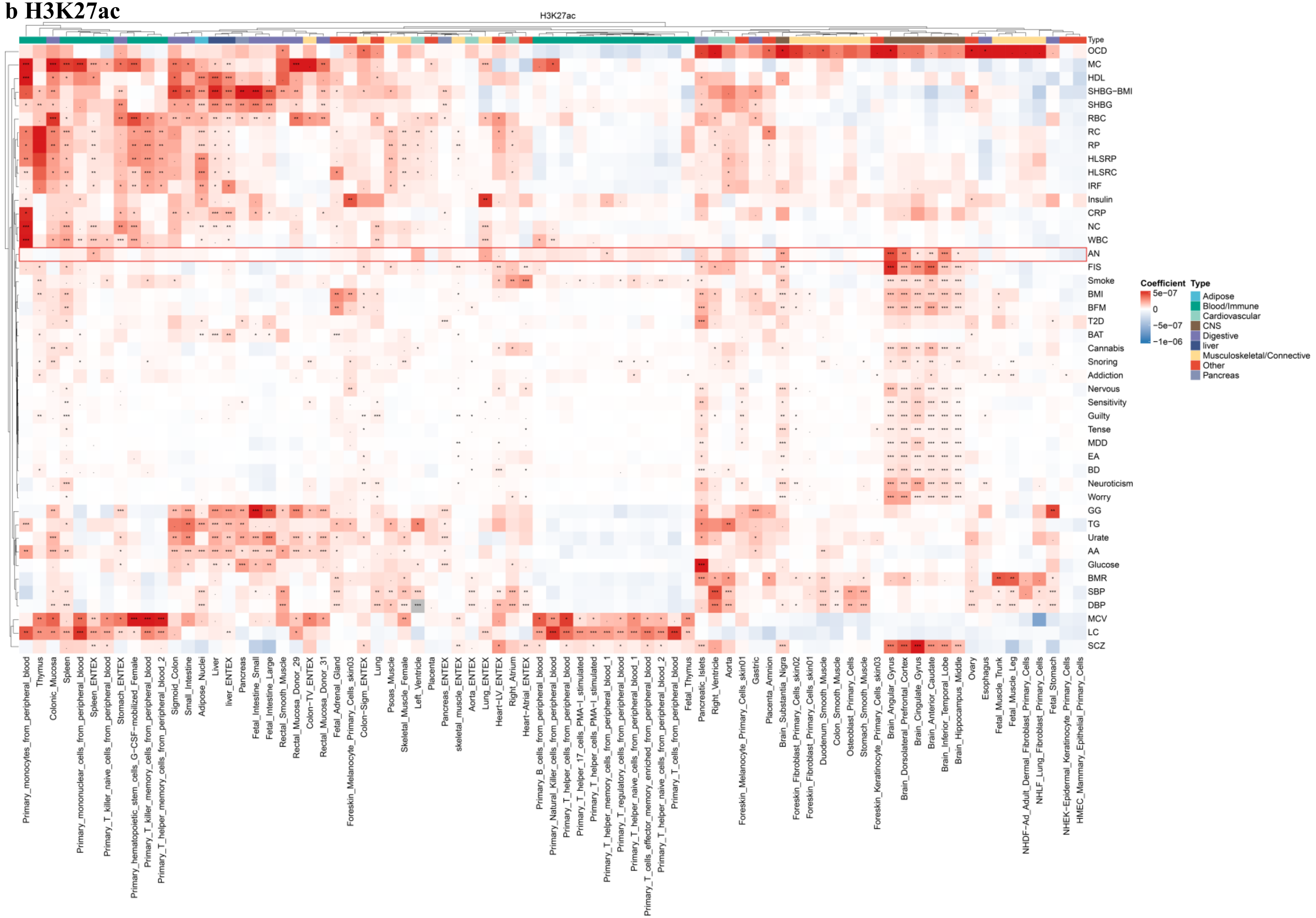

H3K27ac
Type
OCD
MC
HDL
SHBG−BMI
SHBG
RBC
RC
RP
HLSRP
HLSRC
IRF
Insulin
CRP
NC
WBC
AN
FIS
Smoke
BMI
BFM
T2D
BAT
Cannabis
Snoring
Addiction
Nervous
Sensitivity
Guilty
Tense
MDD
EA
BD
Neuroticism
Worry
GG
TG
Urate
AA
Glucose
BMR
SBP
DBP
MCV
LC
SCZ
Coefficient
5e−07
0
−5e−07
−1e−06
Type
Adipose
Blood/Immune
Cardiovascular
CNS
Digestive
liver
Musculoskeletal/Connective
Other
Pancreas
Primary_monocytes_from_peripheral_blood
Thymus
Colonic_Mucosa
Spleen
Primary_mononuclear_cells_from_peripheral_blood
Spleen_ENTEX
Primary_T_killer_naive_cells_from_peripheral_blood
Stomach_ENTEX
Primary_hematopoietic_stem_cells_G−CSF−mobilized_Female
Primary_T_killer_memory_cells_from_peripheral_blood
Primary_T_helper_memory_cells_from_peripheral_blood_2
Sigmoid_Colon
Small_Intestine
Adipose_Nuclei
Liver
liver_ENTEX
Pancreas
Fetal_Intestine_Small
Fetal_Intestine_Large
Rectal_Smooth_Muscle
Rectal_Mucosa_Donor_29
Colon−TV_ENTEX
Rectal_Mucosa_Donor_31
Fetal_Adrenal_Gland
Foreskin_Melanocyte_Primary_Cells_skin03
Colon−Sigm_ENTEX
Lung
Psoas_Muscle
Skeletal_Muscle_Female
Left_Ventricle
Placenta
Pancreas_ENTEX
skeletal_muscle_ENTEX
Aorta_ENTEX
Lung_ENTEX
Heart−LV_ENTEX
Right_Atrium
Heart−Atrial_ENTEX
Primary_B_cells_from_peripheral_blood
Primary_Natural_Killer_cells_from_peripheral_blood
Primary_T_helper_cells_from_peripheral_blood
Primary_T_helper_17_cells_PMA−I_stimulated
Primary_T_helper_cells_PMA−I_stimulated
Primary_T_helper_memory_cells_from_peripheral_blood_1
Primary_T_regulatory_cells_from_peripheral_blood
Primary_T_helper_naive_cells_from_peripheral_blood_1
Primary_T_cells_effector_memory_enriched_from_peripheral_blood
Primary_T_helper_naive_cells_from_peripheral_blood_2
Primary_T_cells_from_peripheral_blood
Fetal_Thymus
Pancreatic_Islets
Right_Ventricle
Aorta
Foreskin_Melanocyte_Primary_Cells_skin01
Gastric
Placenta_Amnion
Brain_Substantia_Nigra
Foreskin_Fibroblast_Primary_Cells_skin02
Foreskin_Fibroblast_Primary_Cells_skin01
Duodenum_Smooth_Muscle
Colon_Smooth_Muscle
Osteoblast_Primary_Cells
Stomach_Smooth_Muscle
Foreskin_Keratinocyte_Primary_Cells_skin03
Brain_Angular_Gyrus
Brain_Dorsolateral_Prefrontal_Cortex
Brain_Cingulate_Gyrus
Brain_Anterior_Caudate
Brain_Inferior_Temporal_Lobe
Brain_Hippocampus_Middle
Ovary
Esophagus
Fetal_Muscle_Trunk
Fetal_Muscle_Leg
NHDF−Ad_Adult_Dermal_Fibroblast_Primary_Cells
NHLF_Lung_Fibroblast_Primary_Cells
Fetal_Stomach
NHEK−Epidermal_Keratinocyte_Primary_Cells
HMEC_Mammary_Epithelial_Primary_Cells

c H3K36me3

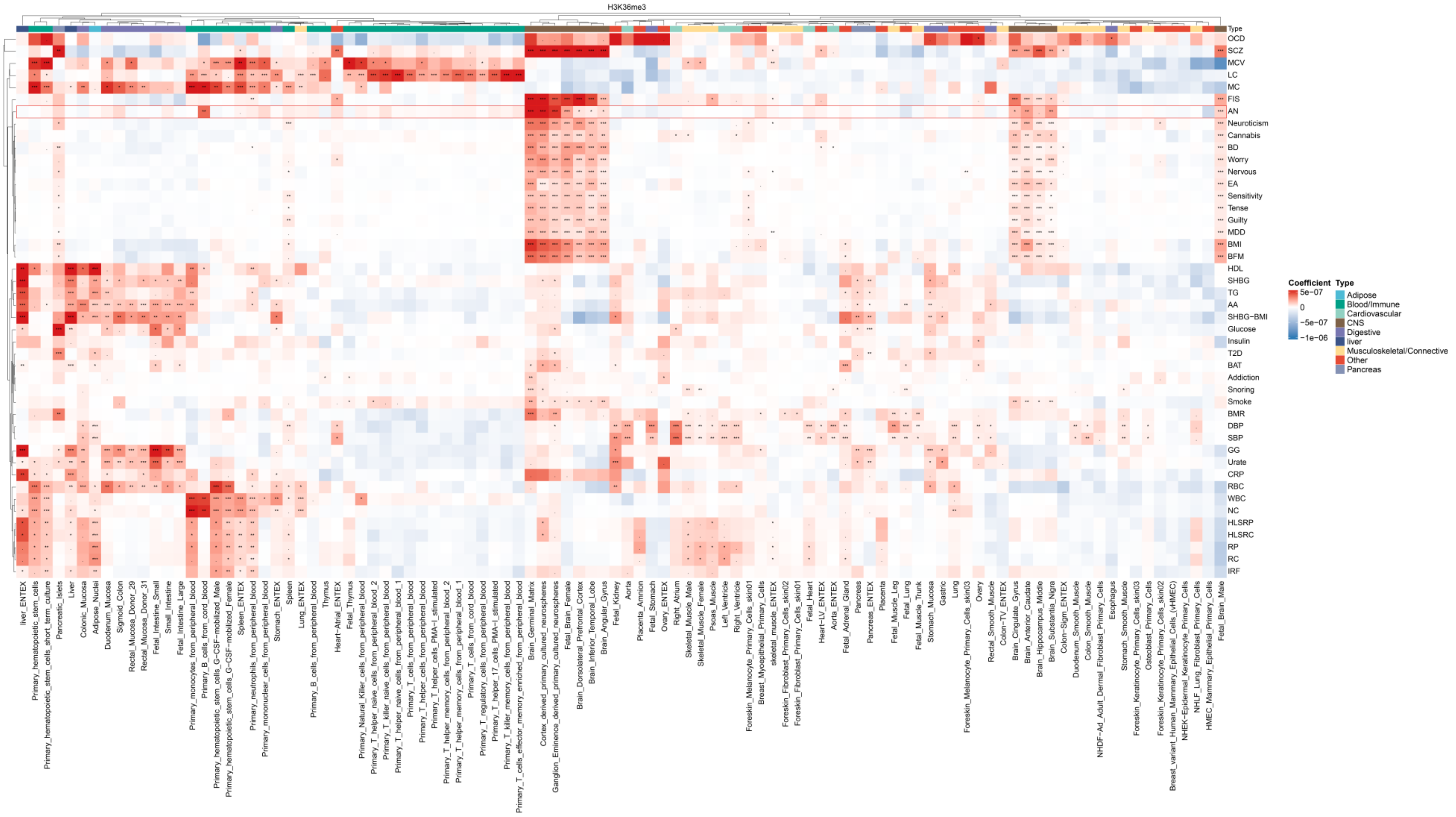

**d H3K4me1**

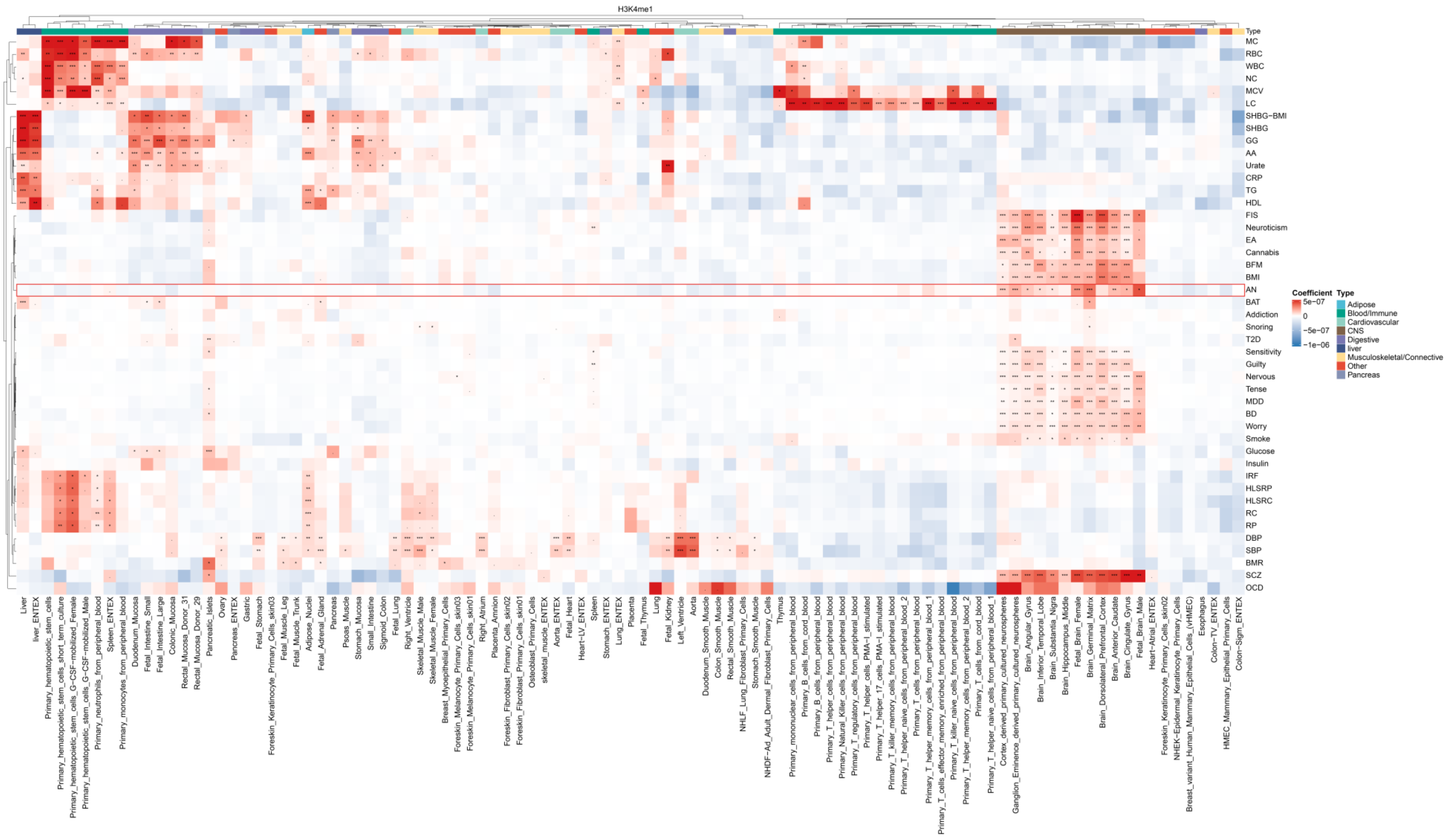
H3K4me1
Type
MC
RBC
WBC
NC
MCV
LC
SHBG−BMI
SHBG
GG
AA
Urate
CRP
TG
HDL
FIS
Neuroticism
EA
Cannabis
BFM
BMI
AN
BAT
Addiction
Snoring
T2D
Sensitivity
Guilty
Nervous
Tense
MDD
BD
Worry
Smoke
Glucose
Insulin
IRF
HLSRP
HLSRC
RC
RP
DBP
SBP
BMR
SCZ
OCD
Coefficient
5e−07
0
−5e−07
−1e−06
Type
Adipose
Blood/Immune
Cardiovascular
CNS
Digestive
liver
Musculoskeletal/Connective
Other
Pancreas

e H3K4me3

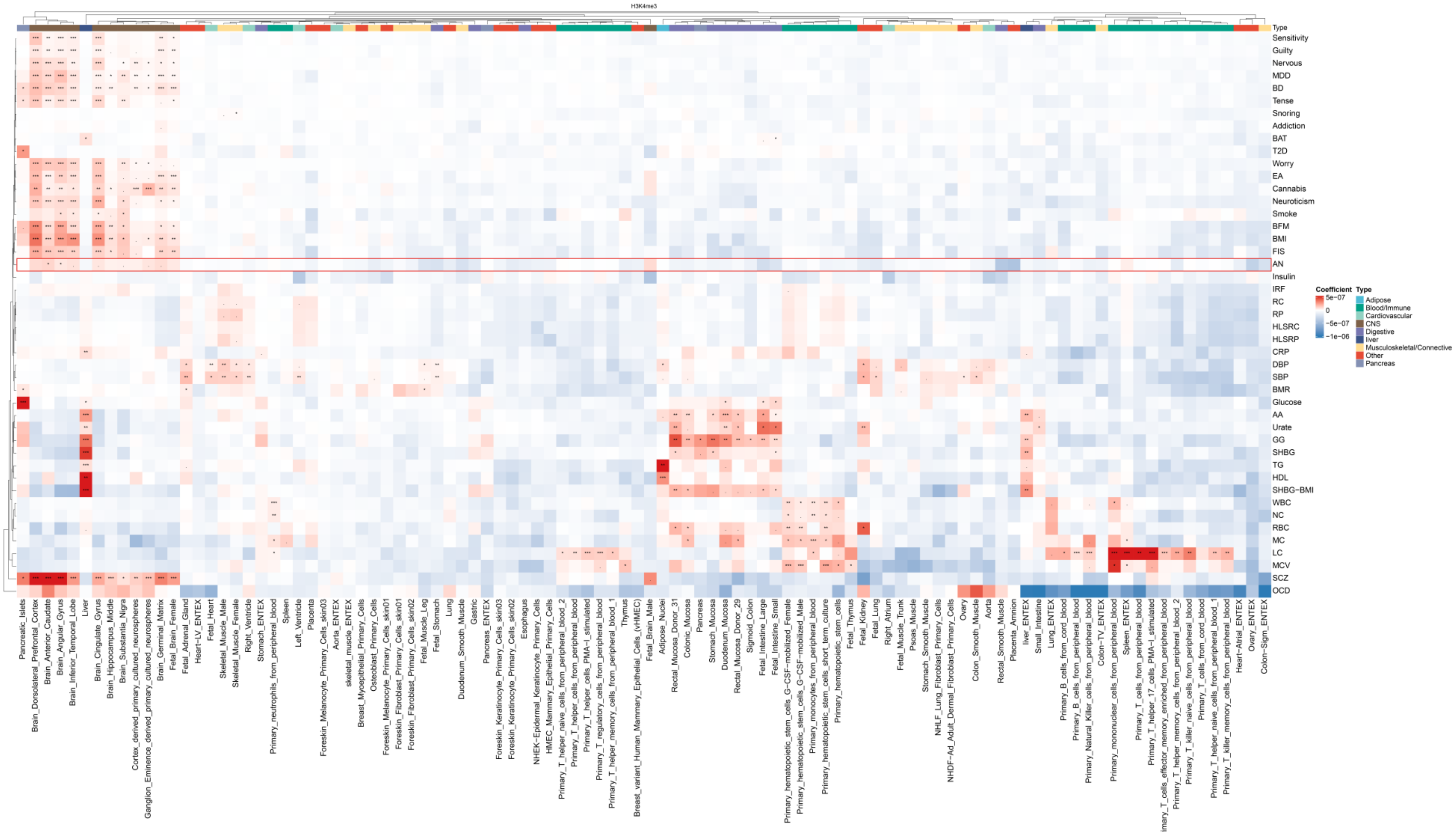
H3K4me3
Type
Sensitivity
Guilty
Nervous
MDD
BD
Tense
Snoring
Addiction
BAT
T2D
Worry
EA
Cannabis
Neuroticism
Smoke
BFM
BMI
FIS
AN
Insulin
IRF
RC
RP
HLSRC
HLSRP
CRP
DBP
SBP
BMR
Glucose
AA
Urate
GG
SHBG
TG
HDL
SHBG-BMI
WBC
NC
RBC
MC
LC
MCV
SCZ
OCD
Coefficient
5e-07
0
-5e-07
-1e-06
Type
Adipose
Blood/Immune
Cardiovascular
CNS
Digestive
liver
Musculoskeletal/Connective
Other
Pancreas

**f H3K9ac**

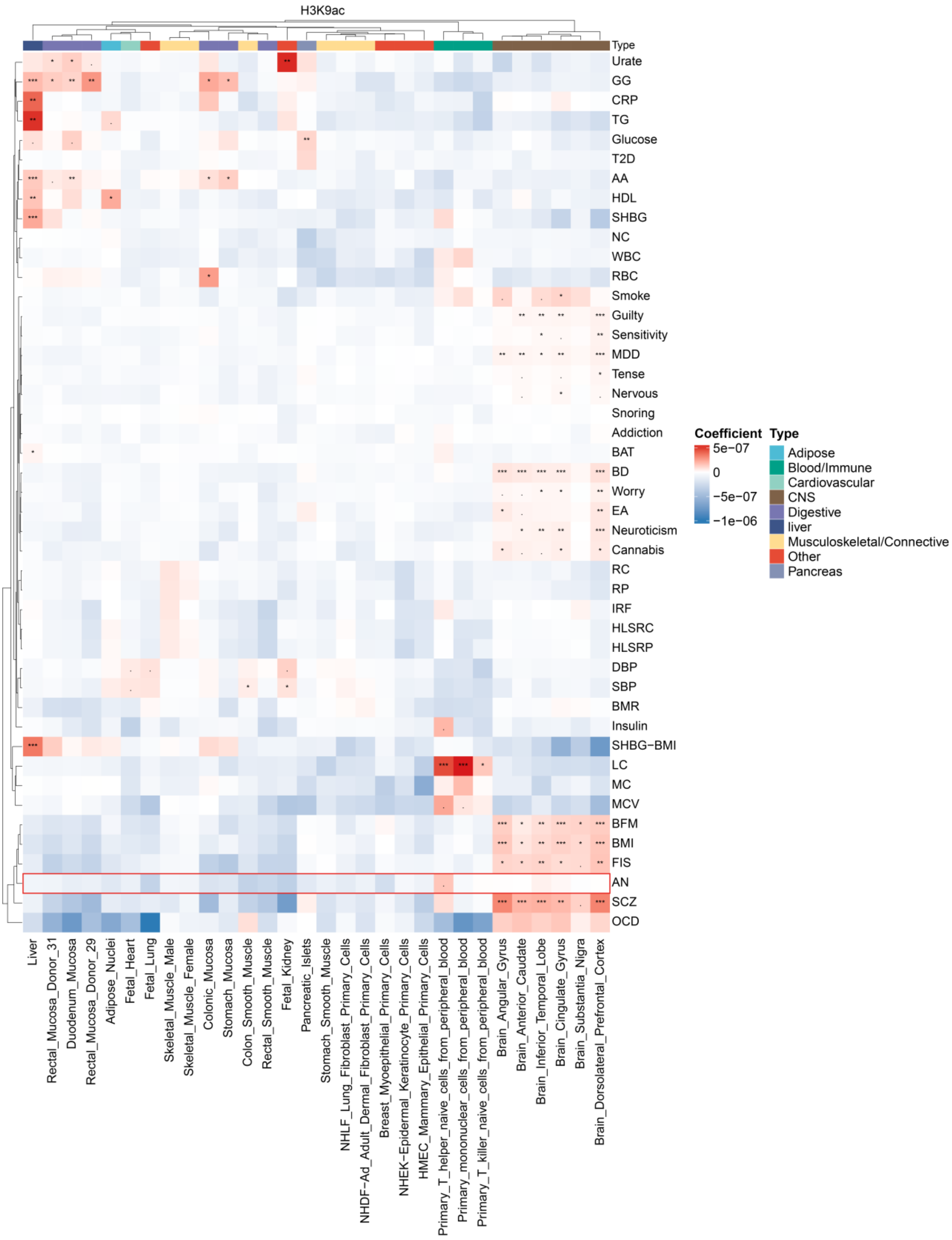
H3K9ac
Type
Urate
GG
CRP
TG
Glucose
T2D
AA
HDL
SHBG
NC
WBC
RBC
Smoke
Guilty
Sensitivity
MDD
Tense
Nervous
Snoring
Addiction
BAT
BD
Worry
EA
Neuroticism
Cannabis
RC
RP
IRF
HLSRC
HLSRP
DBP
SBP
BMR
Insulin
SHBG-BMI
LC
MC
MCV
BFM
BMI
FIS
AN
SCZ
OCD
Liver
Rectal_Mucosa_Donor_31
Duodenum_Mucosa
Rectal_Mucosa_Donor_29
Adipose_Nuclei
Fetal_Heart
Fetal_Lung
Skeletal_Muscle_Male
Skeletal_Muscle_Female
Colonic_Mucosa
Stomach_Mucosa
Colon_Smooth_Muscle
Rectal_Smooth_Muscle
Fetal_Kidney
Pancreatic_Islets
Stomach_Smooth_Muscle
NHLF_Lung_Fibroblast_Primary_Cells
NHDF-Ad_Adult_Dermal_Fibroblast_Primary_Cells
Breast_Myoepithelial_Primary_Cells
NHEK-Epidermal_Keratinocyte_Primary_Cells
HMEC_Mammary_Epithelial_Primary_Cells
Primary_T_helper_naive_cells_from_peripheral_blood
Primary_mononuclear_cells_from_peripheral_blood
Primary_T_killer_naive_cells_from_peripheral_blood
Brain_Angular_Gyrus
Brain_Anterior_Caudate
Brain_Inferior_Temporal_Lobe
Brain_Cingulate_Gyrus
Brain_Substantia_Nigra
Brain_Dorsolateral_Prefrontal_Cortex
Coefficient
5e-07
0
-5e-07
-1e-06
Type
Adipose
Blood/Immune
Cardiovascular
CNS
Digestive
liver
Musculoskeletal/Connective
Other
Pancreas

**Figure S9.** Manhattan plot of the GWAS multi-trait meta-analysis between anorexia nervosa and metabolic traits. The green circle shows the results of multi-trait meta-analysis between anorexia nervosa and basal metabolic rate; the orange circle shows the results of multi-trait meta-analysis between anorexia nervosa and BMI; the purple circle shows the results of multi-trait meta-analysis between anorexia nervosa and HDL cholesterol; the pink circle shows the results of multi-trait meta-analysis between anorexia nervosa and triglycerides; the light green circle shows the results of multi-trait meta-analysis between anorexia nervosa and whole body fat mass.

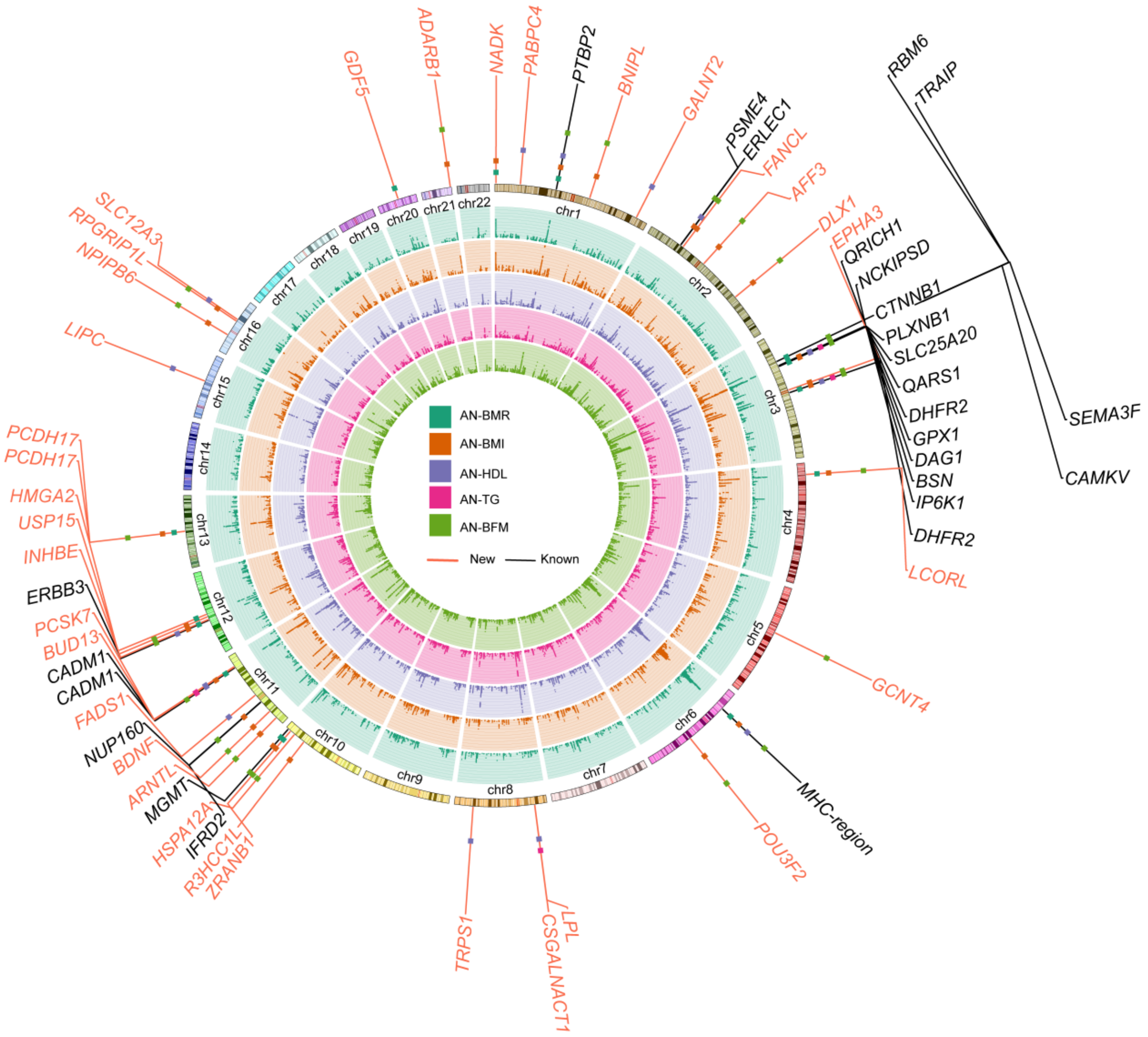

**Figure S10.** Manhattan plot of the GWAS multi-trait meta-analysis between anorexia nervosa and biochemical markers. The green circle shows the results of multi-trait meta-analysis between anorexia nervosa and sex hormone-binding globulin levels; the orange circle shows the results of multi-trait meta-analysis between anorexia nervosa and reticulocyte count; the purple circle shows the results of multi-trait meta-analysis between anorexia nervosa and immature reticulocyte fraction; the pink circle shows the results of multi-trait meta-analysis between anorexia nervosa and high light scatter reticulocyte count.

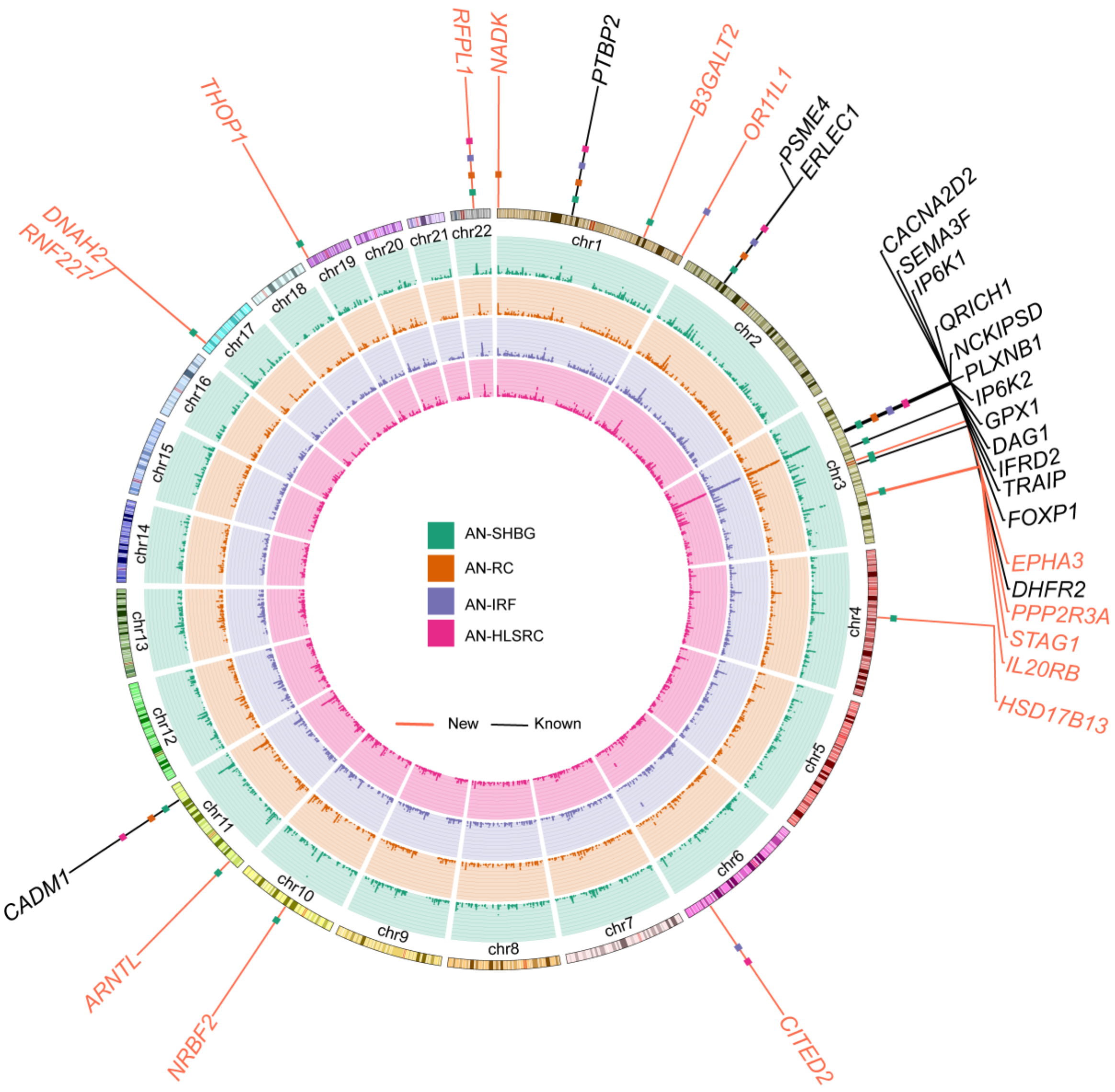
